# Quantifying the contribution of policy measures to COVID-19 dynamics across 126 countries

**DOI:** 10.64898/2026.11.05.26367213

**Authors:** James M Trauer, David S Shipman, Matthew Ryan, Alec S Henderson, Emma S McBryde, Michael T Meehan, Romain Ragonnet

## Abstract

COVID-19 provides an unprecedented opportunity to evaluate the contribution of public health and social measures (PHSMs) to controlling transmission and inform responses to future emerging respiratory pathogens. We analysed SARS-CoV-2 transmission across 126 countries spanning multiple epidemic waves and diverse policy environments during periods when PHSMs were the primary interventions available to influence the epidemic. By explicitly representing key epidemiological drivers of transmission and measuring reductions in residual transmission scaling after incorporating OxCGRT policy time-series data, we isolated the effect of PHSMs on transmission. PHSMs frequently explained a substantial proportion of the observed variation in transmission intensity, with particularly strong effects seen in many high-income countries. In countries with strong evidence for policy effects, reductions in transmission reached 60-80% or above, although the individual policies responsible for these effects varied considerably. Harmonised analyses considering the effect of community mobility showed comparably strong effects in a similar subset of countries.

## Introduction

Preparedness against emerging respiratory pathogens remains a critical global challenge, as increasing environmental disruption and global connectedness create opportunities for pathogen emergence and spread.^1,2^ The COVID-19 pandemic provides the most important recent test of public health responses to such threats and so presents an unprecedented opportunity to evaluate the interventions used to control transmission.^3^ While the effect of medical countermeasures such as vaccination can be more easily quantified through direct clinical investigations, understanding the effect of public health and social measures (PHSMs) must be largely inferred from epidemiological analyses.^4–6^ As such, the scale, duration and geographic breadth of the PHSMs implemented during the COVID-19 pandemic provide an unparalleled opportunity to evaluate their contribution to controlling transmission and inform future responses to emerging respiratory pathogens.

Previous studies have generally concluded that PHSMs contributed to reductions in SARS-CoV-2 transmission, but have differed regarding the estimated magnitude of these effects and the relative importance of specific interventions.^5,7–12^ These studies have variously identified stay-at-home orders, gathering restrictions, school closures and business restrictions as among the most effective measures, despite the inconsistency in the associated effect estimates. These discrepancies may reflect differences in study settings, analysis periods, intervention packages and methodological approaches, including sensitivity to model specification.^13^ Estimating policy effects from epidemic time series remains challenging because interventions are typically implemented in combination, vary across settings and become increasingly difficult to distinguish once medical countermeasures such as vaccination are introduced.^14,15^ Moreover, most previous analyses have been restricted to specific regions, subsets of countries or particular pandemic phases. As a result, analyses that systematically integrate data across diverse geographic settings and throughout the period during which PHSMs constituted the primary response to the pandemic should provide a stronger basis for understanding the contribution of policy measures to transmission dynamics.

To address this challenge, we analysed SARS-CoV-2 transmission across 126 countries, encompassing multiple epidemic waves and diverse policy environments. We developed a mechanistic framework to quantify the explanatory value of policy measures through reductions in residual transmission scaling when time series from the Oxford COVID-19 Government Response Tracker were incorporated into the analysis.^16^ Applying this approach across all countries for which robust inference was possible, we present a globally comprehensive assessment of the extent to which PHSMs influenced transmission dynamics over the course of the pandemic.

## Results

We calibrated a renewal model with and without accounting for time series of policy changes to the COVID-19 epidemics of 126 countries. By examining the extent to which including policy effects in the analysis reduced the need for residual non-mechanistic scaling of transmission, we quantified the evidence that policy changes influenced transmission in these settings. The model incorporated epidemiological features including susceptible depletion,^17^ variant-specific transmissibility,^18^ incomplete immunity to emerging variants^19^ and partial case ascertainment,^20^ and we calibrated it to COVID-19 epidemiological targets that included time series profiles for weekly new deaths, case notifications, up to one hierarchically selected hospitalisation indicator and the proportional prevalence of key variants of concern. For the baseline model, our methods matched those of our previous publication considering the effects of population mobility on transmission and focused on epidemiological contexts with low pre-existing natural immunity, including a later post-vaccination analysis window for Oceania and Singapore.^21^ In the current analysis, we incorporated scaling for the effects of policy measures obtained from the Oxford COVID-19 Government Response Tracker (OxCGRT) database, which provides time series of severity for each of several policy indicators on ordinal scales.^16,22^ We achieved close matches to all calibration targets for each country under six approaches to scaling transmission: a baseline configuration with no scaling of transmission for policy effects, two alternative approaches to scaling for policy measures and three approaches to scaling for mobility inputs (Supplemental Files 2 and 3).

## Effect of policies on transmission

### Policy time series compared to baseline residual transmission scaling

To motivate more formal analyses, we considered whether the OxCGRT time series appeared to track with the residual non-mechanistic scaling of transmission (henceforward “residual scaling”) under the baseline model configuration that did not incorporate policy effects. OxCGRT provides recommended sets of policies that can be combined to create overall indices of government policies, including the Containment and Stringency indices, and we constructed our own custom index for further analyses. Supplemental File 4 and Figure 2 show comparisons of each of these three indices against baseline residual scaling using unweighted averages across policy domains (i.e. 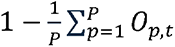, where *0_p,t_* represents the value of policy *p* at time *t* divided by its maximum value). Although some countries showed little correspondence between variation in transmission and these composite policy time series (third column Figure 2), in many countries variation in transmission corresponded closely with the temporal trends of these indices (first two columns).

**Figure 1.**
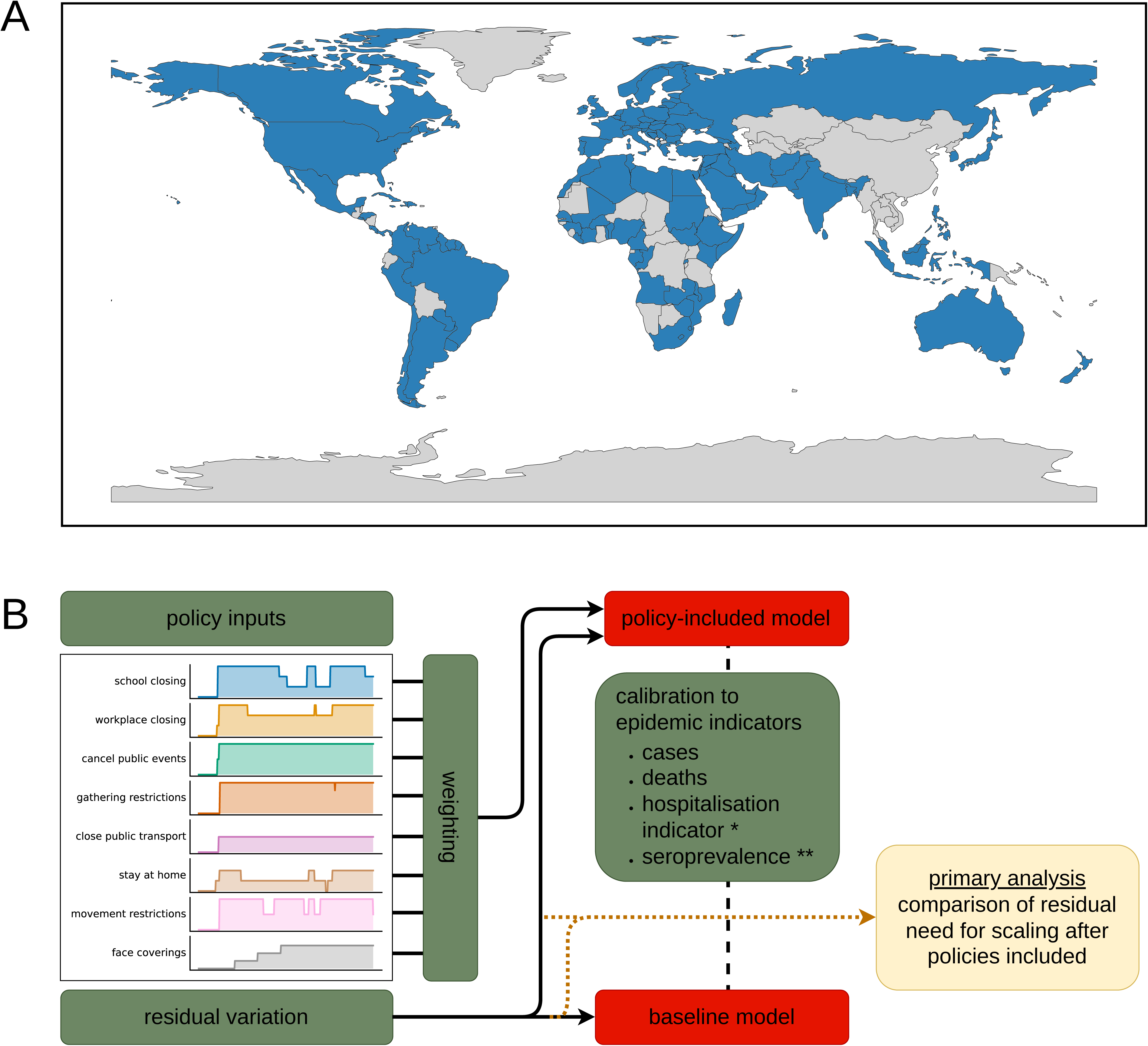
Illustration of study methods. Panel A, country inclusion. Included countries shaded in blue, excluded countries shaded in grey. Panel B, schematic of approach to inference. Data/parameter inputs coloured in green. Candidate model configurations coloured in red (although two different approaches to the inclusion of OxCGRT data were used for the “policy-included model”, along with three approaches to including mobility data). Primary comparison for inference coloured in yellow with dotted arrow indicating main target for comparison. *Only one hospitalisation indicator used for each country, through a hierarchical selection algorithm. **Seroprevalence targets used where available, with lower weight assigned to this indicator compared to the time series indicators listed in the preceding bullets.

**Figure 2.**
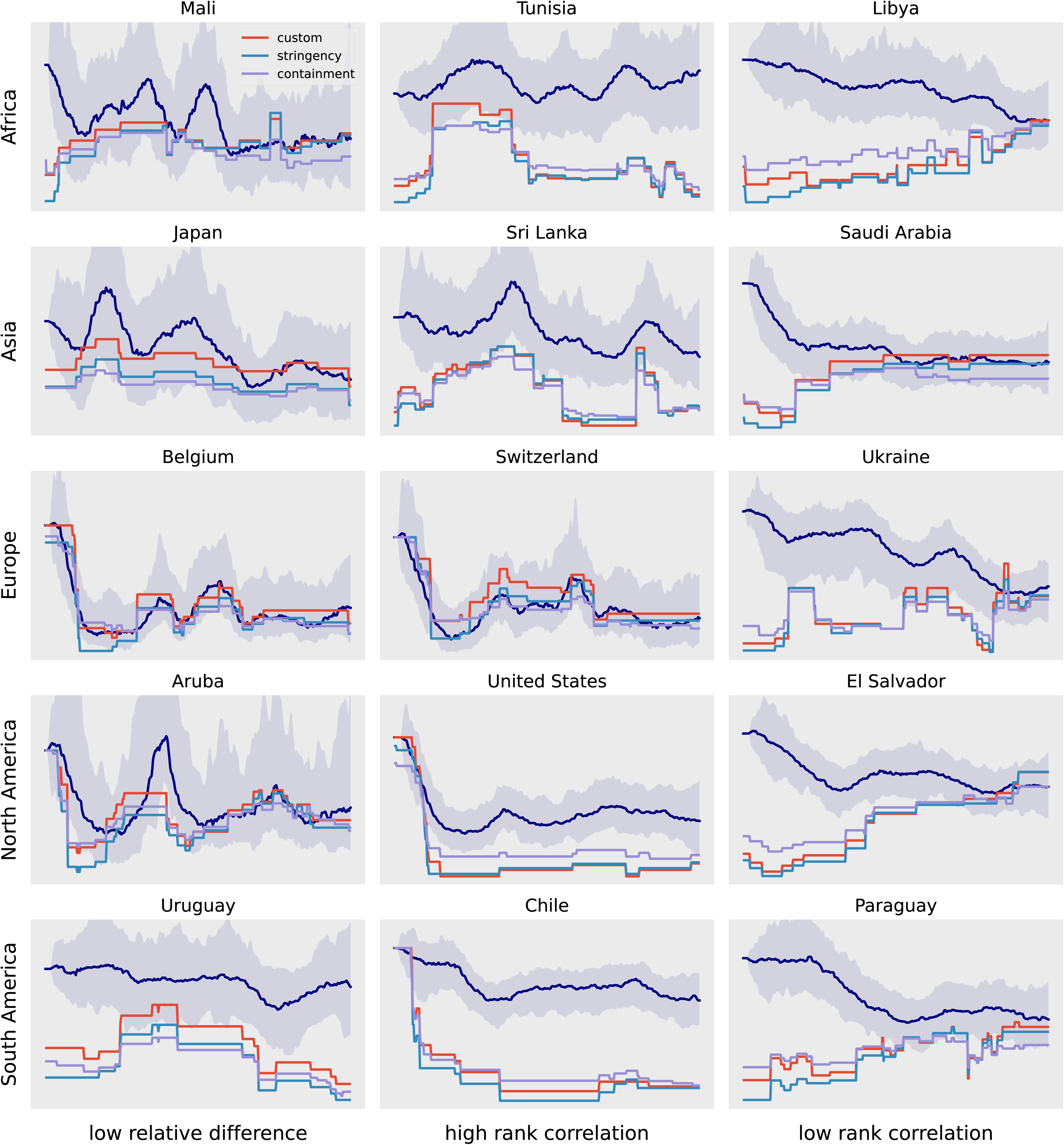
Comparison between the average OxCGRT policy inputs and residual transmission scaling under the baseline model configuration (without scaling for policy changes). OxCGRT series are plotted as an unweighted average applied to the normalised policy indicators. “Stringency” and “containment” policies are the subsets of the OxCGRT policies reported that are recommended by OxCGRT. “Custom” policies are our chosen subset of policies, used throughout our subsequent analyses. Median residual scaling (navy line) and 95% credible interval (navy shaded area) presented. Rows show representative countries from each continent except Oceania. Columns illustrate the range of agreement between the median residual scaling function and the custom-policy average time series. Column 1: lowest mean relative difference over time between residual scaling and the custom policy scaling curve. Columns 2 and 3: highest and lowest Spearman rank correlation between those two series. Y-axis labels shown in full figures in Supplemental File 4; all residual scaling curves start from a value of one.

### Comparison of weighted policy time series to residual transmission scaling

Our primary model configuration with policy effects included (“OxCGRT floored”) incorporated weights for the contribution of each constituent policy to scaling, as well as a floor parameter for the maximum effect of policy changes and an exponential scaling parameter to map from the policy time series to the effect on transmission. Supplemental File 6 and Figure 3 show the comparison between the residual transmission scaling (the same output as shown in Figure 2) and the calibrated policy scaling function after fitting to data. This approach generated functions that closely matched residual transmission scaling in several settings (e.g. Belgium, Aruba), suggesting that much of the unexplained variation in transmission captured by the residual process could alternatively be reproduced using policy information alone.

**Figure 3.**
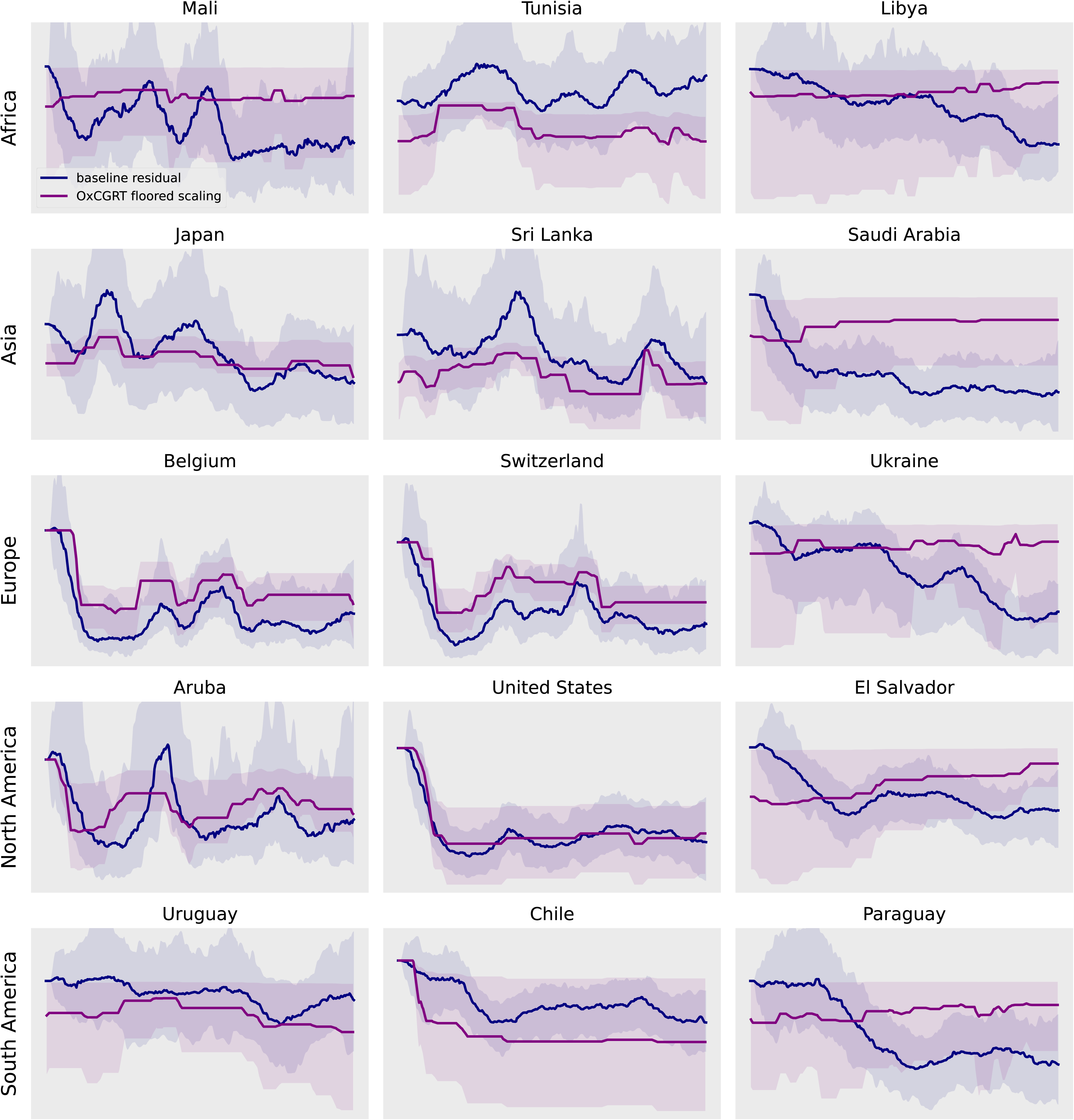
Comparison of weighted OxCGRT scaling function against residual transmission scaling under the baseline model configuration (without scaling for policy changes). Individual policy weights obtained through calibration to epidemic data. Median scaling value (line) and 95% credible interval (shaded area) presented for baseline (navy) and OxCGRT analysis (purple). Same countries selected as for Figure 2. Y-axis labels shown in full figures in Supplemental File 6; all residual scaling curves start from a value of one.

### Reduction in residual scaling with the inclusion of policy inputs

Supplemental File 6 and Figure 4 show the remaining residual scaling required after policy effects were included in the model. If policy measures explain transmission dynamics, inclusion of policy scaling should reduce the residual need to adjust transmission. Consistent with this expectation, the residual process became substantially flatter in most countries, with fewer abrupt fluctuations than under the baseline configuration.

**Figure 4.**
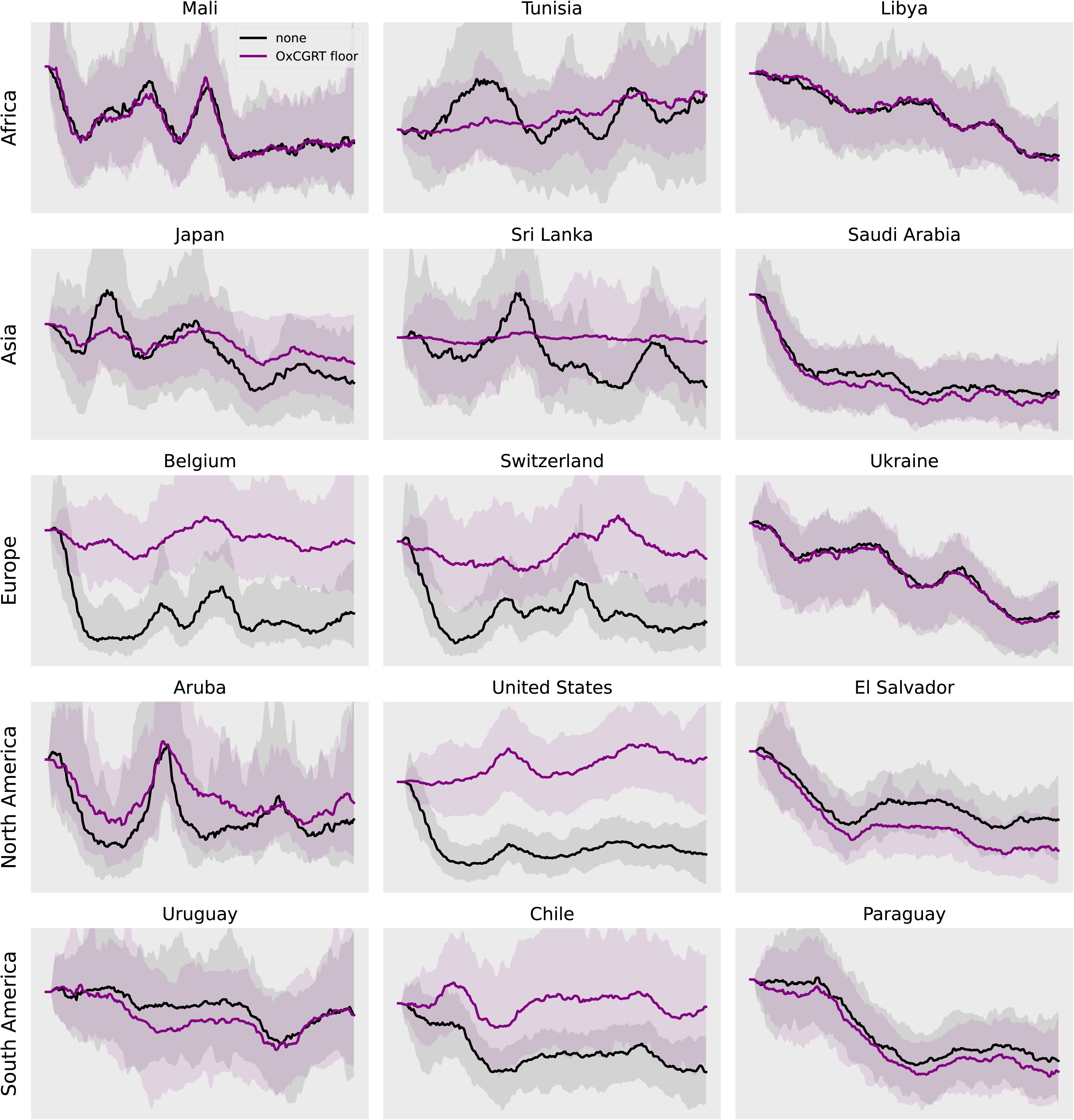
Comparison of residual scaling functions with (purple) and without (black) OxCGRT policy scaling. Median scaling value (line) and 95% credible interval (shaded area) presented. Same countries selected as for Figure 2.

### Evidence of an association between policies and transmission

We estimated the effect of policy changes on transmission by quantifying the reduction in variation in the residual process under the model configuration with scaling for policy effects compared to the baseline configuration without policy scaling. To quantify this formally, we considered the dispersion parameter that governs the prior distributions for each update to the residual process compared to the preceding value. This parameter was regularised by assigning a prior distribution with mode at zero to encourage the model to accept the minimum extent of residual scaling achievable while still matching the calibration targets. Supplemental File 5 and the right-hand panels of Figure 5 show example posterior distributions for countries for which this dispersion parameter reduced substantially (e.g. Norway), showed intermediate reduction (e.g. Estonia) and for which there was little or no reduction (e.g. Angola). Because this parameter quantifies the magnitude of the residual adjustments required to fit observed epidemiological data, reductions in its posterior value indicate that policy data explain variation in transmission that would otherwise need to be captured by the residual process. We therefore interpreted the proportion of calibration iterations under which this quantity reduced as the strength of evidence for an association between policies and transmission.

**Figure 5.**
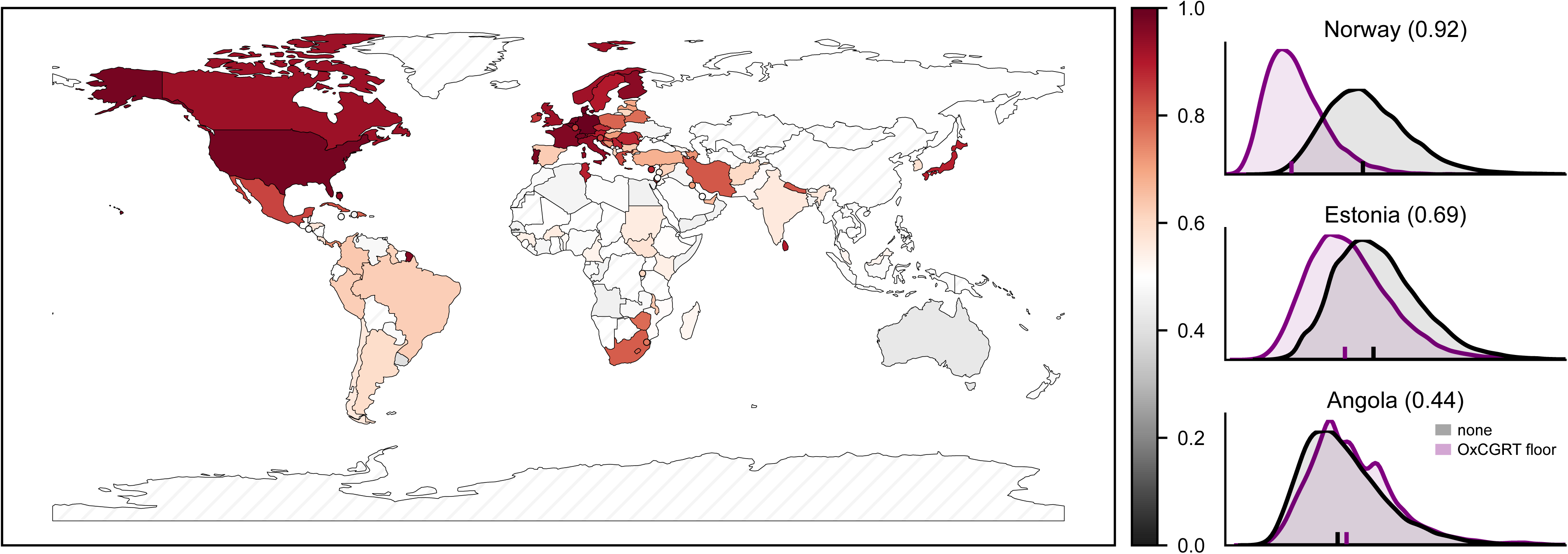
Strength of evidence for an effect of policies on transmission by country, as quantified by the proportion of residual process dispersion parameter values that were greater under the policy scaling analysis than under the baseline analysis. Left panel, countries shaded according to the median ratio of the dispersion parameter without scaling to its value with policy scaling included. Hatched countries not analysed. Right three panels show three representative countries (randomly selected from each ratio tertile) in which there was a largest, intermediate and lowest reduction in the dispersion parameter when scaling was introduced. Median ratio of the residual dispersion value without compared to with scaling is indicated after the country name annotation.

The left-hand panel of Figure 5 presents the results for all countries, showing strong evidence for an effect of policy changes in many high-income countries of Europe and North America, intermediate evidence in several countries of Eastern Europe, South America and Southern Africa, and minimal or no evidence for an effect in many countries of the rest of Africa and Oceania. Among the 52 high-income countries analysed, 29 showed a marked reduction in residual scaling (dispersion ratio reduced in 75% of runs or more), which was not seen in any of the 15 low-income countries. The strongest evidence was observed predominantly in European countries, including Poland, Greece, Slovenia, Luxembourg, Ireland, Czechia, Croatia, Romania, Austria, Sweden, Norway, Andorra, United Kingdom, Italy, Finland, France, Portugal, Switzerland, Netherlands, Denmark, Germany, Belgium, as well as Aruba, Israel, Cyprus, Japan, Canada, Bahamas, United States. However, substantial evidence was also identified in several geographically heterogeneous middle-income countries, (e.g. Eswatini, Belarus, Zimbabwe, Dominican Republic, Lesotho, Kosovo, South Africa, Iran, Nepal, Cuba, Moldova, Mexico, Tunisia, Serbia, Sri Lanka), possibly reflecting greater capacity to characterise transmission dynamics through routine surveillance in some of these countries. We found no evidence for an effect of policy changes in countries for which our analysis spanned the post-vaccination roll-out Omicron period (Australia, New Zealand and Singapore), rather than the earlier pre-vaccination period considered in most of our analyses.

In countries for which there was little evidence for an effect of policy-related scaling of transmission, our alternative policy configuration (OxCGRT independent) performed less well than OxCGRT floored, because its priors were less able to support the absence of an effect.

### Inferred strength of the combined policy package on mitigating transmission

We next quantified the maximum transmission reduction implied by the calibrated policy scaling function, 1 − *f^m^*. Although simultaneous implementation of all policies with the custom suite did not often occur during our analysis windows, we used this theoretical maximum scaling of transmission as our primary metric of the effectiveness of the overall policy package implemented in each country. Figure 6 shows this estimate for the strength of policy effects across all countries analysed. Because this metric is only meaningful when evidence for policy effects was detected, countries without such evidence are displayed in grey. The strongest policy package effects were typically seen in high-income countries of Western and Northern Europe and North America, Japan and some countries of Southern Africa. Intermediate effects were seen in some other African countries and several countries of South America and South Asia. For several of the remaining African countries, as well as Oceania (for which the analysis period and methods differed), no evidence of an effect of policies on transmission was found.

**Figure 6.**
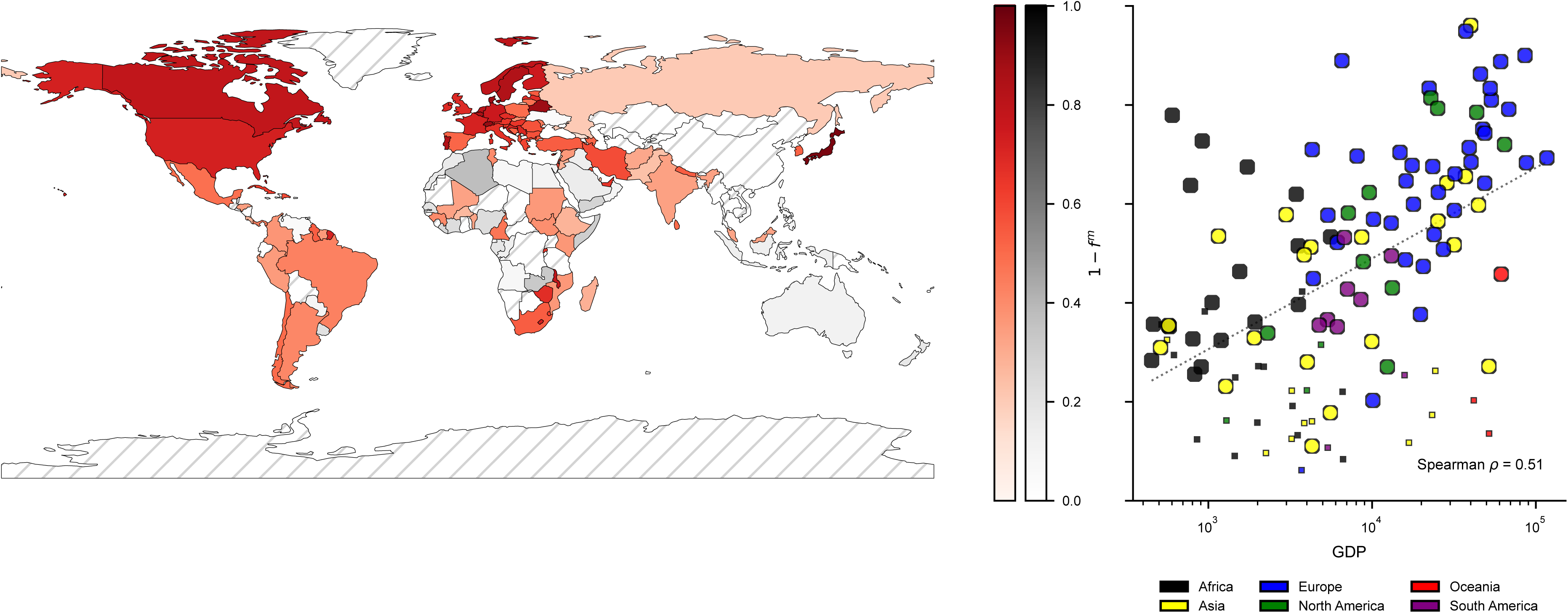
Left panel: policy strength estimates (1 − *f^m^*) by country. Results presented in red for countries for which the majority of model calibration iterations found evidence for policy effects (i.e. a reduction in the dispersion parameter), and presented in grey for countries where this was not the case. Hatched countries not analysed. Right panel: correlation between GDP per capita (2020 US dollars) and policy strength estimates. The vertical axis is on the same scale as the colour bars. Larger circles show countries with evidence of an effect for policies (>50% of calibration runs associated with a reduced residual dispersion parameter), whereas smaller squares show countries for which no such evidence was found. Dotted line shows linear fit between policy strength to natural logarithm of GDP for all countries analysed. GDP; gross domestic product.

### Comparison to mobility analyses

To compare the effect of policy scaling to the effect of scaling for population mobility time series available from Google and Facebook, we adapted our previous mobility analyses for consistency with the policy analyses which are the primary objective of this manuscript. We harmonised the scaling function for the mobility analyses with the function used for the policy analyses to include a “floor” parameter on the maximum reduction in the scaling process prior to the exponential transformation of the scaling function, whereas this floor was not incorporated in our previous mobility-focused analyses. Figure 7 shows that results for the policy and mobility analyses were strongly correlated, with strong effects of policy changes found in a similar group of countries as for those in which strong effects of mobility were observed. Figure 8 compares the floor values between the policy and mobility analyses for the countries in which we found evidence of an effect under both of these analysis configurations. For 39 of 50 countries under the Google comparison and for 36 of 52 under the Facebook comparison, the floor value for the mobility analysis was lower than for the policy analysis (by a median 8.5% and 7.2% respectively for the two analyses). These findings suggest that mobility measures often captured a greater proportion of transmission-related behavioural change than policy measures alone.

**Figure 7.**
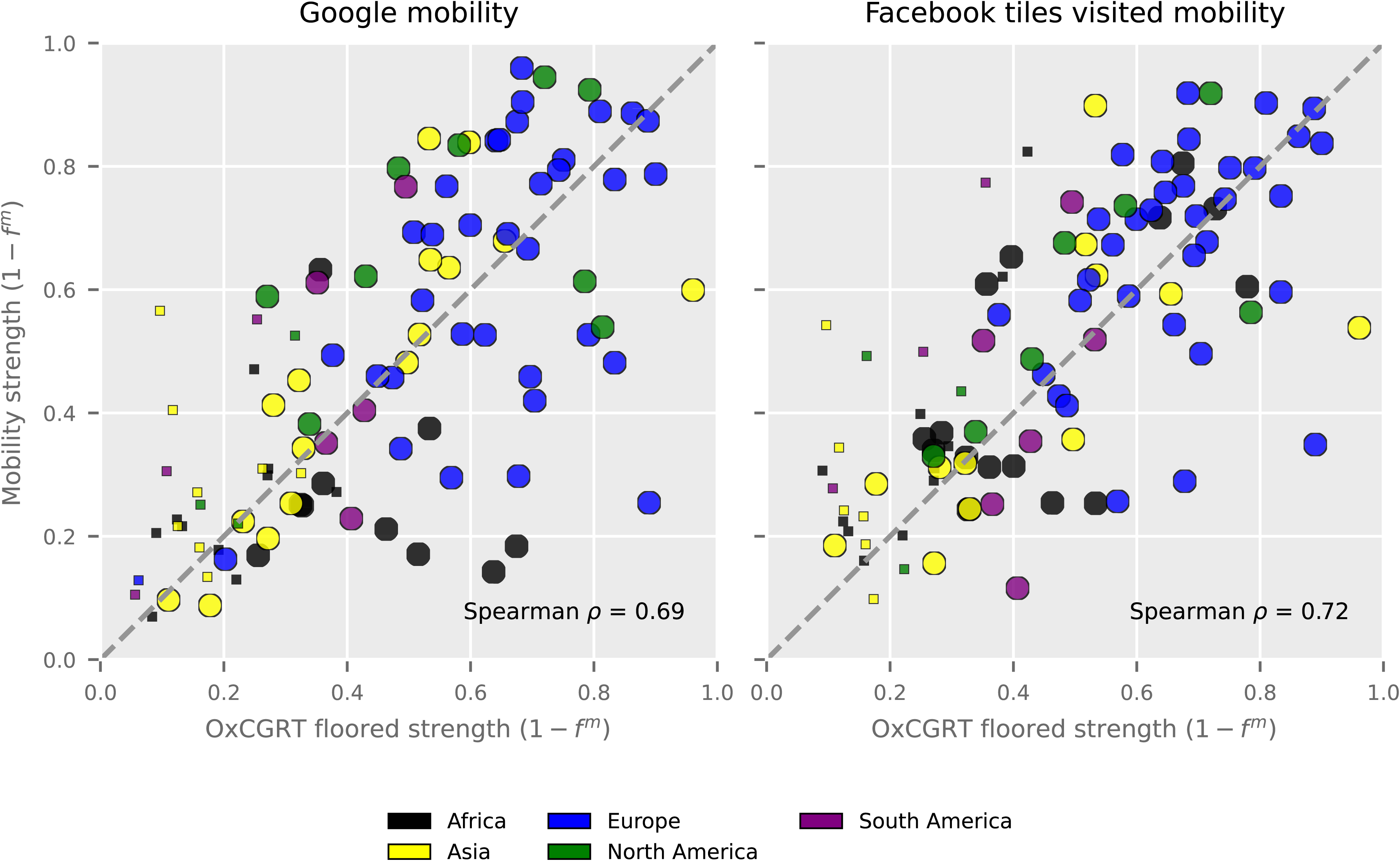
Median policy strength estimate (1 − *f^m^*) plotted against median mobility strength estimate by country. Colours indicate continent of origin. Results presented as circles for countries for which the majority of model calibration iterations found evidence for policy effects (i.e. a reduction in the dispersion parameter), and presented as small squares for countries where this was not the case. Mobility analyses not performed for Oceania.

**Figure 8.**
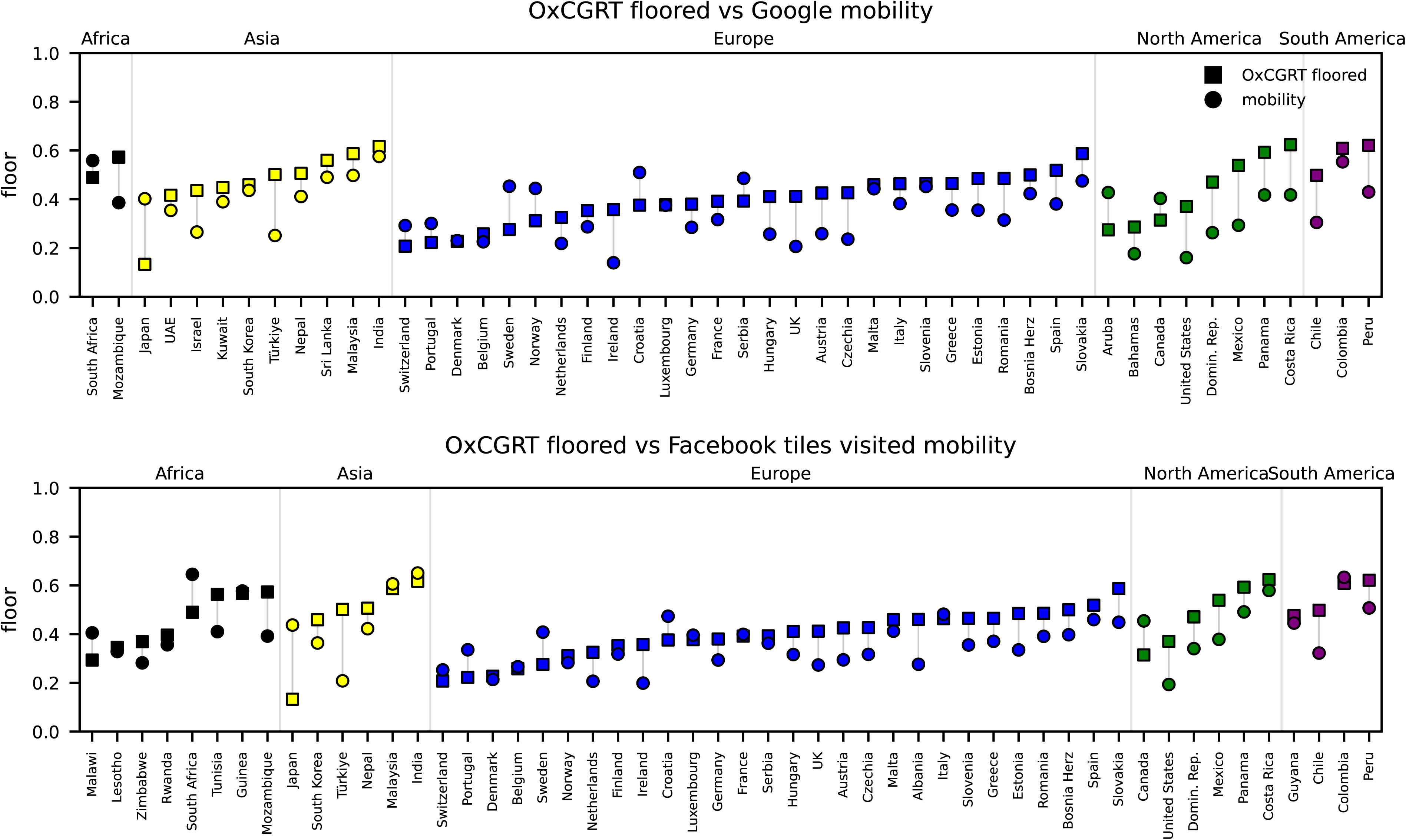
Comparison of median estimate of the floor parameter (*f*) under the policy analysis (squares) to the median estimate for the same parameter under each of two mobility analyses (circles). Countries arranged and coloured by continent. Only countries for which evidence of both policy and mobility effects are displayed.

### Specific policy indicators

Our main analysis (OxCGRT floored) approach assigned individual weights to each of the eight policies of our custom index, which were calibrated to create an overall weighted time series for the effect of policies on scaling transmission. In many countries, specific policies were markedly upweighted by the calibration algorithm, such that the individual policy was assigned greater influence on the overall scaling function. The posterior distributions of each policy weight for each country is shown in Supplemental File 10, with randomly selected country examples presented in Figure 9. The relative extent of up- or down-weighting is presented in Figure 10, with restriction to countries for which there was evidence for an effect of the overall policy package and for which at least one policy was significantly up- or down-weighted. No policy displayed consistently strong effects across all countries, suggesting substantial heterogeneity in policy effectiveness and implementation contexts. However, school closures often upweighted in countries of Europe and Asia, while movement restrictions were down-weighted in countries across several continents.

**Figure 9.**
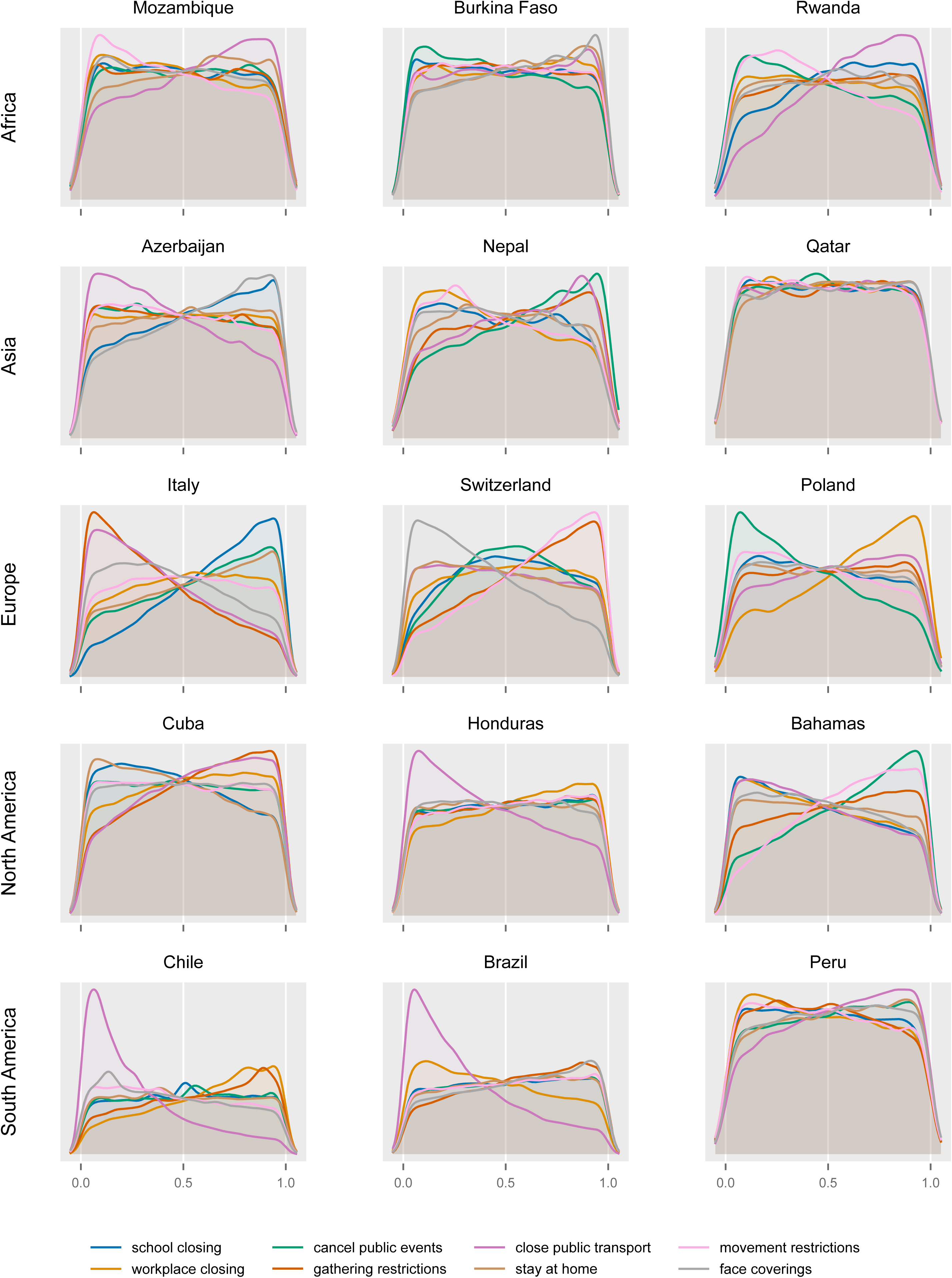
Posterior distributions of pre-normalisation policy weights for each policy for selected countries. Three countries randomly selected from each continent except Oceania (rows) from among all countries for which the majority of calibration iterations found evidence for policy effects.

**Figure 10.**
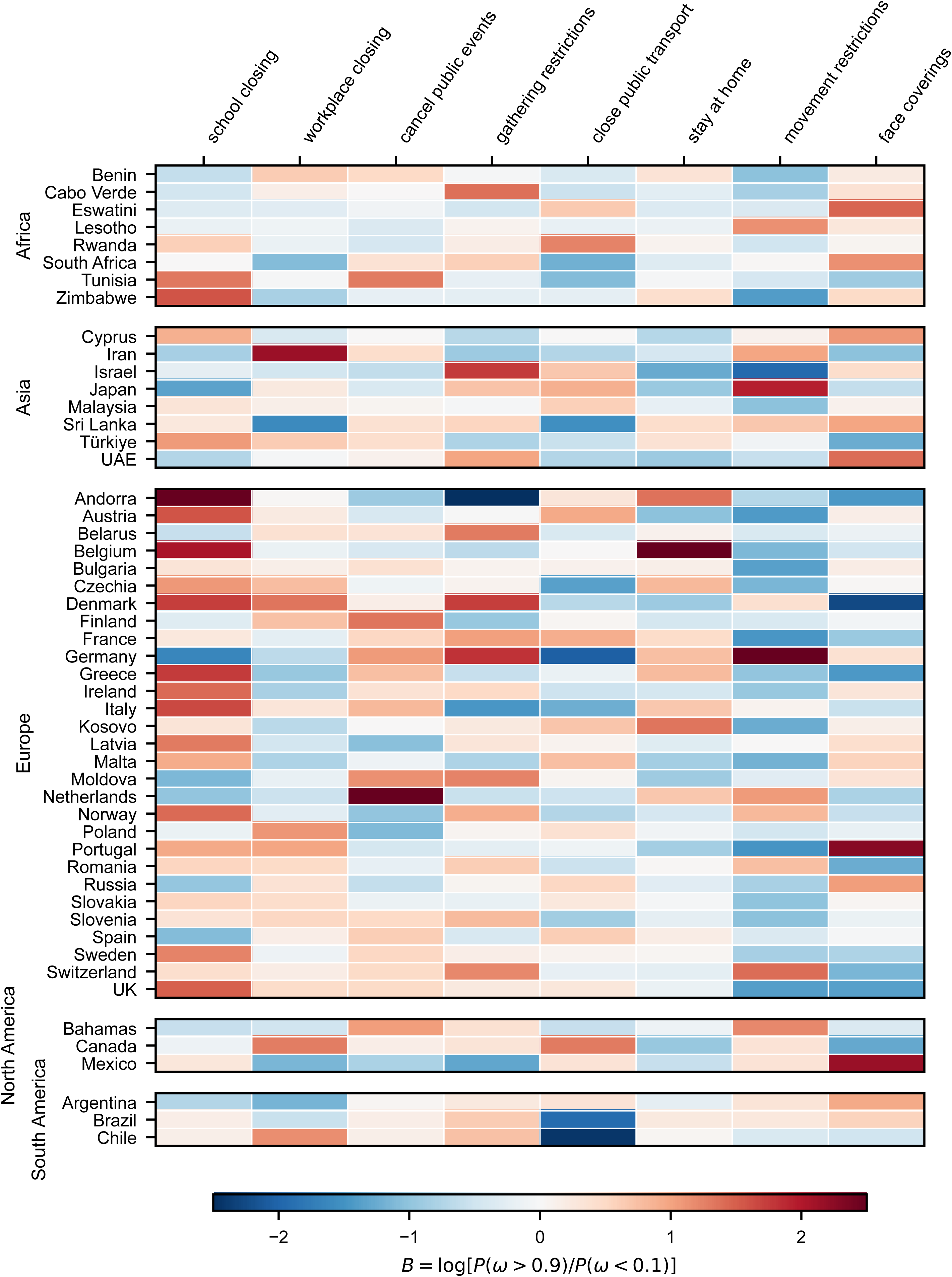
Relative up-weighting (red) and down-weighting (navy) of policies for selected countries (“B” value). All countries presented for which there was both evidence of an effect of policies (i.e. a reduction in the dispersion parameter) and at least one policy weight was up- or down-weighted such that |B|>1.

## Discussion

Across 126 countries spanning diverse policy environments and epidemic trajectories, policy stringency metrics provided substantial information regarding temporal variation in SARS-CoV-2 transmission intensity during periods when PHSMs were the primary pandemic response. Incorporating policy data frequently reduced the model’s need for residual transmission scaling, indicating that policy measures explained a substantial proportion of the variation in transmission beyond what was captured by the established epidemiological drivers we explicitly modelled. Evidence for policy effects was strongest in many high-income countries of Europe and North America, although the magnitude of these effects and the policies most clearly associated with transmission varied considerably between settings. Policy effects were easier to identify when interventions were analysed as an integrated package of measures rather than as independent effects, and the strength of their effects was often comparable to that observed for population mobility under equivalent analyses for the same countries.

Evidence for policy effects was strongly structured by economic setting and region. Excluding the distinct epidemiological context of Oceania, policy signals were clearly identifiable across many high-income European and North American settings, but were largely absent in several low-income countries, particularly in Africa. This inability to detect an effect mirrored our findings for mobility metrics, suggesting that failure to translate policy recommendations into population behavioural changes was unlikely to be the primary explanation. Because the surveillance data typically provided clear epidemic trajectories despite lower case ascertainment estimates, non-detection is more likely to reflect misalignment between formal policy databases and on-the-ground implementation, rather than limitations in estimating transmission intensity itself.

Where policy effects were identifiable, they were often large. In many high-income countries, particularly in Europe, the estimated maximum policy effect exceeded 0.6 and often reached 0.8 or above. Under our modelling framework, this implies that the full package of non-pharmaceutical interventions represented in the OxCGRT database could reduce transmission by 60-80% or more when implemented simultaneously at maximum intensity. These estimates reflect the maximum potential weighted effect of the intervention packages, rather than the independent effect of individual measures. While stronger policy effects were moderately associated with higher gross domestic product and some geographic clustering was observed, these relationships were not homogeneous. This suggests that local country-specific factors, such as differences in background contact structures, policy implementation, community uptake and capacity to adhere to recommendations, played an important role in determining policy effectiveness.

Countries with strong evidence for policy effects generally also showed strong mobility effects. This was particularly evident when mobility was measured using the Facebook metric describing the number of 600 m × 600 m tiles visited, although similar patterns were observed using Google’s location-based mobility indicators. This concordance is intuitive, given that many of the non-pharmaceutical interventions included in our analysis are expected to influence transmission primarily through reductions in population movement and social contact patterns.^23^ Despite this alignment, the estimated transmission floor parameter (*f*) was on average 12% lower under mobility models than policy models. This could plausibly reflect the theoretical bound of mobility data – where near-zero movement would conceptually imply a near absence of transmission – whereas policy models achieve equivalent observed transmission reductions by steepening the scaling response (via the exponent mapping parameter *m*) while maintaining a higher baseline floor of contact.

A further notable finding was that specific interventions were often identified as important contributors to transmission control within a given country, although the specific policies selected varied markedly between settings. This heterogeneity implies that the apparent effectiveness of individual interventions was strongly influenced by the local context and implementation approach, rather than supporting a single set of universally effective policies. Policy indicators were highly correlated on principal component analysis of the raw policy time series values, with the first principal component explaining a median of 58% of temporal variation across countries. This limited the extent to which independent policy effects could be disentangled and reflects the fact that interventions were typically implemented as part of broader policy responses rather than in isolation. We therefore placed greater emphasis on results from the policy-package formulation (OxCGRT floored), which is more consistent with these implementation patterns. Furthermore, the package formulation allowed the overall policy effect to approach zero, whereas the independent policy formulation effectively required all policy effects to approach zero simultaneously to infer no effect. Consequently, our findings provide stronger evidence that changes in the overall policy response influenced transmission than for a single set of intervention-specific effects that generalises across settings.

Early studies of the effects of PHSMs inferred variation in transmission from the first epidemic wave of SARS-CoV-2 and were influential in guiding the initial response to COVID-19.^5,7–10^ Our analysis advances from these works by analysing how PHSMs dynamically influenced multiple waves of transmission that were driven by heterogeneous variants of SARS-CoV-2 and span repeated cycles of policy implementation and relaxation. A study of European countries spanning the first two waves of COVID-19 found effects of PHSMs often declined over these phases of the epidemic, supporting our finding that these effects were not constant over time and depended on the local epidemiological and policy context.^11^ These previous studies frequently estimated substantial effects of PHSMs but arrived at differing conclusions regarding the effectiveness of individual measures. Our findings suggest that this variation may reflect both differences in the ability of available data to capture policy-related effects on transmission across settings and that policy packages are more readily identifiable than the effects of individual measures. Consistent with several past studies we found that mobility and transmission often varied in concert, and further show that the countries with strongest effects of mobility and of policies often coincided.^21,24,25^

Limitations of our study include that the OxCGRT input data pertain to policy recommendations rather than compliance,^26^ which we mitigated by comparing to equivalent analyses of the effects of mobility. Our national-level framework did not capture subnational policy variation, which may be substantial in large or federated countries,^27,28^ though we sought to mitigate this by analysing periods of widespread community transmission rather than localised outbreak responses. Because many interventions were implemented concurrently, our primary analysis considered policies as packages, and we did not find consistent evidence for the independent effects of specific interventions across all countries. Further, the policies we considered were largely restricted to face-covering requirements and measures expected to influence attendance at workplaces, schools and other community venues. Other behaviours that may influence transmission, including voluntary behavioural responses to changing epidemic conditions, were not explicitly represented and so may contribute to residual unexplained variation.^29^

Policy measures substantially influenced SARS-CoV-2 transmission across many countries during periods in which PHSMs formed the primary public health response. Using a framework based on the reduction in residual transmission scaling required to fit epidemiological data, we identified strong evidence that policy measures explained variation in transmission beyond that captured by established epidemiological drivers. The close correspondence between policy- and mobility-based analyses suggests that much of this effect was mediated through changes in population behaviour, although behavioural influences on transmission appear to extend beyond those directly attributable to formal policy measures. While the magnitude of policy effects varied considerably between settings, our findings demonstrate that policy signals remained detectable across multiple epidemic waves, SARS-CoV-2 variants and epidemiological contexts.

## Methods

### Epidemiological model

Our full methods are presented in Supplemental File 1 and summarised as follows. We used a renewal model with daily time innovations and a gamma-distributed generation interval to simulate the COVID-19 epidemic of 126 countries independently. The model incorporated key epidemiological features, including susceptible depletion, multiple SARS-CoV-2 variants of varying transmissibility and severity, partial cross immunity following past infection with a different variant and incomplete case ascertainment. The number of new incident infections *i_p,v,t_* of variant *v* occurring at time *t* in the population with past infection history status *p*, is calculated as:

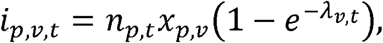

Where *n_p,t,_* represents the number of people with infection history status *p* at time *t* and *x_p,v,_* represents the susceptibility to infection of persons with infection history status *p* to infection with variant *v*. The variant-specific force of infection is calculated as:

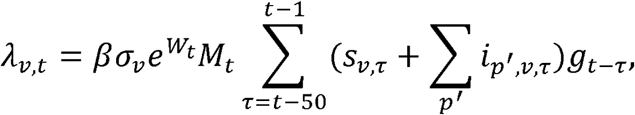

Where *β* represents the baseline transmissibility of SARS-CoV-2, while *σ_v_* captures the relative infectiousness of each variant. *e^W_t_^* is a Wiener process with weekly updates and *M_t_* is a policy scaling function, which is explained further below. *s_v,t_* is the seeding function for variant *v* and *g_t−τ_* is the gamma-distributed generation interval distribution.

### Scaling for policy effects

To incorporate policy effects, we constructed a time-varying transmission scaling function *M_t_* (Figure 1). Under our primary analysis configuration (“OxCGRT floored”), each policy indicator contributed to a combined policy intensity measure, which was then transformed using floor and exponent parameters to produce the final transmission scaling factor.

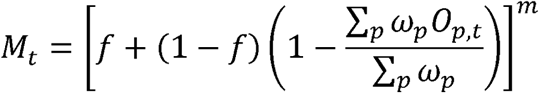

Where *0_p,t_* is the ratio of the policy time series from OxCGRT for the individual policy *p* at time *t* to its maximum ordinal value and *ω_p_* is a policy-specific calibrated weight (such that *0_p,t_* ∈ [0,1]). Our custom policy index comprised the following eight indicators: school closing, workplace closing, cancel public events, gathering restrictions, close public transport, stay at home, movement restrictions and face coverings (OxCGRT codes C1 to C7 and H6). This suite of policies is intended to represent policies that may influence local transmission, except that it excluded public information because this showed almost no variation during the analysis periods considered. As such, our weighted policy indicator provides a per-country modified stringency index, where individual policies contribute to the index based on their empirical influence on case numbers. This differs from the OxCGRT stringency index, which includes international travel and public information but not face coverings and calculates stringency as the simple average of all contributing components.

*f* and *m* are the floor and exponent parameters respectively, which are used to map from the weighted average of policy stringencies to the overall effect on transmission. We also considered an alternative configuration (“OxCGRT independent”) which considered each policy as having an independent effect rather than acting as part of a policy package, and is calculated as 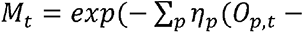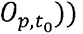, where *η_p_* is a calibrated parameter for the independent effect of policy *p* on transmission and the raw OxCGRT value is normalised to its starting value. Last, we also considered an equivalent analysis considering the effects of mobility instead of policies, but adopted a new approach to calculating *M_t_* from our previous work to align better with the policy scaling approach (incorporating floor and exponent parameters).

### Calibration

Gamma-distributed convolution functions were applied to the infection values calculated from the transmission model above to estimate weekly cases, hospitalisations and deaths, with the parameters to these distributions estimated during calibration from informative literature-based priors. We calculated our likelihood by comparing each of these model outputs against the corresponding calibration targets using a normal distribution in log space. We also fit to estimates of seroprevalence in our calibration algorithm where available to mildly constrain the total epidemic size, but assigned this target a lower weight in the calculation of our overall likelihood. We implemented the model using Google’s jax library and calibrated using numpyro’s NUTS algorithm.^30,31^

### Inference

The primary purpose of our analysis was to estimate the extent to which the need for residual scaling *e^W_t_^* declined with the inclusion of the policy scaling. We assessed policy effects by quantifying changes in the dispersion parameter governing the Wiener process *W_t_*. This parameter determines how freely the residual transmission scaling process can vary over time, with larger values permitting greater temporal fluctuations in residual scaling and smaller values implying less unexplained variation after accounting for mechanistically modelled drivers. We assigned this parameter a half-normal prior distribution with standard deviation 0.5 to penalise greater jumps to each successive value, and so discourage unnecessary non-mechanistic variation. As such, we considered the posterior distribution of the dispersion parameter to represent the strength of evidence for an effect of policies on transmission. Further, when all policies were implemented at their maximum level the transmission scaling function reduces to *f^m^*. As such, we took 1 − *f^m^* to represent the maximum possible policy-related effect on transmission under the floored analysis approach. We interpreted the floor parameter *f* as the proportion of transmission-relevant behaviours that could not be eliminated through changes to policies or mobility. For interpretation of the individual policy weights, we used the quantity 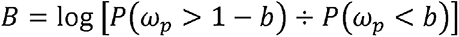 as our primary metric of whether a policy was up- or down-weighted, and set *b* at 0.1. Because each pre-normalisation policy-specific weight *σ_p_* was assigned a uniform prior distribution on [0,1], the prior expectation of *B* is equal to zero, with positive values indicating that the posterior distribution is skewed towards one and negative values indicating skew towards zero.

## Data Availability

The epidemiological, policy and other input data used in generating these analyses are available at https://github.com/monash-emu/renewal/tree/policy_submission/data, with the scripts used to download and process these inputs prior to storage provided at https://github.com/monash-emu/renewal/tree/policy_submission/notebooks/inputs. OxCGRT policy data was downloaded from https://github.com/OxCGRT/covid-policy-dataset/raw/refs/heads/main/data/. Full simulation outputs for all countries have not been deposited due to their size; however, all analyses can be reproduced from the publicly available code, input data and commit identifiers provided in the Supplementary Information. The complete simulation outputs are also available from the authors upon request. The code used in executing the analysis is available at https://github.com/monash-emu/renewal/tree/policy_submission.

https://github.com/monash-emu/renewal/releases/tag/policy_submission

