## Supplemental File 1 for "Quantifying the contribution of policy measures to COVID-19 dynamics across 126 countries"

### Methods

#### Table of contents

**Documentation** 2

**Analysis methods** 2
- Summary equation 2
- Renewal model 4
  - Generation interval 4
  - Renewal process 4
  - Variant seeding 4
  - Immunity 5
  - Outputs 5
- Modelled population size 6
- Analysis period 6
- Transmission scaling for control policies and mobility 7
  - No transmission scaling 7
  - OxCGRT scaling 8
  - Google mobility scaling 8
  - Facebook mobility scaling 9
- Residual transmission process 9
- Calibration targets 10
  - WHO indicators 10
  - Hospitalisations 11
  - Seroprevalence 11
  - Variants 12
- Calibration algorithm 14
  - Initialisation 14
  - Parameter processing 14
  - Algorithm 15

**Selection of countries for inclusion** 15

**Parameter choices and supporting evidence** **24**

**References** **39**

### Documentation

The following document was documented algorithmically from the code used in the analysis. This version was produced from commit da8c990.

The rationale for our methods is provided in the Methods section of the main manuscript.

### Analysis methods

#### Summary equation

The mechanics of the renewal process can be summarised in the following equations, the detail of which is expanded in the following sections.

$$i_{p,v,t} = n_{p,t} x_{p,v} (1 - e^{-\lambda_{v,t}})$$

$$\lambda_{v,t} = \beta \sigma_v e^{W_t} M_t \sum_{\tau=t-50}^{t-1} (s_{v,\tau} + \sum_{p'} i_{p',v,\tau}) g_{t-\tau}$$

$$n_{p,t} = n_{p,t-1} - \sum_{v} i_{p,v,t} + i_{\tilde{p},\tilde{v},t}$$

$$M_t = \begin{cases} 1 & \text{if no scaling} \\ [f + (1-f)\left(1 - \frac{\sum_p \omega_p O_{p,t}}{\sum_p \omega_p}\right)]^m & \text{if OxCGRT floored scaling} \\ \exp\left(-\sum_p \eta_p (O_{p,t} - O_{p,t_0})\right) & \text{if OxCGRT independent scaling} \\ [f + (1-f)\frac{\sum_l \kappa_l G_{l,t}}{\sum_l \kappa_l}]^m & \text{if Google scaling} \\ [f + (1-f) F_t]^m & \text{if Facebook scaling} \end{cases}$$

$$o_t = a \sum_{v} (\prod_{v'=v_1}^{v} y_{v'}) \sum_{p} \sum_{\tau=1}^{50} i_{p,v,t-\tau} \times c_\tau$$

$$h_t = \sum_{\tau=1}^{50} o_{t-\tau} (1 - \sum_{u=0}^{\tau-1} d_u)$$

Where:

- $i_{p,v,t}$ represents the incidence of variant $v$ in immunity status group $p$ at time $t$, where $p$ represents the immunity status prior to infection with variant $v$
- $v$ takes sequential values from $v_1$ to $v_V$, where $V$ is the number of variants included in the analysis
- $p \in \{0,1\}^V$ are the possible combination of past variant exposure statuses with $V$ variants
- $n_{p,t}$ is the size of the population with immunity status $p$ at time $t$

- $x_{p,v}$ is the relative susceptibility of persons with immunity status $p$ to infection with variant $v$
- $x_{\{0\}^V,v} = 1$, full susceptibility for those never previously infected
- $x_{p,v} = 0$ for infection with a variant to which the person has previously been infected with or a preceding variant (e.g. infection with Alpha following infection with Delta)
- $x_{p,v}$ for immunity statuses other than the two preceding cases is the complement of a single calibrated cross-immunity parameter with a beta-distributed prior
- $\beta$ is the infectiousness of the starting variant
- $\sigma_v$ is the infectiousness of variant $v$ relative to the starting variant
- $W_t$ is the non-mechanistic residual transmission process, under which a Wiener sequence of updates from a starting value of zero is linked with a cosine spline at time $t$
- $s_{v,t}$ is the rate of seeding with variant $v$ at time $t$
- $g_t$ is $\int_t^{t+1} g(u)\, du$, where $g$ is the probability density function of the generation interval distribution
- $\tilde{v}$ is the variant for which infection results in transition from immunity status $\tilde{p}$ to immunity status $p$
- $G_{l,t}$ is the Google mobility estimate at location $l$ and time $t$
- $F_t$ is the Facebook mobility estimate (either tiles visited or single tile) at time $t$
- $O_{p,t}$ is the normalised OxCGRT indicator $p$ at time $t$ (zero when unrestricted, one at maximum restriction)
- $\kappa_l$ is a calibrated location-specific weight parameter with support $[0, 1]$ (normalised to sum to one)
- $\omega_p$ is the analogous policy-specific weight in the floored OxCGRT analysis (i.e. the same role as $\kappa_l$)
- $\eta_p$ is a policy-specific effect on $\log M_t$ in the independent OxCGRT analysis, with an independent half-normal prior and no normalisation
- $t_0$ is the first day of the analysis period, to which each policy indicator is referenced in the independent OxCGRT analysis, such that $M_{t_0} = 1$ and $M_t$ may take values either side of one as policies tighten or ease relative to their starting settings
- $f$ is a calibrated floor parameter that applies a lower bound to the scaling signal, thereby limiting the maximum effect of mobility or policy changes
- $o_t$ is any one of the incident indicator quantities calculated from the incidence time series, i.e. either deaths, case notifications, hospital admission or ICU admissions
- $y_v$ is the relative severity of the infecting variant relative to the preceding variant, with $y_1 = 1$ (i.e. the severity of the starting strain is set to one)
- $c_t$ is $\int_t^{t+1} c(u)\, du$, where $c$ represents the probability density function of the distribution of the delay from the onset of the infection episode to the onset of the incident indicator considered
- $a$ is the proportion of infection episodes resulting in the indicator considered
- $h_t$ is either one of the prevalent indicators calculated from the incident indicators, i.e. either hospital occupancy or ICU occupancy
- $d_t$ is $\int_t^{t+1} d(u)\, du$, where $d$ represents the probability density function of the distribution

of the time from the onset of the incident indicator (either hospital or ICU admission) to the end of the prevalent indicator period (either hospital or ICU discharge)

#### Renewal model

We developed a discrete renewal model with daily innovations. The core aspect of our approach was the convolution of the preceding strain-specific incidence rate with the generation interval for subsequent infections, adjusted for past immunity. We then applied convolutions to this model to estimate model outputs that could be compared against our indicator targets described below.

For all analyses, the starting population was assigned to the fully susceptible category.

##### Generation interval

A gamma-distributed generation interval was used for the renewal process. This is consistent with an investigation of household clusters from Germany that showed the profile of serial intervals was well matched by a gamma distribution.[1] The generation interval was truncated at 50 days, because the density reached negligible values beyond this point.

##### Renewal process

The strain-specific incidence array updated for strain seeding was convolved with the generation interval distribution vector to create an array of the effective number of infectious individuals for each strain. These quantities were then multiplied by scalar values representing residual transmission scaling and adjustment for policy or mobility scaling and divided through by the population size (to obtain the scaled per capita infectious population). This was multiplied by the strain-specific vector for the relative infectiousness of each strain to calculate a per capita rate of infection. We then determined the actual force of infection for infection-naive persons for each strain as $1 - e^{-\lambda}$ (where $\lambda$ represents the calculated infection rate) to ensure that the per capita risk of infection could not exceed one in a time step.

##### Variant seeding

Each strain (including the starting strain) was seeded into the model using a triangular pulse of new infections for which the peak per capita seeding rate was specified as a parameter. At each calculation day, the new strain-specific seeding values were added to the most recent value for the strain-specific history of incidence. Infectiousness of each variant was specified with reference to the first modelled variant strain.

#### Immunity

The strain-specific force of infection was then applied to each possible immunological past history of preceding exposure to variant combinations and their associated susceptibility to infection to calculate the number of people infected from each category and transition them to their new immunological states. Persons who had never previously been infected with any strain were considered to have no immunological protection, and the rate of infection was not adjusted further. We considered partial cross immunity was provided by infection with a preceding variant strain to infection with subsequent strains. However, previously infected persons were assumed to derive complete immunity to the infecting strain and to variant strains that had emerged prior to the infecting strain (for example, past infection with Delta conferred complete immunity against future infection with Alpha).

#### Outputs

Once an iteration of the renewal analysis had been run to calculate incidence values over time, the other epidemiological outputs were calculated using a series of convolution operations applied to this series. Cases were calculated by convolving the total incidence summed over strains with a gamma-distributed set of delays from incidence to notification which was truncated after 50 days. This was then multiplied through by the case detection rate (a proportion) and weekly cases were calculated by summing over the preceding 7 days.

By contrast, deaths were calculated from the strain-specific incidence to allow for strain-specific severity for analyses considering the pre-Omicron/pre-vaccination period. The severity of each variant was interpreted with reference to the preceding modelled variant, such that the severity of each variant relative to the starting variant was calculated as the cumulative product of the severity parameter for each modelled emerging variant. Strain-specific variant severity was not included for the Omicron era analyses that considered Omicron subvariants BA.1, BA.2 and BA.5. A separate set of parameters governed the time from incidence to death with the fraction of incident episodes resulting in death given by the infection fatality rate adjusted for strain-specific severity. For Singapore and countries of Oceania, a reduction in the risk of death was applied because this analysis was performed after wide-scale population vaccination had been achieved and Omicron sub-variants were the dominant strains, with severity assumed to be equivalent for all Omicron sub-variants. The relative reduction in the risk of death during the later Omicron/vaccination period was set according to a parameter specifying the protection of vaccination against death given infection and was not varied during calibration, because this would have been collinear with the risk of death parameter.

As for cases and deaths, hospitalisations were estimated through a convolution distribution with independently calibrated parameters, a hospital admission fraction, but the same strain-specific severity parameters as for deaths. As for the approach to deaths, strain-specific severity was included for the pre-Omicron period analyses, and a reduction in the risk of hospitalisation was applied to account for vaccination for the Omicron period analyses. The relative reduction in the risk of hospitalisation was set according to a vaccination protection against hospitalisation given infection parameter. The same vaccination-related reduction was applied to ICU admissions. As for cases, deaths and hospitalisations, the convolution of time to ICU admission was parameterised independently.

Hospital and ICU occupancy were obtained by convolving the time series of hospital and ICU admissions with the complement of the cumulative distribution of the time to hospital or ICU discharge. For comparison to serosurveillance data, seropositivity was calculated as the complement of the proportion of the population remaining in the entirely infection-naïve immunity sub-population. Finally, the proportion of incidence attributable to each variant strain was calculated from the strain-specific incidence.

The parameters to each gamma distribution used in our analysis were parameterised by analytically calculating the "a" (shape) and scale parameters from the mean and standard deviation determined by our literature review.

#### Modelled population size

We used the total population size estimated by the World Bank where a population size was available from this source, and used the estimate provided by the United Nations Department of Economic and Social Affairs otherwise. Population data was downloaded from the World Bank on 1 April 2025. The population size for 2020 was used for all countries except for Singapore and countries of Oceania, for which the population size in 2022 was used (because of the later analysis period for these countries). UN DESA population data was downloaded on 18 March 2025 and where UN DESA population was needed, data for 2020 was used.

#### Analysis period

For all countries, we aimed to place our analysis period during a time period for which the variation in transmission rate may have been substantially attributable to public health and social measures and other changes in population behaviours, although we developed an approach (described below) to address the consideration that the emergence of new SARS-CoV-2 variants likely contributed substantially. Therefore, we aimed to select time periods during which the roll-out of vaccination programs would have contributed less to variations in transmission rate.

For all countries other than Oceania and Singapore, we addressed this by limiting our analysis period to the time prior to vaccination reaching levels that may have had a significant population-level effect.

Given the profile and timing of its epidemic, Singapore was included with the Oceania countries throughout the following analyses.

Mobility analyses were only run for countries other than Singapore and those of Oceania, so that policy and mobility scaling could be compared over a shared pre-Omicron window. For Oceania and Singapore we ran the no-scaling analysis and the OxCGRT analyses only.

For countries other than Singapore and those of Oceania, the start of the calibration period was set to be the time at which the per capita daily rate of deaths passed 2 deaths per million population. However, if this threshold was not reached by 1 June 2020, the simulation commenced at this default time instead. For Singapore and countries of Oceania, the simulation commenced from the time that vaccination coverage reached 90% of its final value.

For all countries other than Singapore and those of Oceania, the end time for the analysis was calculated as the time that population vaccination coverage passed 5%, provided that vaccination coverage did reach this value before the default end time of 31 December 2021. Otherwise, this default end date was used instead. For Oceania and Singapore, the last date in the downloaded OxCGRT series (through 2022) was used, which is the intended end of this later analysis window rather than 31 December 2021. We substituted Germany's data for Switzerland and Ireland because these two countries had almost identical profiles of vaccine doses administered per person in the earliest phases of the roll-out and vaccination data was not available for these two countries through the period when coverage approached the analysis start threshold. Similarly, we substituted Great Britain for Qatar based on the same rationale.

#### Transmission scaling for control policies and mobility

The main purpose of our analysis was to understand the effect of empirically observed changes in policies and mobility on the observed fluctuations of the COVID-19 epidemic.

For each country, we ran one analysis with no scaling to the transmission rate. For countries other than Singapore and those of Oceania, we further ran one analysis in which Google mobility was used to scale the transmission rate, if mobility data was available from Google, and two analyses in which Facebook mobility was used to scale the transmission rate, if mobility data was available from Facebook. We ran two analyses in which OxCGRT policy indicators were used to scale the transmission rate, which differed in how those series were mapped to transmission. For all time series data sources, we smoothed the raw data using a 7-day centred rolling average. For analyses that used a floor and exponent (Google mobility, Facebook mobility, and the floored OxCGRT analysis), we used an exponential scaling parameter (described in more detail below) which was assigned a uniform prior over domain [0, 2].

##### No transmission scaling

For the analysis without scaling for policy or mobility, no empiric data was used to scale the transmission rate over time (which was implemented by setting the scaling value to one throughout the simulation).

##### OxCGRT scaling

Annual OxCGRT files are available for later years, but we only retrieved 2020 to 2022. That extract is the intended analysis window, including for Oceania and Singapore whose run end is the last date in this series. OxCGRT policy indicators were scaled to the unit interval by dividing by each indicator's maximum possible value, so that a value of zero represented no restriction and a value of one represented maximum restriction.

For both OxCGRT analyses, we used eight policy indicators: C1 to C7 (containment and closure policies) and H6 (face coverings). We excluded H1 (public information campaigns) because it was at maximum restriction for almost the entire analysis window in most countries, and where it did vary, the changes did not appear to be credible policy shifts.

##### Floored OxCGRT scaling

This analysis used a weighted combination of OxCGRT policy indicators, with the weights for each indicator normalised to sum to one. Indicators were coded as restriction (zero when unrestricted, one at maximum restriction). The complement of the weighted restriction series was then reduced toward a calibrated floor and exponentiated to the value specified by the scaling exponential parameter, so that greater restriction reduced transmission toward the floor.

##### Independent OxCGRT scaling

This analysis scales transmission by independent multiplicative effects of each OxCGRT policy indicator. Each indicator is coded as zero when unrestricted and one at maximum restriction. Each indicator was then referenced to its value on the first day of the analysis period for the country considered by subtracting this starting value from the series. Transmission is scaled by the exponential of minus the sum over policies of each effect parameter times the referenced indicator. The scaling value is therefore one at the start of the simulation, falling below one as policies tighten relative to the settings in place at that time and rising above one as they ease. This referencing is a reparameterisation that leaves the epidemiological structure of the model unchanged. However, it implies that the transmissibility parameter represents transmission under the policy settings observed at the start of the analysis period, rather than under the counterfactual of a complete absence of policy restrictions, which was not observed for any country during the analysis period and so could only be reached by extrapolation. Each effect parameter was assigned an independent half-normal prior with scale 0.5.

##### Google mobility scaling

We obtained mobility data from the Google Community Mobility Reports on 14 January 2025 and extracted national mobility estimates by Google mobility domain. For this analysis, we used the raw value interpreted as a percentage plus one to scale the transmission rate. This approach to analysis used a set of priors weighting each time series component, with the weights for each time series component normalised to sum to one. Next, a further prior parameter is used to set the floor value for the effect of the time series (i.e. creating a ceiling on the effect of the time series used for scaling). This floored time series is then exponentiated to the value specified by the scaling exponential parameter.

##### Facebook mobility scaling

For each geographic region included in the Facebook data, we estimated a population by intersecting polygons with the centroid of population data grids, and then weighted the resulting series by these calculated populations. For the small proportion of (low-population) time series that had missing data, we imputed population estimates by nearest neighbour interpolation. In general, these series were found to have a negligible contribution to the final outputs.

We used the `all_day_bing_tiles_visited_relative_change` for the first Facebook mobility analysis, and scaled transmission according to one plus this mobility metric. For the second Facebook mobility analysis, we used the `all_day_ratio_single_tile_users` estimate and scaled transmission according to one minus this mobility metric.

This time series approach was used for each of the two analyses that incorporated Facebook data, using both the tiles visited and the within tile estimates.

#### Residual transmission process

We included a non-mechanistic residual process within our model, which was intended to capture any variations in transmission that could not be attributed to the explicitly modelled processes (variant strain emergence, changes in population behaviour and the development of population immunity).

A Wiener variable process was used to capture variation in transmission over time. The starting value for this process was fixed at zero and the subsequent updates to the process were explored in calibration. This exploration was performed in logarithmic space, with the calibrated values for each update exponentiated before being used to scale the transmission rate. Each parameter pertaining to the updates to residual transmission scaling was assigned the same prior centred at zero (i.e. no change), and so can be interpreted as the change in the log-transformed residual transmission scaling relative to the previous value. The cosine function was obtained by translating and scaling a half cosine function (i.e. a cosine function with support $[0, \pi]$), such that it intersected the starting point $(t_1, y_1)$ and finishing point $(t_2, y_2)$ with a gradient of zero at both of these points. This choice of fitting approach ensured that the residual transmission scaling function, its derivative are continuous. Each time point for fitting the residual transmission scaling process was set at intervals through the analysis period spaced by 7 days working backwards from the end of the analysis period. The scaling process was then fit to these points using piecewise cosine functions from a starting value of zero. We compared each scaled analysis to no scaling over the same country-specific window, using the proportion of randomly paired posterior draws in which residual-process dispersion (`dispersion_proc`) was lower under the scaled analysis than under baseline. A value greater than one half indicates that scaling reduced unexplained transmission variation more often than not. Because mobility analyses were not run for Oceania and Singapore, comparison of policy versus mobility scaling was restricted to pre-Omicron countries.

#### Calibration targets

We only included countries for which multiple data streams were available to estimate variations in transmission rates over time. Specifically, we required that a time series for both COVID-19-attributable deaths and case notifications were available. However, we also included one time series for a hospital-related indicator, where this could be sourced. Because we considered that the emergence of new SARS-CoV-2 variants could have substantially affected transmission, we included explicit calibration targets for the strain-specific proportional prevalence of variants where available. Although possibly less epidemiologically relevant to this analysis, we sought to broadly capture the overall size of the epidemic. This was addressed by explicitly incorporating susceptible depletion and including seroprevalence estimates as calibration targets where available. We did not include vaccination explicitly within the model because, for countries other than Oceania and Singapore, our approach to country and time period inclusion was intended to focus on time periods during which vaccination would not have substantially influenced transmission rates. For Oceania and Singapore the analysis window was later, after high vaccination coverage had been reached.

##### WHO indicators

The number of deaths by week reported by WHO was used as a calibration target for all included countries. Any values of zero in this series were replaced with a value of 0.5 to enable comparison of the deaths target to our modelled outputs in logarithmic space. The log of the reported target deaths value was compared against modelled deaths by calculating the density of a normal distribution centred at the log-transformed modelled value. Deaths was one of up to three time-series indicators for which a common dispersion parameter was used for this normal distribution for the target comparison. The value for weighting the deaths indicator time series was set to 20 (a quantity that can be interpreted in relation to the weights of the other indicators, but whose absolute size is arbitrary).

The number of cases by week reported by WHO was included as the second calibration target for all countries. As for deaths, any zero values were replaced with a value of 0.5. We only calibrated to cases from 1 June 2020 onward, because we considered that prior to this time many countries were still rapidly scaling up testing capacity and this indicator may have been less reliable. Cases was the second indicator for which a common dispersion parameter was applied. A total weight was calculated for the overall time-series of cases such that the weight for each individual observation (date) of the cases time-series was equal to that for each death observation.

##### Hospitalisations

A maximum of one hospitalisation-related indicator for calibration was chosen using a hierarchical approach. In selecting the indicator, the number of new admissions was preferred over estimates of total bed occupancy, and total hospital indicators were preferred over ICU indicators. As such, the final hierarchy of indicators was:

1. New hospital admissions
2. Hospital occupancy
3. New ICU admissions
4. ICU occupancy
5. No hospital indicator

That is, the highest ranked indicator was used based on data availability, and no hospital indicator was incorporated if none was available. This indicator was the final calibration target for which a common dispersion parameter was applied. As for the cases target, a weight was applied to the overall hospitalisation time series such that the weight for each observation point was equal to that for each death observation.

##### Seroprevalence

Seroprevalence data was obtained from SeroTracker on 11 December 2024, with the date for each serosurvey estimate calculated as the mid-point between the reported start and end sampling dates for each survey. This date was then lagged earlier by 14 days for comparison to modelled outputs to account for the delay between infection and the subsequent development of detectable antibodies. We filtered the SeroTracker data to include only the estimate reported as primary from Unity-aligned national-level surveys for which the number of participants was at least 600. We also considered only a maximum of one seroprevalence value for any given date (selecting only the largest of three surveys done on the same day for Mexico). For any consecutive estimates for which a lower estimate followed an immediately preceding higher estimate, we pooled these two estimates and placed the pooled estimate at the mid-point of the dates of the two estimates. We recursively applied this process until seroprevalence estimates were monotonically increasing over time.

We compared the modelled proportion ever infected against the seroprevalence reported at least six months (183 days) after the start of the simulation. We chose to exclude seroprevalence estimates from the early months of the pandemic period because the seropositive proportions reported at this time were less likely to be comparable to our model outputs. This is because our modelled seroprevalence would remain close to zero with plausible parameter values for some time after the start of the analysis, whereas seroprevalence estimates could reach higher values due to factors that include low levels of transmission prior to the analysis period and sampling bias. In contrast, we considered that later seroprevalence should provide a broad indication of epidemic size. For countries for which seroprevalence calibration targets were used, we assigned a target weight to this indicator of 5 (which is an arbitrary quantity in absolute terms, but can be interpreted with reference to the deaths indicator weight of 20). The seroprevalence target had a dispersion parameter that was normally distributed and independent of the dispersion parameters for the other targets, including the shared dispersion parameter of the time series indicators.

We discarded seroprevalence estimates that fell less than 0.01 away from a value of zero or one. We also ignored seroprevalence estimates for Oceania and Singapore, for which the analysis was run largely through 2022, during which time seroprevalence values would not reflect the attack rate during the simulation period. We further ignored seroprevalence estimates from low and lower middle income countries of Africa, because we were unable to obtain good fits for several of these countries while also maintaining plausible detection/severity parameters (i.e. case detection rate, hospital admission rate and infection fatality rate). That is, we applied much lower values for these detection-related parameters in these countries, although the modelled attack rate still often remained below seroprevalence estimates for some countries.

Country income classifications were obtained from the World Bank.

#### Variants

##### Data extraction

Reports of the number of isolates of specific variants of SARS-CoV-2 were obtained from the covariants GitHub repository. Each variant-specific file was downloaded and used to create country-specific tables of the variant-specific counts by date. The terms used to identify variants prior to Alpha were: 20A.EU1, 20A.EU2, 20B.S.732A, 21C.Epsilon. The text “Delta” was used to identify Delta variants and the text “21L.Omicron” was used to distinguish the BA.2 variant from later variants (modelled as BA.5). Variant data was considered for dates on which at least 5 sequences were available for the country considered. Further, we required at least 5 such dates be available for that country’s variant data to be used as a calibration target.

To obtain data to use as calibration targets for the strain-specific proportional prevalence of both the Alpha and the Delta variants, we used national data for the country analysed where available. If data was not available for the country considered or if all values for the proportion of the variant were one, we used pooled data from all the other countries from the same continent where available. If insufficient data on a particular variant was available for the country or its continent, prevalence of that variant was not included as a calibration target.

##### Alpha

For countries other than Singapore and countries of Oceania and Africa, a target for the Alpha variant was included in our calibration algorithm. Calibration against data for Alpha was permitted from the beginning of the simulation period (from 1 January 2020) for these countries. The periods for calibration against the Alpha and the Delta variants were set so as to be mutually exclusive in time. Specifically, the date for transitioning from calibrating against available data for the Alpha to calibrating against data for Delta was set as 1 March 2021. Exceptions were made for several Asian countries for which this transition date was set one month earlier and two countries of North America for which it was set six weeks later. (Specifically, the exceptions were Afghanistan: 1 February 2021, India: 1 February 2021, Nepal: 1 December 2020, Indonesia: 1 February 2021, Saudi Arabia: 1 February 2021, Oman: 1 February 2021, Kuwait: 1 February 2021, South Korea: 1 February 2021, Malaysia: 1 February 2021, Honduras: 15 April 2021, Puerto Rico: 15 April 2021.) If this date occurred after the end of the simulation, the Alpha calibration period continued to the end of the simulation. Comparison of the logit-transformed target estimate against the logit-transformed modelled value was undertaken using a normal distribution with a single dispersion parameter that applied to all modelled variants (but was independent of the time series dispersion parameter). Proportions were clipped to the interval from 0.01 to one minus this value before the logit was applied. The target weight for the Alpha target was set to 5.

##### Delta

For countries other than those of Oceania and Singapore, the Delta variant was included if the end date of the calibration fell later than 1 May 2021. The target weight for calibration to Delta was set to 5 for most countries. Exceptions were made if the target time series for Delta began towards the very end of the calibration (last 87 days), in which case a higher weight (of 25) was needed to achieve a plausible fit to the profile of the emergence of this variant with fewer data points.

##### Omicron BA.2

A calibration target for Omicron BA.2 was only included for Oceania and Singapore. As for other variants, the target weight for BA.2 was set to 5. The BA.2 calibration target for these countries used the data available from 1 January 2022 to 15 April 2022.

##### Omicron BA.5

As for BA.2, a calibration target for Omicron BA.5 was only included for Oceania and Singapore. As for other variants, the target weight for BA.5 was set to 5. The BA.5 calibration target for these countries used the data available from 1 April 2022 to 1 September 2022.

#### Calibration algorithm

We used a gradient-based algorithm for model calibration, which efficiently explored our multidimensional parameter space. By exploring all time series in log-space with a common dispersion parameter, we were able to apply an algorithm that provided epidemiologically plausible results for all countries simulated without the need for extensive country-specific adaptations.

##### Initialisation

To initialise the model parameters, we started all the updates to the transmission scaling process from a value of zero (in logarithmic space) to represent no update, such that the initialisation commenced with transmission scaling being constant over time. For the independent OxCGRT policy effects, we started each effect parameter from 0.0001, so that initialisation likewise commenced with no policy-driven change to transmission. For Singapore and countries of Oceania, the starting transmissibility parameter was initialised at 1.0 rather than the centre of its prior, so that initialisation did not commence from a rapidly saturating epidemic. For all other parameters, we used `numpyro`'s `init_to_uniform` method, with a radius of 0.1.

##### Parameter processing

The calibration process estimated parameters for each consecutive update to the transmission scaling process in logarithmic space. The prior for the update for each period of time relative to the previous period was assigned a normal distribution centred at zero, representing no change. The standard deviation of each normal distribution is provided by the (single) dispersion parameter of the transmission scaling process introduced above. The dispersion parameter for transmission scaling was set to a half normal distribution with standard deviation 0.5 under the baseline analyses and each of the analyses that involved scaling transmission according to time series inputs. This approach was adopted in order to favour lesser changes in transmission scaling at each sequential update point.

For all prior distributions representing durations of time (including the generation interval, time from infection to reporting and time from infection to death), we used a truncated normal prior with the lower truncation limit set at 1 day and the upper truncation limit set at 2.5 times the mean value of the prior. For each of the beta-distributed proportions of incidence resulting in the key outputs for calibration, we analytically calculated the parameters to the beta distribution from the desired mean and standard deviation determined from the literature. These parameters comprised the infection fatality rate, the case detection proportion and, where applicable, the hospital or ICU admission proportion. For high-income countries, the mean value used was as presented in the parameter choices section, whereas this was reduced for other countries. The reductions based on income status were: Low income: 0.4, Lower middle income: 0.6, Upper middle income: 0.8, High income: 1.0, with Venezuela (for which a World Bank class was not available) considered a low-income country for this purpose.

The starting transmissibility scaling parameter was assigned a uniform prior distribution on domain 0.5 to 3.5.

The seeding rate for each variant was calibrated from $1 \times 10^{-7}$ to $5 \times 10^{-6}$ cases per capita per day using a uniform distribution in logarithmic space. The seeding offset (i.e. the time from modelled seeding to the first calibration data point) for each variant except for the first one simulated was calibrated from 4.0 to 90.0 days using a uniform distribution in untransformed space.

The relative infectiousness of each variant compared to the preceding variant (other than the first variant included) was calibrated using truncated normal distributions with mean 1.25, standard deviation 0.2, lower truncation limit 1 and upper truncation limit 2.

As for our approach for infectiousness, the relative severity of each variant compared to the preceding variant (other than the first variant included) was calibrated using truncated normal distributions with mean 1.5, standard deviation 0.2, lower truncation limit 1.0 and upper truncation limit 2.0.

The prior for the shared dispersion parameter for all time series data was a half-normal distribution with standard deviation 0.5. The dispersion parameter for all variant incidence proportions (applied on the logit scale) was set at 0.5. The dispersion parameter for all seroprevalence targets was set at 0.2.

##### Algorithm

We ran the calibration algorithm with a warm-up of 10000 iterations for each of 8 chains followed by 10000 evaluated iterations for the main analysis. We used a Hamiltonian Monte Carlo inference approach with the No U-Turn Sampler (NUTS) with adaptive path length and mass matrix adaptation from `numpyro`.

#### Selection of countries for inclusion

Countries were considered for inclusion if they appeared as a national jurisdiction in this OxCGRT extract. Google or Facebook mobility data was not required for inclusion; mobility series were used only for comparative analyses, and only for the pre-Omicron countries, as described above.

We considered the quality of the data for our two main WHO indicators which we required for inclusion in the analysis: `New_cases` and `New_deaths` from the start of data availability through to the end time of the analysis for each country. We considered data quality from 1 November 2021 for countries of Oceania and from 1 April 2020 for the countries of all other continents over the period for which an OxCGRT simulation would apply.

Using this data, we excluded any countries for which no deaths or cases were reported throughout this data availability period. We also excluded any countries for which any negative values were present within the available data. We further excluded any countries for which single marked outliers were present, which we defined as a single value that was more than 2 times greater than the next highest estimate present during the analysis period. Last we excluded any countries for which multiple consecutive missing values were present in the surveillance data because it was unclear whether these should be interpreted as unavailable or as zeroes. Similarly, we excluded any countries (which affected only one country) for which repeated identical values were present (defined as changes within $1 \times 10^{-7}$). For both consecutive missing values and consecutive repeated values, we set the threshold for the number of observations for exclusion to be 15. For Singapore and countries of Oceania, vaccination coverage is required to determine the start of the analysis period. We therefore excluded any such country for which this series was unavailable from Our World in Data.

The final results of the selection are presented in Table 1. The final included countries are illustrated in Figure 1 below.

Table 1: Reasons for country inclusion and exclusion.

| | No death data | No case data | Negative values present | Outlier values present | Absent or repeat values | No Oceania vaccination data | Included |
|---|---|---|---|---|---|---|---|
| Afghanistan | No | No | No | No | No | No | Yes |
| Albania | No | No | No | No | No | No | Yes |
| Algeria | No | No | No | No | No | No | Yes |
| Andorra | No | No | No | No | No | No | Yes |
| Angola | No | No | No | No | No | No | Yes |
| Argentina | No | No | No | No | No | No | Yes |
| Aruba | No | No | No | No | No | No | Yes |
| Australia | No | No | No | No | No | No | Yes |
| Austria | No | No | No | No | No | No | Yes |
| Azerbaijan | No | No | No | No | No | No | Yes |
| Bahamas | No | No | No | No | No | No | Yes |
| Bahrain | No | No | No | No | No | No | Yes |
| Bangladesh | No | No | No | No | No | No | Yes |

Table 1: Reasons for country inclusion and exclusion.

| | No death data | No case data | Negative values present | Outlier values present | Absent or repeat values | No Oceania vaccination data | Incl-uded |
|---|---|---|---|---|---|---|---|
| Barbados | No | No | No | No | Yes | No | No |
| Belarus | No | No | No | No | No | No | Yes |
| Belgium | No | No | No | No | No | No | Yes |
| Belize | No | No | No | No | Yes | No | No |
| Benin | No | No | No | No | No | No | Yes |
| Bermuda | No | No | No | No | Yes | No | No |
| Bhutan | No | No | No | No | Yes | No | No |
| Bolivia, Plurinational State of | No | No | No | Yes | No | No | No |
| Bosnia and Herzegovina | No | No | No | No | No | No | Yes |
| Botswana | No | No | No | No | Yes | No | No |
| Brazil | No | No | No | No | No | No | Yes |
| Brunei Darussalam | No | No | No | No | Yes | No | No |
| Bulgaria | No | No | No | No | No | No | Yes |
| Burkina Faso | No | No | No | No | No | No | Yes |
| Burundi | No | No | No | Yes | Yes | No | No |
| Cabo Verde | No | No | No | No | No | No | Yes |
| Cambodia | No | No | No | Yes | Yes | No | No |
| Cameroon | No | No | No | No | No | No | Yes |
| Canada | No | No | No | No | No | No | Yes |
| Central African Republic | No | No | No | No | Yes | No | No |
| Chad | No | No | No | No | Yes | No | No |
| Chile | No | No | No | No | No | No | Yes |
| China | No | No | No | Yes | No | No | No |
| Colombia | No | No | No | No | No | No | Yes |
| Congo | No | No | No | No | No | No | Yes |
| Congo, The Democratic Republic of the | No | No | No | Yes | No | No | No |
| Costa Rica | No | No | No | No | No | No | Yes |
| Croatia | No | No | No | No | No | No | Yes |

Table 1: Reasons for country inclusion and exclusion.

| | No death data | No case data | Negative values present | Outlier values present | Absent or repeat values | No Oceania vaccination data | Incl-uded |
|---|---|---|---|---|---|---|---|
| Cuba | No | No | No | No | No | No | Yes |
| Cyprus | No | No | No | No | No | No | Yes |
| Czechia | No | No | No | No | No | No | Yes |
| Côte d'Ivoire | No | No | No | No | No | No | Yes |
| Denmark | No | No | No | No | No | No | Yes |
| Djibouti | No | No | No | No | Yes | No | No |
| Dominica | Yes | No | No | No | Yes | No | No |
| Dominican Republic | No | No | No | No | No | No | Yes |
| Ecuador | No | No | Yes | Yes | No | No | No |
| Egypt | No | No | No | No | No | No | Yes |
| El Salvador | No | No | No | No | No | No | Yes |
| Eritrea | No | No | No | No | Yes | No | No |
| Estonia | No | No | No | No | No | No | Yes |
| Eswatini | No | No | No | No | No | No | Yes |
| Ethiopia | No | No | No | No | No | No | Yes |
| Faroe Islands | No | No | No | No | Yes | No | No |
| Fiji | No | No | No | No | Yes | No | No |
| Finland | No | No | No | No | No | No | Yes |
| France | No | No | No | No | No | No | Yes |
| Gabon | No | No | No | No | No | No | Yes |
| Gambia | No | No | No | Yes | Yes | No | No |
| Georgia | No | No | No | No | No | No | Yes |
| Germany | No | No | No | No | No | No | Yes |
| Ghana | No | No | No | Yes | Yes | No | No |
| Greece | No | No | No | No | No | No | Yes |
| Greenland | Yes | No | No | Yes | Yes | No | No |
| Guam | No | No | No | No | No | Yes | No |
| Guatemala | No | No | Yes | No | No | No | No |
| Guinea | No | No | No | No | No | No | Yes |
| Guyana | No | No | No | No | No | No | Yes |
| Haiti | No | No | No | No | No | No | Yes |
| Honduras | No | No | No | No | No | No | Yes |
| Hong Kong | Yes | Yes | No | No | No | No | No |
| Hungary | No | No | No | No | No | No | Yes |
| Iceland | No | No | No | No | Yes | No | No |
| India | No | No | No | No | No | No | Yes |

Table 1: Reasons for country inclusion and exclusion.

| | No death data | No case data | Negative values present | Outlier values present | Absent or repeat values | No Oceania vaccination data | Incl-uded |
|---|---|---|---|---|---|---|---|
| Indonesia | No | No | No | No | No | No | Yes |
| Iran, Islamic Republic of | No | No | No | No | No | No | Yes |
| Iraq | No | No | No | No | No | No | Yes |
| Ireland | No | No | No | No | No | No | Yes |
| Israel | No | No | No | No | No | No | Yes |
| Italy | No | No | No | No | No | No | Yes |
| Jamaica | No | No | No | No | No | No | Yes |
| Japan | No | No | No | No | No | No | Yes |
| Jordan | No | No | No | No | No | No | Yes |
| Kazakhstan | No | No | No | Yes | No | No | No |
| Kenya | No | No | No | No | No | No | Yes |
| Kiribati | No | No | No | Yes | Yes | No | No |
| Korea, Republic of | No | No | No | No | No | No | Yes |
| Kosovo | No | No | No | No | No | No | Yes |
| Kuwait | No | No | No | No | No | No | Yes |
| Kyrgyzstan | No | No | No | No | Yes | No | No |
| Lao People's Democratic Republic | No | No | No | No | Yes | No | No |
| Latvia | No | No | No | No | No | No | Yes |
| Lebanon | No | No | No | No | No | No | Yes |
| Lesotho | No | No | No | No | No | No | Yes |
| Liberia | No | No | No | No | No | No | Yes |
| Libya | No | No | No | No | No | No | Yes |
| Liechtenstein | No | No | No | No | Yes | No | No |
| Lithuania | No | No | No | No | No | No | Yes |
| Luxembourg | No | No | No | No | No | No | Yes |
| Macao | Yes | Yes | No | No | No | No | No |
| Madagascar | No | No | No | No | No | No | Yes |
| Malawi | No | No | No | No | No | No | Yes |
| Malaysia | No | No | No | No | No | No | Yes |
| Mali | No | No | No | No | No | No | Yes |
| Malta | No | No | No | No | No | No | Yes |
| Mauritania | No | No | No | Yes | No | No | No |
| Mauritius | No | No | No | No | Yes | No | No |

Table 1: Reasons for country inclusion and exclusion.

| | No death data | No case data | Negative values present | Outlier values present | Absent or repeat values | No Oceania vaccination data | Incl-uded |
|---|---|---|---|---|---|---|---|
| Mexico | No | No | No | No | No | No | Yes |
| Moldova, Republic of | No | No | No | No | No | No | Yes |
| Monaco | No | No | No | Yes | Yes | No | No |
| Mongolia | No | No | No | No | Yes | No | No |
| Morocco | No | No | No | No | No | No | Yes |
| Mozambique | No | No | No | No | No | No | Yes |
| Myanmar | No | No | No | No | Yes | No | No |
| Namibia | Yes | Yes | No | No | No | No | No |
| Nepal | No | No | No | No | No | No | Yes |
| Netherlands | No | No | No | No | No | No | Yes |
| New Zealand | No | No | No | No | No | No | Yes |
| Nicaragua | No | No | No | No | Yes | No | No |
| Niger | No | No | No | Yes | Yes | No | No |
| Nigeria | No | No | No | No | No | No | Yes |
| Norway | No | No | No | No | No | No | Yes |
| Oman | No | No | No | No | No | No | Yes |
| Pakistan | No | No | No | No | No | No | Yes |
| Palestine, State of | No | No | No | No | No | No | Yes |
| Panama | No | No | No | No | No | No | Yes |
| Papua New Guinea | No | No | No | Yes | No | No | No |
| Paraguay | No | No | No | No | No | No | Yes |
| Peru | No | No | No | No | No | No | Yes |
| Philippines | No | No | No | No | No | No | Yes |
| Poland | No | No | No | No | No | No | Yes |
| Portugal | No | No | No | No | No | No | Yes |
| Puerto Rico | No | No | No | Yes | No | No | No |
| Qatar | No | No | No | No | No | No | Yes |
| Romania | No | No | No | No | No | No | Yes |
| Russian Federation | No | No | No | No | No | No | Yes |
| Rwanda | No | No | No | No | No | No | Yes |
| San Marino | No | No | No | No | Yes | No | No |
| Saudi Arabia | No | No | No | No | No | No | Yes |
| Senegal | No | No | No | No | No | No | Yes |

Table 1: Reasons for country inclusion and exclusion.

| | No death data | No case data | Negative values present | Outlier values present | Absent or repeat values | No Oceania vaccination data | Incl-uded |
|---|---|---|---|---|---|---|---|
| Serbia | No | No | No | No | No | No | Yes |
| Seychelles | No | No | No | No | Yes | No | No |
| Sierra Leone | No | No | No | No | Yes | No | No |
| Singapore | No | No | No | No | No | No | Yes |
| Slovakia | No | No | No | No | No | No | Yes |
| Slovenia | No | No | No | No | No | No | Yes |
| Solomon Islands | No | No | No | No | Yes | No | No |
| Somalia | No | No | No | No | No | No | Yes |
| South Africa | No | No | No | No | No | No | Yes |
| South Sudan | No | No | No | No | No | No | Yes |
| Spain | No | No | No | No | No | No | Yes |
| Sri Lanka | No | No | No | No | No | No | Yes |
| Sudan | No | No | No | No | No | No | Yes |
| Suriname | No | No | No | No | No | No | Yes |
| Sweden | No | No | No | No | No | No | Yes |
| Switzerland | No | No | No | No | No | No | Yes |
| Syrian Arab Republic | No | No | No | No | No | No | Yes |
| Taiwan, Province of China | Yes | Yes | No | No | No | No | No |
| Tajikistan | No | No | No | No | Yes | No | No |
| Tanzania, United Republic of | No | No | No | Yes | Yes | No | No |
| Thailand | No | No | Yes | No | No | No | No |
| Timor-Leste | No | No | No | No | Yes | No | No |
| Togo | No | No | No | No | No | No | Yes |
| Tonga | No | No | No | Yes | Yes | No | No |
| Trinidad and Tobago | No | No | No | No | Yes | No | No |
| Tunisia | No | No | No | No | No | No | Yes |
| Turkmenistan | Yes | Yes | No | No | Yes | No | No |
| Türkiye | No | No | No | No | No | No | Yes |
| Uganda | No | No | No | Yes | Yes | No | No |
| Ukraine | No | No | No | No | No | No | Yes |

Table 1: Reasons for country inclusion and exclusion.

| | No death data | No case data | Negative values present | Outlier values present | Absent or repeat values | No Oceania vaccination data | Incl-uded |
|---|---|---|---|---|---|---|---|
| United Arab Emirates | No | No | No | No | No | No | Yes |
| United Kingdom | No | No | No | No | No | No | Yes |
| United States | No | No | No | No | No | No | Yes |
| Uruguay | No | No | No | No | No | No | Yes |
| Uzbekistan | No | No | No | No | Yes | No | No |
| Vanuatu | No | No | No | Yes | Yes | No | No |
| Venezuela, Bolivarian Republic of | No | No | No | No | No | No | Yes |
| Viet Nam | No | No | No | No | Yes | No | No |
| Virgin Islands, U.S. | No | No | Yes | No | No | No | No |
| Yemen | No | No | No | No | No | No | Yes |
| Zambia | No | No | No | No | No | No | Yes |
| Zimbabwe | No | No | No | No | No | No | Yes |

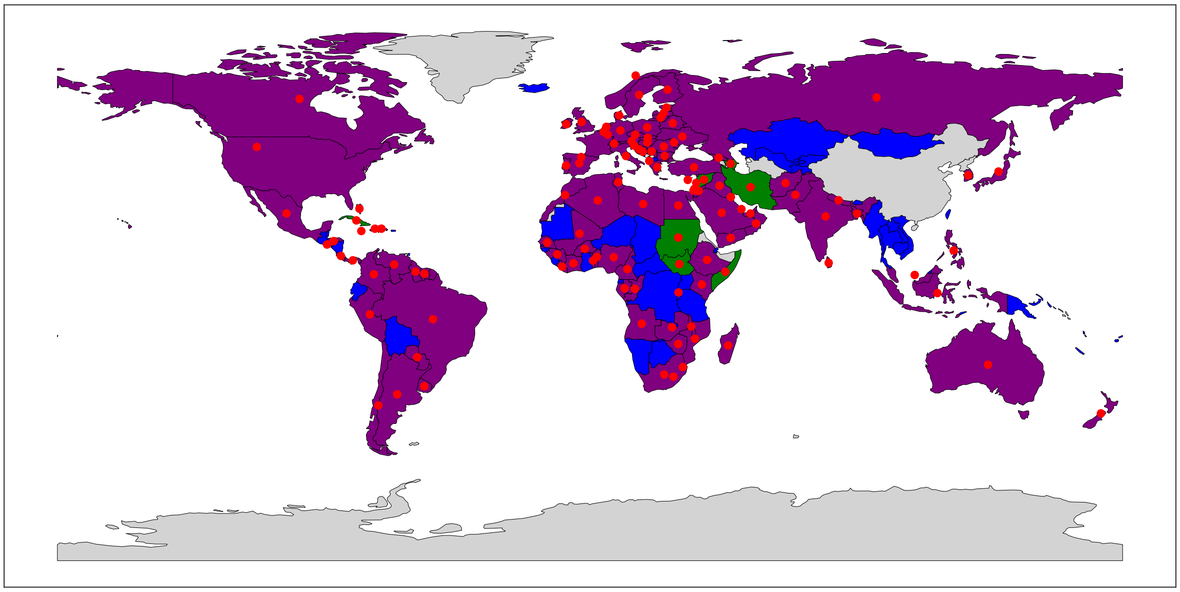

Figure 1: Included countries and mobility availability. Grey, neither included nor mobility data; green, included without mobility; blue, mobility data but not included; purple, included with mobility. Red markers indicate countries included after OxCGRT and surveillance-quality filters. Mobility availability is shown for context; it was not a criterion for inclusion.

#### Parameter choices and supporting evidence

##### Time period parameters

Table 2: Parameters and supporting evidence to time period priors.

| | Mean | SD | Evidence |
|---|---|---|---|
| Generation time mean (days) | 4.25 | 0.75 | We used analyses of case-contact transmission pairs from multiple contexts for SARS-CoV-2 as the main evidence to quantify the generation time distribution.<br><br>Case-contact data from the origin of the pandemic in Wuhan estimated a mean generation time of around seven days prior to the scaling up of population-wide public health and social measures, such as expanded contact tracing and movement restrictions. This led to a notable contraction of the mean generation time to around four to five days after these interventions had been implemented.[2] This study also found marked differences between the generation time and serial interval, due to changes in the epidemic that were sufficiently rapid to result in differences in the incubation period between infectors and infectees. Values of around four to five days are also consistent with an analysis from early 2020 from Tianjin, China and Singapore, in which estimates of the serial interval estimates were close to that of the generation time[3].<br><br>Estimates of the serial interval from household clusters in Germany[1] also found that values of four to five days were appropriate for wild-type, Alpha and Delta SARS-CoV-2 variants, although these appeared less applicable to Omicron. Estimates from close contacts in Spain[4] similarly placed the serial interval of the Delta variant at around five days, while an early study from China similarly placed the mean serial interval for wild-type SARS-CoV-2 at around four to five days.[5]<br><br>A systematic review and meta-analysis of this topic found more studies estimating the serial interval than the generation time, with mean estimates for the serial interval of 4.82 days, 3.59 days and 3.47 days reported for wild-type, Alpha and Delta respectively.[6] Estimates for the generation time from this review were similar, but with wider confidence limits. A rapid review of serial intervals from a different research group reported similar findings.[7] There was notable variation in these findings, with results of individual studies from these systematic reviews estimating mean values for the serial interval of less than three days to more than six days. |

| | Mean | SD | Evidence |
|---|---|---|---|
| Generation time mean (days), Oceania and Singapore | 3 | 0.75 | An analysis from China estimated that the generation time fell from 5.2 days to 2.2 days over the course of an Omicron outbreak in Hangzhou in 2022.[8] The systematic review of generation times by variant cited above estimated a mean generation time of 2.96 days (95% confidence interval 2.54 to 3.38 days).[6] A rapid review and meta-analysis of generation times by variant found a pooled estimate of 3.3 days for Omicron.[7]<br><br>As such, we chose a generation interval mean for the Oceania and Singapore analyses of three days. |
| Generation time standard deviation (days) | 2 | 0.4 | For the standard deviation of the generation interval, we generally used the same sources of evidence as for the mean. However, the systematic reviews and many of the individual studies presented their results as point estimates with confidence intervals, rather than reporting the standard deviations of the data of the individual studies, making these results less applicable to this parameter.<br><br>In studies that reported standard deviations, estimates included two to four days for early Wuhan[2] and 1.5 to 2.5 days from Tianjin and Singapore.[3] |

| | Mean | SD | Evidence |
|---|---|---|---|
| Infection to reporting mean (days) | 8 | 0.5 | This quantity is a composite of the incubation period and the delay from symptom onset to diagnosis, although the latter of these quantities is not widely reported. The review cited above in estimating the generation time found estimates of the incubation period of 6.5, 4.92 and 4.63 days for the wild-type, Alpha and Delta variants respectively.[6] A meta-analysis of early cases reported a mean incubation period of 5.68 days[9], while an analysis of early cases in China found a mean incubation period of 5.2 days.[10] As such, although the incubation period was not an explicit model parameter, we considered five days to be a plausible central estimate when considering parameters required for the convolutions applied in our analysis, including this one.<br><br>Regarding the delay from symptom onset to case notification, an analysis from Australia found that this quantity was often in the range of one to seven days after the first wave of cases in early 2020 had declined.[11] This is consistent with a survey from the USA that reported a median of three days unwell before testing.[12] A study of case notifications in New York City found a mean delay from testing to case notification of 3.3 days.[13] WHO reported that the median time from symptom onset to laboratory confirmation had decreased to three days by early February 2020.[14] Our previous analysis of the Australian epidemic found that our prior estimate of three to five days was not markedly influenced through calibration.[15]<br><br>Studies of the time from exposure to diagnosis include a study from South Korea which found an average of 3.7 days,[16] although this very short interval likely reflects proactive testing for these quarantined close contacts. An analysis from the USA estimated that infections could peak anywhere from six to 41 days before peaks in case notification.[17]<br><br>Overall, we considered that a mean time from infection to case notification of eight days would be consistent with a five-day incubation period followed by a three-day average delay from symptom onset to case notification. |

| | Mean | SD | Evidence |
|---|---|---|---|
| Infection to reporting standard deviation (days) | 3 | 0.5 | Although person-to-person variation in the incubation period was not typically quantified in systematic reviews,[6] individual studies reported standard deviations of around three days[10].<br><br>The survey from the USA reported an interquartile range for the time to testing from feeling unwell that spanned five days.[12] Most delays in the study from New York fell in the range from one to five days,[13] while the WHO report cited above described an interquartile range that spanned six days.[14]<br><br>Given this background, we considered that a standard deviation for the time from infection to case notification of three days was plausible, and set the standard deviation for the prior of this parameter distribution to 0.5 days. |
| Infection to death mean (days) | 22 | 5 | The meta-analysis of early studies from China cited above[9] estimated the mean duration from symptom onset to death to be 15.93 days. A study leveraging individual-level data from Hubei, China in combination with early cases outside of China (not included in the meta-analysis) found a mean time from symptom onset to death of 17.8 days.[18] This was similar to a report from a single centre study from Wuhan (Hubei, China) during a time of major health system strain, which found a median time to death of 15 days,[19] and another single centre study of early cases from Wuhan which reported a median time to death of 16 days.[20] A cohort study from a north-eastern state of Brazil found a median 19 days from symptom onset to death.[21] An analysis of deaths from Victoria, Australia over the course of 2020 found a mean time to death of 18.1 days.[22]<br><br>We therefore considered 17 days to be a reasonable estimate for the time from symptom onset to death, which we increased by five days to account for the time from infection to symptom onset. |

| | Mean | SD | Evidence |
|---|---|---|---|
| Infection to death standard deviation (days) | 3 | 0.5 | The first single centre study from Wuhan cited above reported an interquartile range which spanned nine days,[19] while the second reported a span of eight days.[20] The interquartile range from the study in north-eastern Brazil spanned 11 days.[21]<br><br>We considered that the standard deviation for the infection to death prior would be broadly of the order of one quarter the ranges reported by these individual studies, although inflated slightly to account for the uncertainty in the incubation period. We therefore set this parameter to three days, with a standard deviation to the distribution of 0.5 days. |
| Infection to hospitalisation mean (days) | 10 | 3 | One of the earliest studies of cases from China found a median time from onset of symptoms to hospital admission of seven days,[23] while a single centre study from Wuhan reported a median time from symptom onset to hospital admission of ten days.[20] A cohort of adult patients admitted to an Italian hospital found a mean time from onset of symptoms to hospital admission of 3.6 days,[24] while a study from Belgium reported the mean duration from symptom onset to hospitalisation to be 5.6 days.[25] The final report of the ISARIC collaboration reported a mean of 5.4 days from symptom onset to admission.[26]<br><br>Given this evidence, we chose a mean value for the time from symptom onset to hospitalisation prior of five days. As for the time from infection to death parameter, we inflated this by five days to allow for the incubation period. |
| Infection to hospitalisation standard deviation (days) | 2 | 0.5 | The early study of cases from China reported an interquartile range that spanned four days,[23] while the single centre study from Wuhan reported an interquartile range spanning six days.[20] The Italian cohort of hospital admissions reported the standard deviation of the time from symptom onset to hospitalisation to be 3.2 days.[24] The study from Belgium cited above found considerable variation in the time to admission, with the interquartile range spanning six days.[25]<br><br>As with previous parameters, we selected a prior standard deviation of slightly more than one quarter of the spread in the data in the constituent studies. We therefore chose a central estimate for this standard deviation parameter of two days, with a standard deviation to this distribution of 0.5 days. |

| | Mean | SD | Evidence |
|---|---|---|---|
| Hospital stay duration mean (days) | 10 | 3 | An analysis of surveillance data from Belgium reported a mean length of hospital stay of 11.0 days,[25] while a multi-centre study from the USA reported a mean length of hospital stay of 14.7 days.[27] The single centre study from Wuhan cited above reported a median time from hospital admission to discharge of 16 days, with a median time from admission to death of five days.[20]<br><br>A systematic review of hospital admission durations found a median length of stay of 14 days in China, but only five days outside of China.[28], while a scoping review of infectious durations reported that the mean time from symptom onset to hospital discharge or death was 18.1 days.[29] Another systematic review undertaken later in the pandemic period found similar durations, with a mean length of stay of 15.35 days.[30]<br><br>The final report of the ISARIC collaboration reported a mean 9.7 days (median seven days) length of hospital stay.[26] |
| Hospital stay duration standard deviation (days) | 2.5 | 0.5 | The analysis from Belgium cited above reported an interquartile range that spanned ten days,[25] while the multi-centre study from the USA reported a standard deviation of 13.7 days.[27]<br><br>The interquartile range reported in the first systematic review above[28] was ten to 19 days, while the individual-level standard deviation identified in the later systematic review above[30] was 7.92 days.<br><br>As such, we implemented a standard deviation for hospital admission duration of 2.5 days, with a standard deviation to this parameter prior of 0.5 days. |

| | Mean | SD | Evidence |
|---|---|---|---|
| Infection to ICU admission mean (days) | 12 | 3 | A single centre study from Wuhan found a median duration from symptom onset to ICU admission of 9.5 days,[31] while an early study of cases from China found a median time from onset of symptoms to ICU admission of 10.5 days.[23] A study from two hospitals in China reported a median time from illness onset to ICU admission of 12.0 days.[32] The study from north-eastern Brazil reported a median six days from symptom onset to ICU admission.[21]<br><br>The final report from the ISARIC collaboration estimated a mean time from symptom onset to hospital admission of 5.4 days and a mean time from hospital admission to ICU admission of 1.8 days.[26]<br><br>Given this evidence, we implemented a mean for our prior distribution of time from infection to ICU admission of twelve days to allow for a five day incubation period. |
| Infection to ICU admission standard deviation (days) | 2.5 | 0.5 | The single centre study from Wuhan cited above reported an interquartile range around the time from admission to discharge that spanned five days, while the time to death spanned six days.[20] The study of early cases from China reported an interquartile range that spanned nine days,[23] while the study from two hospitals in China reported an interquartile range that spanned 7.0 days.[32] The second single centre study from Wuhan reported an interquartile range from the onset of symptoms to ICU admission that spanned 5.5 days.[31] The analysis from Belgium cited above reported an interquartile range for ICU admission that spanned 13 days.[25] The multi-centre study from the USA reported a standard deviation for the ICU admission duration of 8.3 days.[27] The study in north-eastern Brazil reported an interquartile range around the time to ICU admission that spanned 6.5 days.[21]<br><br>We therefore implemented a standard deviation for our time from infection to ICU admission parameter of 2.5 days, with a standard deviation to this distribution of 0.5 days. |

| | Mean | SD | Evidence |
|---|---|---|---|
| ICU stay duration mean (days) | 10 | 3 | The analysis from Belgium cited above reported a mean length of ICU stay of 12.2 days,[25] while the multi-centre study from the USA cited above reported a mean ICU admission duration of 8.3 days.[27] The study from multiple regions of Brazil indicated the median length of ICU stay was five days.[33]<br><br>The systematic review introduced above[28] reported a median ICU length of stay outside of China of seven days. The final report from ISARIC gave a mean duration of stay in ICU of 10.3 days.[26]<br><br>We therefore chose a mean for the prior distribution of the duration of ICU stay of ten days. |
| ICU stay duration standard deviation (days) | 2.5 | 0.5 | The study from multiple regions of Brazil reported an interquartile range that spanned eight days,[33] while the final ISARIC report provided a standard deviation for ICU duration of 10.4 days.[26]<br><br>We therefore chose a standard deviation of 2.5 days for this prior distribution and assigned a standard deviation for the spread of this parameter of 0.5 days. |

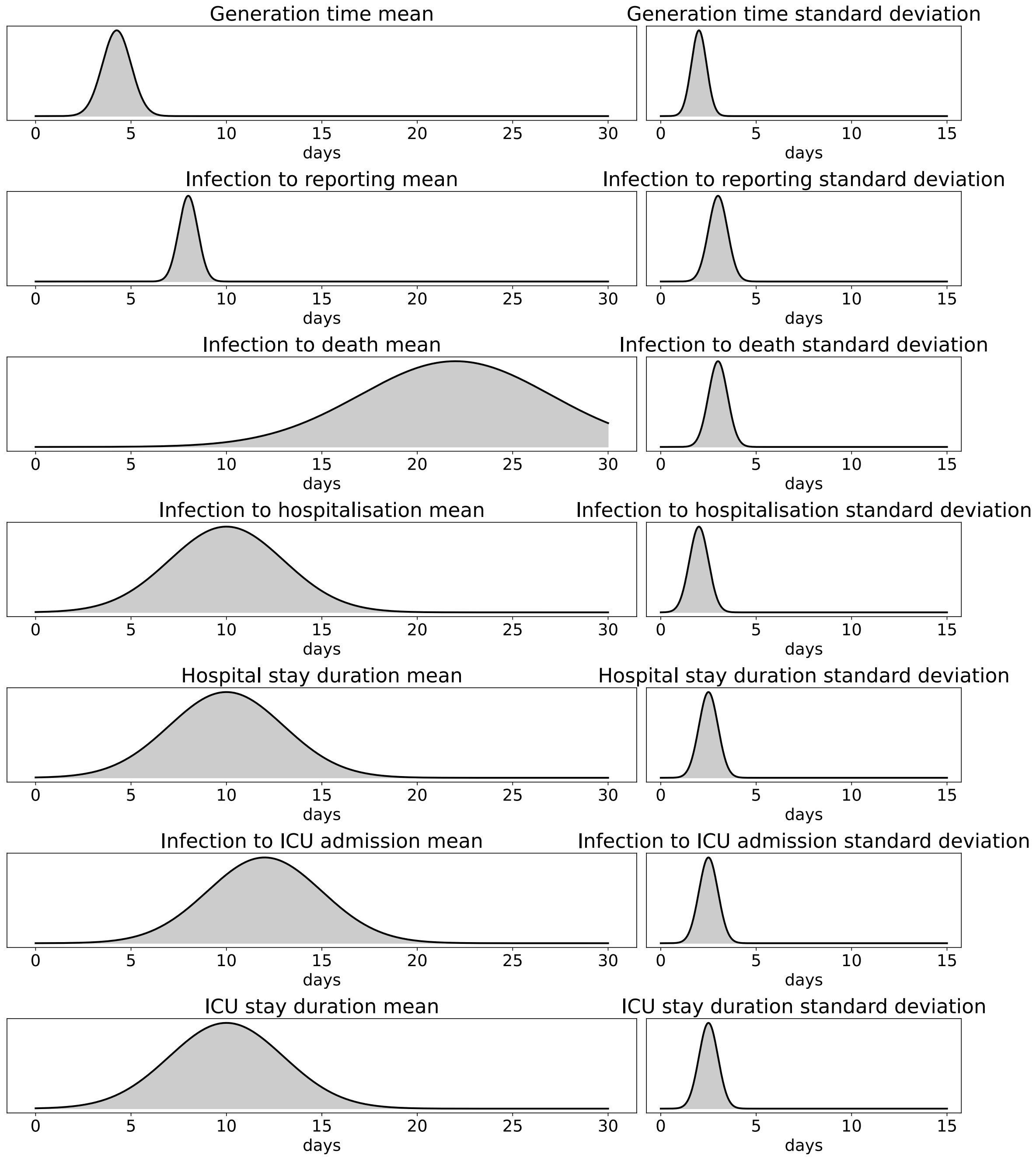

Figure 2: Duration-related parameter prior distributions.

#### Proportion parameters

Table 3: Parameters and supporting evidence to beta-distributed priors.

| | Mean | SD | Evidence |
|---|---|---|---|
| Case detection proportion | 0.4 | 0.1 | An analysis from France in early to mid-2020 of the proportion of cases detected estimated that case ascertainment increased from 7% to 38% (95% confidence interval 35 to 44%) over time.[34] A study of multiple countries found that case detection was highly dependent on the country considered, but reached 20 to 60% in several European countries by the middle of 2020.[35] Estimates from the UK Health Security Agency placed the proportion of cases detected over time at around 30 to 60% from the latter half of 2020.[36] These estimates are consistent with our previous estimate that one in two to one in six cases were detected from the early stages of Australia's Omicron outbreak.[15] An analysis from Africa estimated generally lower case ascertainment proportions, which also varied significantly by country.[37]<br><br>Given the difficulty in determining an appropriate prior for this parameter and the likelihood that the quantity is highly setting-dependent, we selected a broad prior distribution for this parameter. |
| Cross immunity | 0.5 | 0.1 | A pre-print describing a large dataset from the USA in 2020 found an 87% lower rate of reinfection compared to primary infection over a median five months of follow-up.[38]<br><br>A systematic review and meta-analysis found that protection against re-infection declined with time and varied with the variant considered, but was estimated at 78.6% after 40 weeks from infection (95% uncertainty interval 49.8 to 93.6%).[39] Another systematic review and meta-analysis of the same topic found 65% protection (credible interval 53 to 74%) overall, with the past infecting variant and time from infection appearing to modify the extent of protection.[40] A systematic review and meta-regression of reinfections with Omicron variants found that protection from previous infections waned to lower levels (25%) by 12 months from infection.[41] |

| | Mean | SD | Evidence |
|---|---|---|---|
| Infection hospitalisation proportion | 0.03 | 0.006 | This quantity was estimated to be 3.39% (credible interval 2.79 to 3.97%) from officially reported data in England prior to the marked declines that occurred as population-wide vaccination was rolled out.[42] An analysis from Indiana, USA estimated a value of 2.1% overall (credible interval 1.6 to 3.0%), with substantial variation by demographic factors, particularly age.[43] A study from the USA estimated that one in three adults aged 70 years and above who were infected were hospitalised, while one in 21.6 infections in those aged 18 to 49 years resulted in hospitalisation.[44] |
| Infection fatality proportion | 0.008 | 0.001 | An analysis that integrated serosurveillance with age-specific death rates from 45 countries estimated an infection fatality rate of 0.79% (credible interval 0.68 to 0.92%) for the reference population distribution of France.[45] Other groups have found similar estimates using a combination of modelling and serosurvey results, including an overall IFR estimated at 0.49 to 2.53%.[46] The analysis from England of the hospitalisation risk estimated a peak infection fatality risk of 0.97% in January 2021, which fell to 0.06% by April 2022.[42]<br><br>We selected a prior distribution with a central estimate of 0.8%. Our prior estimate is relatively broadly distributed and so should be applicable to a range of higher income settings. |
| ICU admission proportion | 0.01 | 0.001 | The analysis from Indiana cited above found an infection ICU admission risk of 0.51% (credible interval 0.38% to 0.70%).[43] An analysis from Chile found a risk of ICU admission given infection of 0.33, 1.18, 3.59 and 7.18% for the age groups under 40, 40 to 49, 50 to 59 and 60 to 69 respectively.[47] A systematic review and meta-analysis from early in the pandemic period found that 32% of hospital admissions resulted in ICU admission.[48]<br><br>We therefore chose a central estimate for this parameter of 1%. |

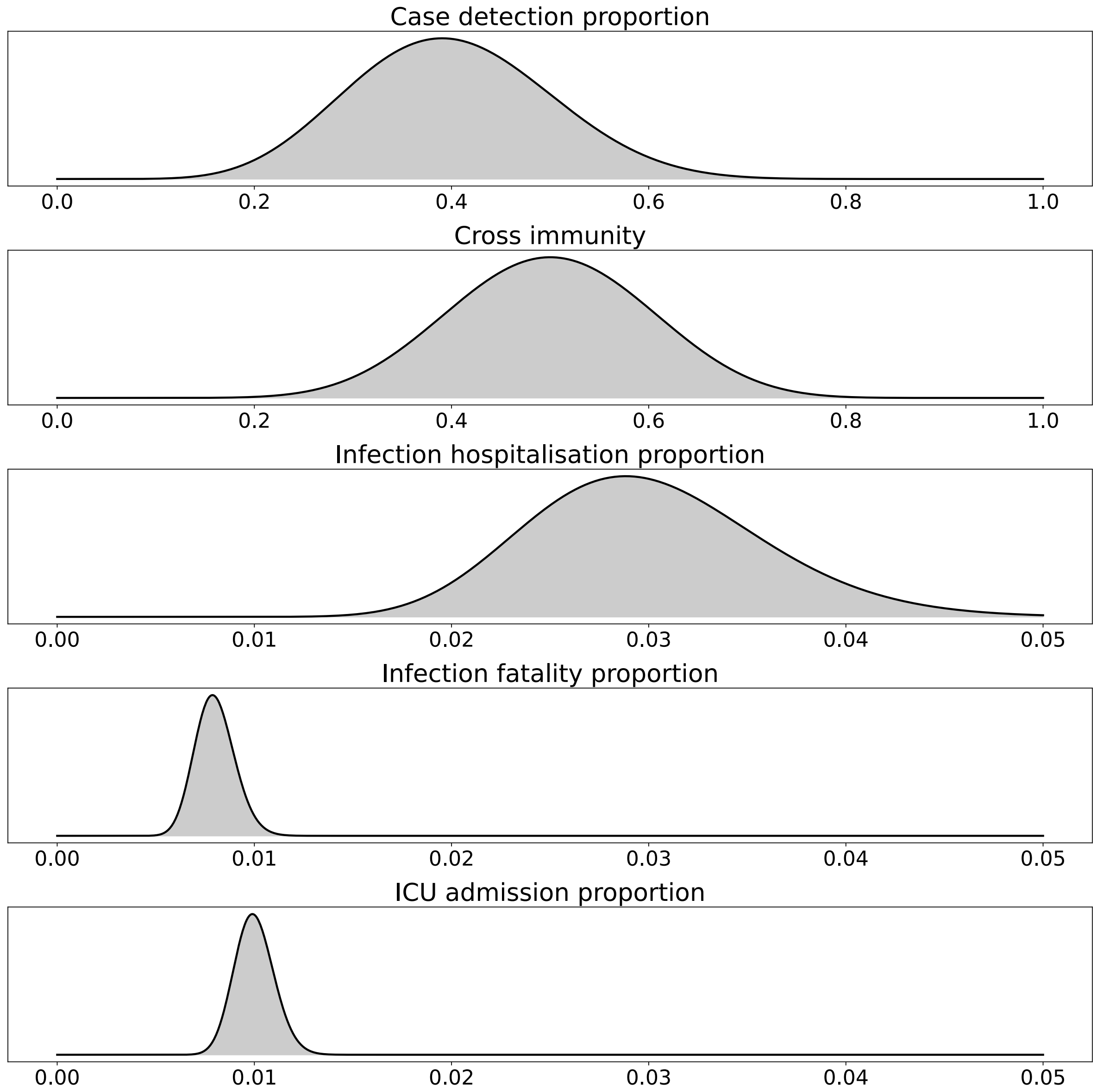

Figure 3: Proportion parameter prior distributions. Note horizontal axis range differs between upper two panels and lower three panels.

#### Fixed parameters

Table 4: Fixed parameters and supporting evidence.

| | Value | Evidence |
|---|---|---|
| Vaccination protection against hospitalisation | 0.5 | The first estimate of the efficacy of BNT162b2 mRNA vaccine that was widely used in Australia, found a total efficacy of 95% protection against COVID-19.[49]<br><br>However, protection against infection will be captured by scaling the modelled transmission parameters, including the residual transmission. As such, the evidence needed for this quantity is the real-world reduction in the risk of hospitalisation and death given infection.<br><br>Estimates of the real-world protection of vaccination against SARS-CoV-2 infections found 85% protection (95% confidence interval 81 to 89%) against infection, but higher protection against severe outcomes, including hospitalisation (93%), ICU admission (98%) and death (95%).[50]<br><br>As such, we estimated the reduction in hospitalisation given infection to be 50%. |
| Vaccination protection against death | 0.67 | Using the same evidence as for the protection against hospitalisation, we estimated this parameter at 67%. |

#### Between-variant relative parameters

Estimates of the relative severity of Alpha compared to preceding strains include a large community-based study from England, which reported a 1.73-fold adjusted hazard of death and a 1.62-fold hazard of hospital admission for Alpha versus wild-type[51]. In Scotland, a genomics-linked cohort analysis incorporating graded clinical severity found a 1.40-fold increase in overall disease severity associated with Alpha[52].

For Delta compared with Alpha, a cohort study from the UK estimated an adjusted hazard ratio of 2.26 for hospital admission and 1.45 for the composite outcome of hospital admission or emergency care[53], with similar estimates observed among unvaccinated individuals. However, an independent national analysis from Norway did not identify an increased risk of hospitalisation for Delta relative to Alpha[54].
