## Supplemental File 2 for "Quantifying the contribution of policy measures to COVID-19 dynamics across 126 countries"

### Calibration results

**Commits used for analyses** **133**

#### Purpose

This document presents the comparison of model outputs to the target indicators used for fitting for each country-analysis combination.

#### Country grouping

For this document and the following supplemental files, we grouped countries according to the following continent groupings.

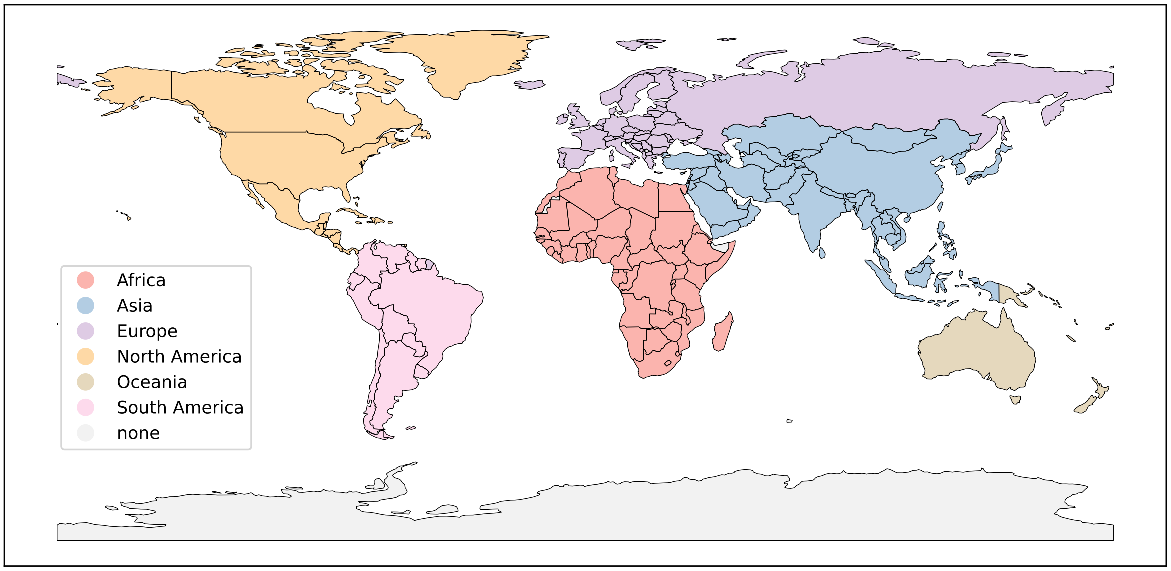

#### Results by continent and country

### Africa

#### Algeria

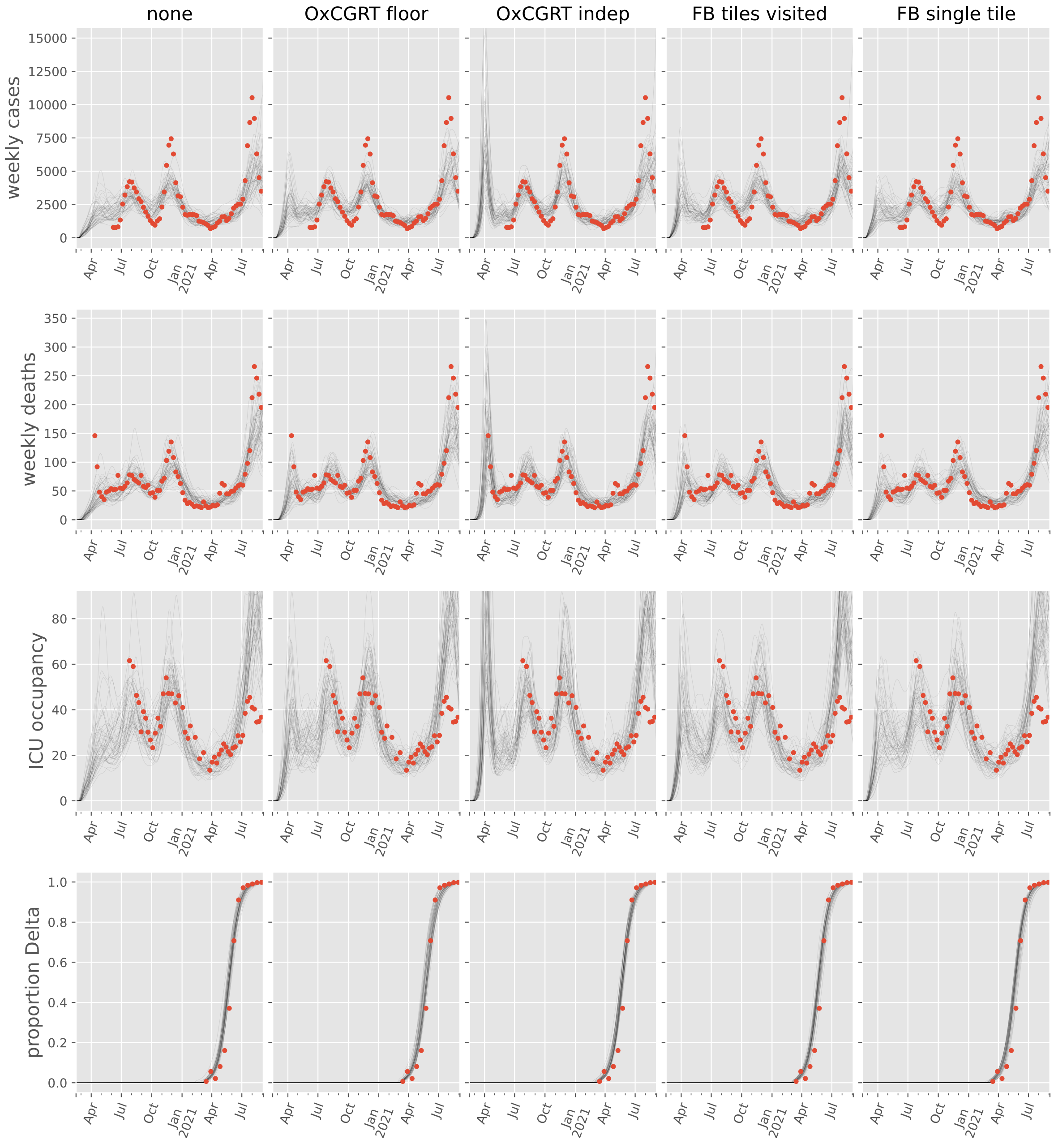

#### Angola

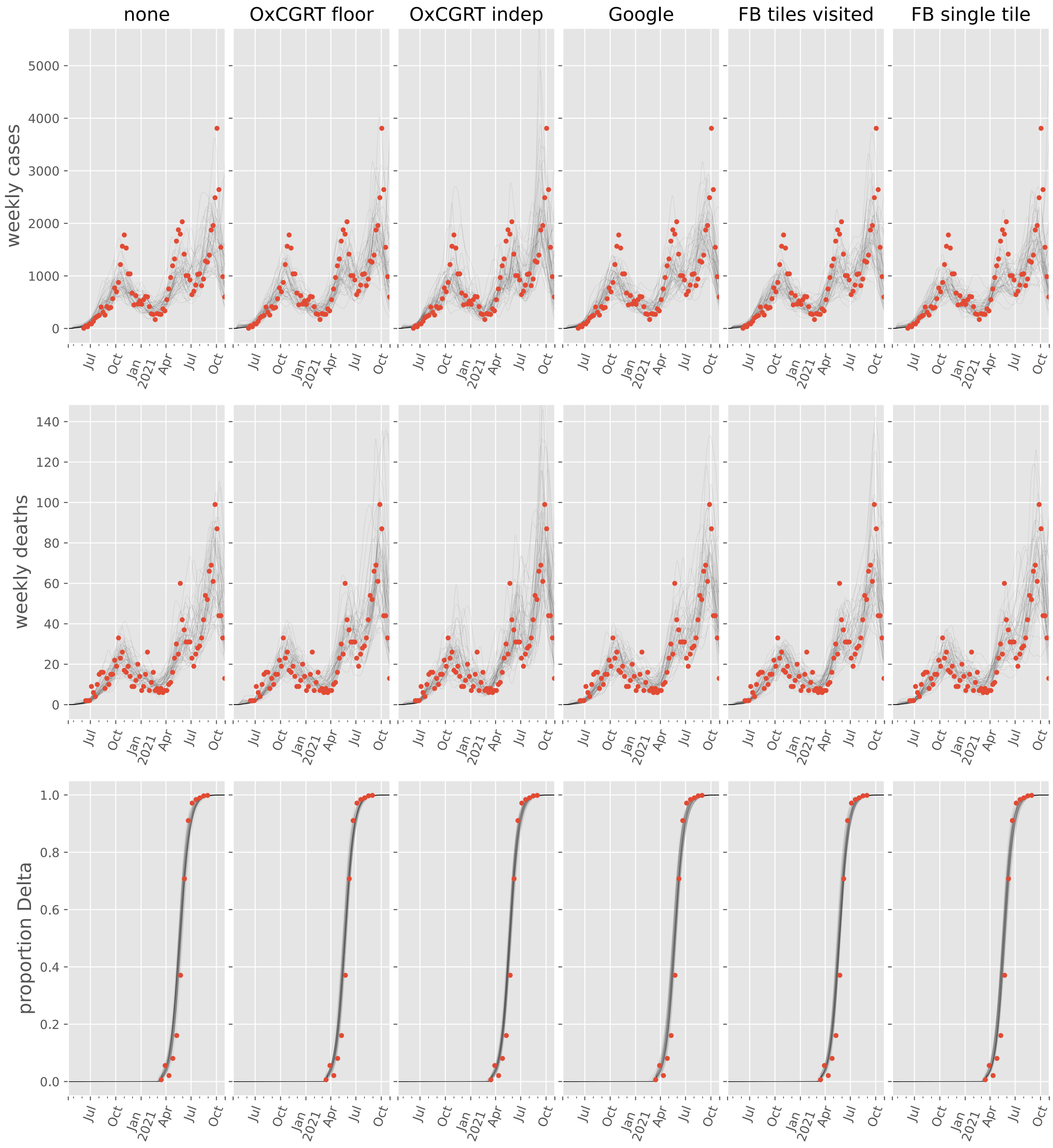
none
OxCGRT floor
OxCGRT indep
Google
FB tiles visited
FB single tile
weekly cases
weekly deaths
proportion Delta
0
1000
2000
3000
4000
5000
0
20
40
60
80
100
120
140
0.0
0.2
0.4
0.6
0.8
1.0
Jul
Oct
Jan
2021
Apr
Jul
Oct

#### Benin

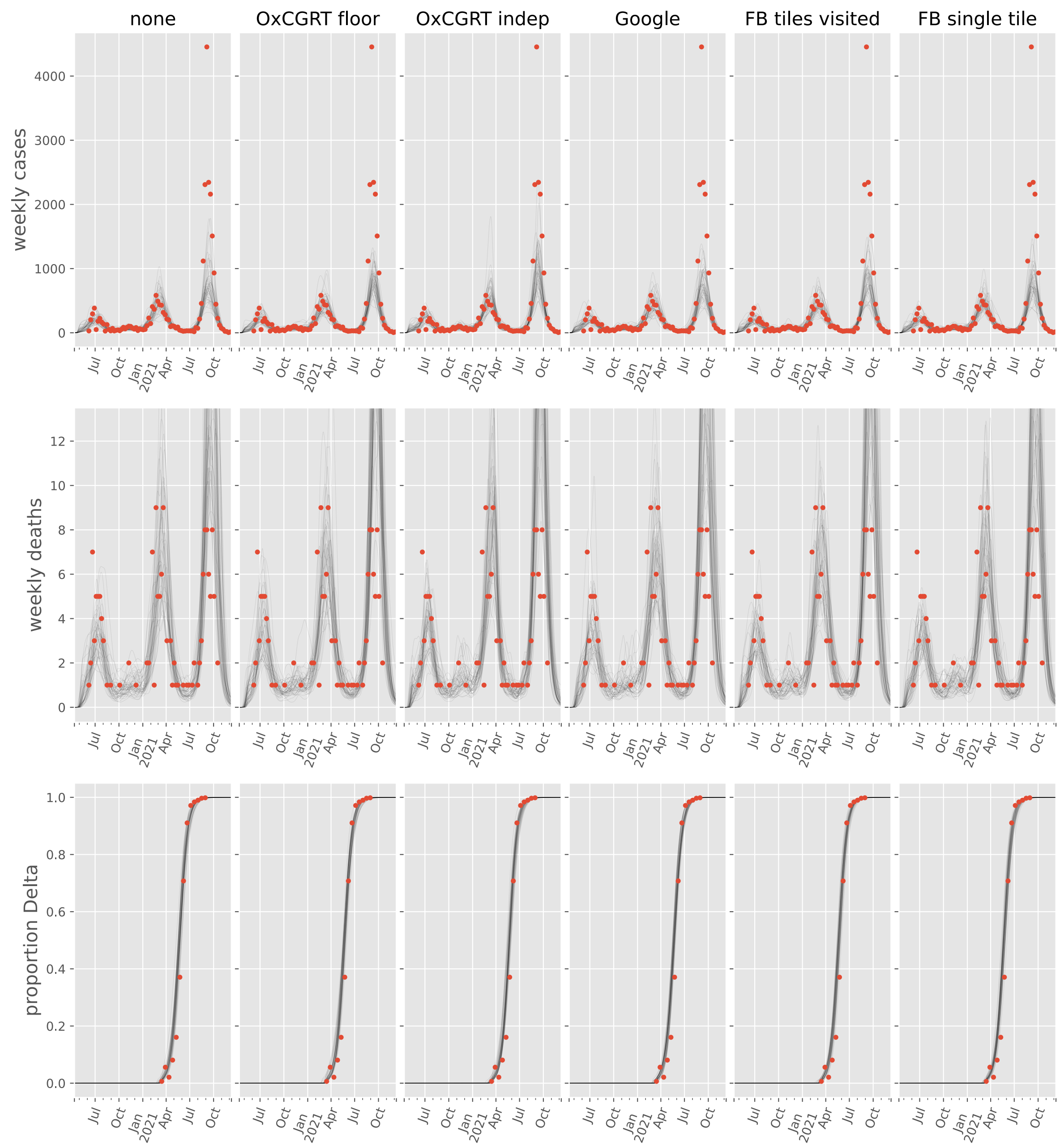
none
OxCGRT floor
OxCGRT indep
Google
FB tiles visited
FB single tile
weekly cases
weekly deaths
proportion Delta
Jul
Oct
Jan
2021
Apr
Jul
Oct

#### Burkina Faso

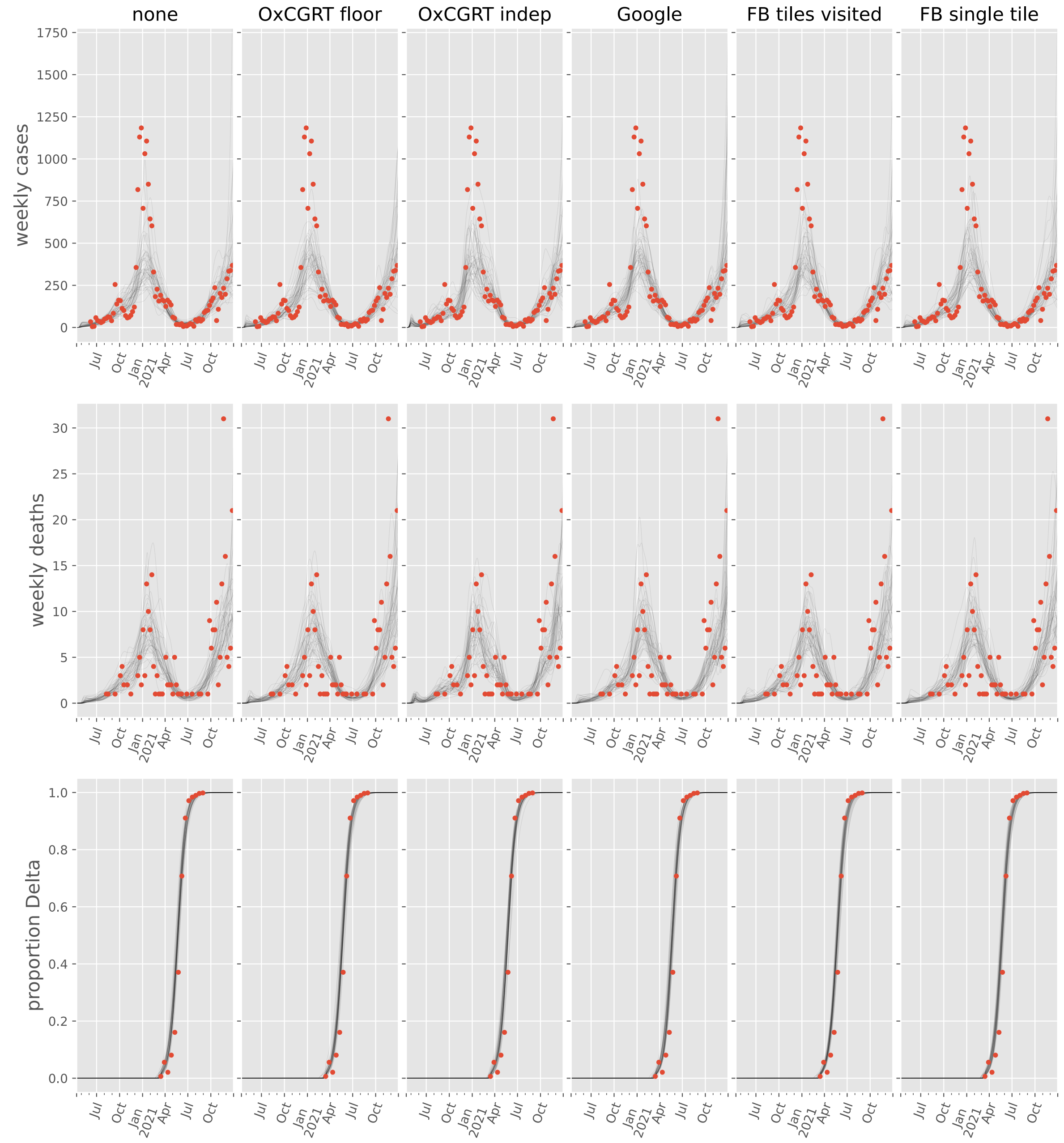

#### Cabo Verde

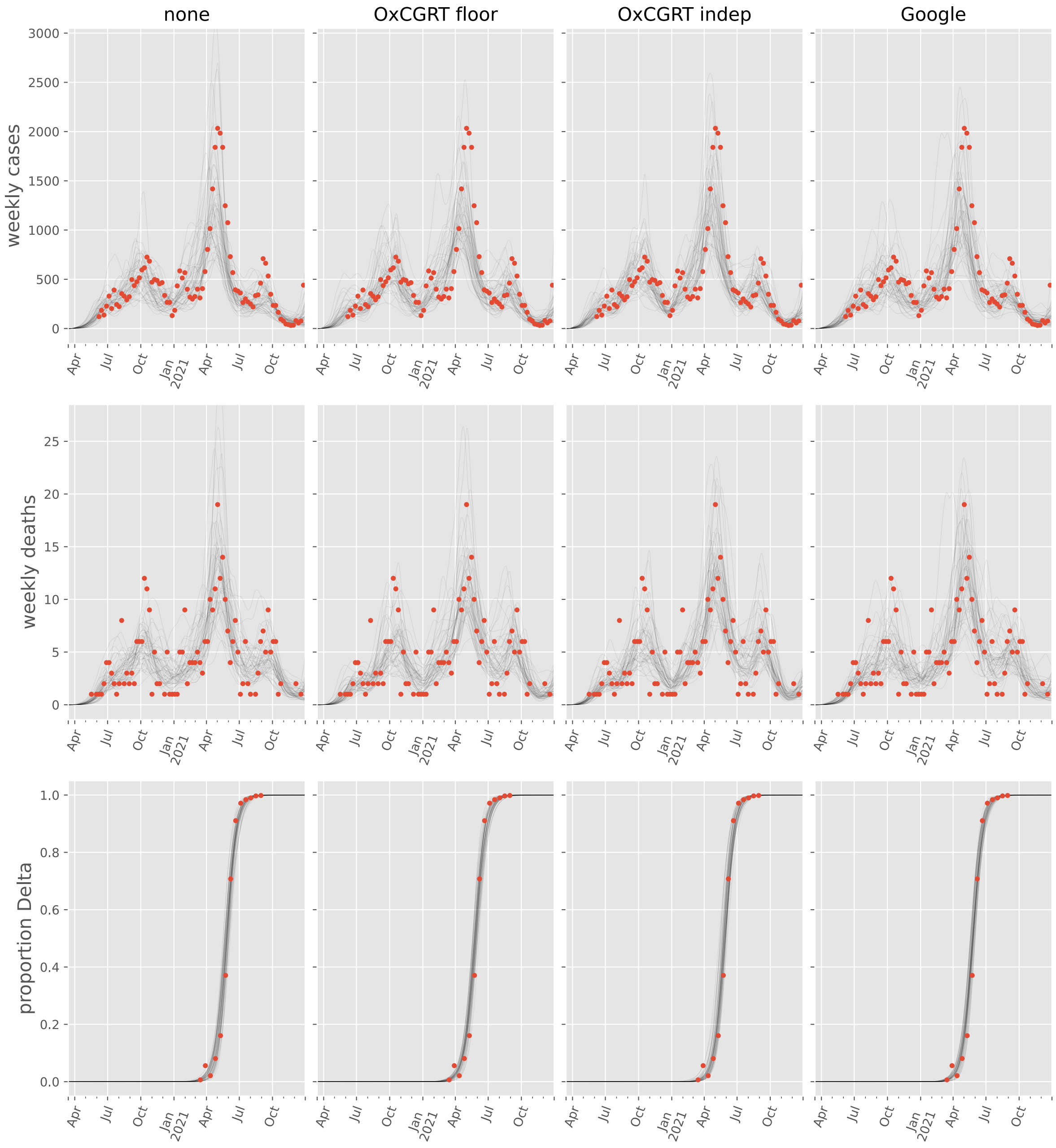

none
OxCGRT floor
OxCGRT indep
Google
weekly cases
weekly deaths
proportion Delta
3000
2500
2000
1500
1000
500
0
25
20
15
10
5
1.0
0.8
0.6
0.4
0.2
0.0
Apr
Jul
Oct
Jan 2021
Apr
Jul
Oct

#### Cameroon

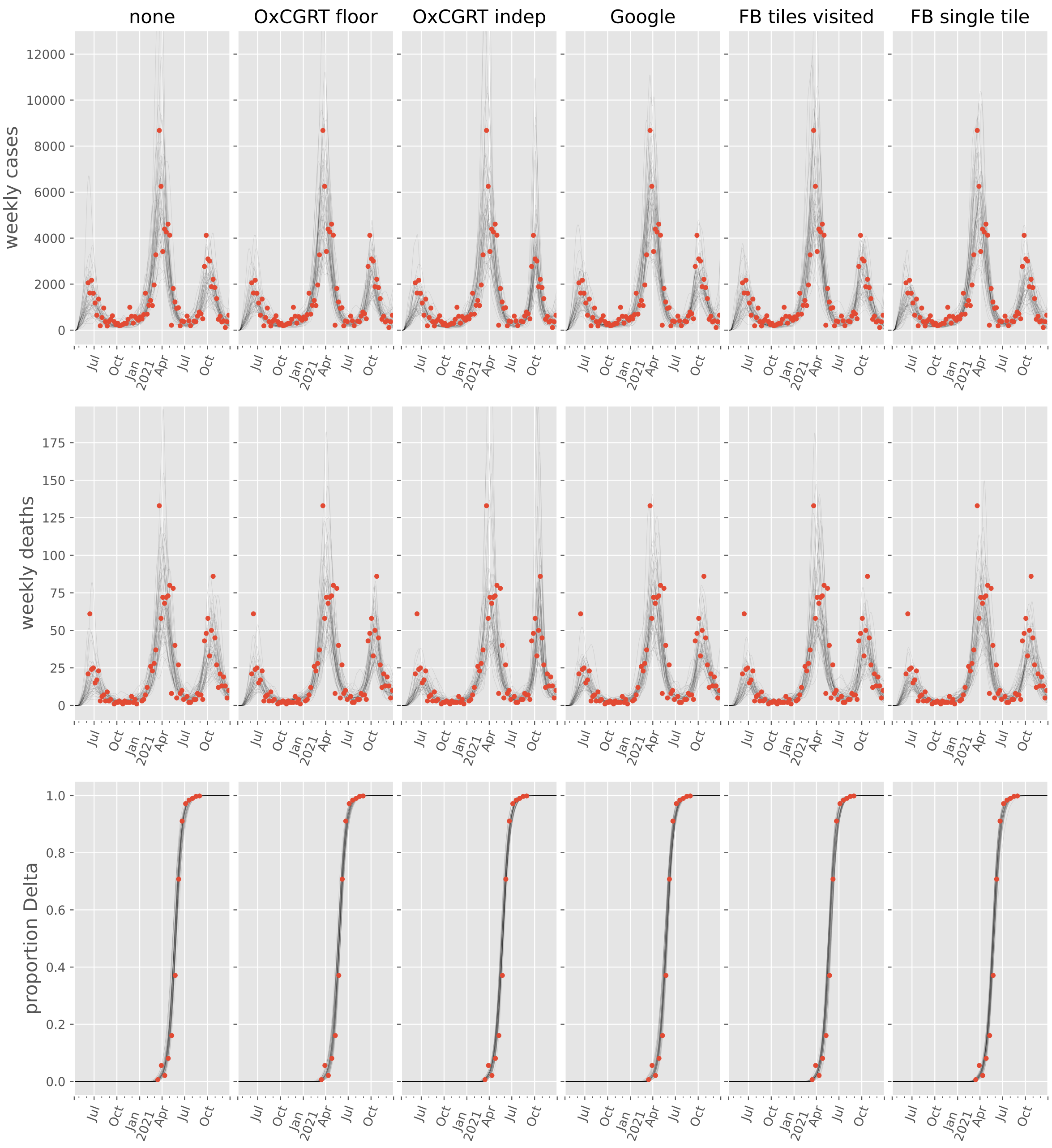

none
OxCGRT floor
OxCGRT indep
Google
FB tiles visited
FB single tile
weekly cases
weekly deaths
proportion Delta
12000
10000
8000
6000
4000
2000
0
175
150
125
100
75
50
25
1.0
0.8
0.6
0.4
0.2
0.0
Jul
Oct
Jan
2021
Apr

#### Congo

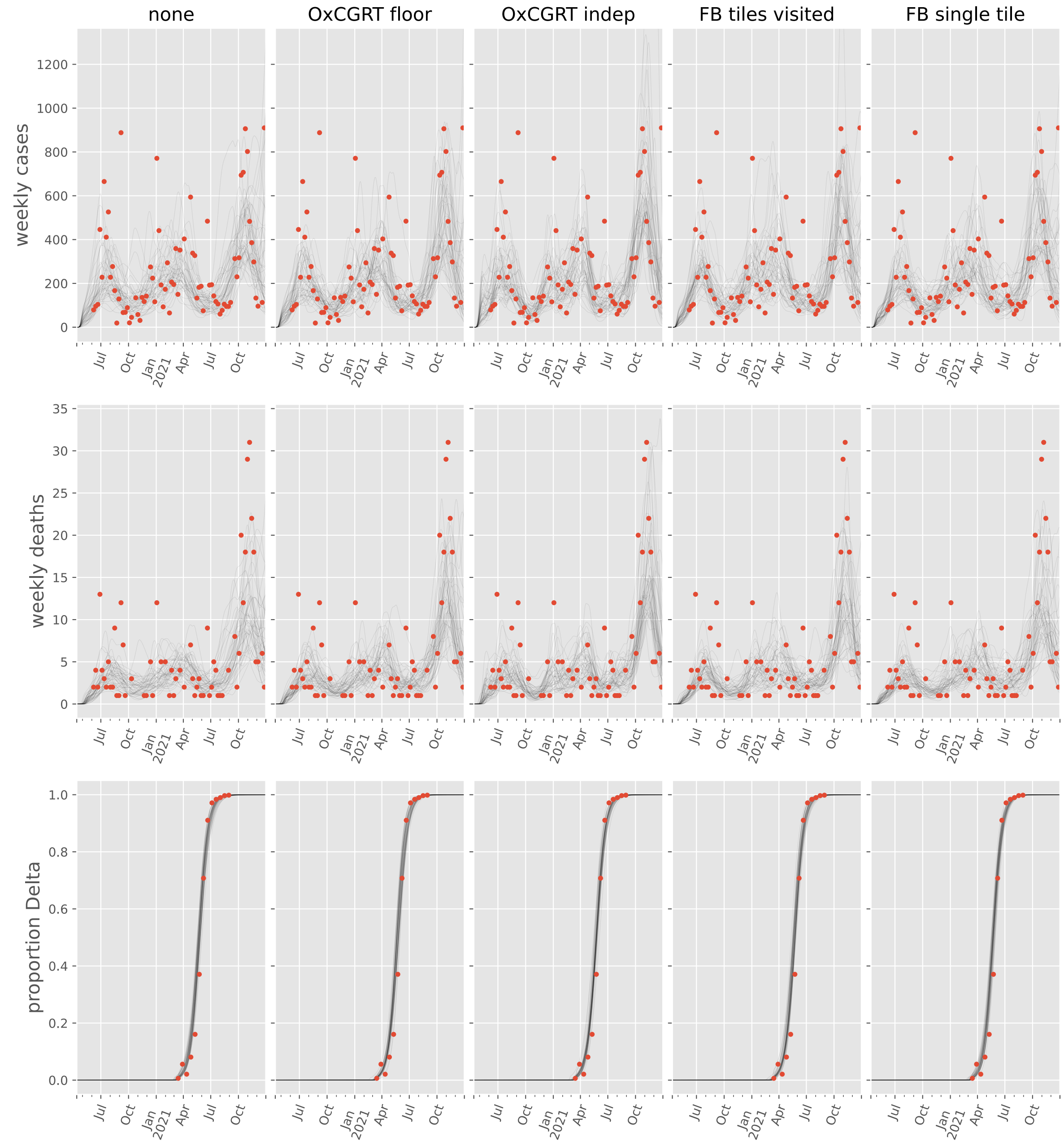

none
OxCGRT floor
OxCGRT indep
FB tiles visited
FB single tile
weekly cases
weekly deaths
proportion Delta
0
200
400
600
800
1000
1200
0
5
10
15
20
25
30
35
0.0
0.2
0.4
0.6
0.8
1.0
Jul
Oct
Jan 2021
Apr
Jul
Oct

#### Côte d'Ivoire

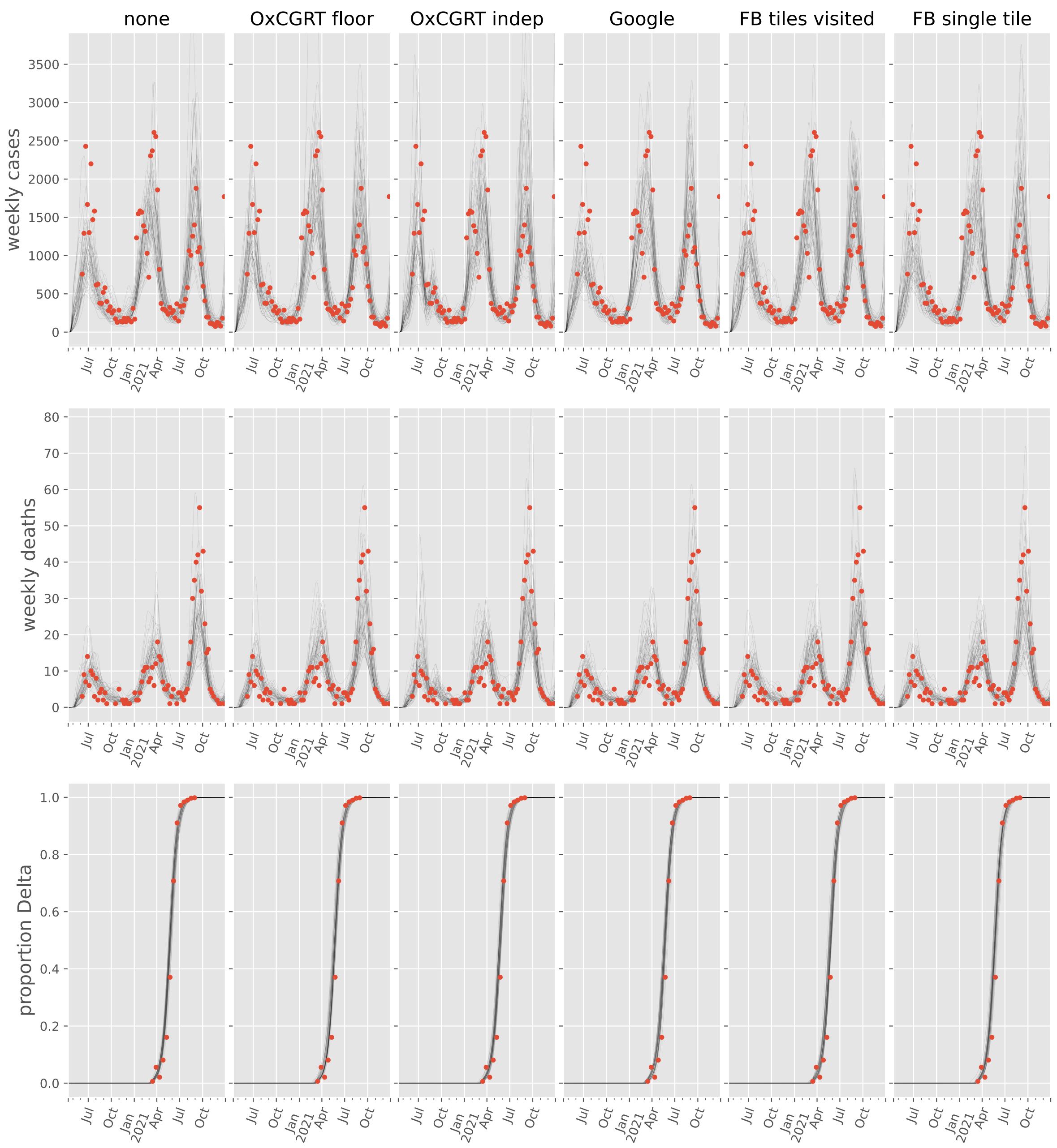

#### Egypt

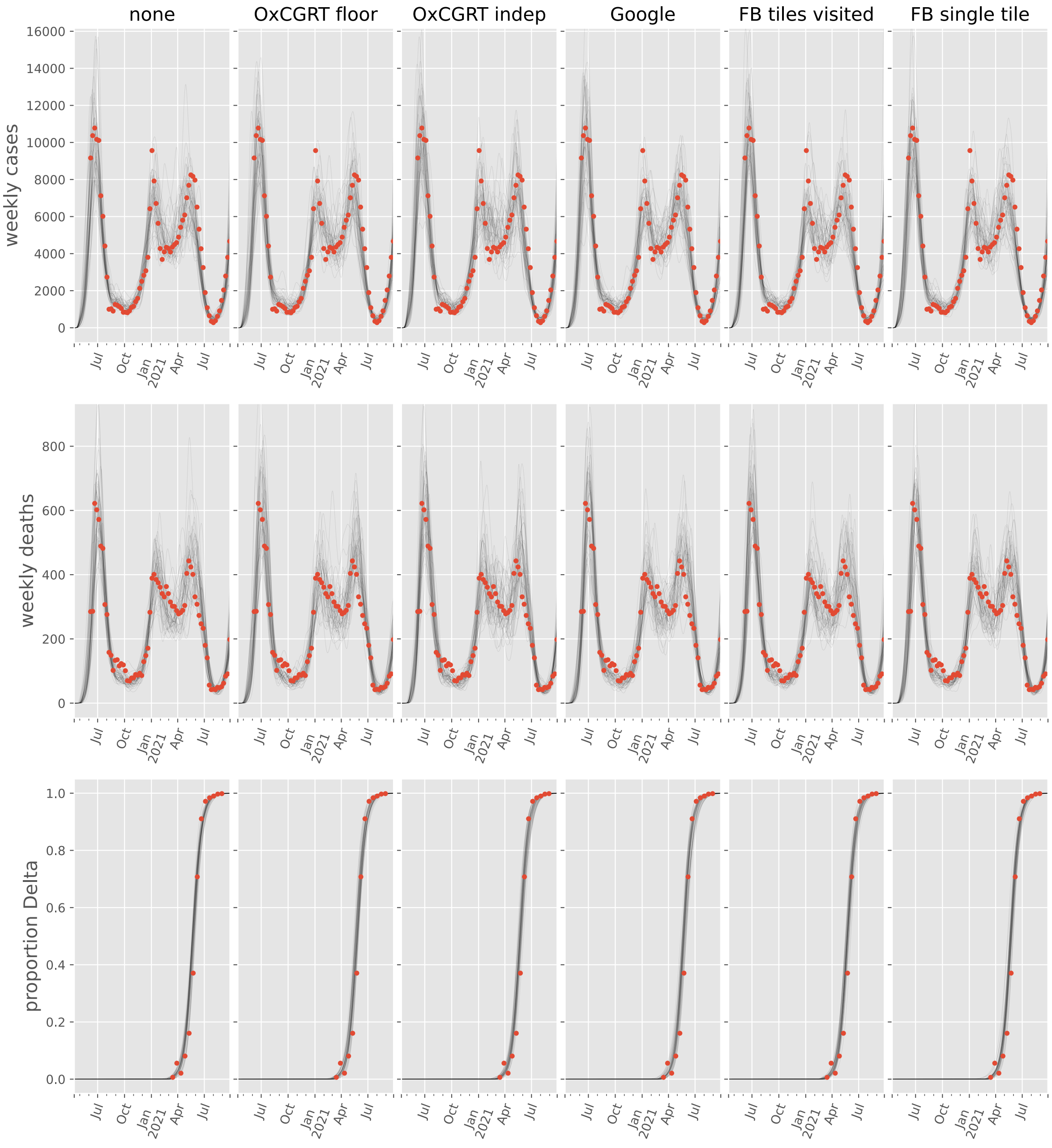
none
OxCGRT floor
OxCGRT indep
Google
FB tiles visited
FB single tile
weekly cases
weekly deaths
proportion Delta
0
2000
4000
6000
8000
10000
12000
14000
16000
200
400
600
800
0.0
0.2
0.4
0.6
0.8
1.0
Jul
Oct
Jan 2021
Apr
Jul

#### Eswatini

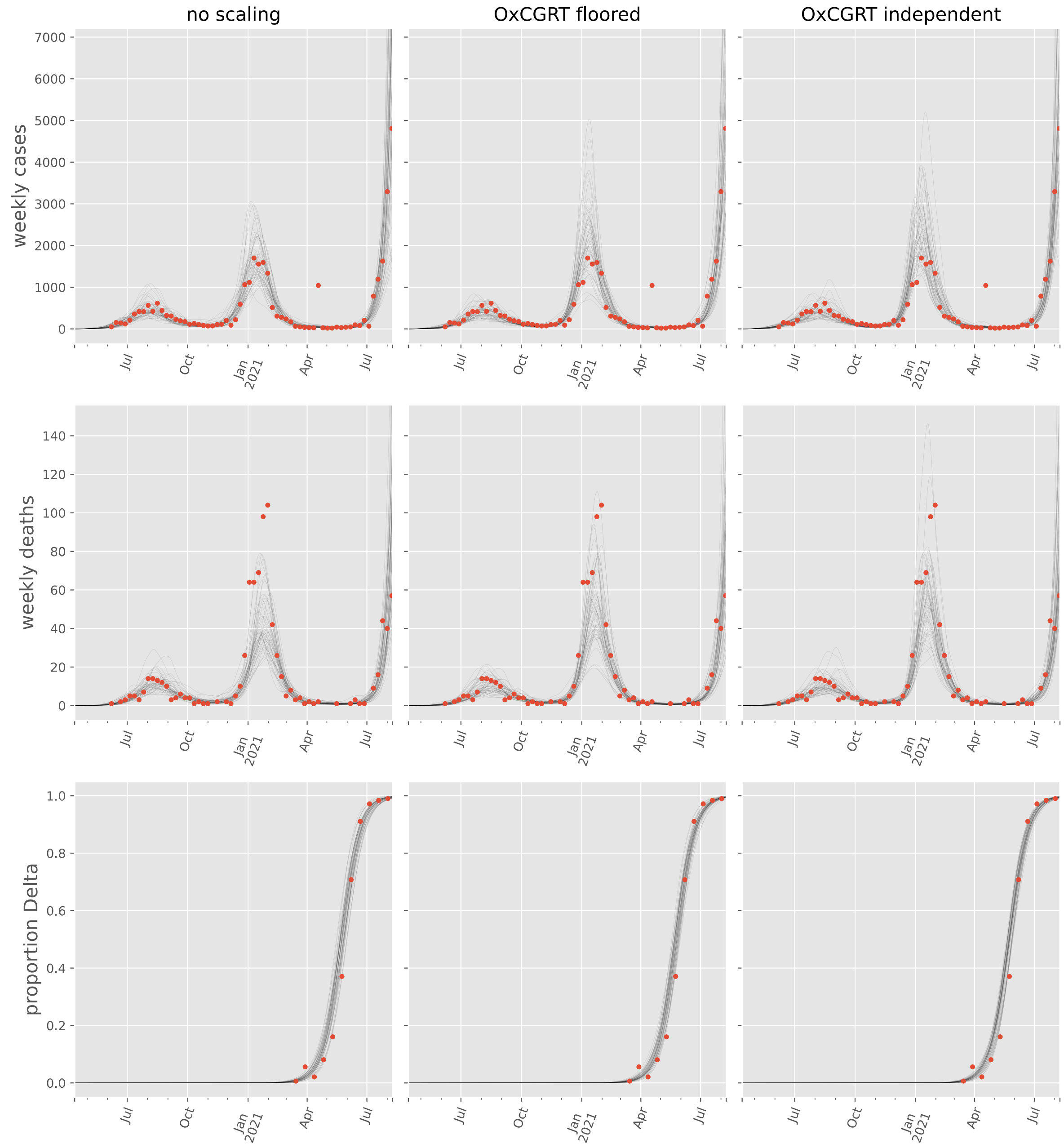

no scaling
OxCGRT floored
OxCGRT independent
weekly cases
weekly deaths
proportion Delta
7000
6000
5000
4000
3000
2000
1000
0
140
120
100
80
60
40
20
1.0
0.8
0.6
0.4
0.2
0.0
Jul
Oct
Jan 2021
Apr
Jul

#### Ethiopia

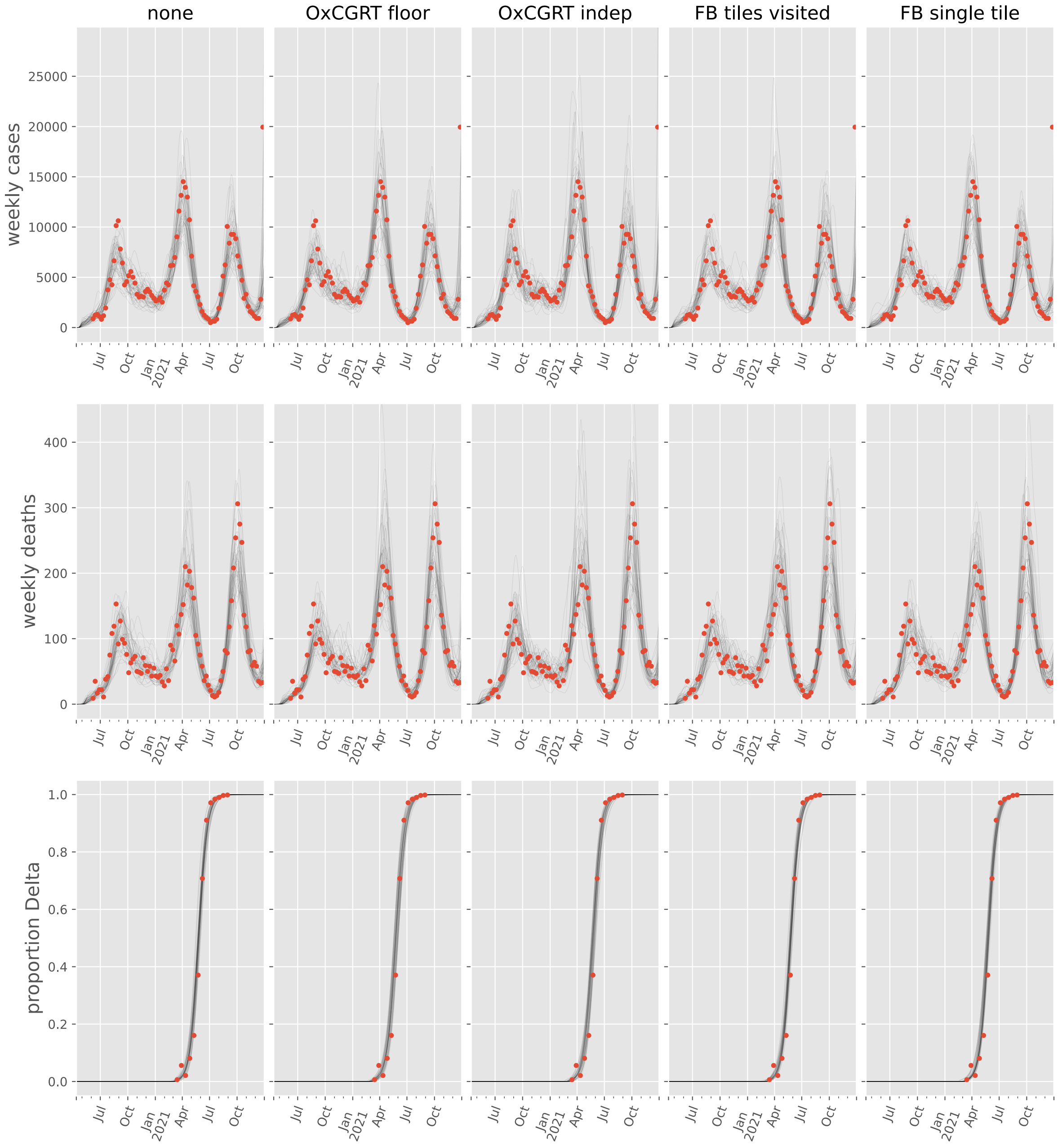
none
OxCGRT floor
OxCGRT indep
FB tiles visited
FB single tile
weekly cases
weekly deaths
proportion Delta
25000
20000
15000
10000
5000
0
400
300
200
100
1.0
0.8
0.6
0.4
0.2
0.0
Jul
Oct
Jan 2021
Apr
Jul
Oct

#### Gabon

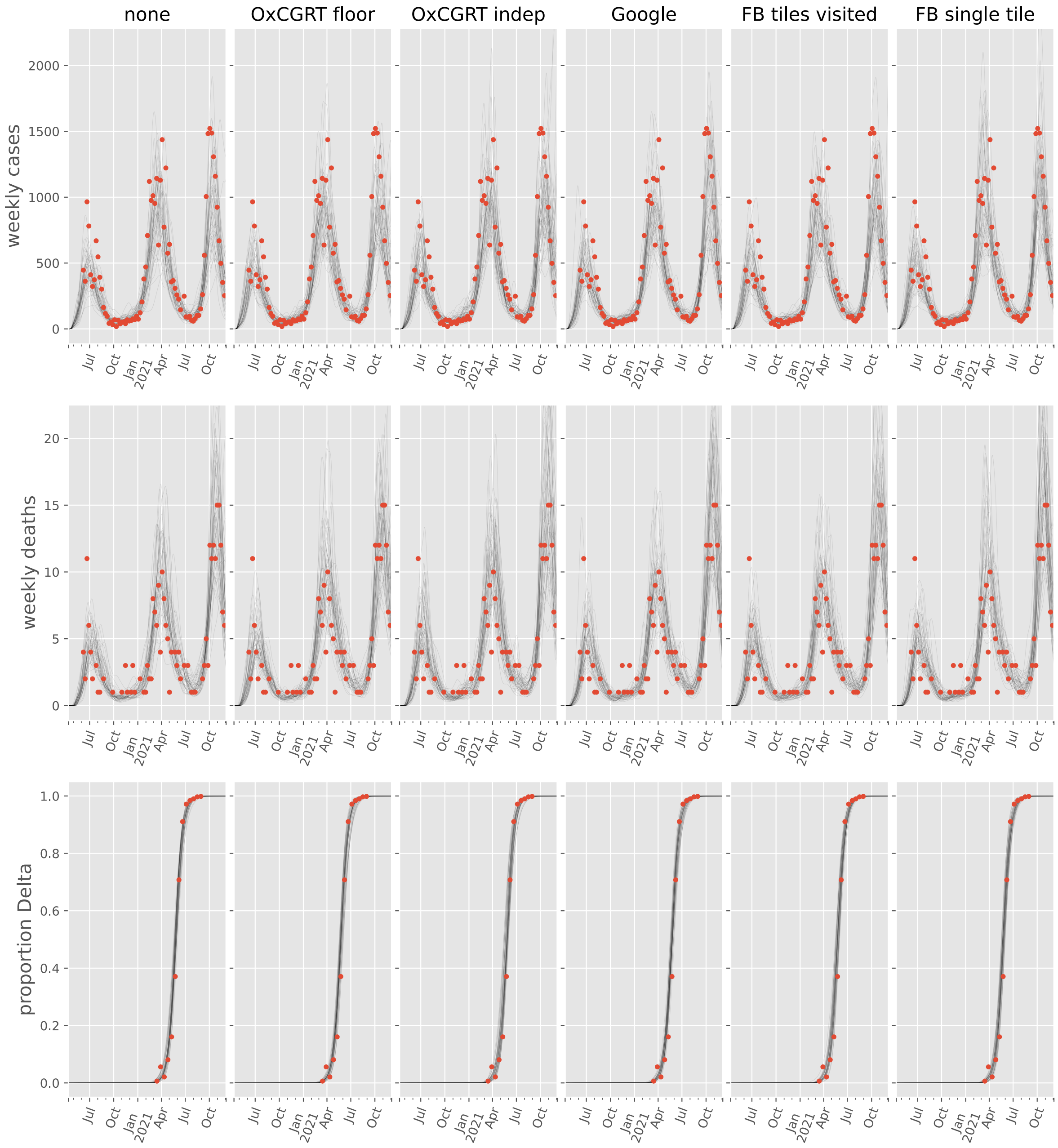

none
OxCGRT floor
OxCGRT indep
Google
FB tiles visited
FB single tile
weekly cases
weekly deaths
proportion Delta
2000
1500
1000
500
0
20
15
10
5
1.0
0.8
0.6
0.4
0.2
0.0
Jul
Oct
Jan
2021
Apr

#### Guinea

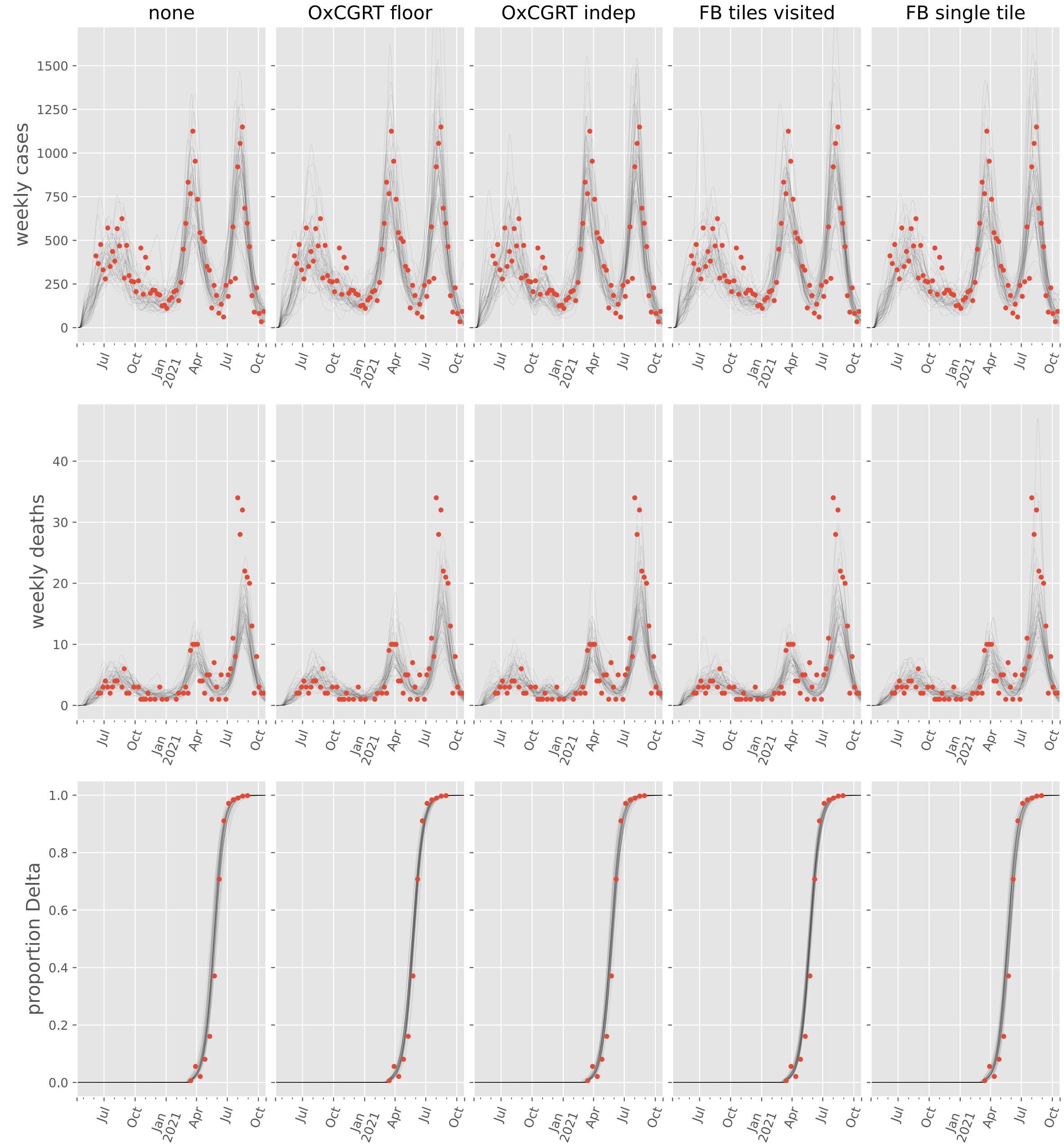

none
OxCGRT floor
OxCGRT indep
FB tiles visited
FB single tile
weekly cases
weekly deaths
proportion Delta
0
250
500
750
1000
1250
1500
0
10
20
30
40
0.0
0.2
0.4
0.6
0.8
1.0
Jul
Oct
Jan
2021
Apr
Jul
Oct

#### Kenya

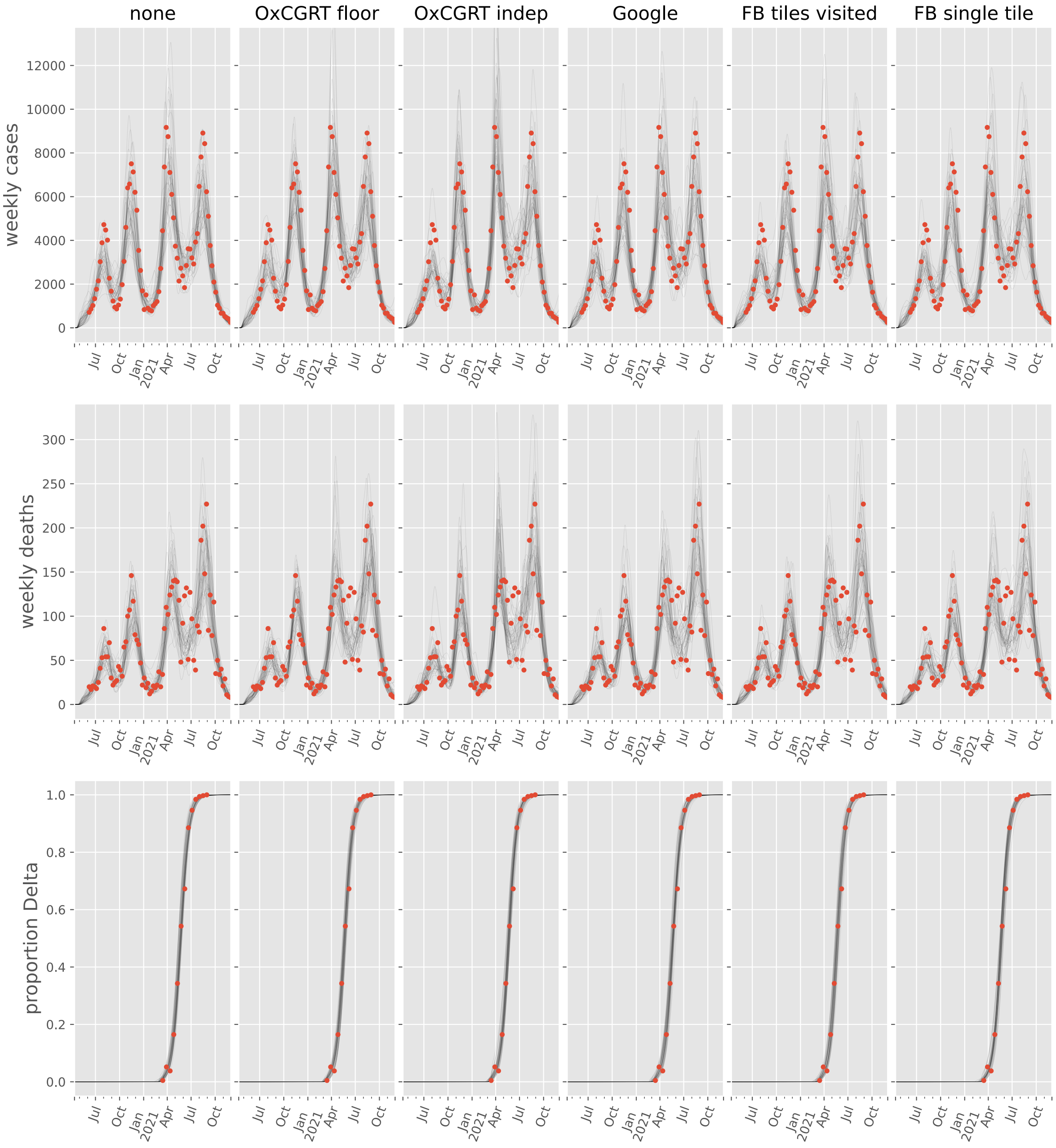

none
OxCGRT floor
OxCGRT indep
Google
FB tiles visited
FB single tile
weekly cases
weekly deaths
proportion Delta
Jul
Oct
Jan 2021
Apr

#### Lesotho

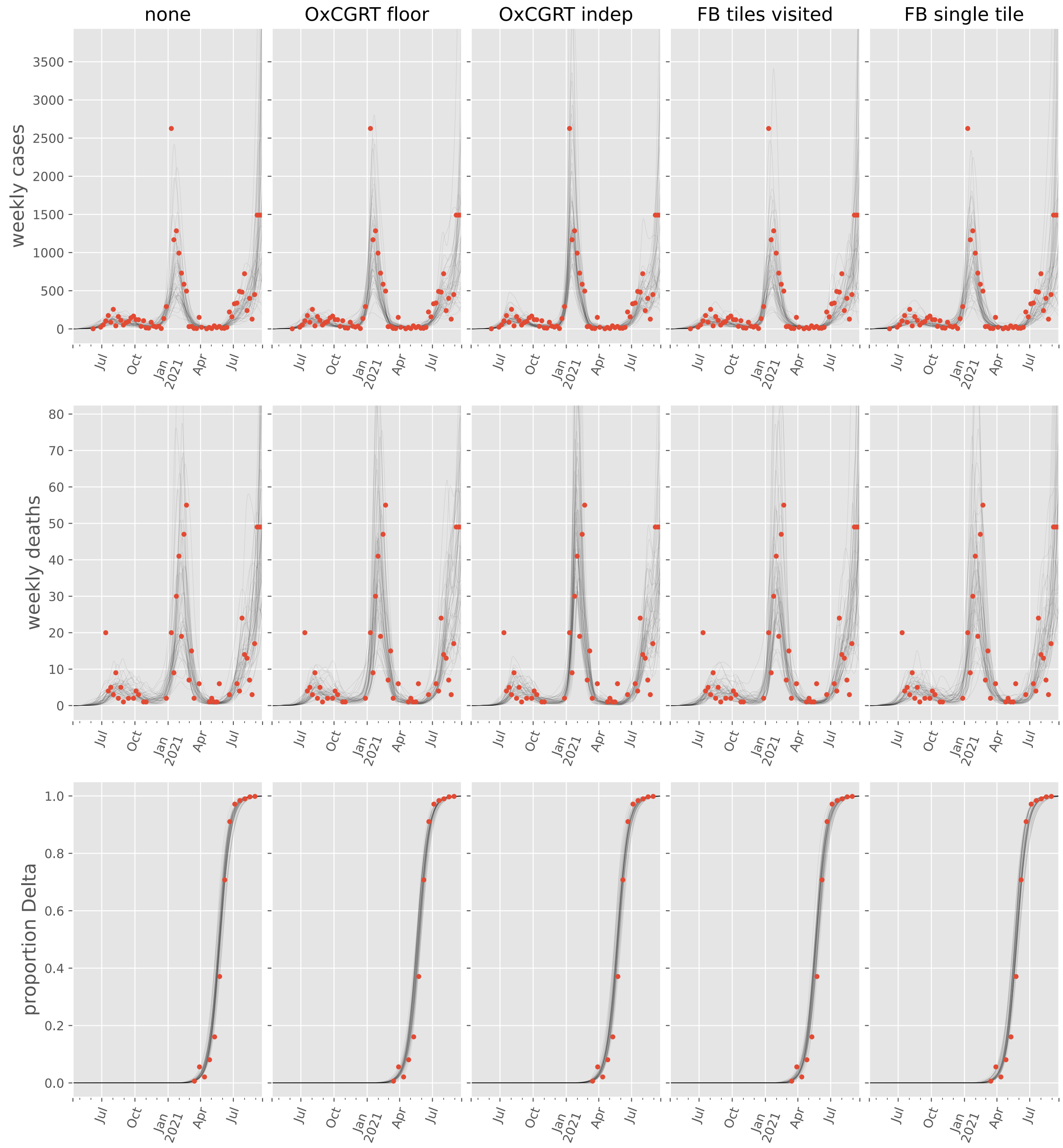

none
OxCGRT floor
OxCGRT indep
FB tiles visited
FB single tile
weekly cases
weekly deaths
proportion Delta
Jul
Oct
Jan
2021
Apr
Jul

#### Liberia

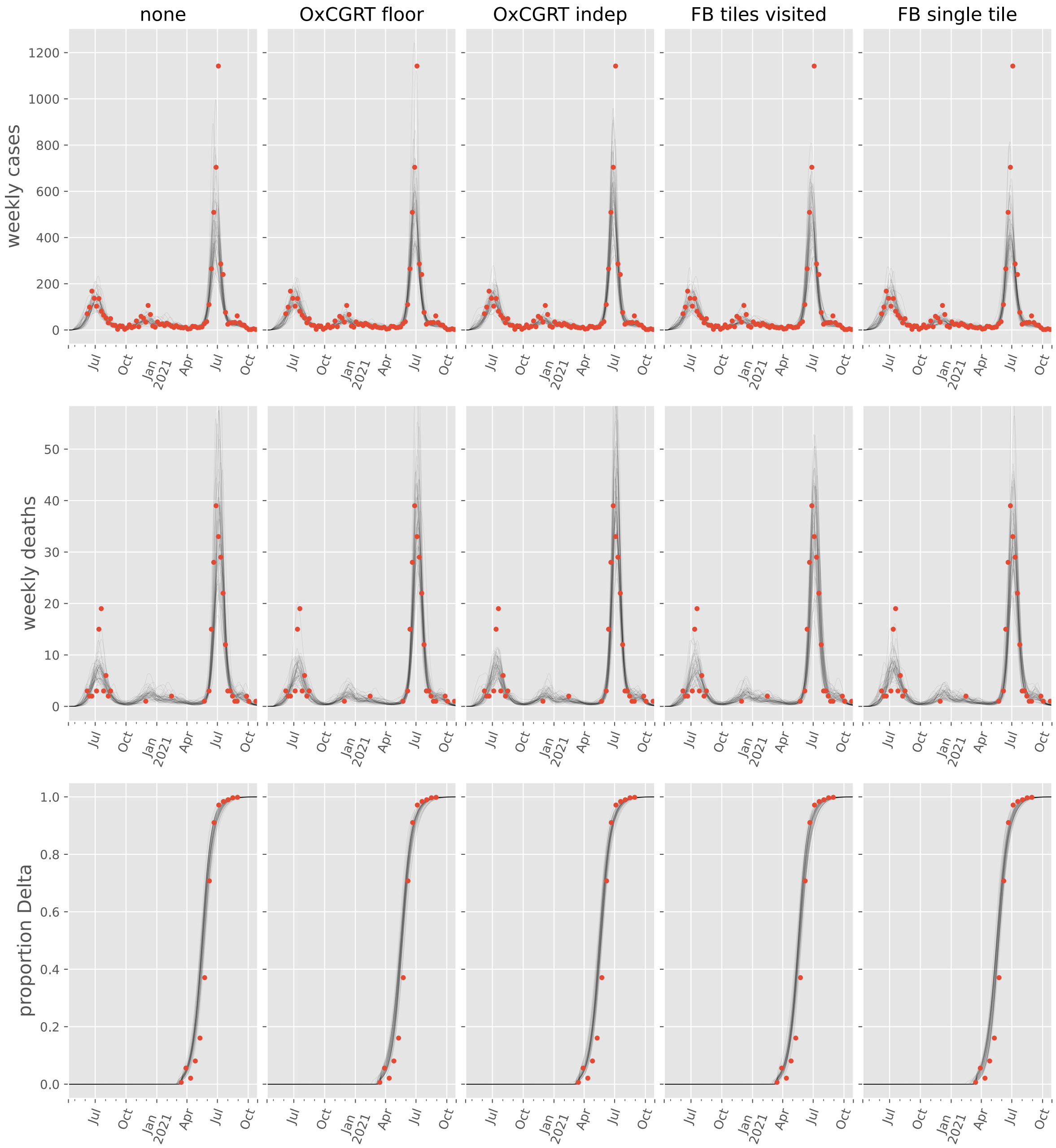

none
OxCGRT floor
OxCGRT indep
FB tiles visited
FB single tile
weekly cases
weekly deaths
proportion Delta
0
200
400
600
800
1000
1200
0
10
20
30
40
50
0.0
0.2
0.4
0.6
0.8
1.0
Jul
Oct
Jan
2021
Apr
Jul
Oct

#### Libya

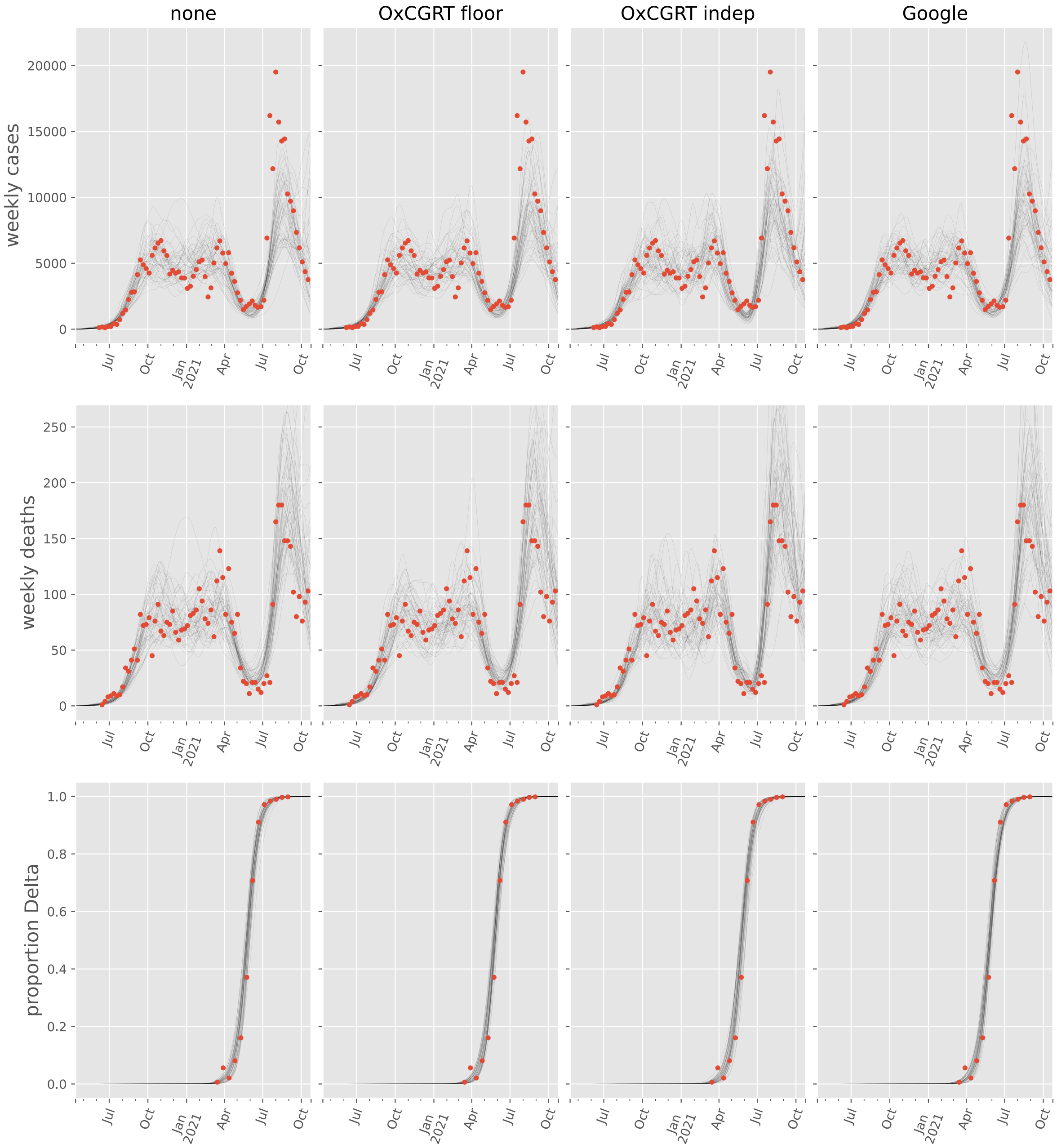

none
OxCGRT floor
OxCGRT indep
Google
weekly cases
weekly deaths
proportion Delta
20000
15000
10000
5000
0
250
200
150
100
50
1.0
0.8
0.6
0.4
0.2
0.0
Jul
Oct
Jan 2021
Apr

#### Madagascar

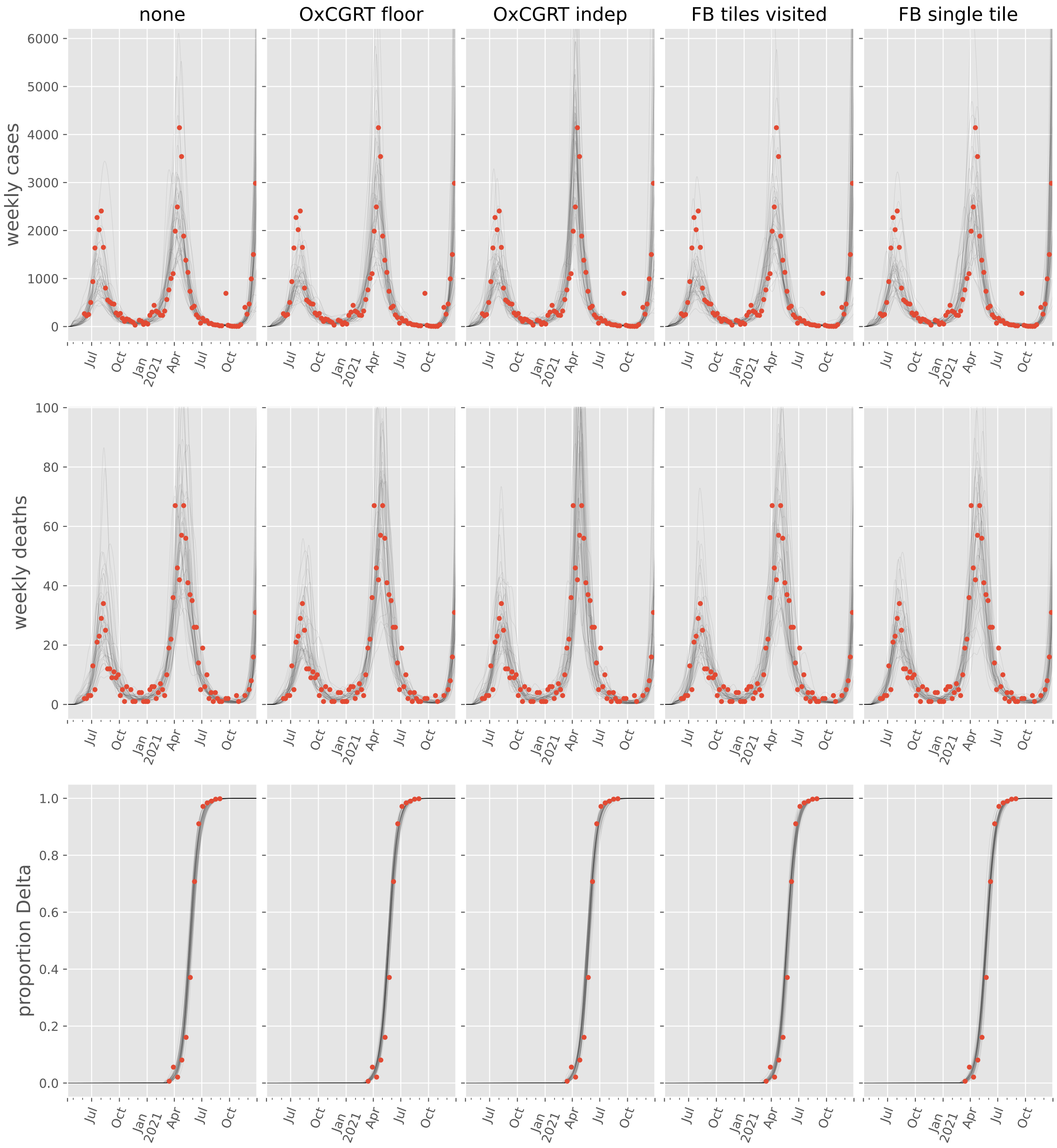

none
OxCGRT floor
OxCGRT indep
FB tiles visited
FB single tile
weekly cases
weekly deaths
proportion Delta
0
1000
2000
3000
4000
5000
6000
20
40
60
80
100
0.0
0.2
0.4
0.6
0.8
1.0
Jul
Oct
Jan
2021
Apr
Jul
Oct

#### Malawi

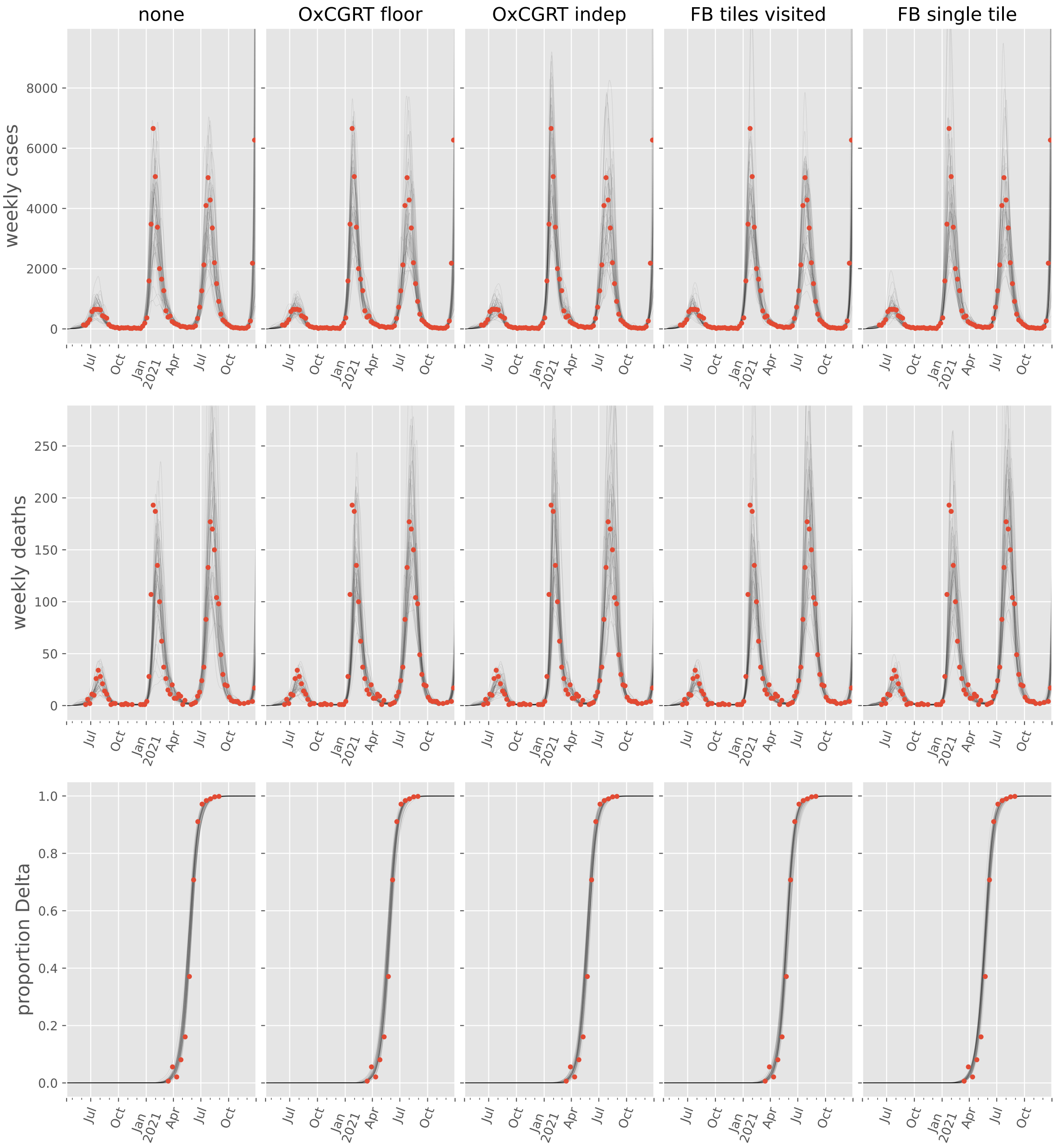

none
OxCGRT floor
OxCGRT indep
FB tiles visited
FB single tile
weekly cases
weekly deaths
proportion Delta
Jul
Oct
Jan
2021
Apr

#### Mali

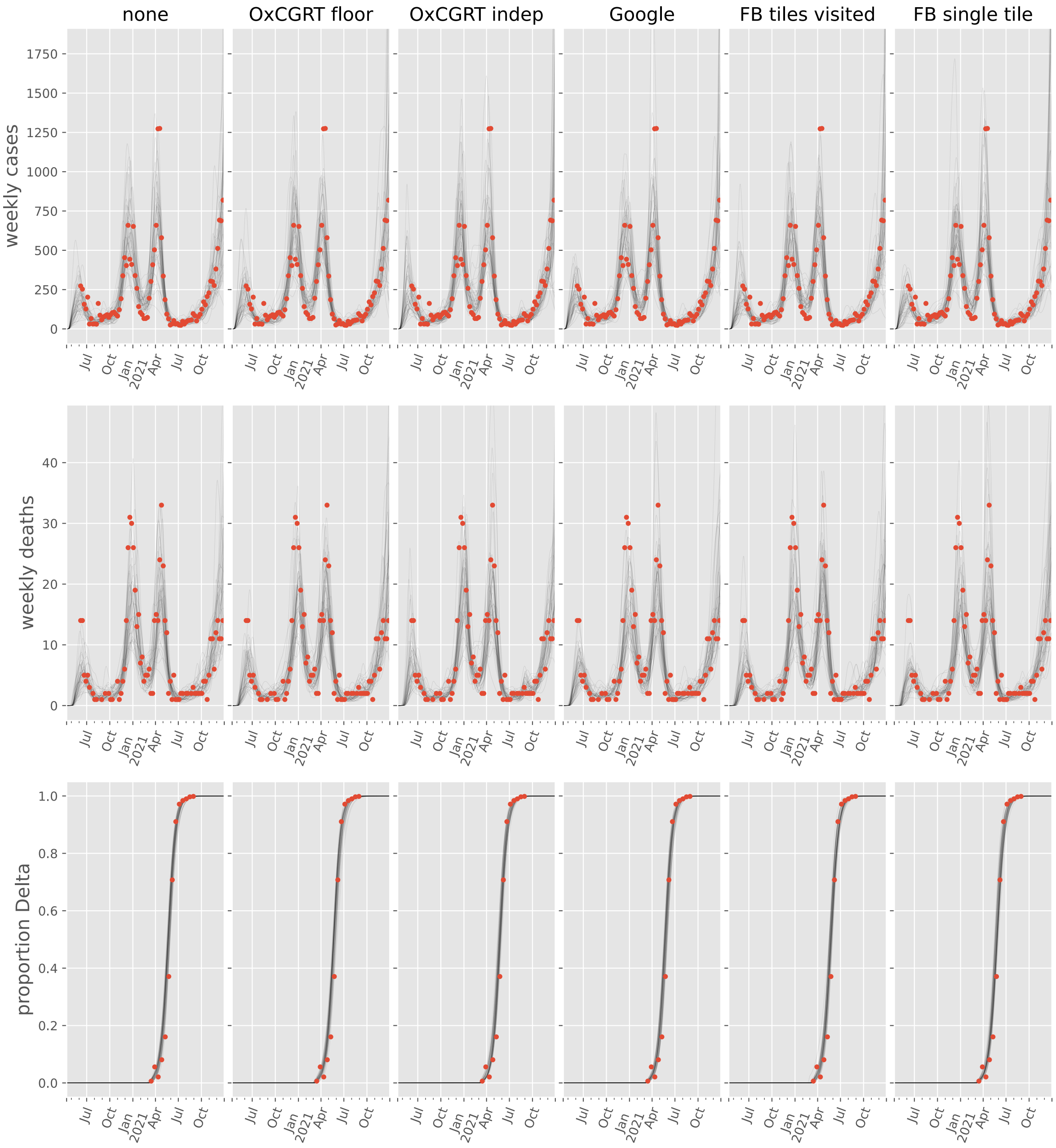

none
OxCGRT floor
OxCGRT indep
Google
FB tiles visited
FB single tile
weekly cases
weekly deaths
proportion Delta
0
250
500
750
1000
1250
1500
1750
10
20
30
40
0.0
0.2
0.4
0.6
0.8
1.0
Jul
Oct
Jan 2021
Apr
Jul
Oct

#### Morocco

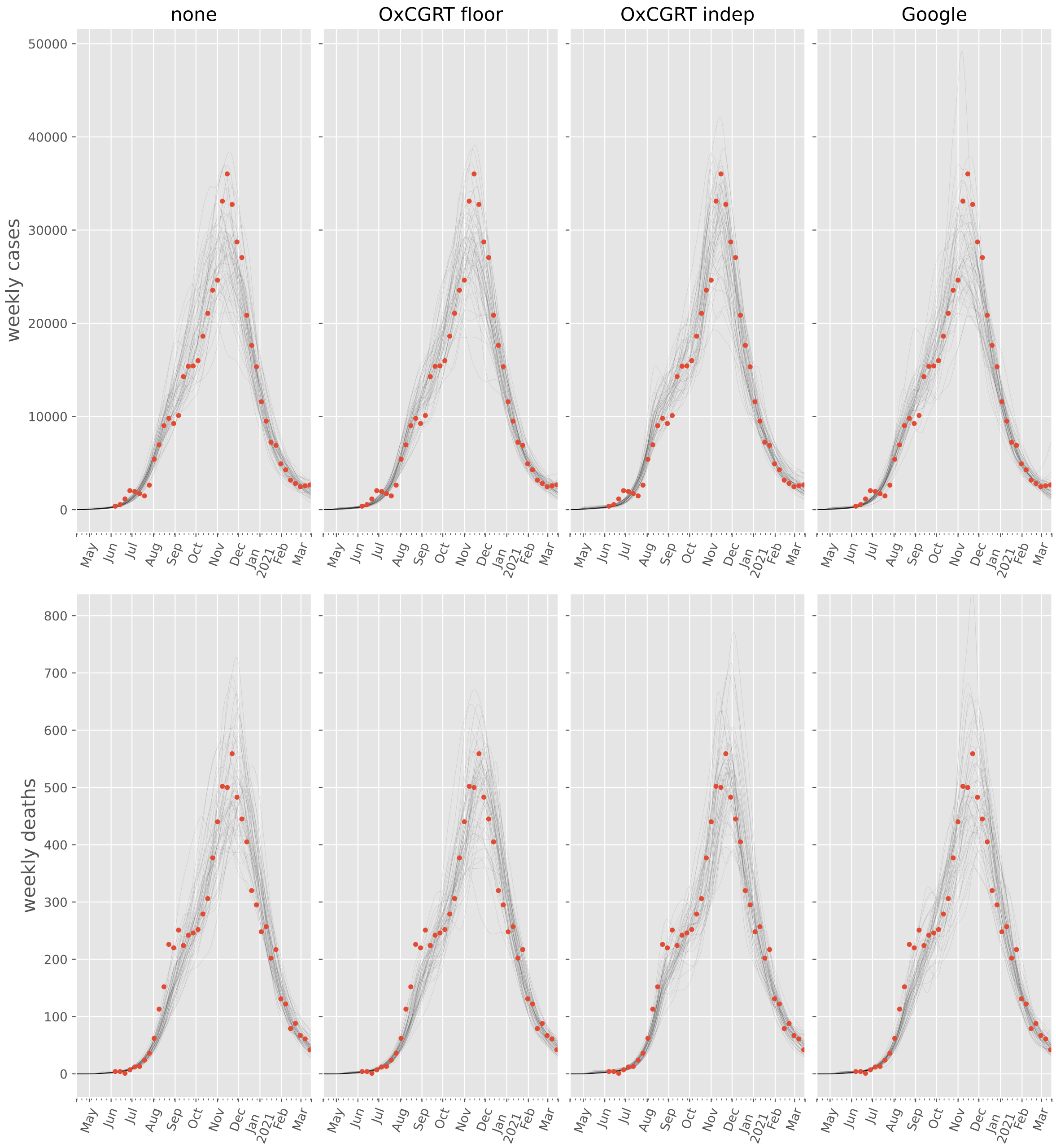

none
OxCGRT floor
OxCGRT indep
Google
weekly cases
weekly deaths
May
Jun
Jul
Aug
Sep
Oct
Nov
Dec
Jan
2021
Feb
Mar

#### Mozambique

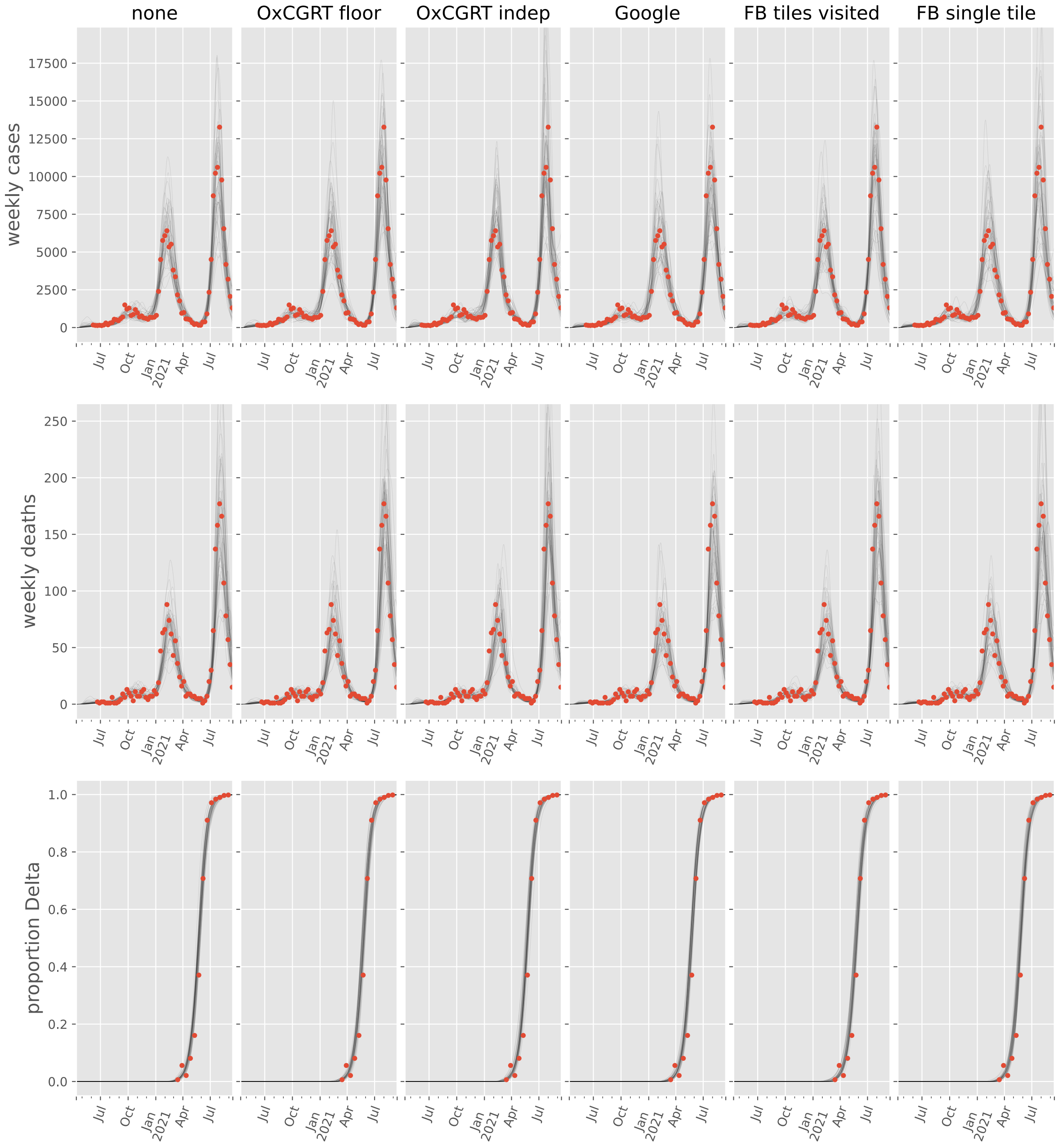
none
OxCGRT floor
OxCGRT indep
Google
FB tiles visited
FB single tile
weekly cases
17500
15000
12500
10000
7500
5000
2500
0
weekly deaths
250
200
150
100
50
0
proportion Delta
1.0
0.8
0.6
0.4
0.2
0.0
Jul
Oct
Jan
2021
Apr
Jul

#### Nigeria

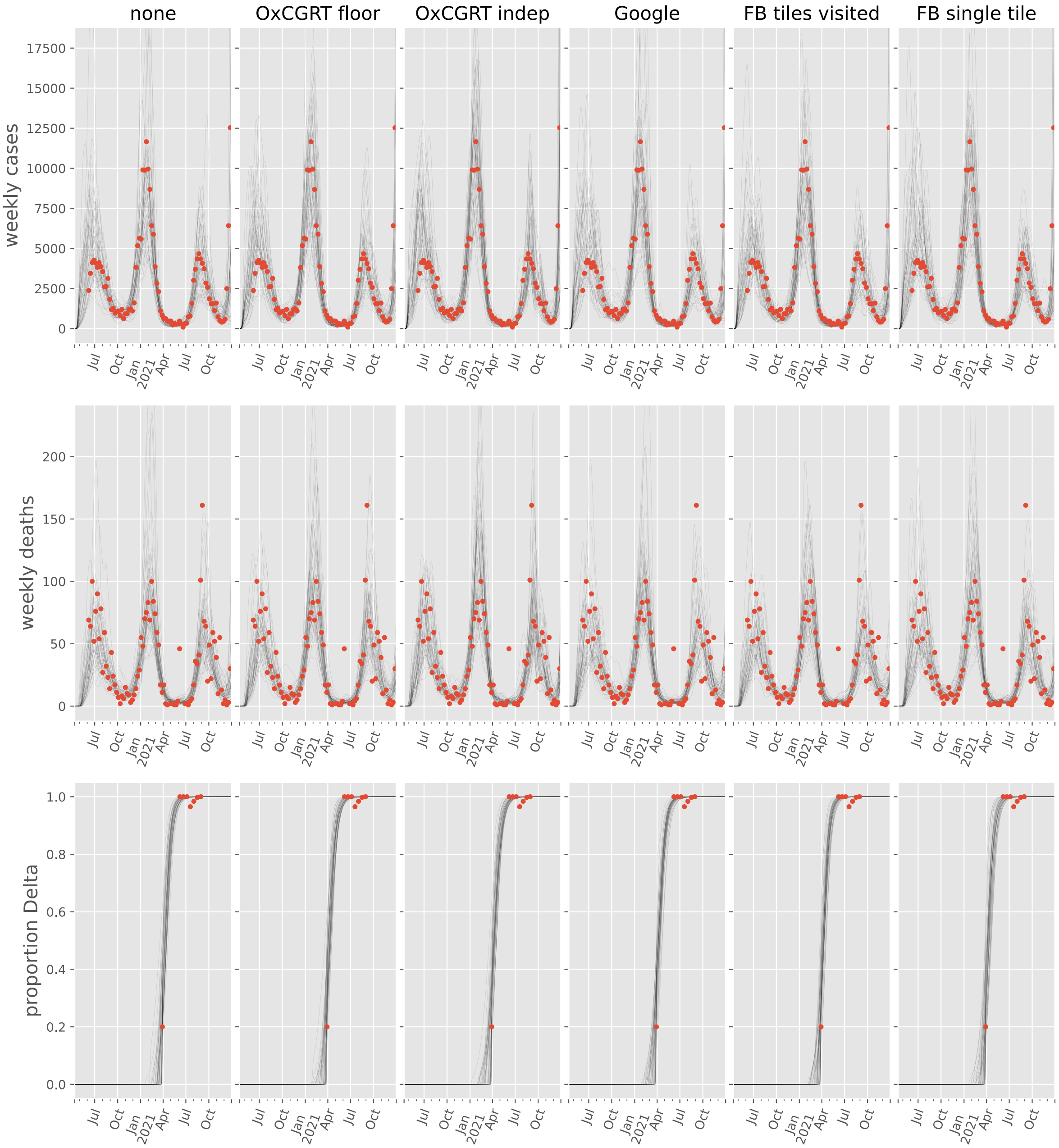

none
OxCGRT floor
OxCGRT indep
Google
FB tiles visited
FB single tile
weekly cases
17500
15000
12500
10000
7500
5000
2500
0
Jul
Oct
Jan 2021
Apr
Jul
Oct
weekly deaths
200
150
100
50
0
proportion Delta
1.0
0.8
0.6
0.4
0.2
0.0

#### Rwanda

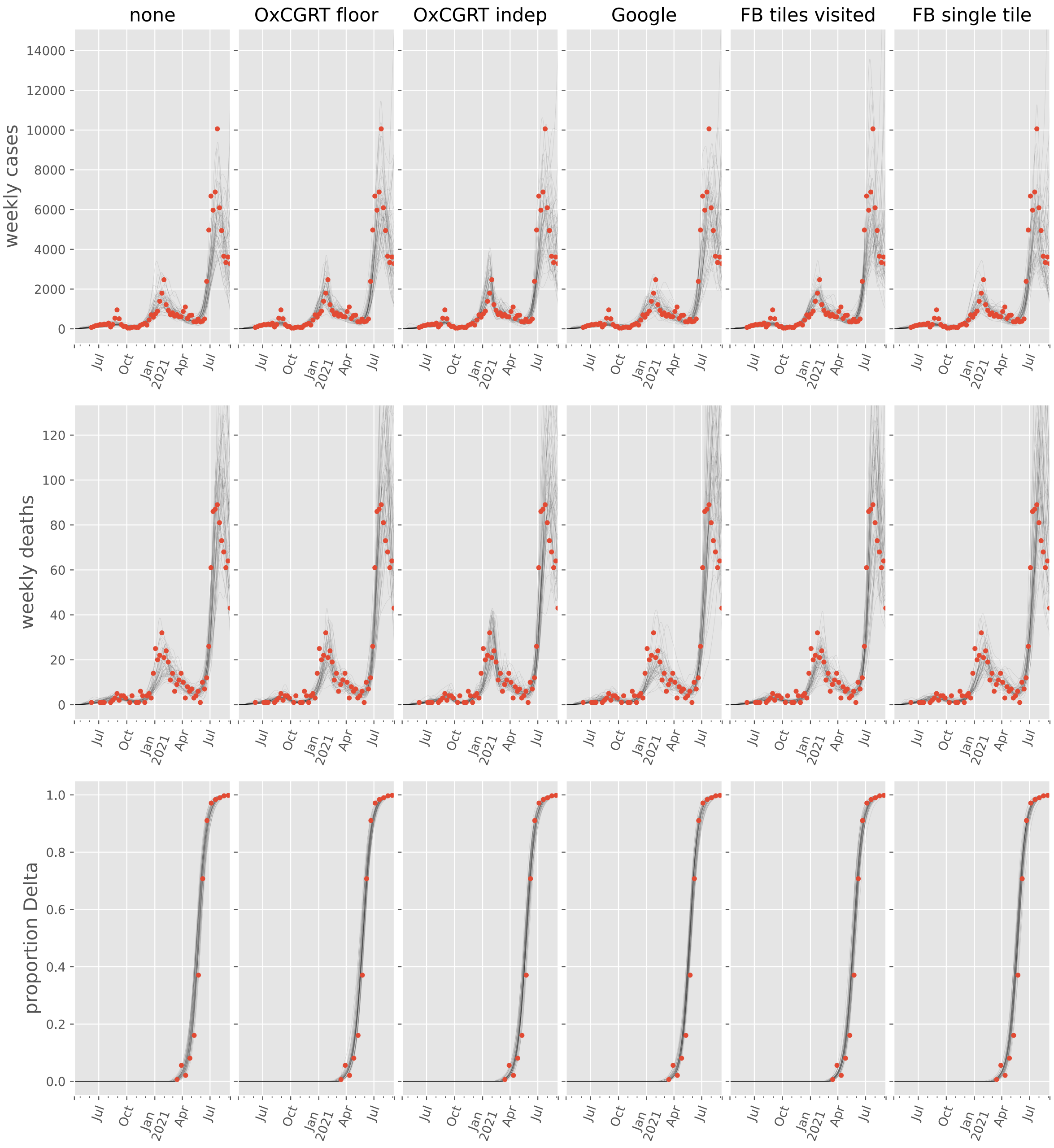

none
OxCGRT floor
OxCGRT indep
Google
FB tiles visited
FB single tile
weekly cases
weekly deaths
proportion Delta
Jul
Oct
Jan
2021
Apr

#### Senegal

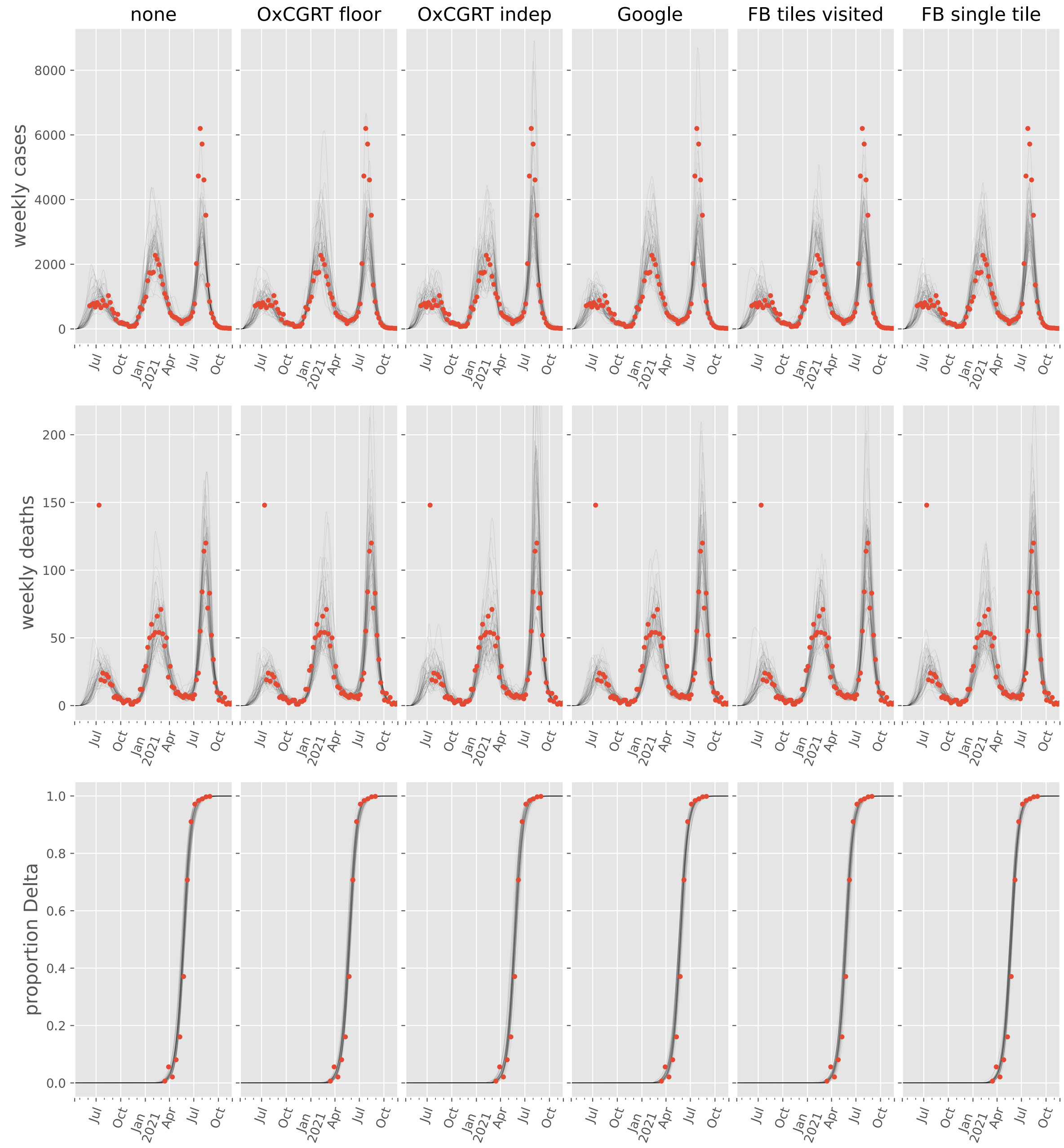
none
OxCGRT floor
OxCGRT indep
Google
FB tiles visited
FB single tile
weekly cases
weekly deaths
proportion Delta
0
2000
4000
6000
8000
0
50
100
150
200
0.0
0.2
0.4
0.6
0.8
1.0
Jul
Oct
Jan
2021
Apr
Jul
Oct

#### Somalia

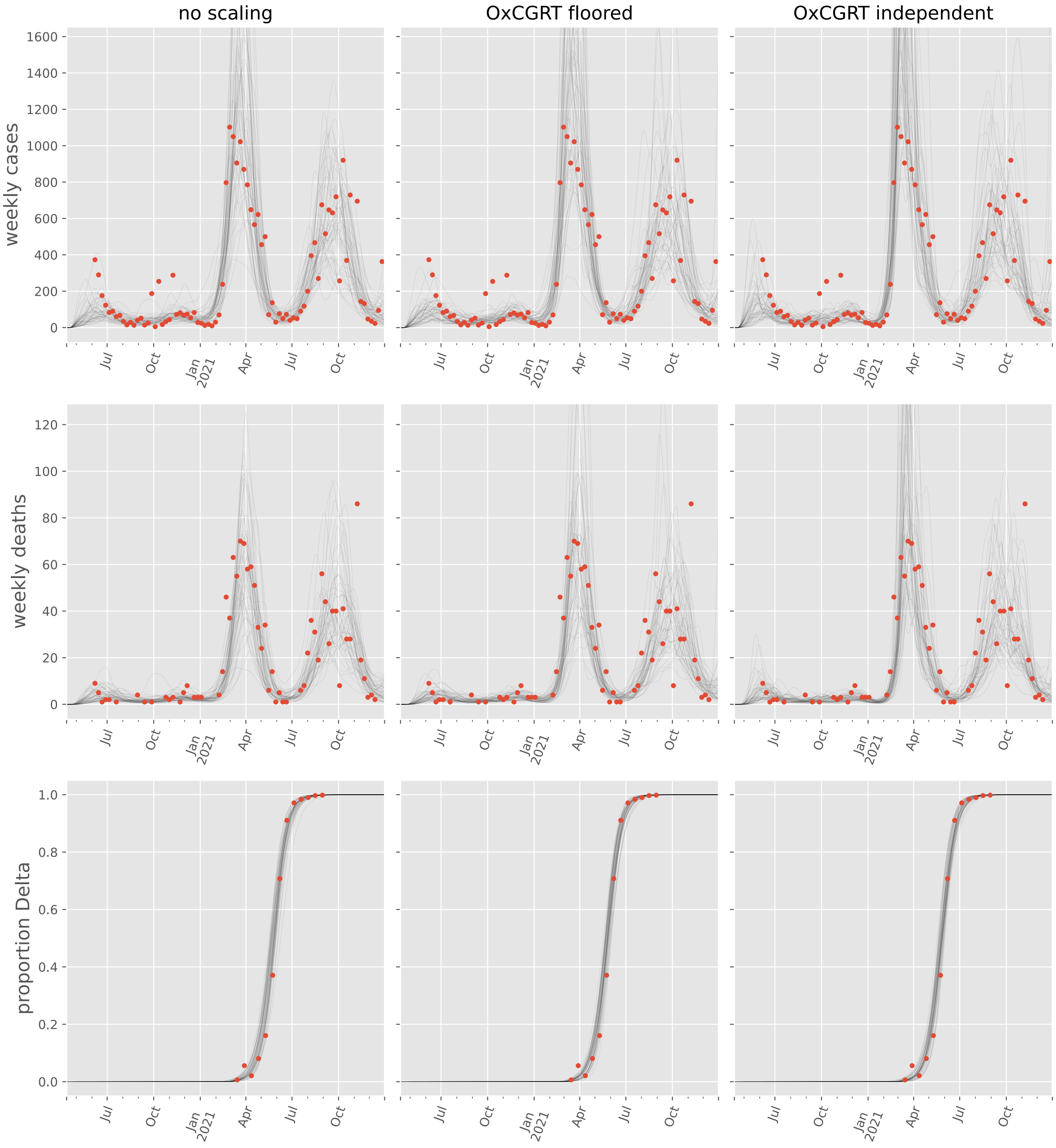
no scaling
OxCGRT floored
OxCGRT independent
weekly cases
weekly deaths
proportion Delta
1600
1400
1200
1000
800
600
400
200
0
120
100
80
60
40
20
0
1.0
0.8
0.6
0.4
0.2
0.0
Jul
Oct
Jan
2021
Apr
Jul
Oct

#### South Africa

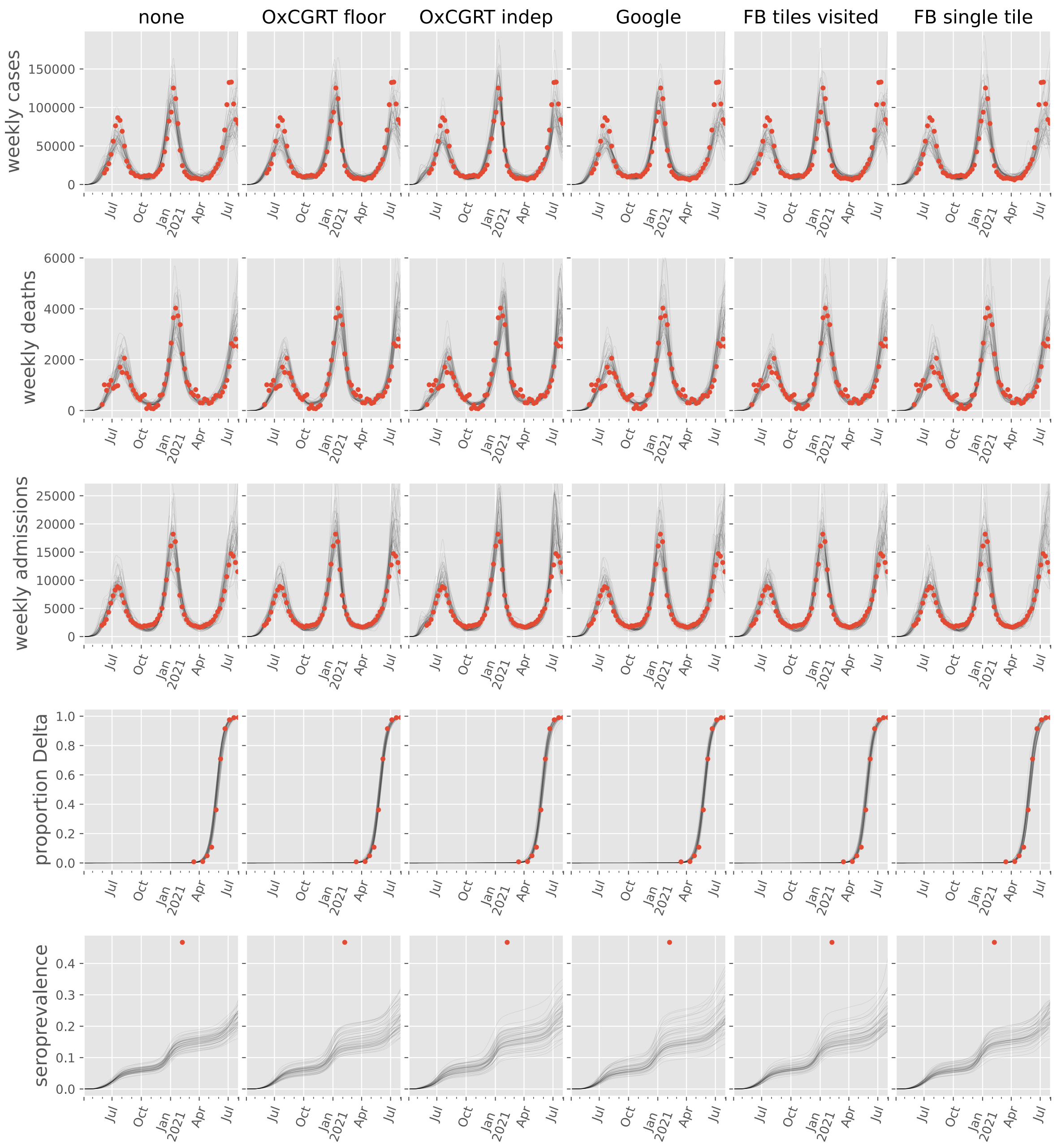

#### South Sudan

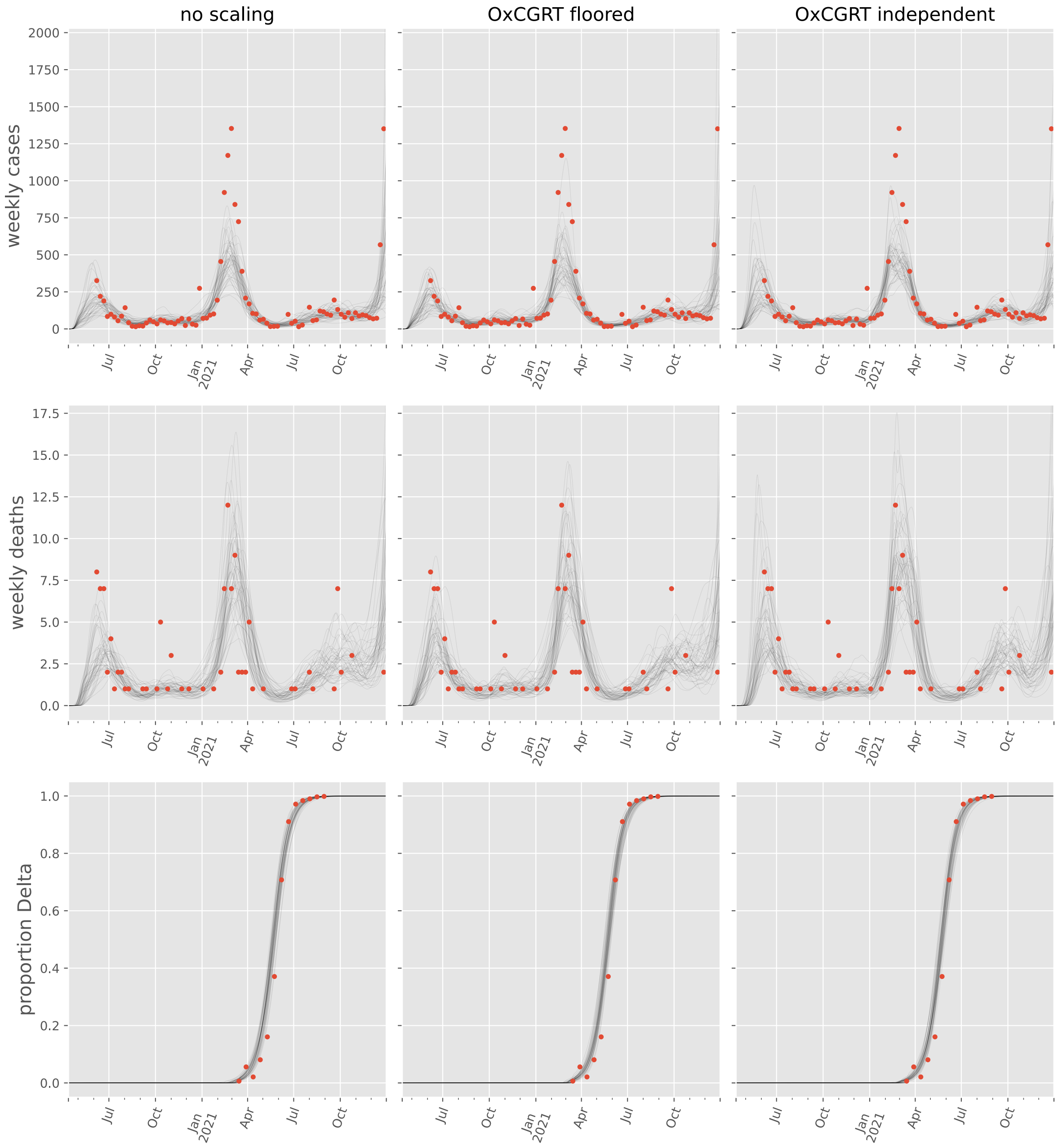

no scaling
OxCGRT floored
OxCGRT independent
weekly cases
weekly deaths
proportion Delta
2000
1750
1500
1250
1000
750
500
250
0
17.5
15.0
12.5
10.0
7.5
5.0
2.5
0.0
1.0
0.8
0.6
0.4
0.2
0.0
Jul
Oct
Jan 2021
Apr
Jul
Oct

#### Sudan

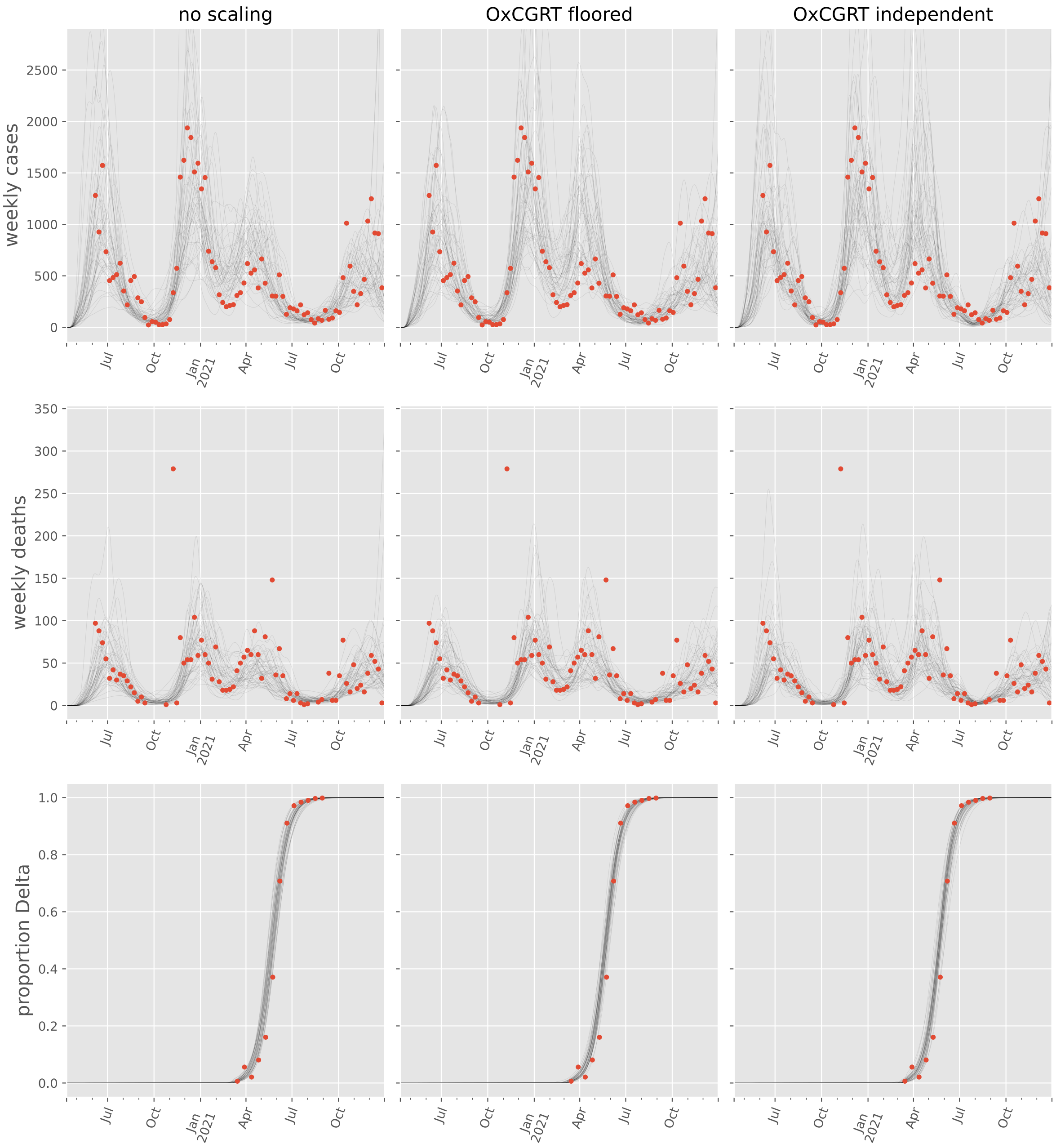

no scaling
OxCGRT floored
OxCGRT independent
weekly cases
weekly deaths
proportion Delta
0
500
1000
1500
2000
2500
50
100
150
200
250
300
350
0.0
0.2
0.4
0.6
0.8
1.0
Jul
Oct
Jan
2021
Apr

#### Togo

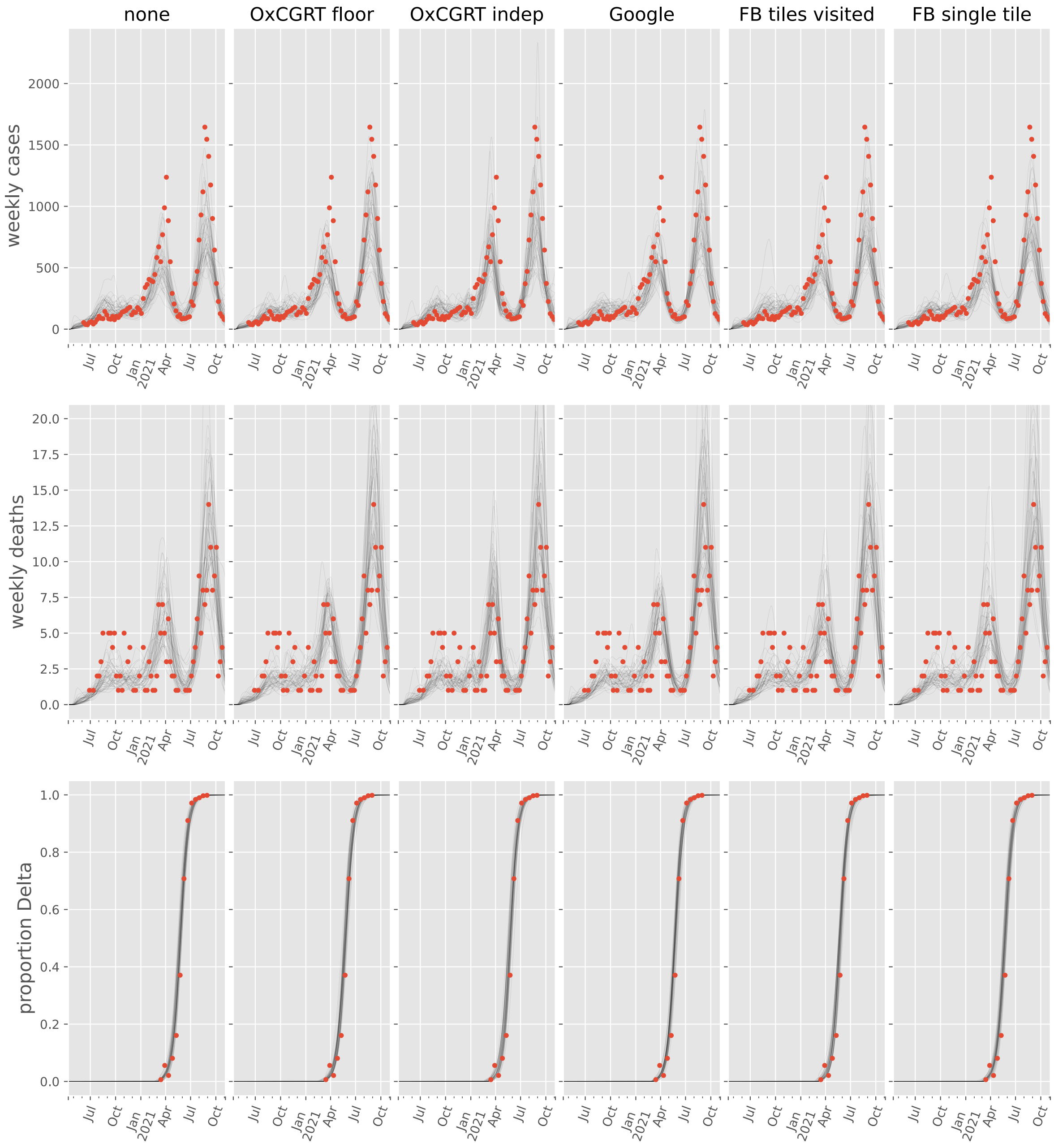
none
OxCGRT floor
OxCGRT indep
Google
FB tiles visited
FB single tile
weekly cases
weekly deaths
proportion Delta
Jul
Oct
Jan
2021
Apr
Jul
Oct

#### Tunisia

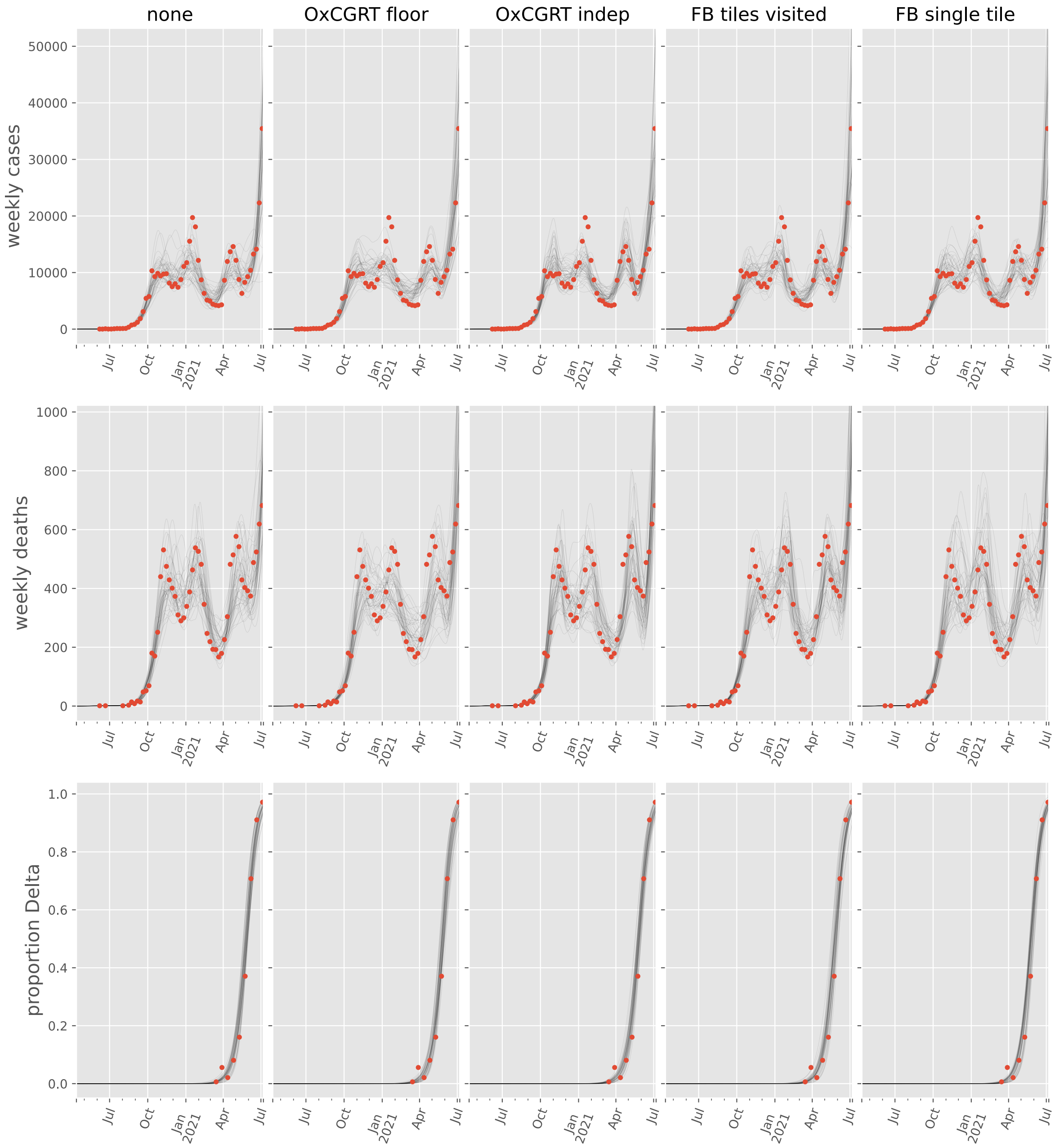

none
OxCGRT floor
OxCGRT indep
FB tiles visited
FB single tile
weekly cases
weekly deaths
proportion Delta
50000
40000
30000
20000
10000
0
1000
800
600
400
200
0
1.0
0.8
0.6
0.4
0.2
0.0
Jul
Oct
Jan 2021
Apr
Jul

#### Zambia

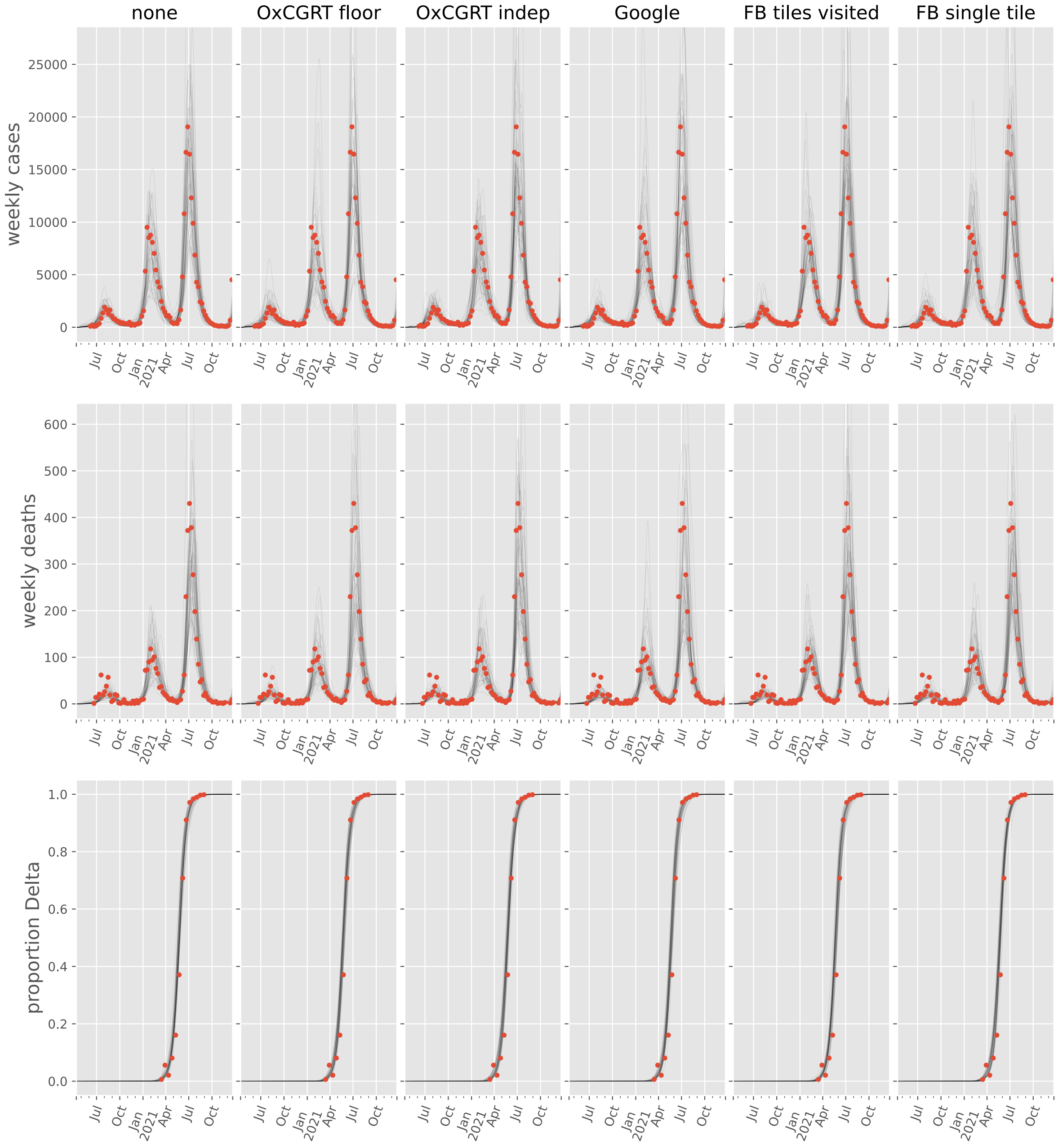

none
OxCGRT floor
OxCGRT indep
Google
FB tiles visited
FB single tile
weekly cases
weekly deaths
proportion Delta
25000
20000
15000
10000
5000
0
600
500
400
300
200
100
1.0
0.8
0.6
0.4
0.2
0.0
Jul
Oct
Jan
2021
Apr

#### Zimbabwe

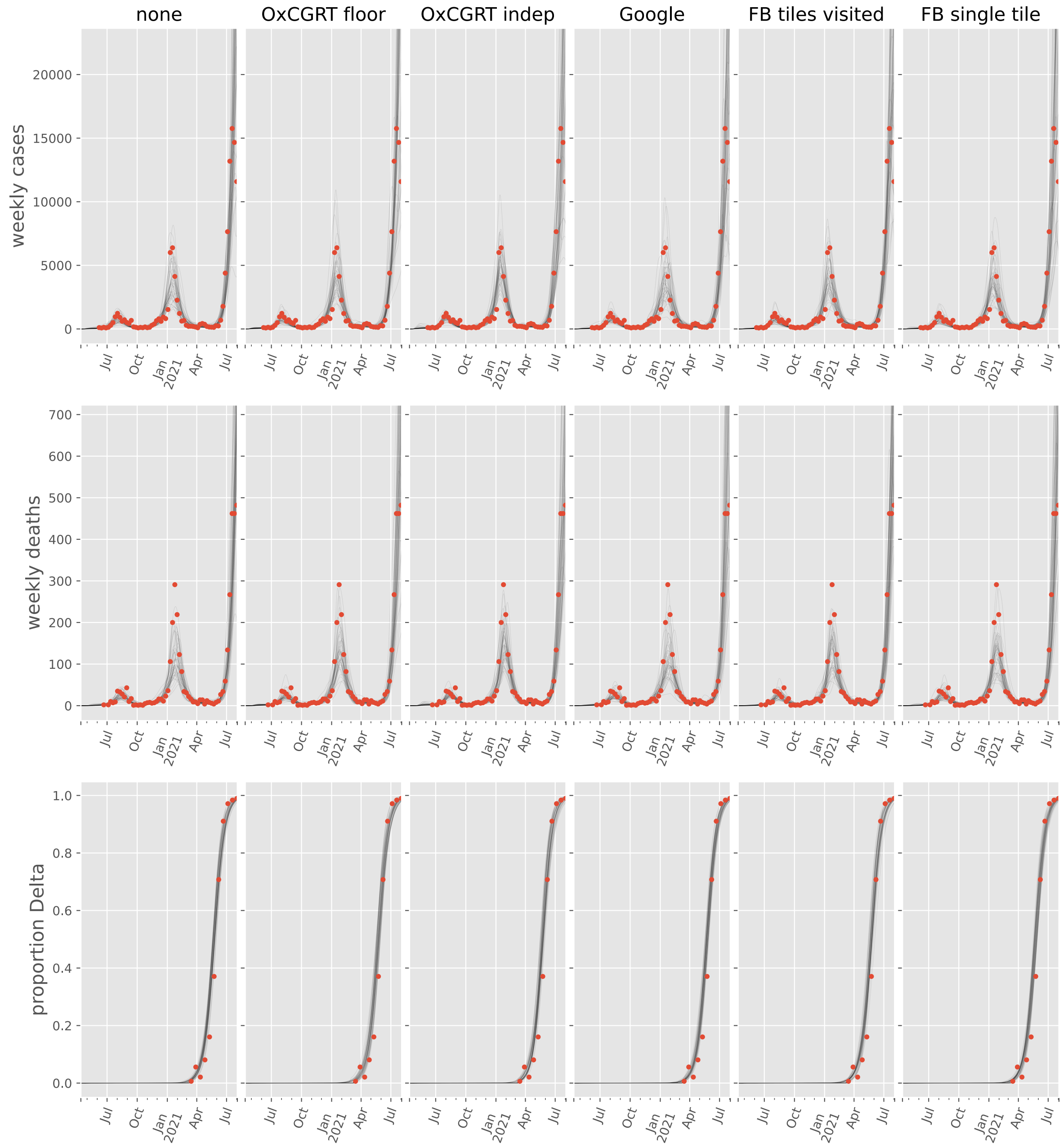

none
OxCGRT floor
OxCGRT indep
Google
FB tiles visited
FB single tile
weekly cases
weekly deaths
proportion Delta
20000
15000
10000
5000
0
700
600
500
400
300
200
100
1.0
0.8
0.6
0.4
0.2
0.0
Jul
Oct
Jan
2021
Apr

### Asia

#### Afghanistan

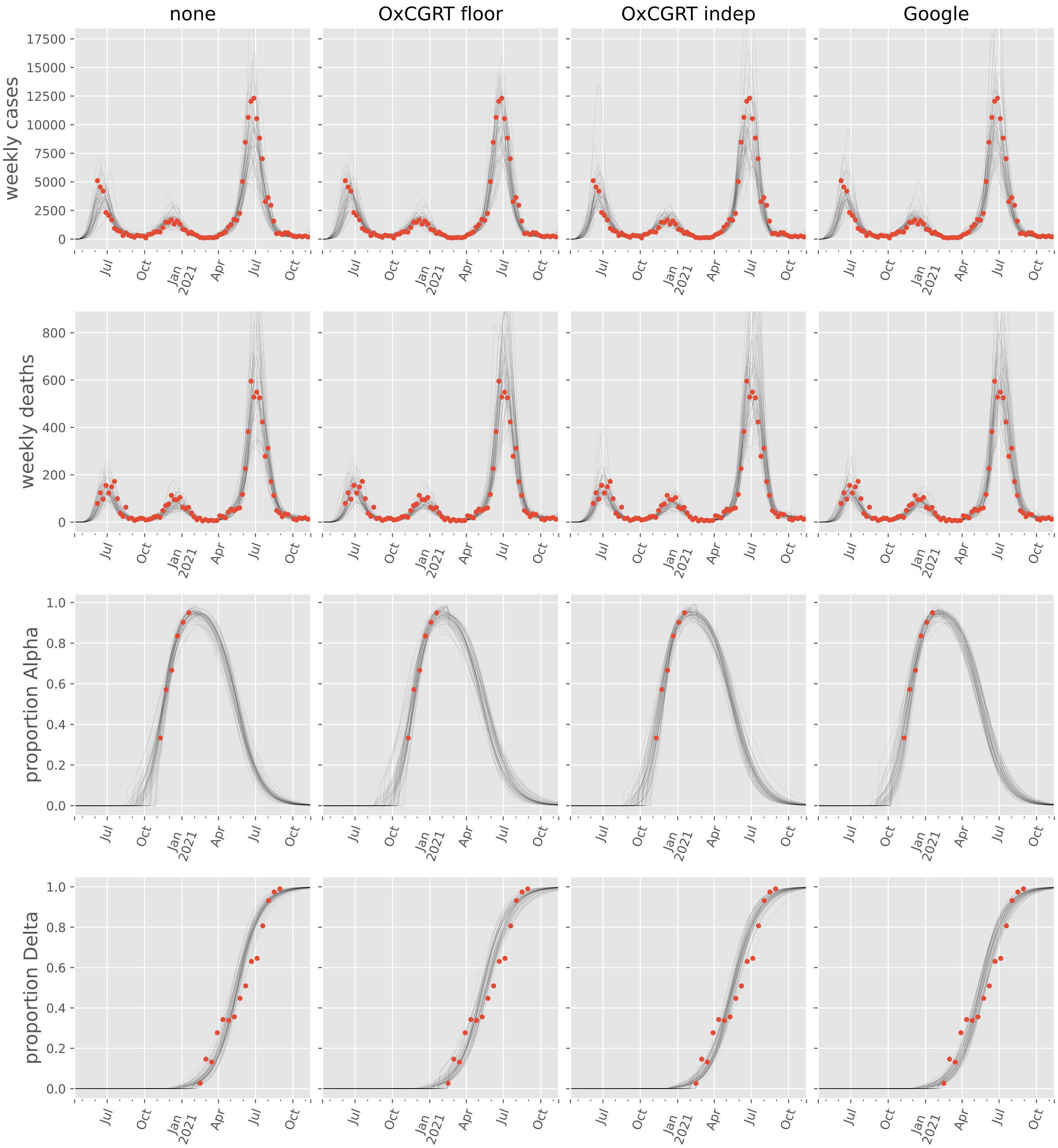

#### Azerbaijan

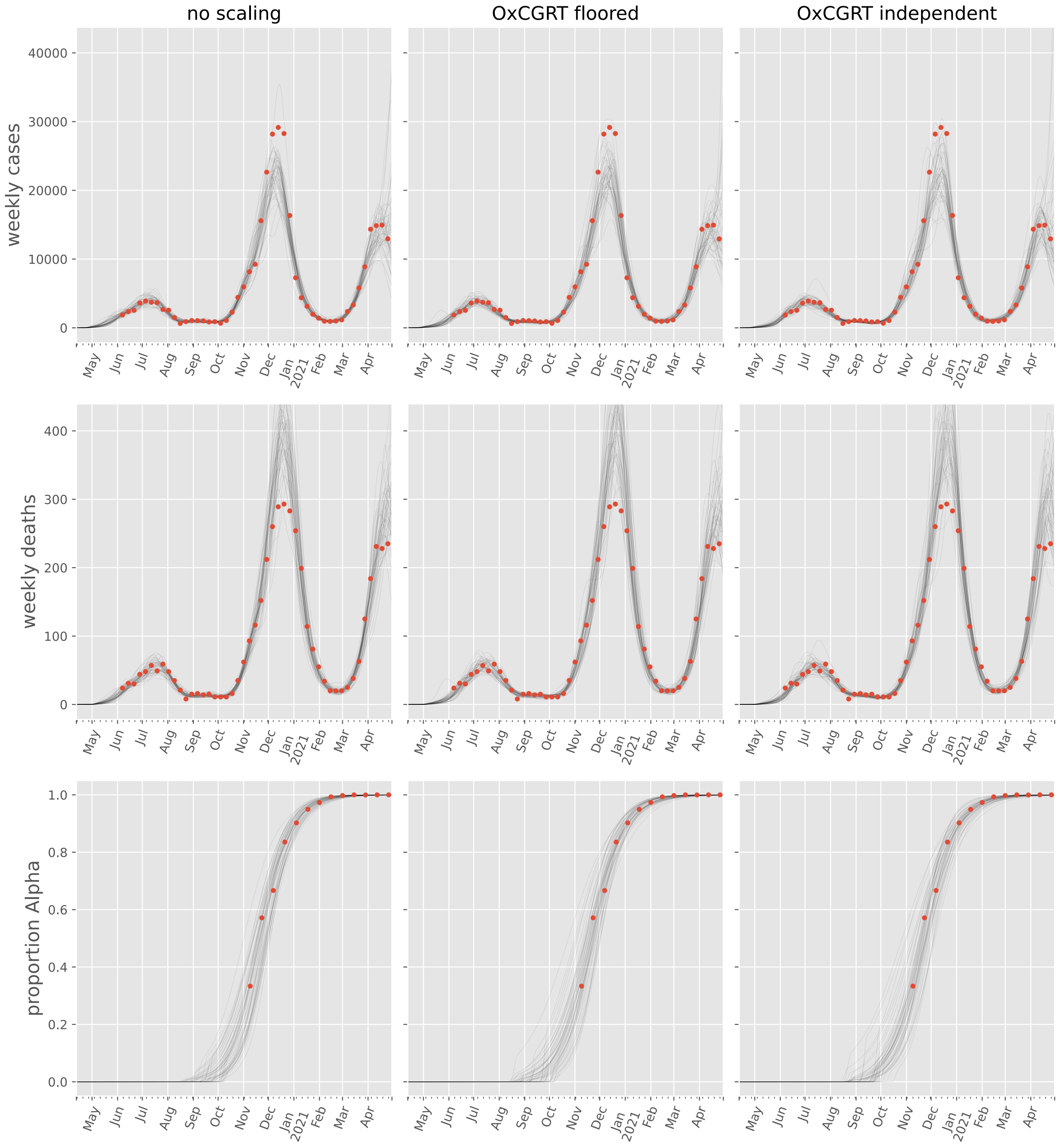

no scaling
OxCGRT floored
OxCGRT independent
weekly cases
weekly deaths
proportion Alpha
40000
30000
20000
10000
0
400
300
200
100
1.0
0.8
0.6
0.4
0.2
0.0
May
Jun
Jul
Aug
Sep
Oct
Nov
Dec
Jan 2021
Feb
Mar
Apr

#### Bahrain

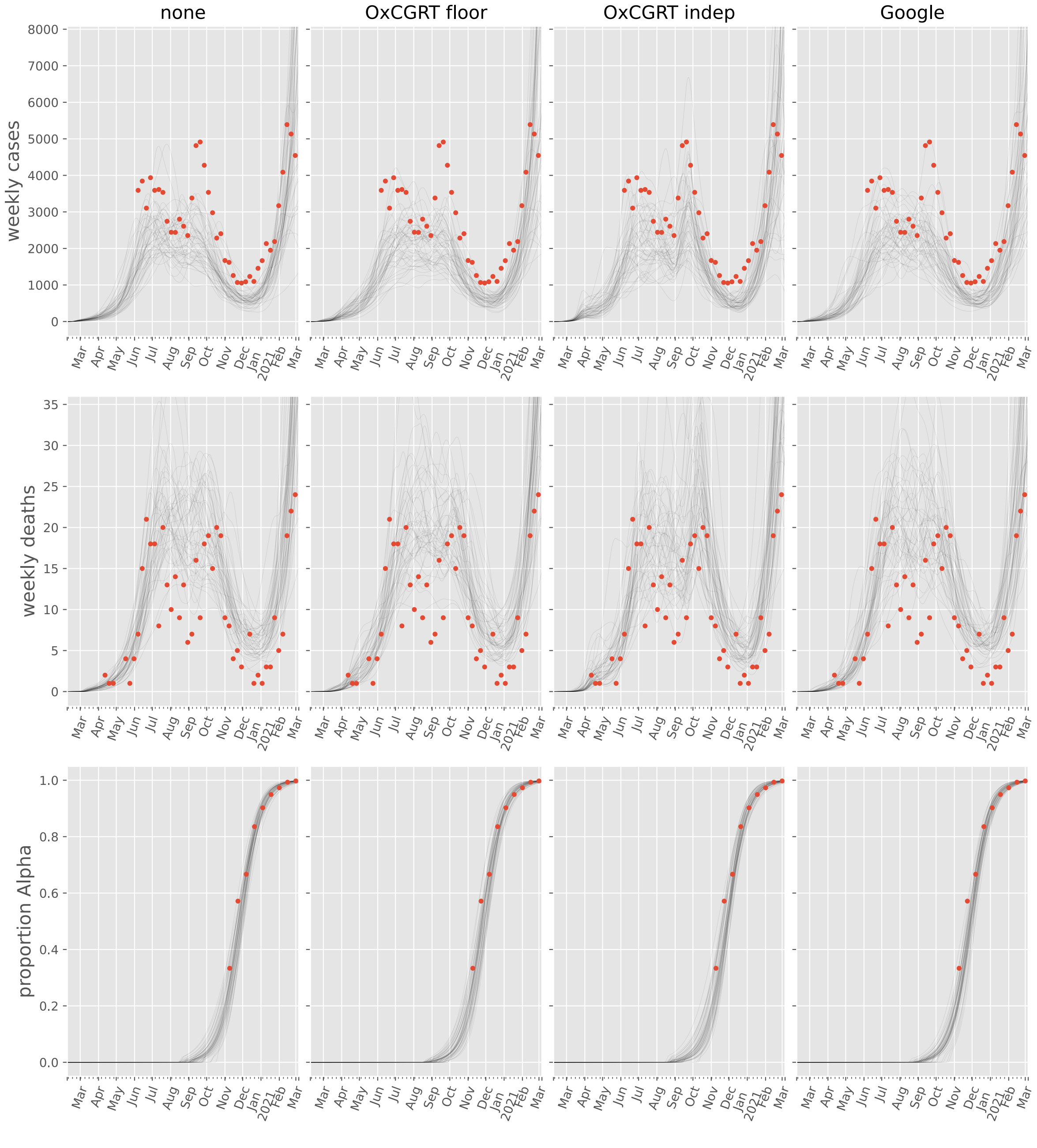

none
OxCGRT floor
OxCGRT indep
Google
weekly cases
weekly deaths
proportion Alpha
8000
7000
6000
5000
4000
3000
2000
1000
0
35
30
25
20
15
10
5
0
1.0
0.8
0.6
0.4
0.2
0.0
Mar
Apr
May
Jun
Jul
Aug
Sep
Oct
Nov
Dec
Jan
2021
Feb
Mar

#### Bangladesh

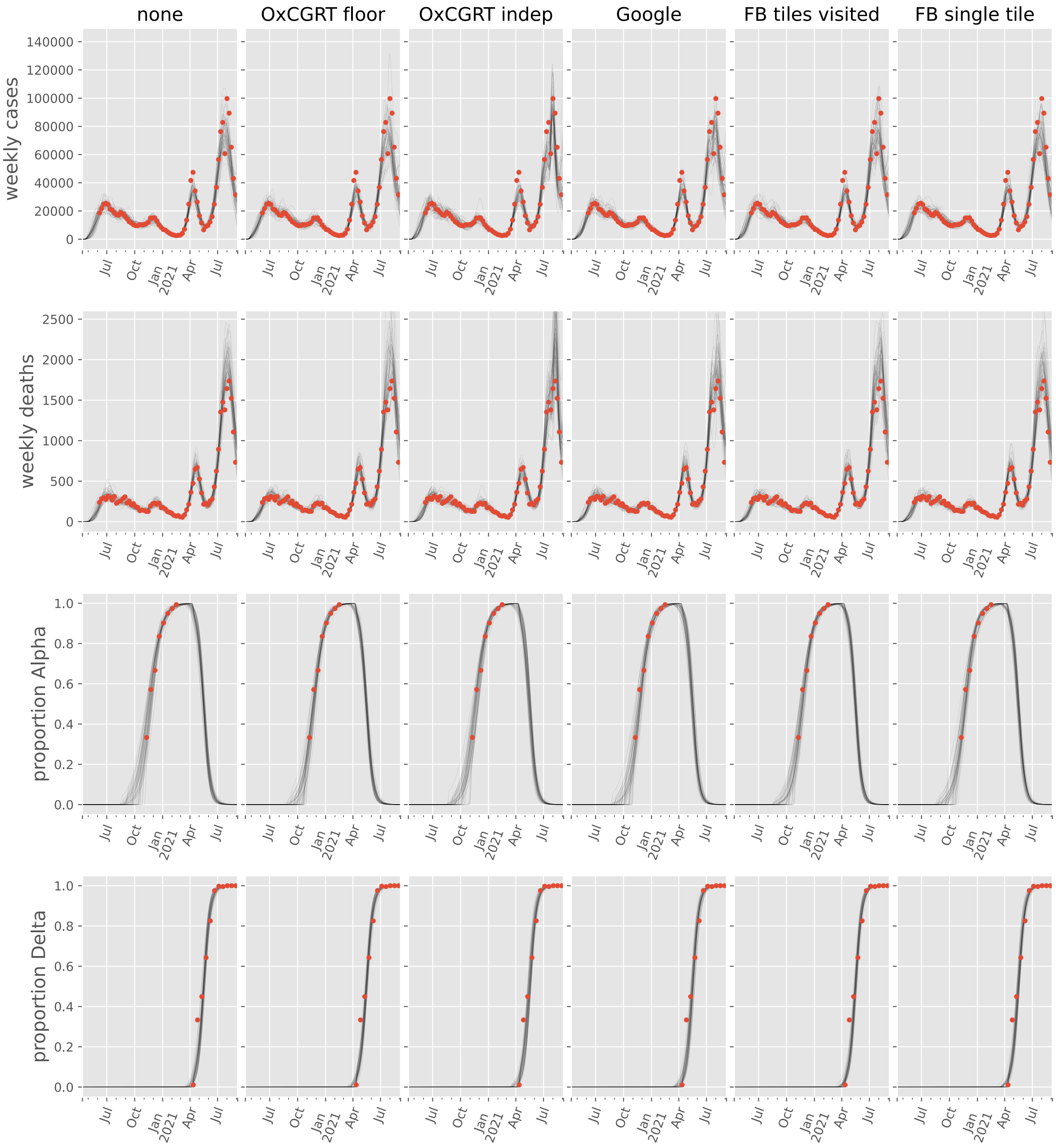

none
OxCGRT floor
OxCGRT indep
Google
FB tiles visited
FB single tile
weekly cases
weekly deaths
proportion Alpha
proportion Delta
Jul
Oct
Jan
2021
Apr
Jul

#### Cyprus

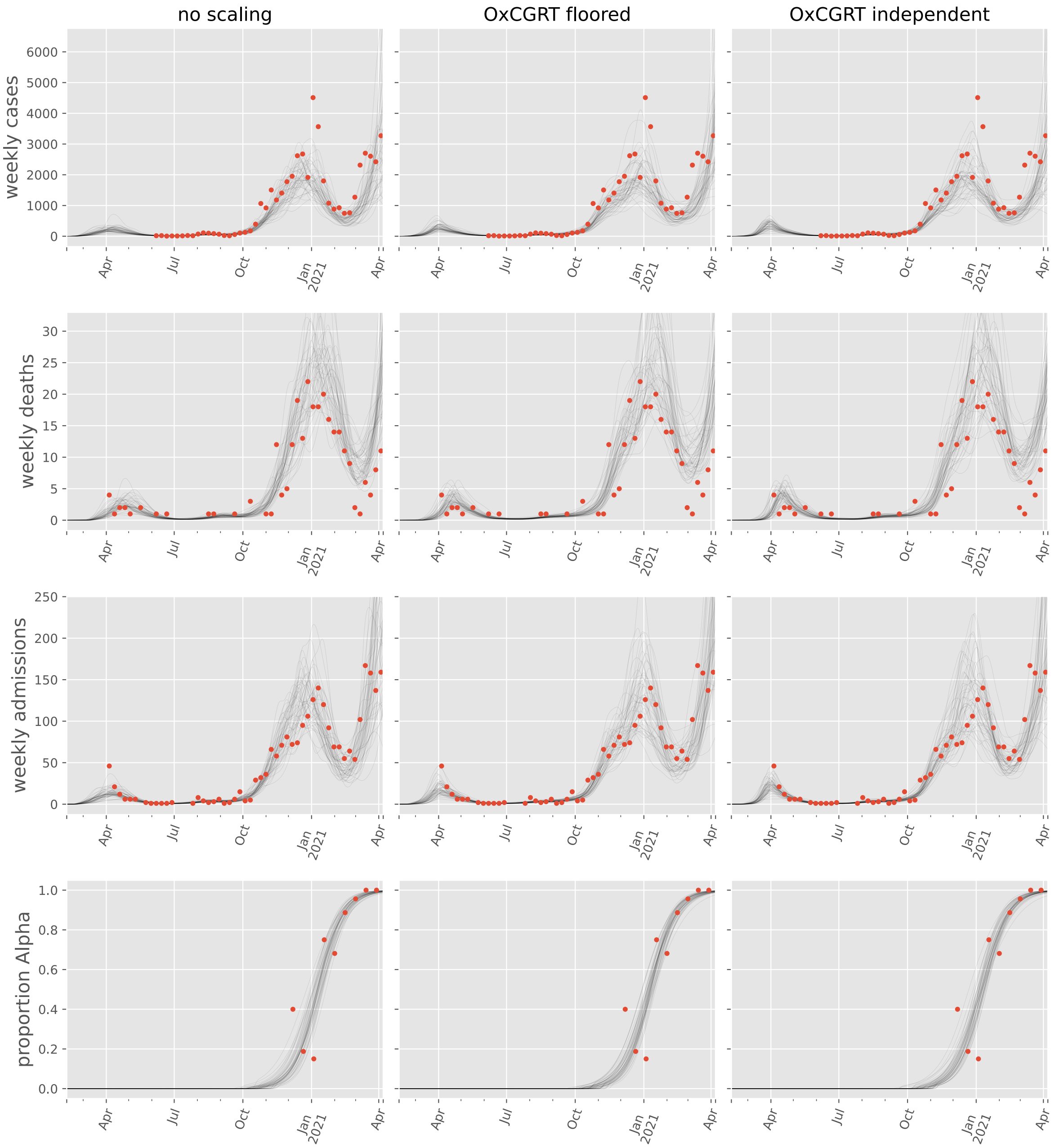

no scaling
OxCGRT floored
OxCGRT independent
weekly cases
weekly deaths
weekly admissions
proportion Alpha
6000
5000
4000
3000
2000
1000
0
30
25
20
15
10
5
250
200
150
100
50
1.0
0.8
0.6
0.4
0.2
0.0
Apr
Jul
Oct
Jan
2021

#### Georgia

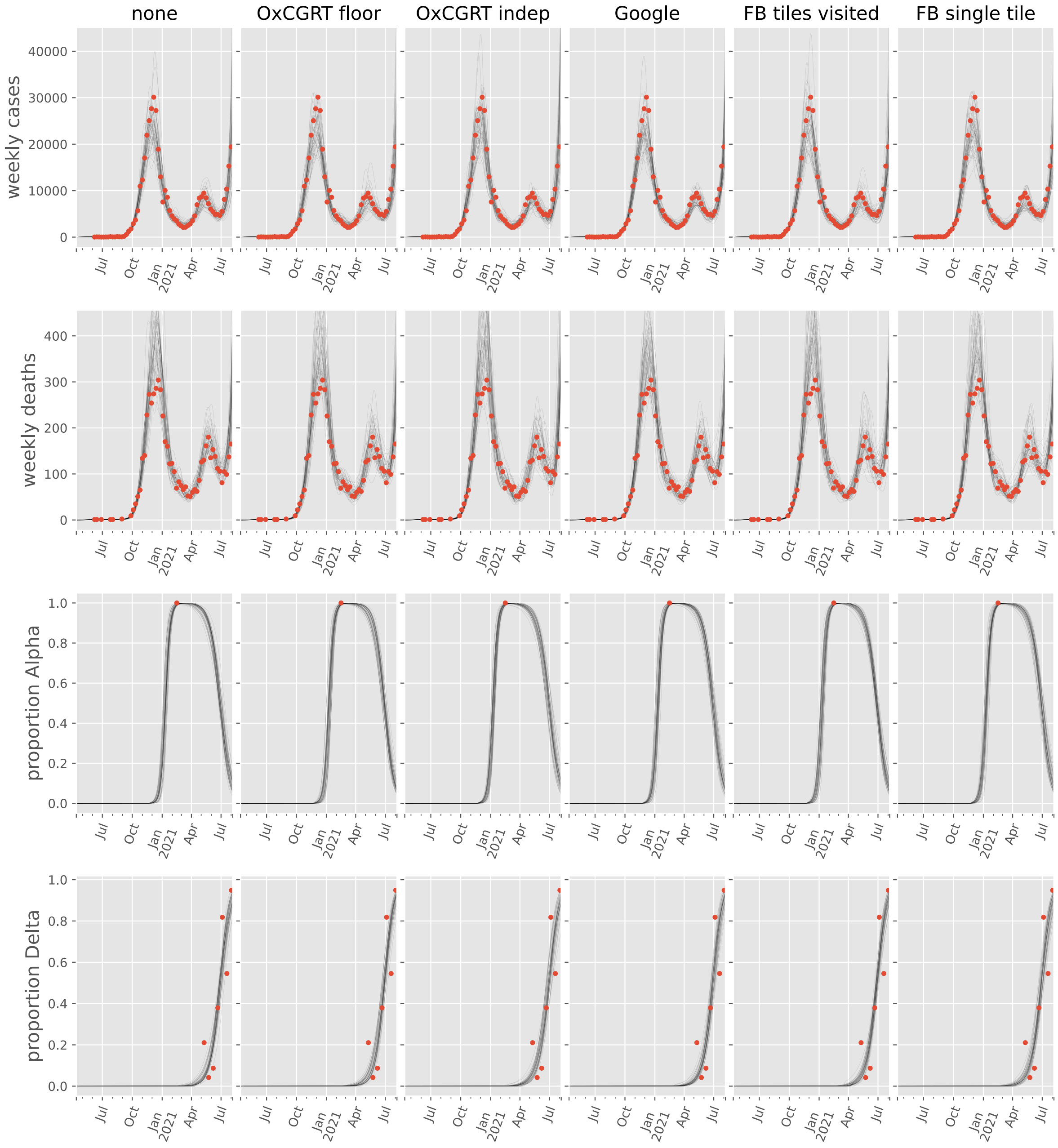

none
OxCGRT floor
OxCGRT indep
Google
FB tiles visited
FB single tile
weekly cases
weekly deaths
proportion Alpha
proportion Delta
40000
30000
20000
10000
0
400
300
200
100
1.0
0.8
0.6
0.4
0.2
0.0
Jul
Oct
Jan
2021
Apr

#### India

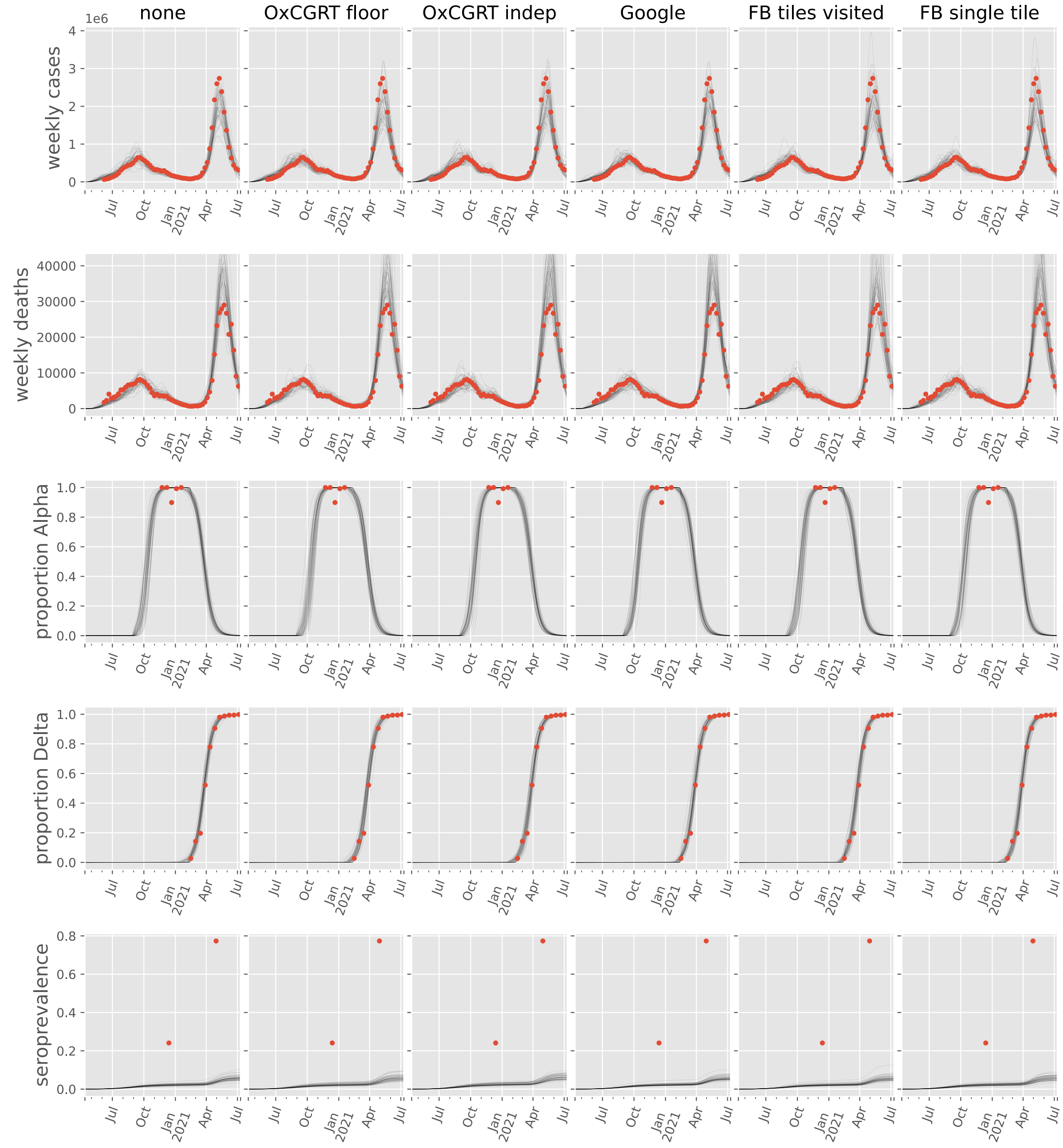

none
OxCGRT floor
OxCGRT indep
Google
FB tiles visited
FB single tile
weekly cases
weekly deaths
proportion Alpha
proportion Delta
seroprevalence
1e6
Jul
Oct
Jan 2021
Apr

#### Indonesia

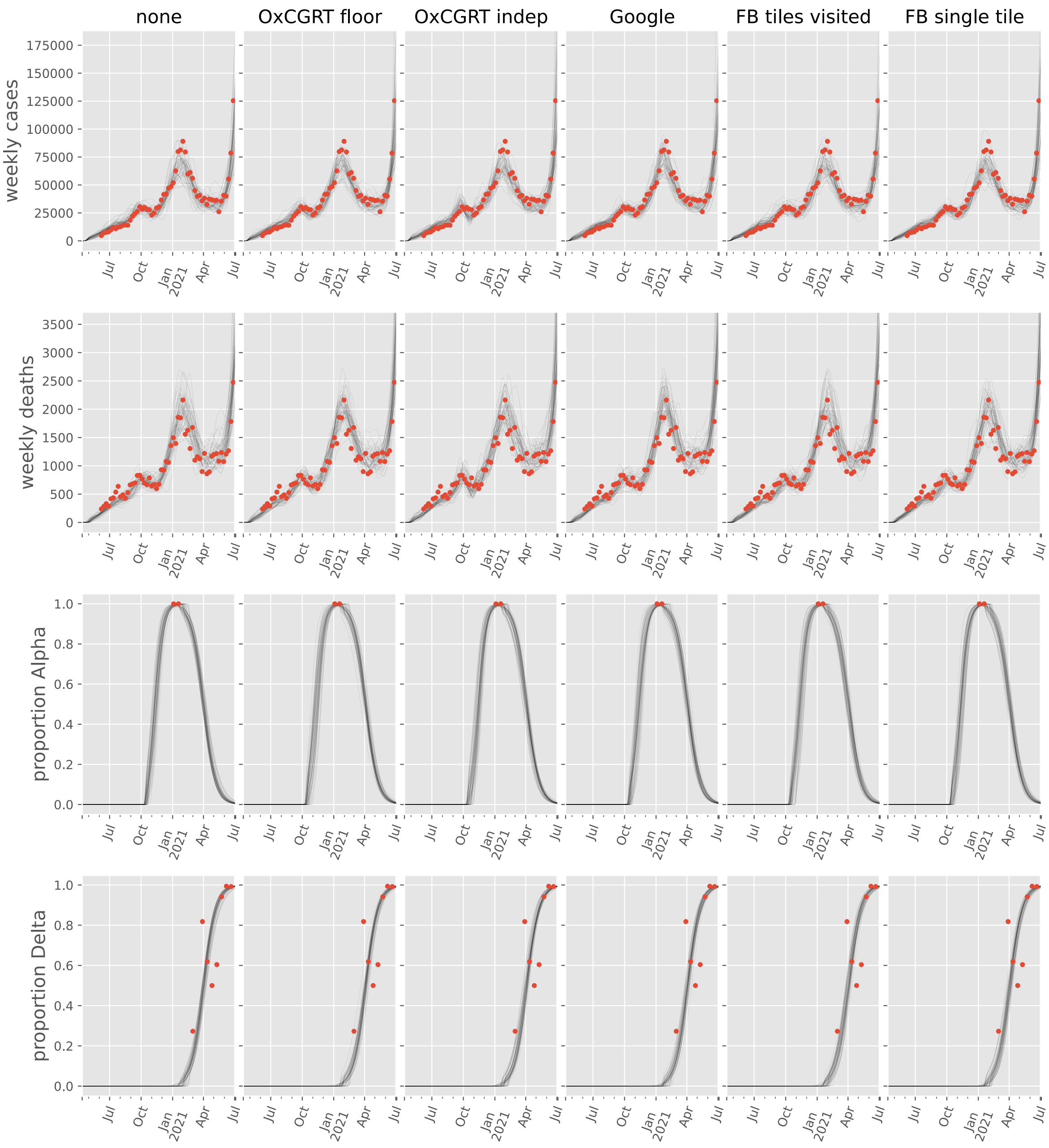

none
OxCGRT floor
OxCGRT indep
Google
FB tiles visited
FB single tile
weekly cases
weekly deaths
proportion Alpha
proportion Delta
175000
150000
125000
100000
75000
50000
25000
0
3500
3000
2500
2000
1500
1000
500
1.0
0.8
0.6
0.4
0.2
0.0
Jul
Oct
Jan 2021
Apr

#### Iran, Islamic Republic of

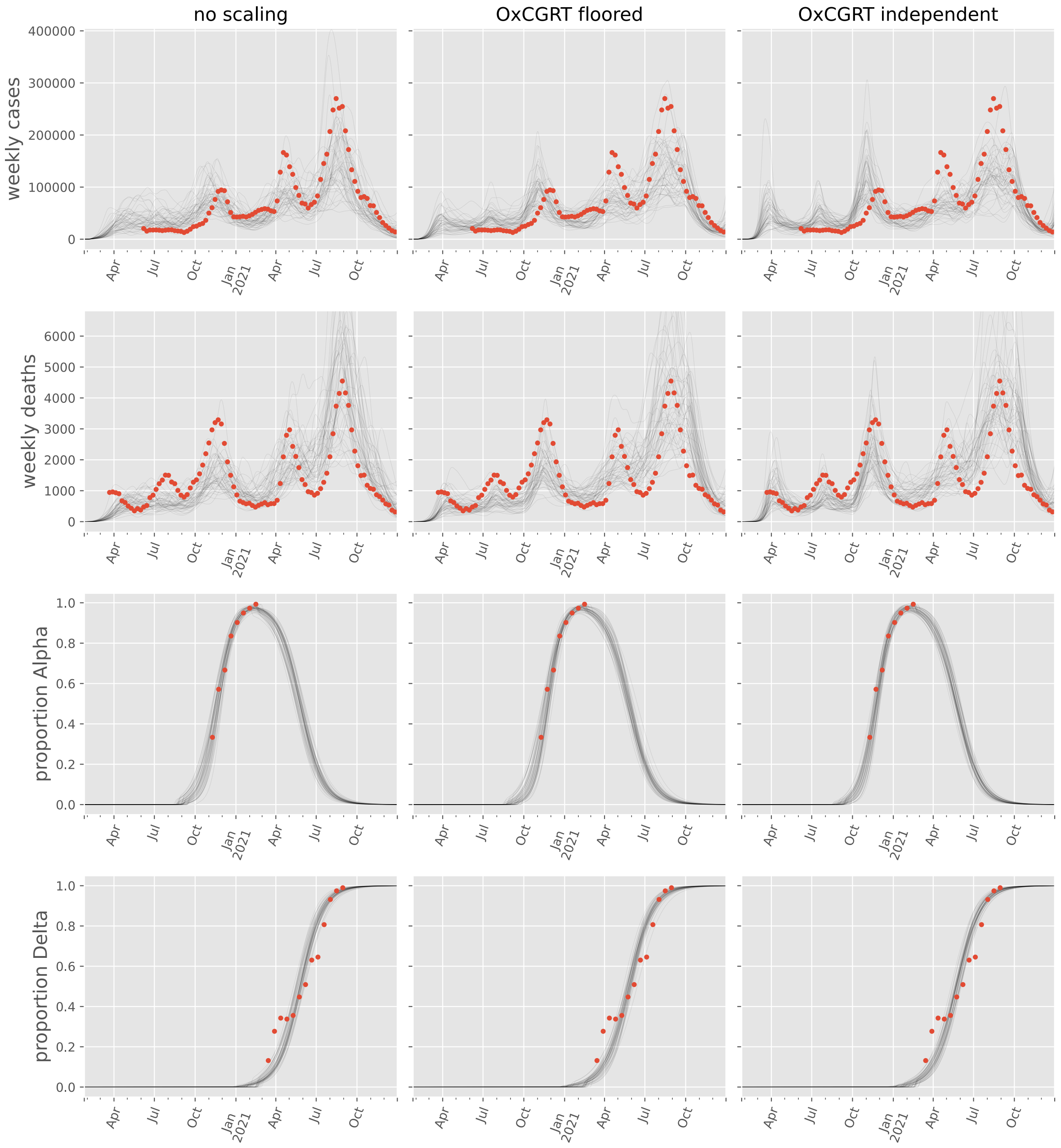

#### Iraq

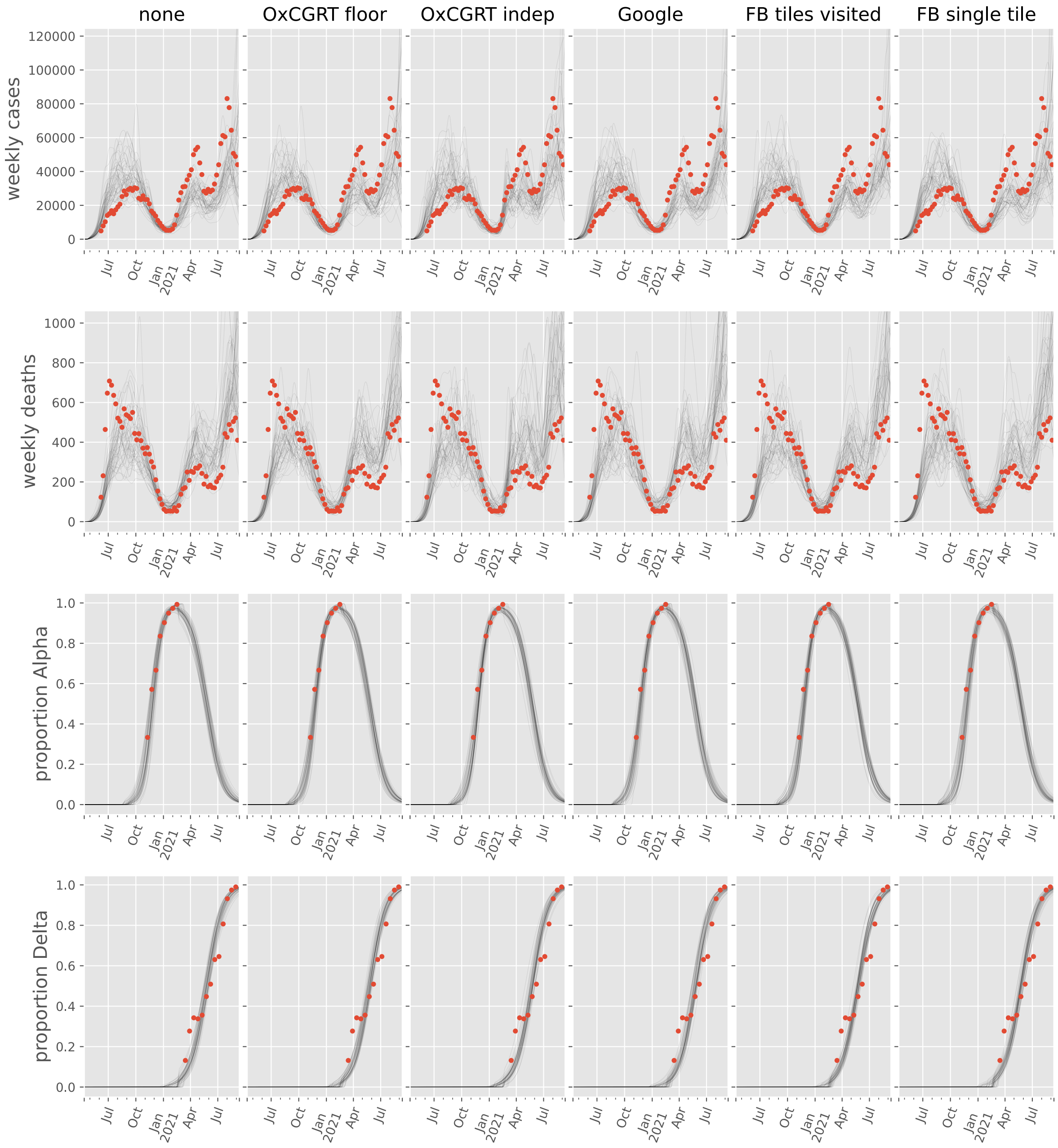
none
OxCGRT floor
OxCGRT indep
Google
FB tiles visited
FB single tile
weekly cases
weekly deaths
proportion Alpha
proportion Delta
Jul
Oct
Jan
2021
Apr
Jul

#### Israel

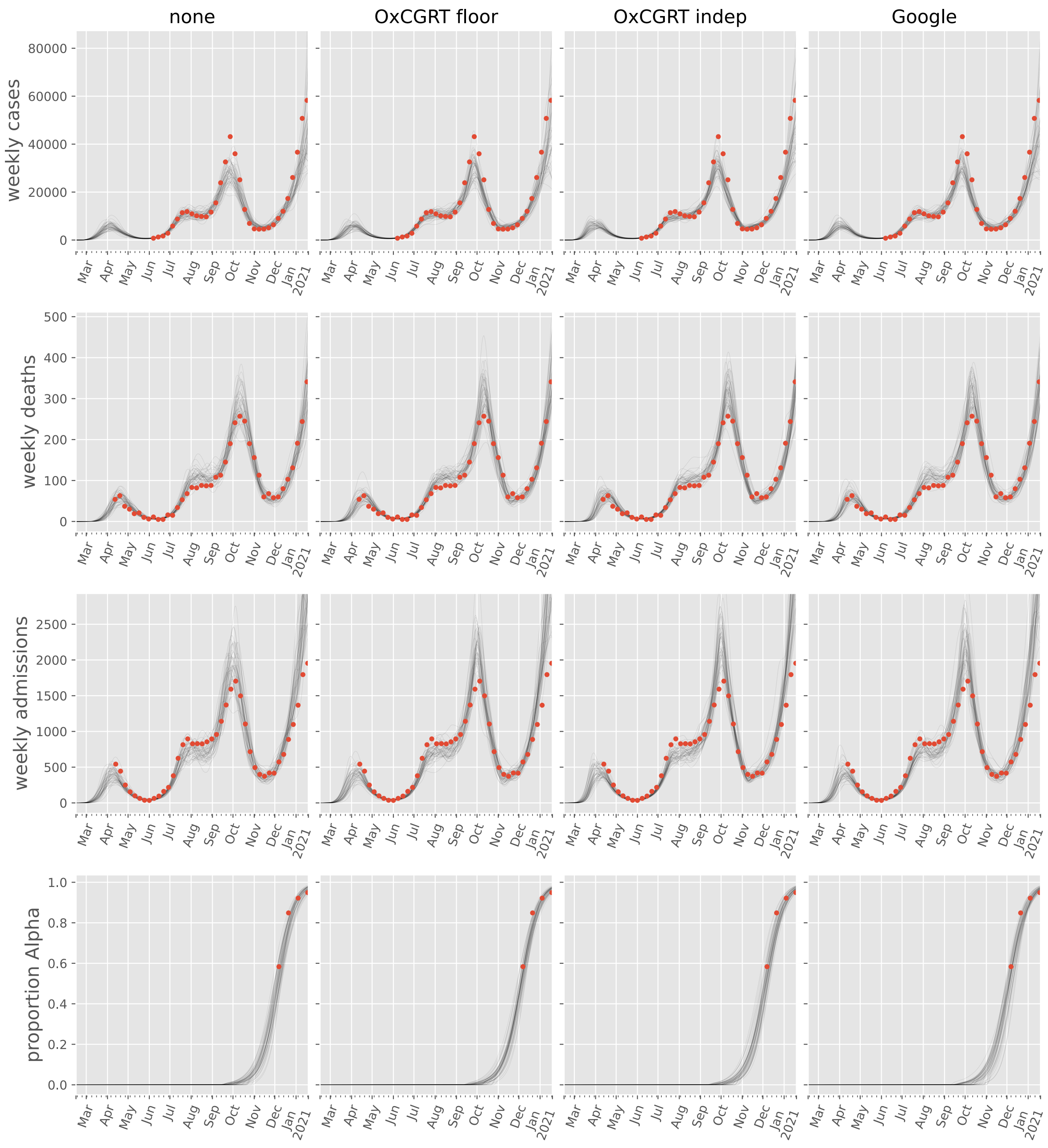
none
OxCGRT floor
OxCGRT indep
Google
weekly cases
weekly deaths
weekly admissions
proportion Alpha
Mar
Apr
May
Jun
Jul
Aug
Sep
Oct
Nov
Dec
Jan
2021

#### Japan

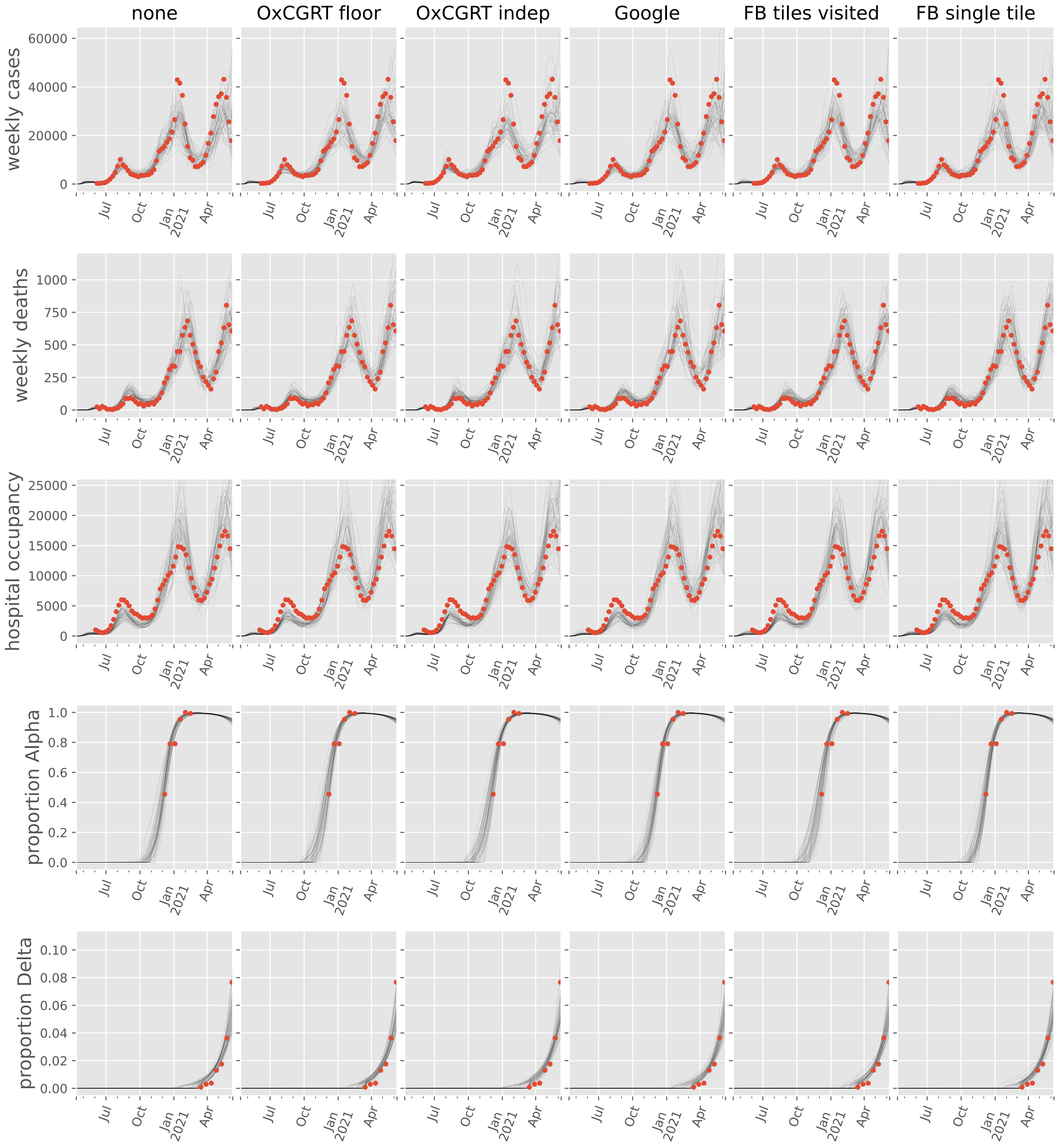
none
OxCGRT floor
OxCGRT indep
Google
FB tiles visited
FB single tile
weekly cases
weekly deaths
hospital occupancy
proportion Alpha
proportion Delta
Jul
Oct
Jan
2021
Apr

#### Jordan

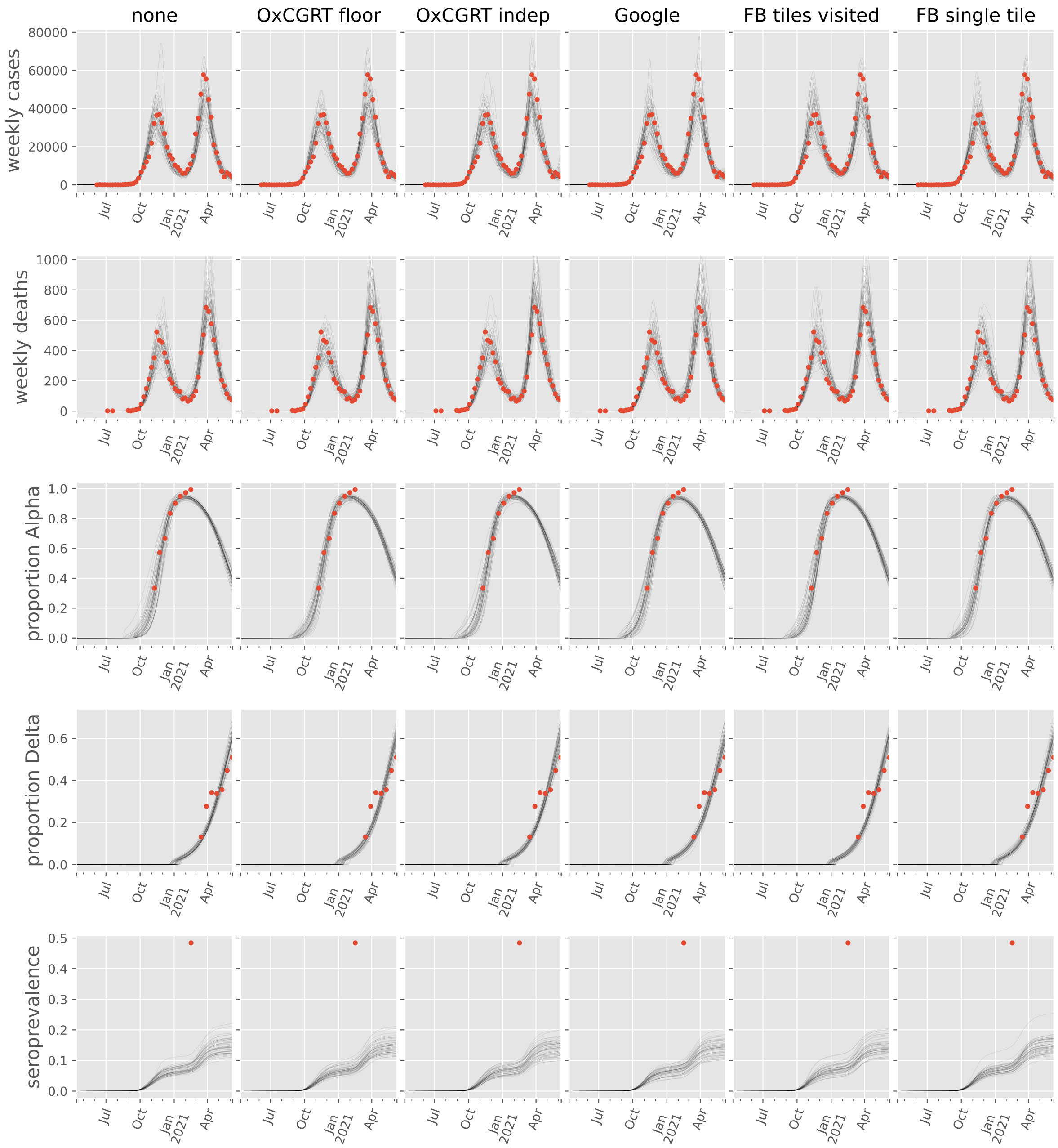

none
OxCGRT floor
OxCGRT indep
Google
FB tiles visited
FB single tile
weekly cases
weekly deaths
proportion Alpha
proportion Delta
seroprevalence

#### Korea, Republic of

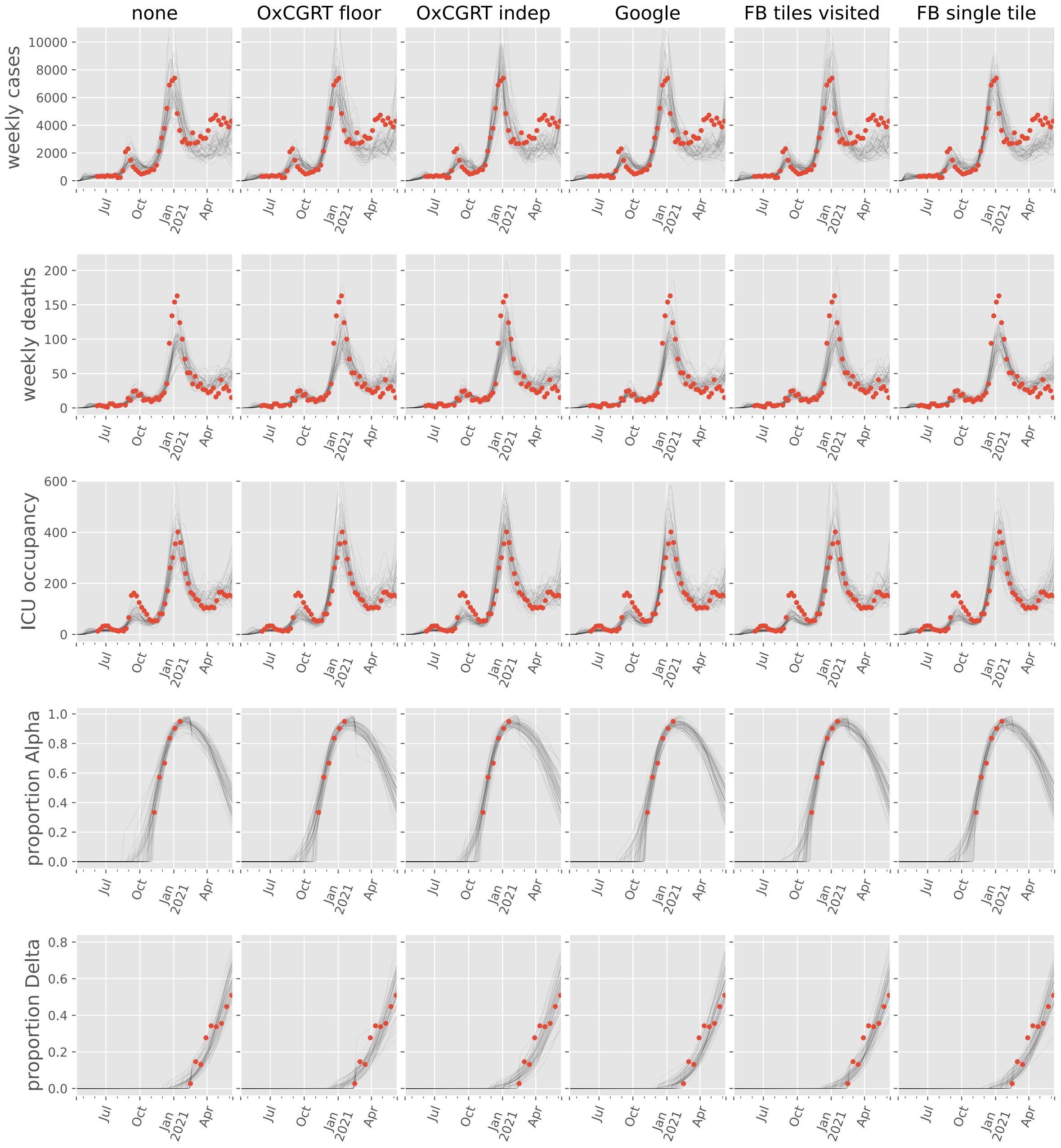

#### Kuwait

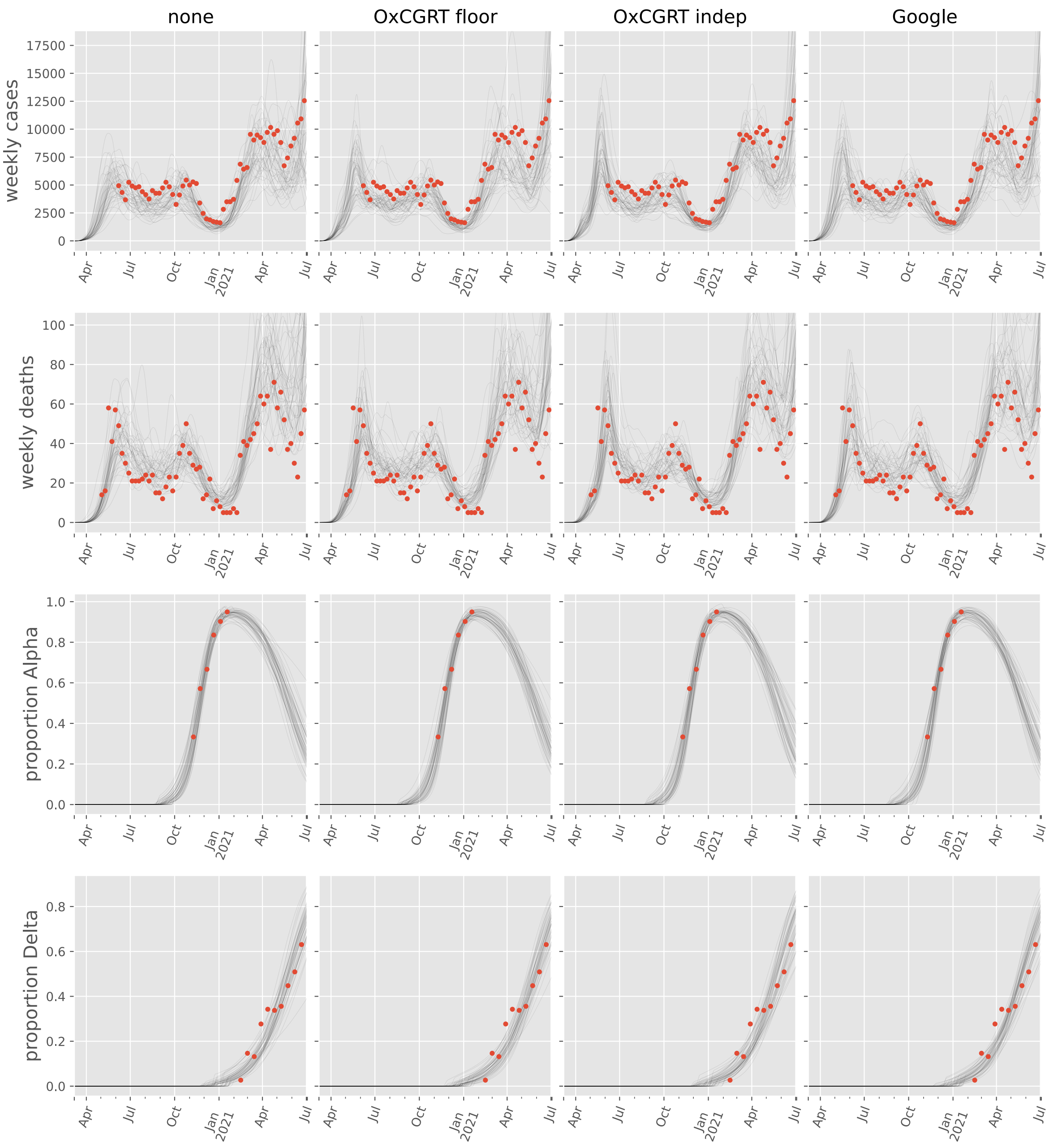

none
OxCGRT floor
OxCGRT indep
Google
weekly cases
weekly deaths
proportion Alpha
proportion Delta
17500
15000
12500
10000
7500
5000
2500
0
100
80
60
40
20
1.0
0.8
0.6
0.4
0.2
0.0
Apr
Jul
Oct
Jan
2021

#### Lebanon

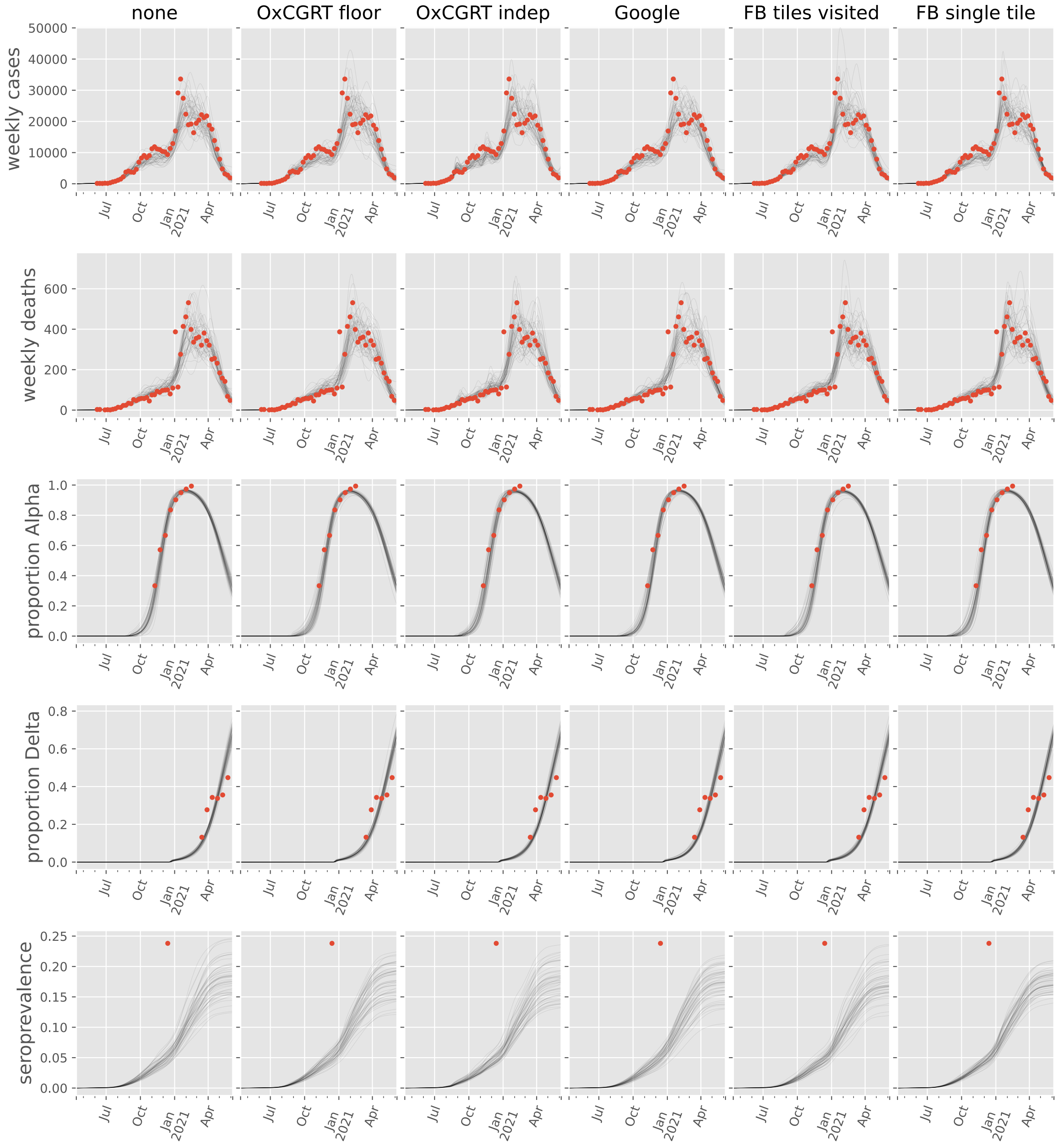
none
OxCGRT floor
OxCGRT indep
Google
FB tiles visited
FB single tile
weekly cases
weekly deaths
proportion Alpha
proportion Delta
seroprevalence
Jul
Oct
Jan
2021
Apr

#### Malaysia

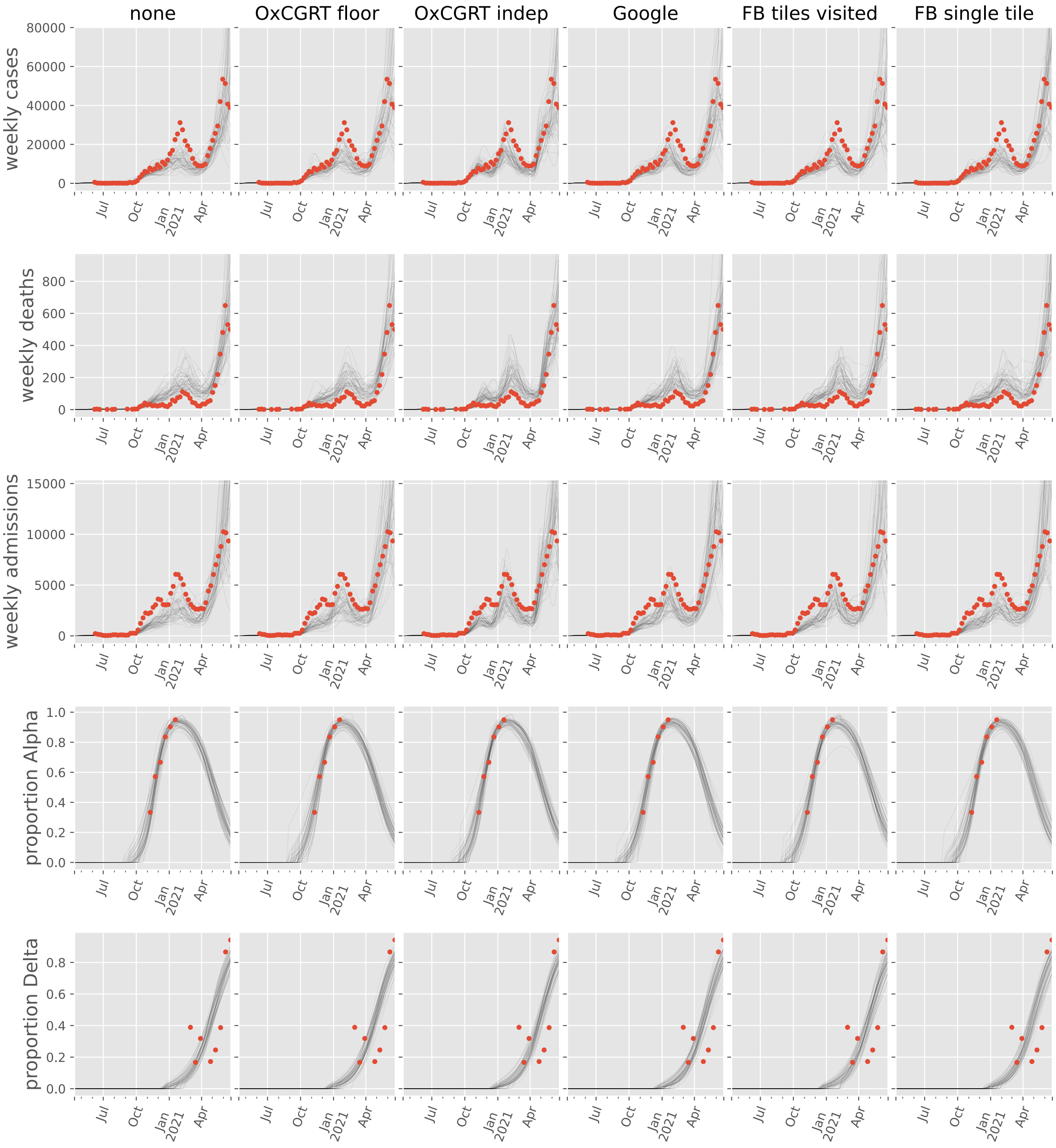
none
OxCGRT floor
OxCGRT indep
Google
FB tiles visited
FB single tile
weekly cases
weekly deaths
weekly admissions
proportion Alpha
proportion Delta
80000
60000
40000
20000
0
800
600
400
200
15000
10000
5000
1.0
0.8
0.6
0.4
0.2
0.0
Jul
Oct
Jan
2021
Apr

#### Nepal

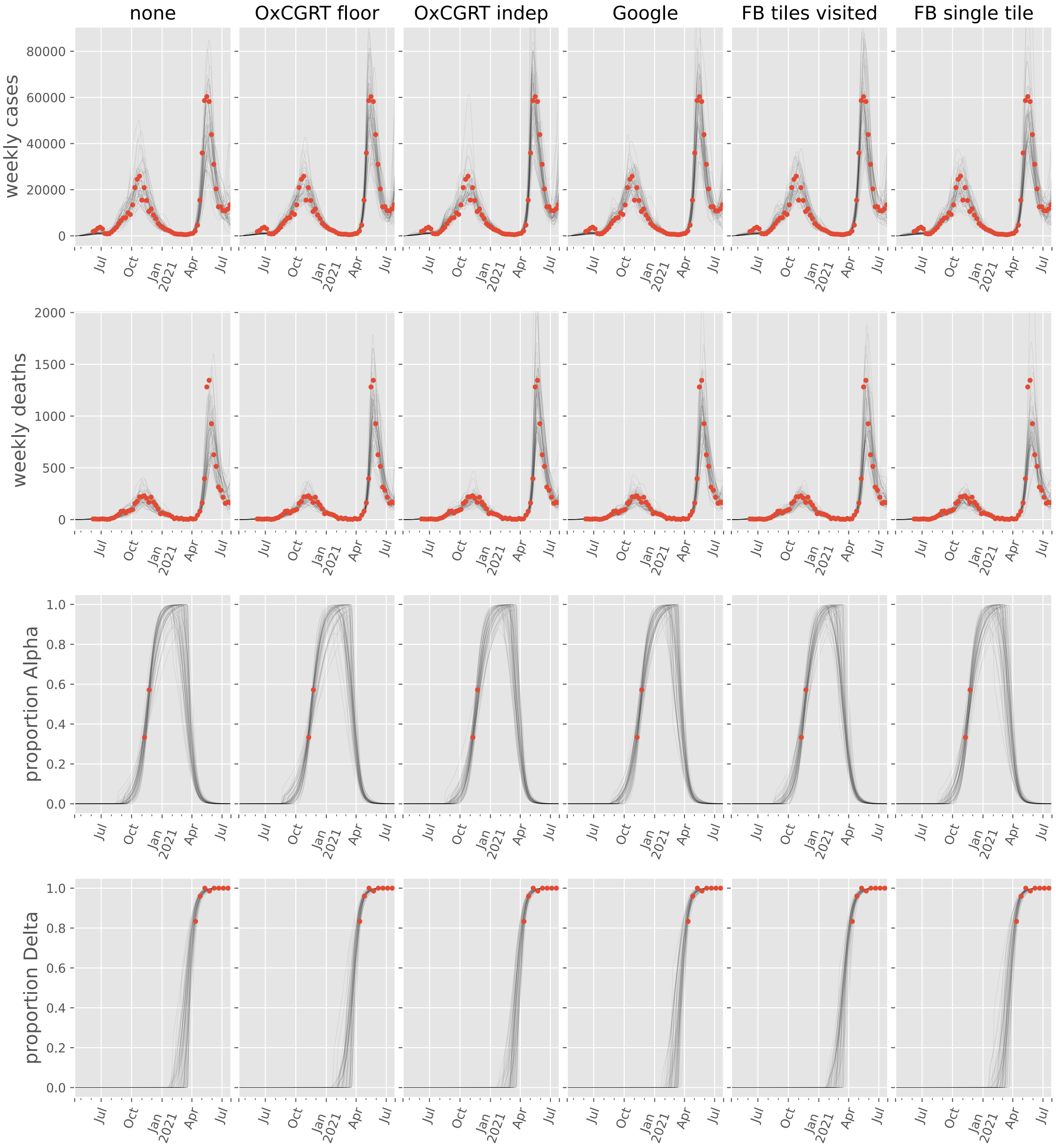
none
OxCGRT floor
OxCGRT indep
Google
FB tiles visited
FB single tile
weekly cases
weekly deaths
proportion Alpha
proportion Delta
80000
60000
40000
20000
0
2000
1500
1000
500
1.0
0.8
0.6
0.4
0.2
0.0
Jul
Oct
Jan
2021
Apr

#### Oman

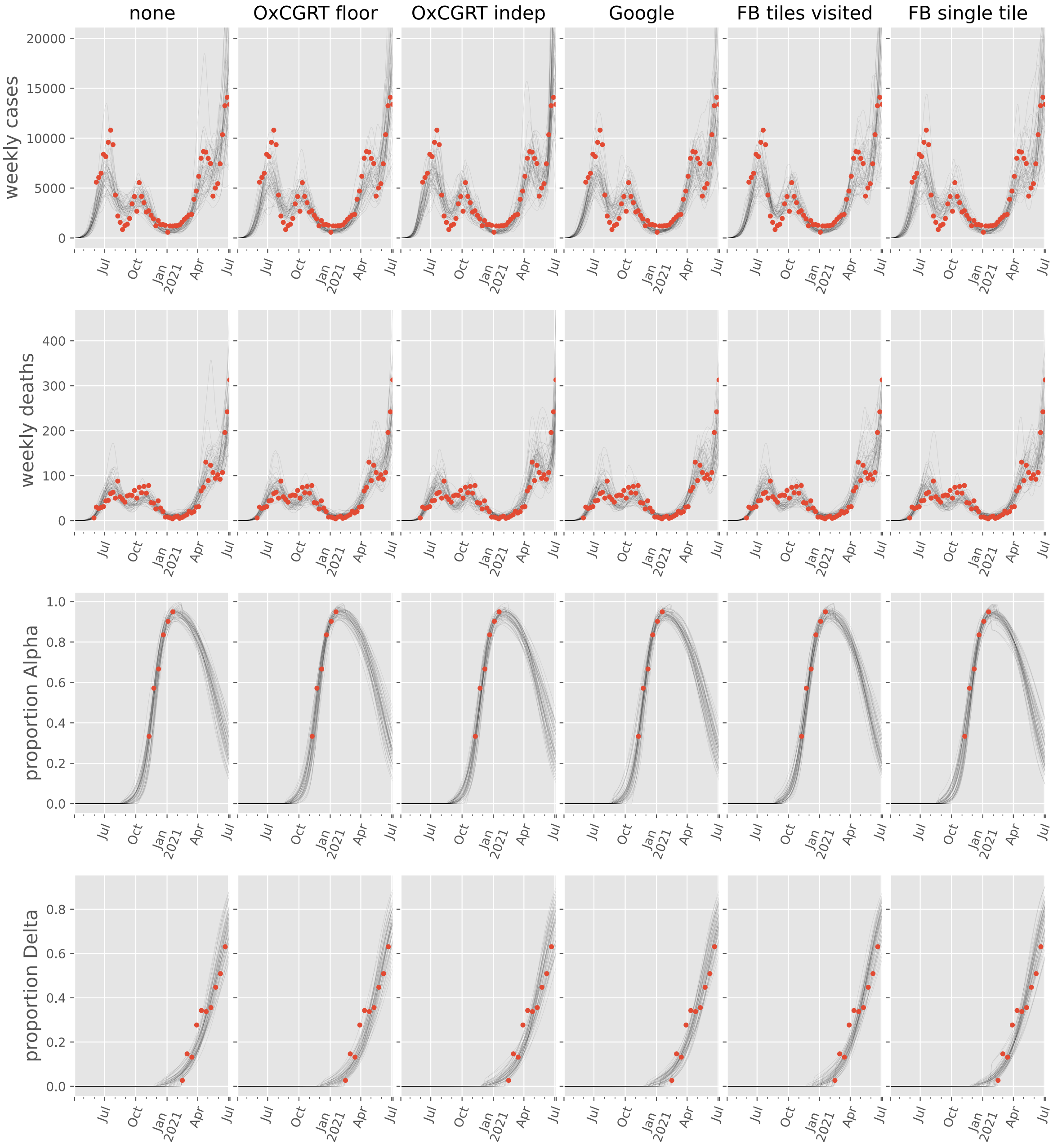
none
OxCGRT floor
OxCGRT indep
Google
FB tiles visited
FB single tile
weekly cases
weekly deaths
proportion Alpha
proportion Delta
Jul
Oct
Jan 2021
Apr

#### Pakistan

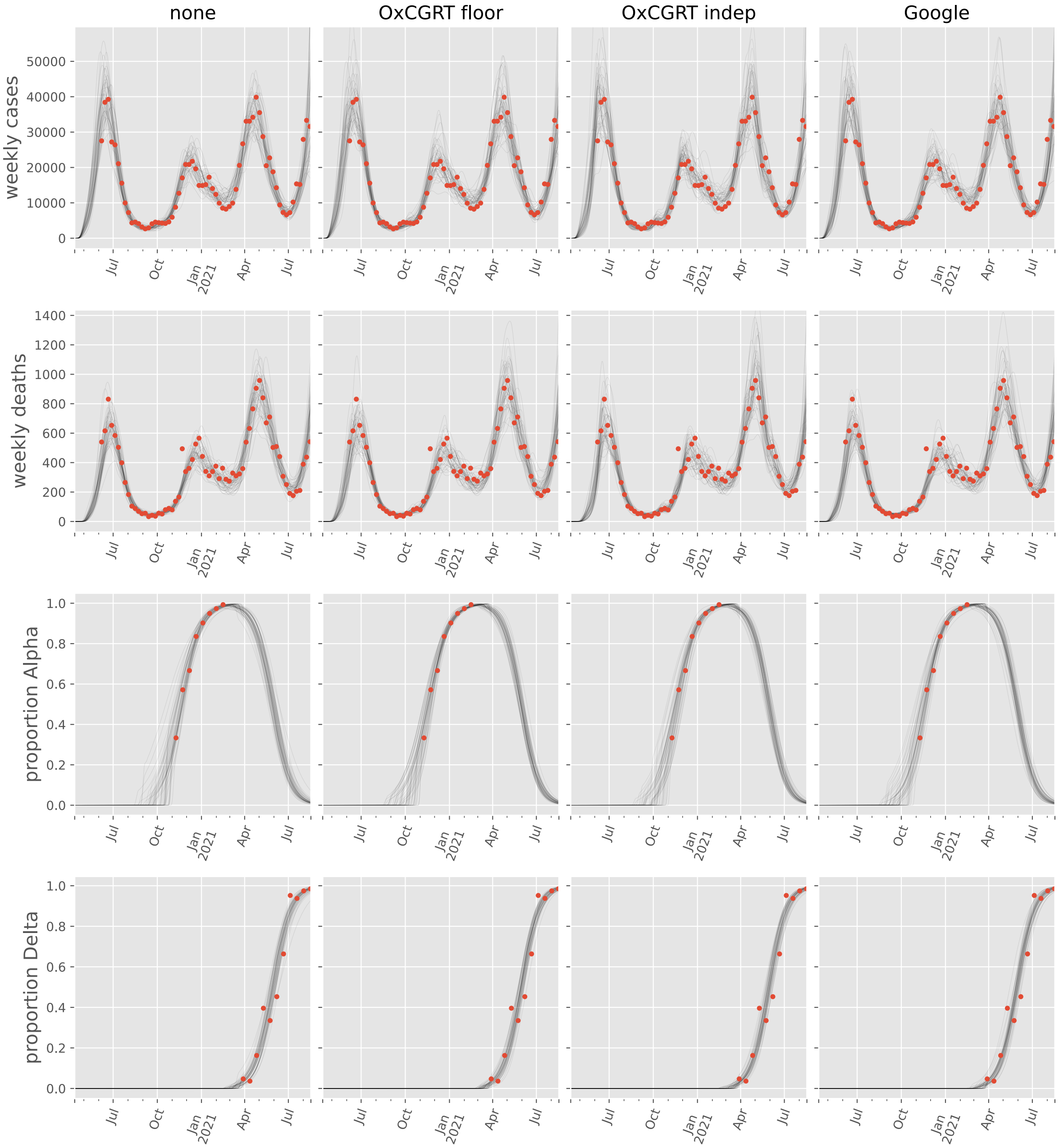

none
OxCGRT floor
OxCGRT indep
Google
weekly cases
weekly deaths
proportion Alpha
proportion Delta
Jul
Oct
Jan
2021
Apr
Jul

#### Palestine, State of

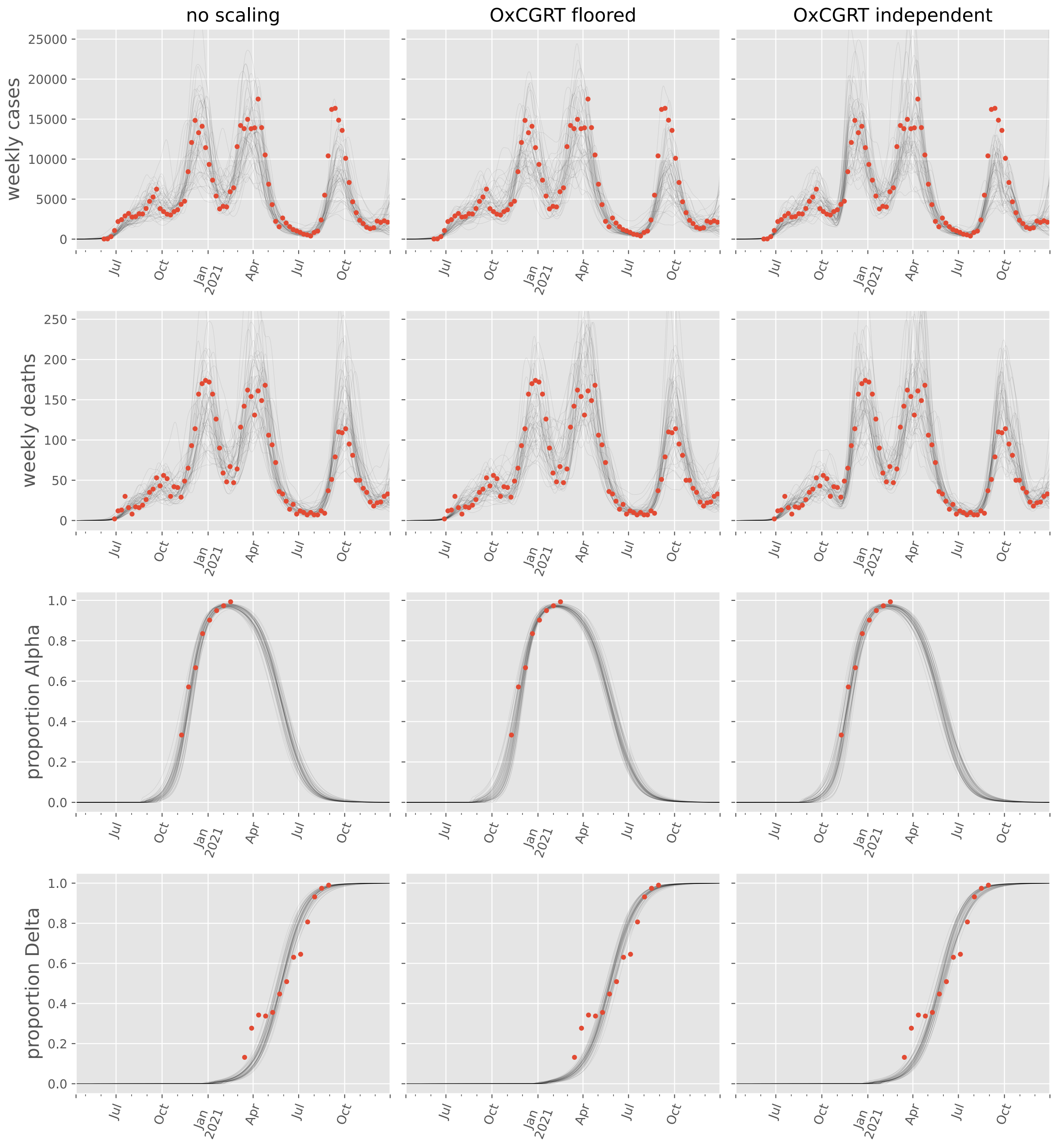

#### Philippines

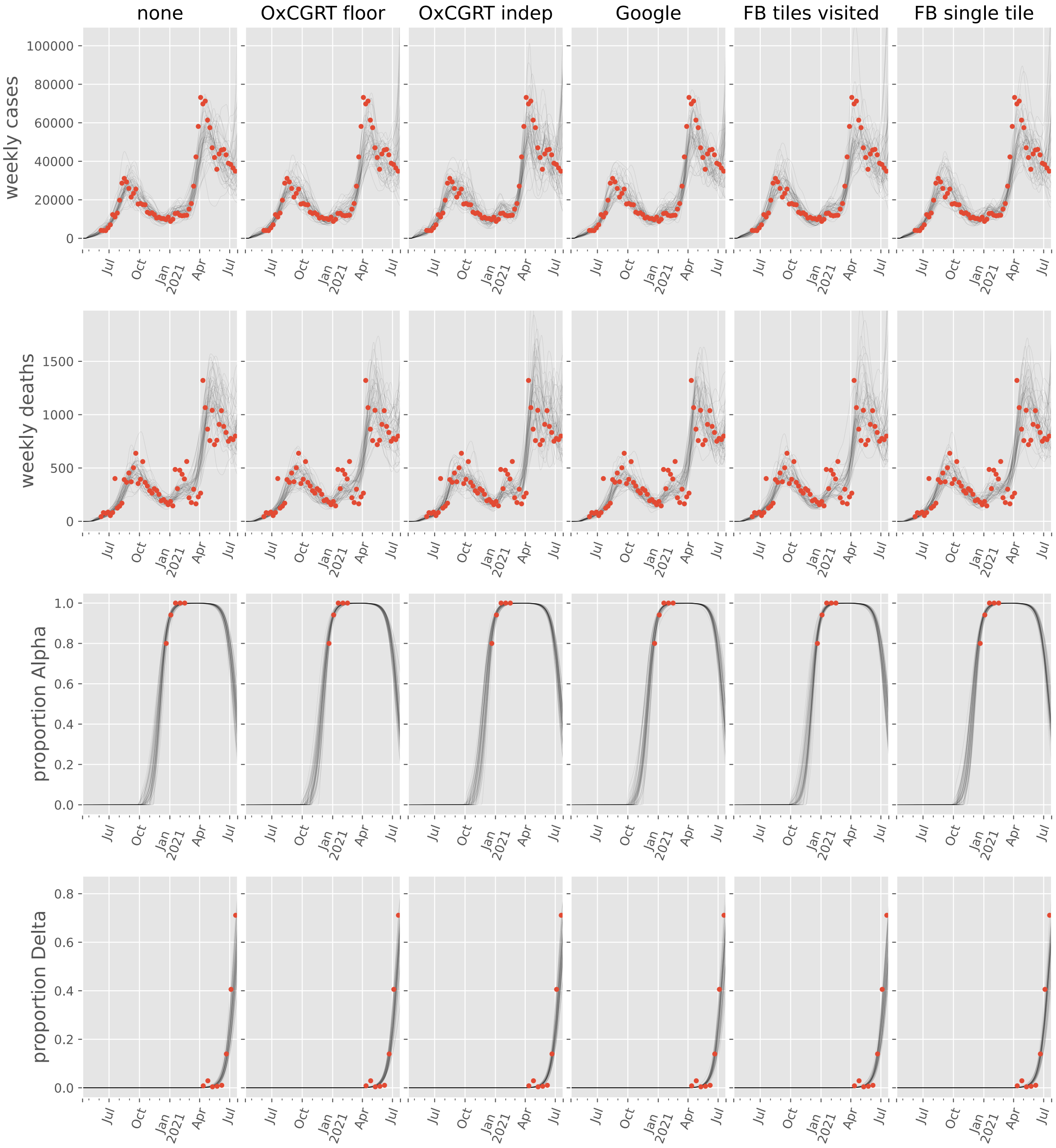

#### Qatar

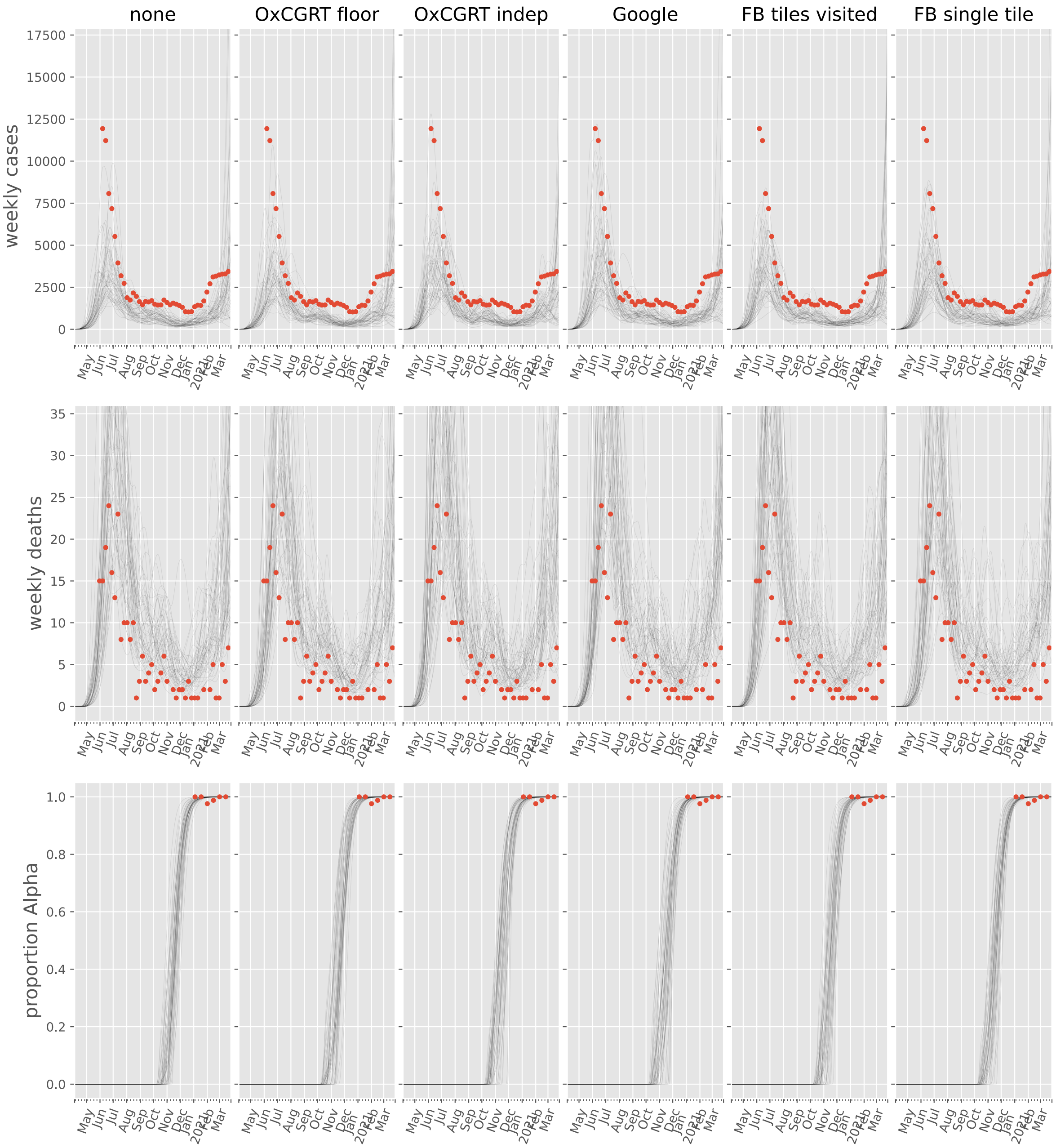

none
OxCGRT floor
OxCGRT indep
Google
FB tiles visited
FB single tile
weekly cases
weekly deaths
proportion Alpha
May
Jun
Jul
Aug
Sep
Oct
Nov
Dec
Jan
2021
Feb
Mar

#### Saudi Arabia

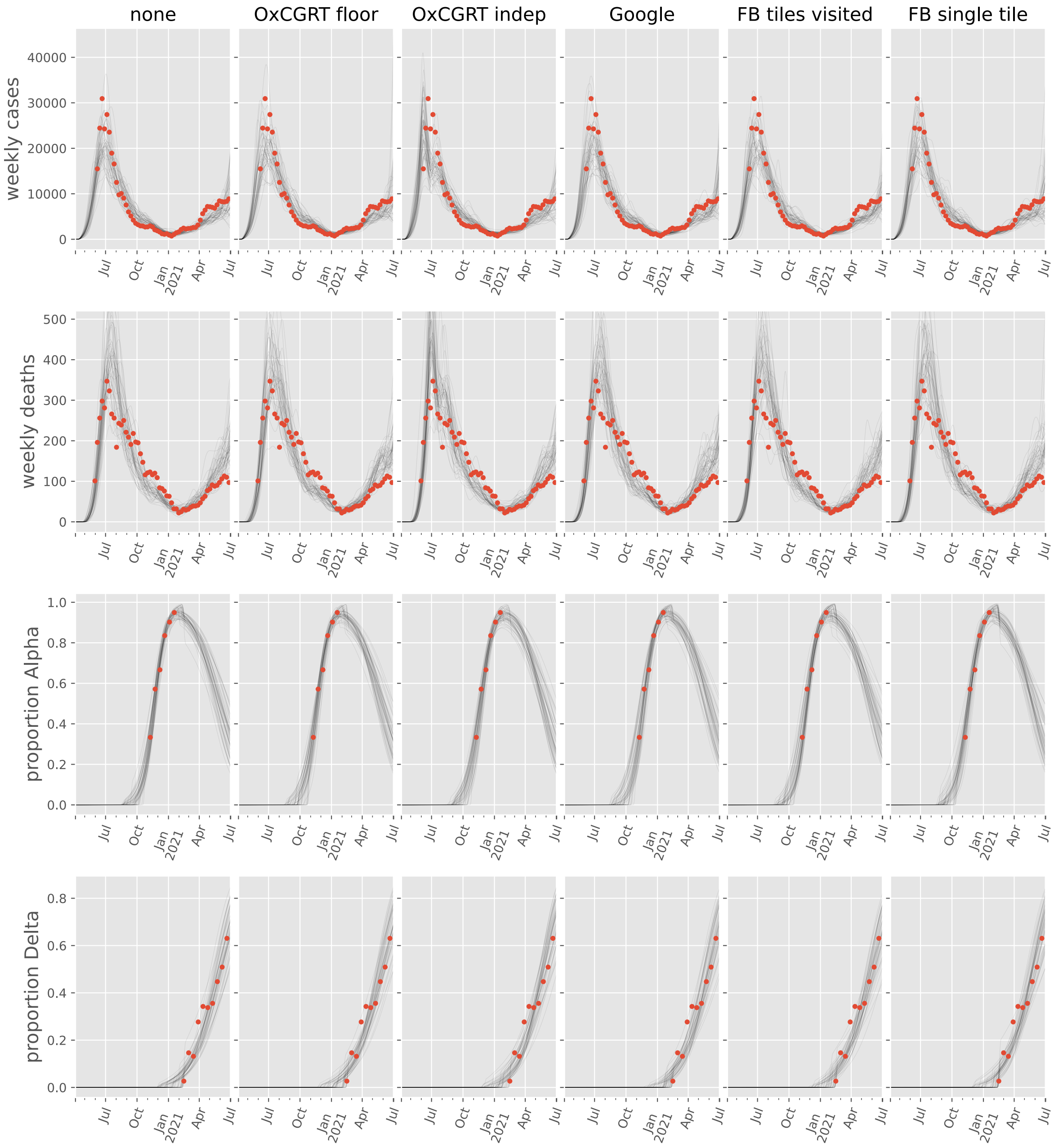

#### Sri Lanka

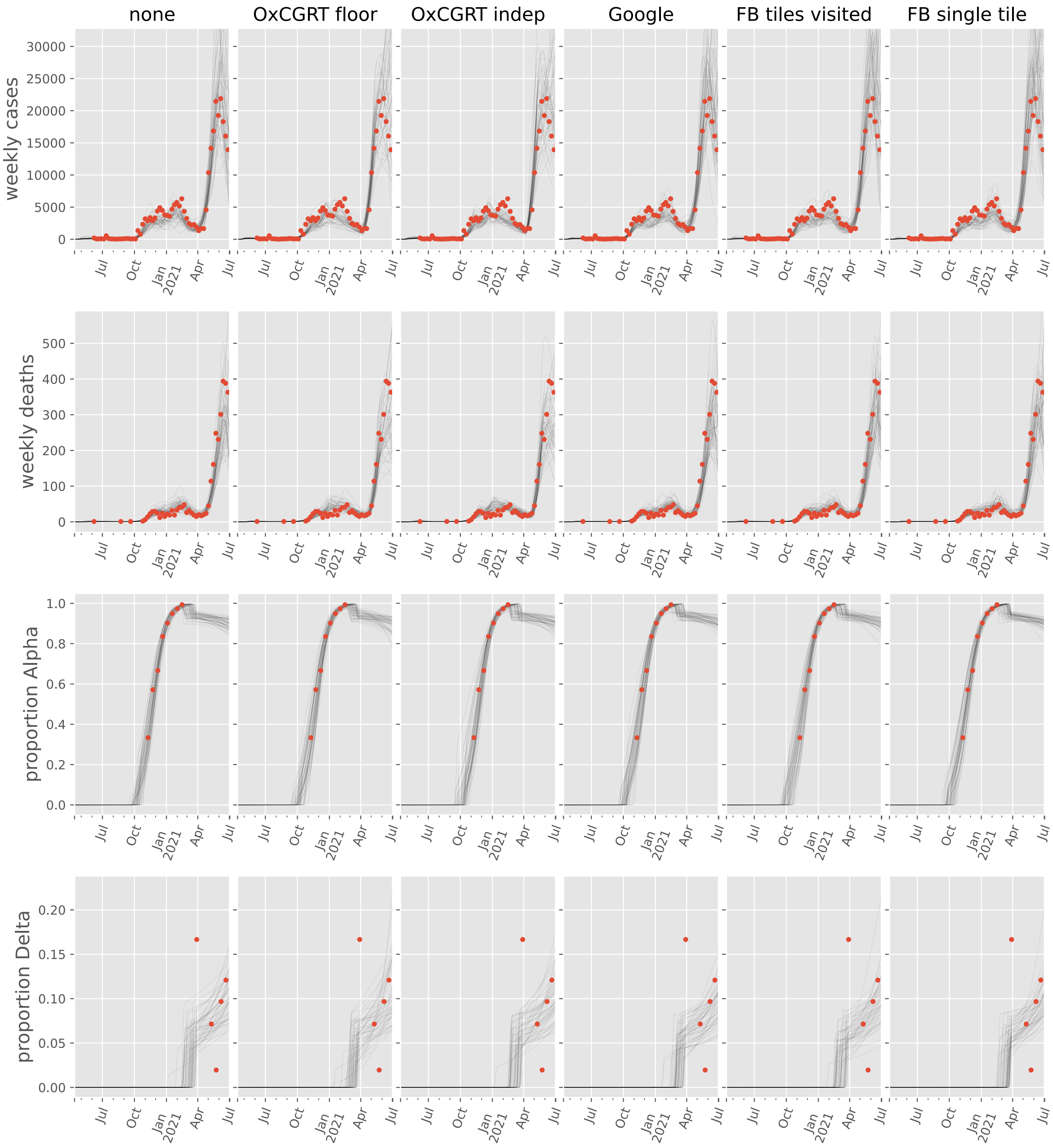

none
OxCGRT floor
OxCGRT indep
Google
FB tiles visited
FB single tile
weekly cases
weekly deaths
proportion Alpha
proportion Delta
30000
25000
20000
15000
10000
5000
0
500
400
300
200
100
1.0
0.8
0.6
0.4
0.2
0.0
0.20
0.15
0.10
0.05
0.00
Jul
Oct
Jan 2021
Apr

#### Syrian Arab Republic

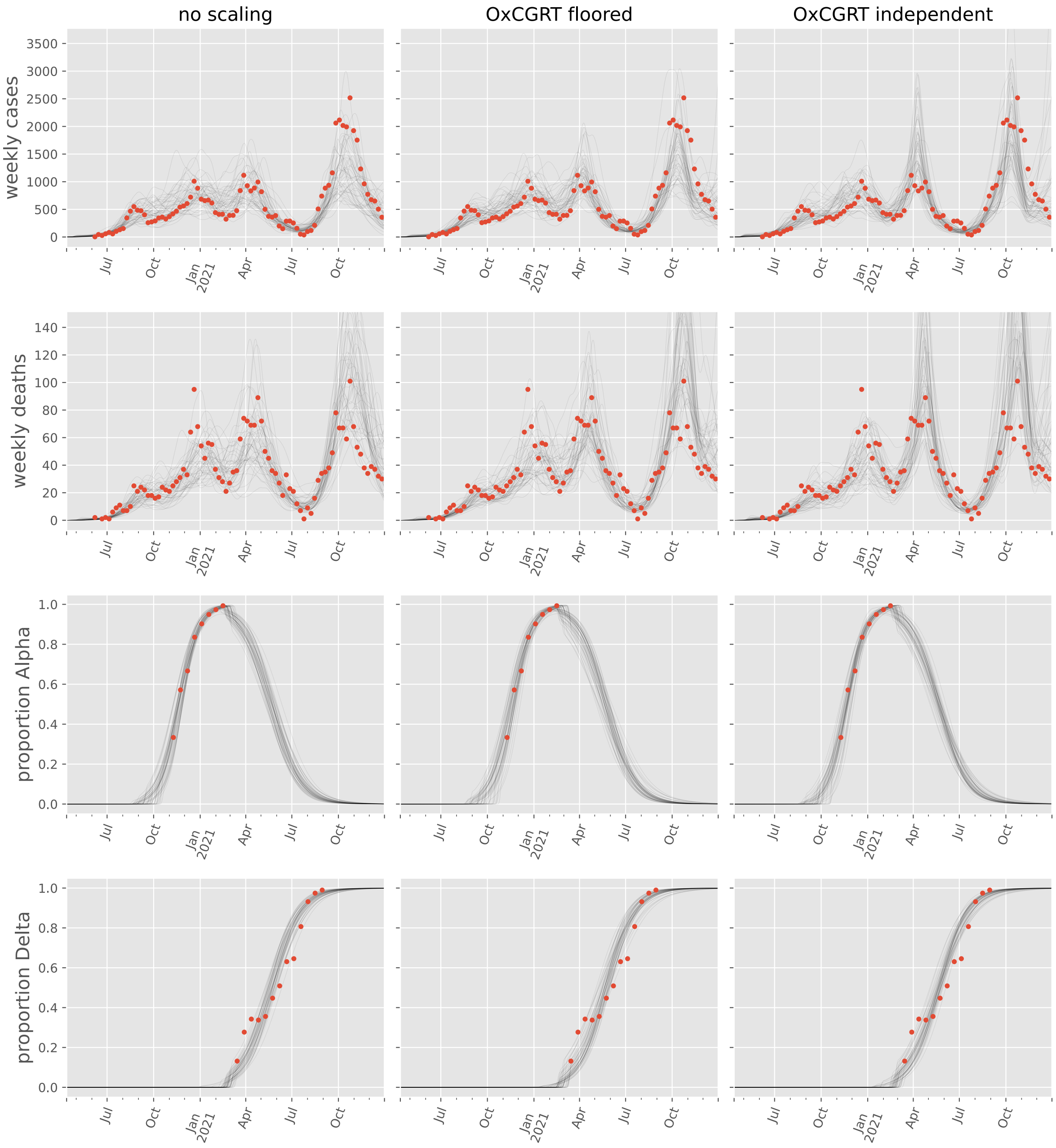

#### Türkiye

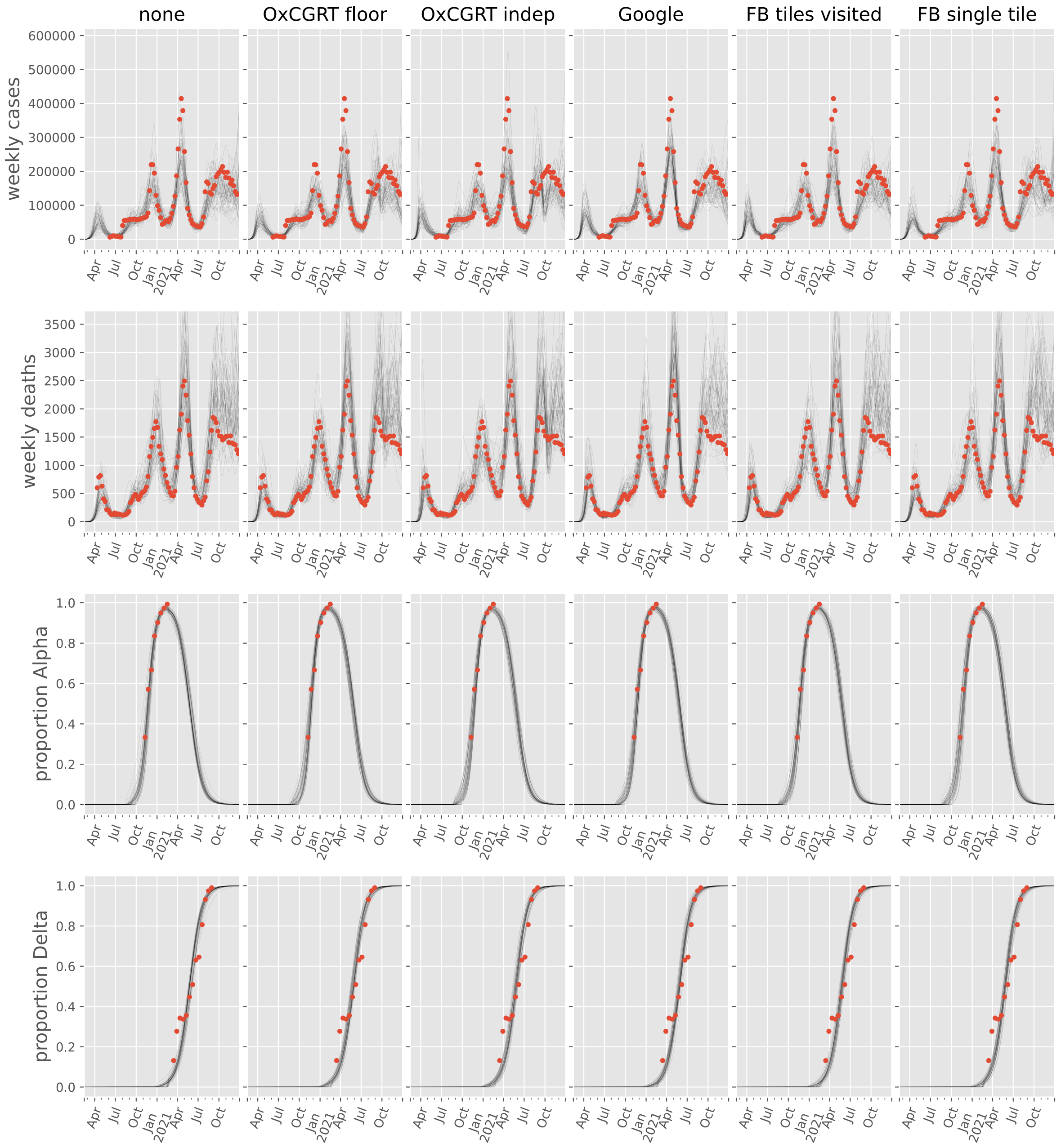
none
OxCGRT floor
OxCGRT indep
Google
FB tiles visited
FB single tile
weekly cases
weekly deaths
proportion Alpha
proportion Delta
Apr
Jul
Oct
Jan
2021
Apr
Jul
Oct

#### United Arab Emirates

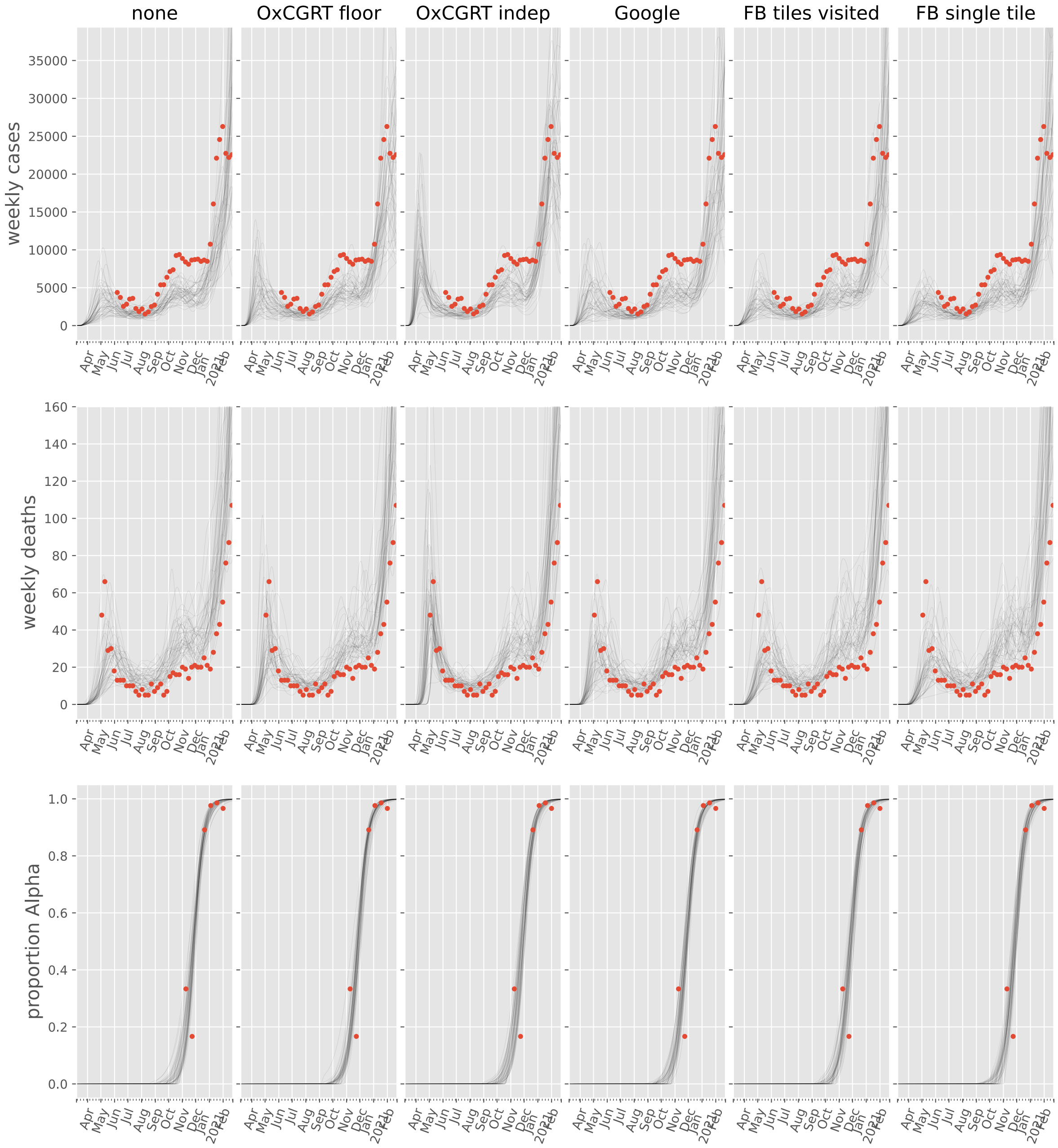

#### Yemen

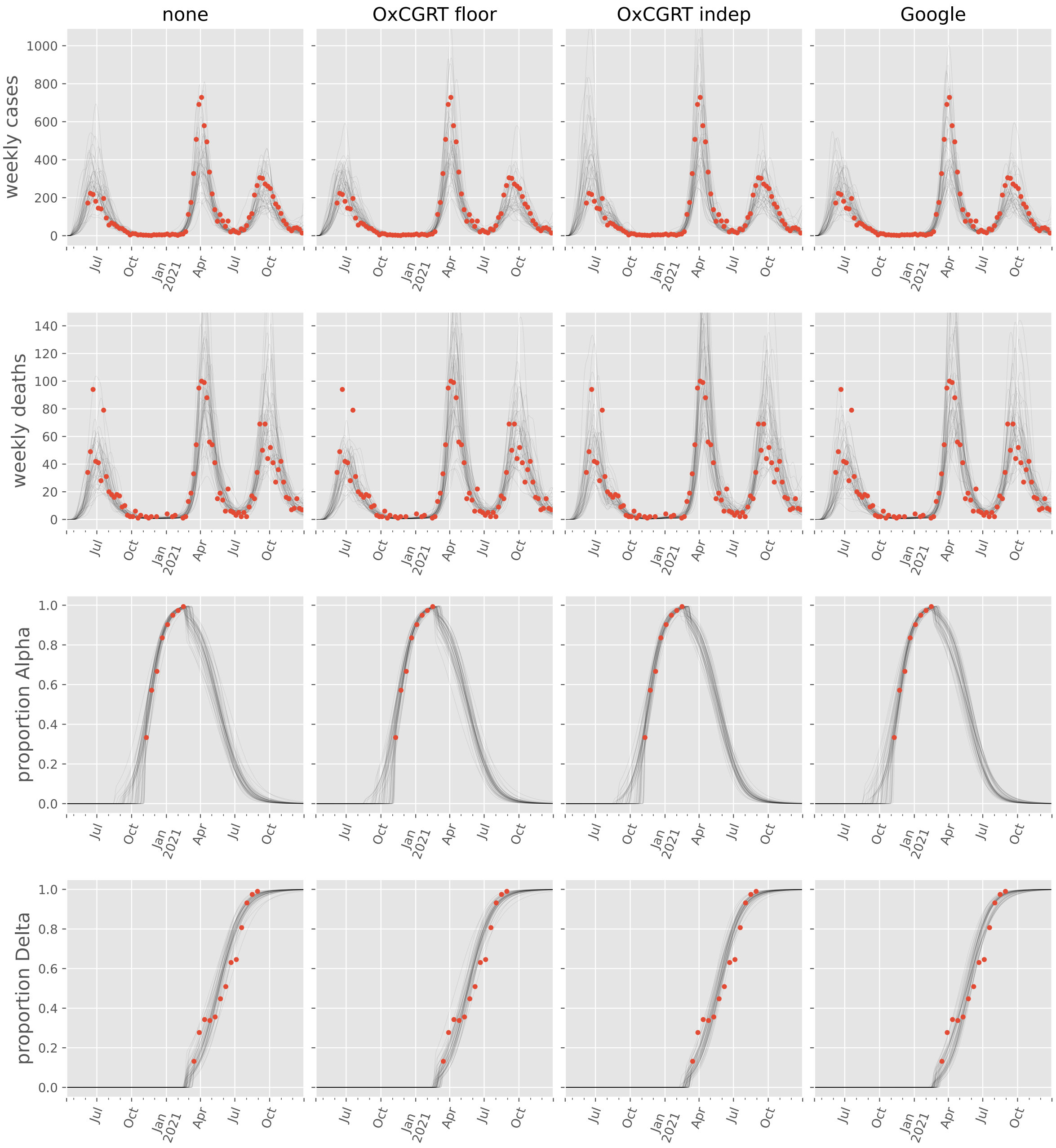

none
OxCGRT floor
OxCGRT indep
Google
weekly cases
weekly deaths
proportion Alpha
proportion Delta
Jul
Oct
Jan
2021
Apr

### Europe

#### Albania

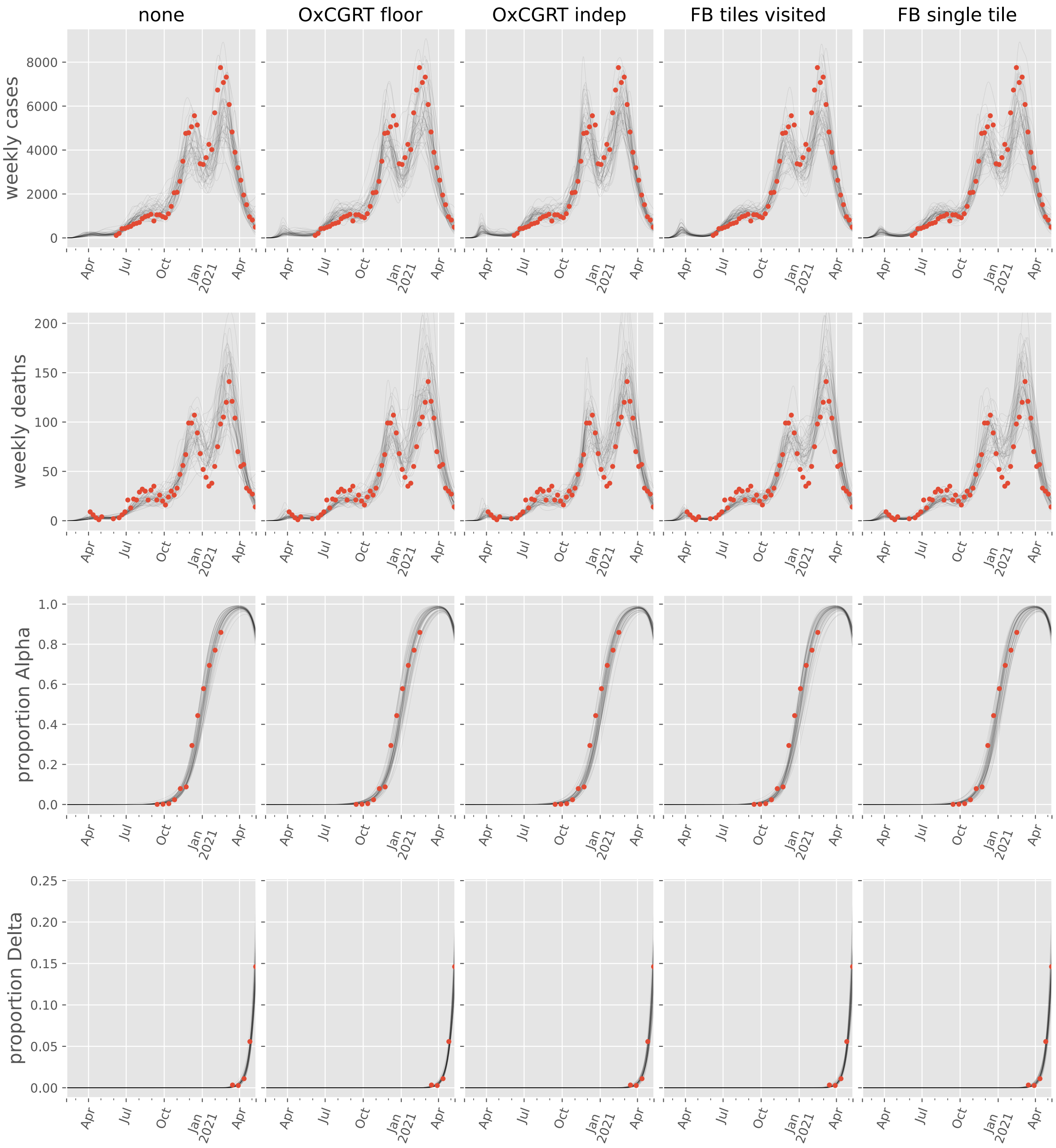

#### Andorra

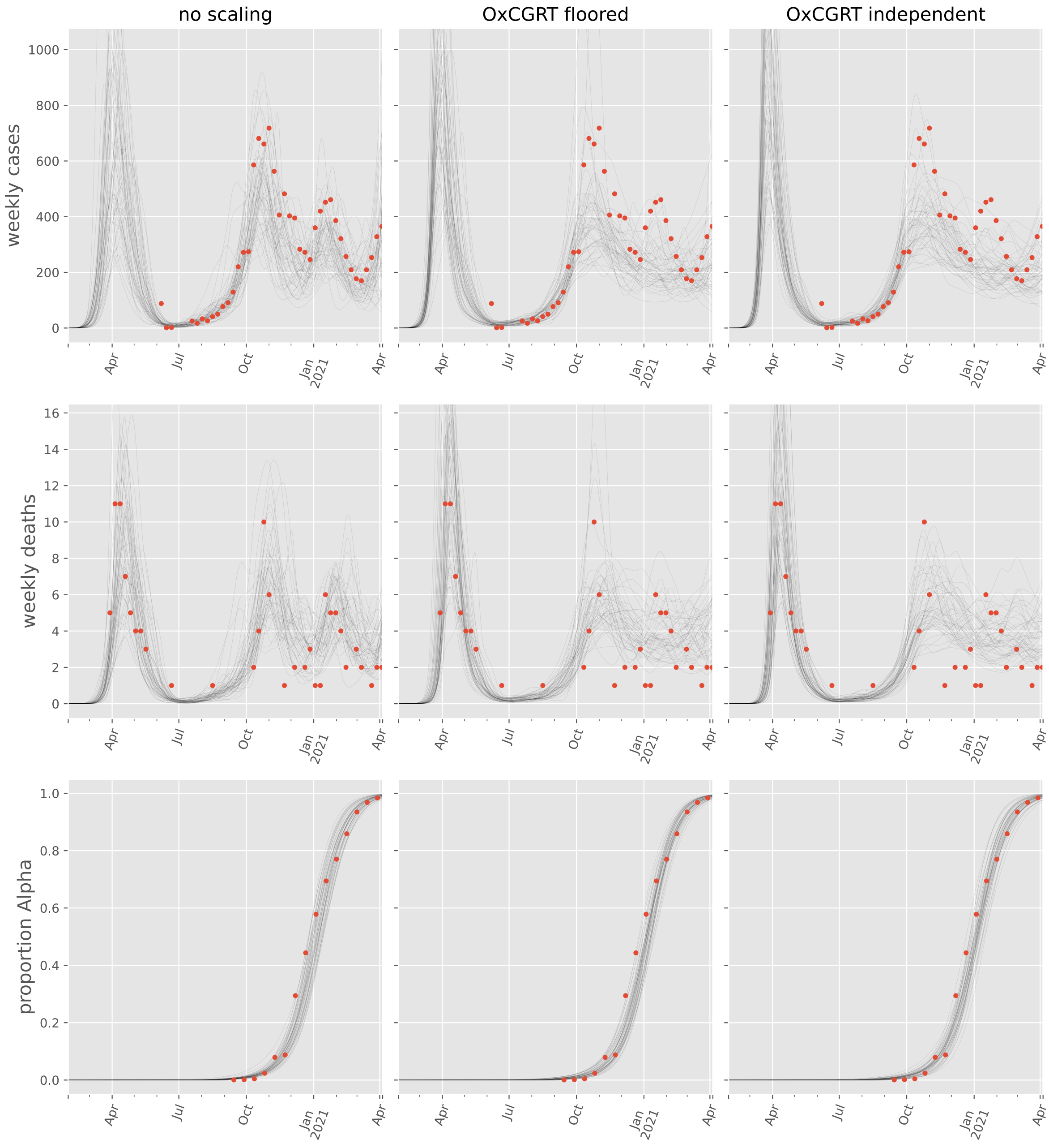

no scaling
OxCGRT floored
OxCGRT independent
weekly cases
weekly deaths
proportion Alpha
1000
800
600
400
200
0
16
14
12
10
8
6
4
2
0
1.0
0.8
0.6
0.4
0.2
0.0
Apr
Jul
Oct
Jan
2021
Apr

#### Austria

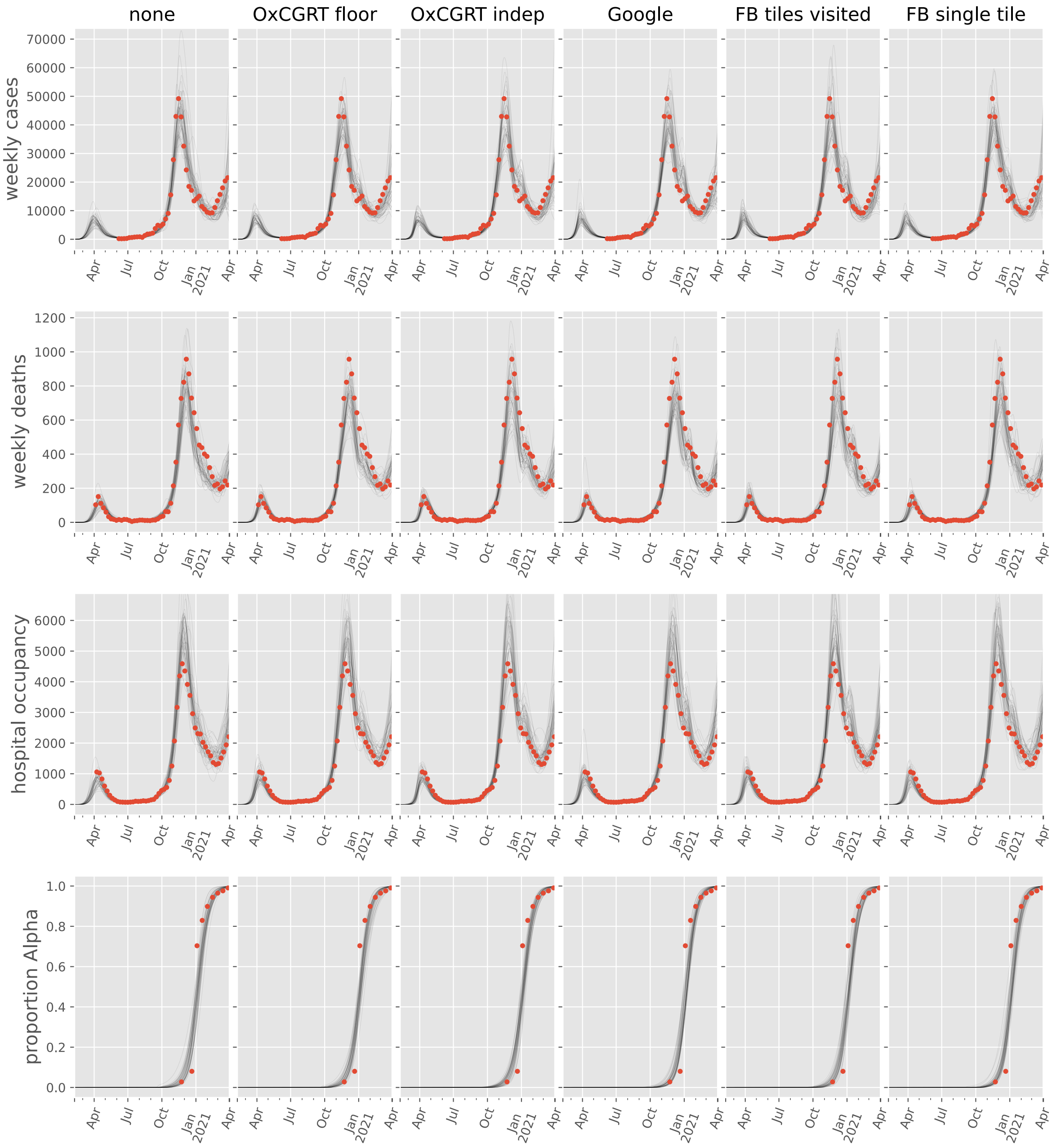
none
OxCGRT floor
OxCGRT indep
Google
FB tiles visited
FB single tile
weekly cases
weekly deaths
hospital occupancy
proportion Alpha
Apr
Jul
Oct
Jan
2021
Apr

#### Belarus

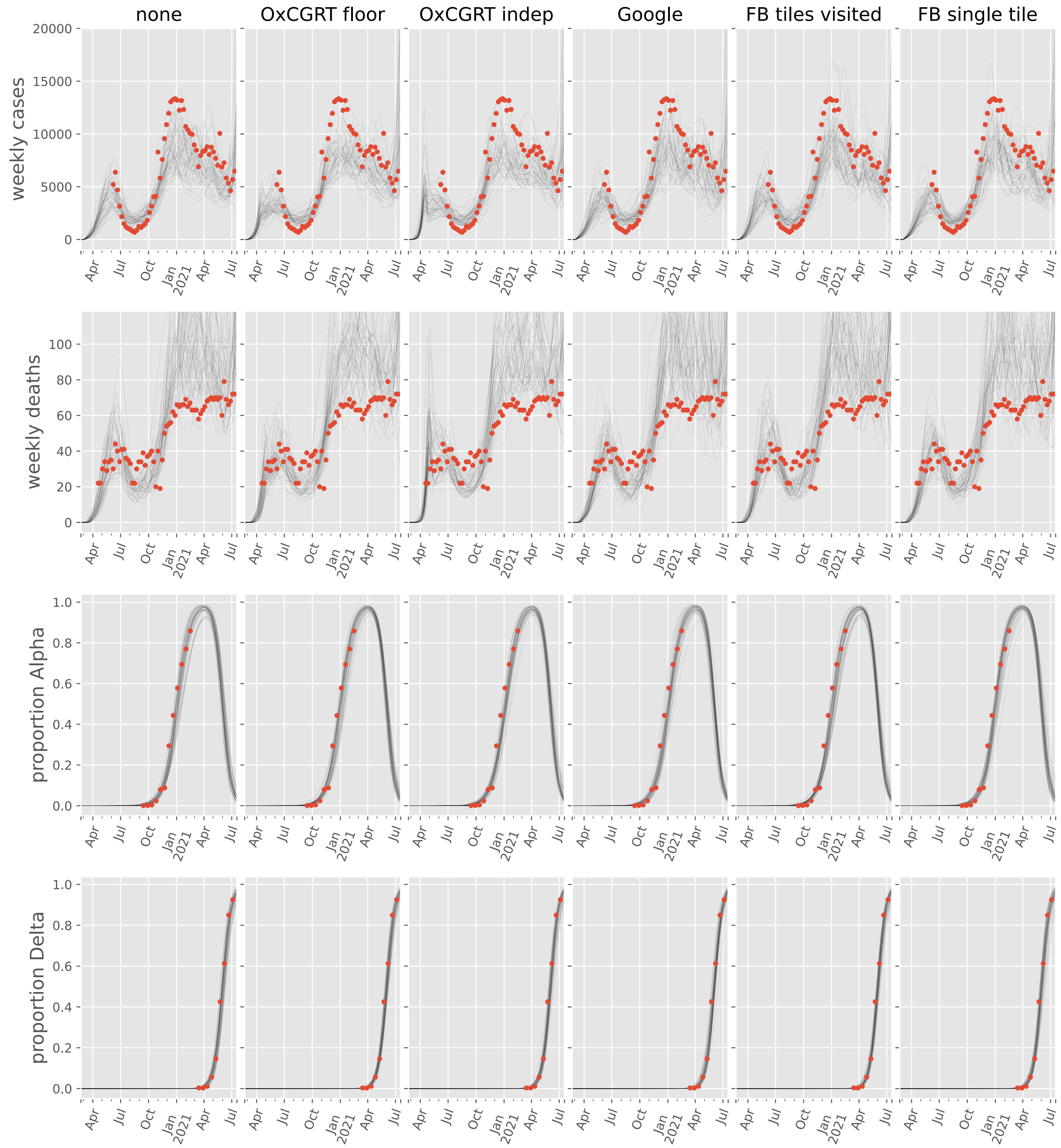
none
OxCGRT floor
OxCGRT indep
Google
FB tiles visited
FB single tile
weekly cases
weekly deaths
proportion Alpha
proportion Delta
Apr
Jul
Oct
Jan
2021
Apr
Jul

#### Belgium

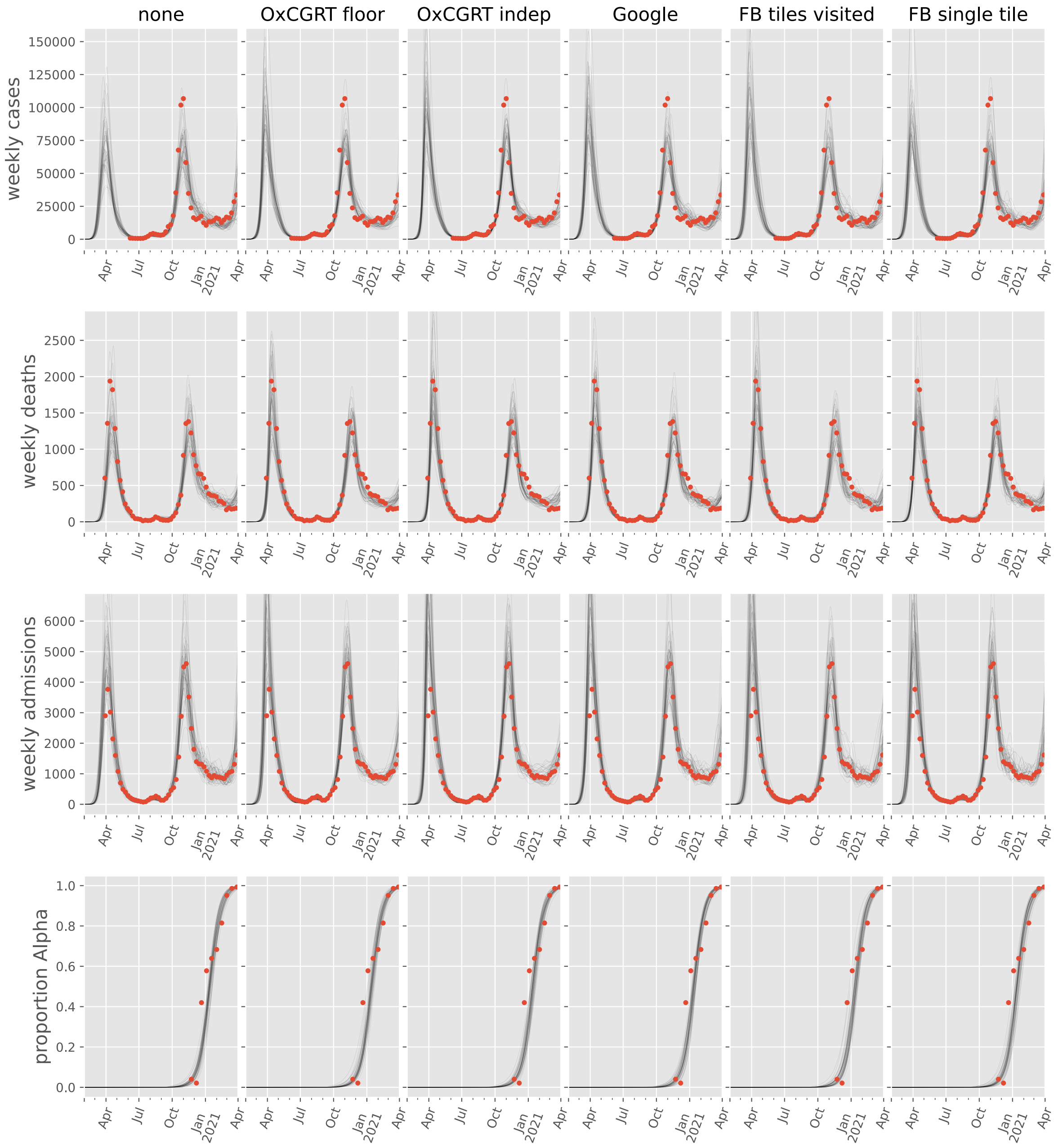

none
OxCGRT floor
OxCGRT indep
Google
FB tiles visited
FB single tile
weekly cases
weekly deaths
weekly admissions
proportion Alpha
Apr
Jul
Oct
Jan
2021

#### Bosnia and Herzegovina

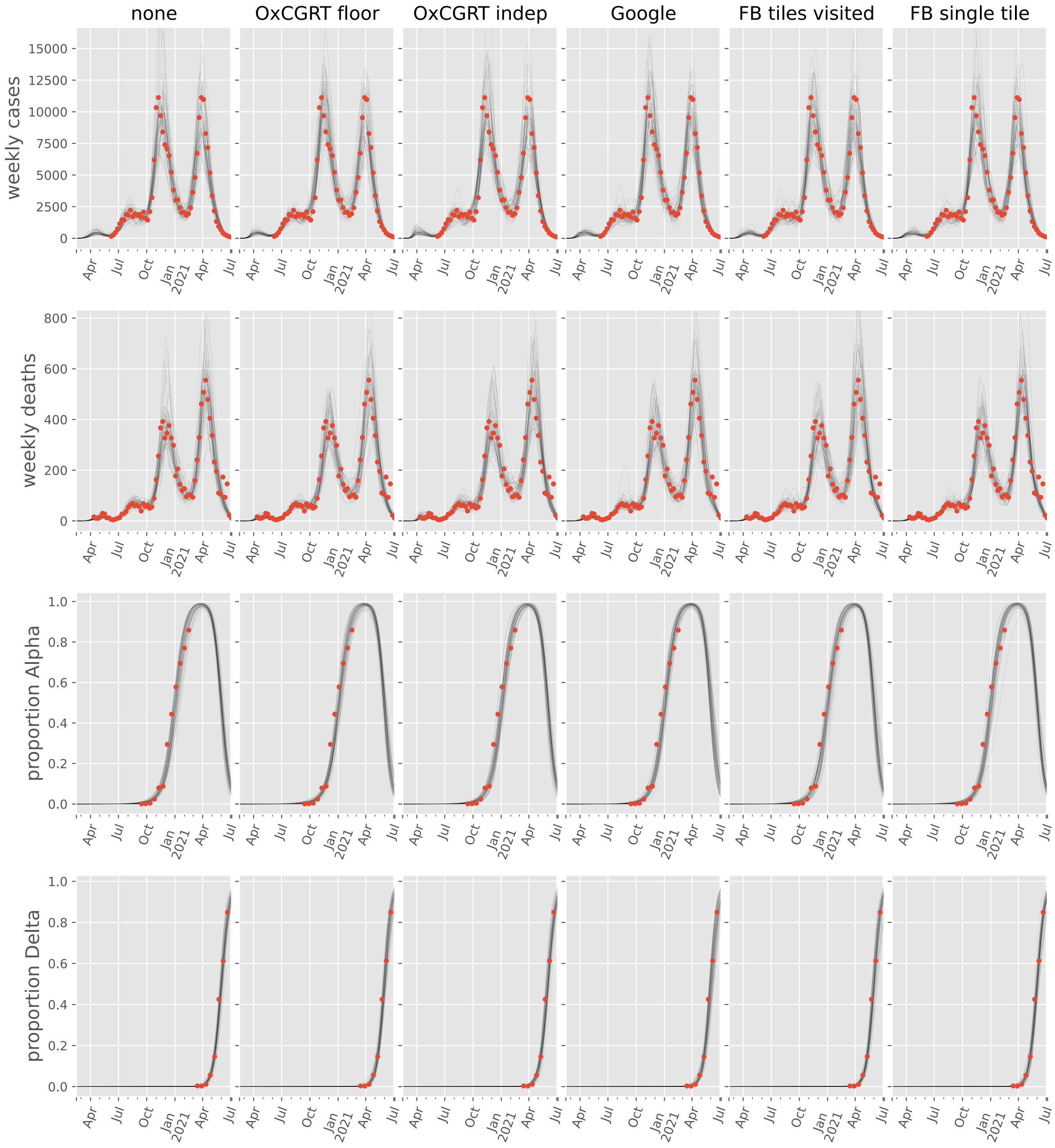

#### Bulgaria

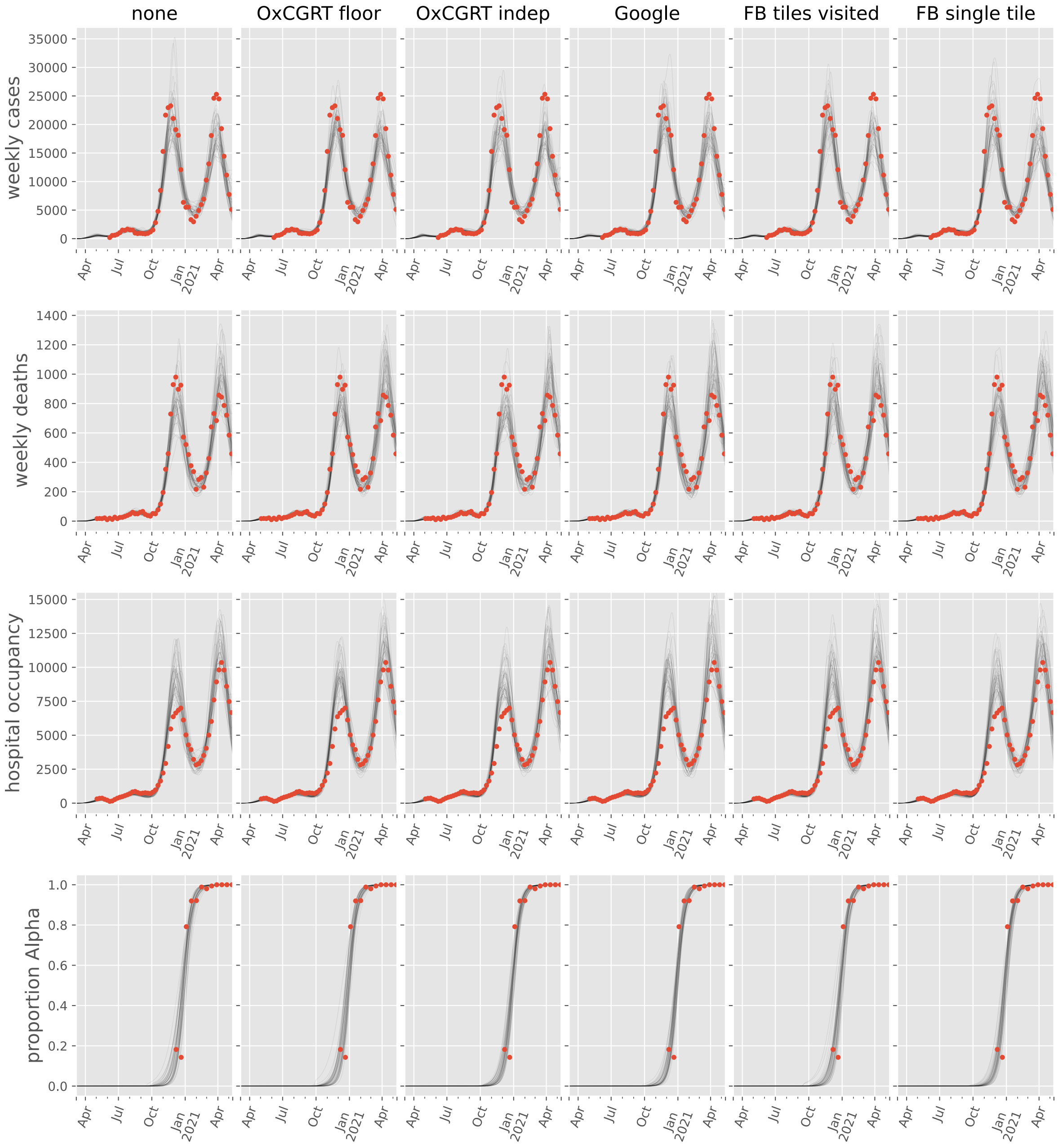

none
OxCGRT floor
OxCGRT indep
Google
FB tiles visited
FB single tile
weekly cases
weekly deaths
hospital occupancy
proportion Alpha

#### Croatia

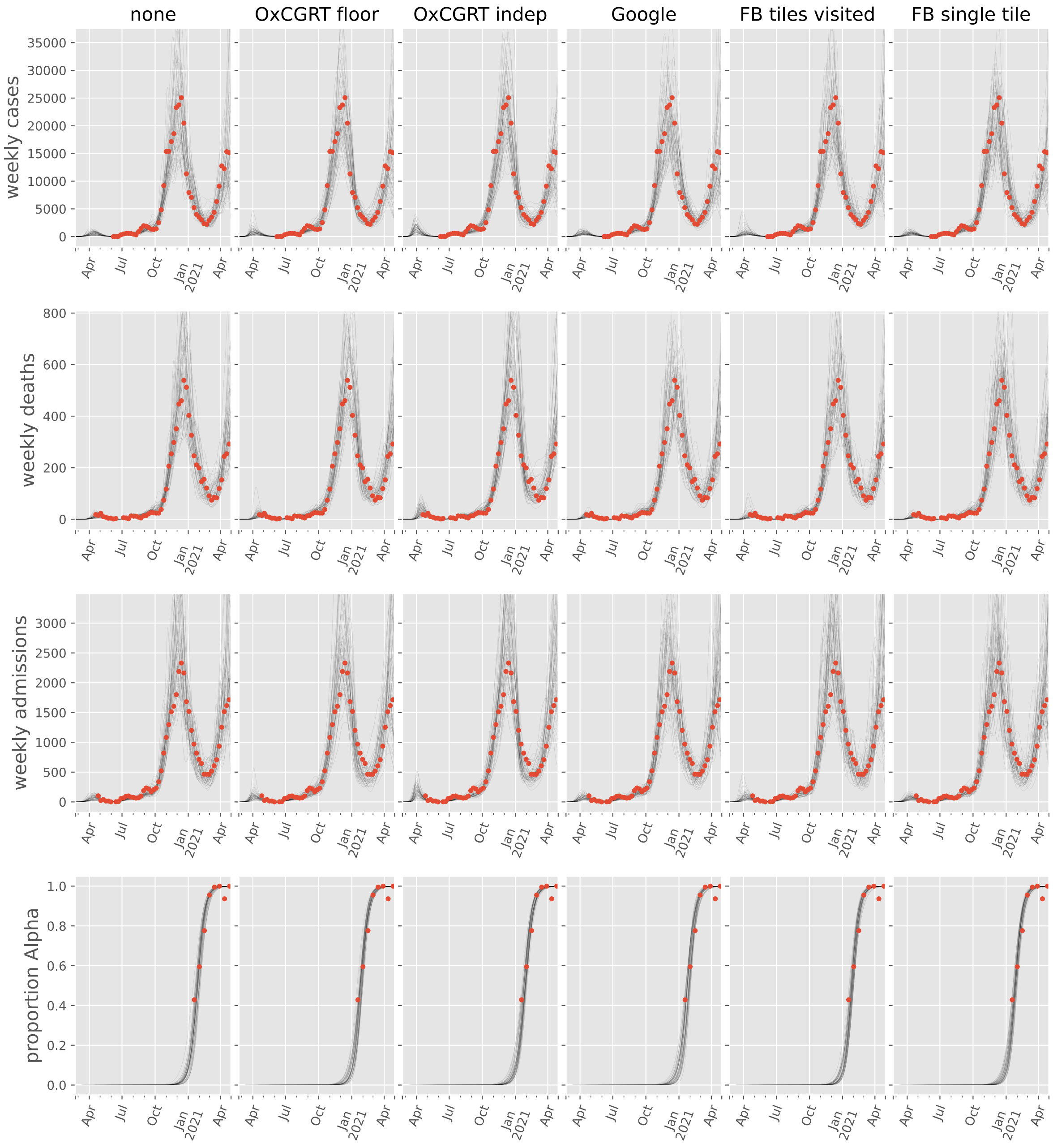

none
OxCGRT floor
OxCGRT indep
Google
FB tiles visited
FB single tile
weekly cases
weekly deaths
weekly admissions
proportion Alpha
35000
30000
25000
20000
15000
10000
5000
0
800
600
400
200
3000
2500
2000
1500
1000
500
1.0
0.8
0.6
0.4
0.2
0.0
Apr
Jul
Oct
Jan 2021
Apr

#### Czechia

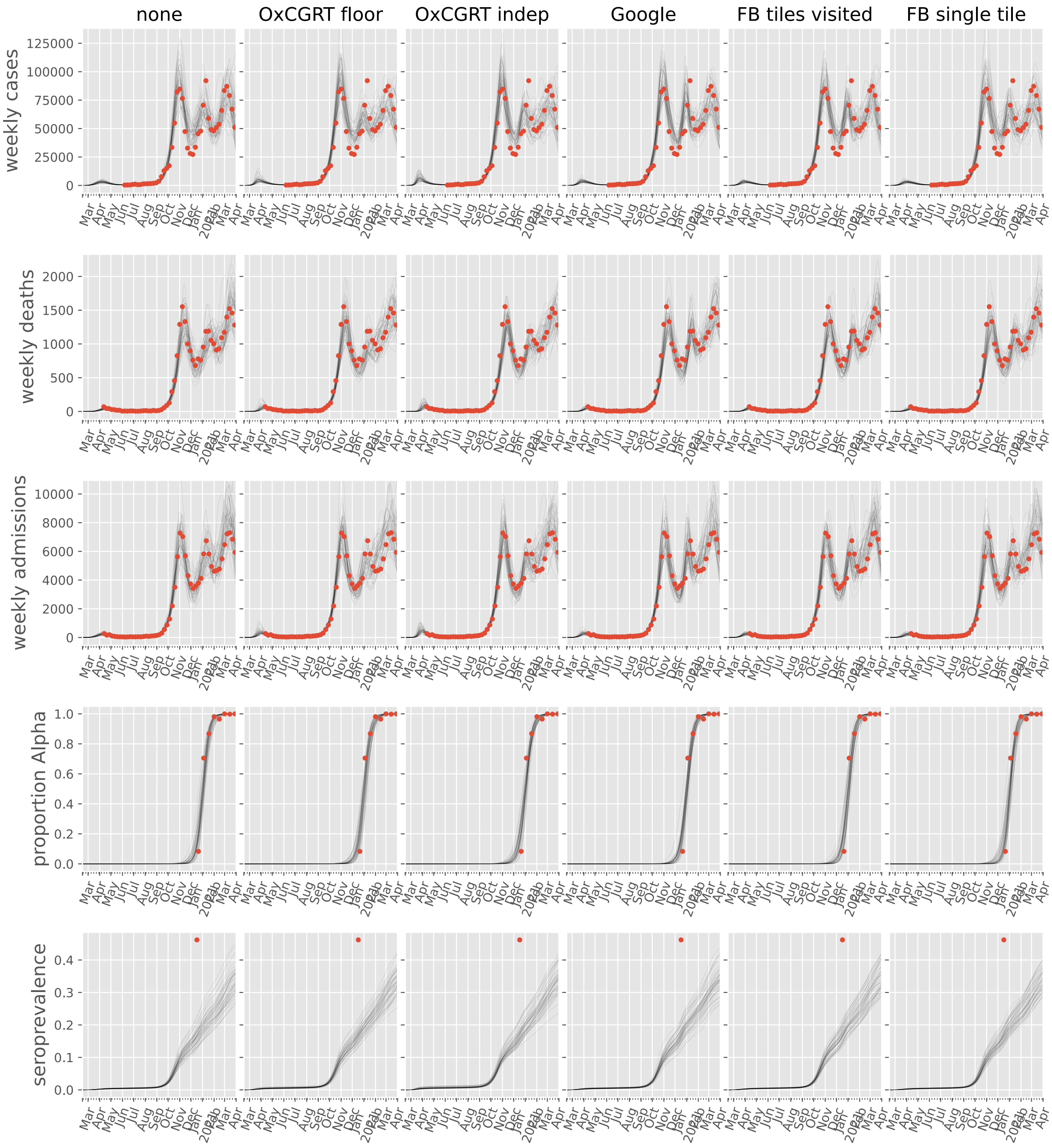

none
OxCGRT floor
OxCGRT indep
Google
FB tiles visited
FB single tile
weekly cases
weekly deaths
weekly admissions
proportion Alpha
seroprevalence

#### Denmark

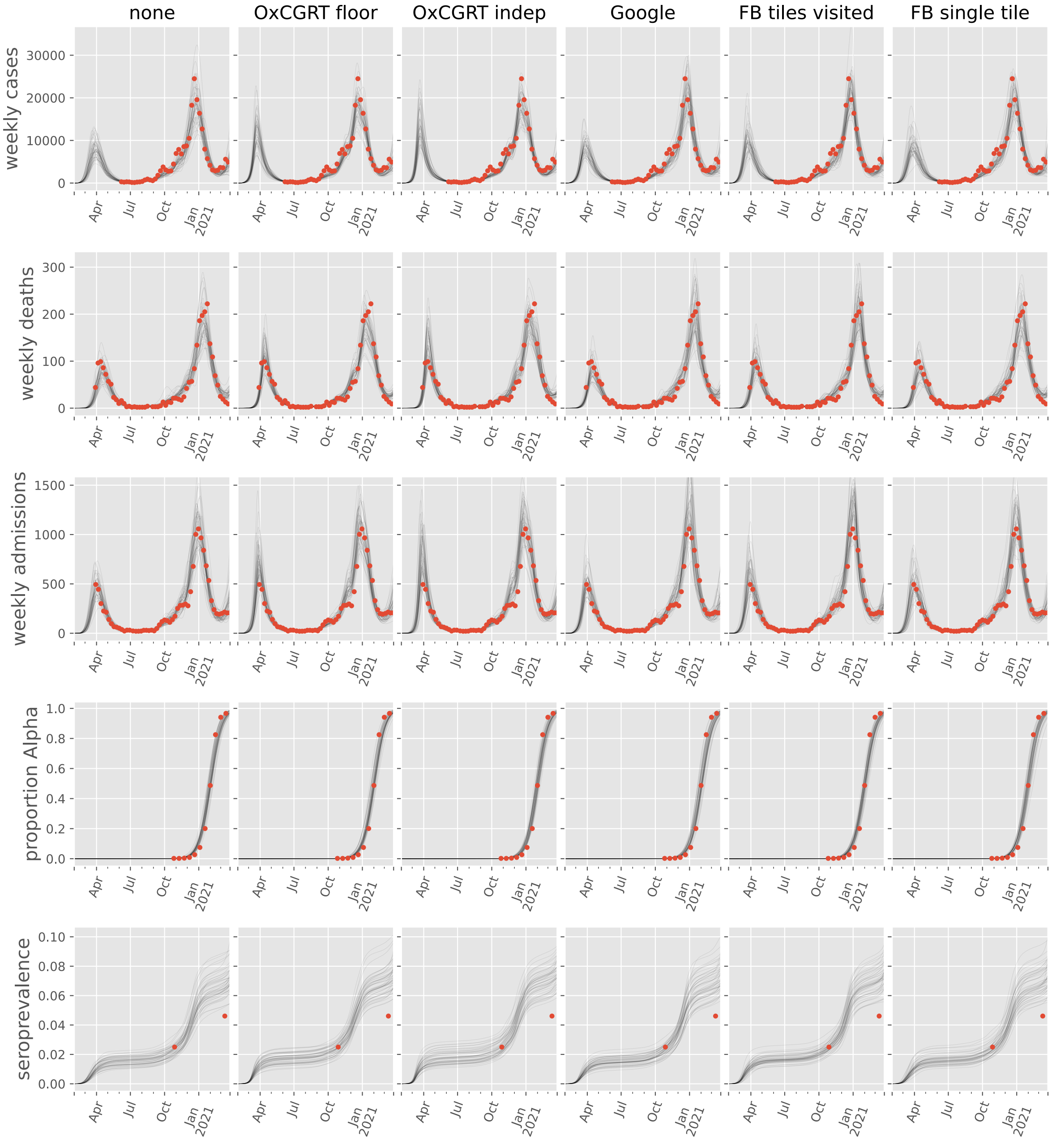

none
OxCGRT floor
OxCGRT indep
Google
FB tiles visited
FB single tile
weekly cases
weekly deaths
weekly admissions
proportion Alpha
seroprevalence
30000
20000
10000
0
300
200
100
1500
1000
500
1.0
0.8
0.6
0.4
0.2
0.0
0.10
0.08
0.06
0.04
0.02
0.00
Apr
Jul
Oct
Jan
2021

#### Estonia

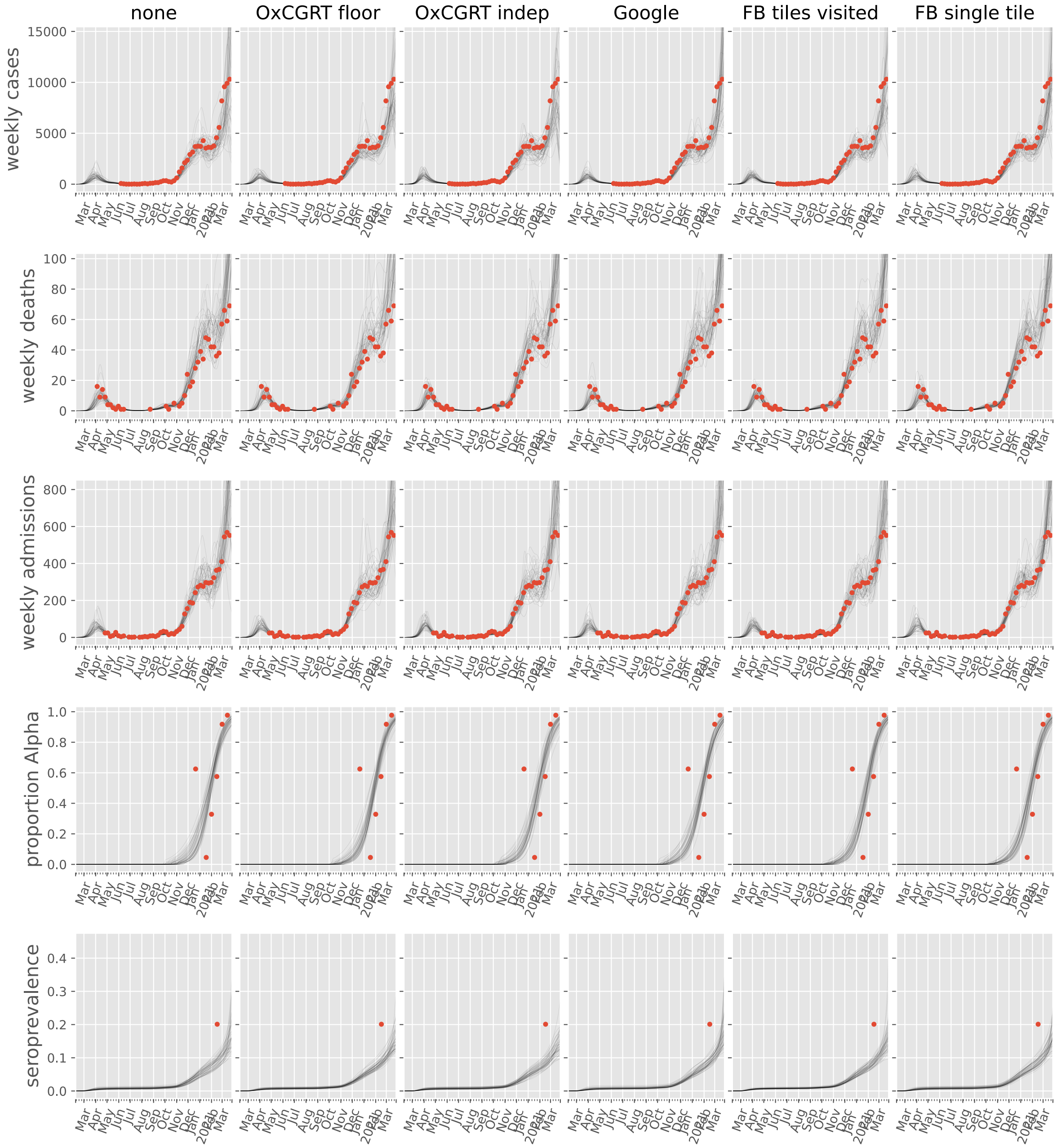

none
OxCGRT floor
OxCGRT indep
Google
FB tiles visited
FB single tile
weekly cases
weekly deaths
weekly admissions
proportion Alpha
seroprevalence

#### Finland

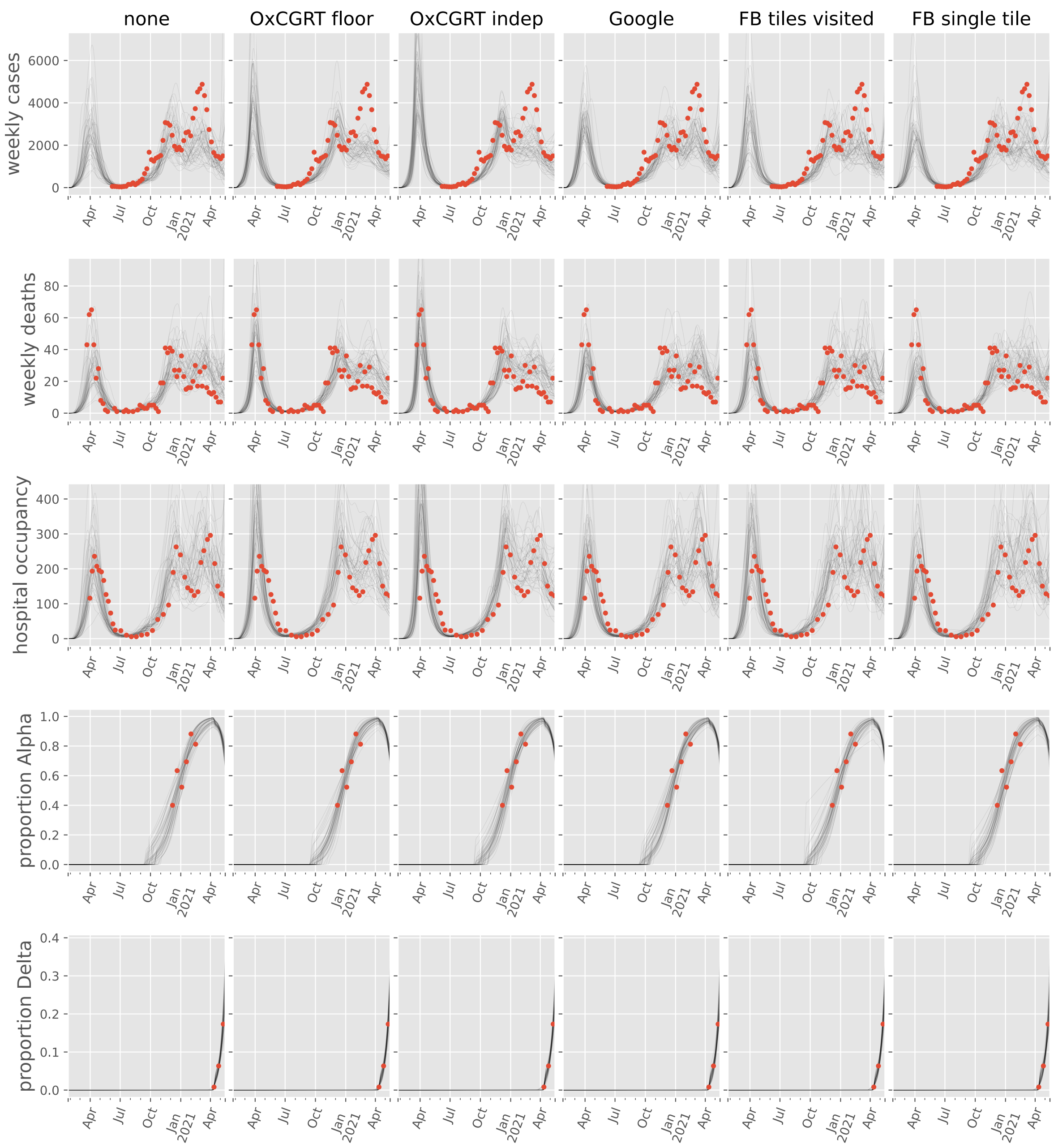

none
OxCGRT floor
OxCGRT indep
Google
FB tiles visited
FB single tile
weekly cases
weekly deaths
hospital occupancy
proportion Alpha
proportion Delta
Apr
Jul
Oct
Jan
2021
Apr

#### France

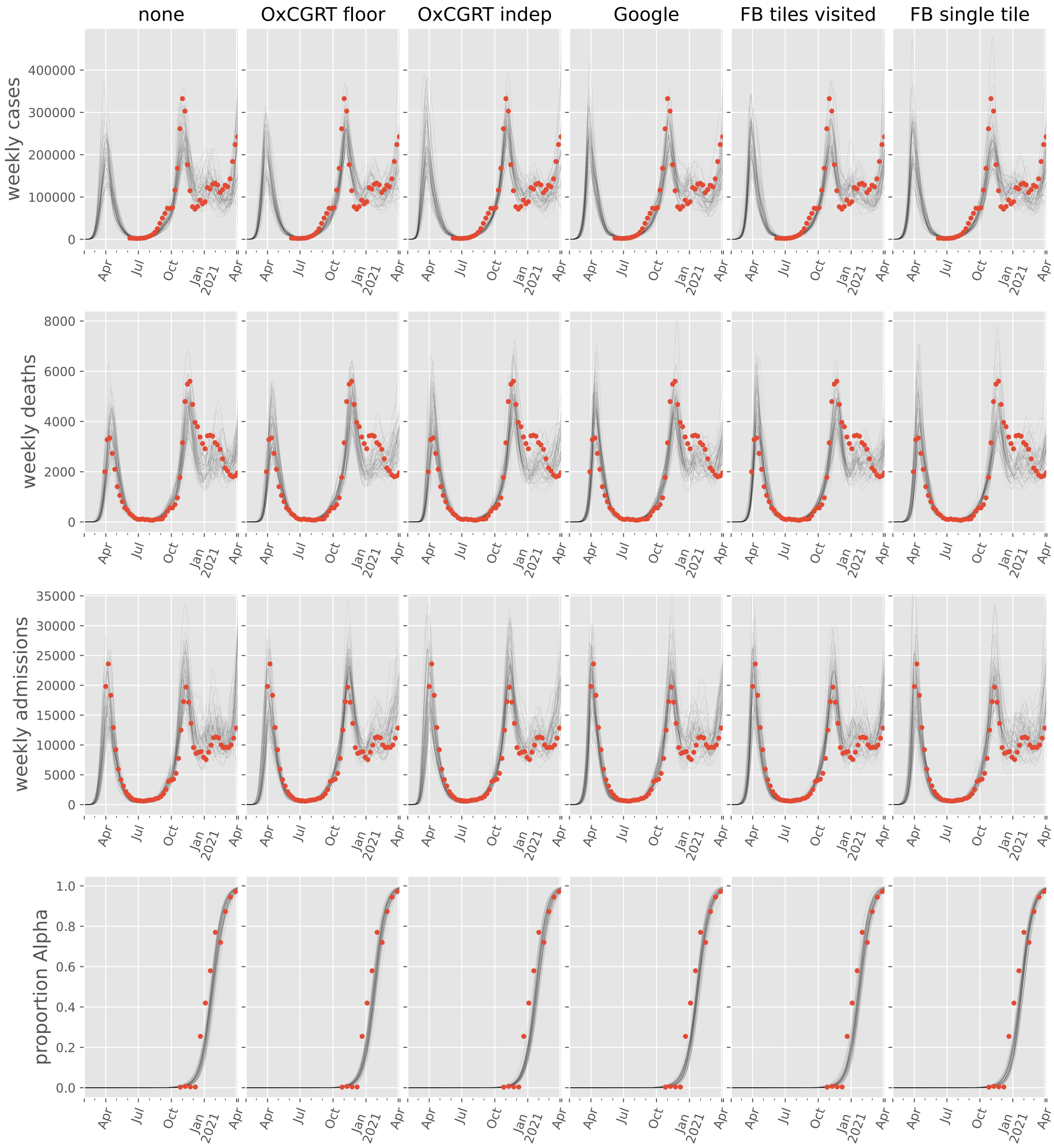
none
OxCGRT floor
OxCGRT indep
Google
FB tiles visited
FB single tile
weekly cases
weekly deaths
weekly admissions
proportion Alpha
0
100000
200000
300000
400000
2000
4000
6000
8000
5000
10000
15000
20000
25000
30000
35000
0.0
0.2
0.4
0.6
0.8
1.0
Apr
Jul
Oct
Jan
2021

#### Germany

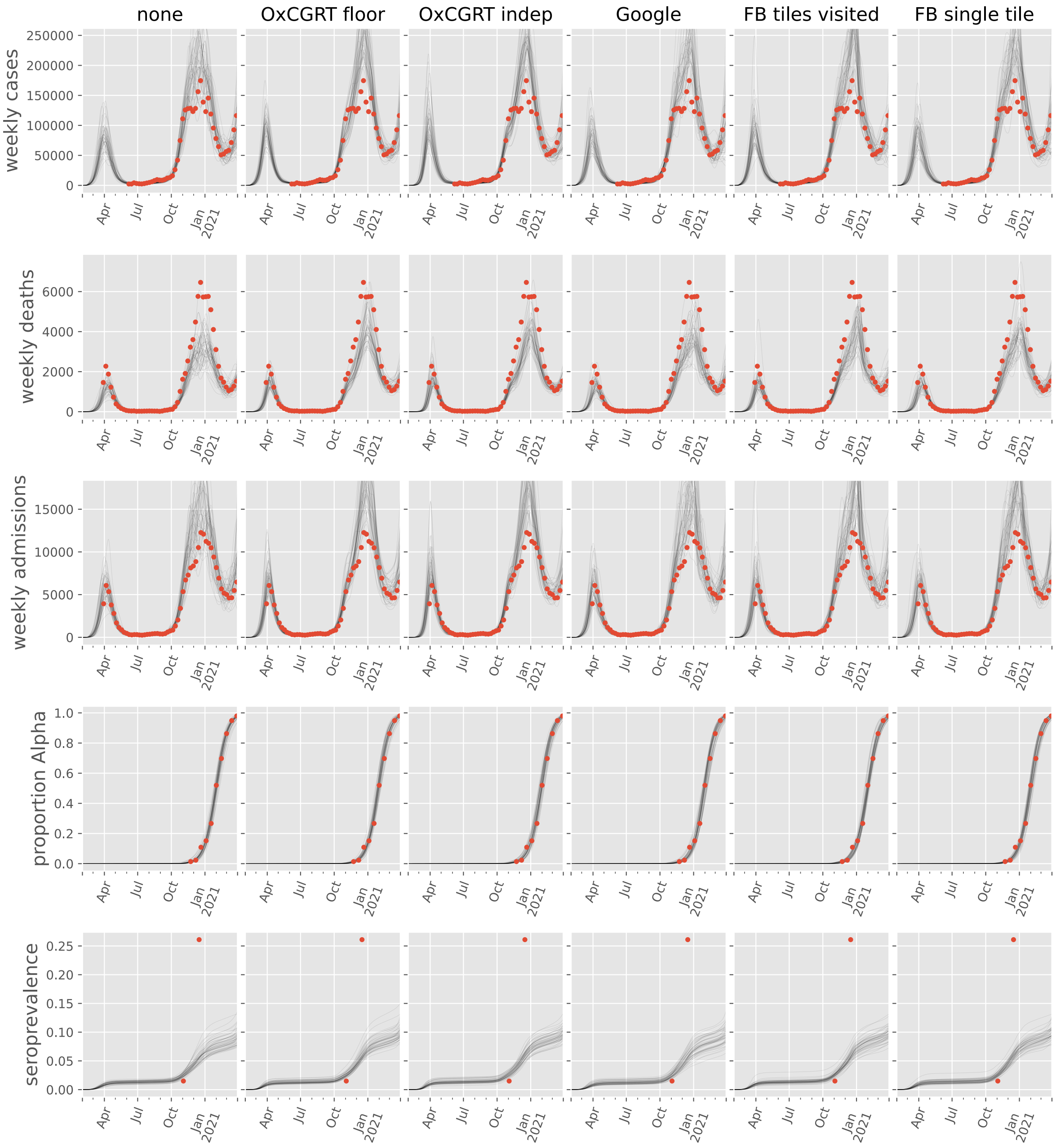
none
OxCGRT floor
OxCGRT indep
Google
FB tiles visited
FB single tile
weekly cases
weekly deaths
weekly admissions
proportion Alpha
seroprevalence
250000
200000
150000
100000
50000
0
6000
4000
2000
0
15000
10000
5000
0
1.0
0.8
0.6
0.4
0.2
0.0
0.25
0.20
0.15
0.10
0.05
0.00
Apr
Jul
Oct
Jan
2021

#### Greece

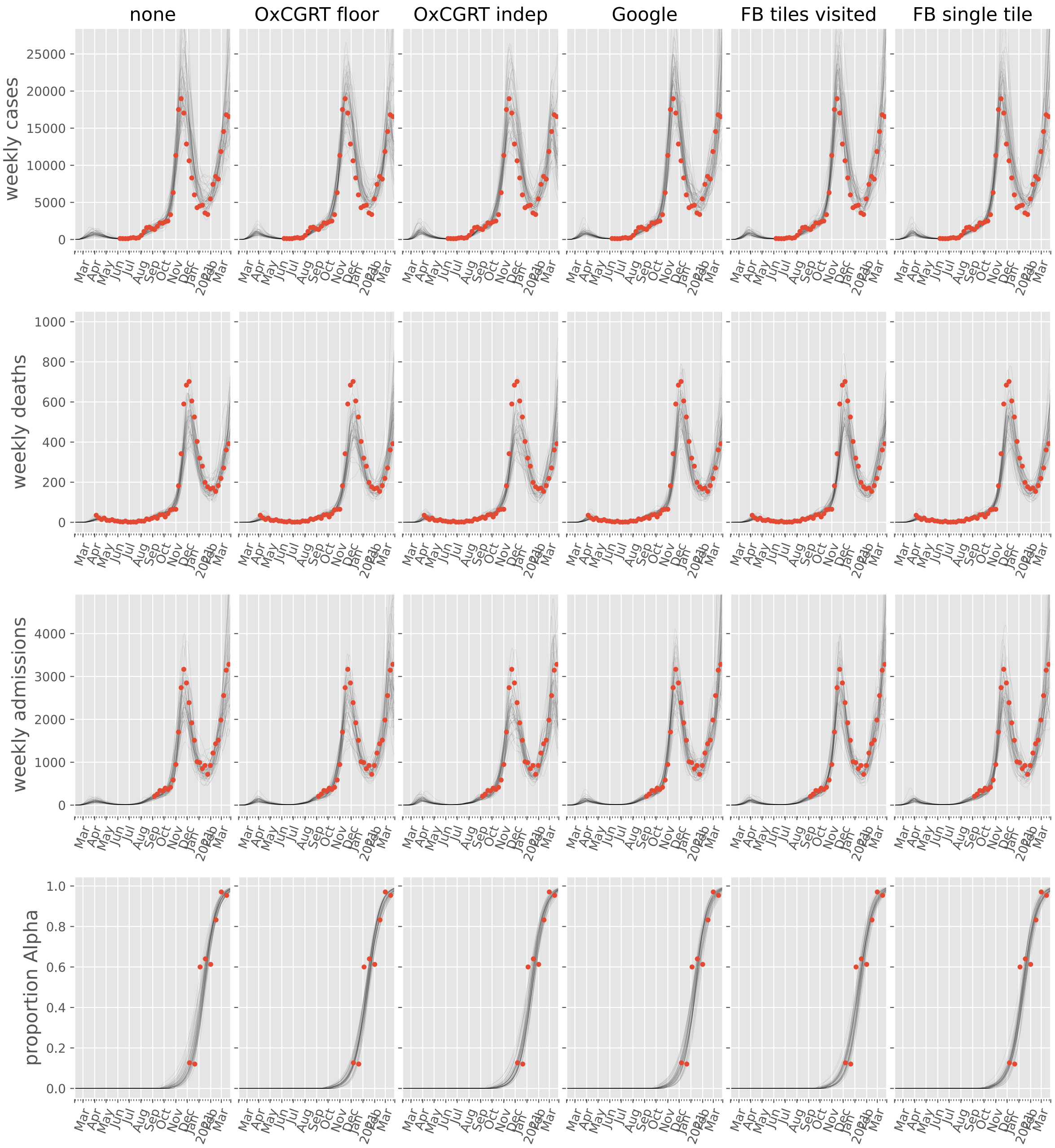

none
OxCGRT floor
OxCGRT indep
Google
FB tiles visited
FB single tile
weekly cases
weekly deaths
weekly admissions
proportion Alpha

#### Hungary

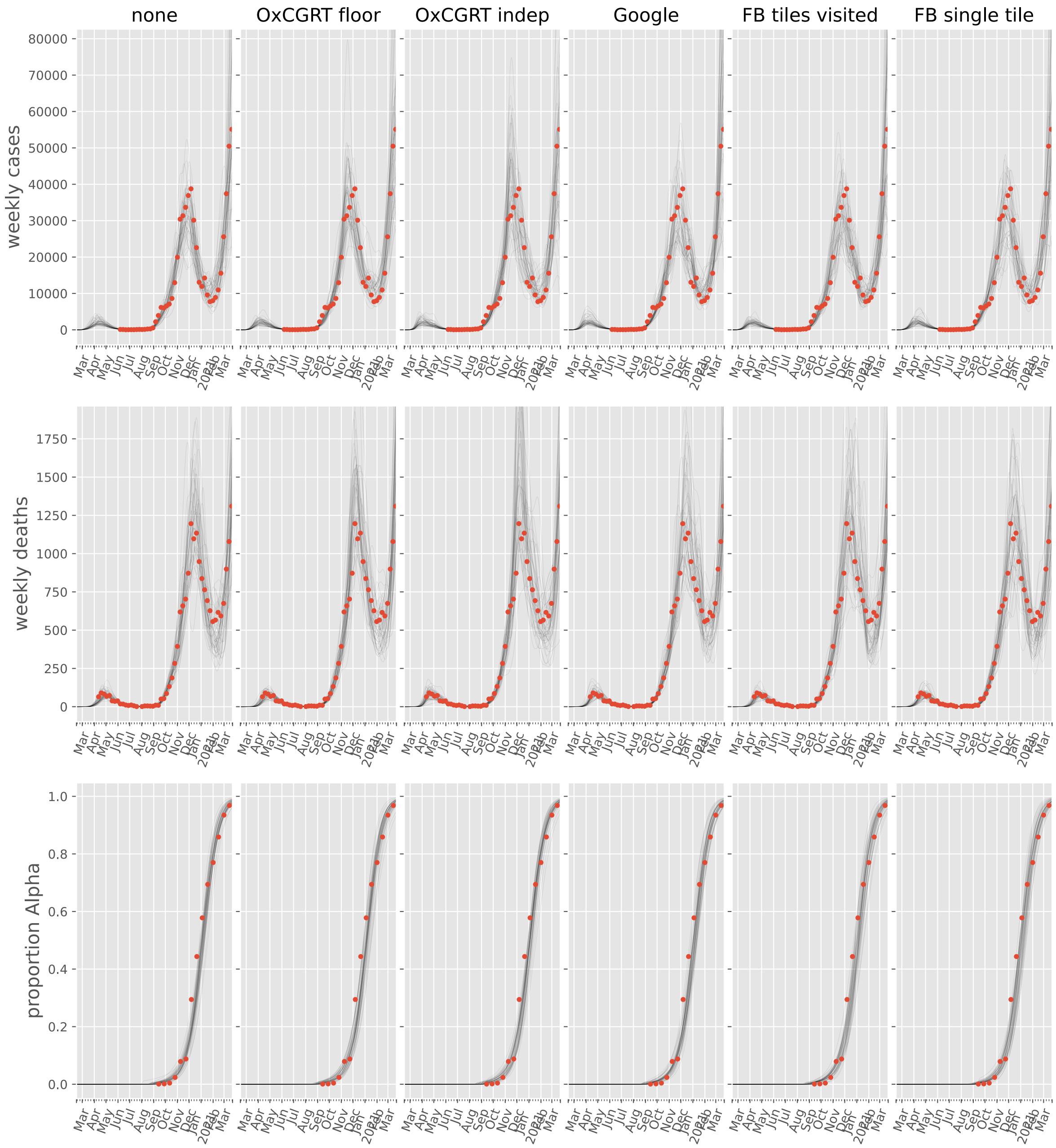
none
OxCGRT floor
OxCGRT indep
Google
FB tiles visited
FB single tile
weekly cases
weekly deaths
proportion Alpha

#### Ireland

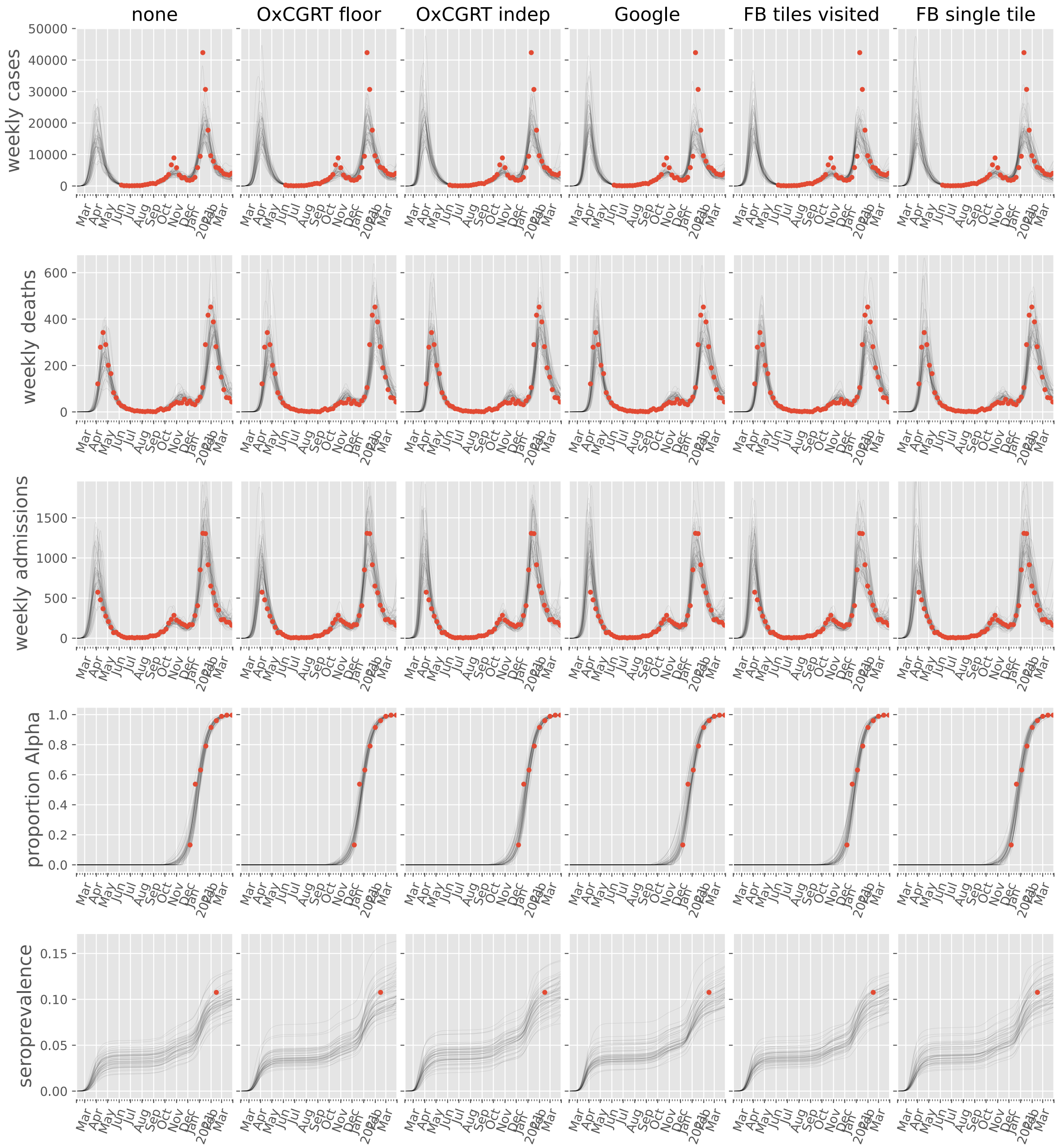

none
OxCGRT floor
OxCGRT indep
Google
FB tiles visited
FB single tile
weekly cases
weekly deaths
weekly admissions
proportion Alpha
seroprevalence

#### Italy

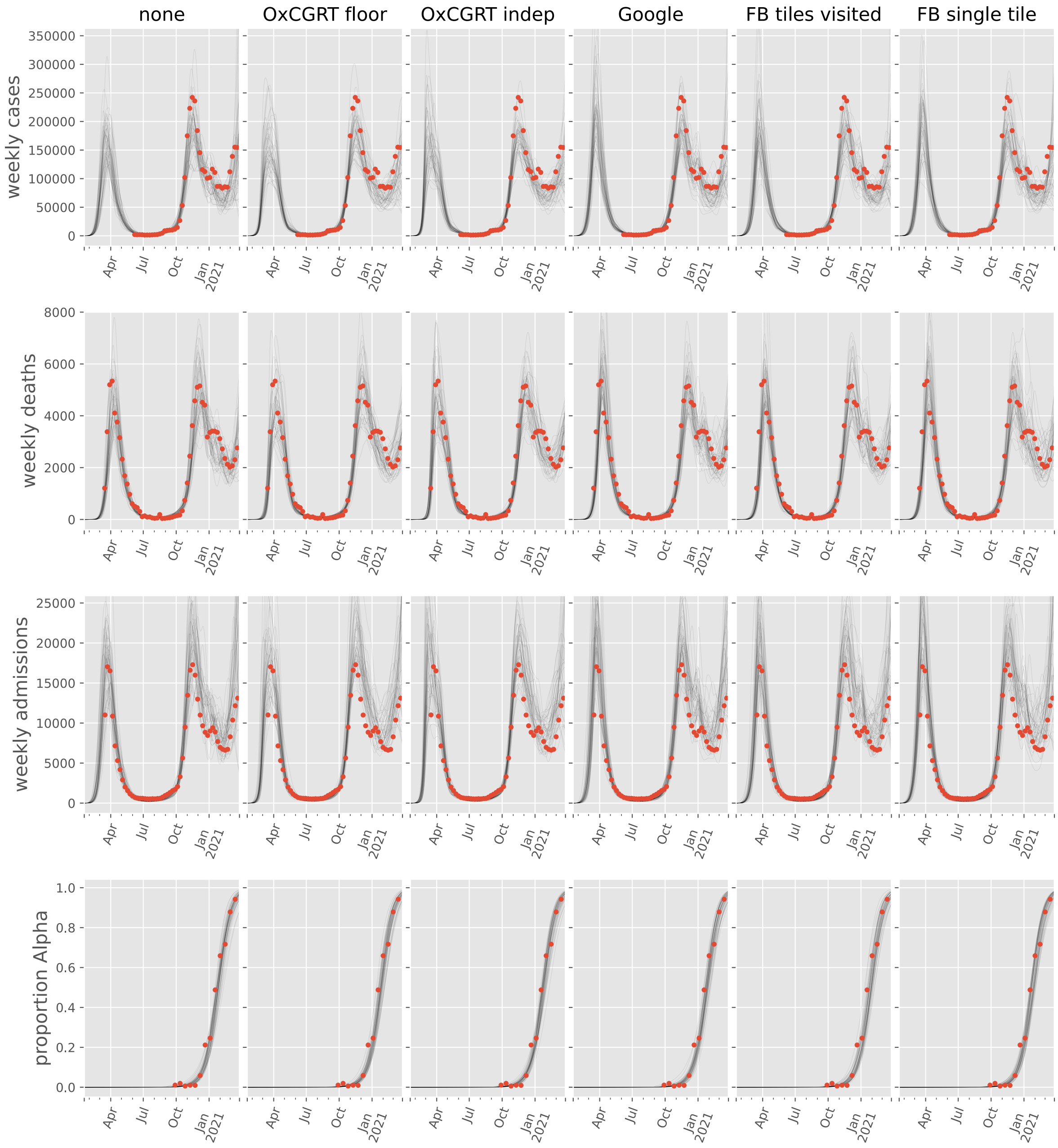

none
OxCGRT floor
OxCGRT indep
Google
FB tiles visited
FB single tile
weekly cases
weekly deaths
weekly admissions
proportion Alpha
Apr
Jul
Oct
Jan 2021

#### Kosovo

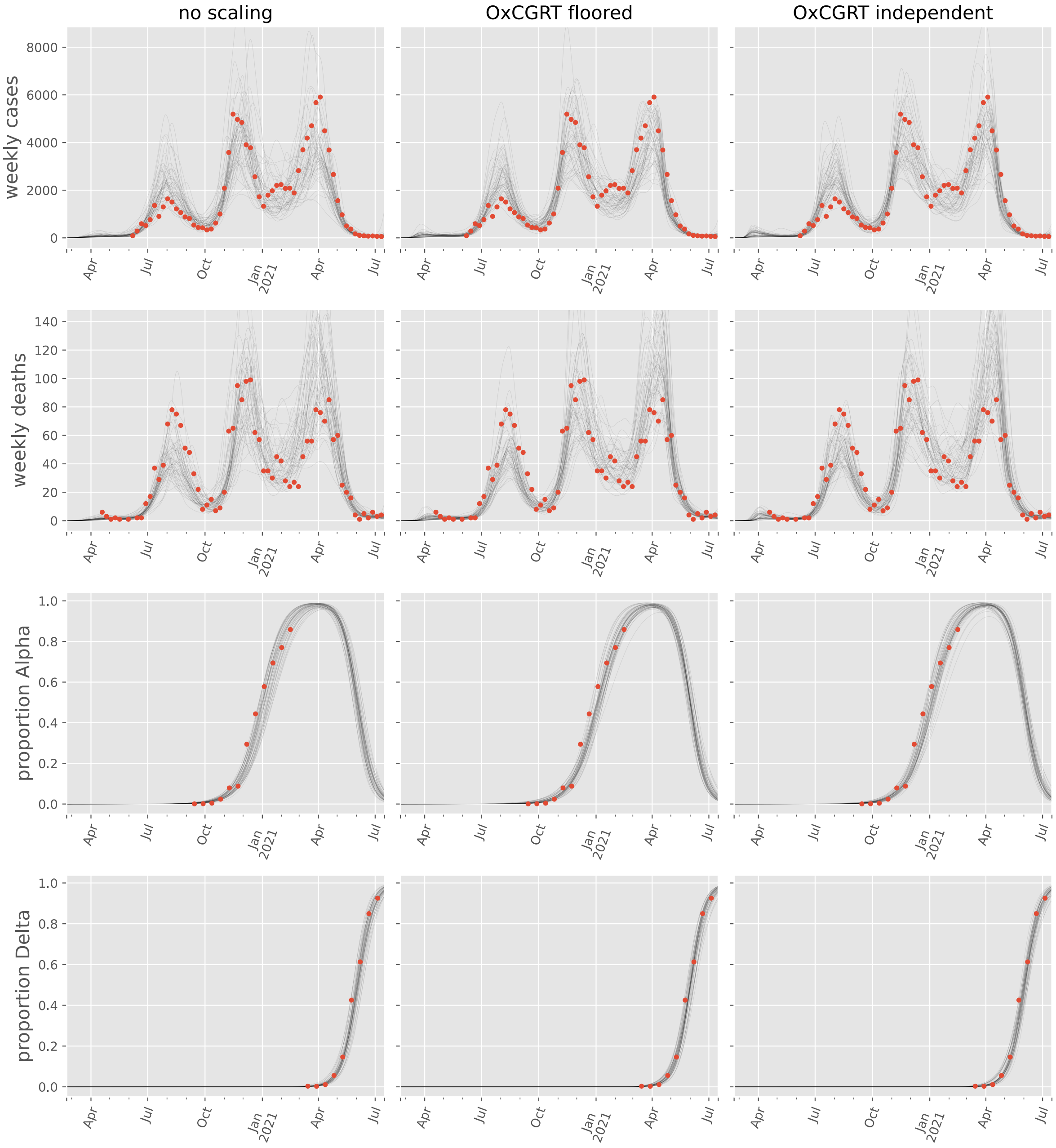

no scaling
OxCGRT floored
OxCGRT independent
weekly cases
weekly deaths
proportion Alpha
proportion Delta
0
2000
4000
6000
8000
0
20
40
60
80
100
120
140
0.0
0.2
0.4
0.6
0.8
1.0
Apr
Jul
Oct
Jan
2021

#### Latvia

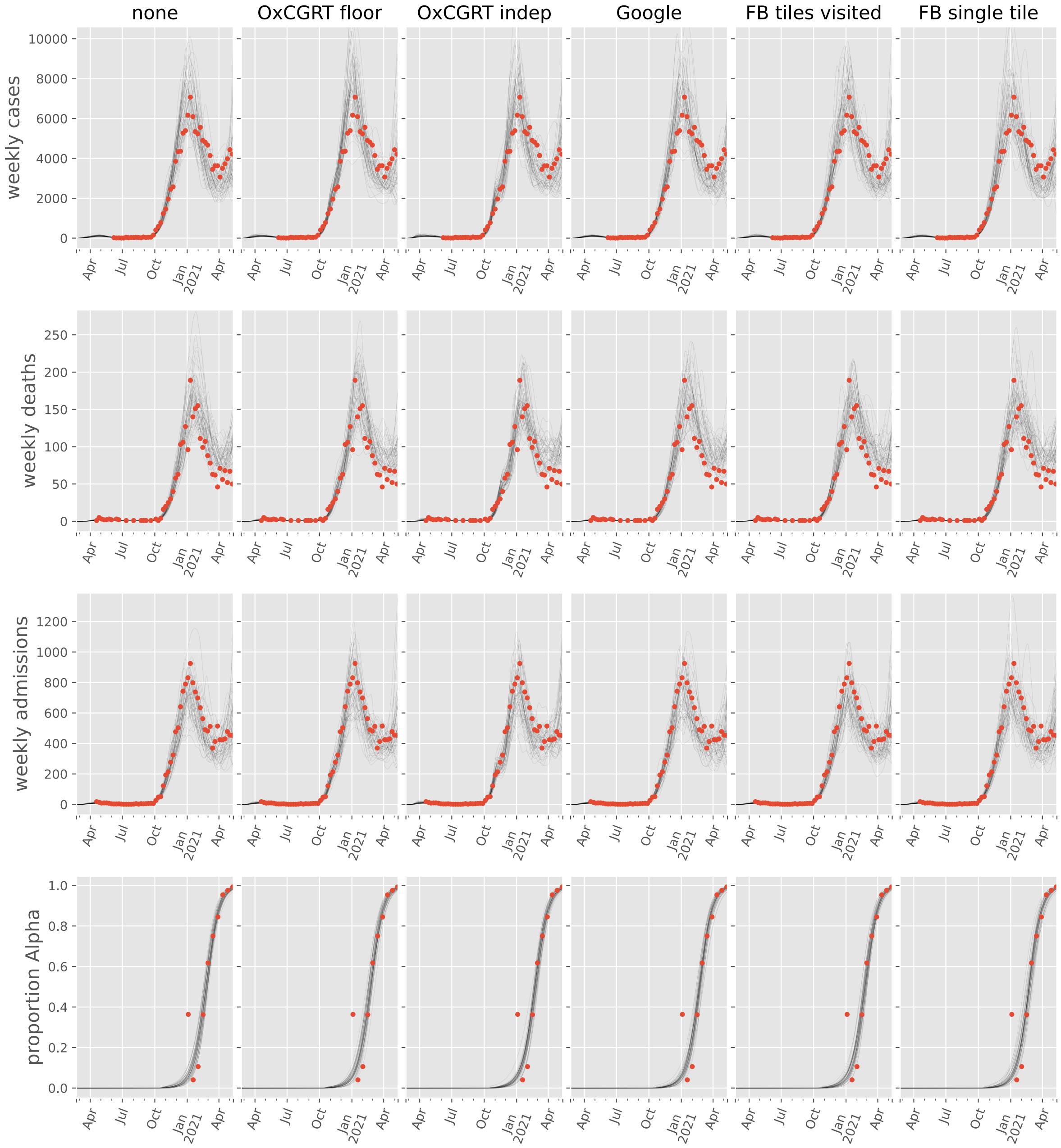

none
OxCGRT floor
OxCGRT indep
Google
FB tiles visited
FB single tile
weekly cases
weekly deaths
weekly admissions
proportion Alpha
Apr
Jul
Oct
Jan
2021

#### Lithuania

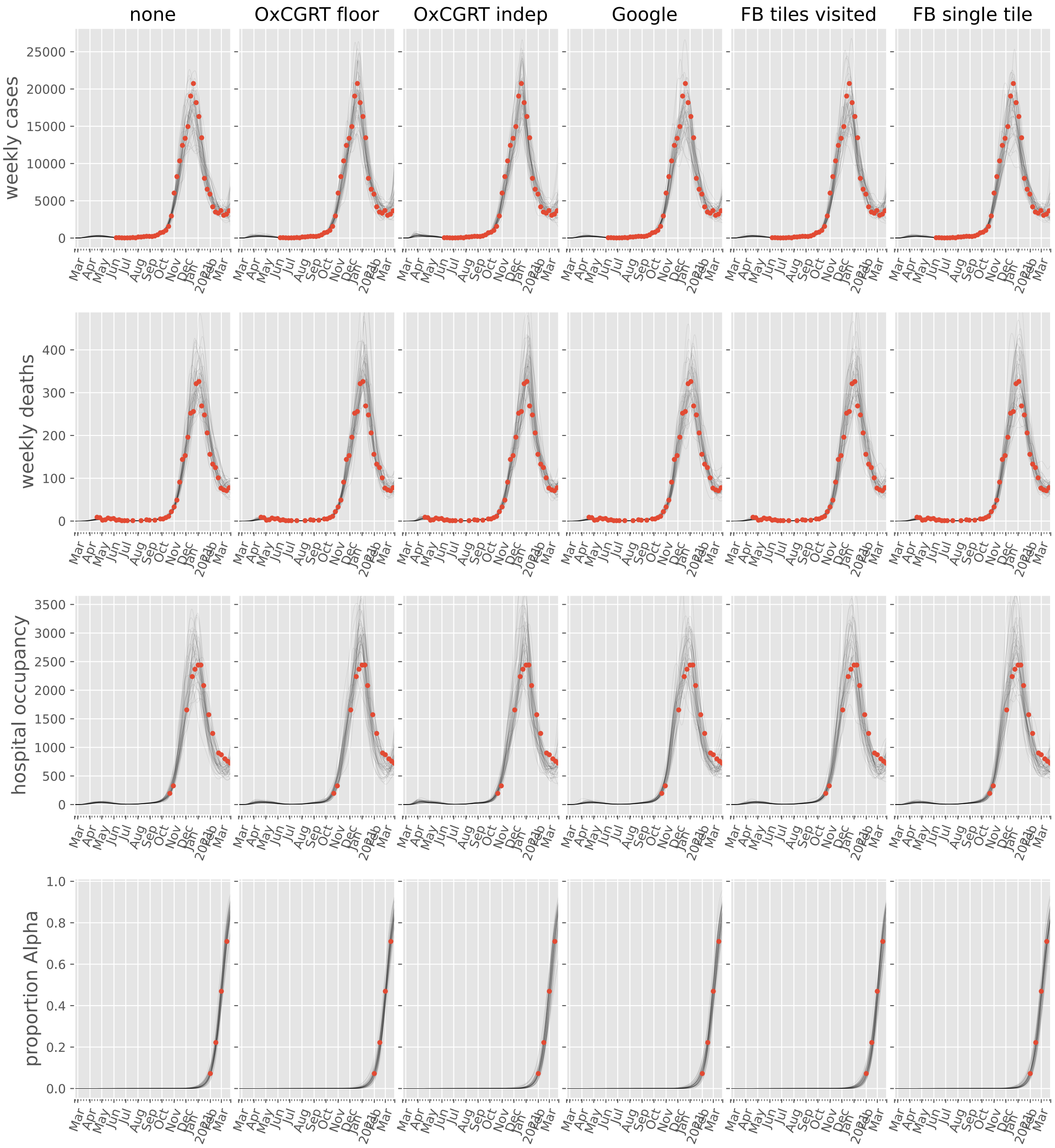
none
OxCGRT floor
OxCGRT indep
Google
FB tiles visited
FB single tile
weekly cases
weekly deaths
hospital occupancy
proportion Alpha

#### Luxembourg

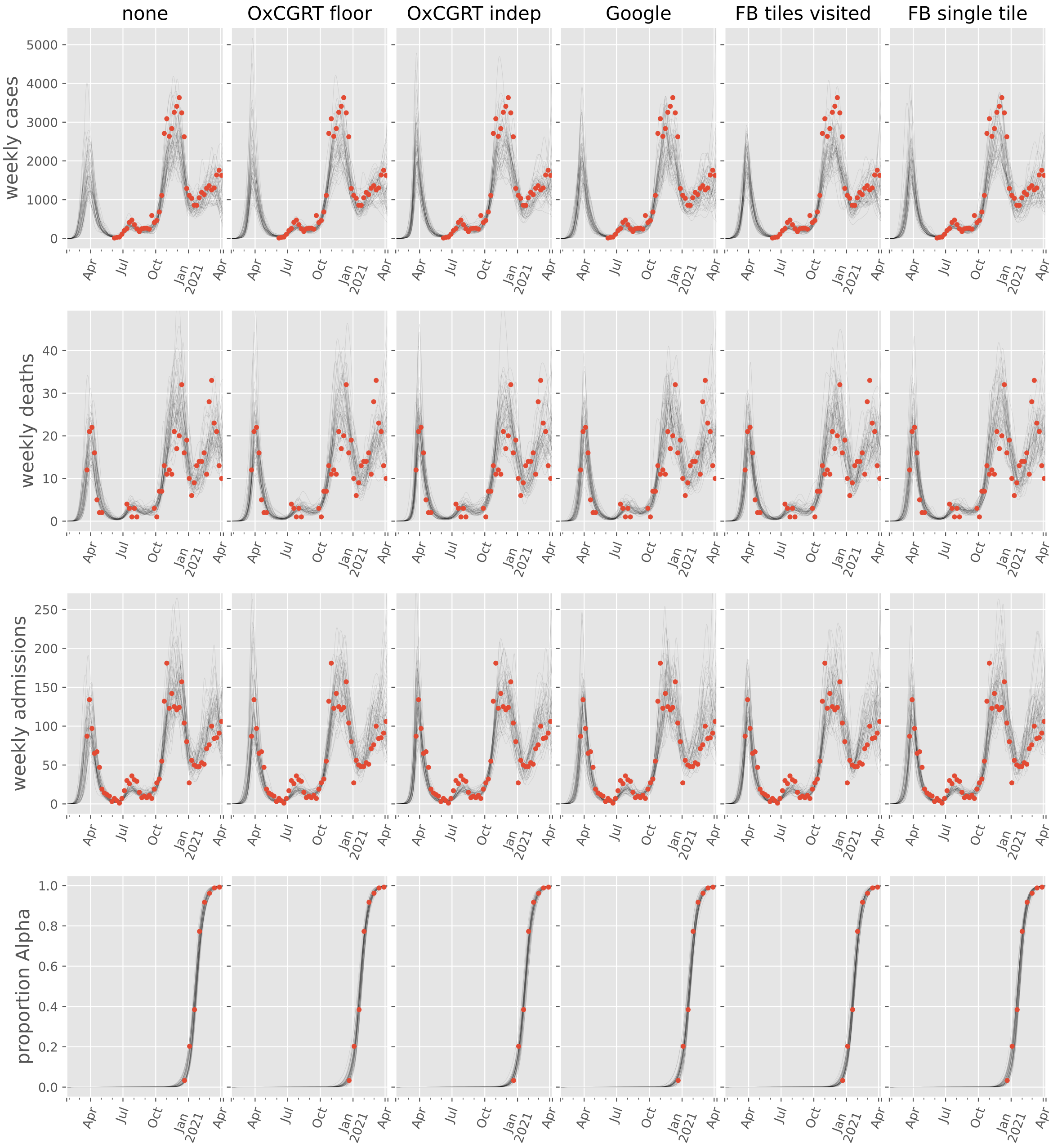
none
OxCGRT floor
OxCGRT indep
Google
FB tiles visited
FB single tile
weekly cases
weekly deaths
weekly admissions
proportion Alpha
Apr
Jul
Oct
Jan 2021
Apr

#### Malta

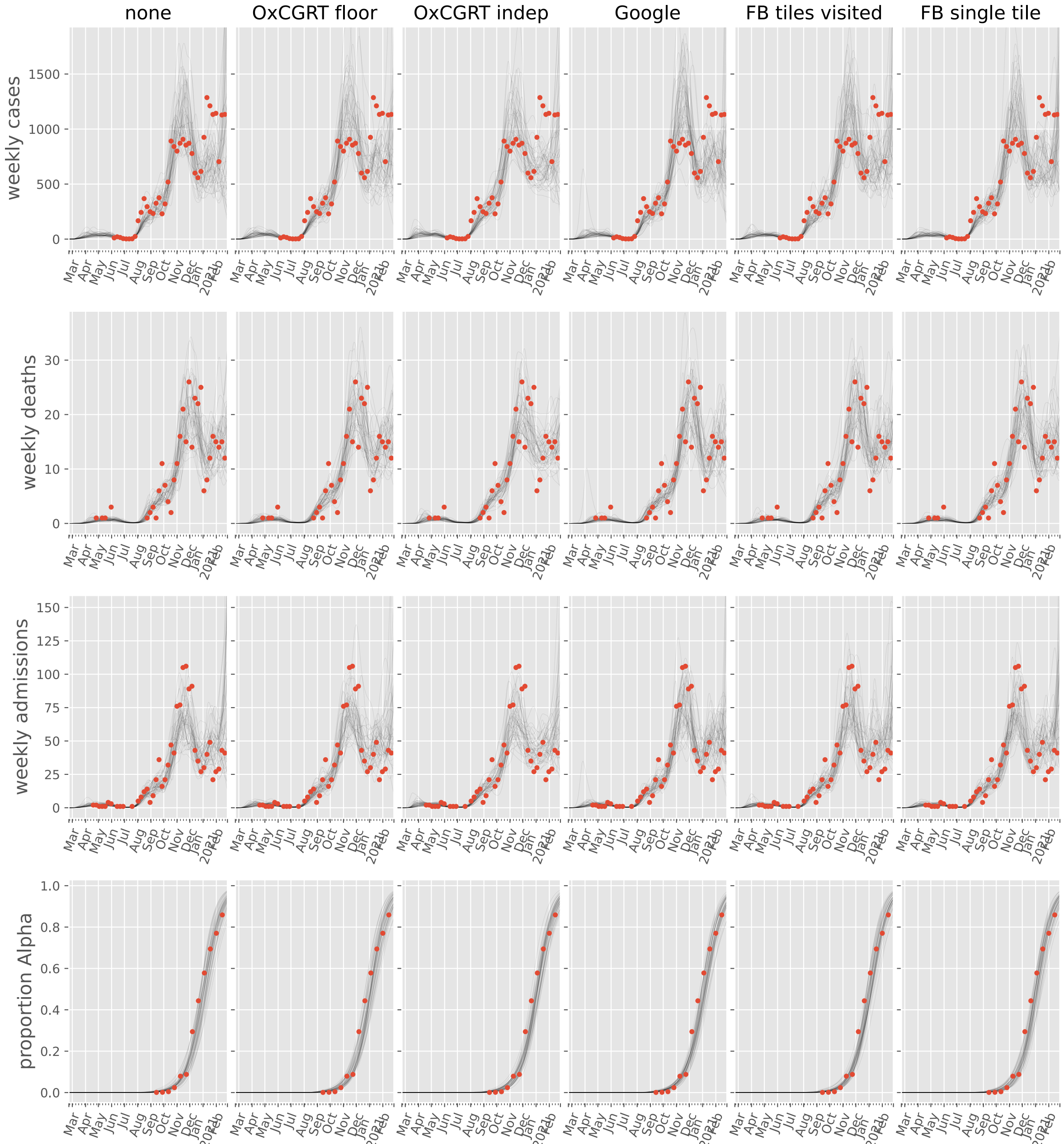

none
OxCGRT floor
OxCGRT indep
Google
FB tiles visited
FB single tile
weekly cases
weekly deaths
weekly admissions
proportion Alpha

#### Moldova, Republic of

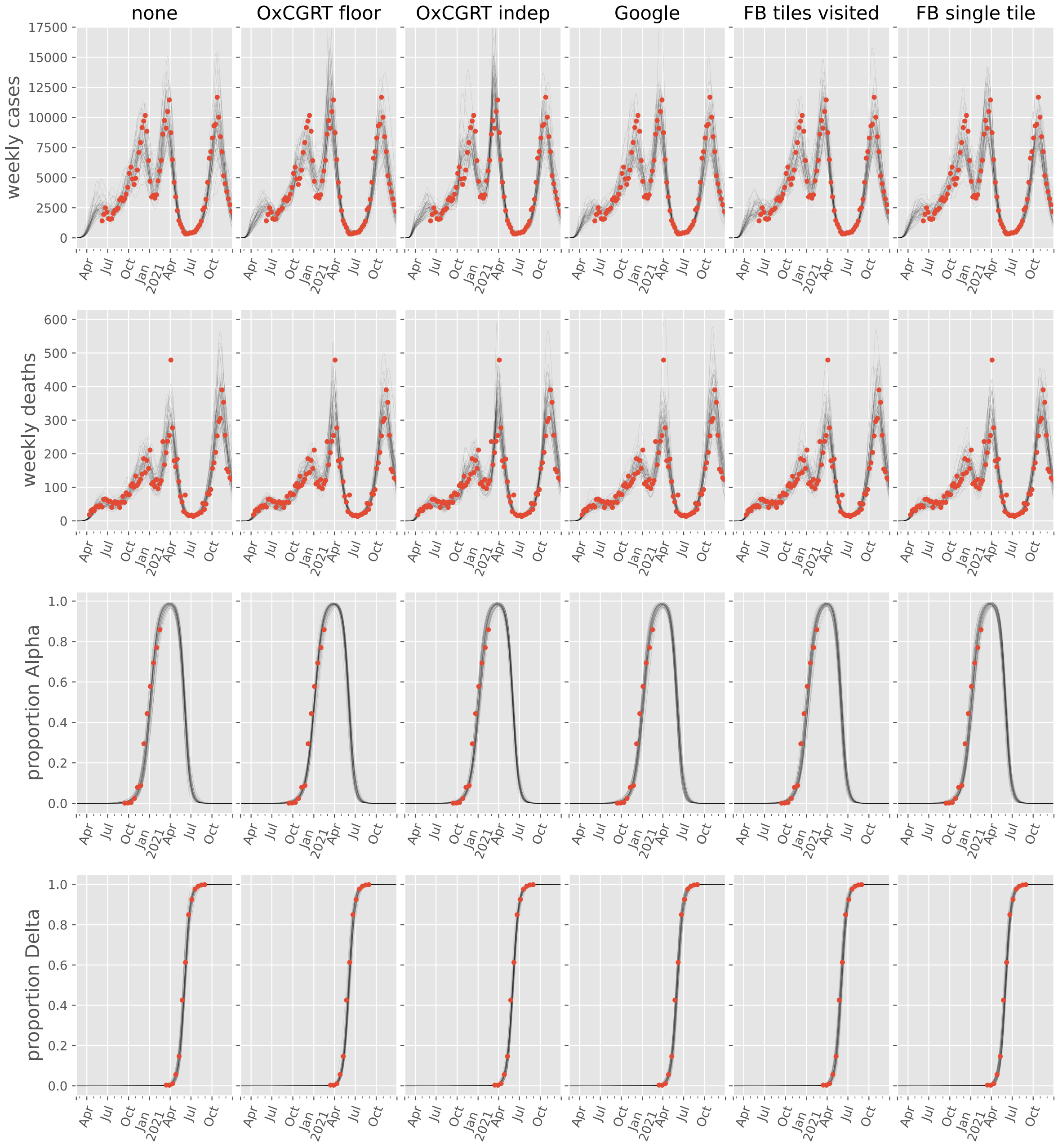

#### Netherlands

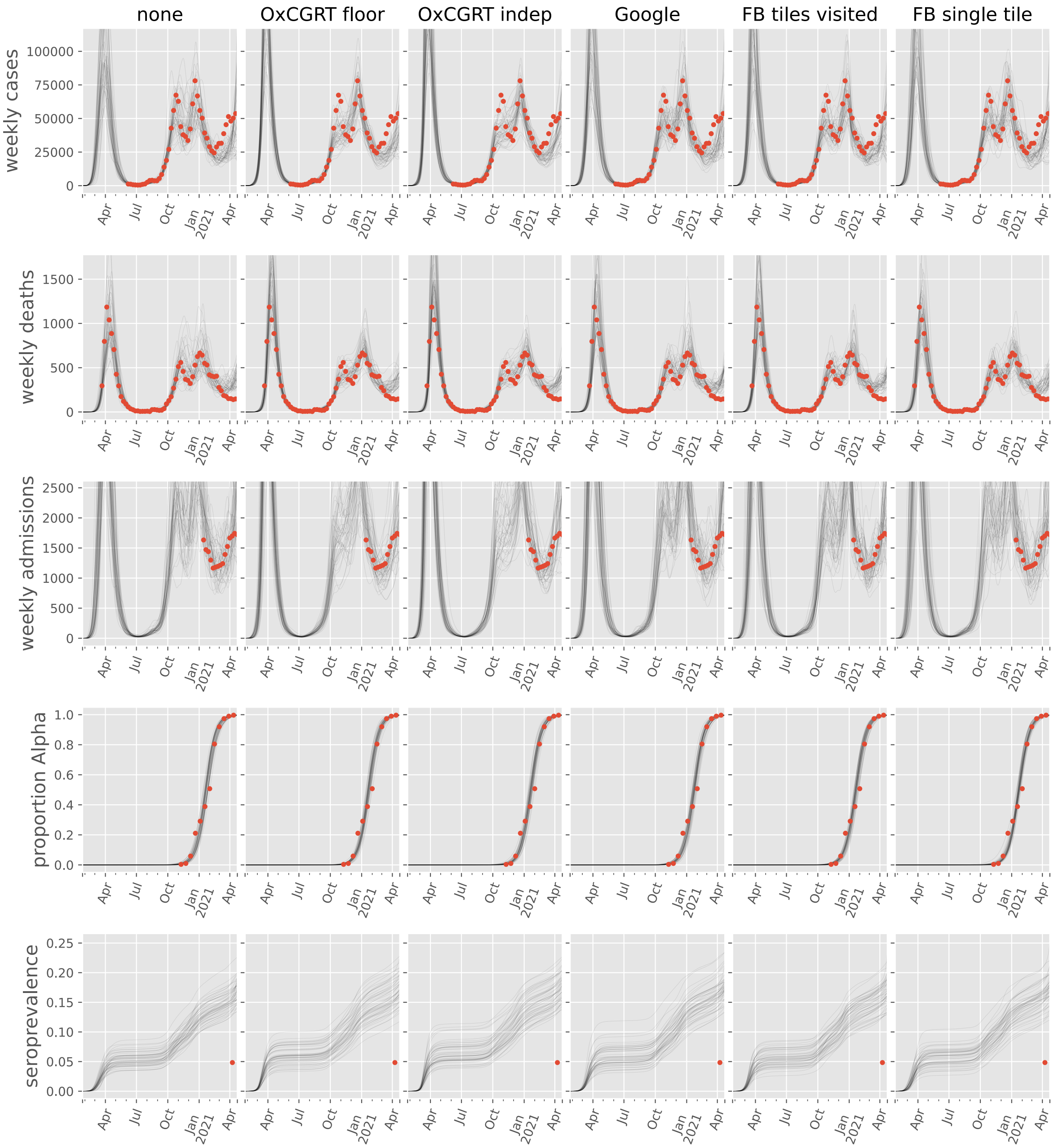

#### Norway

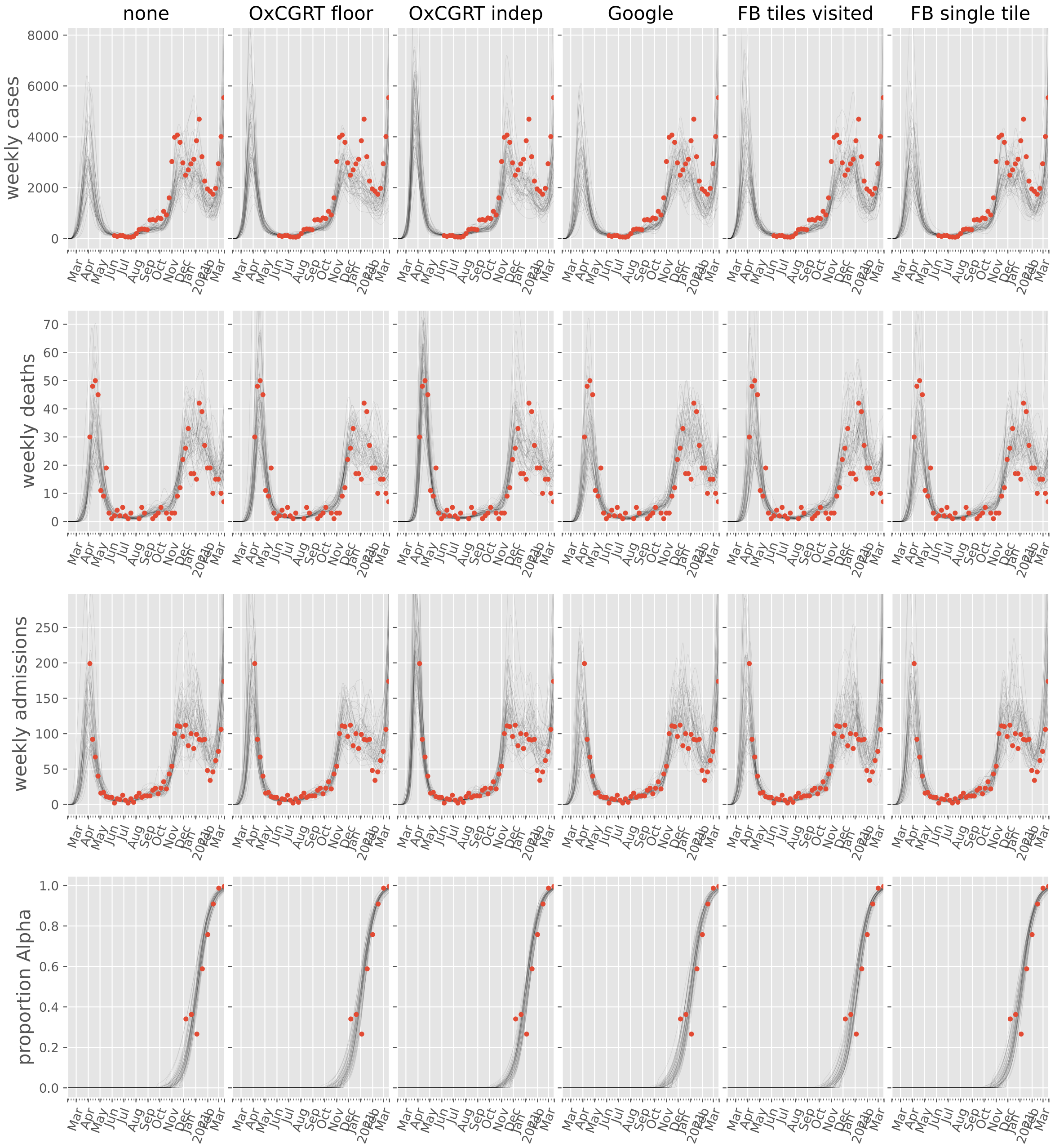
none
OxCGRT floor
OxCGRT indep
Google
FB tiles visited
FB single tile
weekly cases
weekly deaths
weekly admissions
proportion Alpha
Mar
Apr
May
Jun
Jul
Aug
Sep
Oct
Nov
Dec
Jan
2021
Feb
Mar

#### Poland

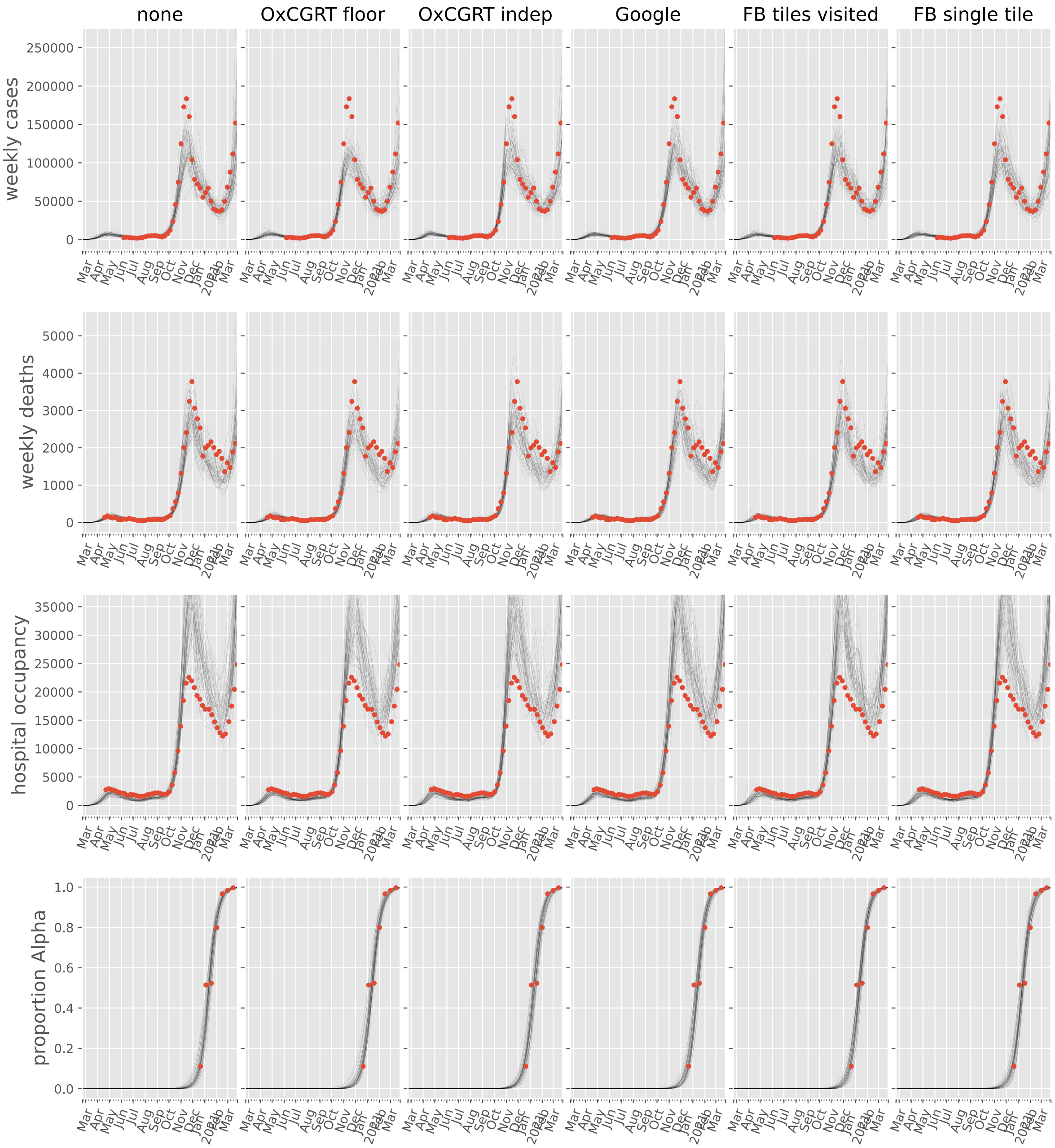
none
OxCGRT floor
OxCGRT indep
Google
FB tiles visited
FB single tile
weekly cases
weekly deaths
hospital occupancy
proportion Alpha

#### Portugal

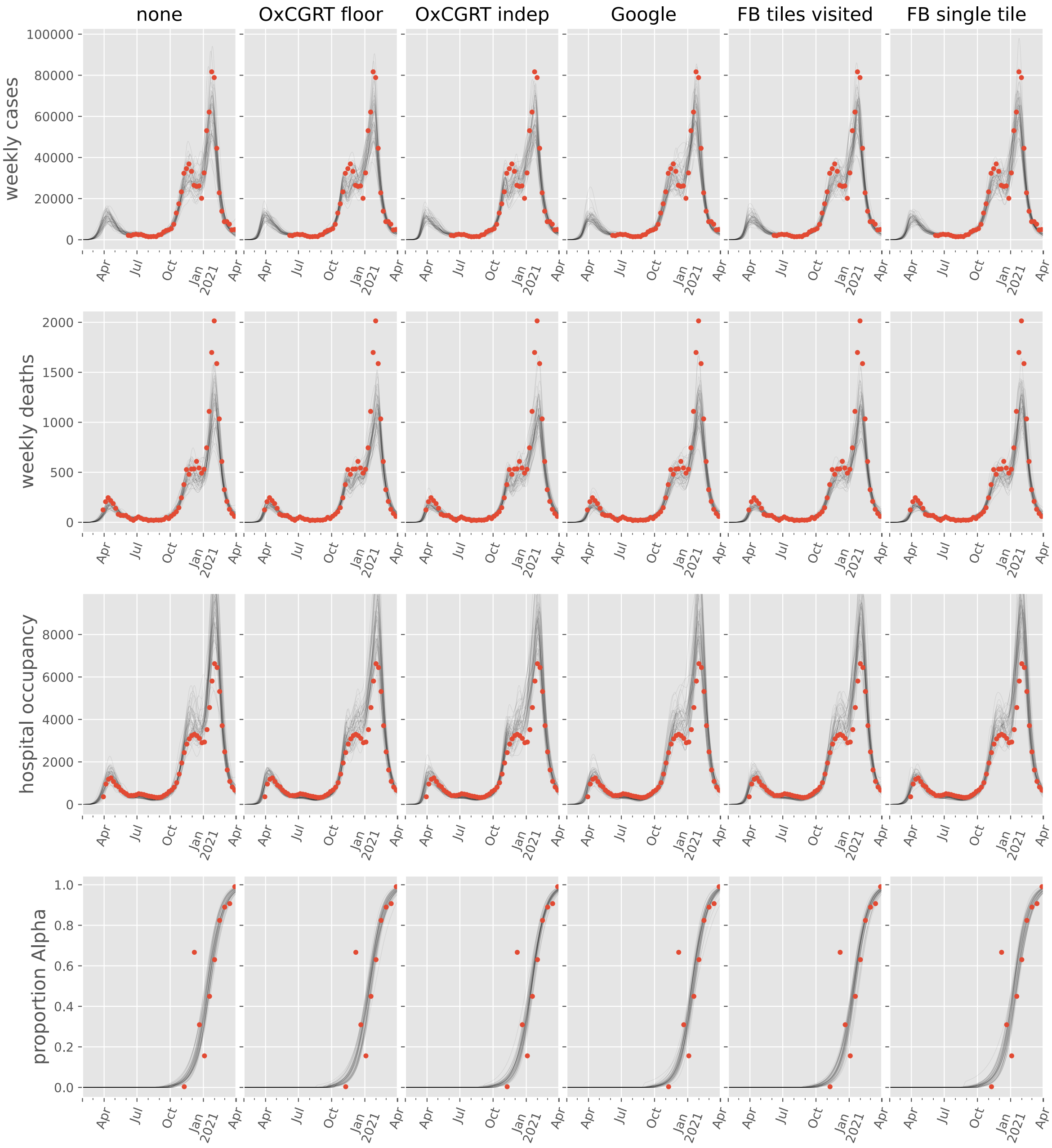
none
OxCGRT floor
OxCGRT indep
Google
FB tiles visited
FB single tile
weekly cases
weekly deaths
hospital occupancy
proportion Alpha
100000
80000
60000
40000
20000
0
2000
1500
1000
500
8000
6000
4000
2000
1.0
0.8
0.6
0.4
0.2
0.0
Apr
Jul
Oct
Jan
2021

#### Romania

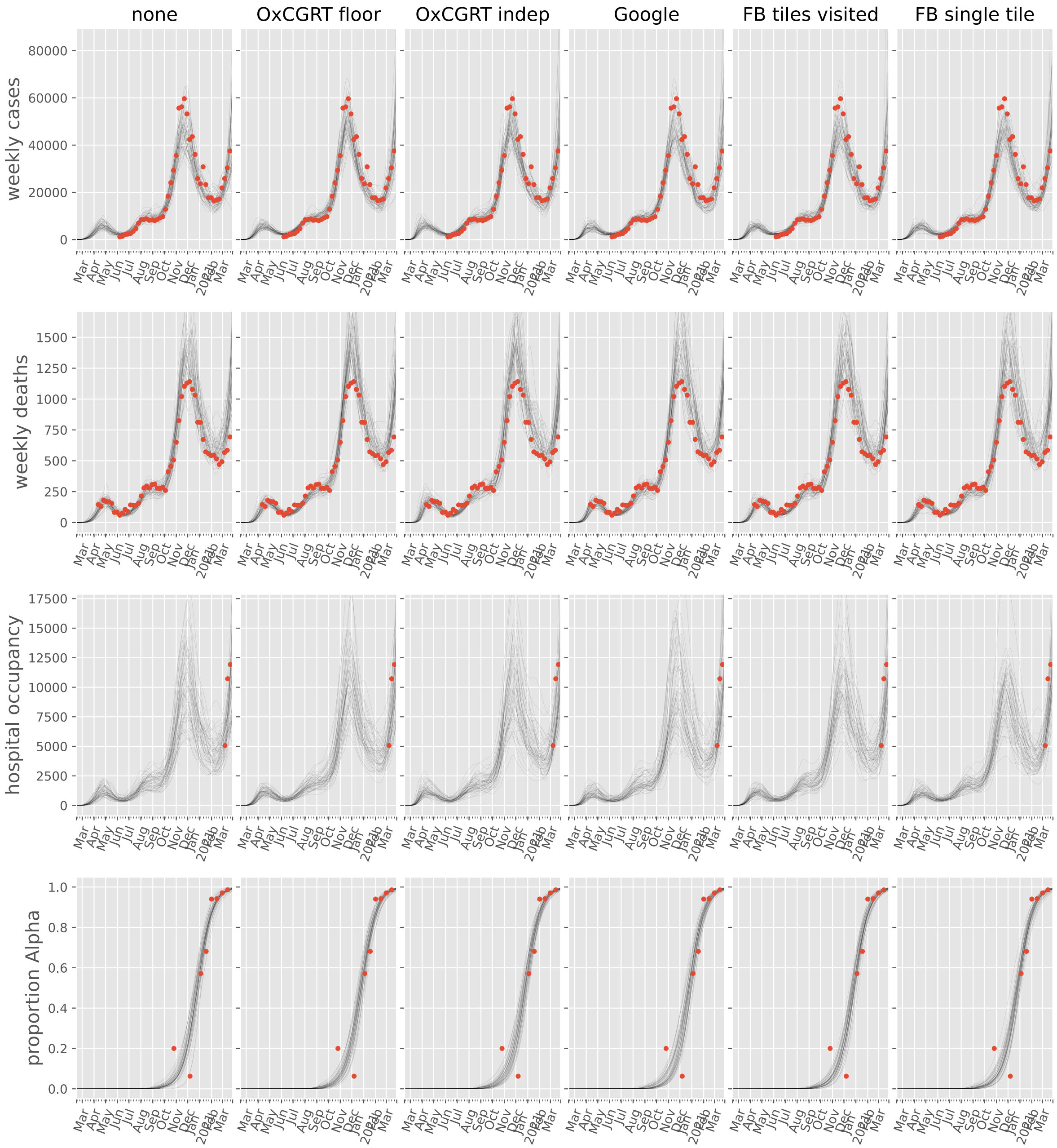

none
OxCGRT floor
OxCGRT indep
Google
FB tiles visited
FB single tile
weekly cases
weekly deaths
hospital occupancy
proportion Alpha

#### Russian Federation

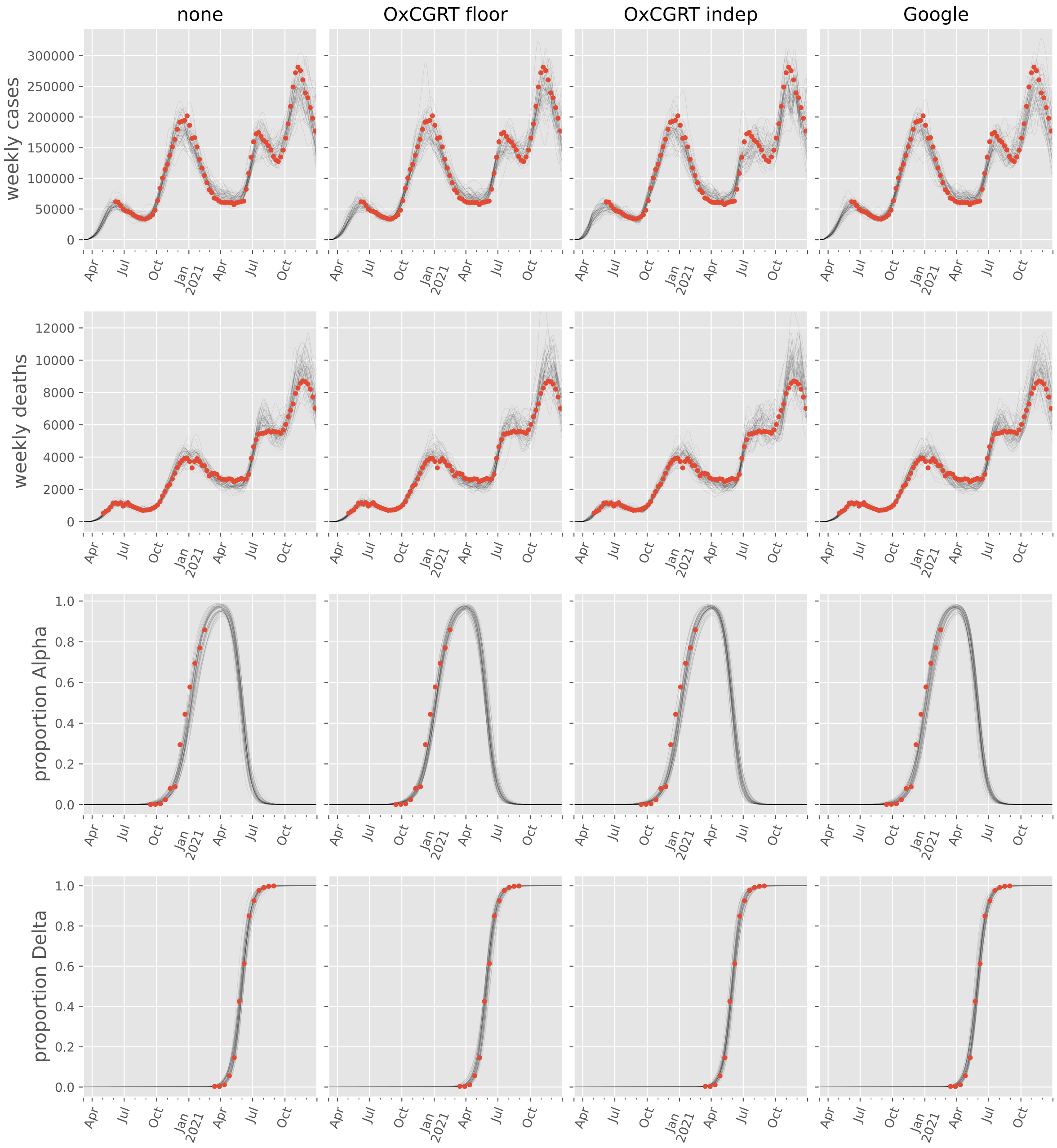

#### Serbia

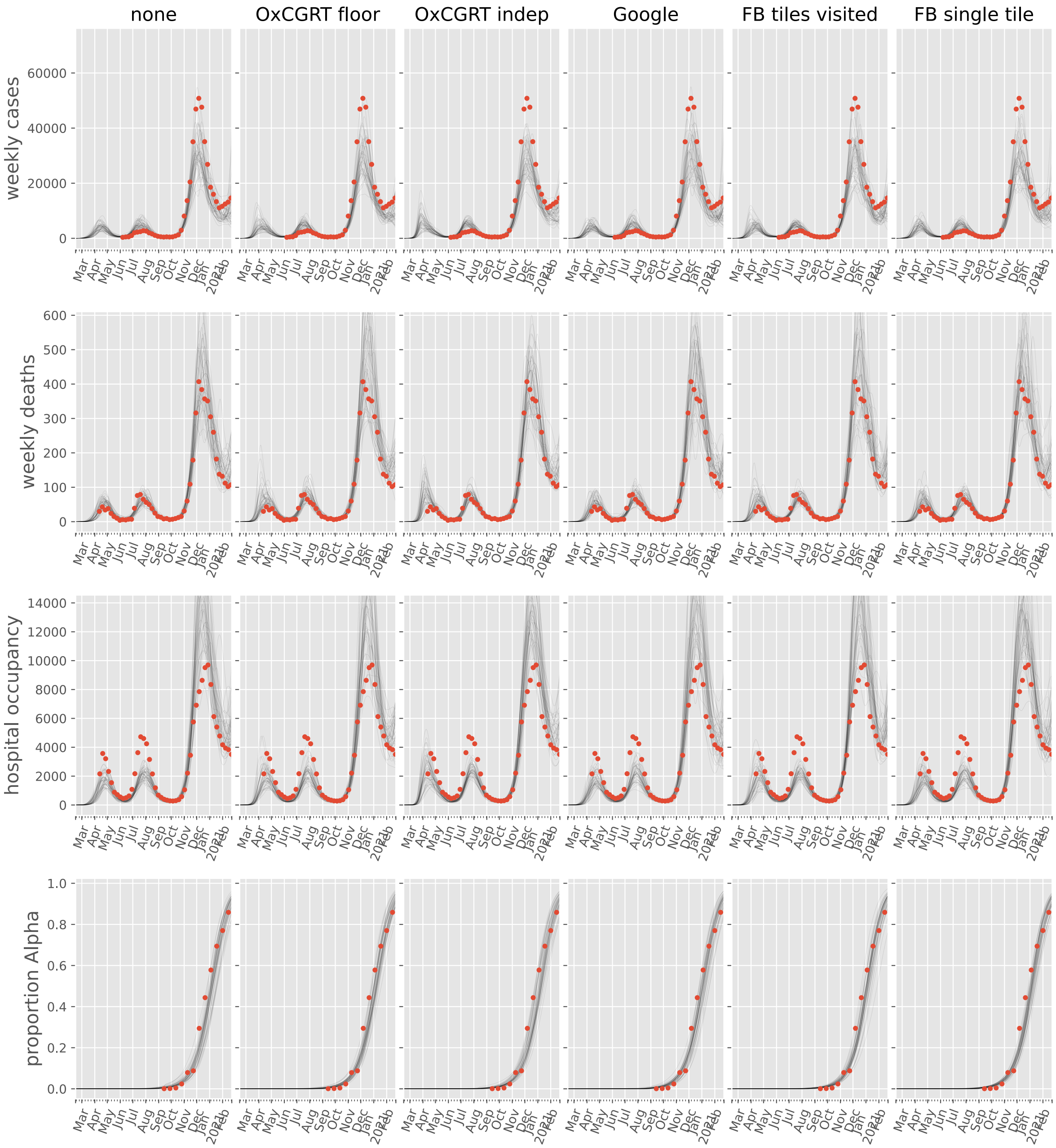

none
OxCGRT floor
OxCGRT indep
Google
FB tiles visited
FB single tile
weekly cases
weekly deaths
hospital occupancy
proportion Alpha

#### Slovakia

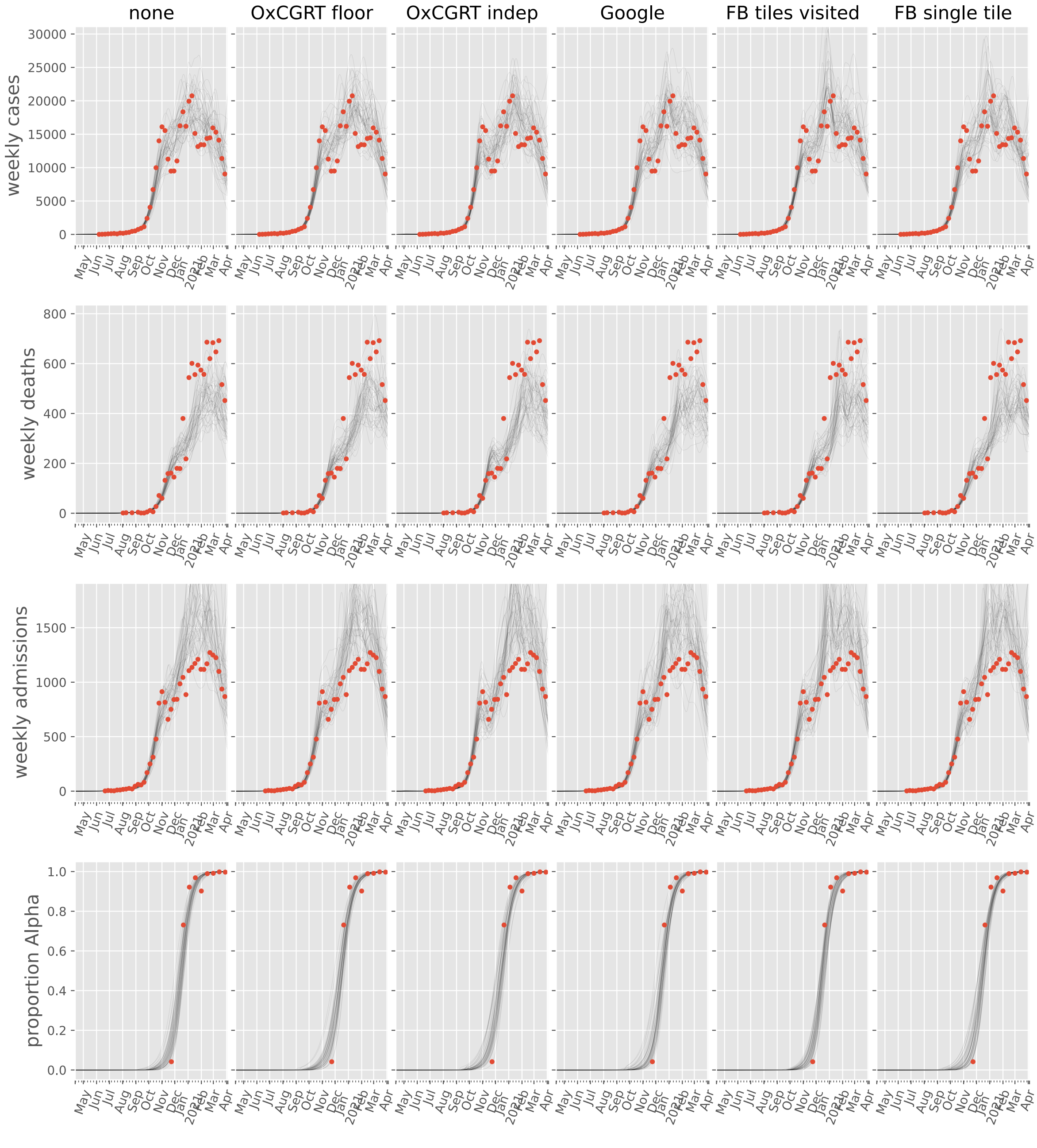

none
OxCGRT floor
OxCGRT indep
Google
FB tiles visited
FB single tile
weekly cases
weekly deaths
weekly admissions
proportion Alpha
May
Jun
Jul
Aug
Sep
Oct
Nov
Dec
Jan
2021
Feb
Mar
Apr

#### Slovenia

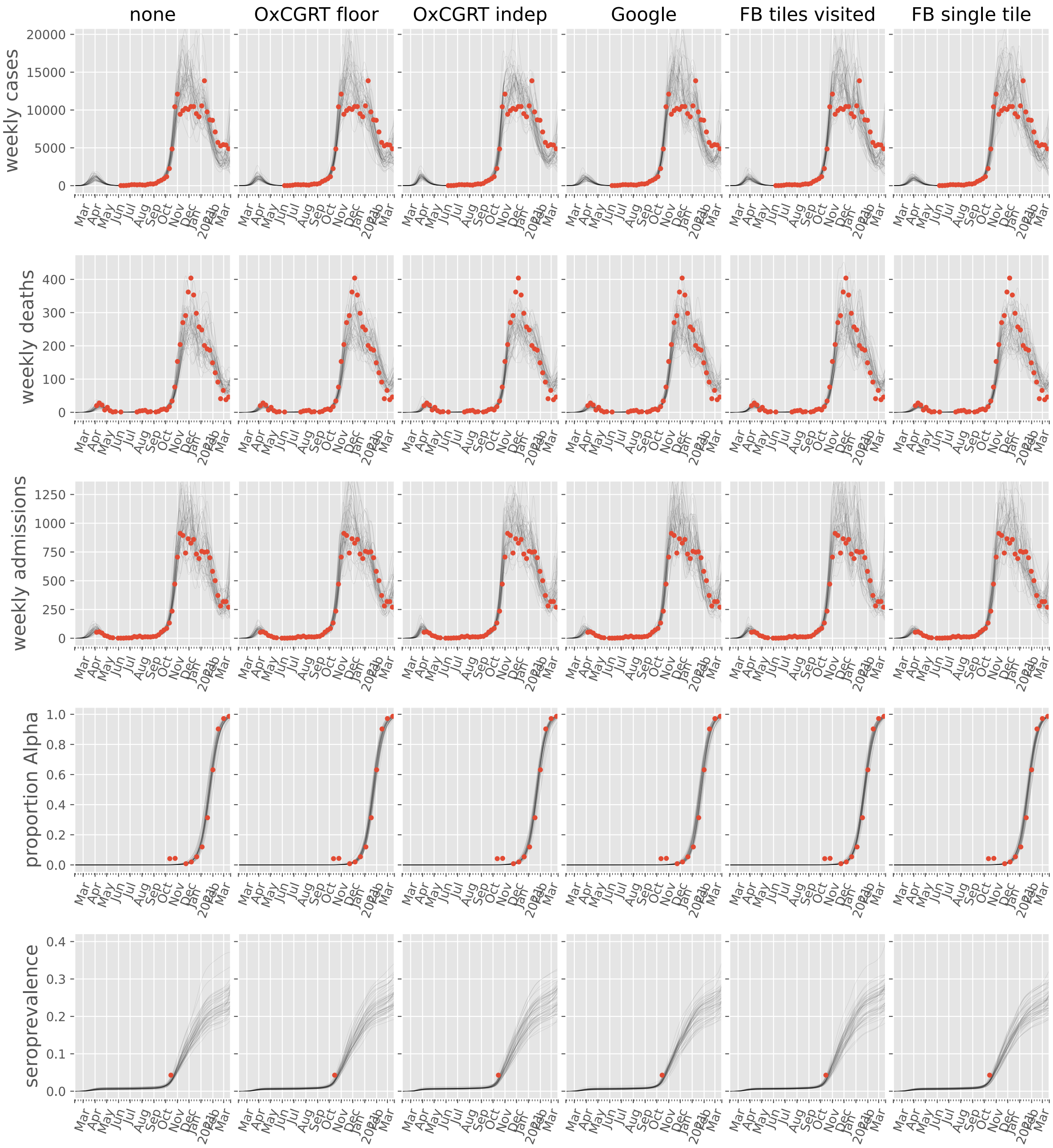
none
OxCGRT floor
OxCGRT indep
Google
FB tiles visited
FB single tile
weekly cases
weekly deaths
weekly admissions
proportion Alpha
seroprevalence
Mar Apr May Jun Jul Aug Sep Oct Nov Dec Jan Feb Mar
2021

#### Spain

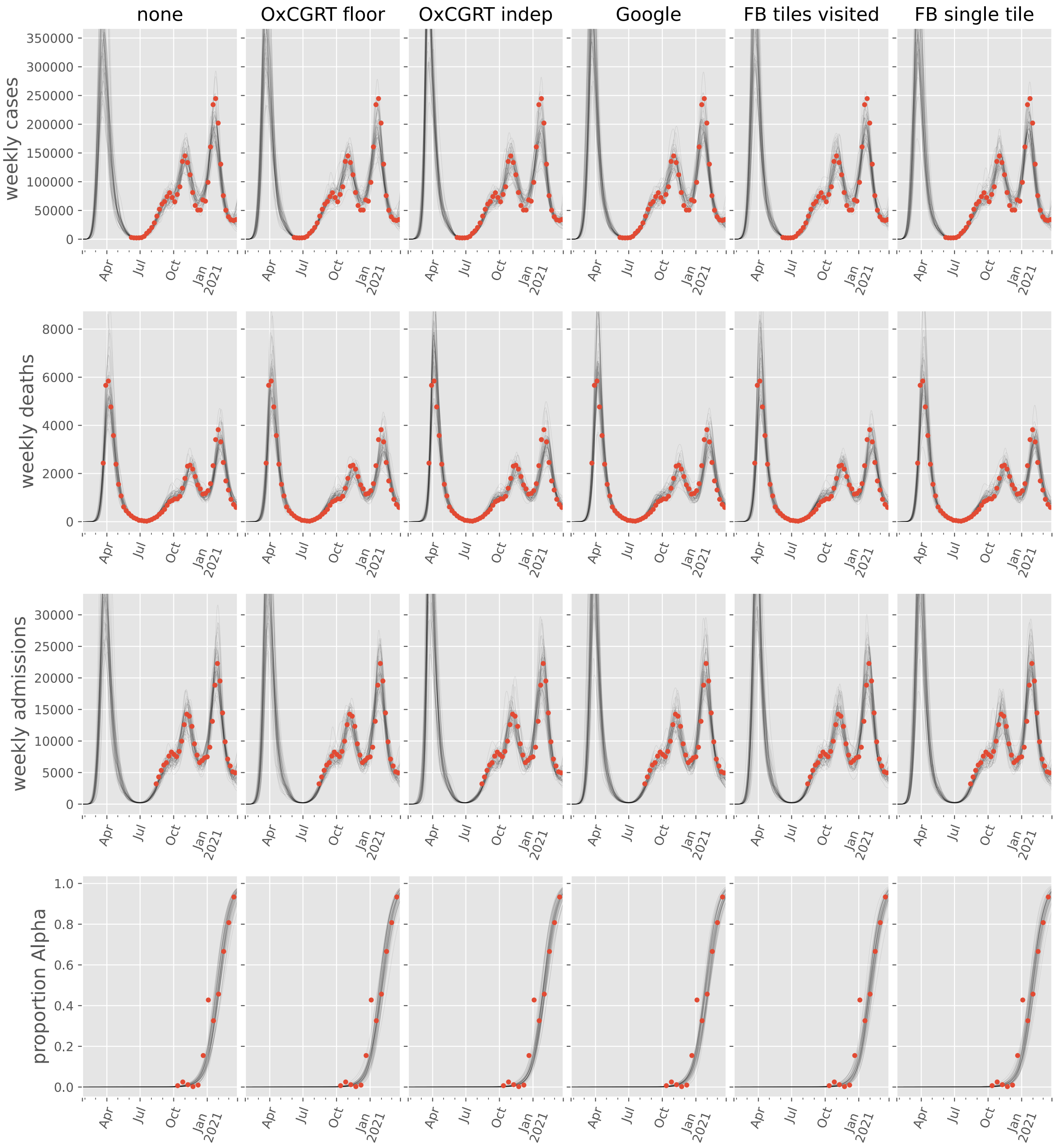

none
OxCGRT floor
OxCGRT indep
Google
FB tiles visited
FB single tile
weekly cases
weekly deaths
weekly admissions
proportion Alpha
Apr
Jul
Oct
Jan
2021

#### Sweden

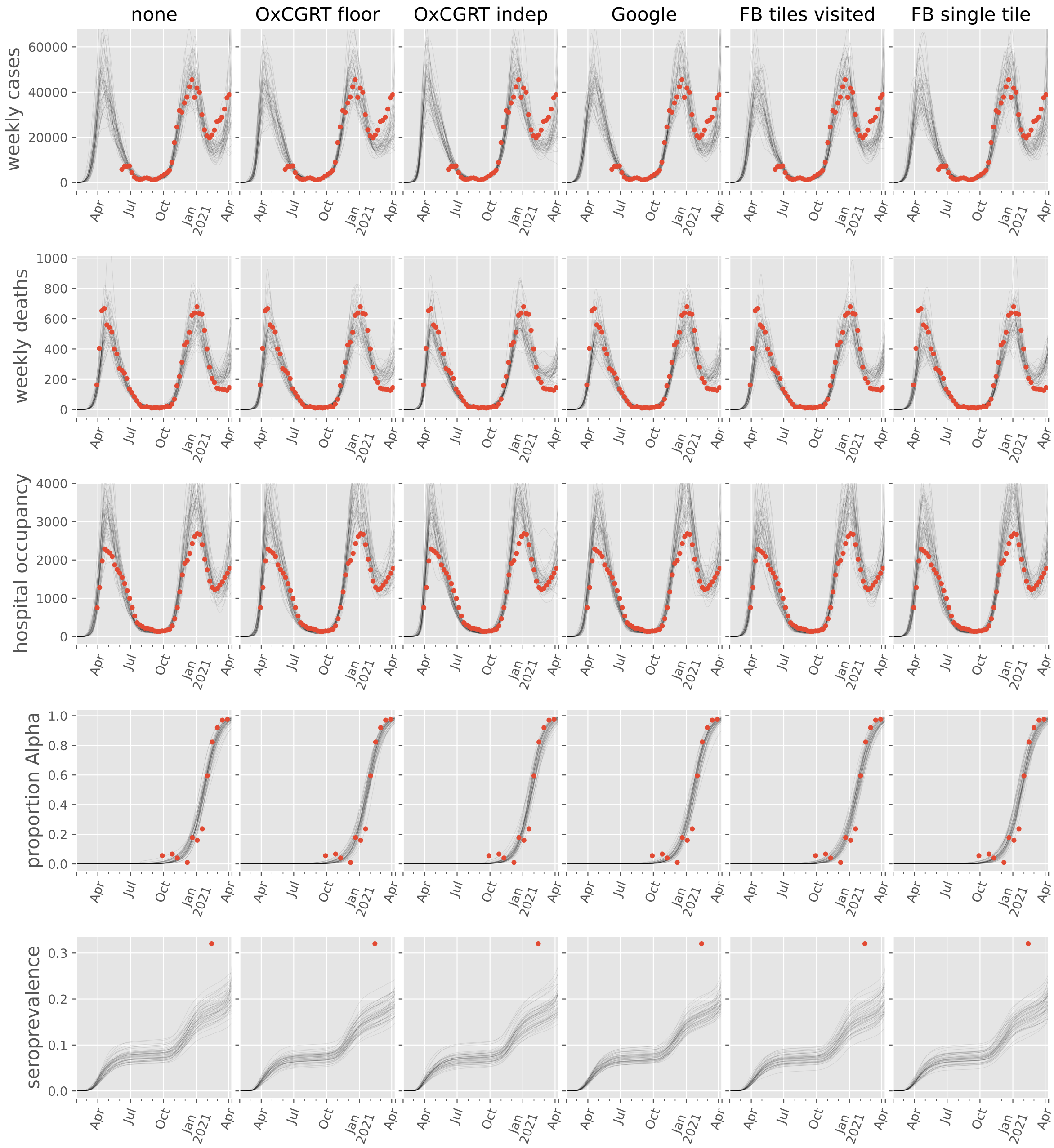

none
OxCGRT floor
OxCGRT indep
Google
FB tiles visited
FB single tile
weekly cases
weekly deaths
hospital occupancy
proportion Alpha
seroprevalence
Apr
Jul
Oct
Jan 2021
Apr

#### Switzerland

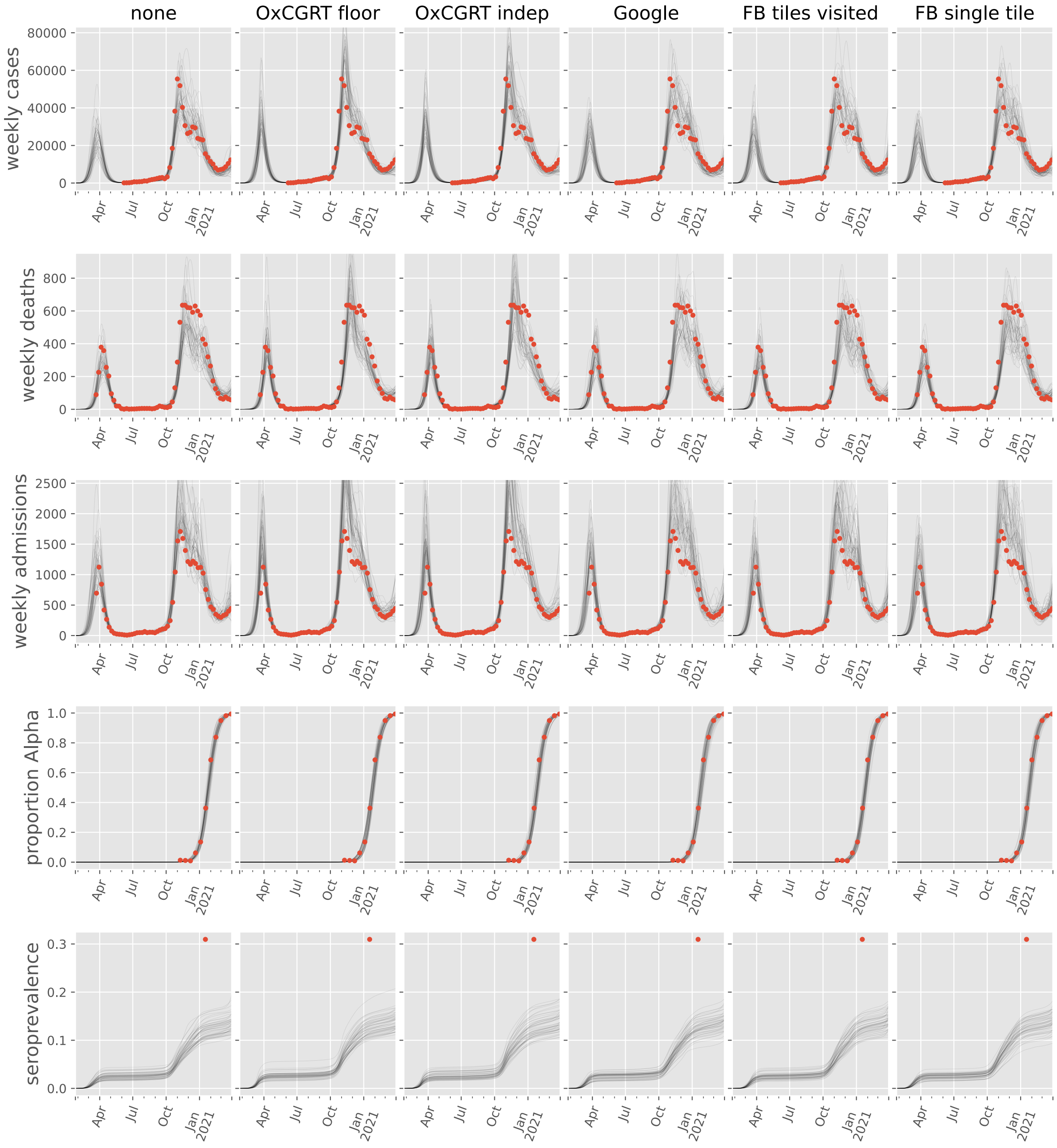

#### Ukraine

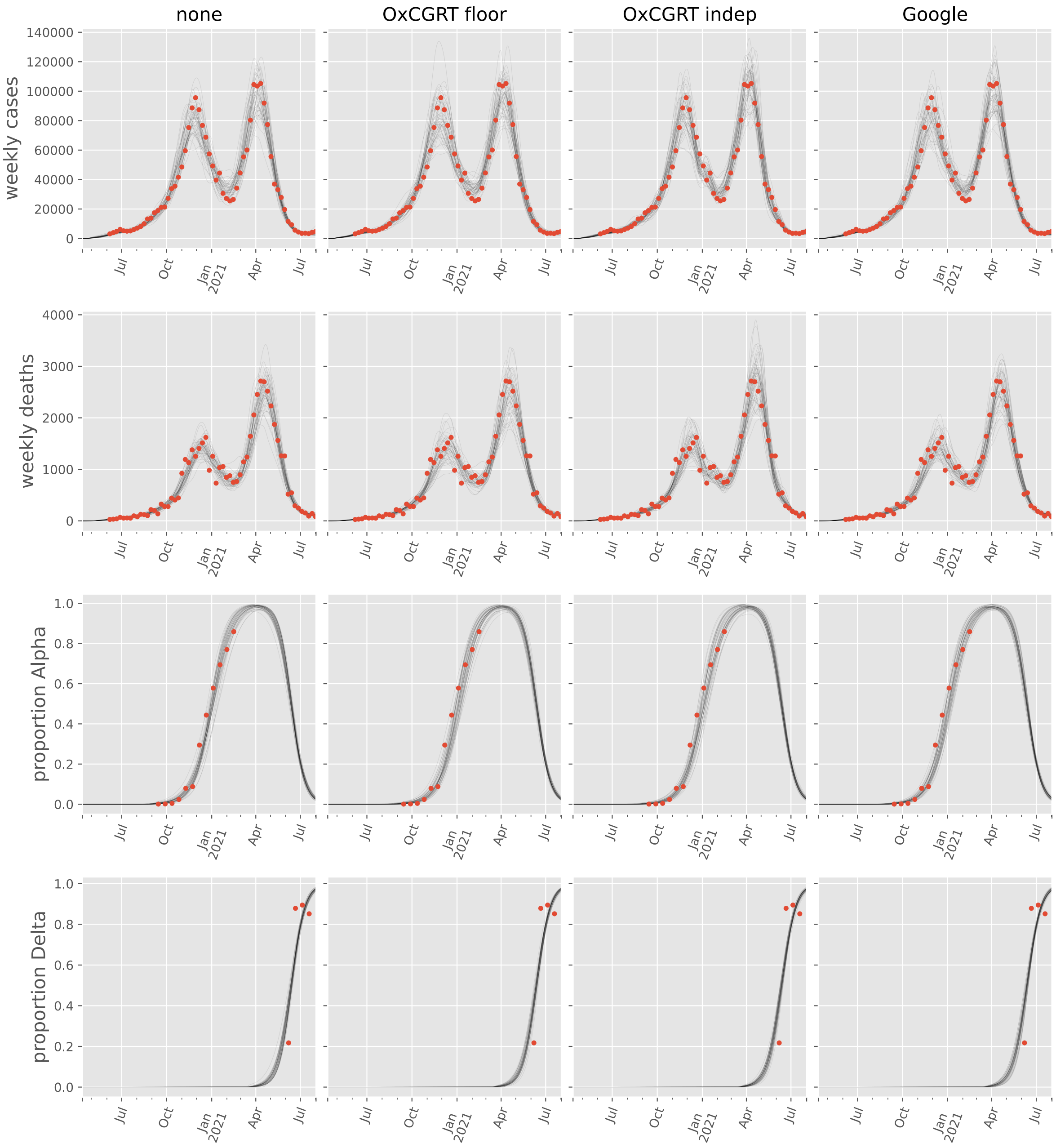

none
OxCGRT floor
OxCGRT indep
Google
weekly cases
weekly deaths
proportion Alpha
proportion Delta
Jul
Oct
Jan
2021
Apr
Jul

#### United Kingdom

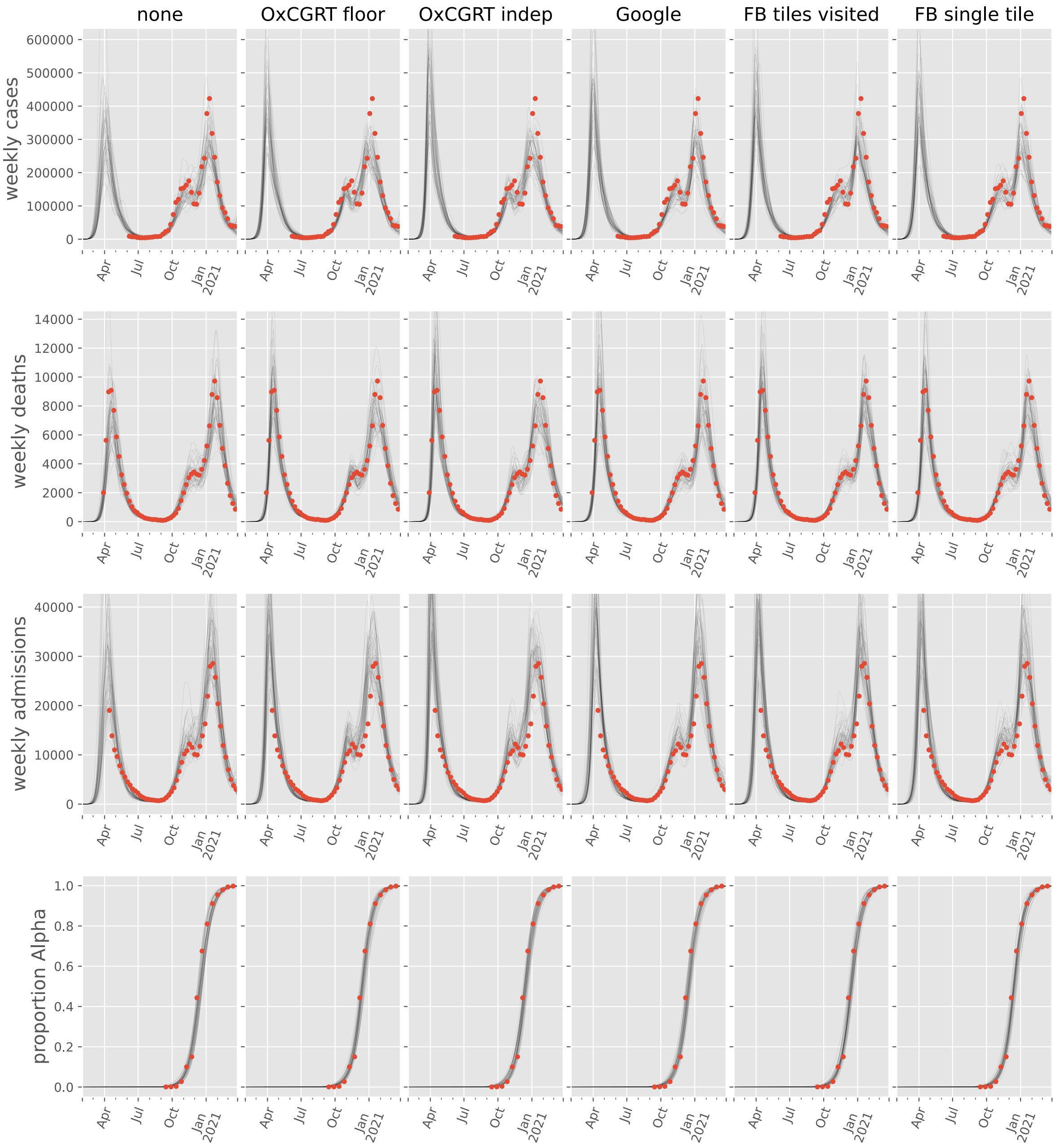

### North America

#### Aruba

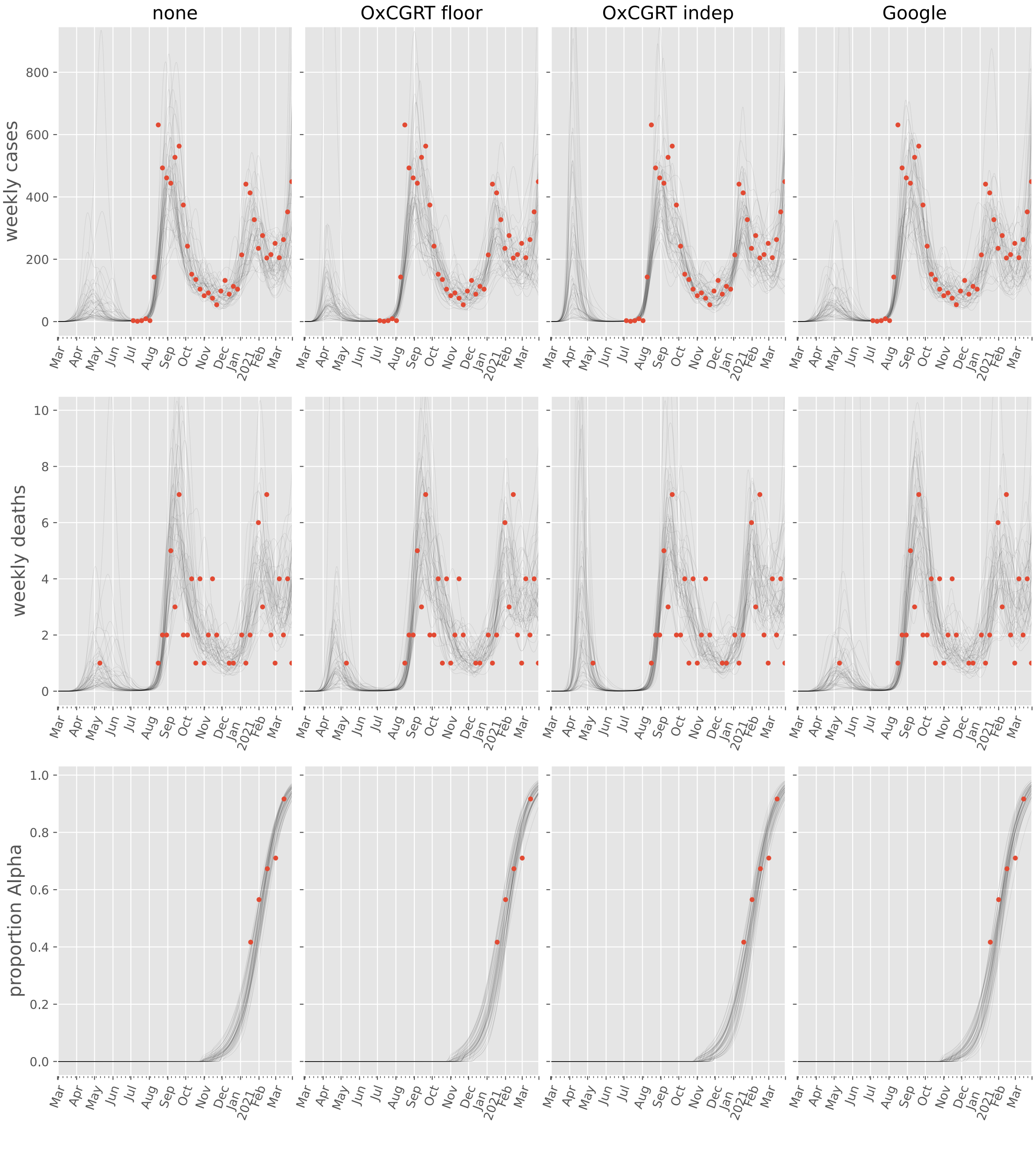

#### Bahamas

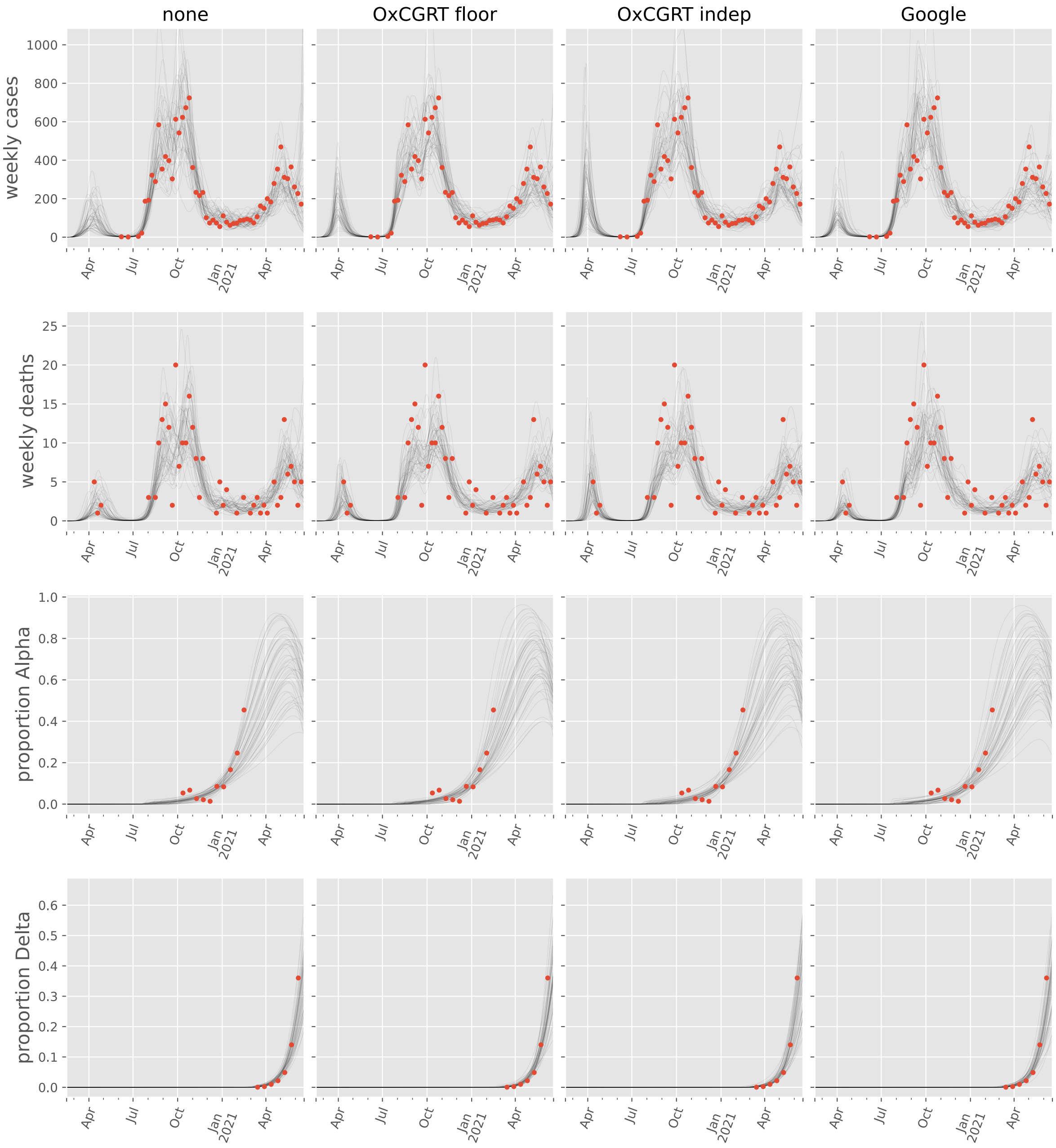

none
OxCGRT floor
OxCGRT indep
Google
weekly cases
weekly deaths
proportion Alpha
proportion Delta
Apr
Jul
Oct
Jan
2021

#### Canada

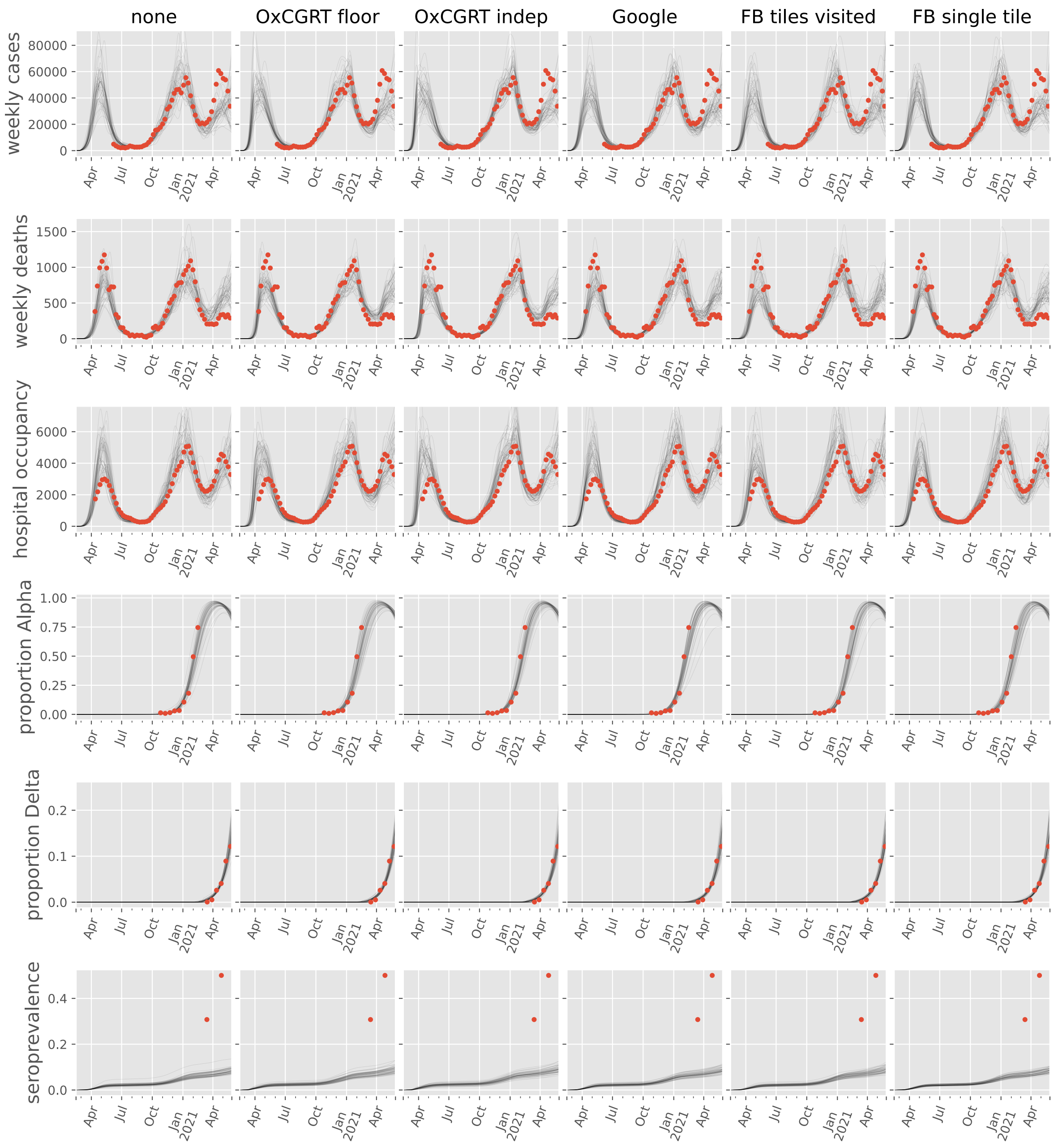
none
OxCGRT floor
OxCGRT indep
Google
FB tiles visited
FB single tile
weekly cases
weekly deaths
hospital occupancy
proportion Alpha
proportion Delta
seroprevalence
Apr
Jul
Oct
Jan
2021
Apr

#### Costa Rica

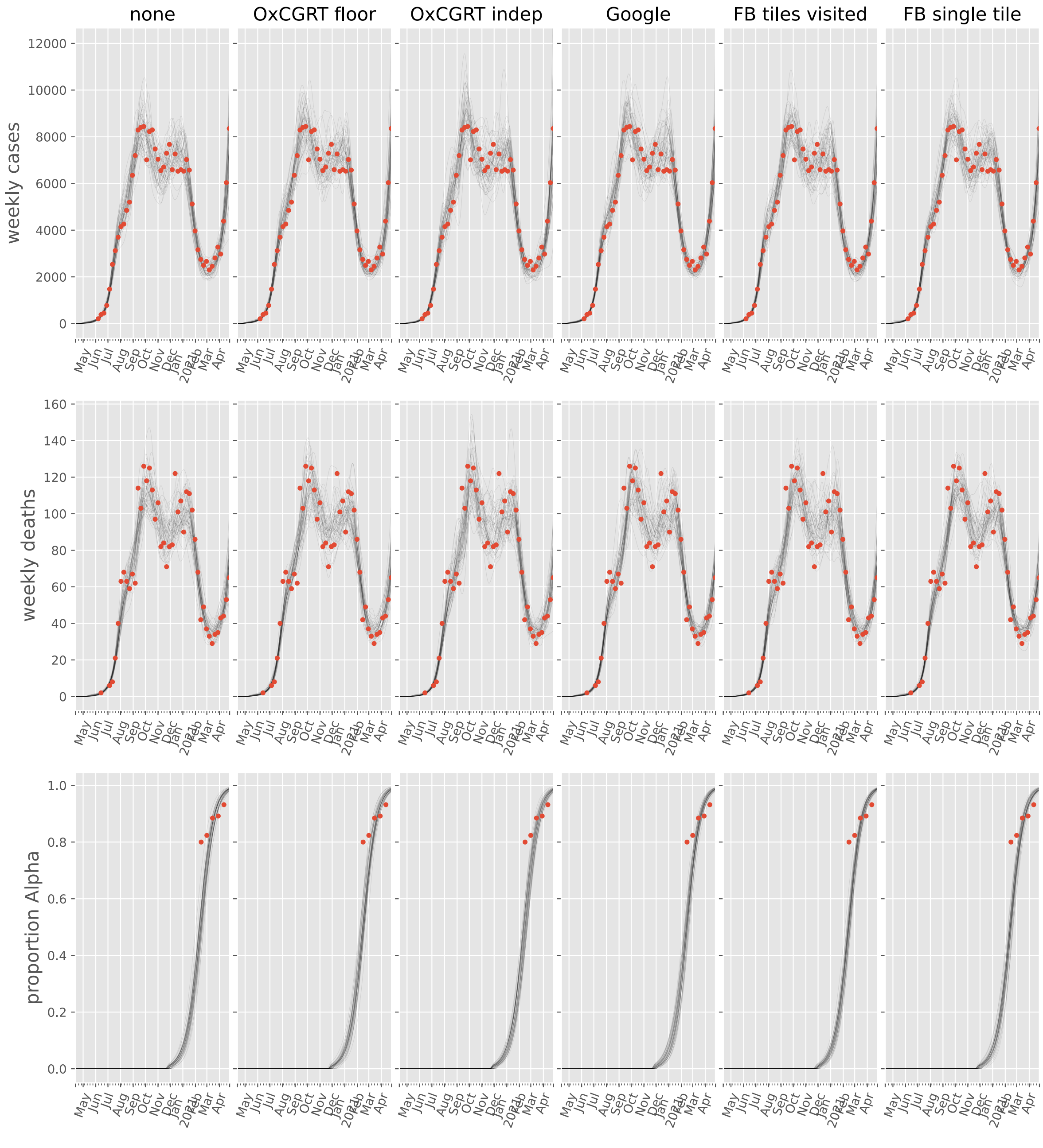
none
OxCGRT floor
OxCGRT indep
Google
FB tiles visited
FB single tile
weekly cases
weekly deaths
proportion Alpha
0
2000
4000
6000
8000
10000
12000
0
20
40
60
80
100
120
140
160
0.0
0.2
0.4
0.6
0.8
1.0
May
Jun
Jul
Aug
Sep
Oct
Nov
Dec
Jan
2021
Feb
Mar
Apr

#### Cuba

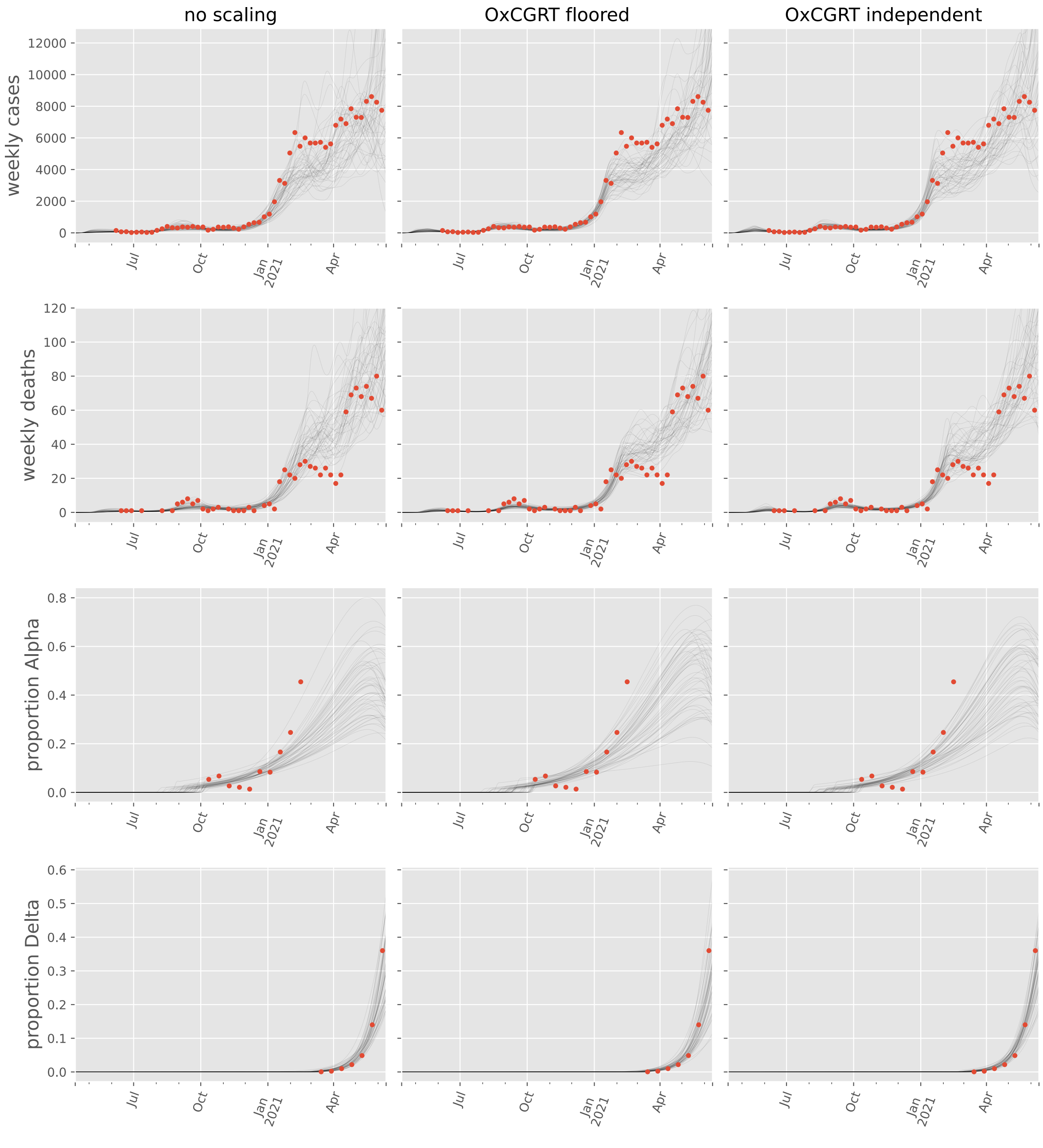

no scaling
OxCGRT floored
OxCGRT independent
weekly cases
weekly deaths
proportion Alpha
proportion Delta
Jul
Oct
Jan
2021
Apr

#### Dominican Republic

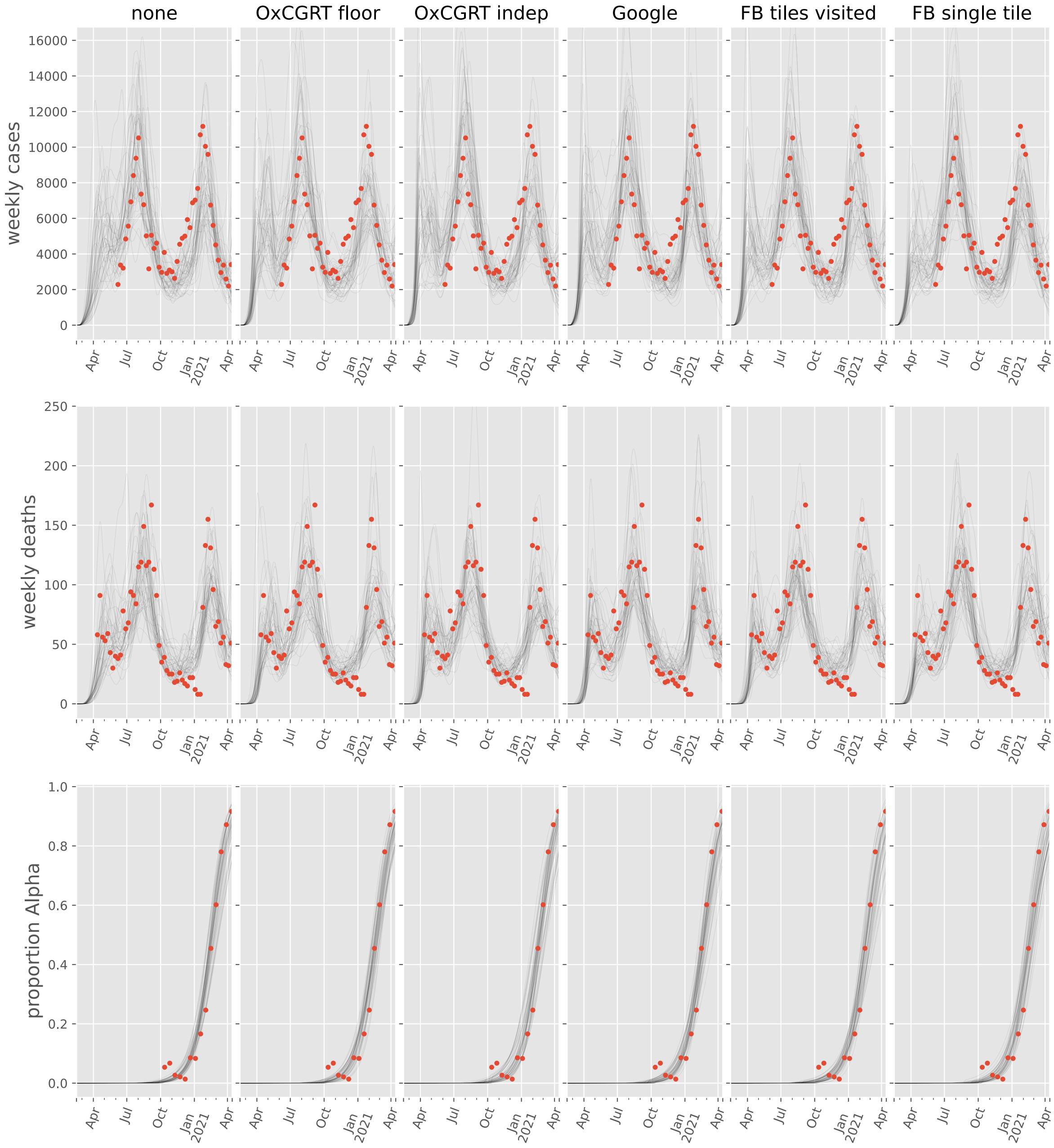

#### El Salvador

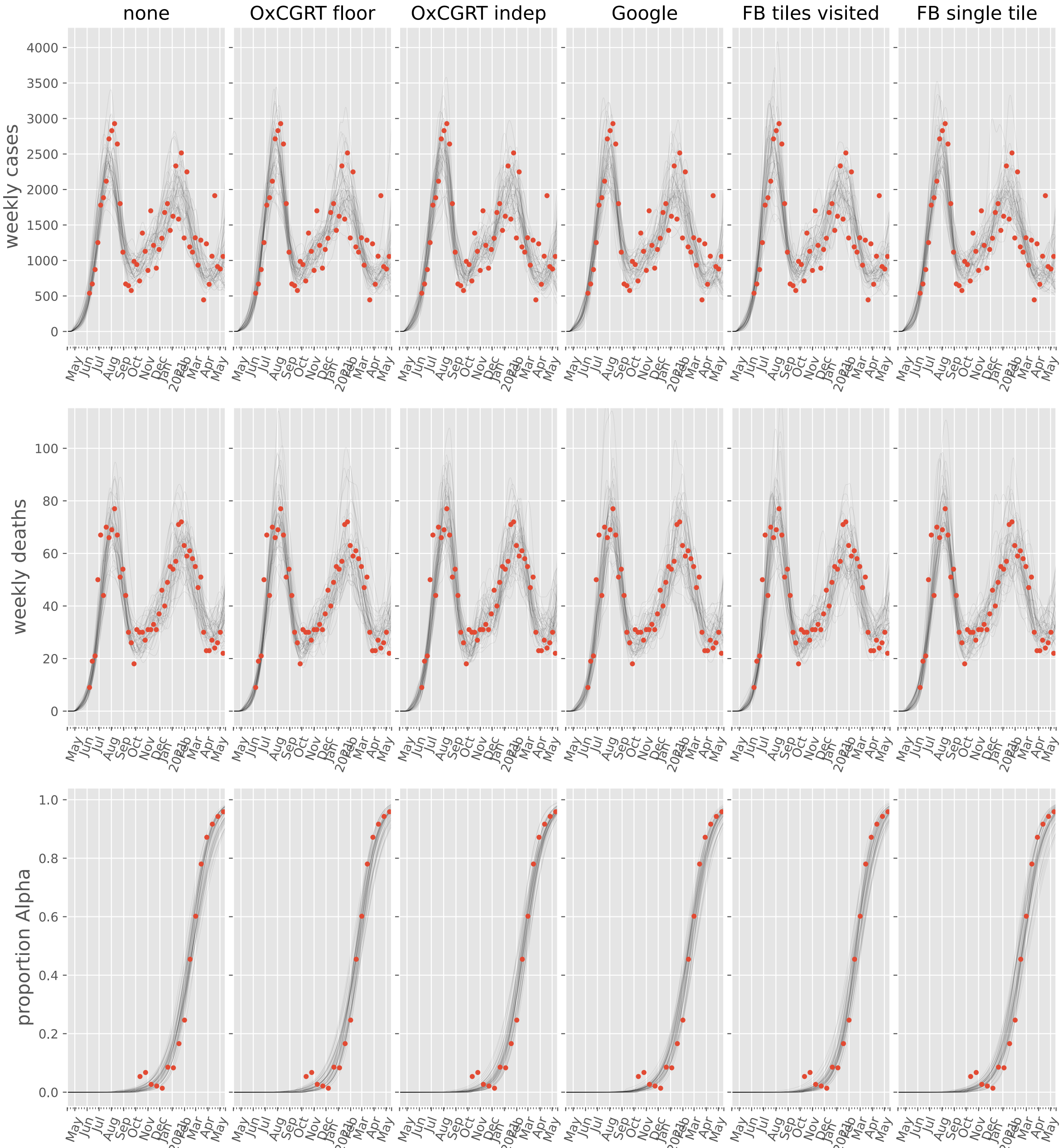
none
OxCGRT floor
OxCGRT indep
Google
FB tiles visited
FB single tile
weekly cases
weekly deaths
proportion Alpha
May Jun Jul Aug Sep Oct Nov Dec Jan 2021 Feb Mar Apr May

#### Haiti

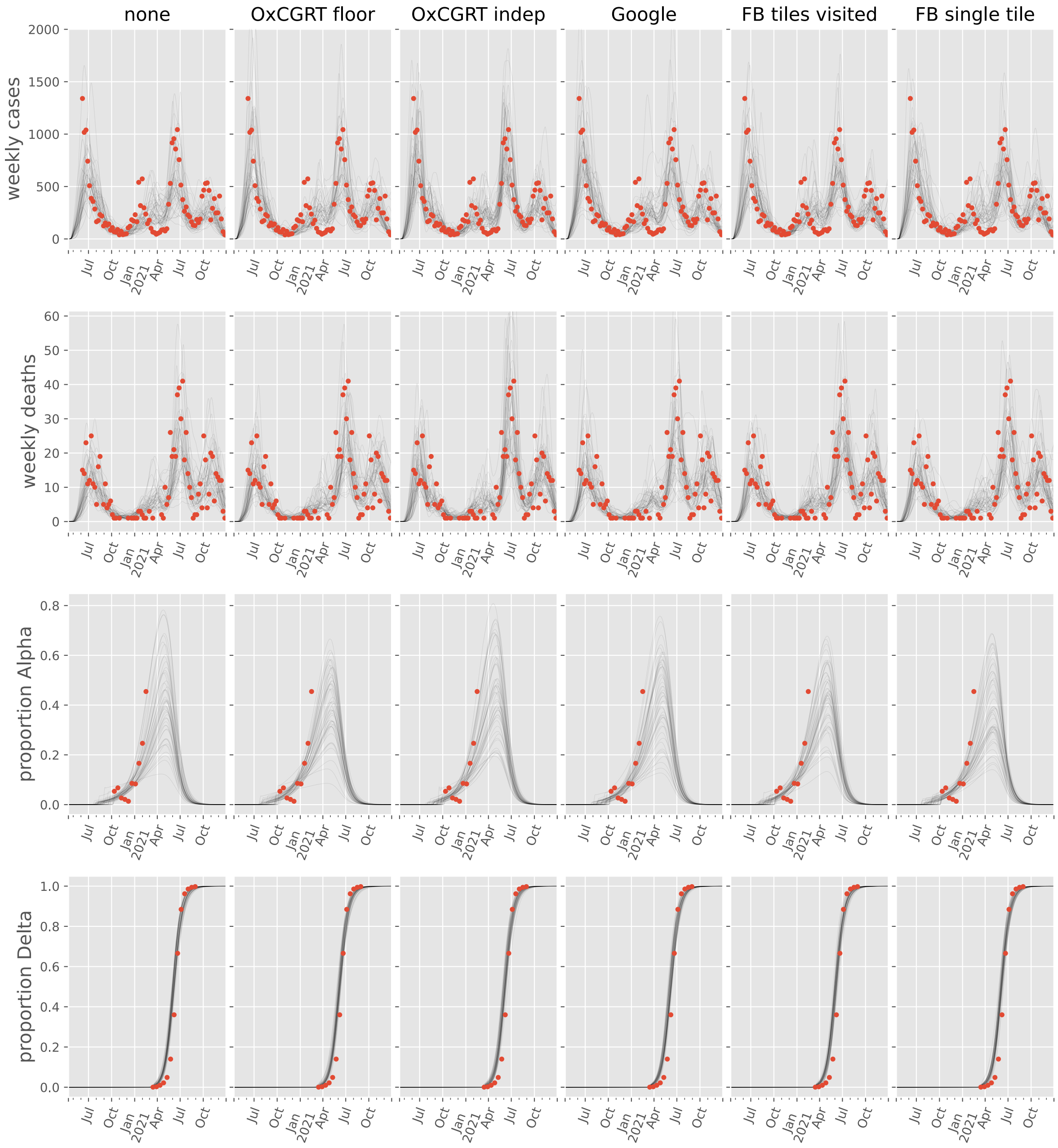

none
OxCGRT floor
OxCGRT indep
Google
FB tiles visited
FB single tile
weekly cases
weekly deaths
proportion Alpha
proportion Delta
2000
1500
1000
500
0
60
50
40
30
20
10
0.8
0.6
0.4
0.2
0.0
1.0
Jul
Oct
Jan
2021
Apr

#### Honduras

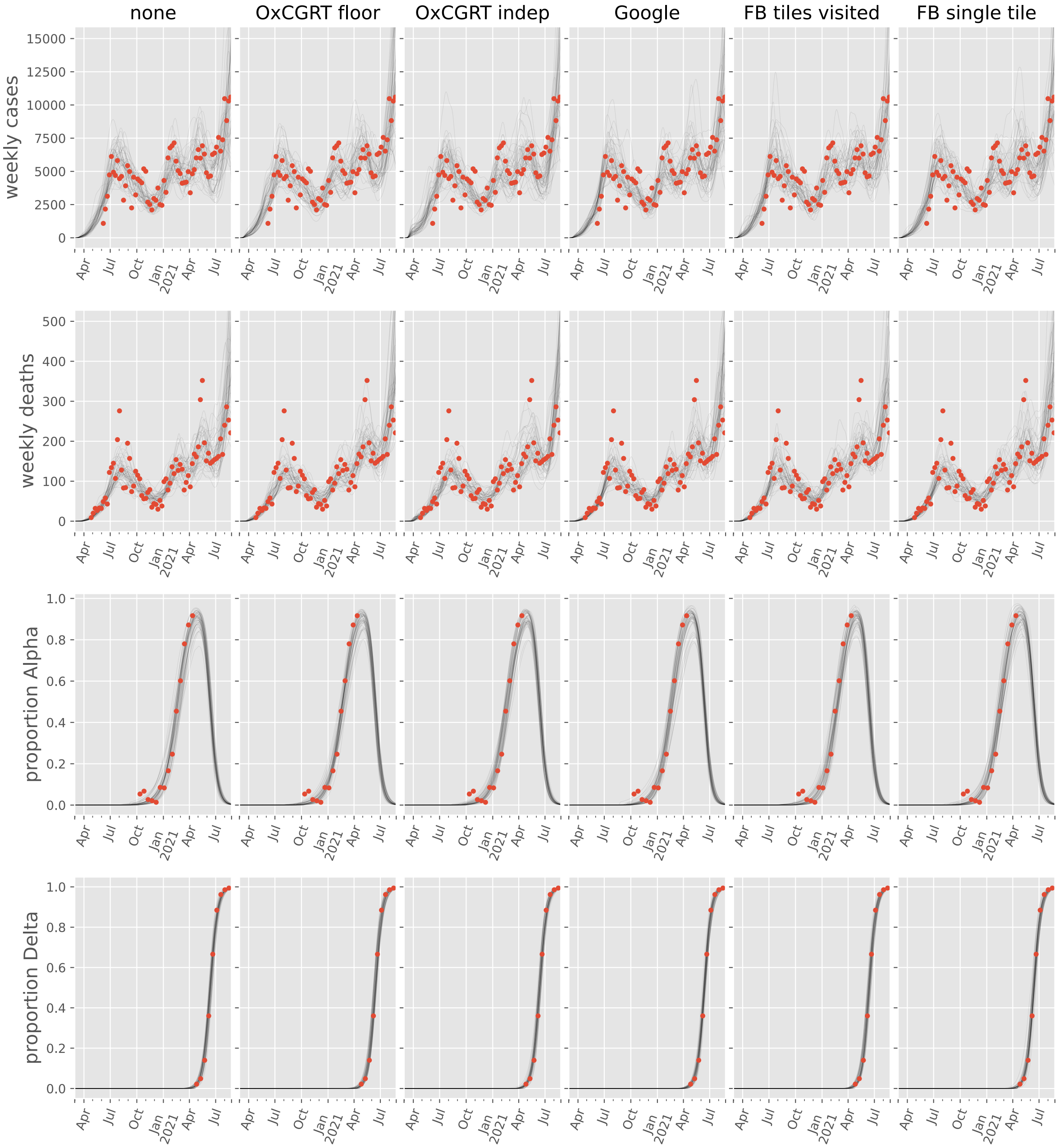
none
OxCGRT floor
OxCGRT indep
Google
FB tiles visited
FB single tile
weekly cases
weekly deaths
proportion Alpha
proportion Delta
Apr
Jul
Oct
Jan 2021
Apr
Jul

#### Jamaica

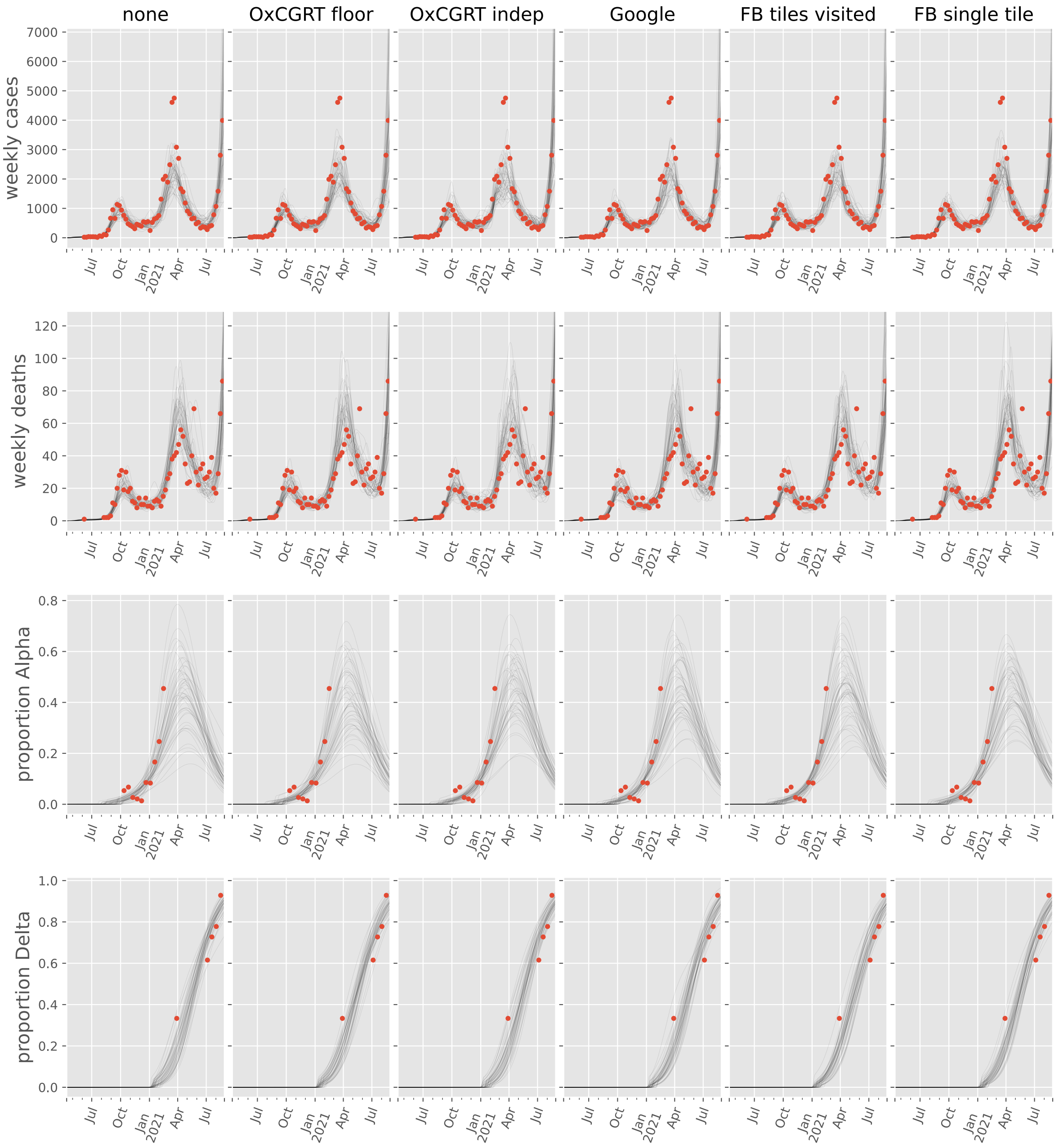

none
OxCGRT floor
OxCGRT indep
Google
FB tiles visited
FB single tile
weekly cases
weekly deaths
proportion Alpha
proportion Delta
Jul
Oct
Jan
2021
Apr
Jul

#### Mexico

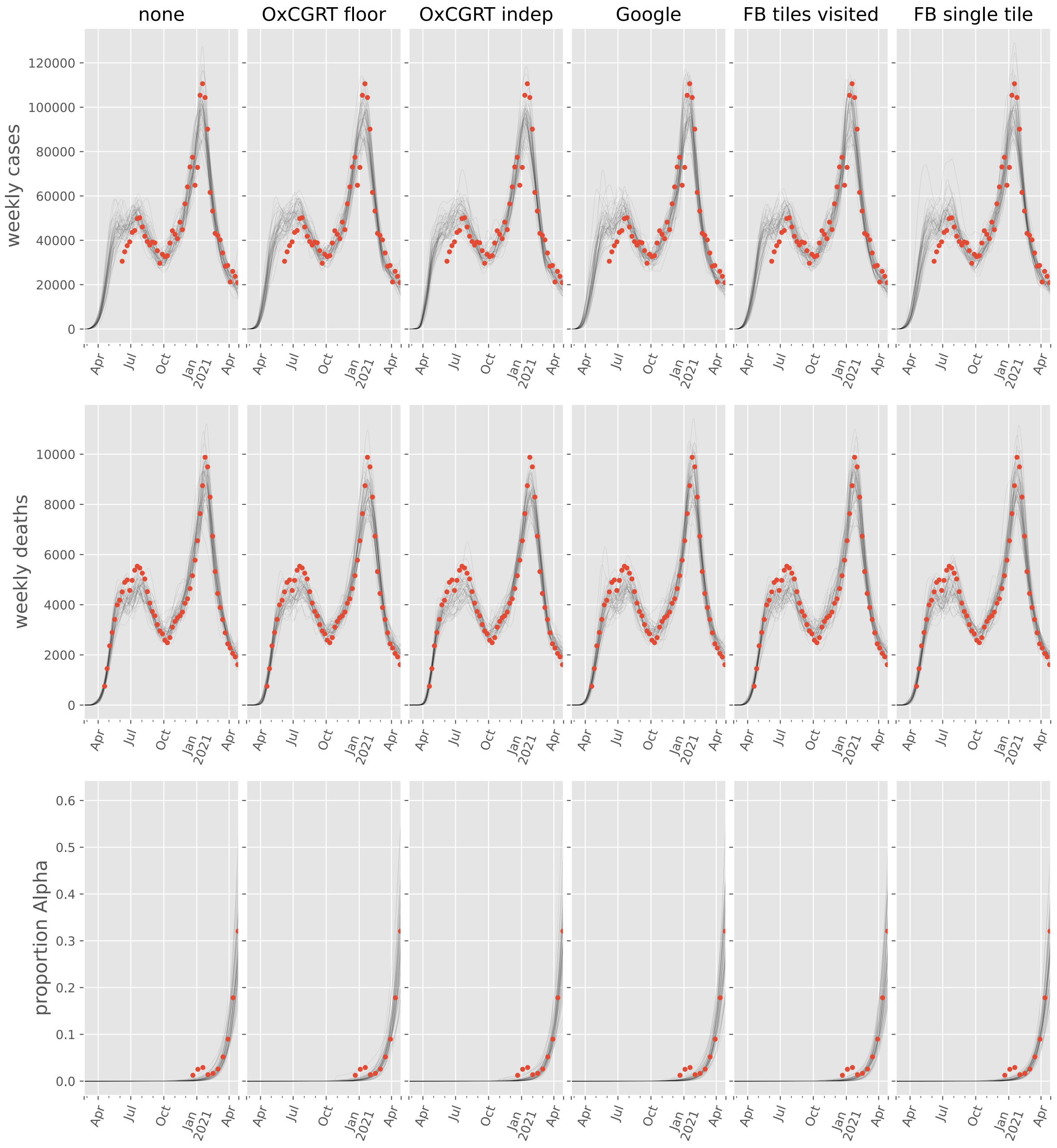
none
OxCGRT floor
OxCGRT indep
Google
FB tiles visited
FB single tile
weekly cases
weekly deaths
proportion Alpha
0
20000
40000
60000
80000
100000
120000
2000
4000
6000
8000
10000
0.0
0.1
0.2
0.3
0.4
0.5
0.6
Apr
Jul
Oct
Jan
2021
Apr

#### Panama

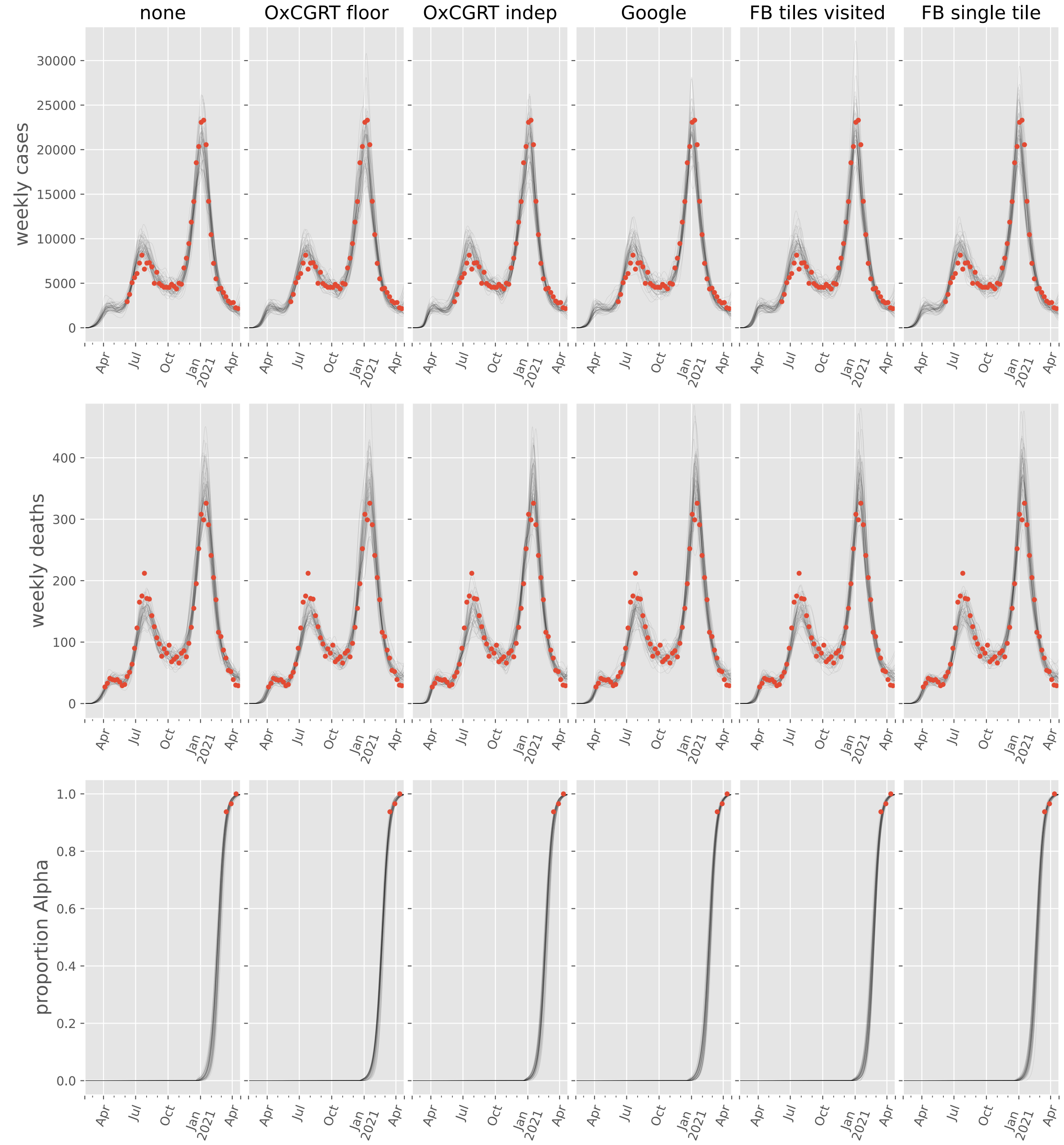
none
OxCGRT floor
OxCGRT indep
Google
FB tiles visited
FB single tile
weekly cases
weekly deaths
proportion Alpha
30000
25000
20000
15000
10000
5000
0
400
300
200
100
0
1.0
0.8
0.6
0.4
0.2
0.0
Apr
Jul
Oct
Jan
2021
Apr

#### United States

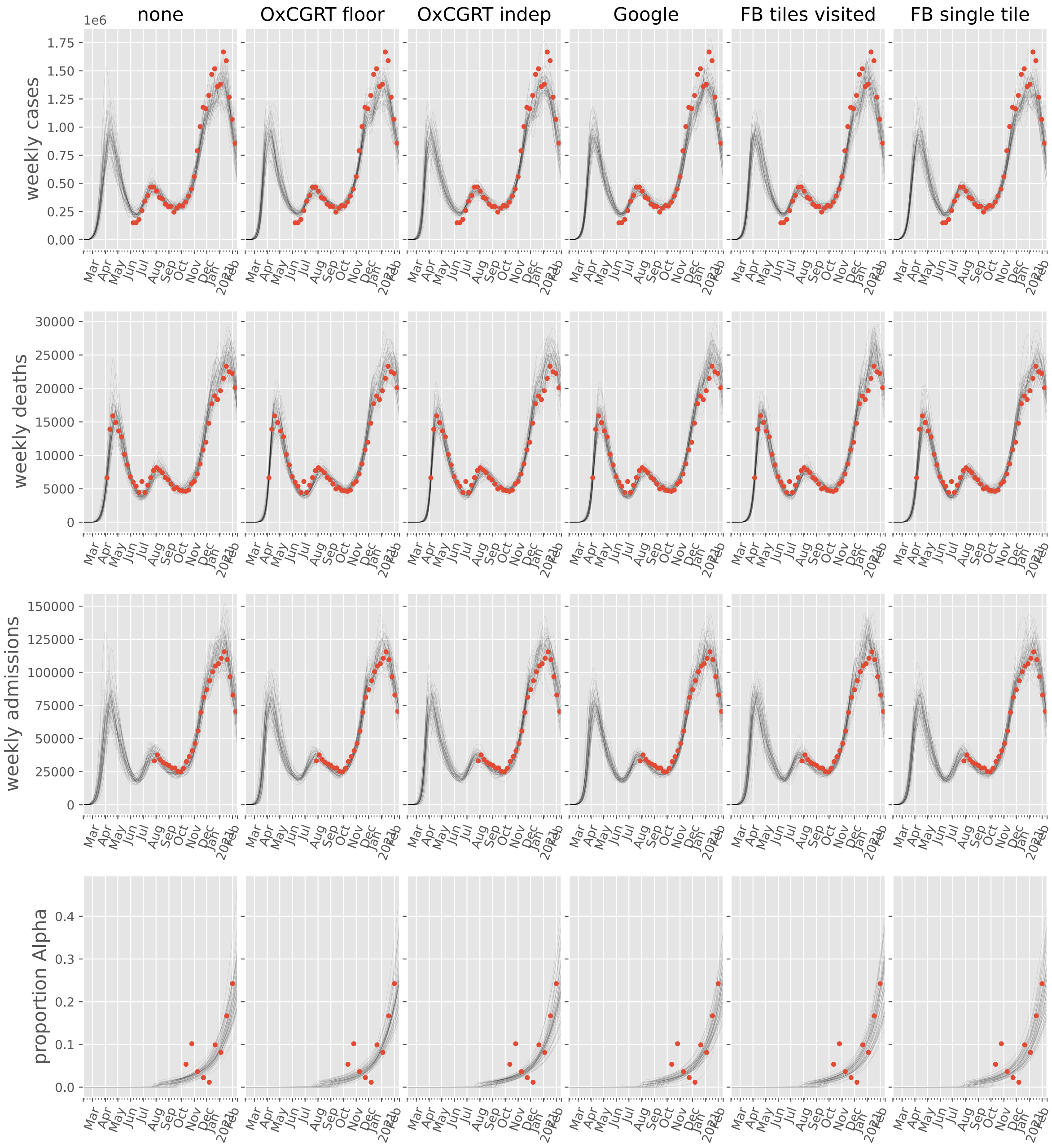

### Oceania

#### Australia

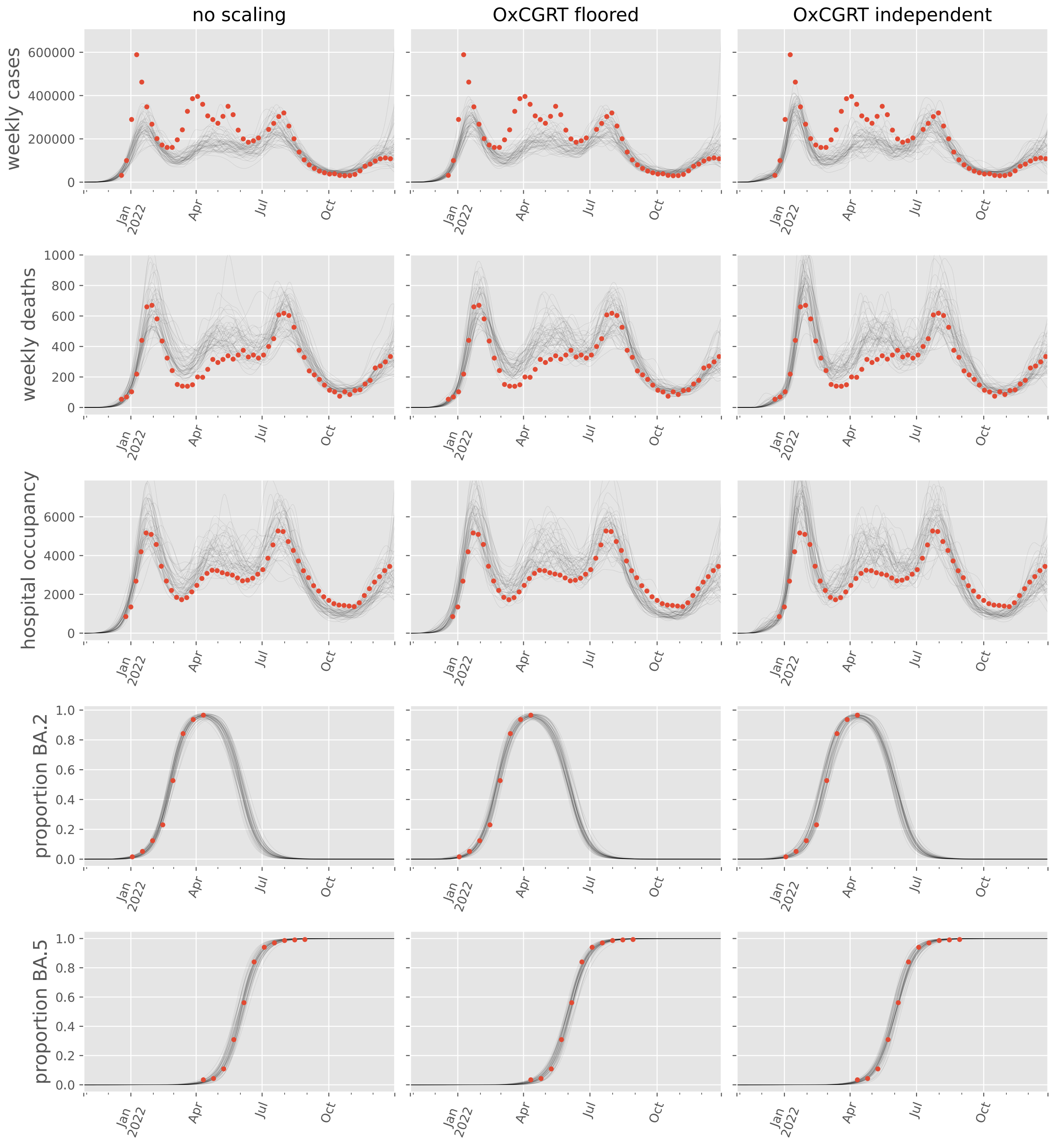

#### New Zealand

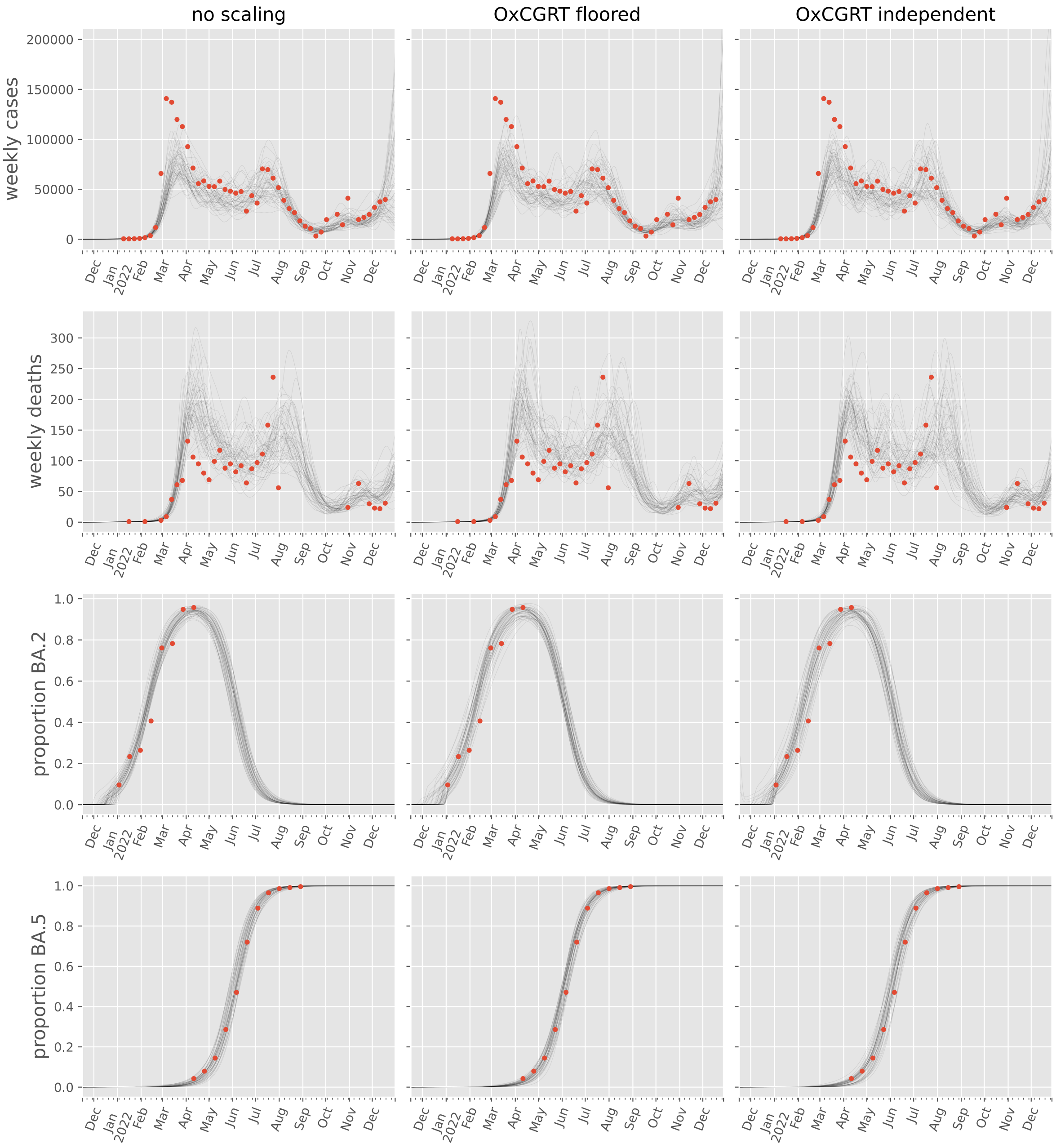

no scaling
OxCGRT floored
OxCGRT independent
weekly cases
weekly deaths
proportion BA.2
proportion BA.5
200000
150000
100000
50000
0
300
250
200
150
100
50
1.0
0.8
0.6
0.4
0.2
0.0
Dec
Jan
2022
Feb
Mar
Apr
May
Jun
Jul
Aug
Sep
Oct
Nov
Dec

#### Singapore

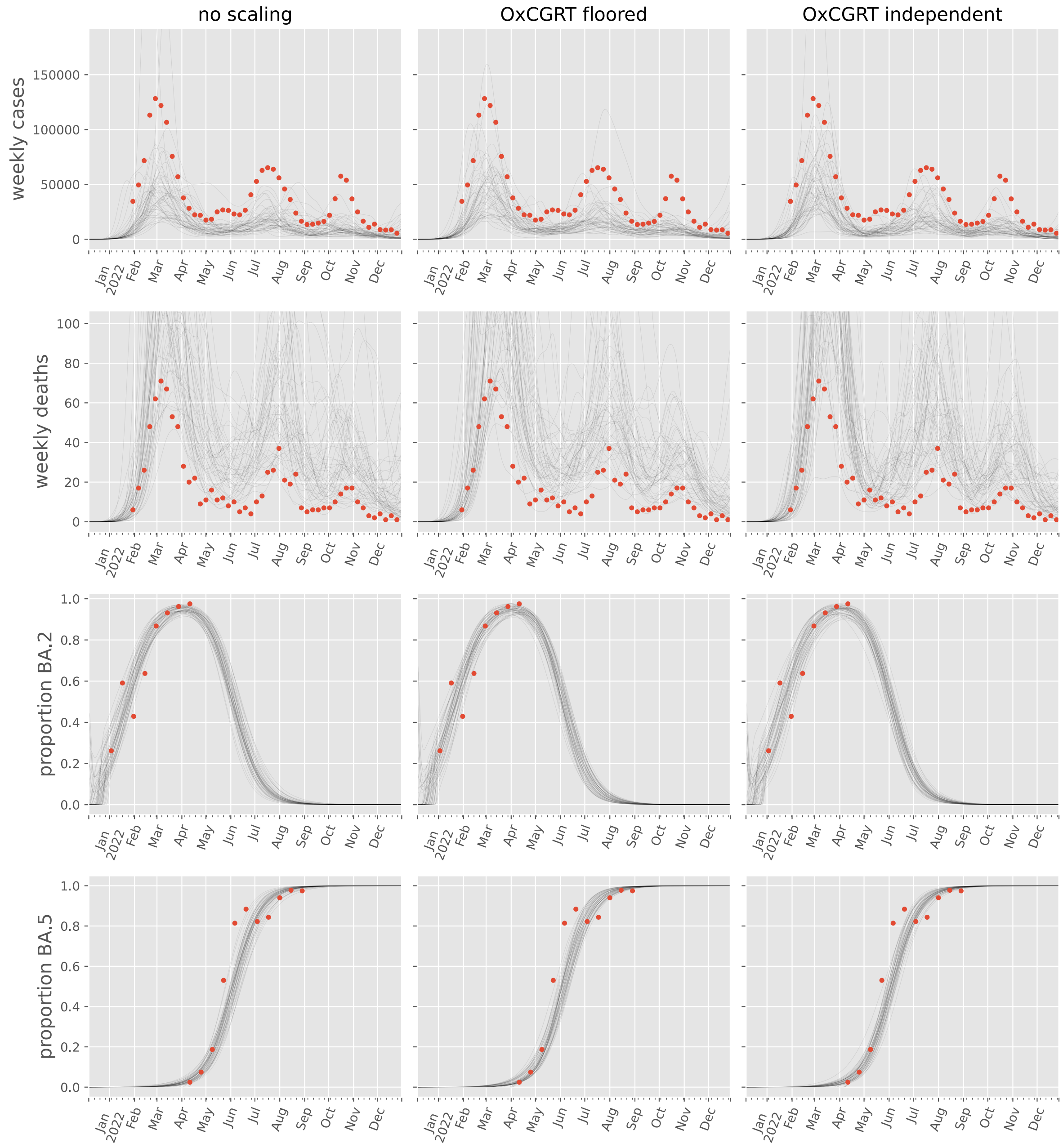

no scaling
OxCGRT floored
OxCGRT independent
weekly cases
weekly deaths
proportion BA.2
proportion BA.5
Jan 2022
Feb
Mar
Apr
May
Jun
Jul
Aug
Sep
Oct
Nov
Dec

### South America

#### Argentina

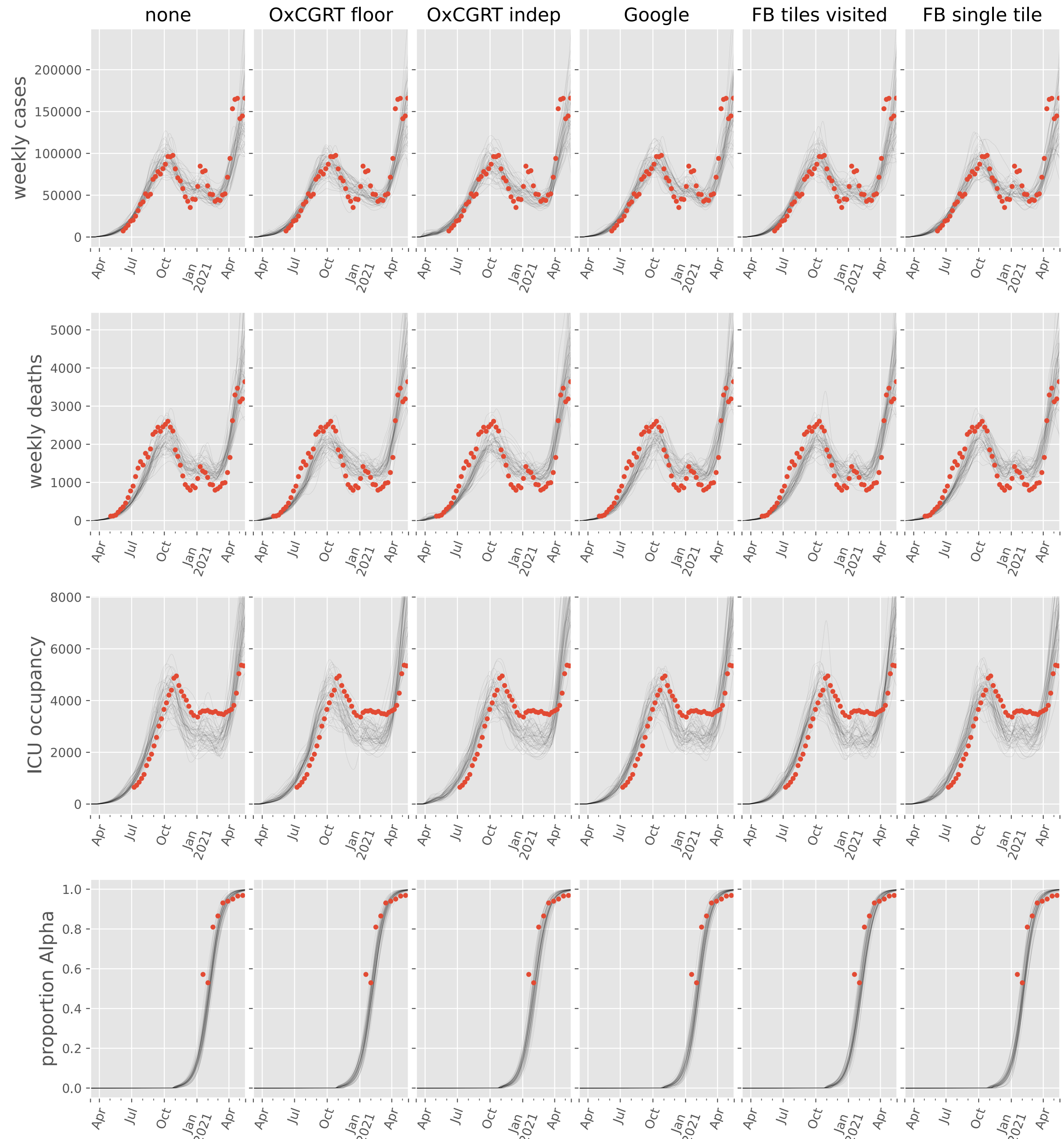

#### Brazil

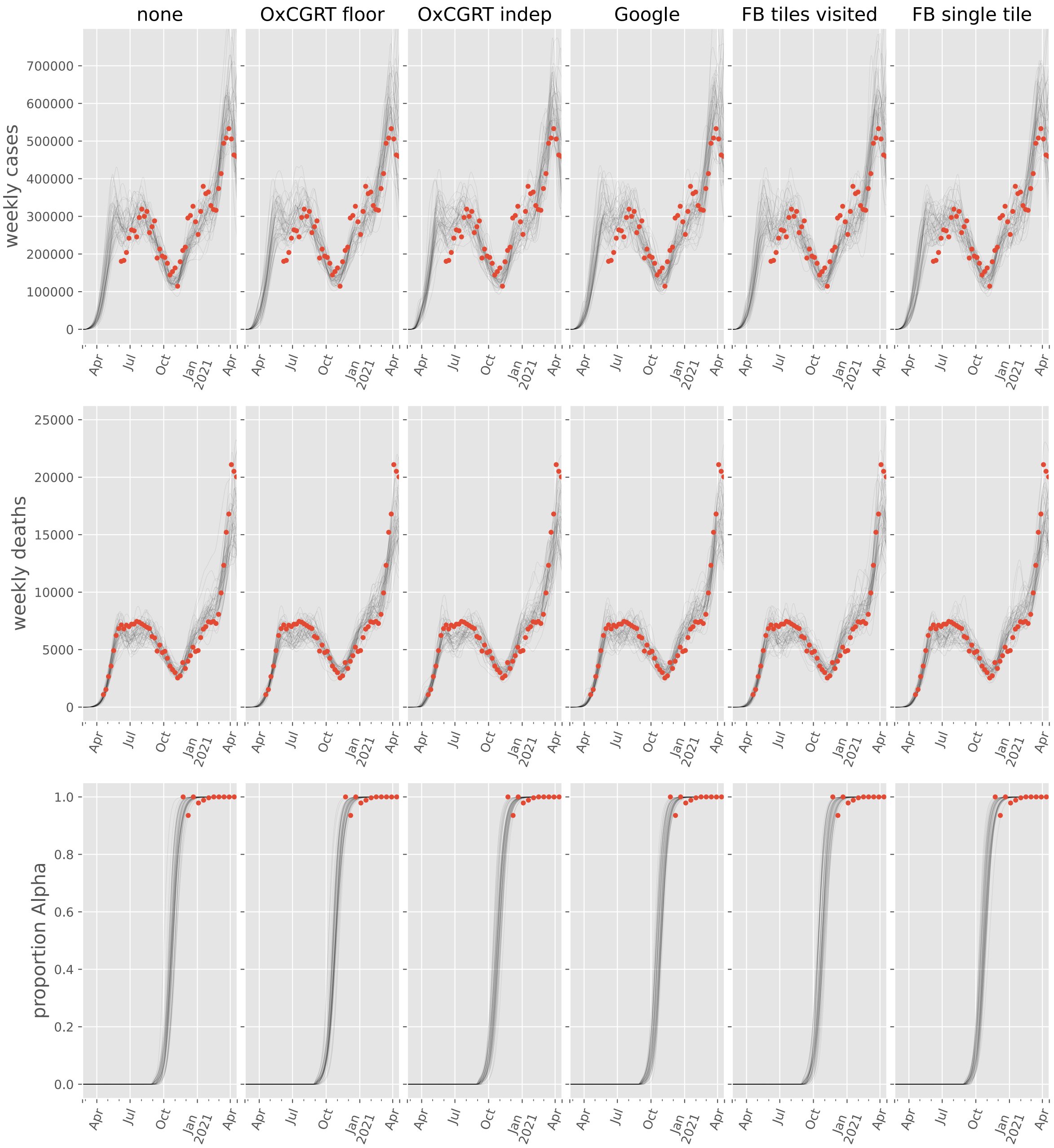
none
OxCGRT floor
OxCGRT indep
Google
FB tiles visited
FB single tile
weekly cases
weekly deaths
proportion Alpha
Apr
Jul
Oct
Jan 2021
Apr

#### Chile

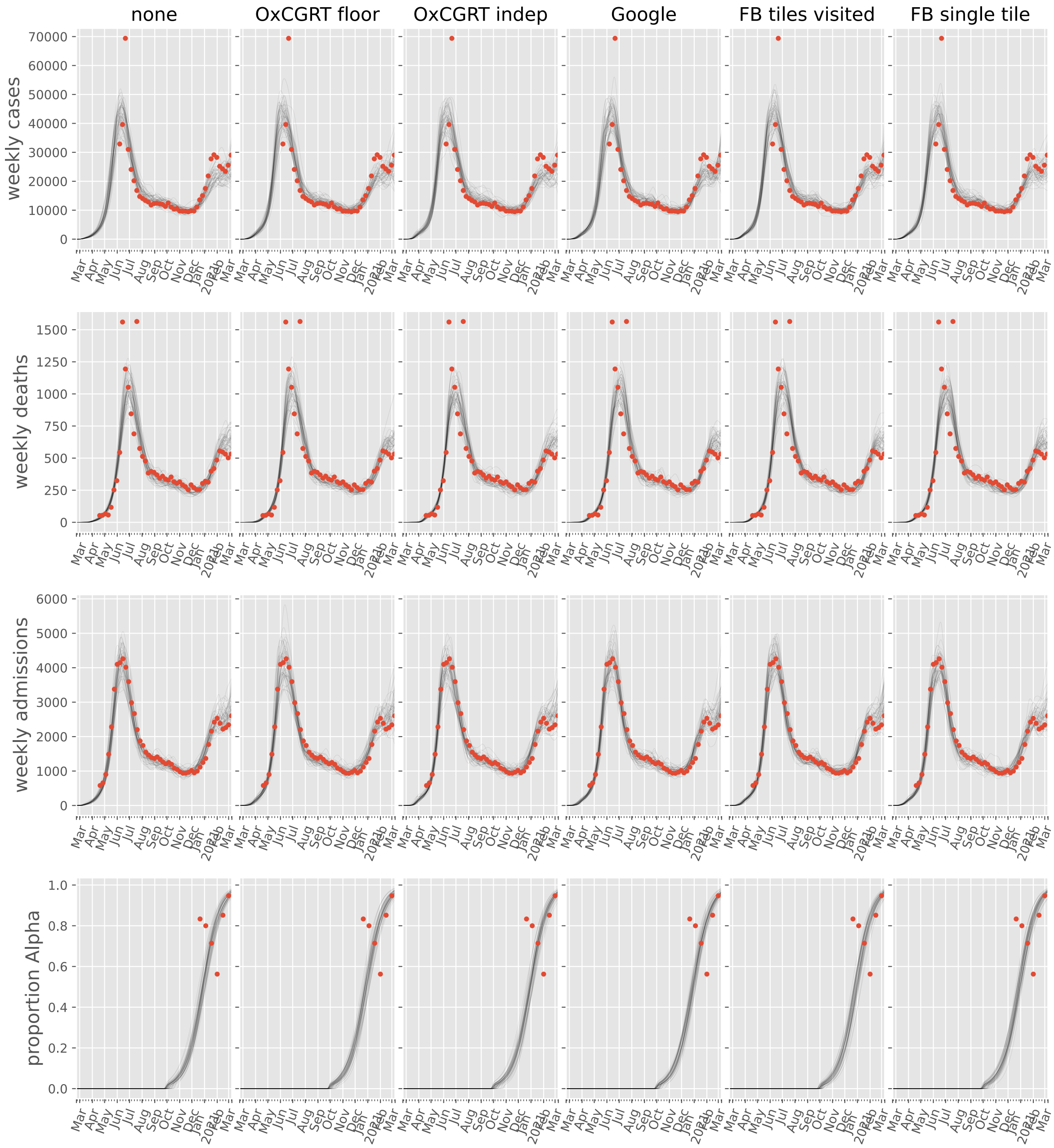

none
OxCGRT floor
OxCGRT indep
Google
FB tiles visited
FB single tile
weekly cases
weekly deaths
weekly admissions
proportion Alpha

#### Colombia

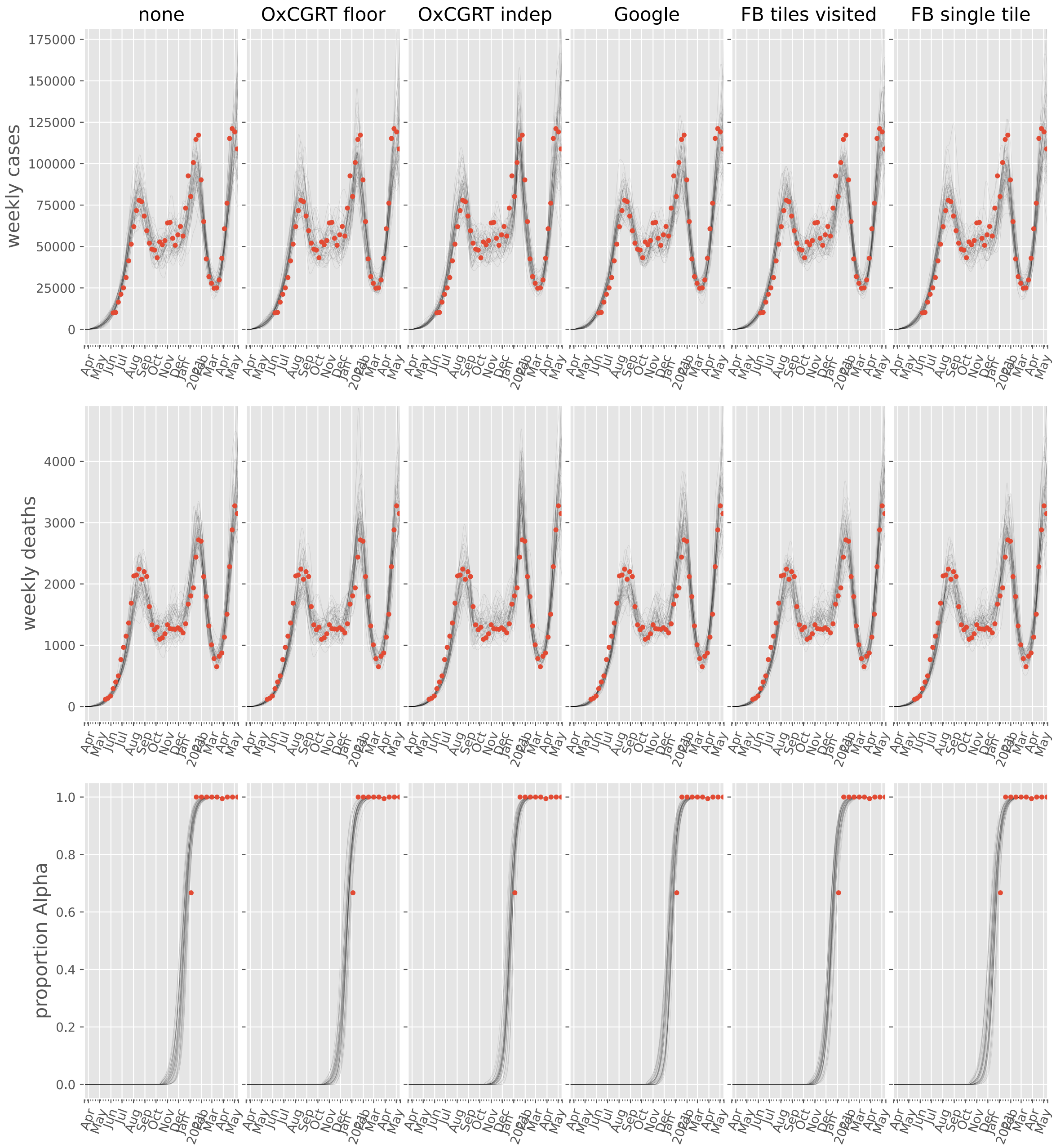

none
OxCGRT floor
OxCGRT indep
Google
FB tiles visited
FB single tile
weekly cases
weekly deaths
proportion Alpha
Apr May Jun Jul Aug Sep Oct Nov Dec Jan 2021 Feb Mar Apr May

#### Guyana

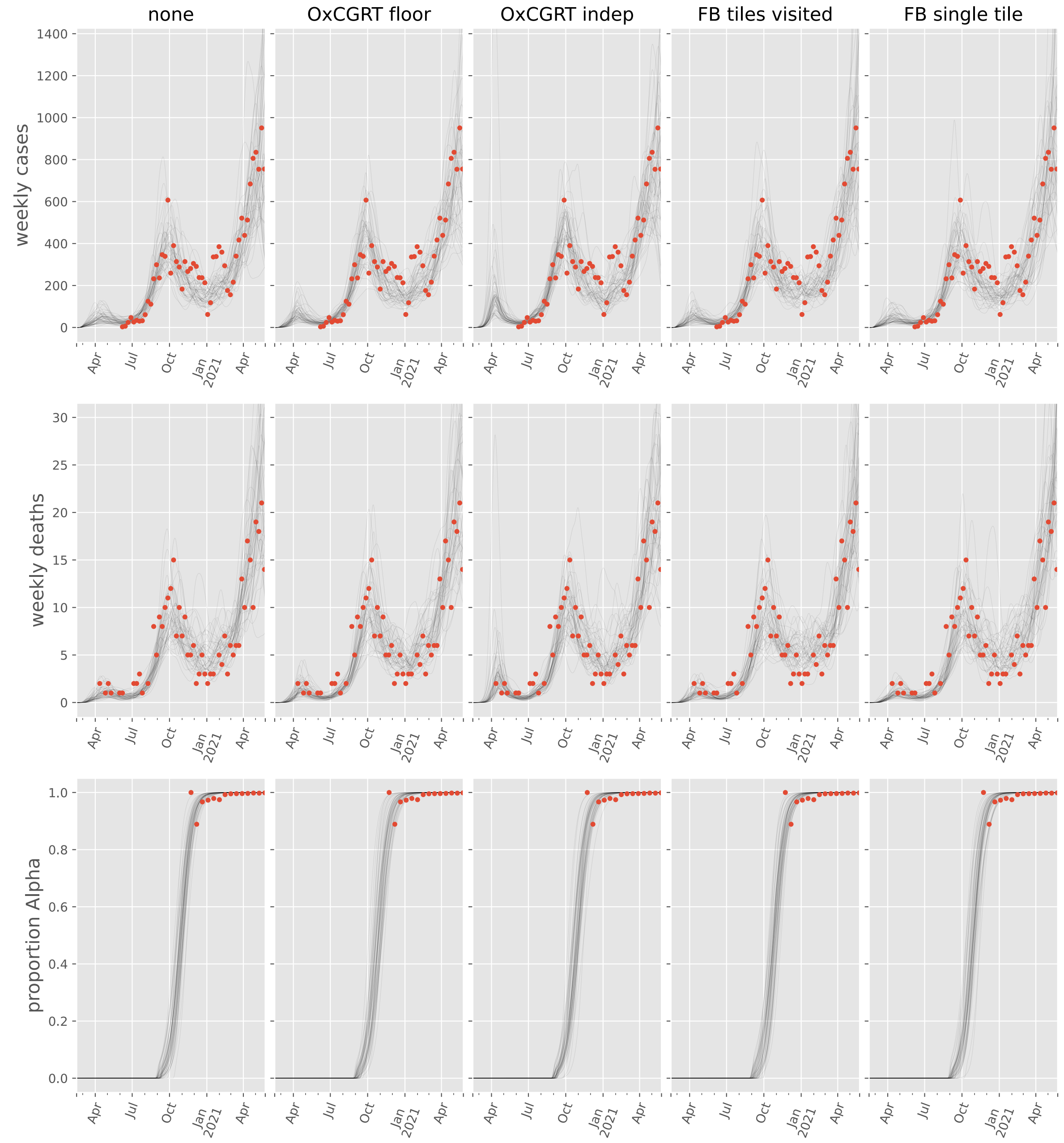

none
OxCGRT floor
OxCGRT indep
FB tiles visited
FB single tile
weekly cases
weekly deaths
proportion Alpha
0
200
400
600
800
1000
1200
1400
0
5
10
15
20
25
30
0.0
0.2
0.4
0.6
0.8
1.0
Apr
Jul
Oct
Jan
2021
Apr

#### Paraguay

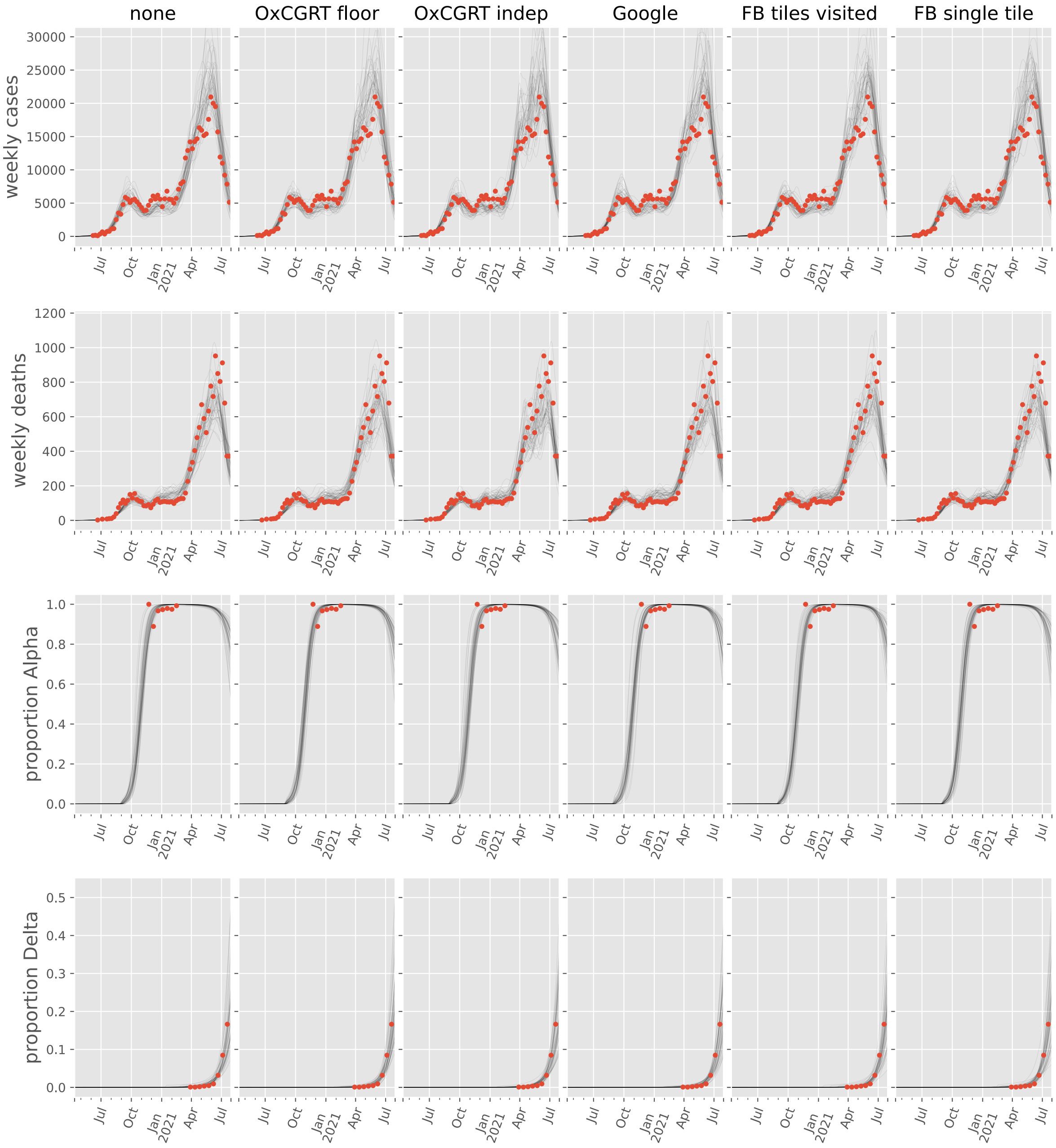

none
OxCGRT floor
OxCGRT indep
Google
FB tiles visited
FB single tile
weekly cases
weekly deaths
proportion Alpha
proportion Delta
Jul
Oct
Jan 2021
Apr

#### Peru

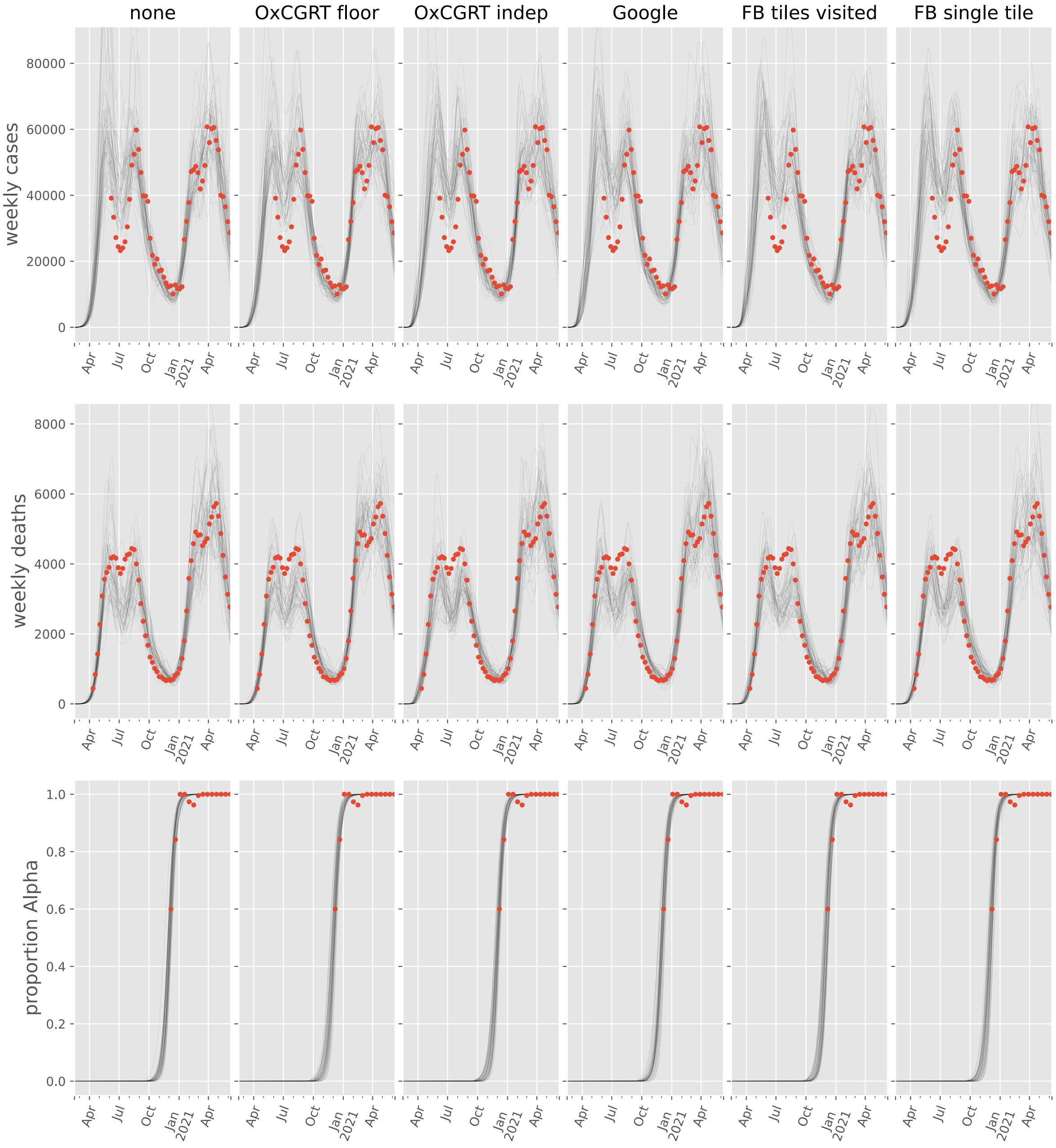

none
OxCGRT floor
OxCGRT indep
Google
FB tiles visited
FB single tile
weekly cases
80000
60000
40000
20000
0
weekly deaths
8000
6000
4000
2000
0
proportion Alpha
1.0
0.8
0.6
0.4
0.2
0.0
Apr
Jul
Oct
Jan 2021
Apr

#### Suriname

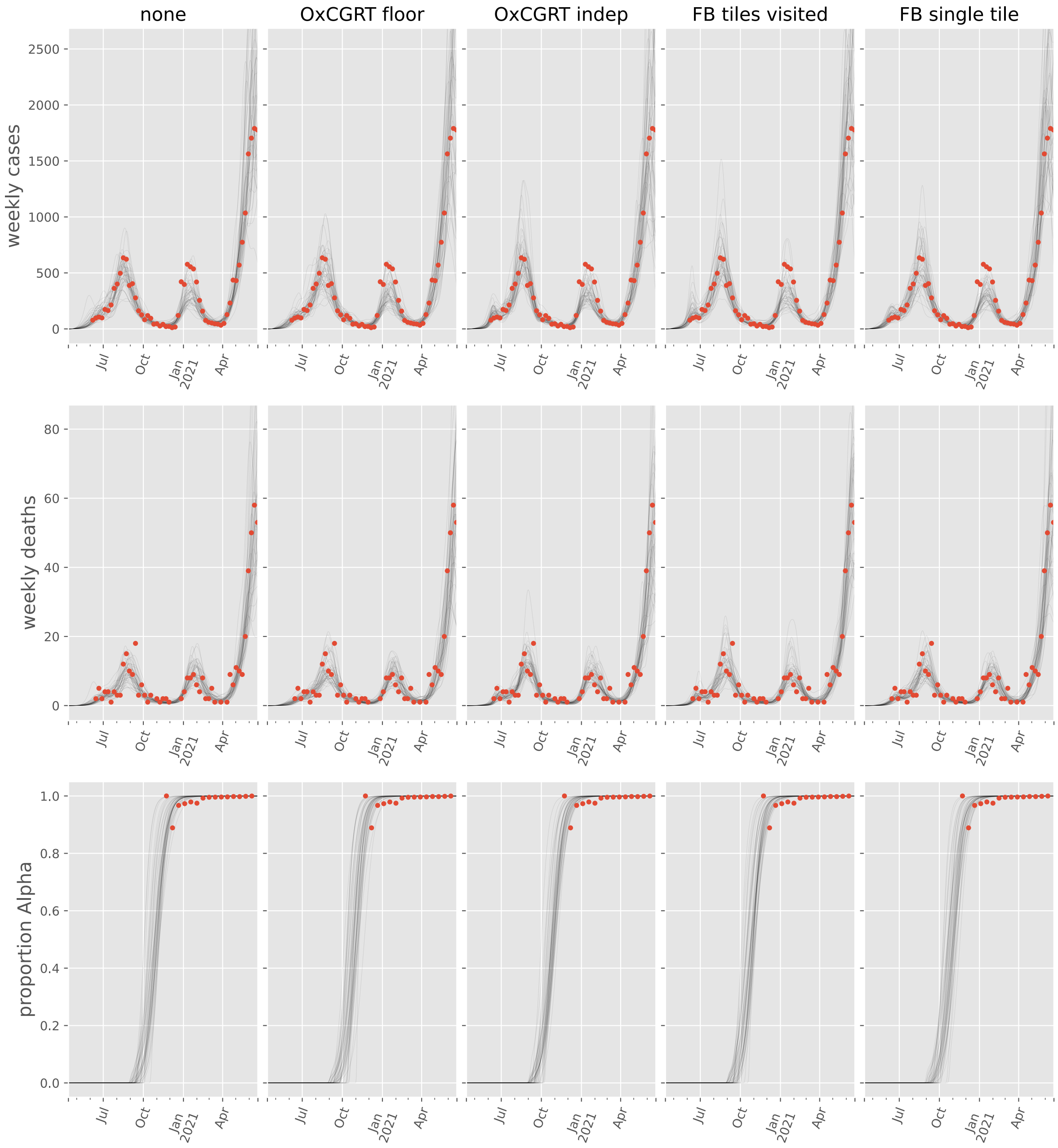
none
OxCGRT floor
OxCGRT indep
FB tiles visited
FB single tile
weekly cases
weekly deaths
proportion Alpha
Jul
Oct
Jan
2021
Apr

#### Uruguay

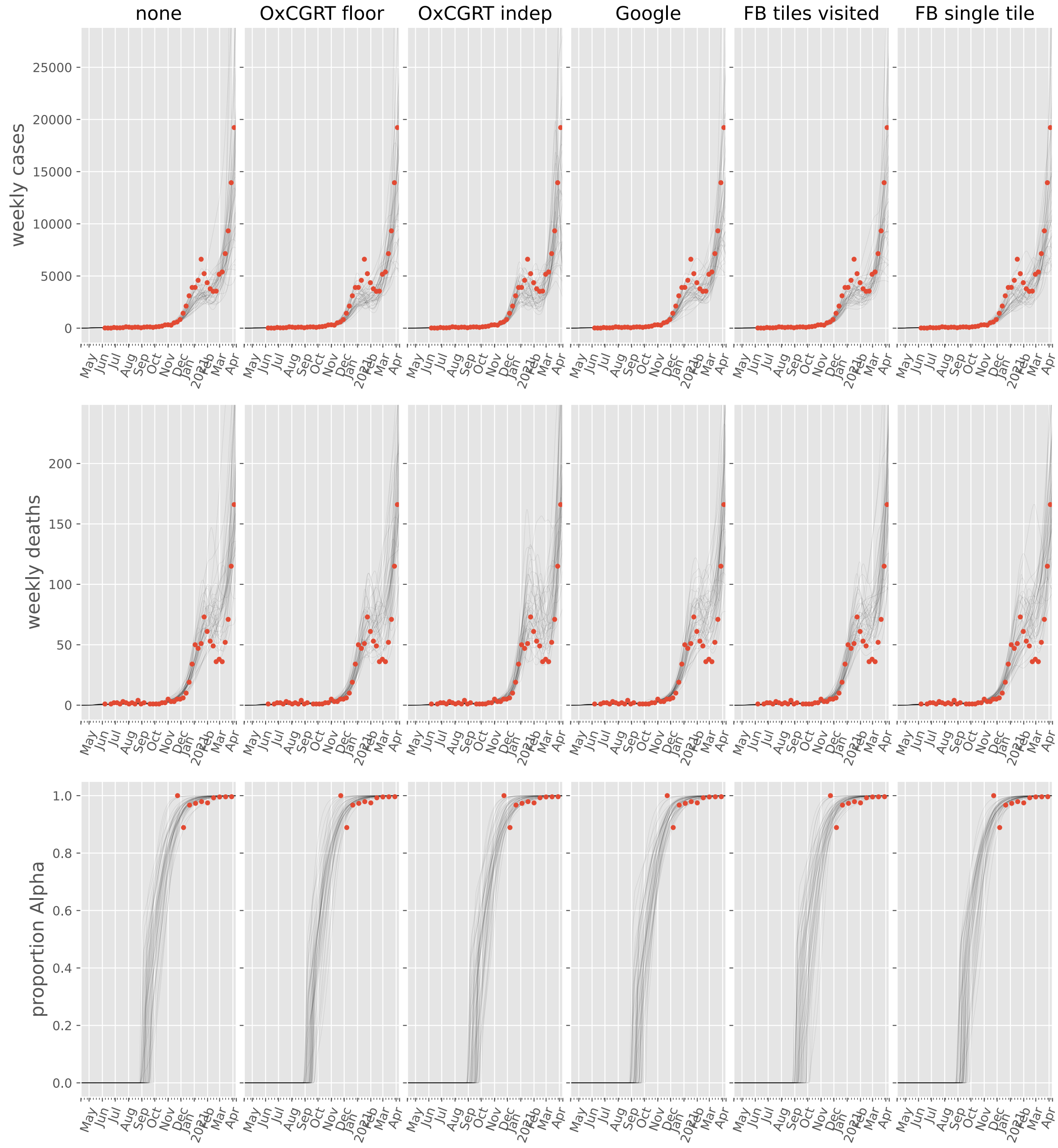

none
OxCGRT floor
OxCGRT indep
Google
FB tiles visited
FB single tile
weekly cases
25000
20000
15000
10000
5000
0
weekly deaths
200
150
100
50
0
proportion Alpha
1.0
0.8
0.6
0.4
0.2
0.0
May
Jun
Jul
Aug
Sep
Oct
Nov
Dec
Jan
2021
Feb
Mar
Apr

#### Venezuela, Bolivarian Republic of

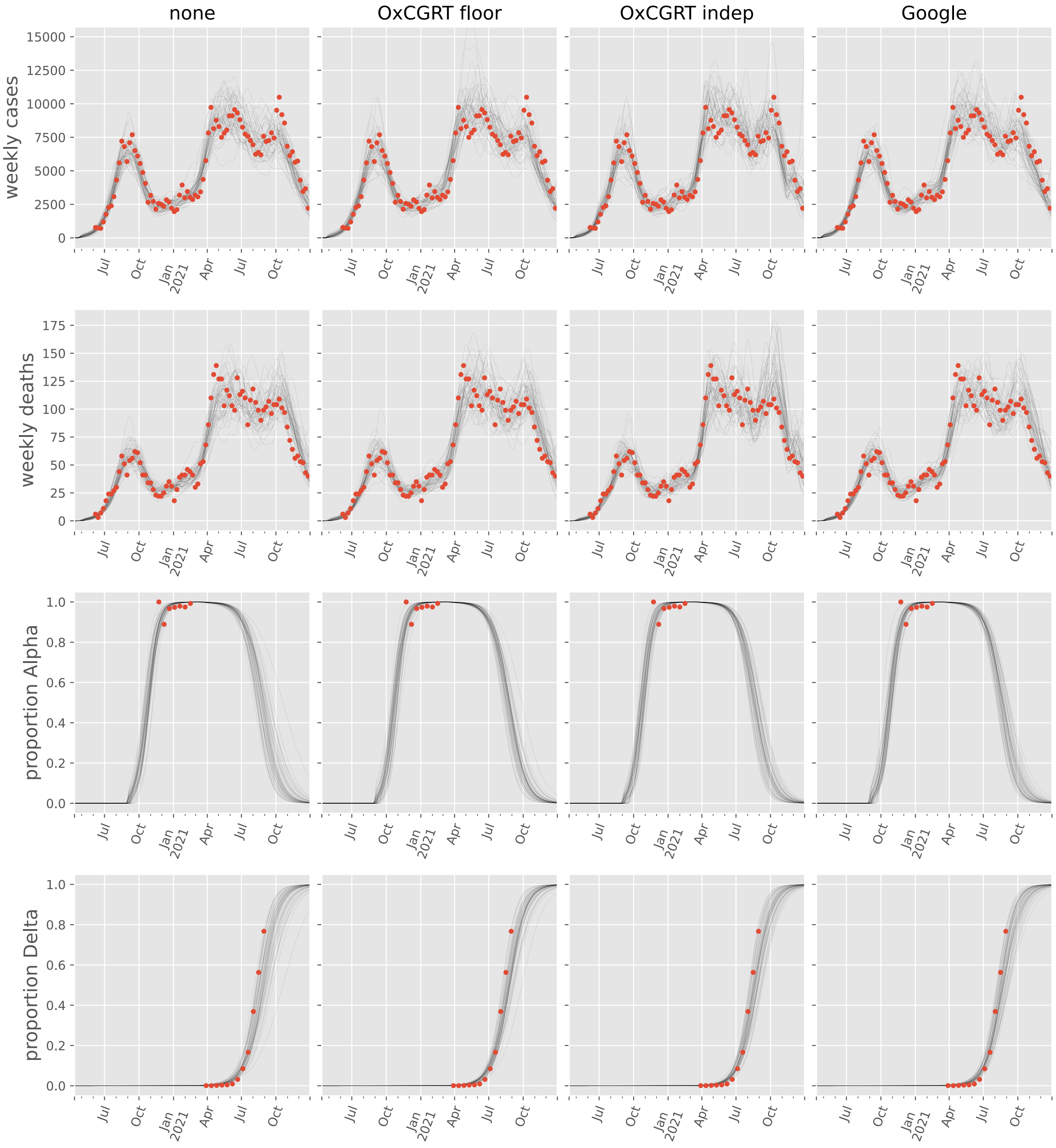

#### Commits used for analyses

For reproducibility, the following table gives the (short) commit SHA for each analysis.

| | no scaling | OxCGRT floored | OxCGRT independent | Google mobility | Facebook tiles visited mobility | Facebook single tile mobility |
|---|---|---|---|---|---|---|
| Afghanistan | 4f57571 | b934196 | b934196 | b934196 | no analysis | no analysis |
| Albania | 4f57571 | b934196 | b934196 | no analysis | b934196 | b934196 |
| Algeria | 4f57571 | b934196 | b934196 | no analysis | b934196 | b934196 |
| Andorra | 4f57571 | b934196 | b934196 | no analysis | no analysis | no analysis |
| Angola | 4f57571 | b934196 | b934196 | cb8324a | cb8324a | cb8324a |
| Argentina | 4f57571 | b934196 | b934196 | b934196 | b934196 | b934196 |
| Aruba | 4f57571 | b934196 | b934196 | 4f57571 | no analysis | no analysis |
| Australia | b934196 | b934196 | b934196 | no analysis | no analysis | no analysis |
| Austria | 4f57571 | b934196 | b934196 | b934196 | b934196 | b934196 |
| Azerbaijan | 4f57571 | b934196 | b934196 | no analysis | no analysis | no analysis |
| Bahamas | 4f57571 | b934196 | b934196 | b934196 | no analysis | no analysis |
| Bahrain | 4f57571 | b934196 | b934196 | 4f57571 | no analysis | no analysis |
| Bangladesh | 4f57571 | b934196 | b934196 | b934196 | b934196 | b934196 |
| Belarus | 4f57571 | b934196 | b934196 | cb8324a | cb8324a | b934196 |
| Belgium | 4f57571 | b934196 | b934196 | 4f57571 | 4f57571 | 4f57571 |
| Benin | 4f57571 | b934196 | b934196 | 4f57571 | 4f57571 | b934196 |
| Bosnia and Herzegovina | 4f57571 | b934196 | b934196 | b934196 | b934196 | b934196 |
| Brazil | 4f57571 | b934196 | b934196 | 4f57571 | 4f57571 | 4f57571 |
| Bulgaria | 4f57571 | b934196 | b934196 | b934196 | b934196 | b934196 |
| Burkina Faso | 4f57571 | b934196 | b934196 | 4f57571 | 4f57571 | 4f57571 |
| Cabo Verde | 4f57571 | b934196 | b934196 | 4f57571 | no analysis | no analysis |
| Cameroon | 4f57571 | b934196 | b934196 | b934196 | b934196 | b934196 |
| Canada | 4f57571 | b934196 | b934196 | b934196 | b934196 | b934196 |
| Chile | 4f57571 | b934196 | b934196 | 4f57571 | b934196 | b934196 |
| Colombia | 4f57571 | b934196 | b934196 | 4f57571 | 4f57571 | b934196 |
| Congo | 4f57571 | b934196 | b934196 | no analysis | 4f57571 | b934196 |
| Costa Rica | 4f57571 | b934196 | b934196 | 4f57571 | b934196 | b934196 |
| Croatia | 4f57571 | b934196 | b934196 | b934196 | b934196 | b934196 |

| | no scaling | OxCGRT floored | OxCGRT independent | Google mobility | Facebook tiles visited mobility | Facebook single tile mobility |
|---|---|---|---|---|---|---|
| Cuba | 4f57571 | b934196 | b934196 | no analysis | no analysis | no analysis |
| Cyprus | 4f57571 | b934196 | b934196 | no analysis | no analysis | no analysis |
| Czechia | 4f57571 | b934196 | b934196 | 4f57571 | b934196 | b934196 |
| Côte d'Ivoire | 4f57571 | b934196 | b934196 | b934196 | b934196 | b934196 |
| Denmark | 4f57571 | b934196 | b934196 | 4f57571 | 4f57571 | 4f57571 |
| Dominican Republic | 4f57571 | b934196 | b934196 | 4f57571 | b934196 | b934196 |
| Egypt | 4f57571 | b934196 | b934196 | b934196 | b934196 | b934196 |
| El Salvador | 4f57571 | 4c513a7 | 4c513a7 | 4f57571 | 4f57571 | 4f57571 |
| Estonia | 4f57571 | b934196 | b934196 | 4f57571 | 4f57571 | 4f57571 |
| Eswatini | 4f57571 | 4c513a7 | missing git information | no analysis | no analysis | no analysis |
| Ethiopia | 4f57571 | b934196 | b934196 | no analysis | b934196 | b934196 |
| Finland | 4f57571 | b934196 | b934196 | b934196 | b934196 | b934196 |
| France | 4f57571 | b934196 | b934196 | b934196 | b934196 | b934196 |
| Gabon | 4f57571 | b934196 | b934196 | b934196 | b934196 | b934196 |
| Georgia | 4f57571 | b934196 | b934196 | 4f57571 | b934196 | b934196 |
| Germany | 4f57571 | b934196 | b934196 | b934196 | b934196 | b934196 |
| Greece | 4f57571 | b934196 | b934196 | 4f57571 | 4f57571 | b934196 |
| Guinea | 4f57571 | 4c513a7 | b934196 | no analysis | b934196 | b934196 |
| Guyana | 4f57571 | b934196 | b934196 | no analysis | 4f57571 | 4f57571 |
| Haiti | 4f57571 | b934196 | 4c513a7 | b934196 | b934196 | b934196 |
| Honduras | 4f57571 | b934196 | b934196 | b934196 | b934196 | b934196 |
| Hungary | 4f57571 | b934196 | b934196 | 4f57571 | 4f57571 | b934196 |
| India | 4f57571 | b934196 | b934196 | b934196 | b934196 | b934196 |
| Indonesia | 4f57571 | b934196 | b934196 | b934196 | b934196 | b934196 |
| Iran, Islamic Republic of | 4f57571 | b934196 | b934196 | no analysis | no analysis | no analysis |
| Iraq | 4f57571 | b934196 | b934196 | 4f57571 | 4f57571 | 4f57571 |
| Ireland | 4f57571 | b934196 | b934196 | b934196 | b934196 | b934196 |
| Israel | 4f57571 | b934196 | b934196 | b934196 | no analysis | no analysis |
| Italy | 4f57571 | b934196 | b934196 | 4f57571 | 4f57571 | b934196 |
| Jamaica | 4f57571 | b934196 | cb8324a | b934196 | b934196 | b934196 |

| | no scaling | OxCGRT floored | OxCGRT independent | Google mobility | Facebook tiles visited mobility | Facebook single tile mobility |
|---|---|---|---|---|---|---|
| Japan | cb8324a | cb8324a | cb8324a | cb8324a | b934196 | cb8324a |
| Jordan | 4f57571 | b934196 | b934196 | 4f57571 | 4f57571 | 4f57571 |
| Kenya | 4f57571 | b934196 | b934196 | b934196 | b934196 | b934196 |
| Korea, Republic of | 4f57571 | b934196 | cb8324a | b934196 | b934196 | cb8324a |
| Kosovo | 4f57571 | 4c513a7 | 4c513a7 | no analysis | no analysis | no analysis |
| Kuwait | 4f57571 | 4c513a7 | 4c513a7 | 4f57571 | no analysis | no analysis |
| Latvia | 4f57571 | 4c513a7 | 4c513a7 | 4f57571 | 4f57571 | 4f57571 |
| Lebanon | 4f57571 | 4c513a7 | b934196 | 4f57571 | 4f57571 | 4f57571 |
| Lesotho | 4f57571 | b934196 | 4c513a7 | no analysis | 4f57571 | 4f57571 |
| Liberia | 4f57571 | 4c513a7 | 4c513a7 | no analysis | 4f57571 | 4f57571 |
| Libya | cb8324a | b934196 | cb8324a | cb8324a | no analysis | no analysis |
| Lithuania | 4f57571 | 4c513a7 | 4c513a7 | 4f57571 | 4f57571 | 4f57571 |
| Luxembourg | 4f57571 | 4c513a7 | 4c513a7 | 4f57571 | cb8324a | cb8324a |
| Madagascar | 4f57571 | 4c513a7 | 4c513a7 | no analysis | 4f57571 | cb8324a |
| Malawi | 4f57571 | 4c513a7 | cb8324a | no analysis | cb8324a | cb8324a |
| Malaysia | 4f57571 | 1cab74a | 4c513a7 | 4f57571 | 4f57571 | 4f57571 |
| Mali | 4f57571 | 4c513a7 | 4c513a7 | 4f57571 | 4f57571 | 4f57571 |
| Malta | cb8324a | cb8324a | cb8324a | cb8324a | cb8324a | cb8324a |
| Mexico | 4f57571 | 4c513a7 | 4c513a7 | 4f57571 | 4f57571 | 4f57571 |
| Moldova, Republic of | 4f57571 | 4c513a7 | 4c513a7 | cb8324a | cb8324a | cb8324a |
| Morocco | 4f57571 | 4c513a7 | 4c513a7 | 4f57571 | no analysis | no analysis |
| Mozambique | 4f57571 | 4c513a7 | 4c513a7 | 4f57571 | cb8324a | cb8324a |
| Nepal | 4f57571 | 4c513a7 | 4c513a7 | cb8324a | cb8324a | cb8324a |
| Netherlands | 4f57571 | 4c513a7 | 4c513a7 | 4f57571 | cb8324a | cb8324a |
| New Zealand | 4c513a7 | 4c513a7 | 4c513a7 | no analysis | no analysis | no analysis |
| Nigeria | 4f57571 | 4c513a7 | 4c513a7 | 4f57571 | 4f57571 | 4f57571 |
| Norway | 4f57571 | 4c513a7 | 4c513a7 | cb8324a | cb8324a | cb8324a |
| Oman | 4f57571 | 4c513a7 | 4c513a7 | 4f57571 | 4f57571 | 4f57571 |
| Pakistan | 4f57571 | 4c513a7 | cb8324a | cb8324a | no analysis | no analysis |
| Palestine, State of | cb8324a | cb8324a | cb8324a | no analysis | no analysis | no analysis |

| | no scaling | OxCGRT floored | OxCGRT independent | Google mobility | Facebook tiles visited mobility | Facebook single tile mobility |
|---|---|---|---|---|---|---|
| Panama | 4f57571 | 4c513a7 | 4c513a7 | 4f57571 | cb8324a | cb8324a |
| Paraguay | 4f57571 | 4c513a7 | cb8324a | cb8324a | cb8324a | cb8324a |
| Peru | 4f57571 | 4c513a7 | 4c513a7 | 4f57571 | 4f57571 | 4f57571 |
| Philippines | 4f57571 | 4c513a7 | cb8324a | cb8324a | cb8324a | cb8324a |
| Poland | 4f57571 | 4c513a7 | 4c513a7 | 4f57571 | 4f57571 | cb8324a |
| Portugal | 4f57571 | 4c513a7 | 4c513a7 | cb8324a | cb8324a | cb8324a |
| Qatar | 4f57571 | 4c513a7 | 4c513a7 | 4f57571 | 4f57571 | 4f57571 |
| Romania | 4f57571 | 4c513a7 | 4c513a7 | 4f57571 | cb8324a | cb8324a |
| Russian Federation | 4f57571 | 4c513a7 | cb8324a | cb8324a | no analysis | no analysis |
| Rwanda | 4f57571 | 4c513a7 | 4c513a7 | 4f57571 | cb8324a | cb8324a |
| Saudi Arabia | 4f57571 | 4c513a7 | 4c513a7 | 4f57571 | 4f57571 | cb8324a |
| Senegal | 4f57571 | 4c513a7 | 4c513a7 | 4f57571 | 4f57571 | 4f57571 |
| Serbia | 4f57571 | 4c513a7 | 4c513a7 | cb8324a | cb8324a | cb8324a |
| Singapore | 4c513a7 | cb8324a | 4c513a7 | no analysis | no analysis | no analysis |
| Slovakia | cb8324a | cb8324a | cb8324a | cb8324a | cb8324a | cb8324a |
| Slovenia | cb8324a | cb8324a | cb8324a | cb8324a | cb8324a | cb8324a |
| Somalia | 4f57571 | 4c513a7 | 4c513a7 | no analysis | no analysis | no analysis |
| South Africa | 4f57571 | 4c513a7 | 4c513a7 | 4f57571 | 4f57571 | cb8324a |
| South Sudan | 4f57571 | cb8324a | cb8324a | no analysis | no analysis | no analysis |
| Spain | cb8324a | b934196 | b934196 | b934196 | b934196 | b934196 |
| Sri Lanka | cb8324a | cb8324a | cb8324a | cb8324a | cb8324a | b934196 |
| Sudan | 4f57571 | 4c513a7 | 4c513a7 | no analysis | no analysis | no analysis |
| Suriname | 4f57571 | 4c513a7 | 4c513a7 | no analysis | 4f57571 | 4f57571 |
| Sweden | 4f57571 | 4c513a7 | 4c513a7 | 4f57571 | 4f57571 | 4f57571 |
| Switzerland | 4f57571 | b934196 | b934196 | 4f57571 | b934196 | b934196 |
| Syrian Arab Republic | 4f57571 | cb8324a | cb8324a | no analysis | no analysis | no analysis |
| Togo | 4f57571 | 4c513a7 | 4c513a7 | 4f57571 | 4f57571 | 4f57571 |
| Tunisia | 4f57571 | 4c513a7 | 4c513a7 | no analysis | 4f57571 | 4f57571 |
| Türkiye | cb8324a | cb8324a | cb8324a | cb8324a | cb8324a | cb8324a |

| | no scaling | OxCGRT floored | OxCGRT independent | Google mobility | Facebook tiles visited mobility | Facebook single tile mobility |
|---|---|---|---|---|---|---|
| Ukraine | 4f57571 | missing git information | 4c513a7 | 4f57571 | no analysis | no analysis |
| United Arab Emirates | 4f57571 | b934196 | b934196 | 4f57571 | 4f57571 | 4f57571 |
| United Kingdom | 4f57571 | b934196 | b934196 | 4f57571 | b934196 | b934196 |
| United States | 4f57571 | 4c513a7 | 4c513a7 | cb8324a | cb8324a | cb8324a |
| Uruguay | 4f57571 | 4c513a7 | 4c513a7 | 4f57571 | 4c513a7 | 4f57571 |
| Venezuela, Bolivarian Republic of | 4f57571 | 4c513a7 | cb8324a | cb8324a | no analysis | no analysis |
| Yemen | 4f57571 | 4c513a7 | 4c513a7 | 4f57571 | no analysis | no analysis |
| Zambia | cb8324a | cb8324a | cb8324a | cb8324a | cb8324a | cb8324a |
| Zimbabwe | 4f57571 | 4c513a7 | 4c513a7 | 4f57571 | cb8324a | cb8324a |
