## Supplemental File 3 for "Quantifying the contribution of policy measures to COVID-19 dynamics across 126 countries"

### Prior-posterior comparisons

#### Table of contents

**Purpose** 4

**Prior posterior comparisons by continent and country** 5

Africa 5

Algeria 5
Angola 6
Benin 7
Burkina Faso 8
Cabo Verde 9
Cameroon 10
Congo 11
Côte d'Ivoire 12
Egypt 13
Eswatini 14
Ethiopia 15
Gabon 16
Guinea 17
Kenya 18
Lesotho 19
Liberia 20
Libya 21
Madagascar 22
Malawi 23
Mali 24
Morocco 25
Mozambique 26
Nigeria 27
Rwanda 28
Senegal 29

**Commits used for analyses** **131**

#### Purpose

This document presents comparisons of parameter prior distributions to their corresponding posterior distributions. Only parameters used by the "no scaling" baseline run are presented here, with the parameters specifically pertaining to the additional scaling of transmission to capture the effects of policy or mobility effects presented in later documents.

### Prior posterior comparisons by continent and country

#### Africa

##### Algeria

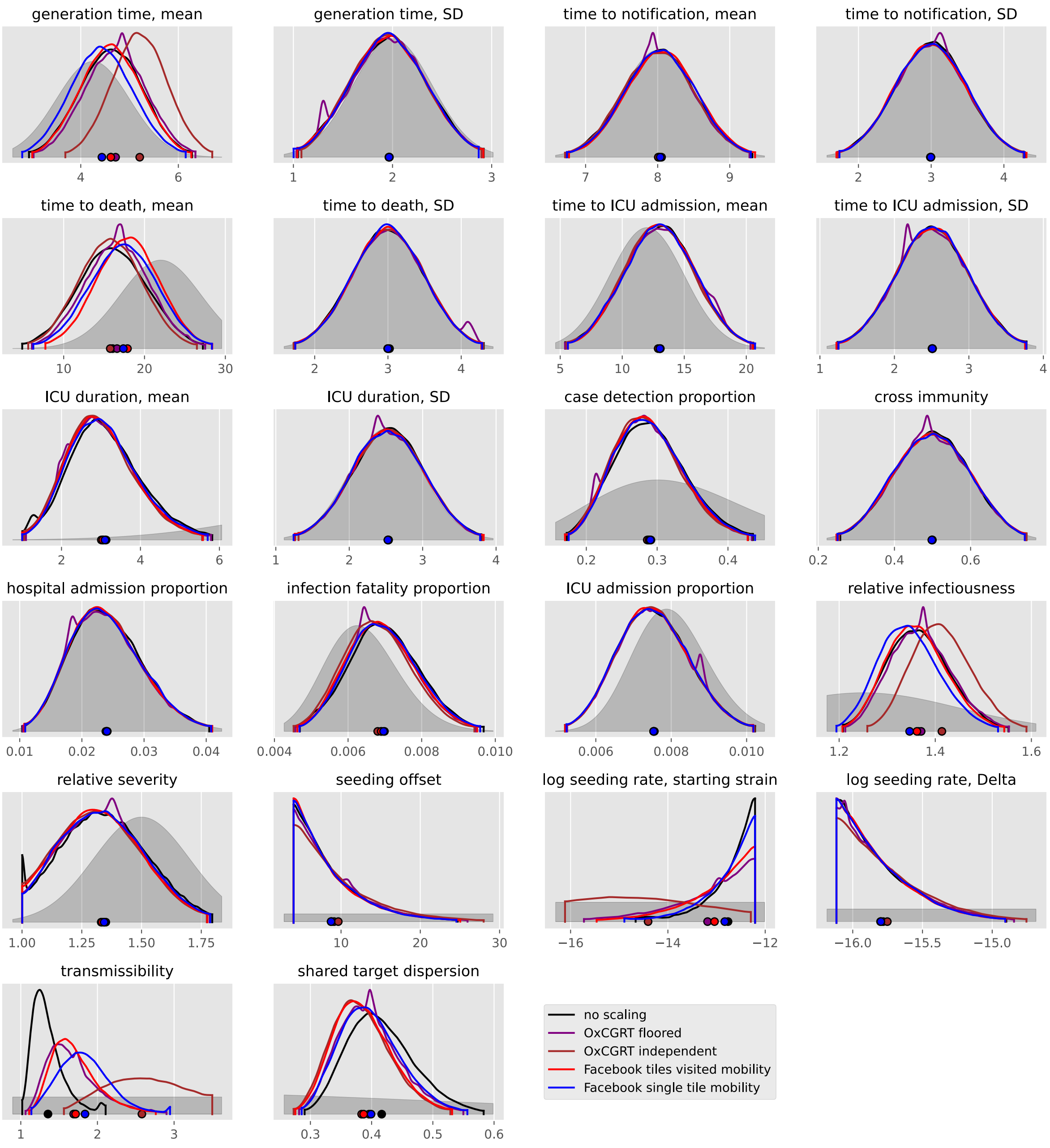

#### Angola

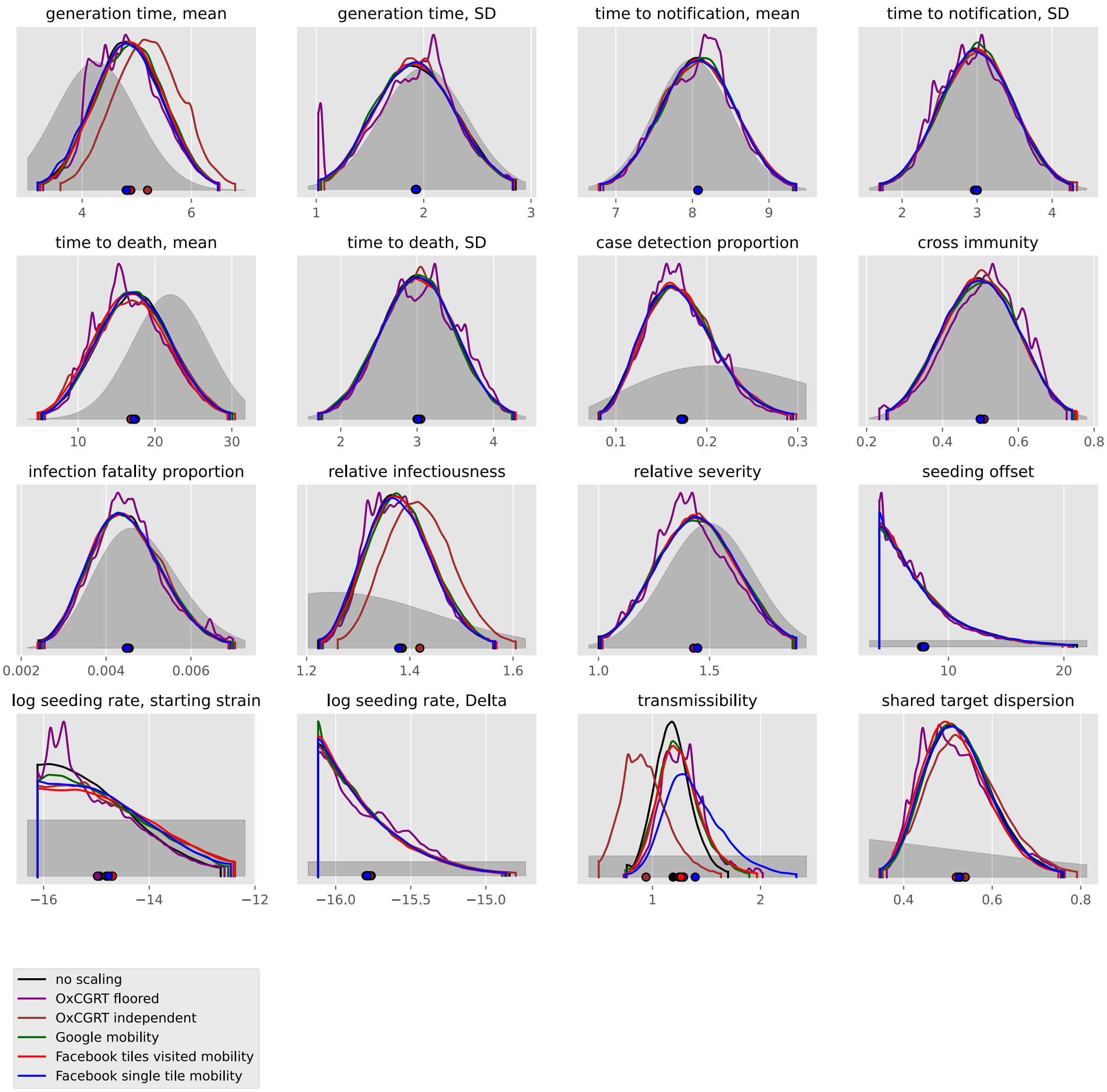
generation time, mean
generation time, SD
time to notification, mean
time to notification, SD
time to death, mean
time to death, SD
case detection proportion
cross immunity
infection fatality proportion
relative infectiousness
relative severity
seeding offset
log seeding rate, starting strain
log seeding rate, Delta
transmissibility
shared target dispersion
no scaling
OxCGRT floored
OxCGRT independent
Google mobility
Facebook tiles visited mobility
Facebook single tile mobility

#### Benin

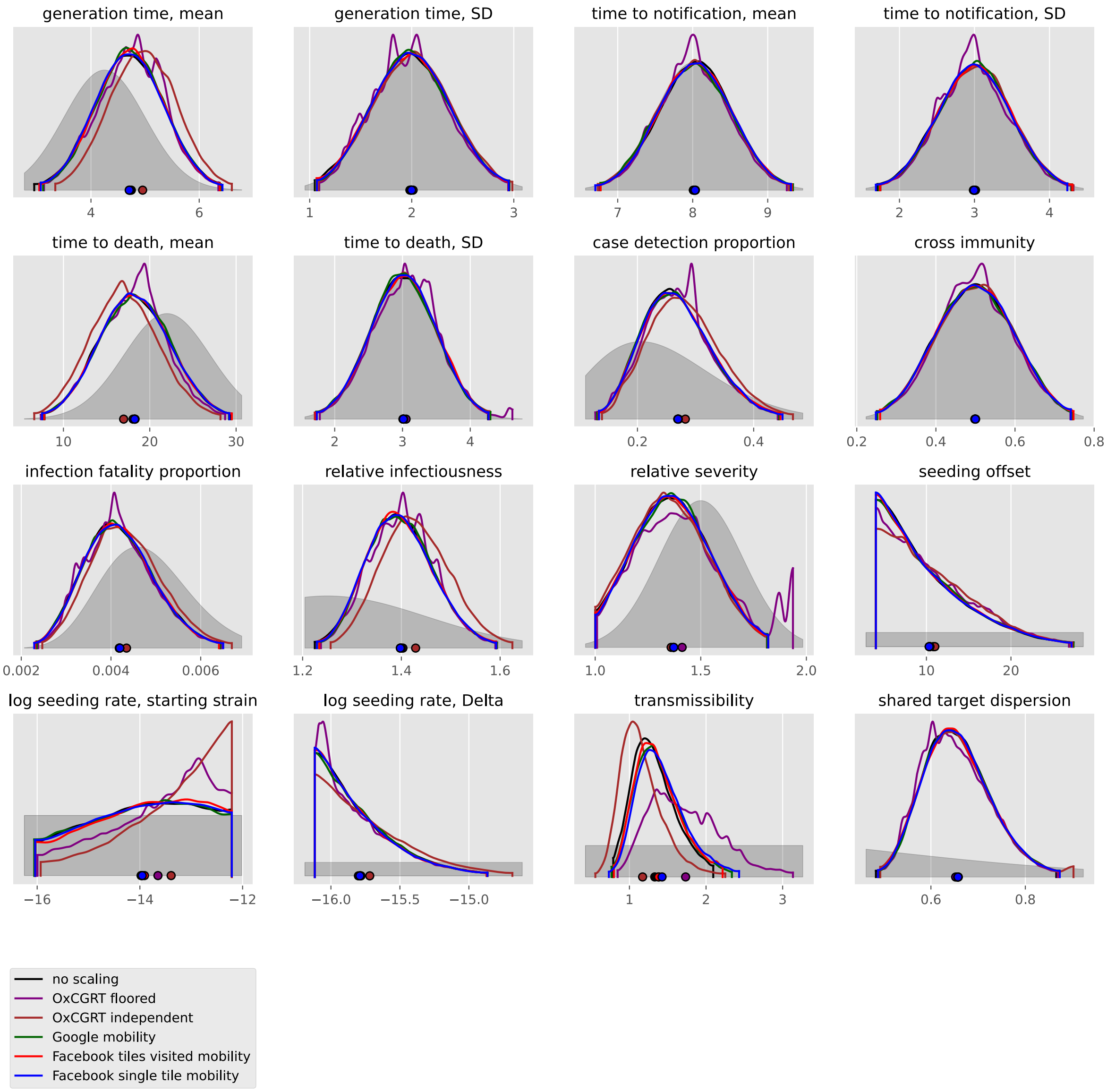
generation time, mean
generation time, SD
time to notification, mean
time to notification, SD
time to death, mean
time to death, SD
case detection proportion
cross immunity
infection fatality proportion
relative infectiousness
relative severity
seeding offset
log seeding rate, starting strain
log seeding rate, Delta
transmissibility
shared target dispersion
no scaling
OxCGRT floored
OxCGRT independent
Google mobility
Facebook tiles visited mobility
Facebook single tile mobility

#### Burkina Faso

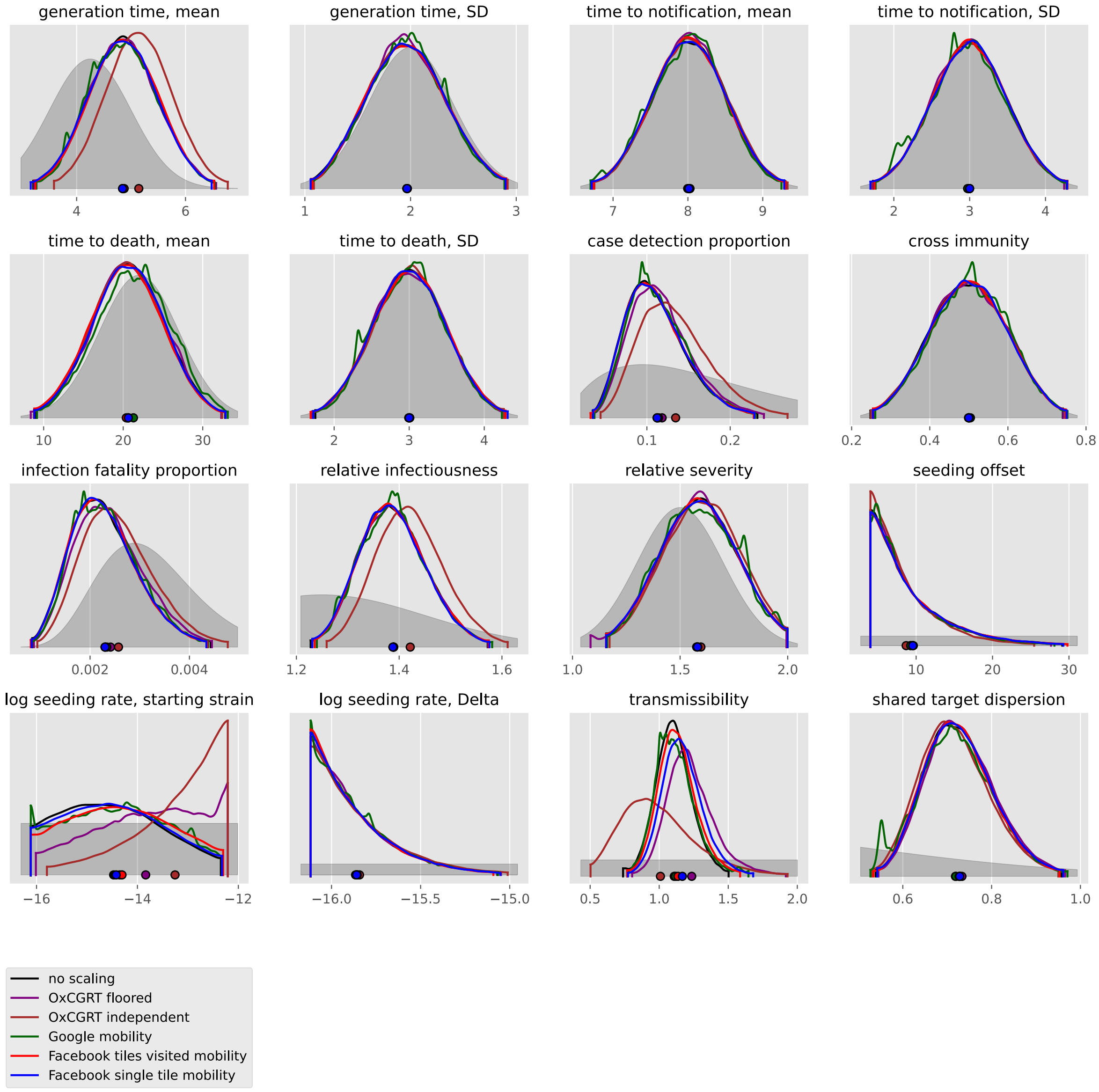

#### Cabo Verde

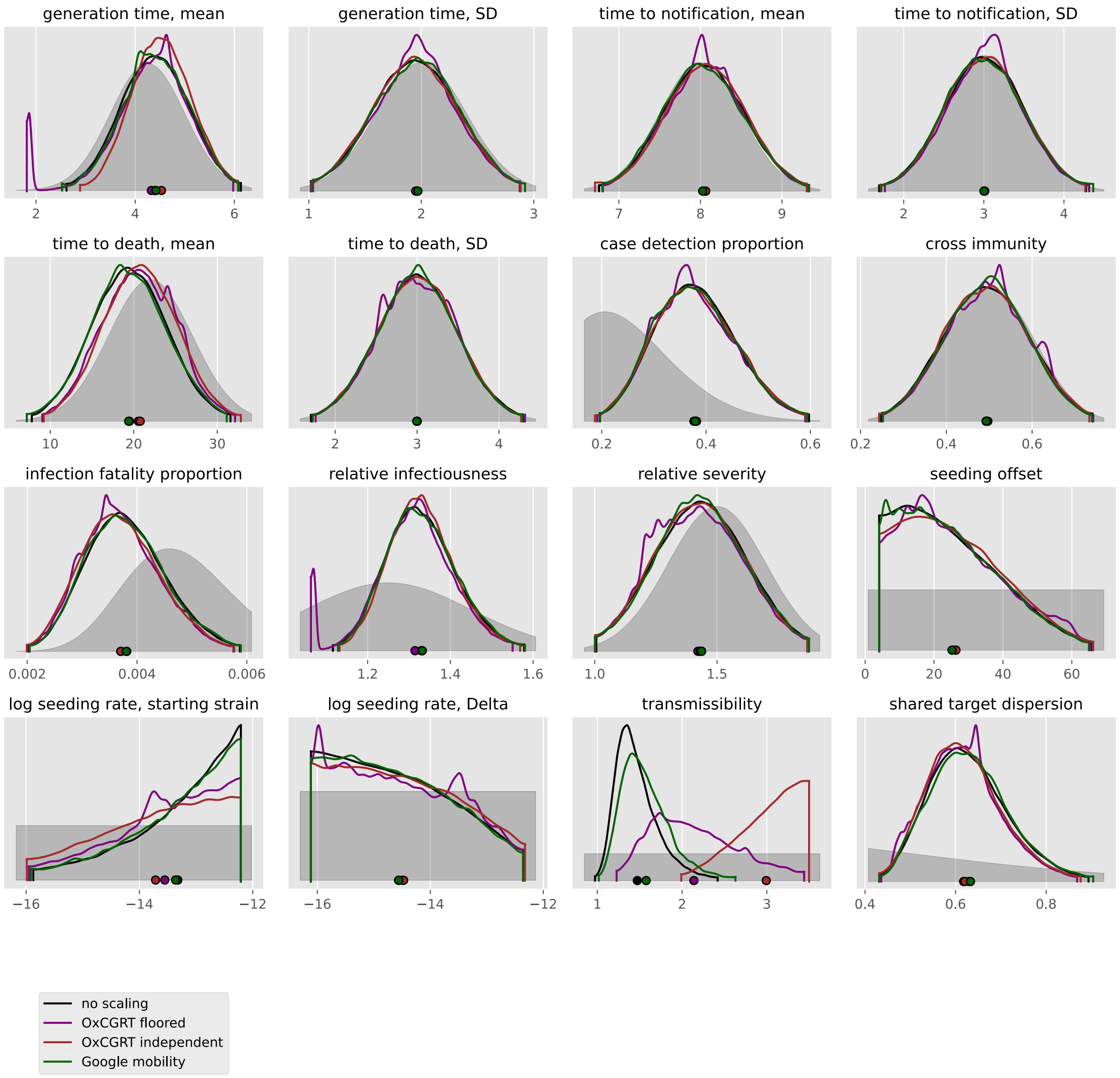
generation time, mean
generation time, SD
time to notification, mean
time to notification, SD
time to death, mean
time to death, SD
case detection proportion
cross immunity
infection fatality proportion
relative infectiousness
relative severity
seeding offset
log seeding rate, starting strain
log seeding rate, Delta
transmissibility
shared target dispersion
no scaling
OxCGRT floored
OxCGRT independent
Google mobility

#### Cameroon

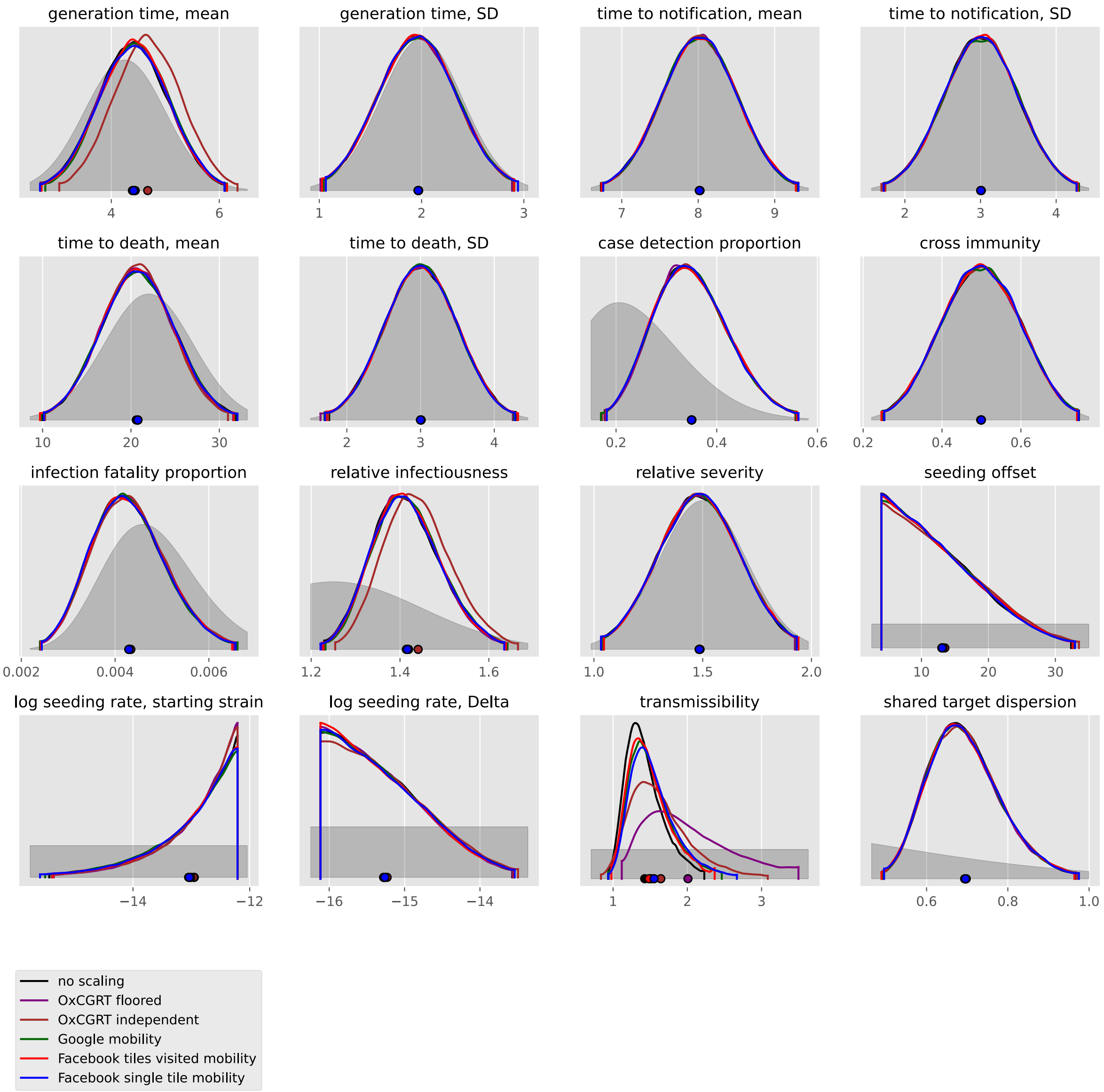

generation time, mean
4
6
generation time, SD
1
2
3
time to notification, mean
7
8
9
time to notification, SD
2
3
4
time to death, mean
10
20
30
time to death, SD
2
3
4
case detection proportion
0.2
0.4
0.6
cross immunity
0.2
0.4
0.6
infection fatality proportion
0.002
0.004
0.006
relative infectiousness
1.2
1.4
1.6
relative severity
1.0
1.5
2.0
seeding offset
10
20
30
log seeding rate, starting strain
−14
−12
log seeding rate, Delta
−16
−15
−14
transmissibility
1
2
3
shared target dispersion
0.6
0.8
1.0
no scaling
OxCGRT floored
OxCGRT independent
Google mobility
Facebook tiles visited mobility
Facebook single tile mobility

#### Congo

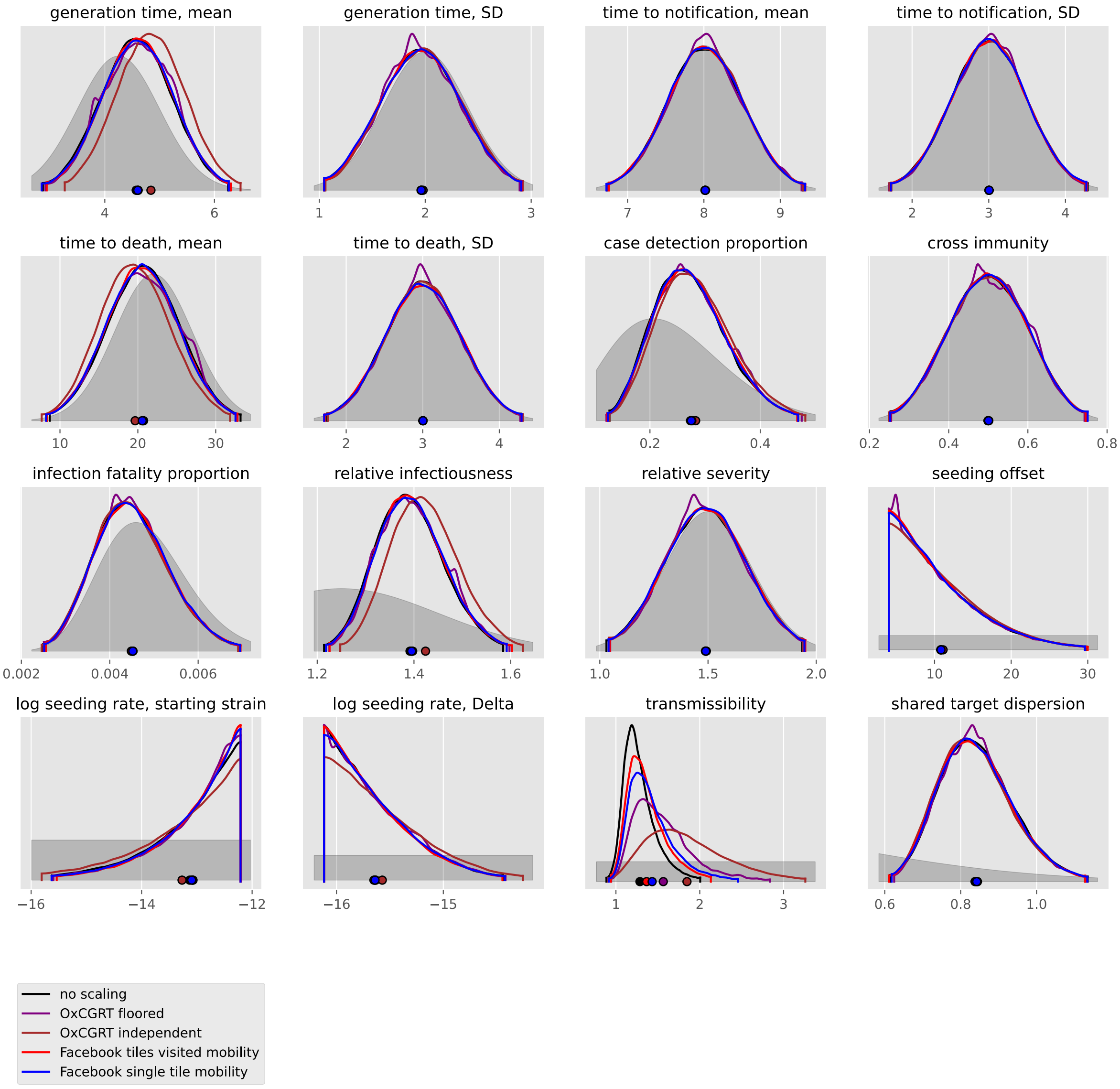
generation time, mean
generation time, SD
time to notification, mean
time to notification, SD
time to death, mean
time to death, SD
case detection proportion
cross immunity
infection fatality proportion
relative infectiousness
relative severity
seeding offset
log seeding rate, starting strain
log seeding rate, Delta
transmissibility
shared target dispersion
no scaling
OxCGRT floored
OxCGRT independent
Facebook tiles visited mobility
Facebook single tile mobility

#### Côte d'Ivoire

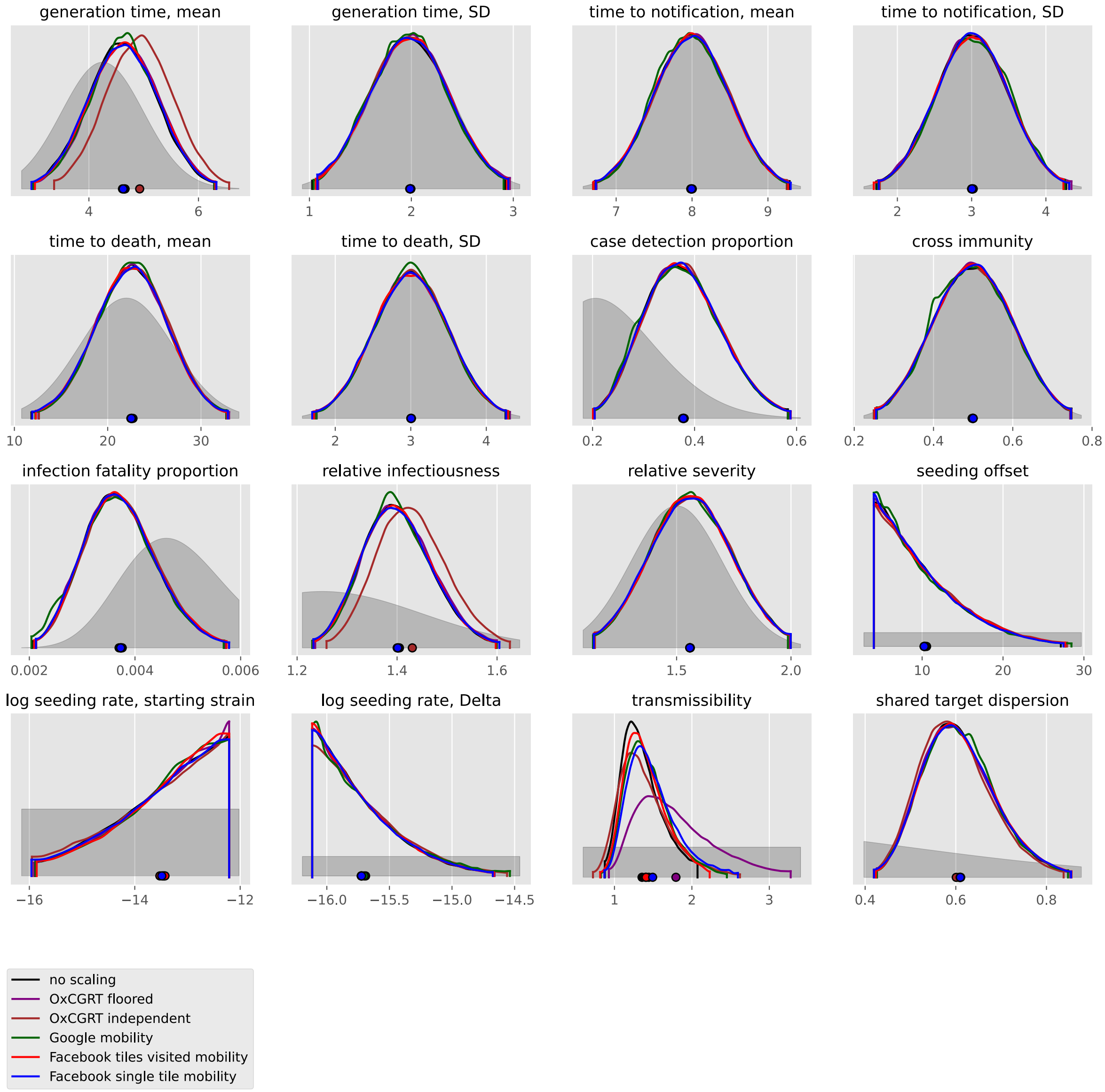

#### Egypt

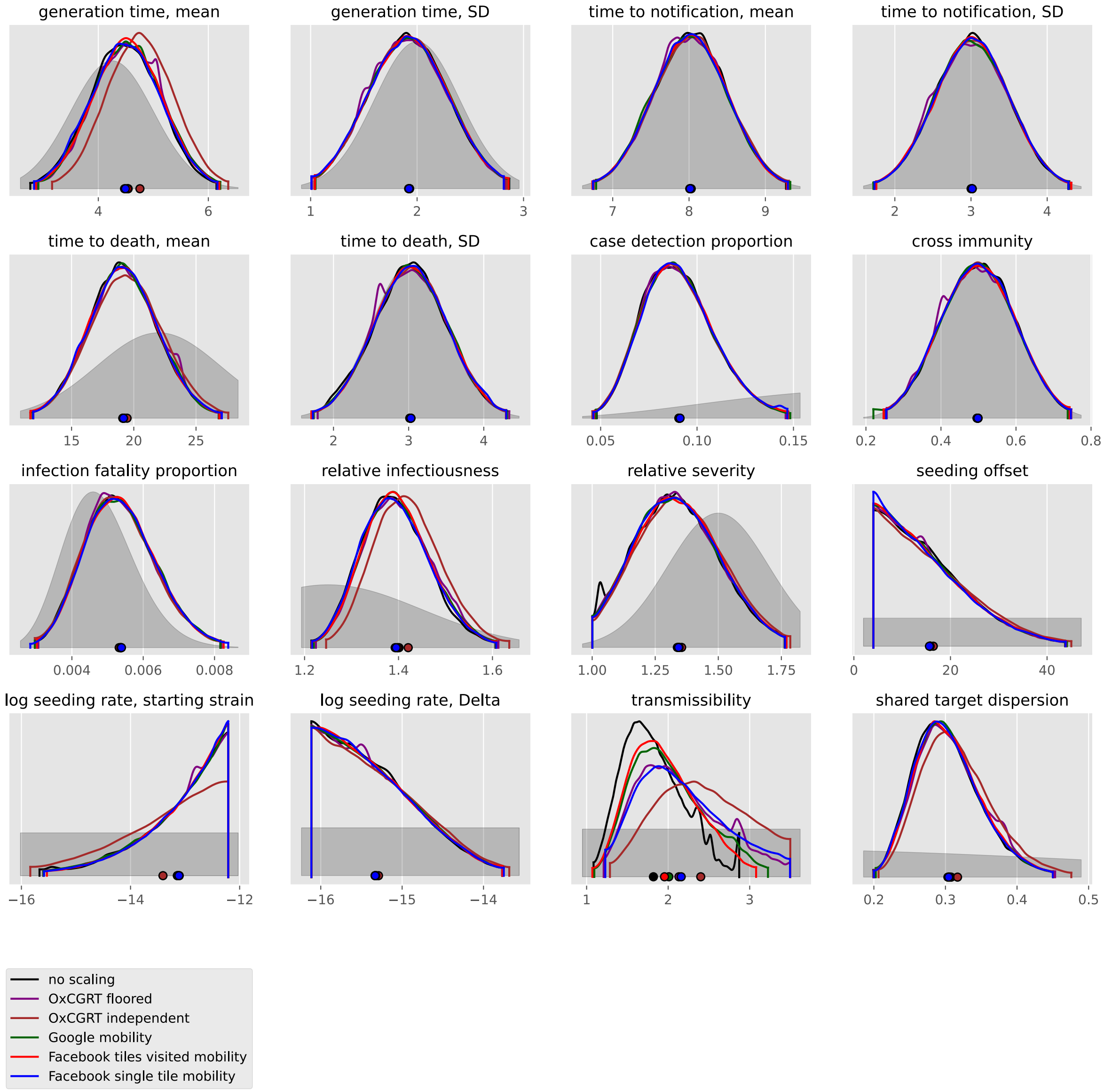

generation time, mean
4
6
generation time, SD
1
2
3
time to notification, mean
7
8
9
time to notification, SD
2
3
4
time to death, mean
15
20
25
time to death, SD
2
3
4
case detection proportion
0.05
0.10
0.15
cross immunity
0.2
0.4
0.6
0.8
infection fatality proportion
0.004
0.006
0.008
relative infectiousness
1.2
1.4
1.6
relative severity
1.00
1.25
1.50
1.75
seeding offset
0
20
40
log seeding rate, starting strain
−16
−14
−12
log seeding rate, Delta
−16
−15
−14
transmissibility
1
2
3
shared target dispersion
0.2
0.3
0.4
0.5
no scaling
OxCGRT floored
OxCGRT independent
Google mobility
Facebook tiles visited mobility
Facebook single tile mobility

#### Eswatini

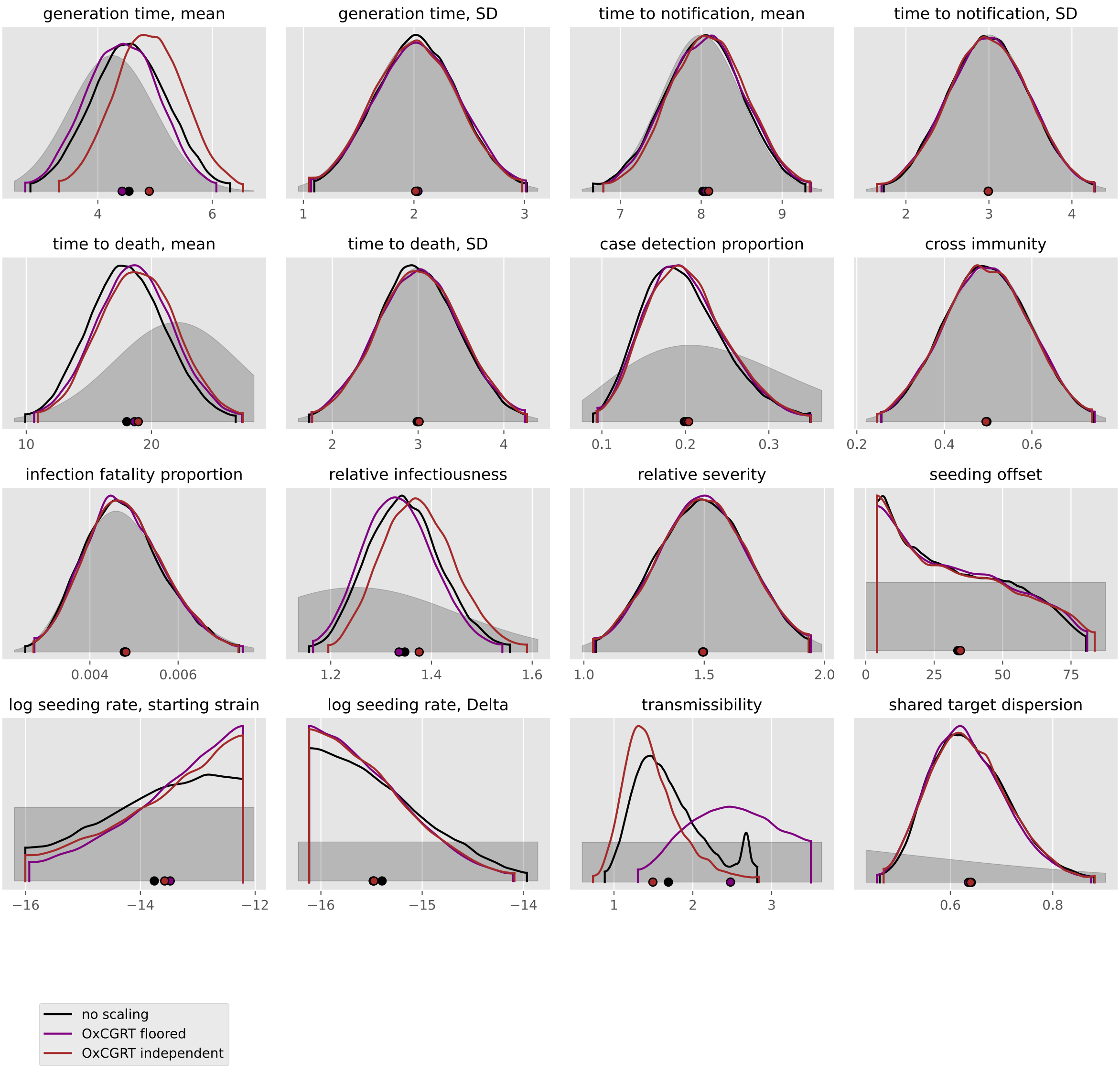

generation time, mean
4
6
generation time, SD
1
2
3
time to notification, mean
7
8
9
time to notification, SD
2
3
4
time to death, mean
10
20
time to death, SD
2
3
4
case detection proportion
0.1
0.2
0.3
cross immunity
0.2
0.4
0.6
infection fatality proportion
0.004
0.006
relative infectiousness
1.2
1.4
1.6
relative severity
1.0
1.5
2.0
seeding offset
0
25
50
75
log seeding rate, starting strain
−16
−14
−12
log seeding rate, Delta
−16
−15
−14
transmissibility
1
2
3
shared target dispersion
0.6
0.8
no scaling
OxCGRT floored
OxCGRT independent

### Ethiopia

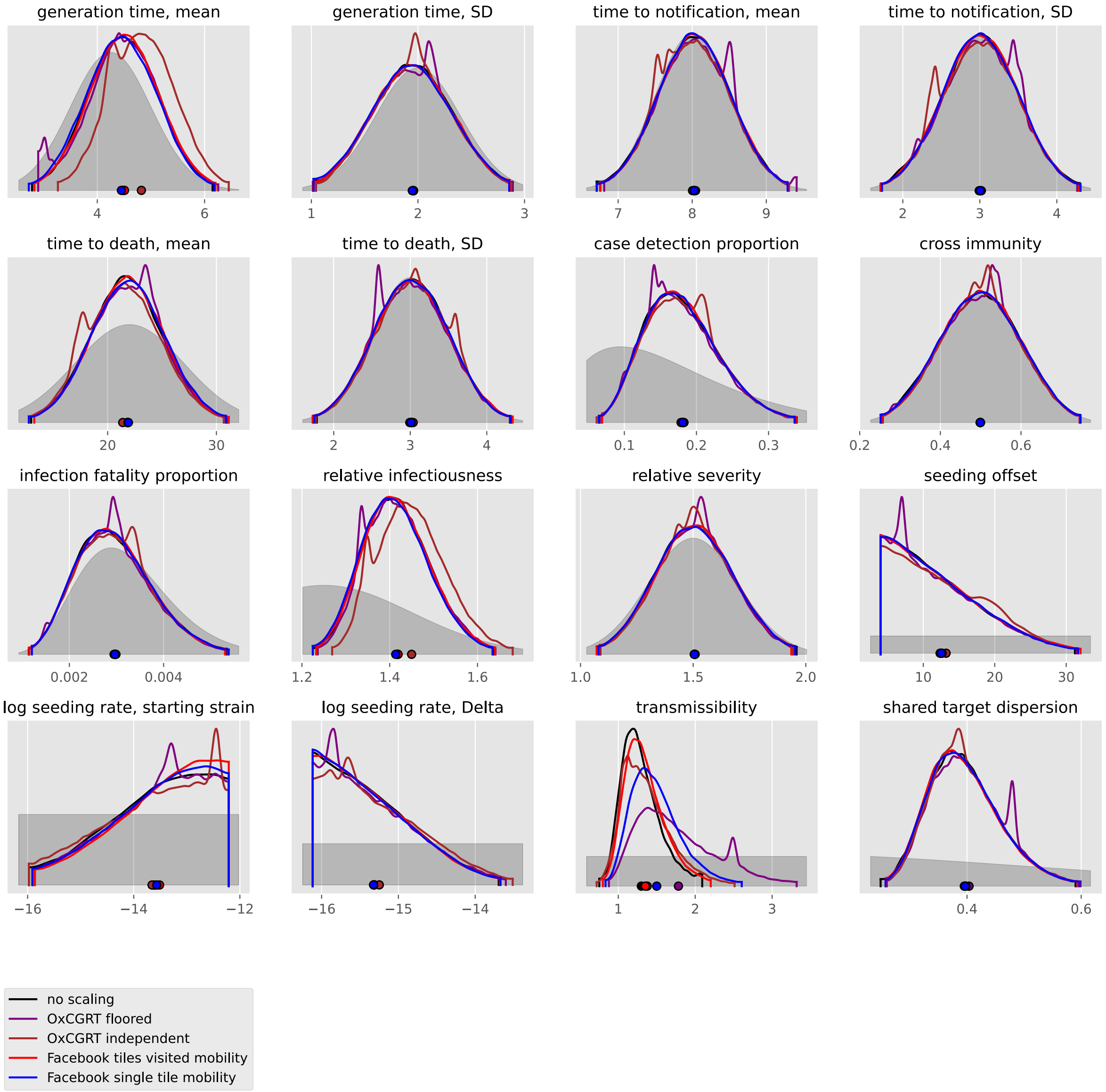
generation time, mean
generation time, SD
time to notification, mean
time to notification, SD
time to death, mean
time to death, SD
case detection proportion
cross immunity
infection fatality proportion
relative infectiousness
relative severity
seeding offset
log seeding rate, starting strain
log seeding rate, Delta
transmissibility
shared target dispersion
no scaling
OxCGRT floored
OxCGRT independent
Facebook tiles visited mobility
Facebook single tile mobility

#### Gabon

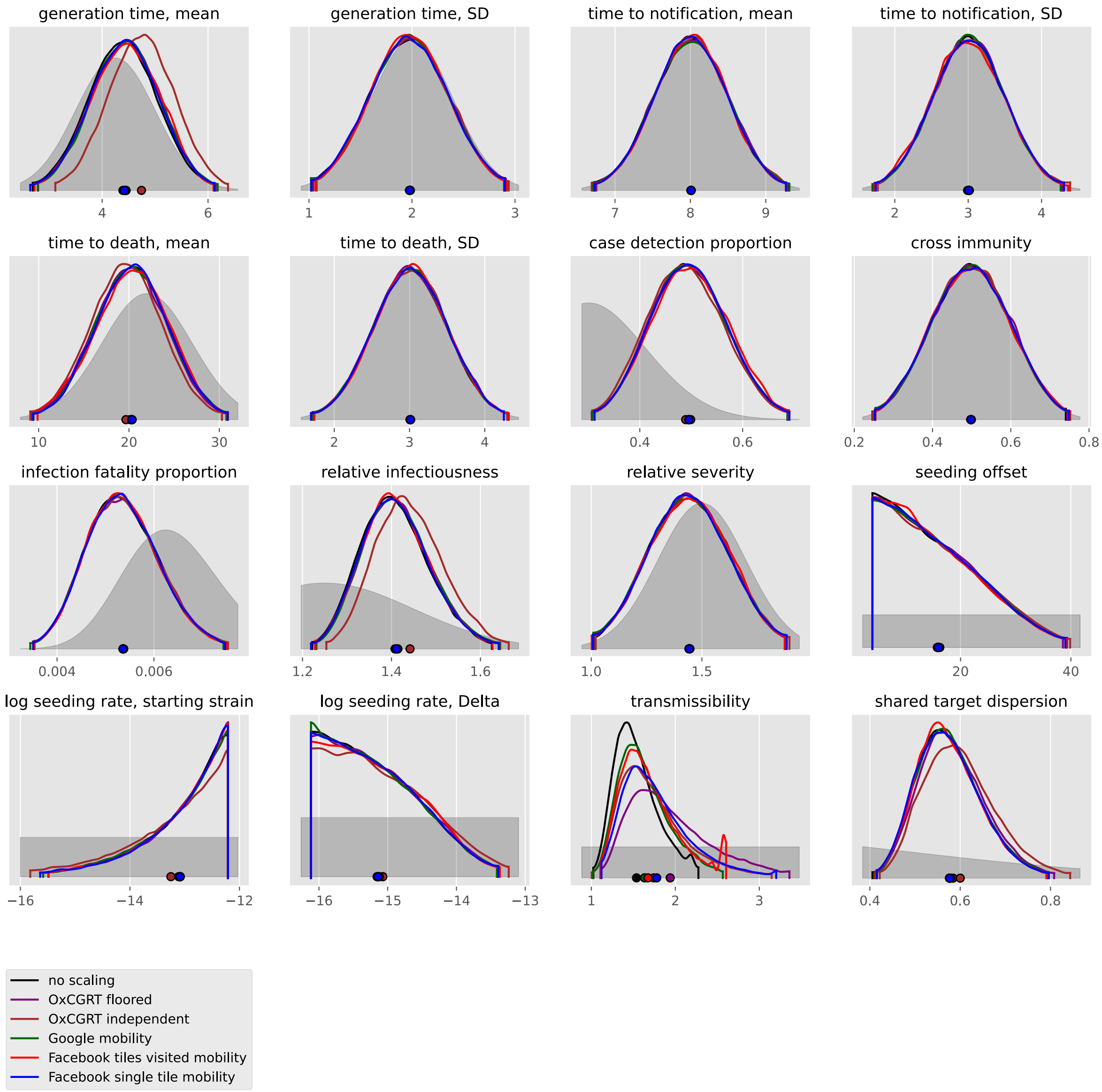

generation time, mean
generation time, SD
time to notification, mean
time to notification, SD
time to death, mean
time to death, SD
case detection proportion
cross immunity
infection fatality proportion
relative infectiousness
relative severity
seeding offset
log seeding rate, starting strain
log seeding rate, Delta
transmissibility
shared target dispersion
no scaling
OxCGRT floored
OxCGRT independent
Google mobility
Facebook tiles visited mobility
Facebook single tile mobility

#### Guinea

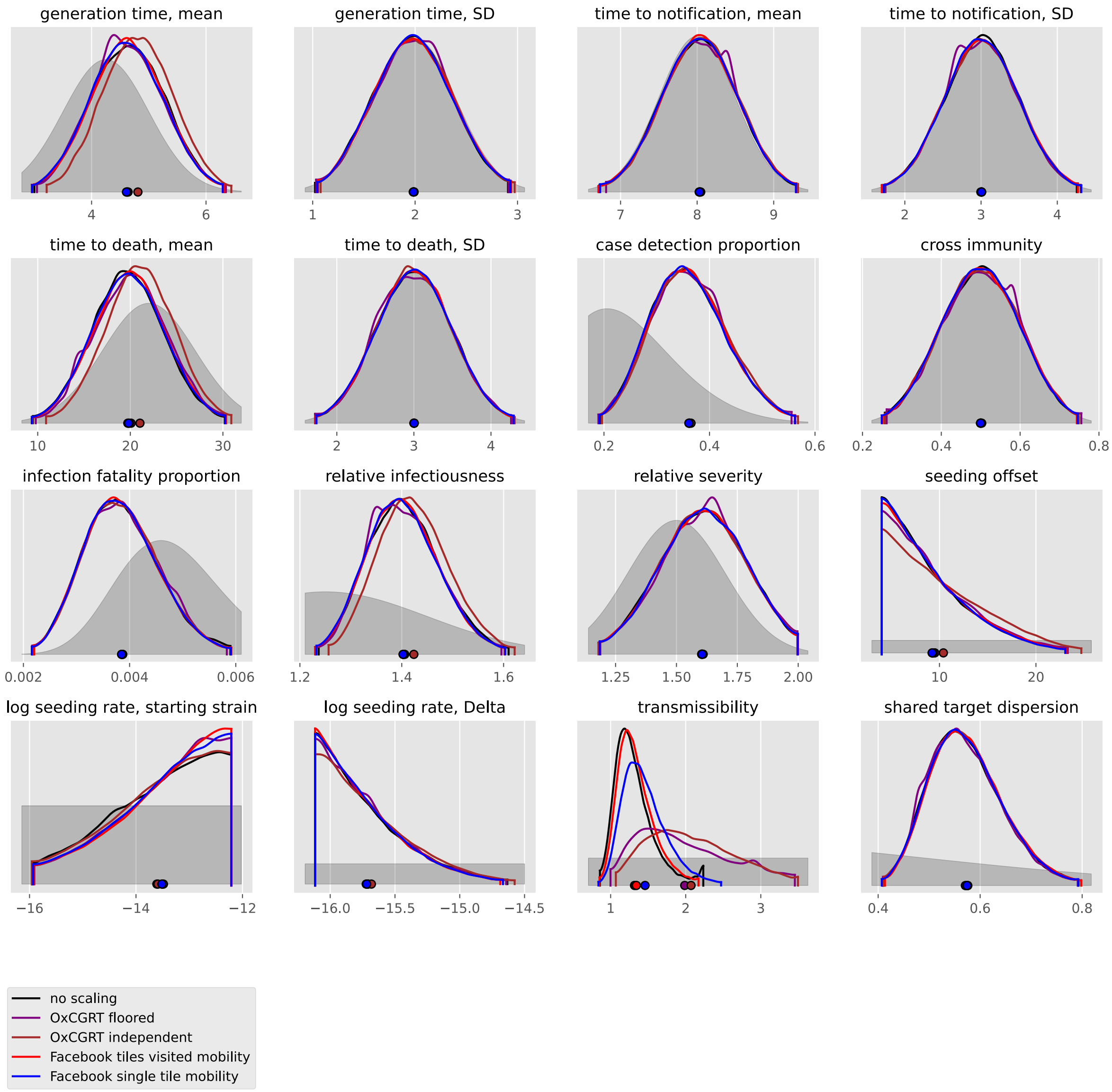
generation time, mean
generation time, SD
time to notification, mean
time to notification, SD
time to death, mean
time to death, SD
case detection proportion
cross immunity
infection fatality proportion
relative infectiousness
relative severity
seeding offset
log seeding rate, starting strain
log seeding rate, Delta
transmissibility
shared target dispersion
no scaling
OxCGRT floored
OxCGRT independent
Facebook tiles visited mobility
Facebook single tile mobility

#### Kenya

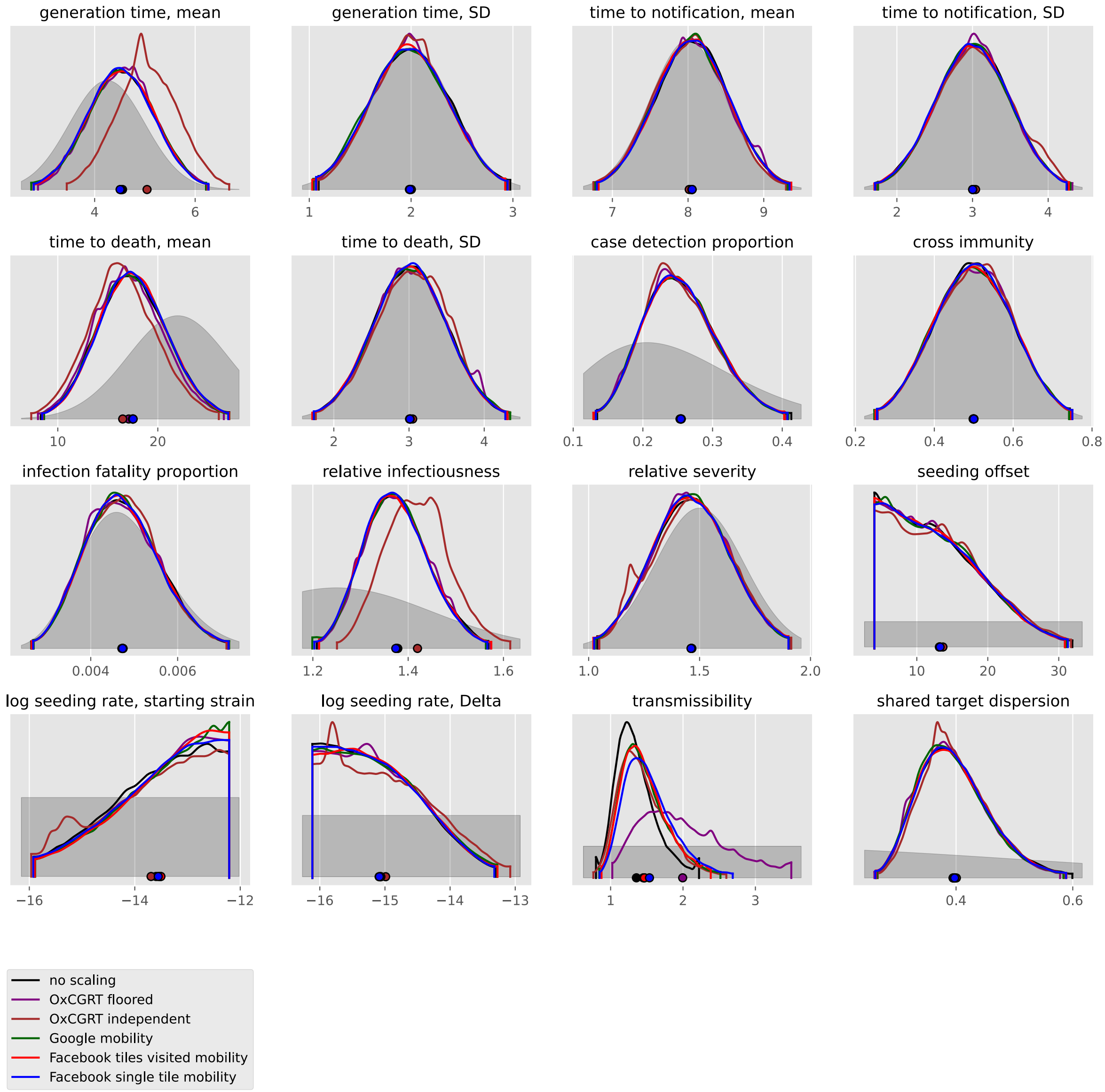

generation time, mean
generation time, SD
time to notification, mean
time to notification, SD
time to death, mean
time to death, SD
case detection proportion
cross immunity
infection fatality proportion
relative infectiousness
relative severity
seeding offset
log seeding rate, starting strain
log seeding rate, Delta
transmissibility
shared target dispersion
no scaling
OxCGRT floored
OxCGRT independent
Google mobility
Facebook tiles visited mobility
Facebook single tile mobility

#### Lesotho

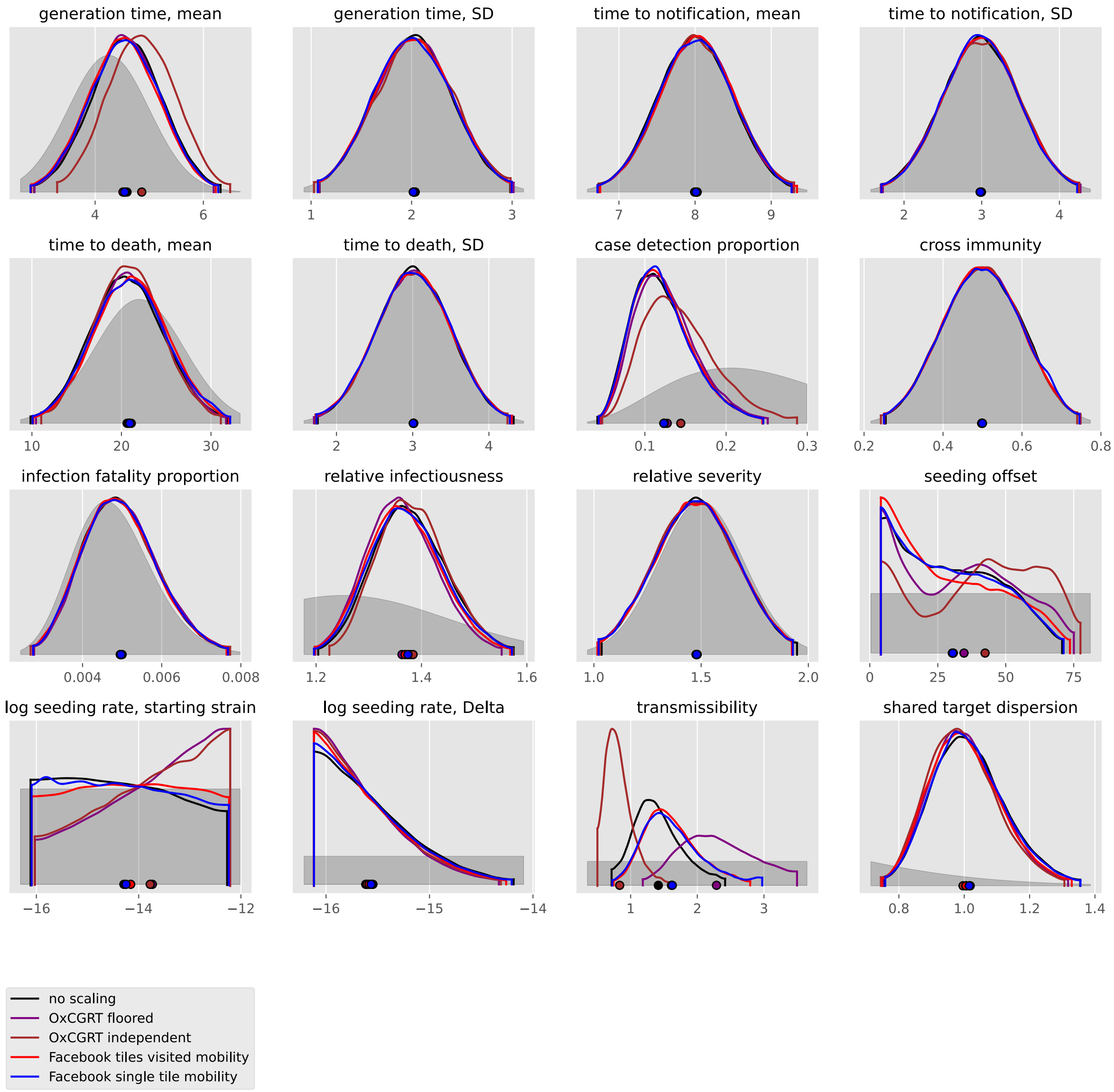
generation time, mean
generation time, SD
time to notification, mean
time to notification, SD
time to death, mean
time to death, SD
case detection proportion
cross immunity
infection fatality proportion
relative infectiousness
relative severity
seeding offset
log seeding rate, starting strain
log seeding rate, Delta
transmissibility
shared target dispersion
no scaling
OxCGRT floored
OxCGRT independent
Facebook tiles visited mobility
Facebook single tile mobility

#### Liberia

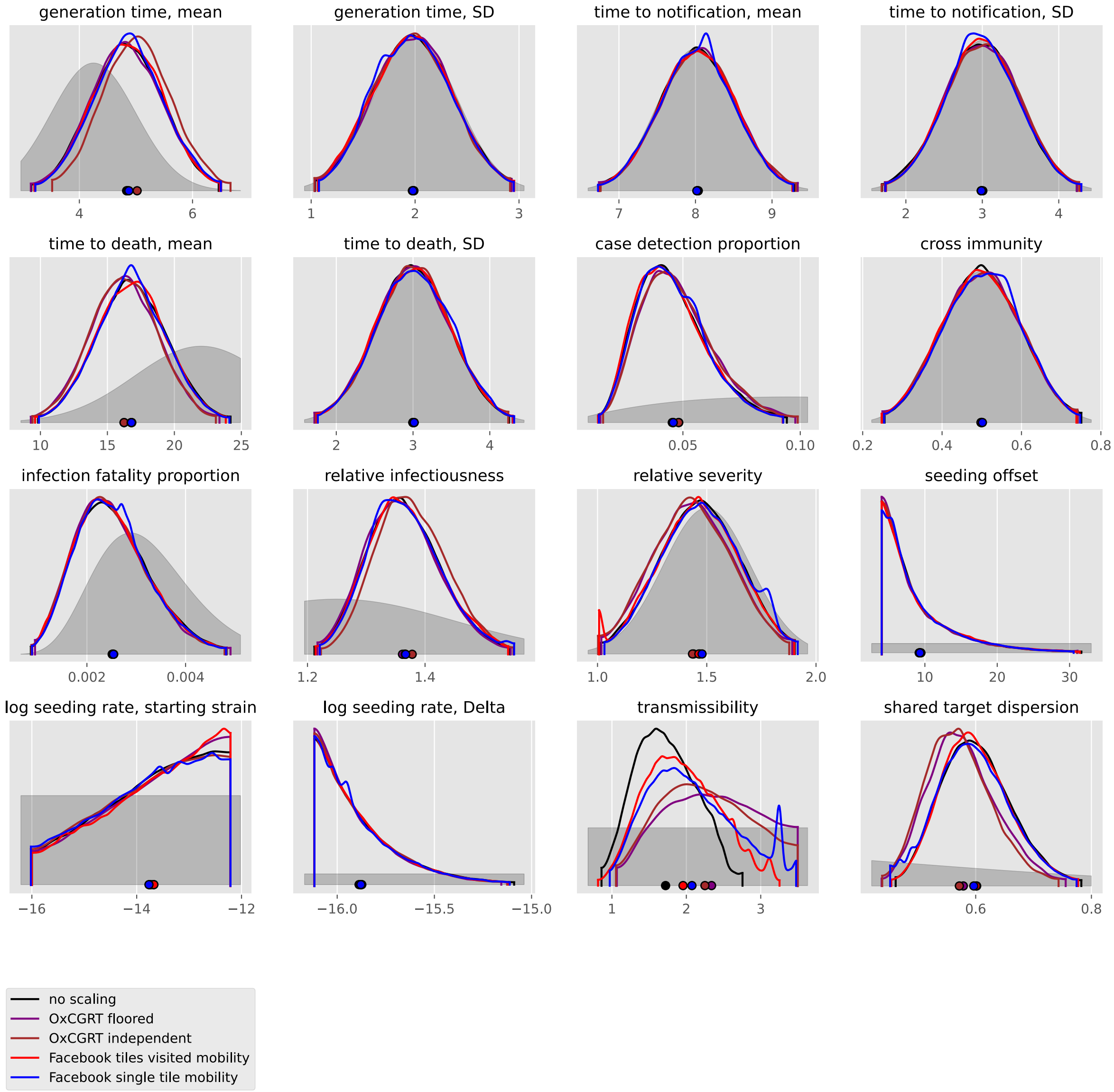
generation time, mean
generation time, SD
time to notification, mean
time to notification, SD
time to death, mean
time to death, SD
case detection proportion
cross immunity
infection fatality proportion
relative infectiousness
relative severity
seeding offset
log seeding rate, starting strain
log seeding rate, Delta
transmissibility
shared target dispersion
no scaling
OxCGRT floored
OxCGRT independent
Facebook tiles visited mobility
Facebook single tile mobility

#### Libya

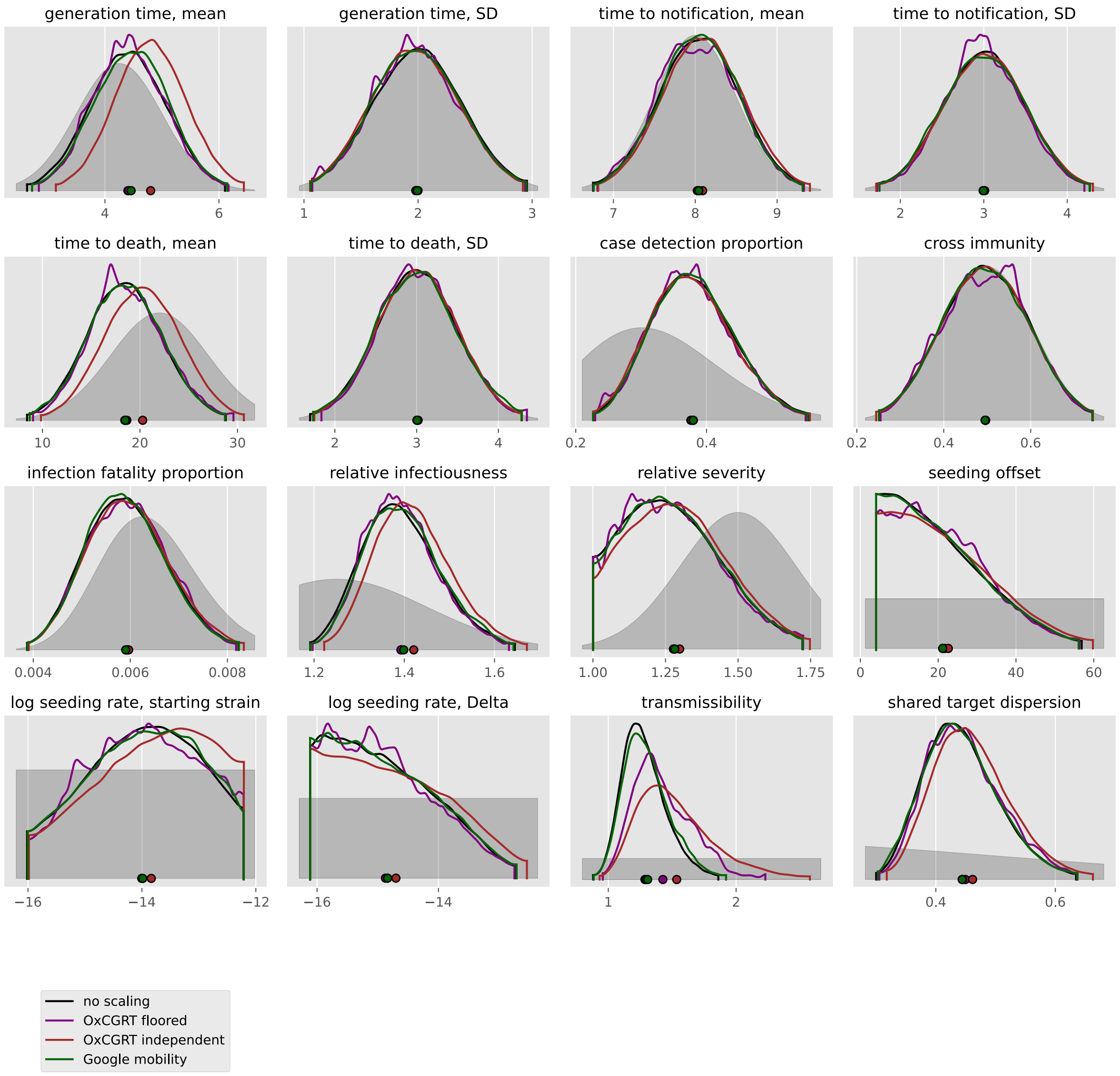

generation time, mean
generation time, SD
time to notification, mean
time to notification, SD
time to death, mean
time to death, SD
case detection proportion
cross immunity
infection fatality proportion
relative infectiousness
relative severity
seeding offset
log seeding rate, starting strain
log seeding rate, Delta
transmissibility
shared target dispersion
no scaling
OxCGRT floored
OxCGRT independent
Google mobility

#### Madagascar

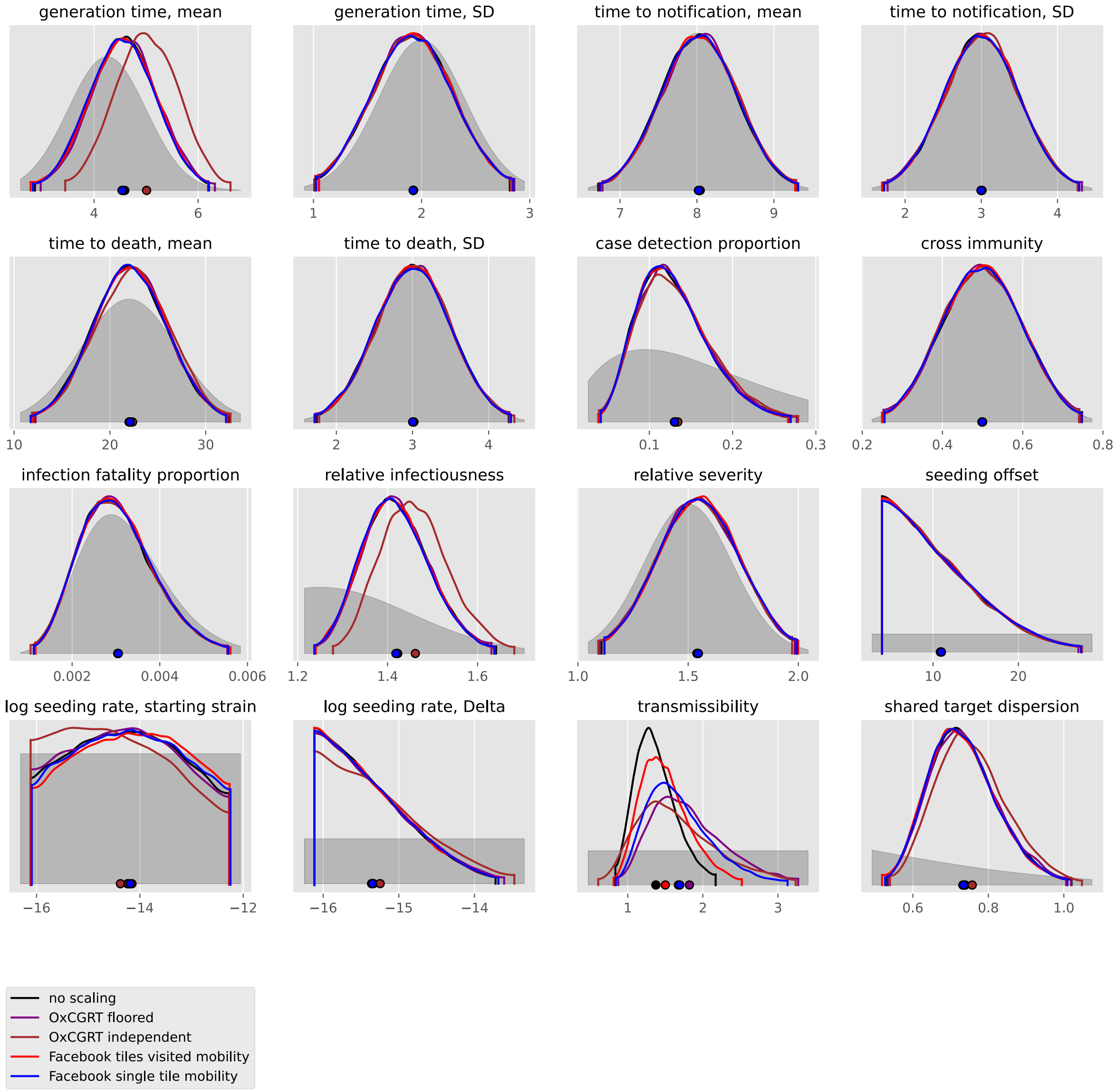
generation time, mean
generation time, SD
time to notification, mean
time to notification, SD
time to death, mean
time to death, SD
case detection proportion
cross immunity
infection fatality proportion
relative infectiousness
relative severity
seeding offset
log seeding rate, starting strain
log seeding rate, Delta
transmissibility
shared target dispersion
no scaling
OxCGRT floored
OxCGRT independent
Facebook tiles visited mobility
Facebook single tile mobility

#### Malawi

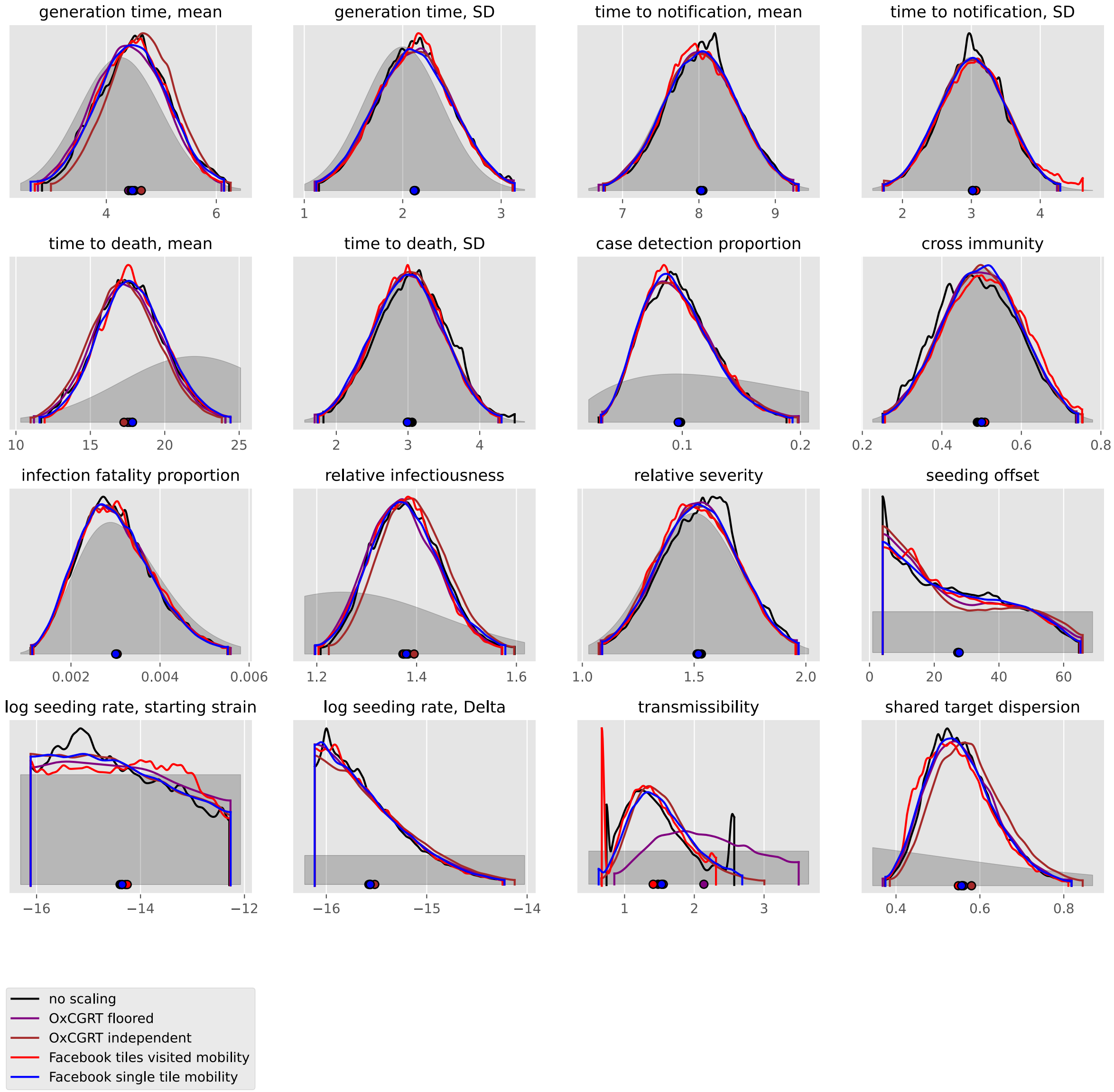

generation time, mean
4
6
generation time, SD
1
2
3
time to notification, mean
7
8
9
time to notification, SD
2
3
4
time to death, mean
10
15
20
25
time to death, SD
2
3
4
case detection proportion
0.1
0.2
cross immunity
0.2
0.4
0.6
0.8
infection fatality proportion
0.002
0.004
0.006
relative infectiousness
1.2
1.4
1.6
relative severity
1.0
1.5
2.0
seeding offset
0
20
40
60
log seeding rate, starting strain
−16
−14
−12
log seeding rate, Delta
−16
−15
−14
transmissibility
1
2
3
shared target dispersion
0.4
0.6
0.8
no scaling
OxCGRT floored
OxCGRT independent
Facebook tiles visited mobility
Facebook single tile mobility

#### Mali

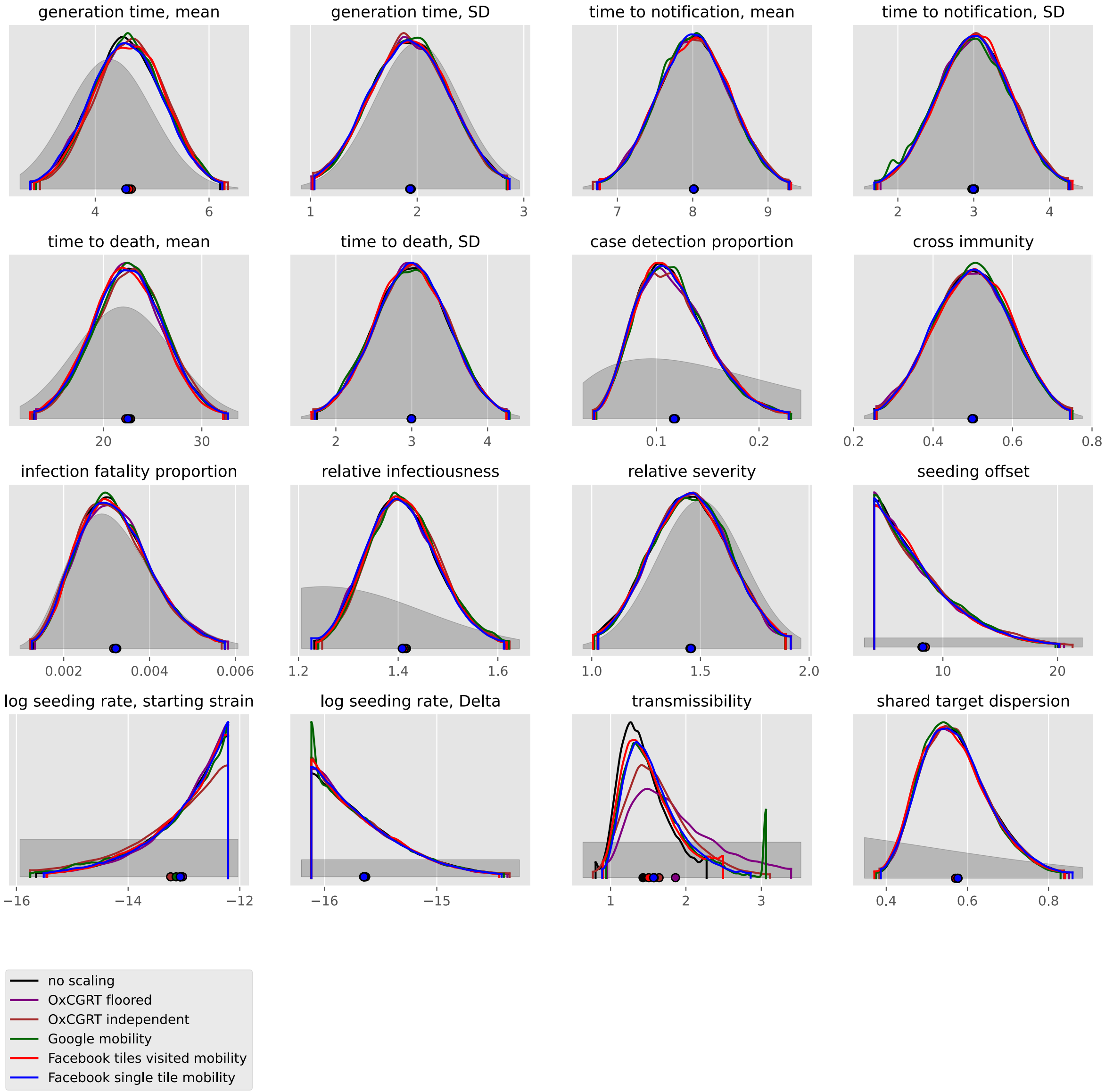
generation time, mean
generation time, SD
time to notification, mean
time to notification, SD
time to death, mean
time to death, SD
case detection proportion
cross immunity
infection fatality proportion
relative infectiousness
relative severity
seeding offset
log seeding rate, starting strain
log seeding rate, Delta
transmissibility
shared target dispersion
no scaling
OxCGRT floored
OxCGRT independent
Google mobility
Facebook tiles visited mobility
Facebook single tile mobility

#### Morocco

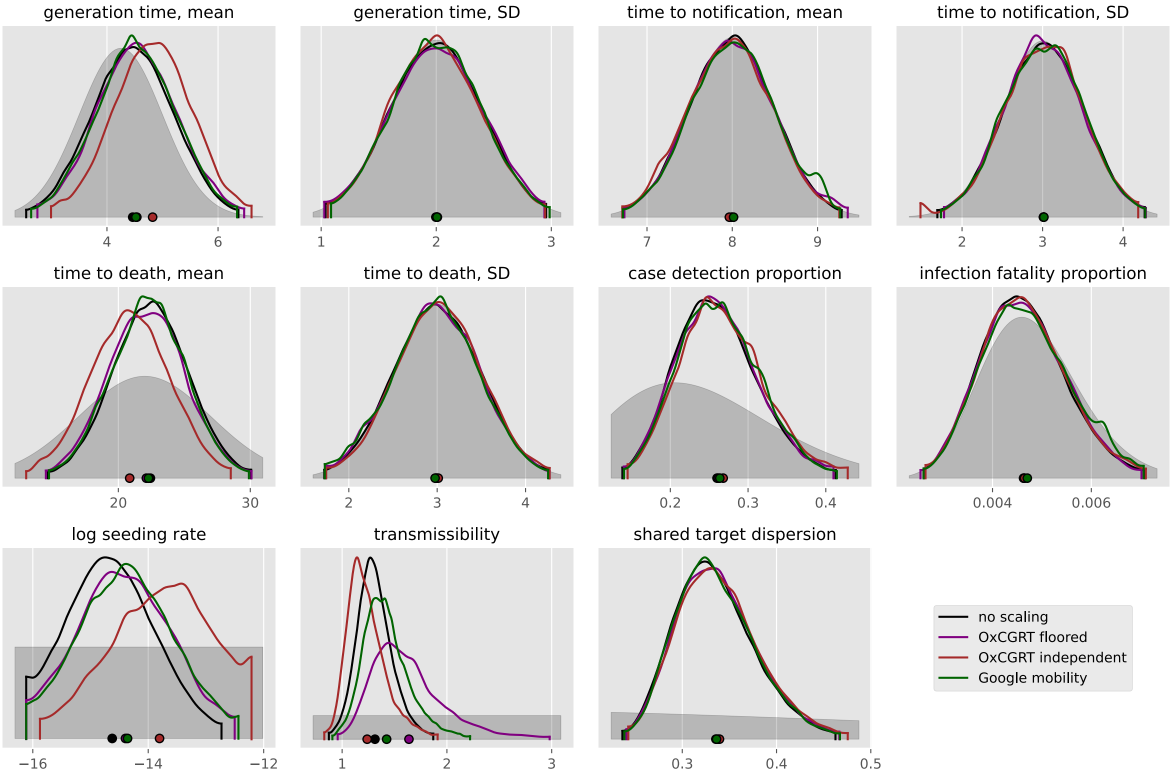

generation time, mean
4
6
generation time, SD
1
2
3
time to notification, mean
7
8
9
time to notification, SD
2
3
4
time to death, mean
20
30
time to death, SD
2
3
4
case detection proportion
0.2
0.3
0.4
infection fatality proportion
0.004
0.006
log seeding rate
−16
−14
−12
transmissibility
1
2
3
shared target dispersion
0.3
0.4
0.5
no scaling
OxCGRT floored
OxCGRT independent
Google mobility

#### Mozambique

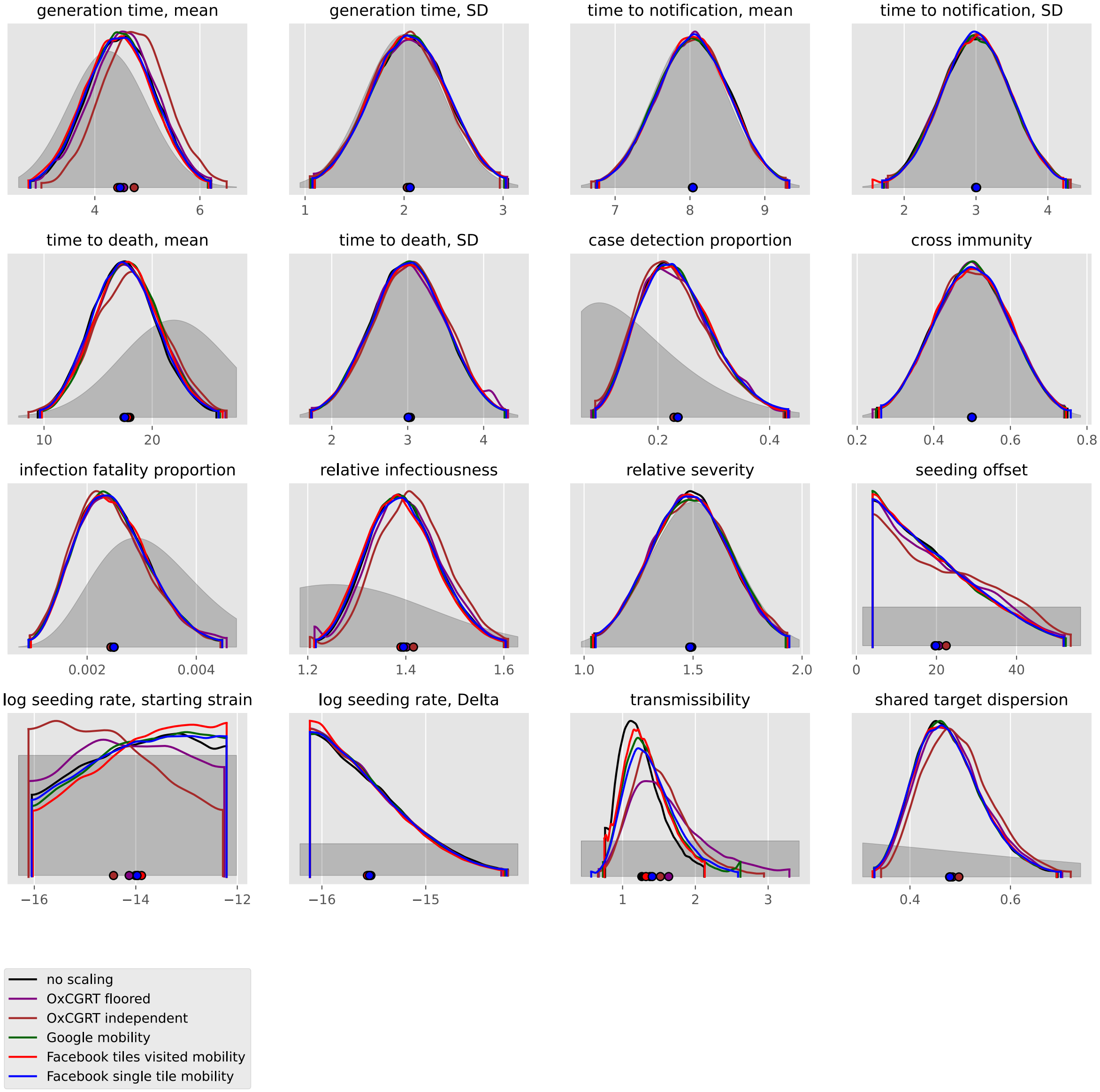

generation time, mean
generation time, SD
time to notification, mean
time to notification, SD
time to death, mean
time to death, SD
case detection proportion
cross immunity
infection fatality proportion
relative infectiousness
relative severity
seeding offset
log seeding rate, starting strain
log seeding rate, Delta
transmissibility
shared target dispersion
no scaling
OxCGRT floored
OxCGRT independent
Google mobility
Facebook tiles visited mobility
Facebook single tile mobility

#### Nigeria

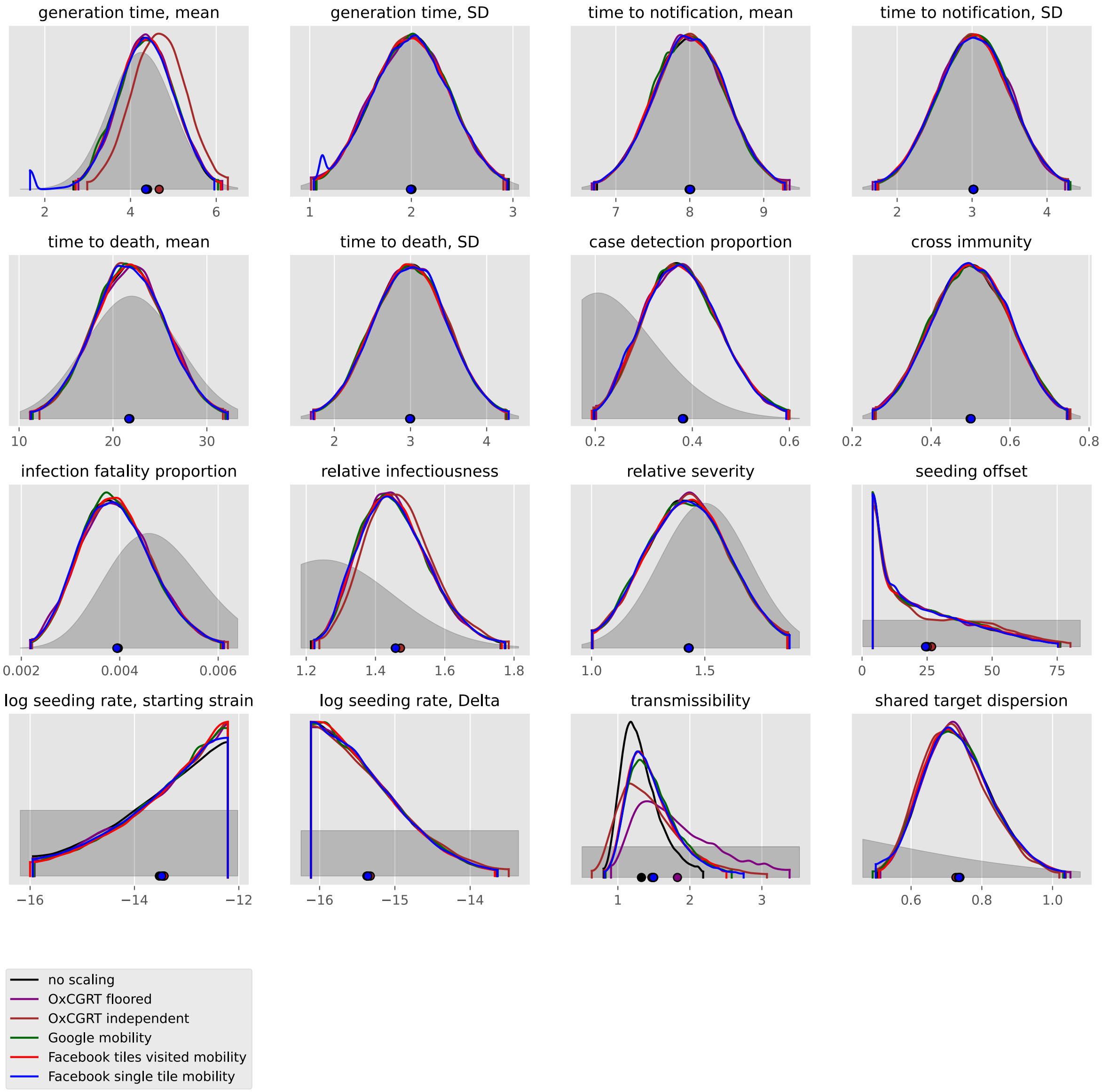
generation time, mean
generation time, SD
time to notification, mean
time to notification, SD
time to death, mean
time to death, SD
case detection proportion
cross immunity
infection fatality proportion
relative infectiousness
relative severity
seeding offset
log seeding rate, starting strain
log seeding rate, Delta
transmissibility
shared target dispersion
no scaling
OxCGRT floored
OxCGRT independent
Google mobility
Facebook tiles visited mobility
Facebook single tile mobility

#### Rwanda

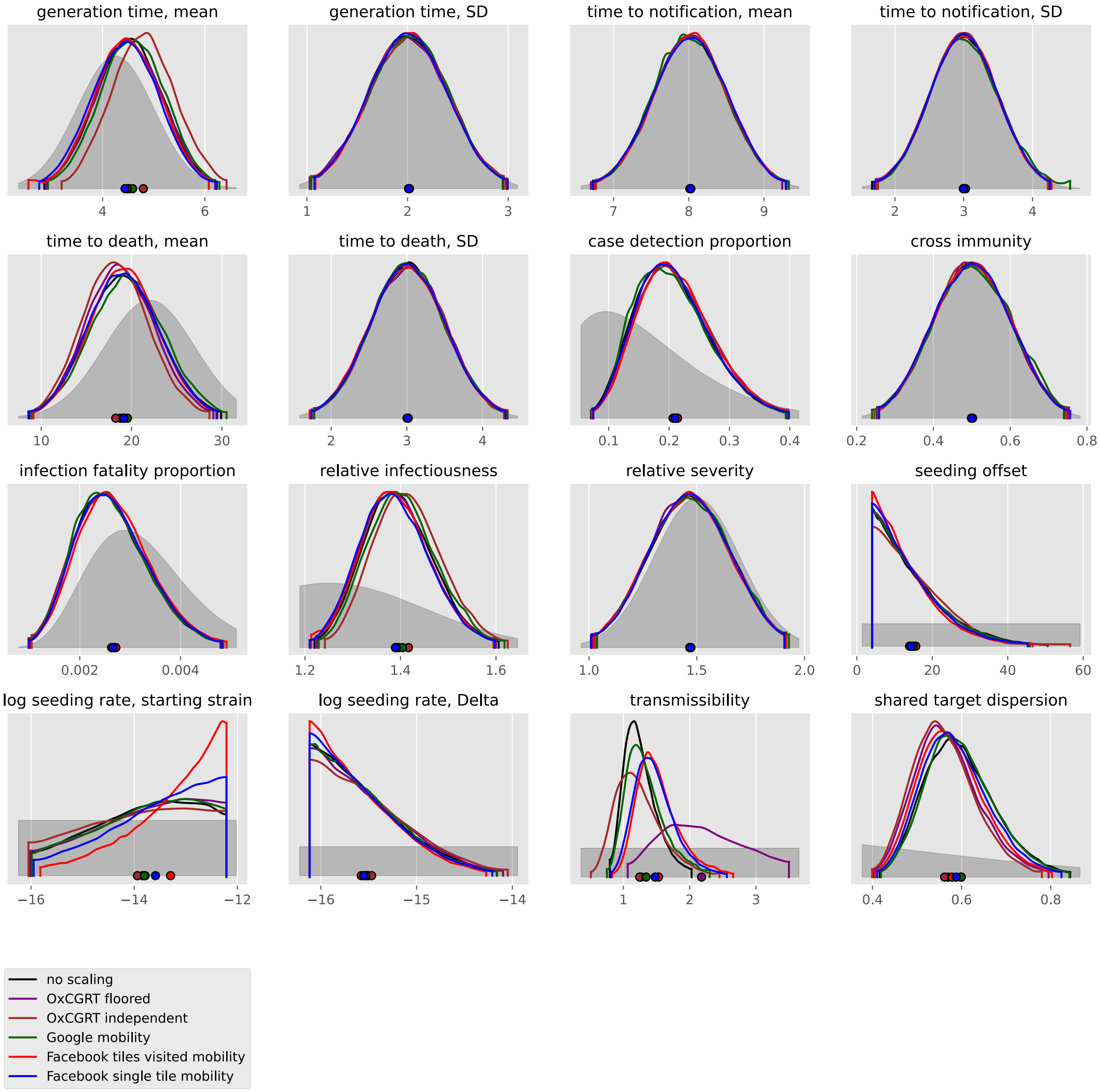
generation time, mean
4
6
generation time, SD
1
2
3
time to notification, mean
7
8
9
time to notification, SD
2
3
4
time to death, mean
10
20
30
time to death, SD
2
3
4
case detection proportion
0.1
0.2
0.3
0.4
cross immunity
0.2
0.4
0.6
0.8
infection fatality proportion
0.002
0.004
relative infectiousness
1.2
1.4
1.6
relative severity
1.0
1.5
2.0
seeding offset
0
20
40
60
log seeding rate, starting strain
−16
−14
−12
log seeding rate, Delta
−16
−15
−14
transmissibility
1
2
3
shared target dispersion
0.4
0.6
0.8
no scaling
OxCGRT floored
OxCGRT independent
Google mobility
Facebook tiles visited mobility
Facebook single tile mobility

#### Senegal

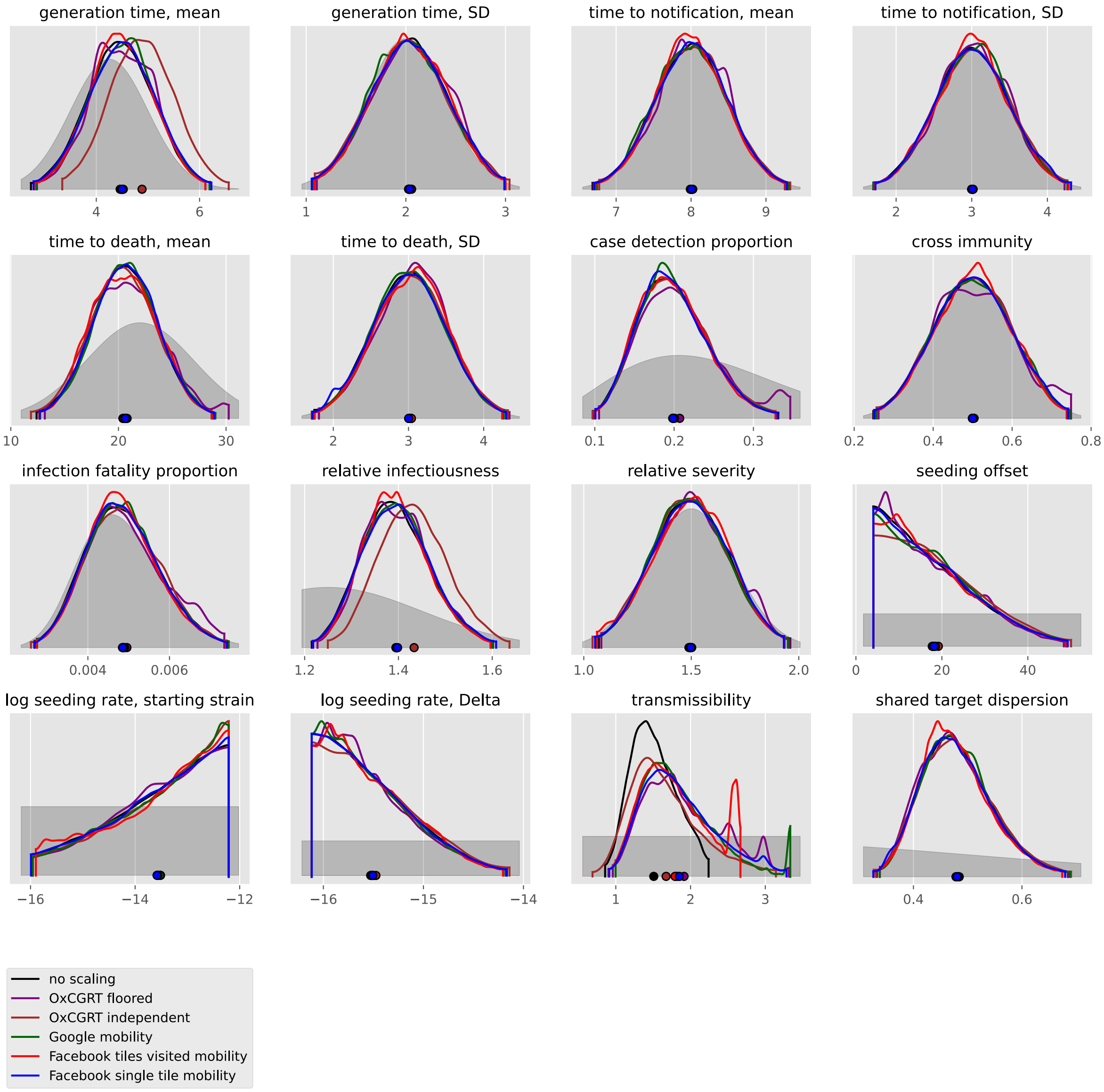

generation time, mean
4
6
generation time, SD
1
2
3
time to notification, mean
7
8
9
time to notification, SD
2
3
4
time to death, mean
10
20
30
time to death, SD
2
3
4
case detection proportion
0.1
0.2
0.3
cross immunity
0.2
0.4
0.6
0.8
infection fatality proportion
0.004
0.006
relative infectiousness
1.2
1.4
1.6
relative severity
1.0
1.5
2.0
seeding offset
0
20
40
log seeding rate, starting strain
−16
−14
−12
log seeding rate, Delta
−16
−15
−14
transmissibility
1
2
3
shared target dispersion
0.4
0.6
no scaling
OxCGRT floored
OxCGRT independent
Google mobility
Facebook tiles visited mobility
Facebook single tile mobility

#### Somalia

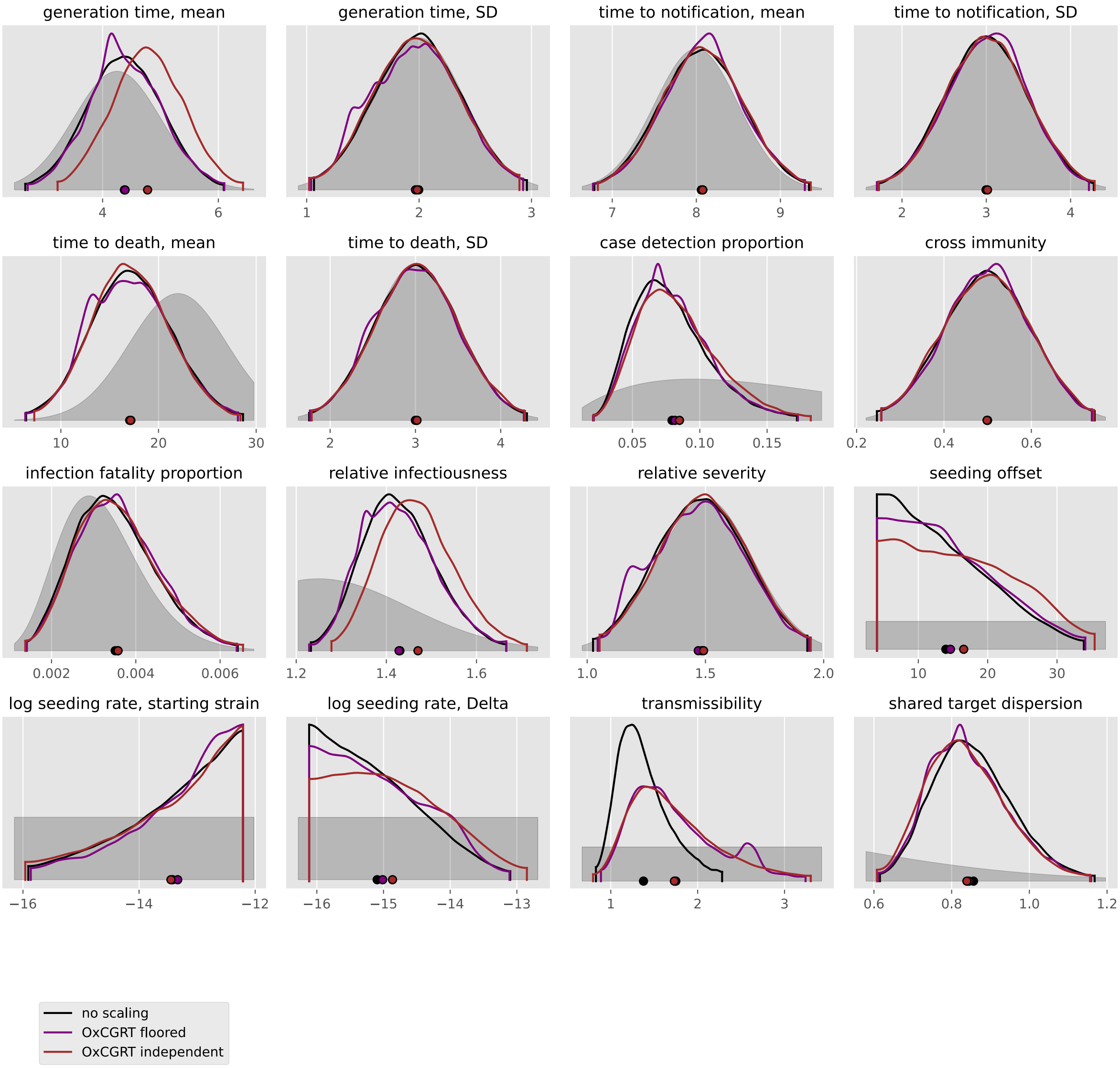

generation time, mean
generation time, SD
time to notification, mean
time to notification, SD
time to death, mean
time to death, SD
case detection proportion
cross immunity
infection fatality proportion
relative infectiousness
relative severity
seeding offset
log seeding rate, starting strain
log seeding rate, Delta
transmissibility
shared target dispersion
no scaling
OxCGRT floored
OxCGRT independent

#### South Africa

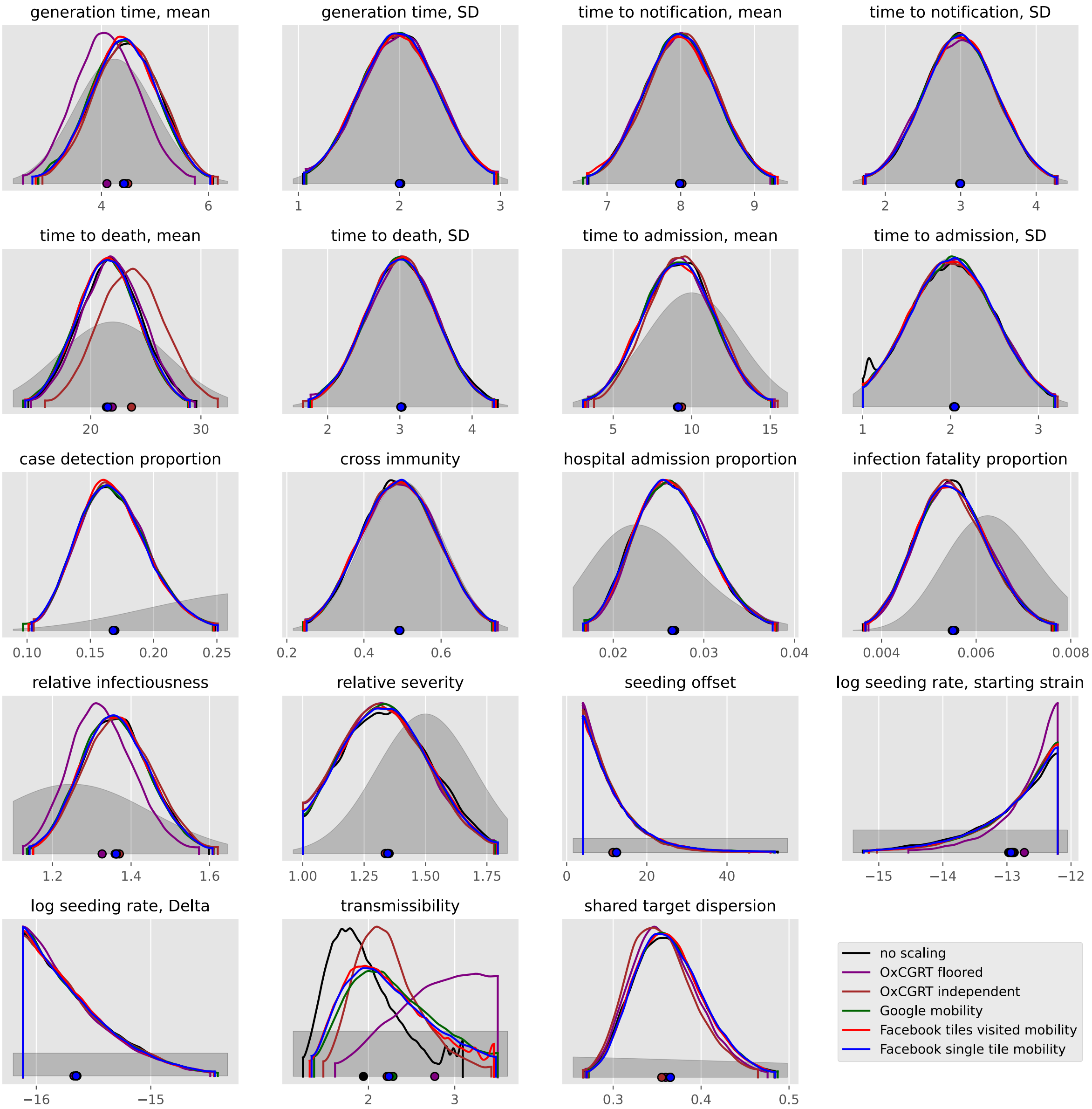

#### South Sudan

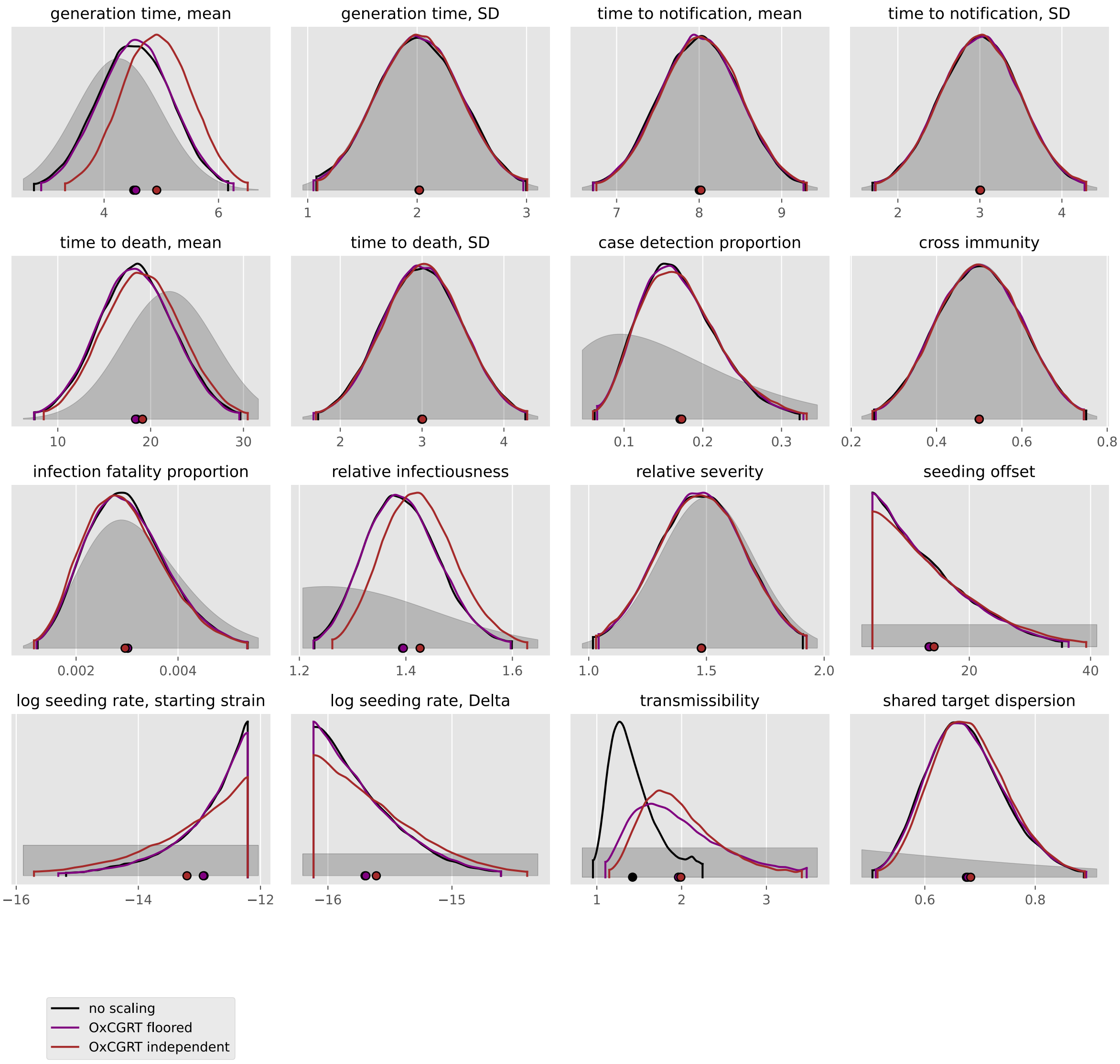

generation time, mean
generation time, SD
time to notification, mean
time to notification, SD
time to death, mean
time to death, SD
case detection proportion
cross immunity
infection fatality proportion
relative infectiousness
relative severity
seeding offset
log seeding rate, starting strain
log seeding rate, Delta
transmissibility
shared target dispersion
no scaling
OxCGRT floored
OxCGRT independent

#### Sudan

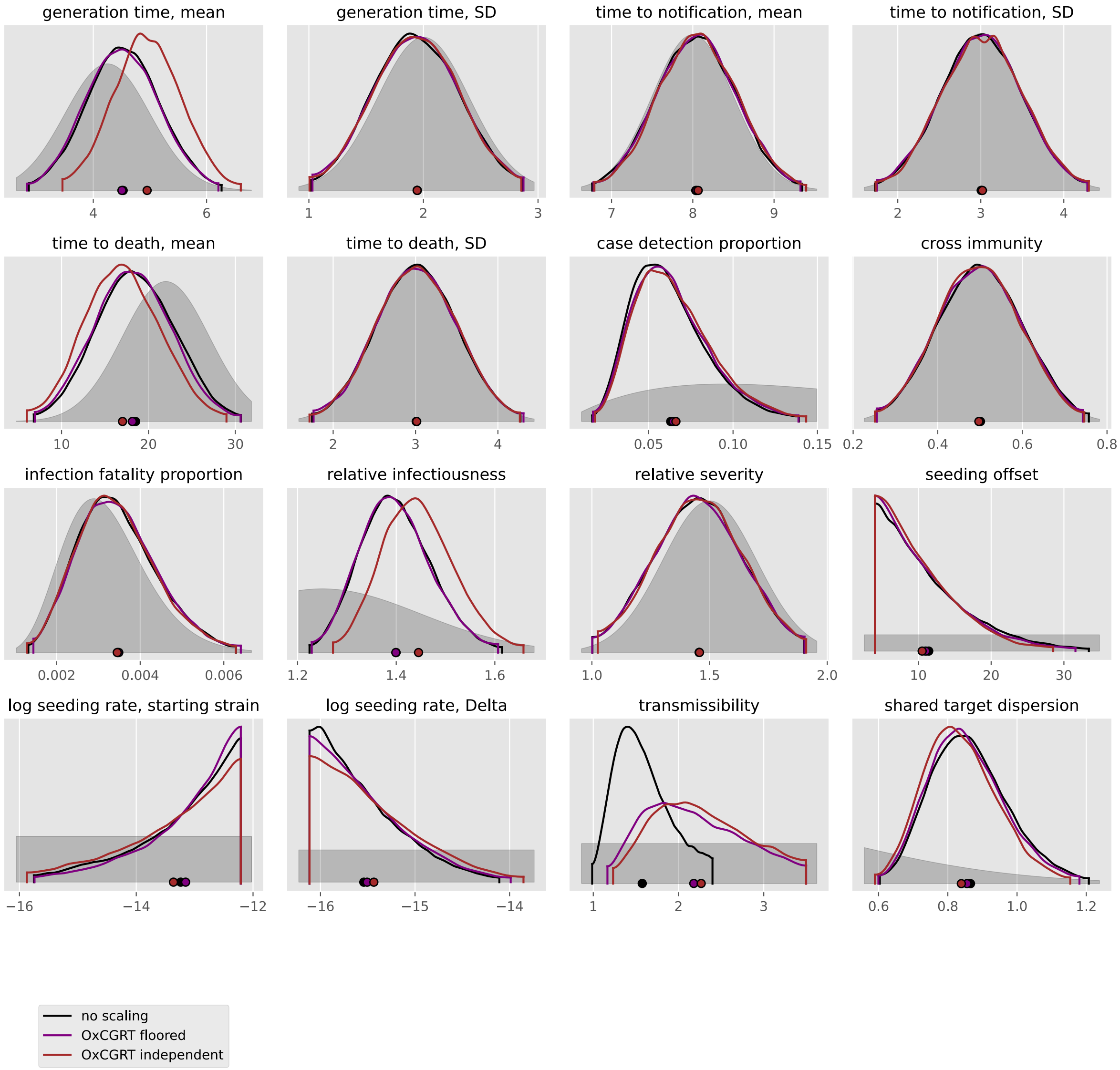

generation time, mean
generation time, SD
time to notification, mean
time to notification, SD
time to death, mean
time to death, SD
case detection proportion
cross immunity
infection fatality proportion
relative infectiousness
relative severity
seeding offset
log seeding rate, starting strain
log seeding rate, Delta
transmissibility
shared target dispersion
no scaling
OxCGRT floored
OxCGRT independent

#### Togo

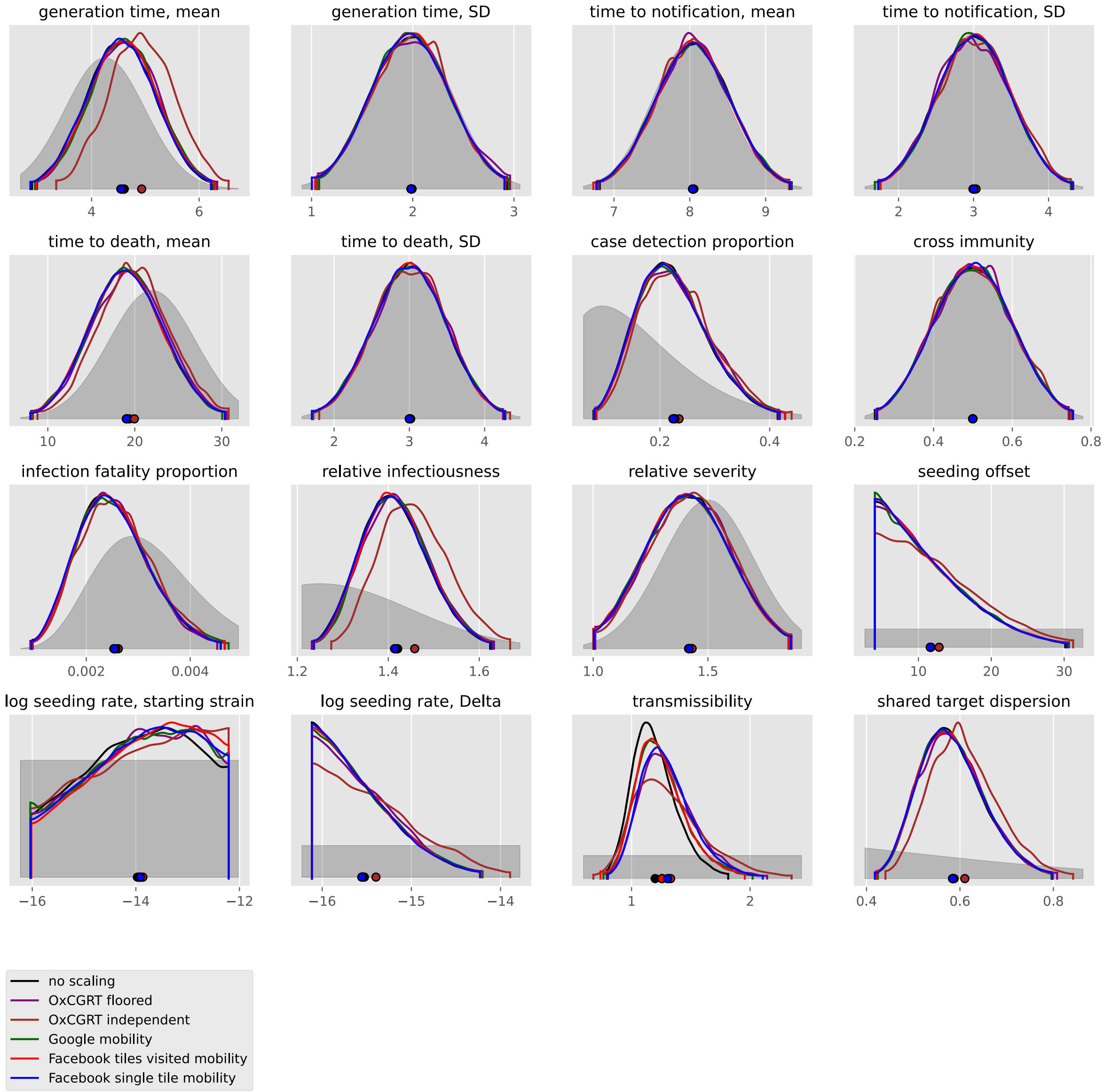
generation time, mean
4
6
generation time, SD
1
2
3
time to notification, mean
7
8
9
time to notification, SD
2
3
4
time to death, mean
10
20
30
time to death, SD
2
3
4
case detection proportion
0.2
0.4
cross immunity
0.2
0.4
0.6
0.8
infection fatality proportion
0.002
0.004
relative infectiousness
1.2
1.4
1.6
relative severity
1.0
1.5
seeding offset
10
20
30
log seeding rate, starting strain
−16
−14
−12
log seeding rate, Delta
−16
−15
−14
transmissibility
1
2
shared target dispersion
0.4
0.6
0.8
no scaling
OxCGRT floored
OxCGRT independent
Google mobility
Facebook tiles visited mobility
Facebook single tile mobility

#### Tunisia

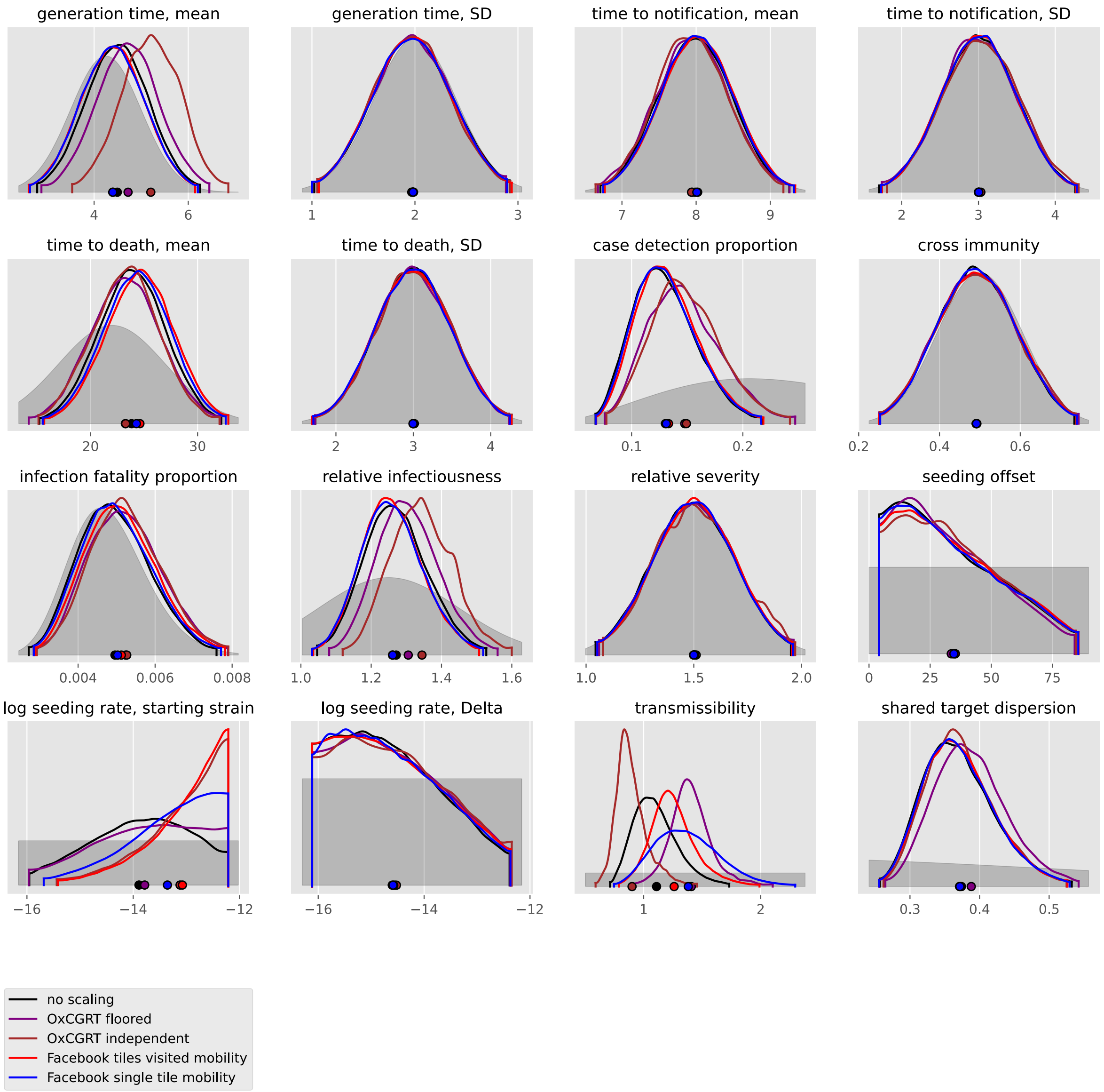

generation time, mean
4
6
generation time, SD
1
2
3
time to notification, mean
7
8
9
time to notification, SD
2
3
4
time to death, mean
20
30
time to death, SD
2
3
4
case detection proportion
0.1
0.2
cross immunity
0.2
0.4
0.6
infection fatality proportion
0.004
0.006
0.008
relative infectiousness
1.0
1.2
1.4
1.6
relative severity
1.0
1.5
2.0
seeding offset
0
25
50
75
log seeding rate, starting strain
−16
−14
−12
log seeding rate, Delta
−16
−14
−12
transmissibility
1
2
shared target dispersion
0.3
0.4
0.5
no scaling
OxCGRT floored
OxCGRT independent
Facebook tiles visited mobility
Facebook single tile mobility

#### Zambia

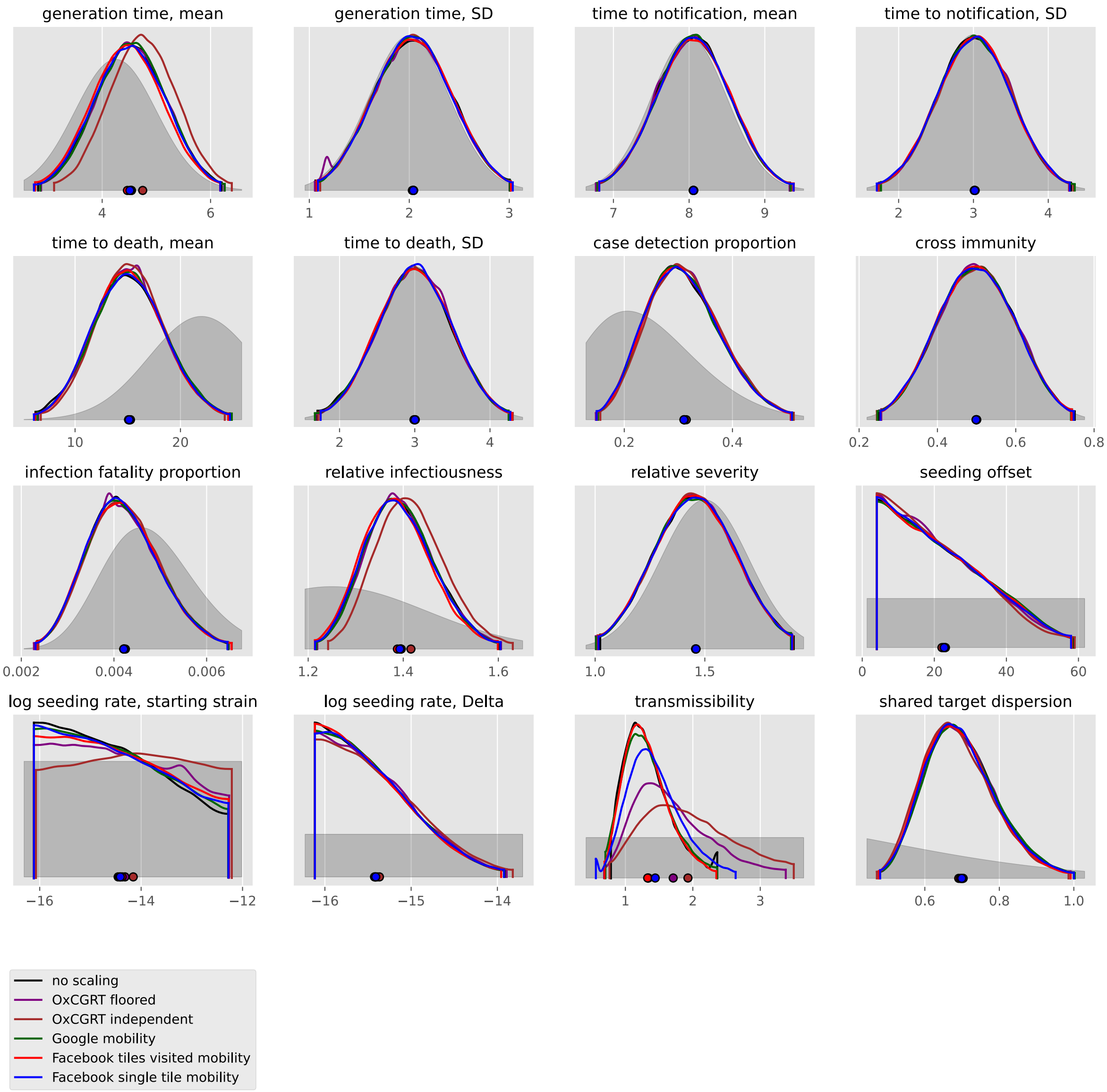

generation time, mean
generation time, SD
time to notification, mean
time to notification, SD
time to death, mean
time to death, SD
case detection proportion
cross immunity
infection fatality proportion
relative infectiousness
relative severity
seeding offset
log seeding rate, starting strain
log seeding rate, Delta
transmissibility
shared target dispersion
no scaling
OxCGRT floored
OxCGRT independent
Google mobility
Facebook tiles visited mobility
Facebook single tile mobility

#### Zimbabwe

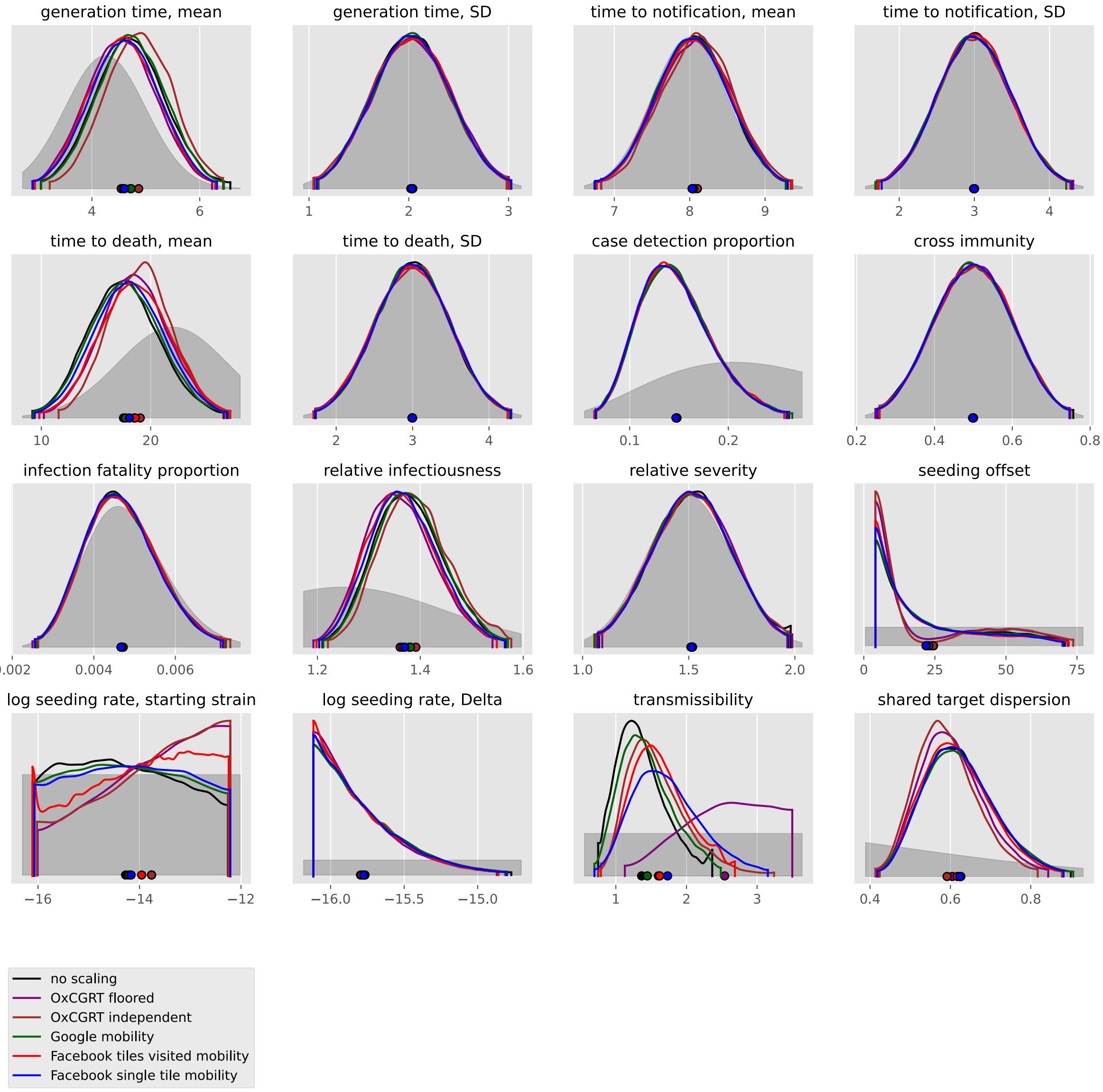
generation time, mean
4
6
generation time, SD
1
2
3
time to notification, mean
7
8
9
time to notification, SD
2
3
4
time to death, mean
10
20
time to death, SD
2
3
4
case detection proportion
0.1
0.2
cross immunity
0.2
0.4
0.6
0.8
infection fatality proportion
0.002
0.004
0.006
relative infectiousness
1.2
1.4
1.6
relative severity
1.0
1.5
2.0
seeding offset
0
25
50
75
log seeding rate, starting strain
−16
−14
−12
log seeding rate, Delta
−16.0
−15.5
−15.0
transmissibility
1
2
3
shared target dispersion
0.4
0.6
0.8
no scaling
OxCGRT floored
OxCGRT independent
Google mobility
Facebook tiles visited mobility
Facebook single tile mobility

### Asia

#### Afghanistan

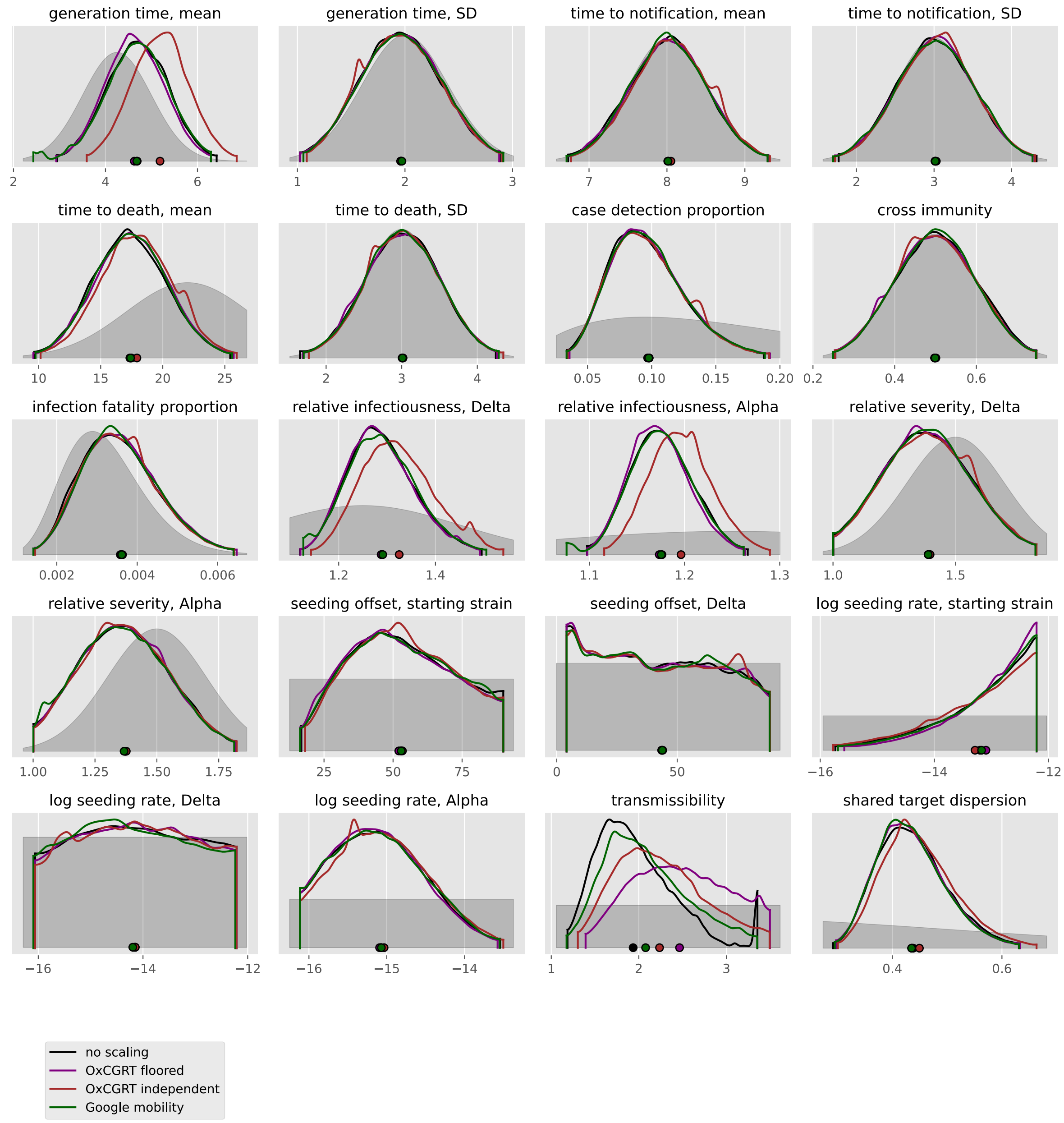

#### Azerbaijan

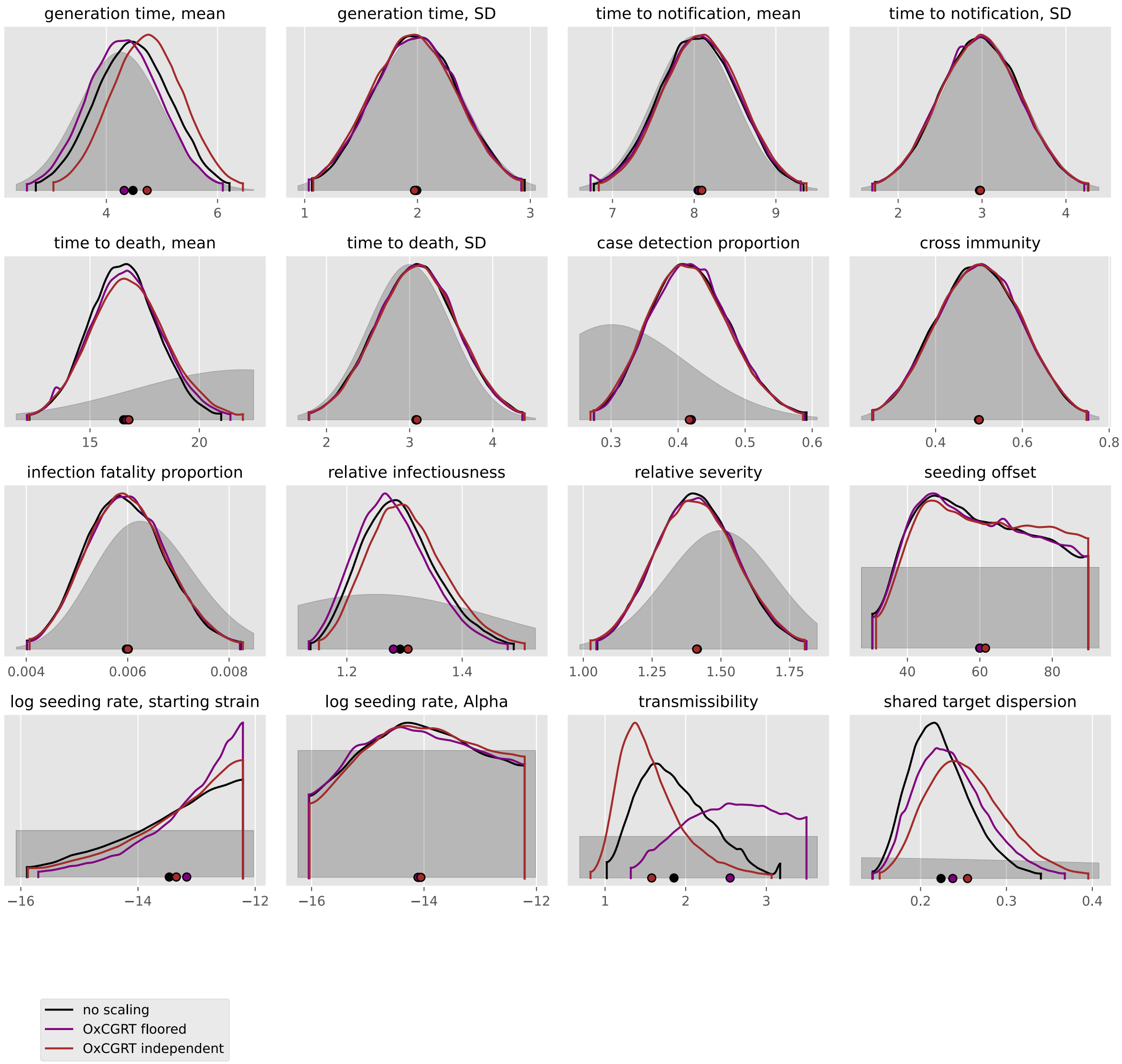

generation time, mean
generation time, SD
time to notification, mean
time to notification, SD
time to death, mean
time to death, SD
case detection proportion
cross immunity
infection fatality proportion
relative infectiousness
relative severity
seeding offset
log seeding rate, starting strain
log seeding rate, Alpha
transmissibility
shared target dispersion
no scaling
OxCGRT floored
OxCGRT independent

#### Bahrain

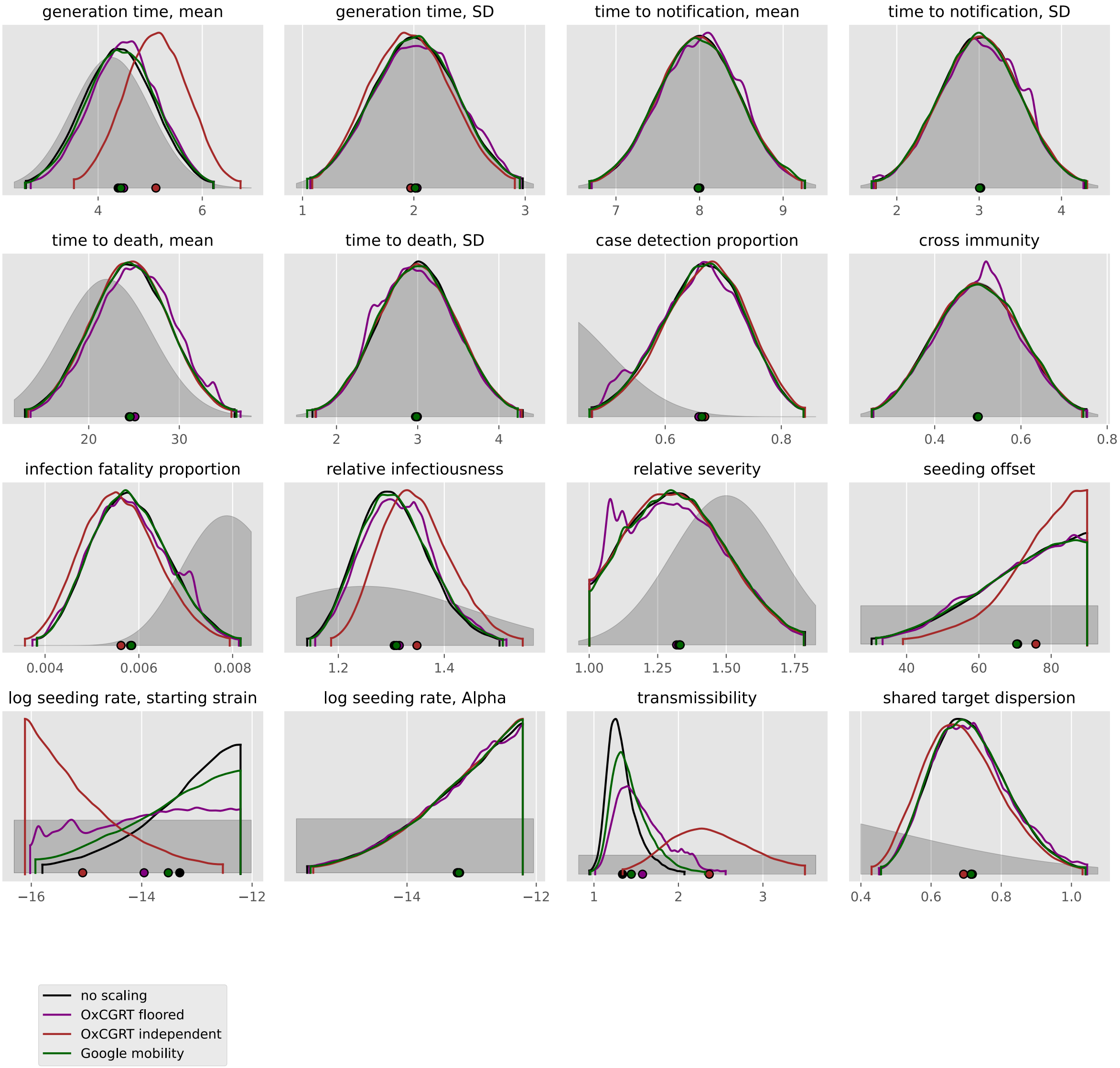

generation time, mean
generation time, SD
time to notification, mean
time to notification, SD
time to death, mean
time to death, SD
case detection proportion
cross immunity
infection fatality proportion
relative infectiousness
relative severity
seeding offset
log seeding rate, starting strain
log seeding rate, Alpha
transmissibility
shared target dispersion
no scaling
OxCGRT floored
OxCGRT independent
Google mobility

#### Bangladesh

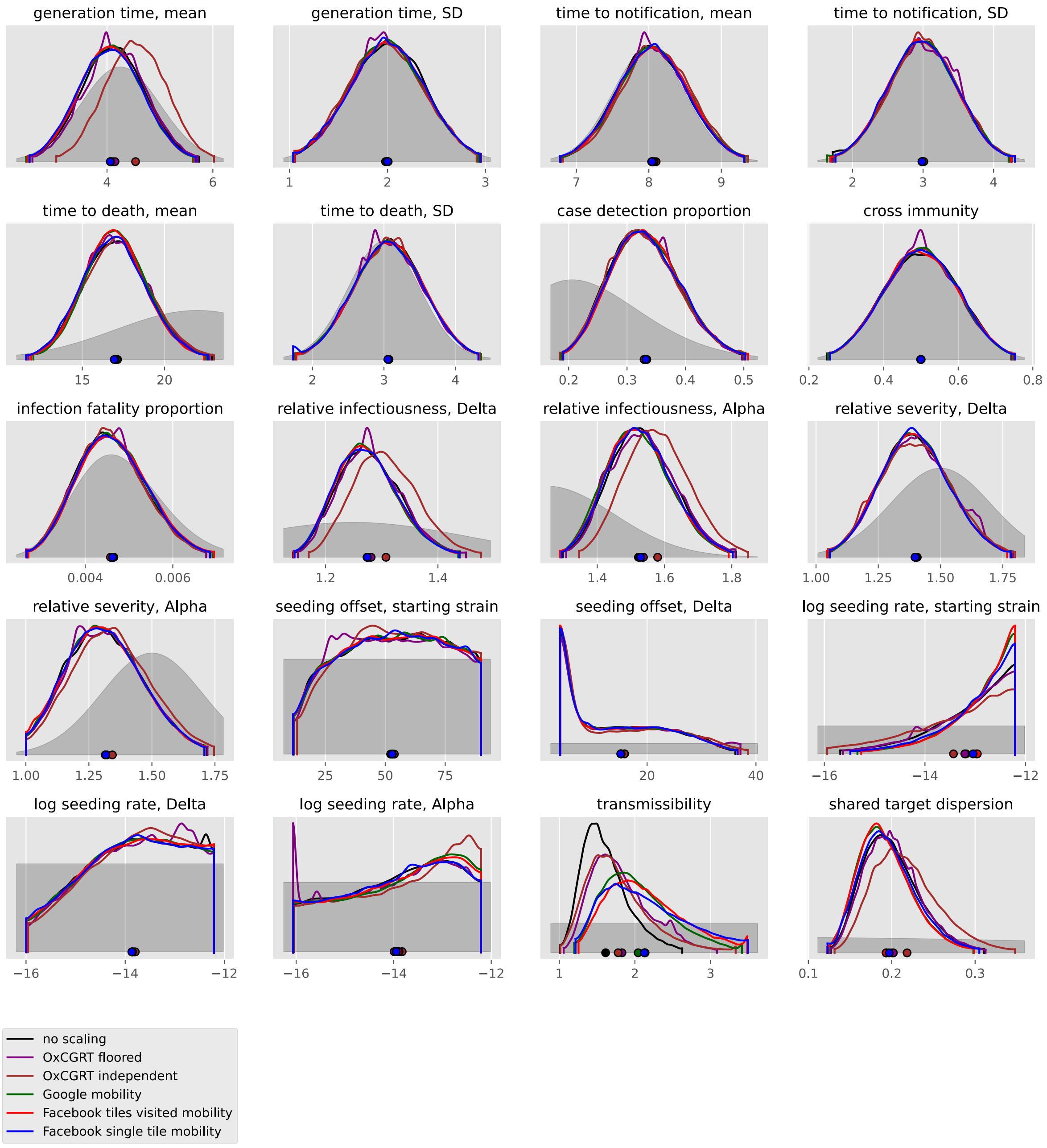
generation time, mean
generation time, SD
time to notification, mean
time to notification, SD
time to death, mean
time to death, SD
case detection proportion
cross immunity
infection fatality proportion
relative infectiousness, Delta
relative infectiousness, Alpha
relative severity, Delta
relative severity, Alpha
seeding offset, starting strain
seeding offset, Delta
log seeding rate, starting strain
log seeding rate, Delta
log seeding rate, Alpha
transmissibility
shared target dispersion
no scaling
OxCGRT floored
OxCGRT independent
Google mobility
Facebook tiles visited mobility
Facebook single tile mobility

#### Cyprus

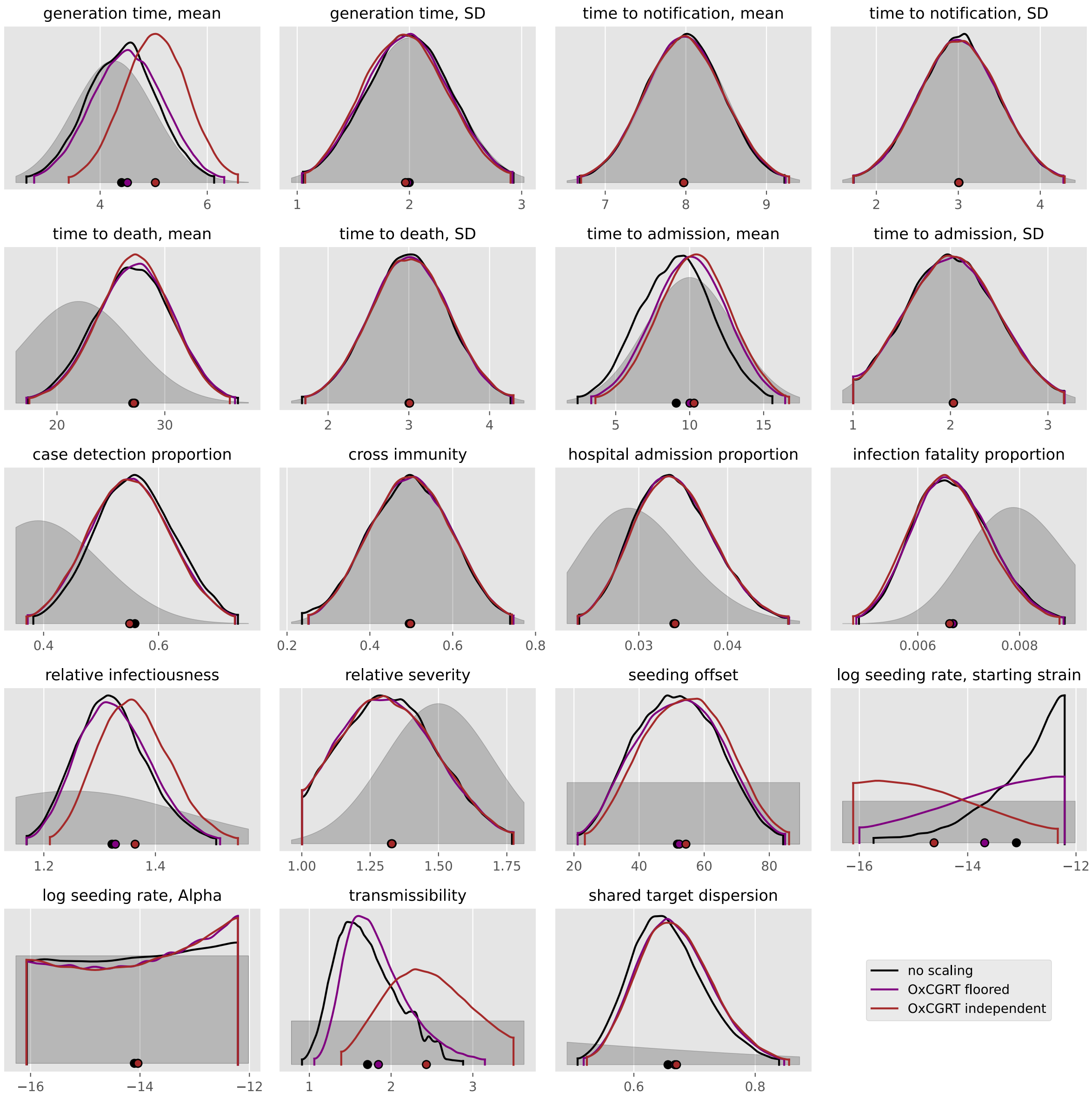

generation time, mean
4
6
generation time, SD
1
2
3
time to notification, mean
7
8
9
time to notification, SD
2
3
4
time to death, mean
20
30
time to death, SD
2
3
4
time to admission, mean
5
10
15
time to admission, SD
1
2
3
case detection proportion
0.4
0.6
cross immunity
0.2
0.4
0.6
0.8
hospital admission proportion
0.03
0.04
infection fatality proportion
0.006
0.008
relative infectiousness
1.2
1.4
relative severity
1.00
1.25
1.50
1.75
seeding offset
20
40
60
80
log seeding rate, starting strain
−16
−14
−12
log seeding rate, Alpha
−16
−14
−12
transmissibility
1
2
3
shared target dispersion
0.6
0.8
no scaling
OxCGRT floored
OxCGRT independent

#### Georgia

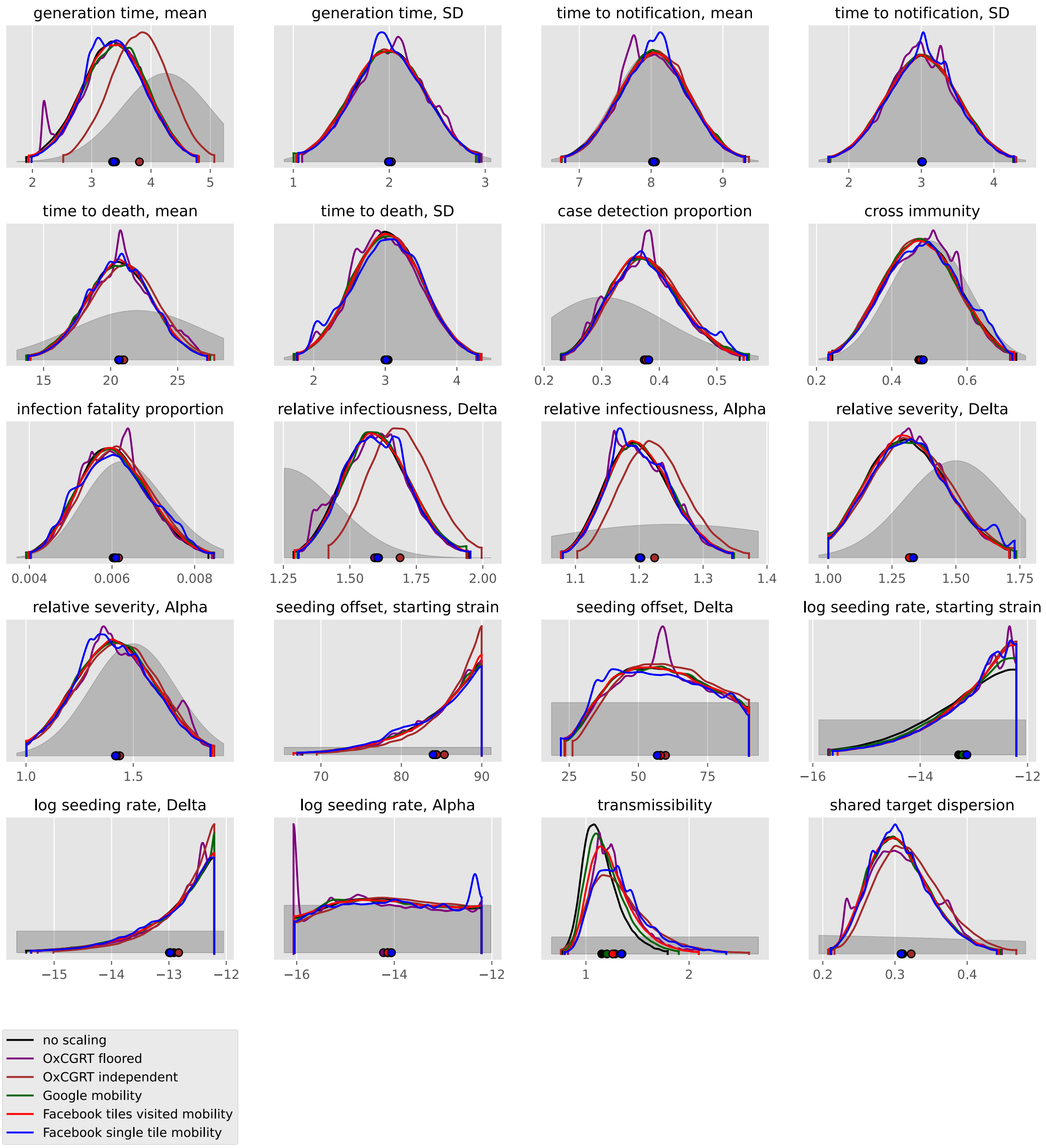
generation time, mean
generation time, SD
time to notification, mean
time to notification, SD
time to death, mean
time to death, SD
case detection proportion
cross immunity
infection fatality proportion
relative infectiousness, Delta
relative infectiousness, Alpha
relative severity, Delta
relative severity, Alpha
seeding offset, starting strain
seeding offset, Delta
log seeding rate, starting strain
log seeding rate, Delta
log seeding rate, Alpha
transmissibility
shared target dispersion
no scaling
OxCGRT floored
OxCGRT independent
Google mobility
Facebook tiles visited mobility
Facebook single tile mobility

#### India

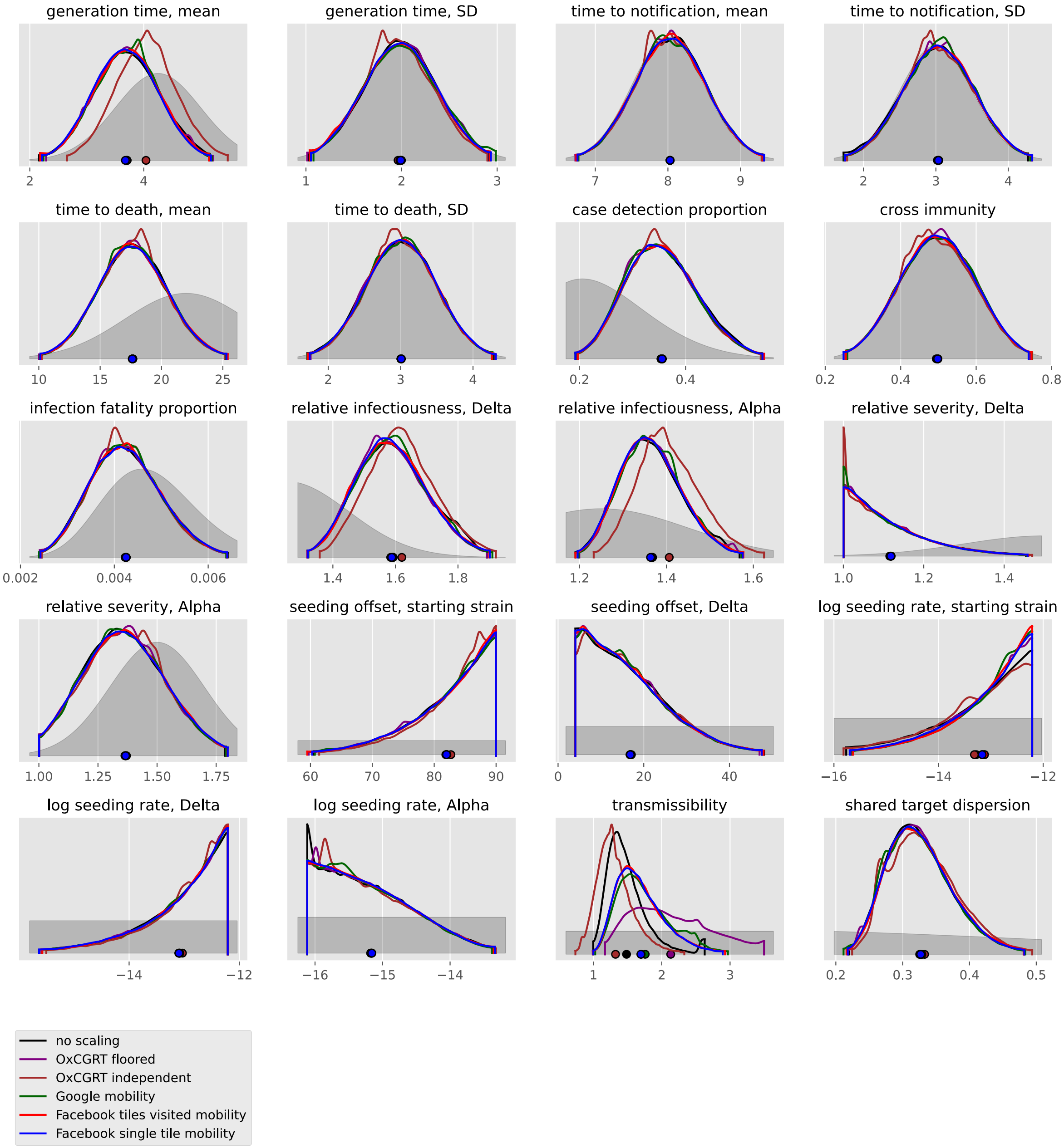
generation time, mean
generation time, SD
time to notification, mean
time to notification, SD
time to death, mean
time to death, SD
case detection proportion
cross immunity
infection fatality proportion
relative infectiousness, Delta
relative infectiousness, Alpha
relative severity, Delta
relative severity, Alpha
seeding offset, starting strain
seeding offset, Delta
log seeding rate, starting strain
log seeding rate, Delta
log seeding rate, Alpha
transmissibility
shared target dispersion
no scaling
OxCGRT floored
OxCGRT independent
Google mobility
Facebook tiles visited mobility
Facebook single tile mobility

#### Indonesia

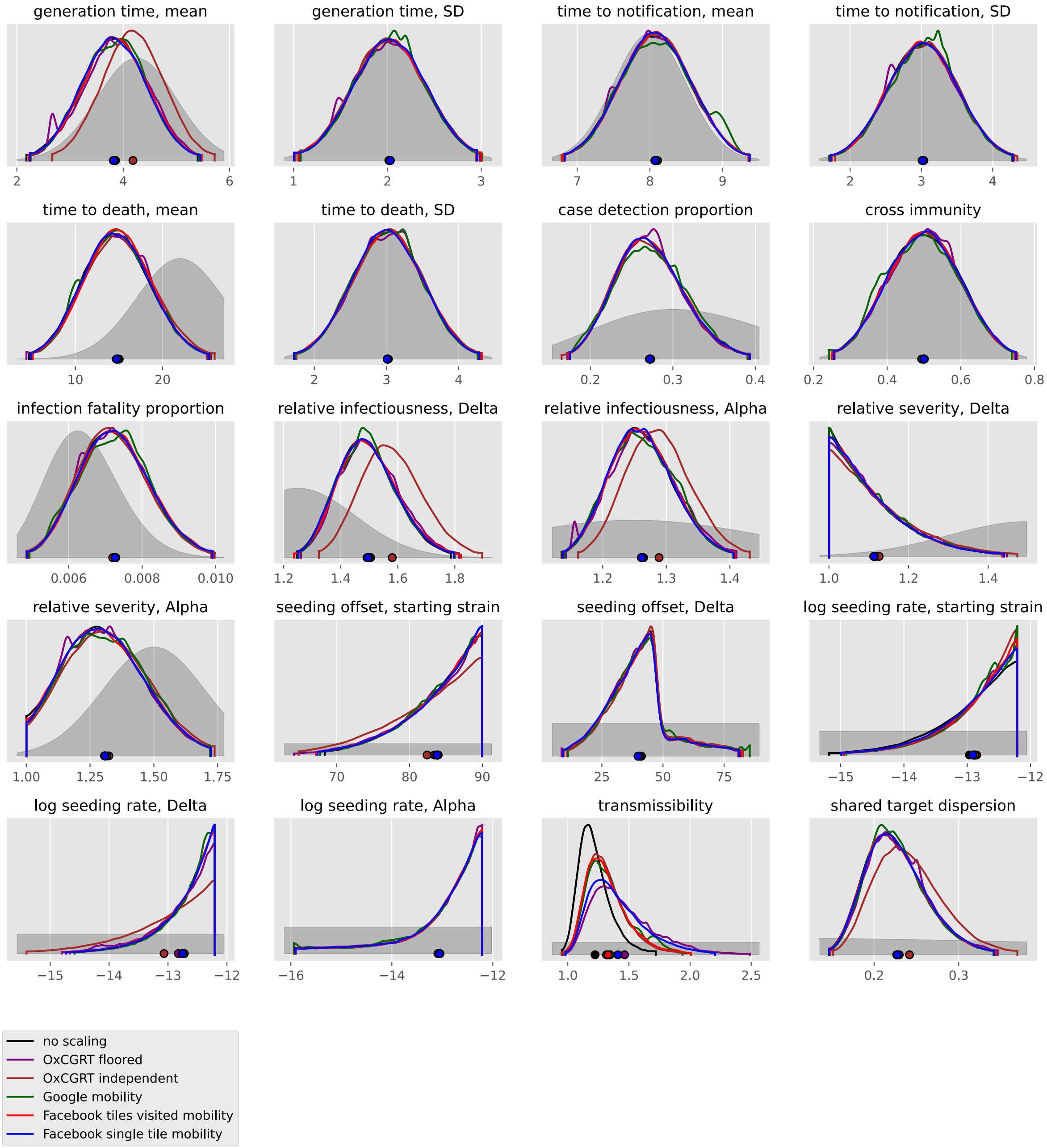

generation time, mean
generation time, SD
time to notification, mean
time to notification, SD
time to death, mean
time to death, SD
case detection proportion
cross immunity
infection fatality proportion
relative infectiousness, Delta
relative infectiousness, Alpha
relative severity, Delta
relative severity, Alpha
seeding offset, starting strain
seeding offset, Delta
log seeding rate, starting strain
log seeding rate, Delta
log seeding rate, Alpha
transmissibility
shared target dispersion
no scaling
OxCGRT floored
OxCGRT independent
Google mobility
Facebook tiles visited mobility
Facebook single tile mobility

#### Iran, Islamic Republic of

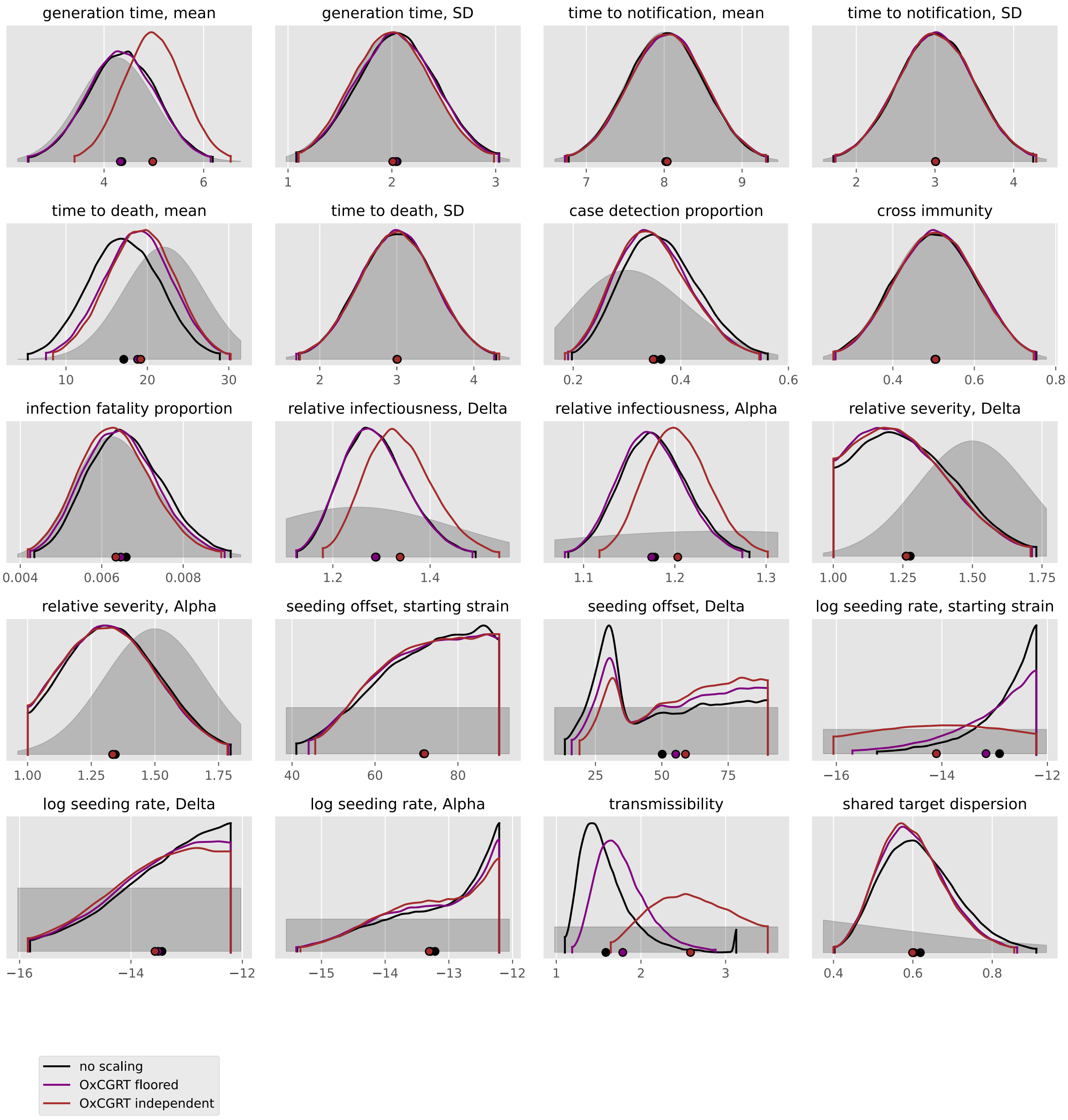

#### Iraq

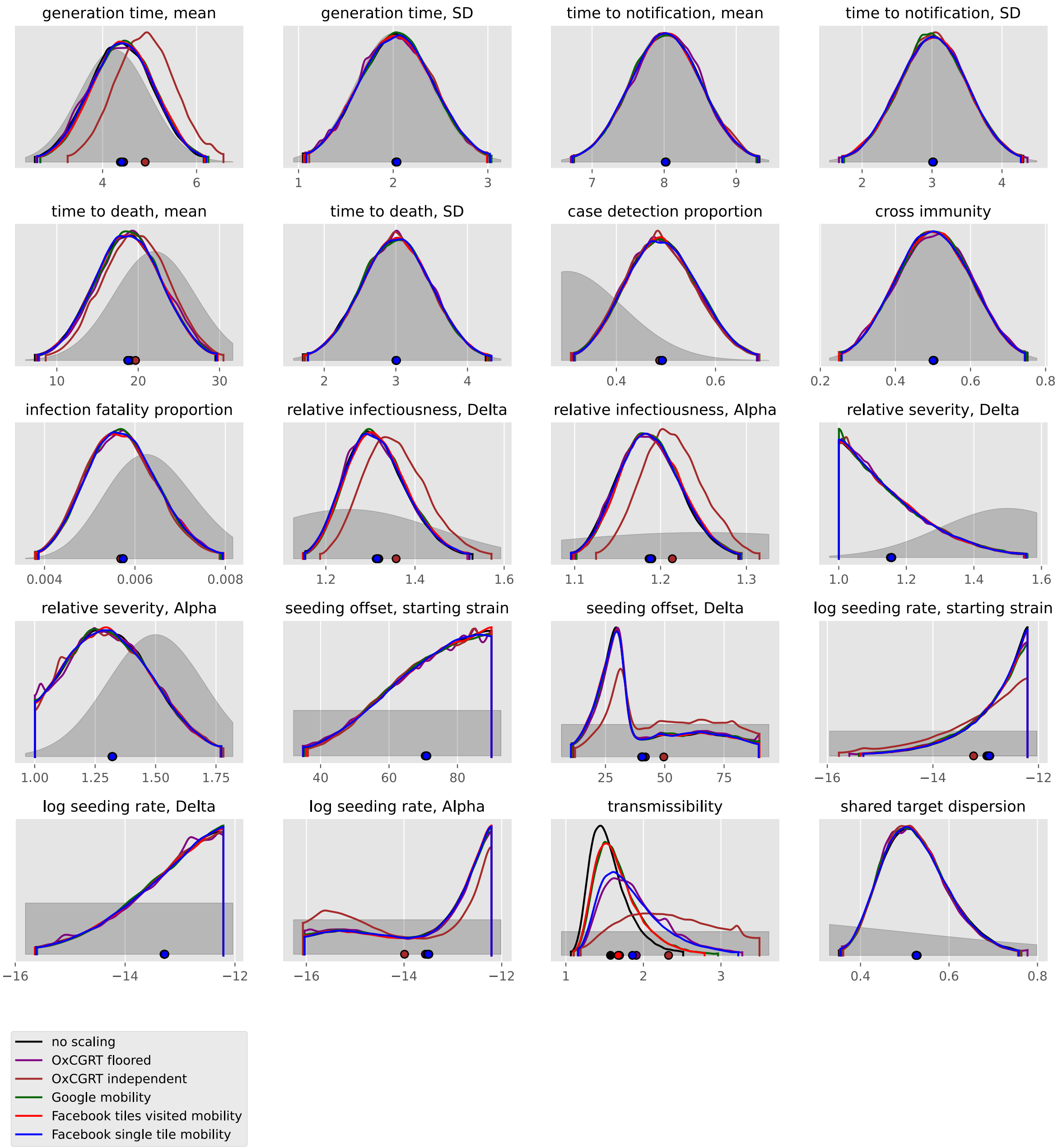
generation time, mean
generation time, SD
time to notification, mean
time to notification, SD
time to death, mean
time to death, SD
case detection proportion
cross immunity
infection fatality proportion
relative infectiousness, Delta
relative infectiousness, Alpha
relative severity, Delta
relative severity, Alpha
seeding offset, starting strain
seeding offset, Delta
log seeding rate, starting strain
log seeding rate, Delta
log seeding rate, Alpha
transmissibility
shared target dispersion
no scaling
OxCGRT floored
OxCGRT independent
Google mobility
Facebook tiles visited mobility
Facebook single tile mobility

#### Israel

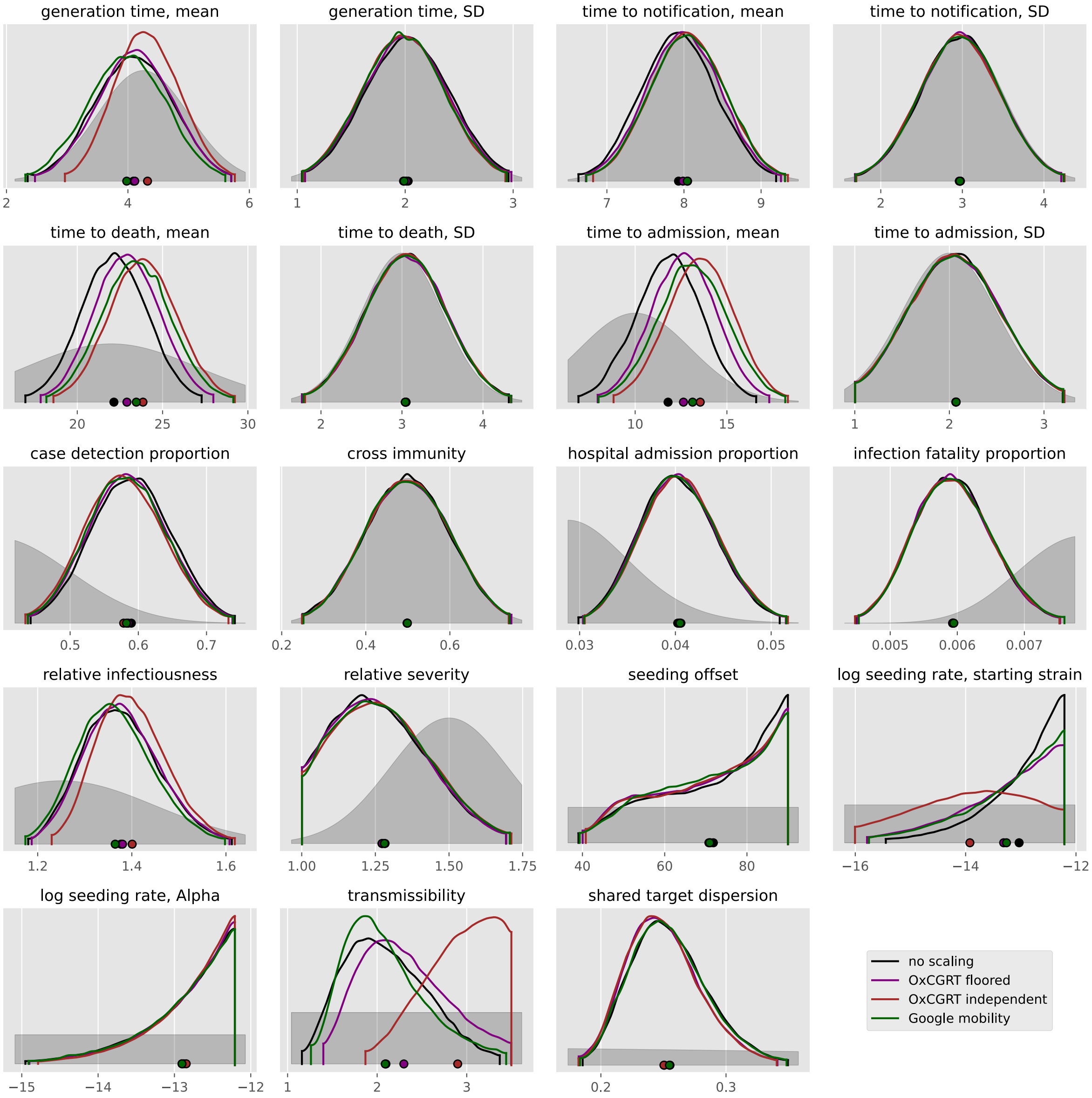
generation time, mean
generation time, SD
time to notification, mean
time to notification, SD
time to death, mean
time to death, SD
time to admission, mean
time to admission, SD
case detection proportion
cross immunity
hospital admission proportion
infection fatality proportion
relative infectiousness
relative severity
seeding offset
log seeding rate, starting strain
log seeding rate, Alpha
transmissibility
shared target dispersion
no scaling
OxCGRT floored
OxCGRT independent
Google mobility

#### Japan

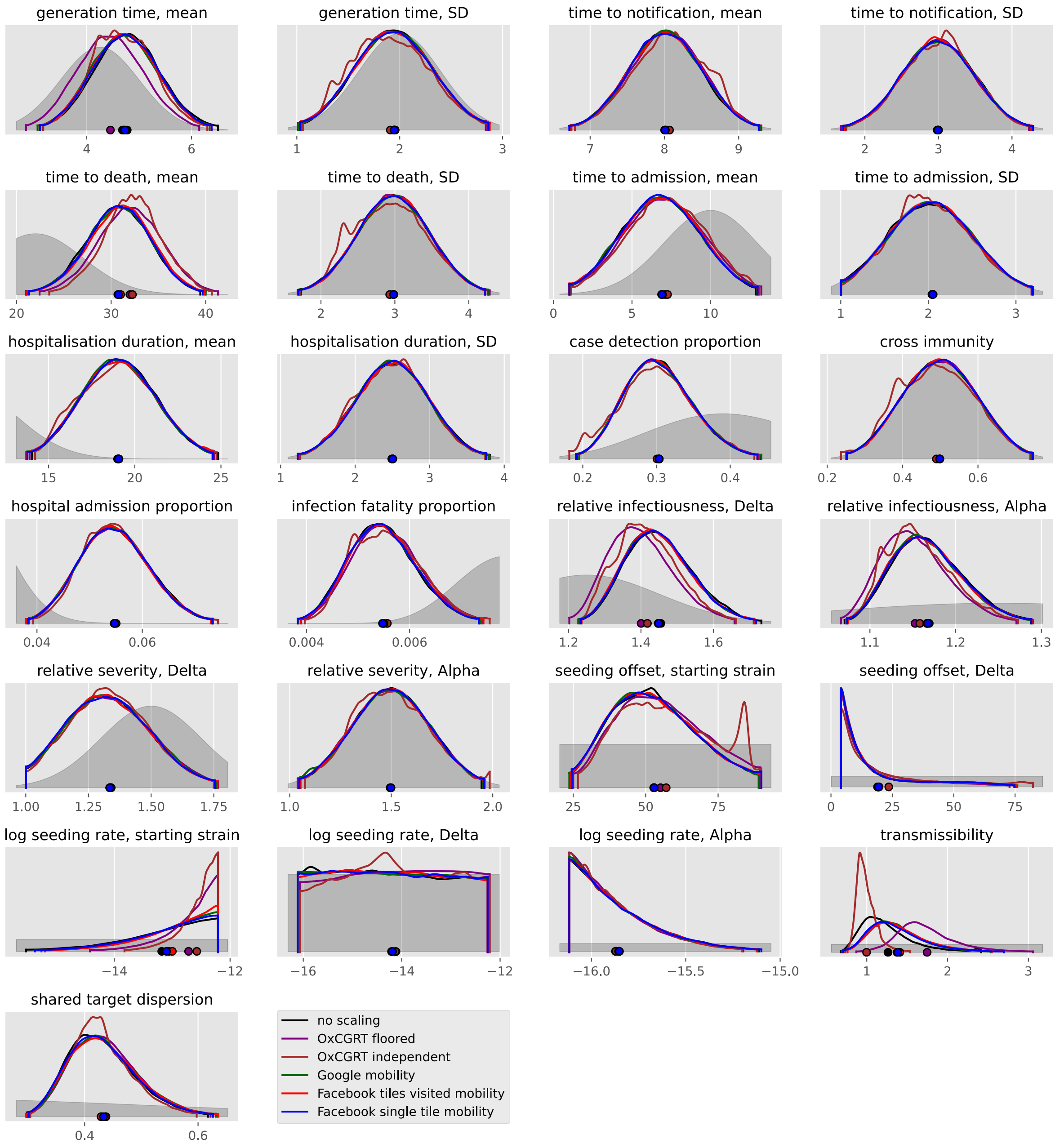

generation time, mean
generation time, SD
time to notification, mean
time to notification, SD
time to death, mean
time to death, SD
time to admission, mean
time to admission, SD
hospitalisation duration, mean
hospitalisation duration, SD
case detection proportion
cross immunity
hospital admission proportion
infection fatality proportion
relative infectiousness, Delta
relative infectiousness, Alpha
relative severity, Delta
relative severity, Alpha
seeding offset, starting strain
seeding offset, Delta
log seeding rate, starting strain
log seeding rate, Delta
log seeding rate, Alpha
transmissibility
shared target dispersion
no scaling
OxCGRT floored
OxCGRT independent
Google mobility
Facebook tiles visited mobility
Facebook single tile mobility

#### Jordan

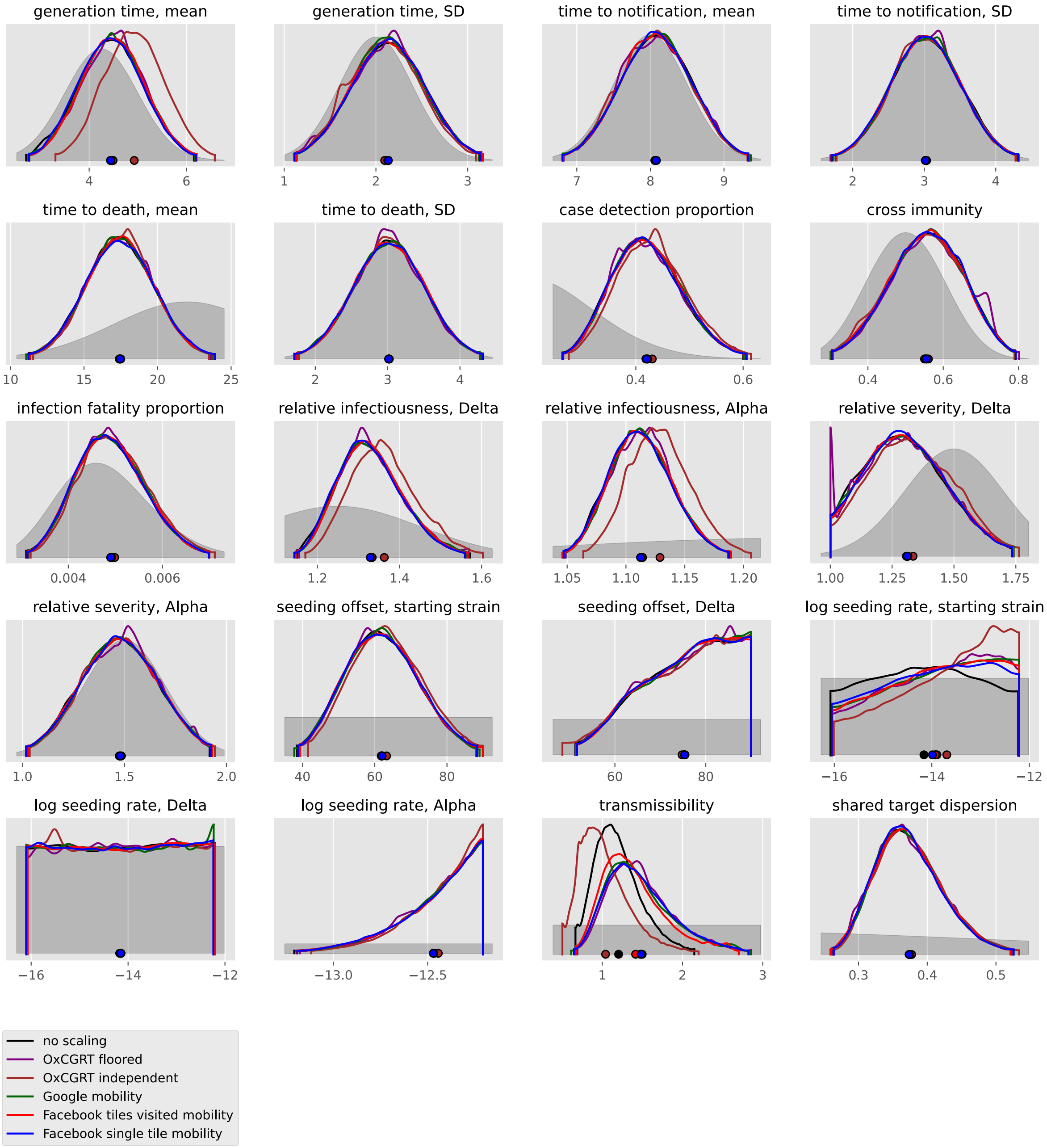

generation time, mean
generation time, SD
time to notification, mean
time to notification, SD
time to death, mean
time to death, SD
case detection proportion
cross immunity
infection fatality proportion
relative infectiousness, Delta
relative infectiousness, Alpha
relative severity, Delta
relative severity, Alpha
seeding offset, starting strain
seeding offset, Delta
log seeding rate, starting strain
log seeding rate, Delta
log seeding rate, Alpha
transmissibility
shared target dispersion
no scaling
OxCGRT floored
OxCGRT independent
Google mobility
Facebook tiles visited mobility
Facebook single tile mobility

#### Korea, Republic of

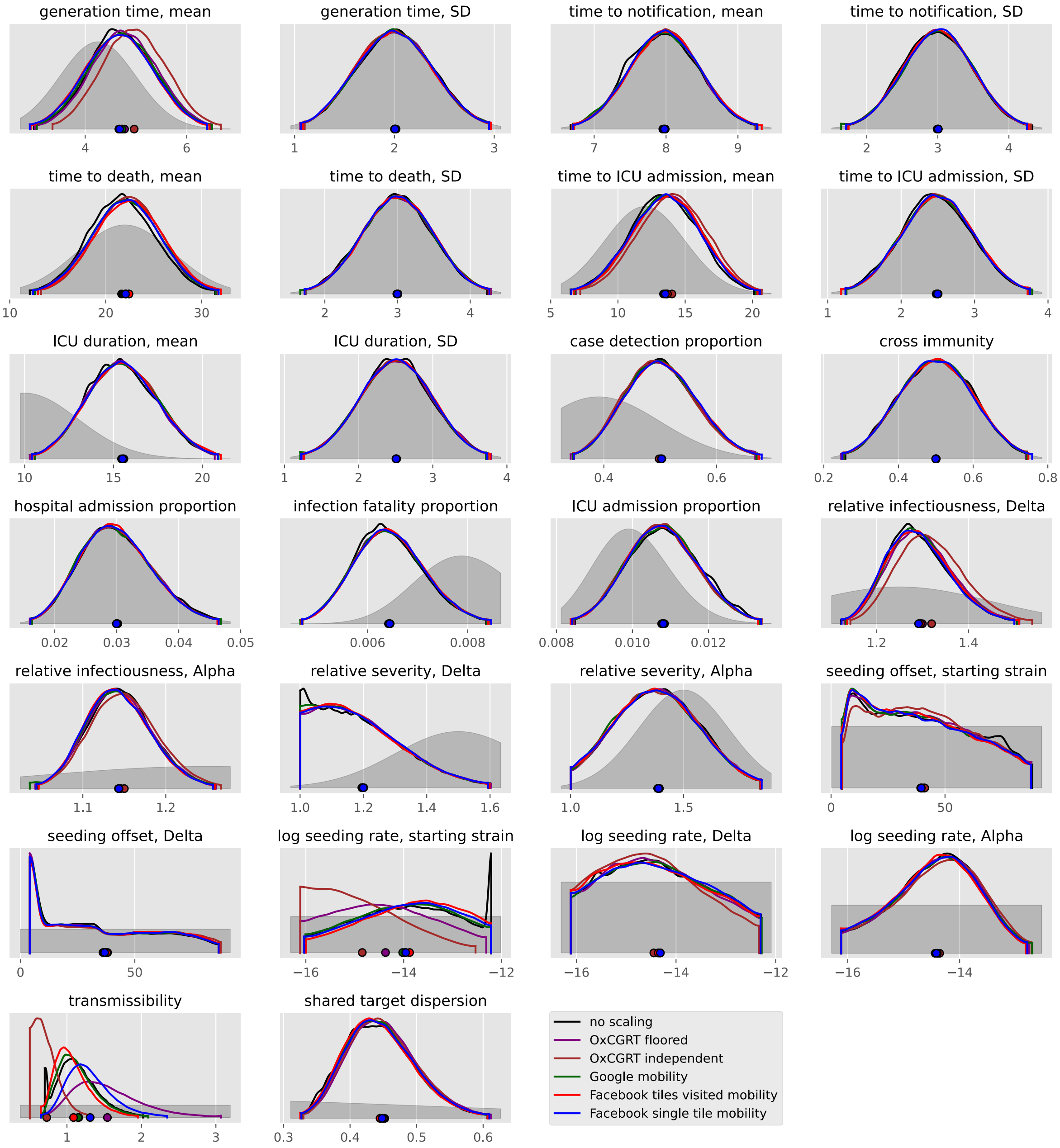

#### Kuwait

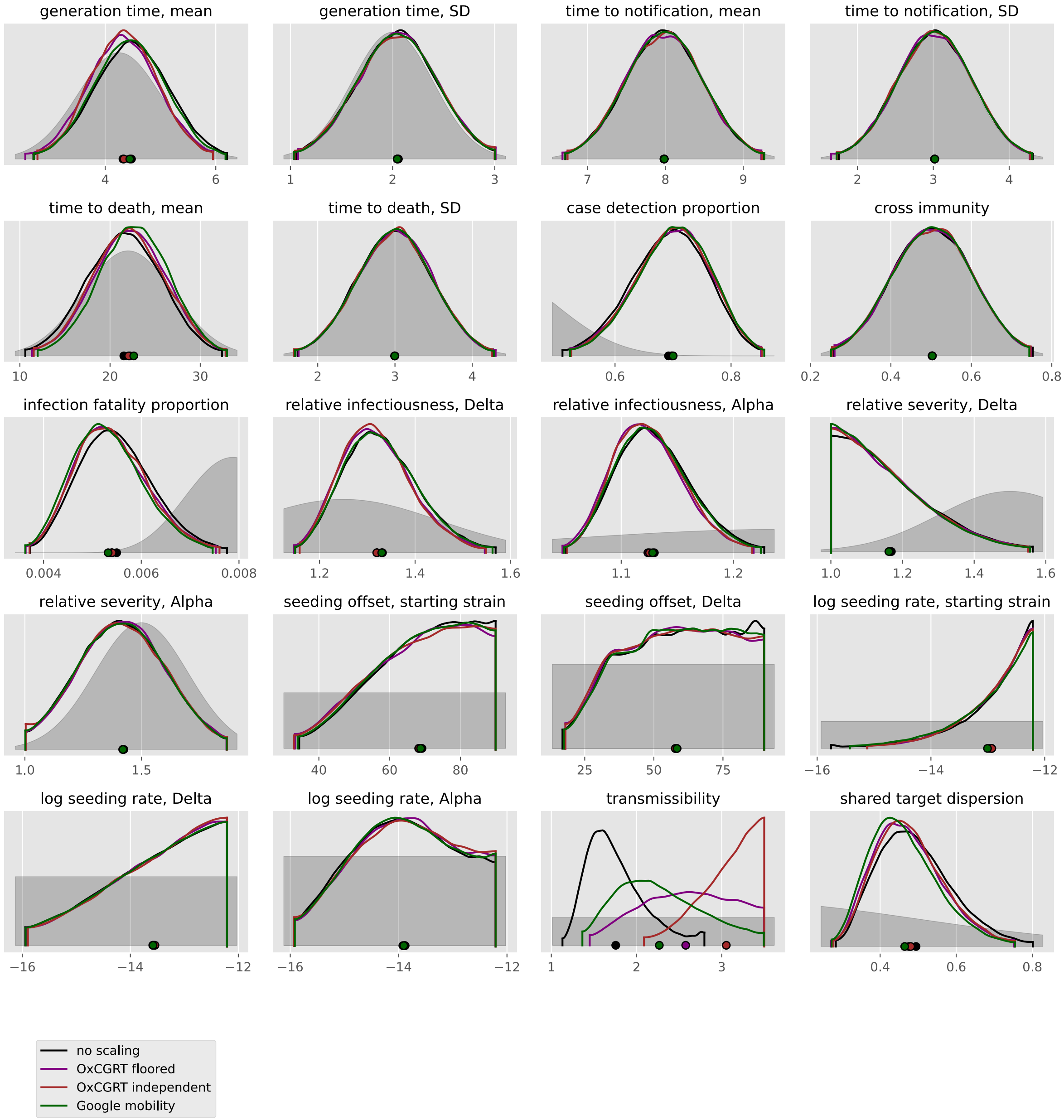

generation time, mean
generation time, SD
time to notification, mean
time to notification, SD
time to death, mean
time to death, SD
case detection proportion
cross immunity
infection fatality proportion
relative infectiousness, Delta
relative infectiousness, Alpha
relative severity, Delta
relative severity, Alpha
seeding offset, starting strain
seeding offset, Delta
log seeding rate, starting strain
log seeding rate, Delta
log seeding rate, Alpha
transmissibility
shared target dispersion
no scaling
OxCGRT floored
OxCGRT independent
Google mobility

#### Lebanon

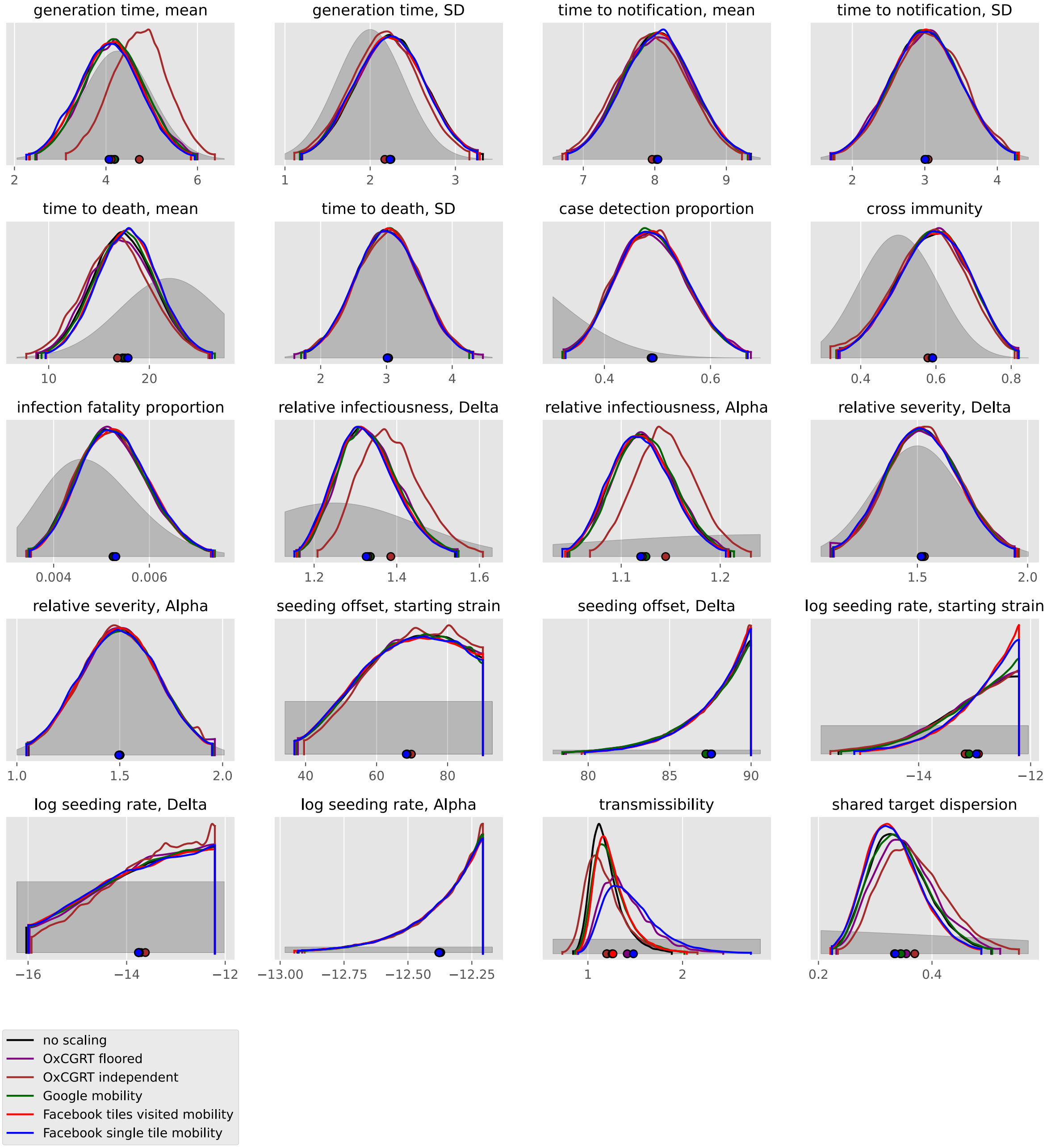
generation time, mean
2
4
6
generation time, SD
1
2
3
time to notification, mean
7
8
9
time to notification, SD
2
3
4
time to death, mean
10
20
time to death, SD
2
3
4
case detection proportion
0.4
0.6
cross immunity
0.4
0.6
0.8
infection fatality proportion
0.004
0.006
relative infectiousness, Delta
1.2
1.4
1.6
relative infectiousness, Alpha
1.1
1.2
relative severity, Delta
1.5
2.0
relative severity, Alpha
1.0
1.5
2.0
seeding offset, starting strain
40
60
80
seeding offset, Delta
80
85
90
log seeding rate, starting strain
−14
−12
log seeding rate, Delta
−16
−14
−12
log seeding rate, Alpha
−13.00
−12.75
−12.50
−12.25
transmissibility
1
2
shared target dispersion
0.2
0.4
no scaling
OxCGRT floored
OxCGRT independent
Google mobility
Facebook tiles visited mobility
Facebook single tile mobility

#### Malaysia

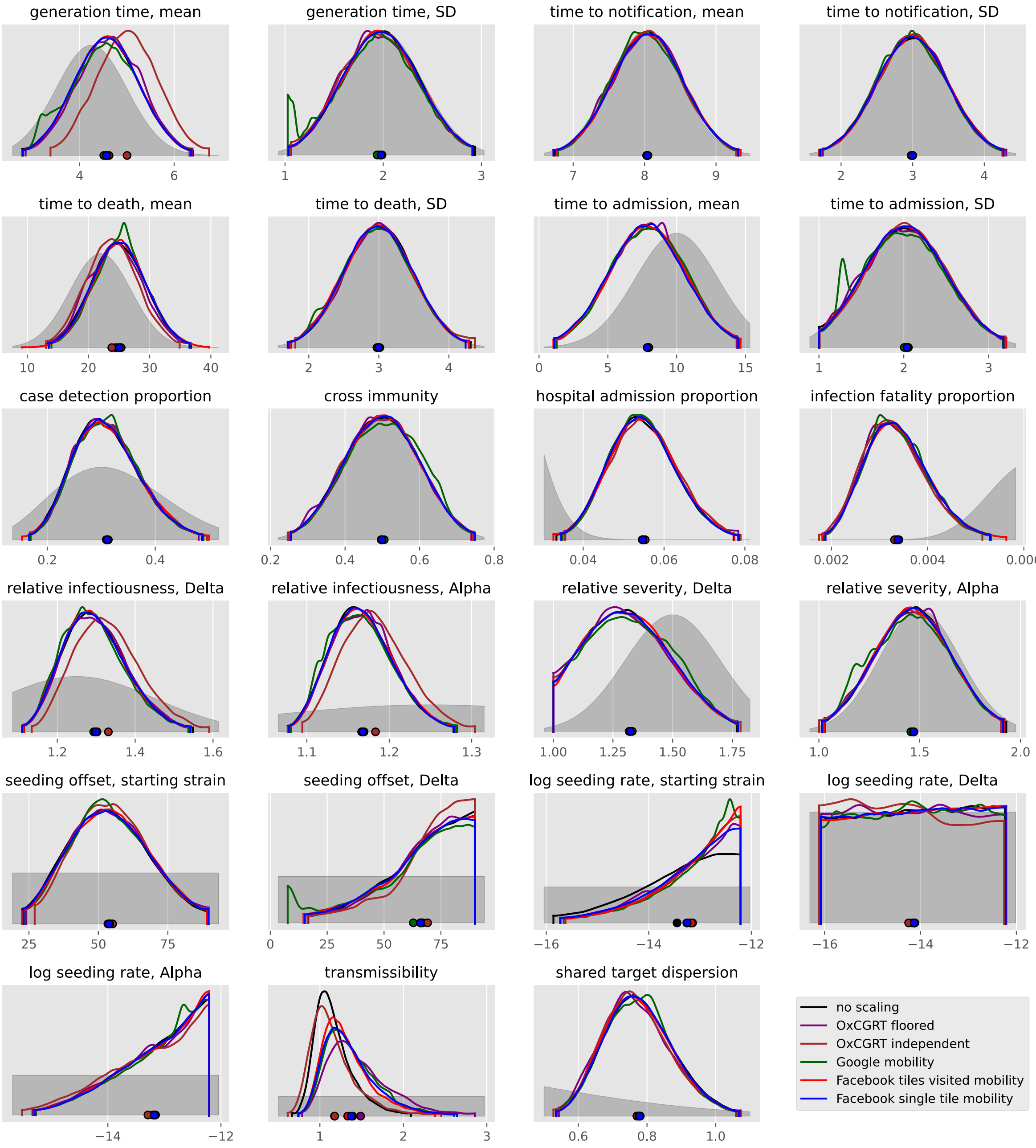

generation time, mean
4
6
generation time, SD
1
2
3
time to notification, mean
7
8
9
time to notification, SD
2
3
4
time to death, mean
10
20
30
40
time to death, SD
2
3
4
time to admission, mean
0
5
10
15
time to admission, SD
1
2
3
case detection proportion
0.2
0.4
cross immunity
0.2
0.4
0.6
0.8
hospital admission proportion
0.04
0.06
0.08
infection fatality proportion
0.002
0.004
0.006
relative infectiousness, Delta
1.2
1.4
1.6
relative infectiousness, Alpha
1.1
1.2
1.3
relative severity, Delta
1.00
1.25
1.50
1.75
relative severity, Alpha
1.0
1.5
2.0
seeding offset, starting strain
25
50
75
seeding offset, Delta
0
25
50
75
log seeding rate, starting strain
−16
−14
−12
log seeding rate, Delta
−16
−14
−12
log seeding rate, Alpha
−14
−12
transmissibility
1
2
3
shared target dispersion
0.6
0.8
1.0
no scaling
OxCGRT floored
OxCGRT independent
Google mobility
Facebook tiles visited mobility
Facebook single tile mobility

#### Nepal

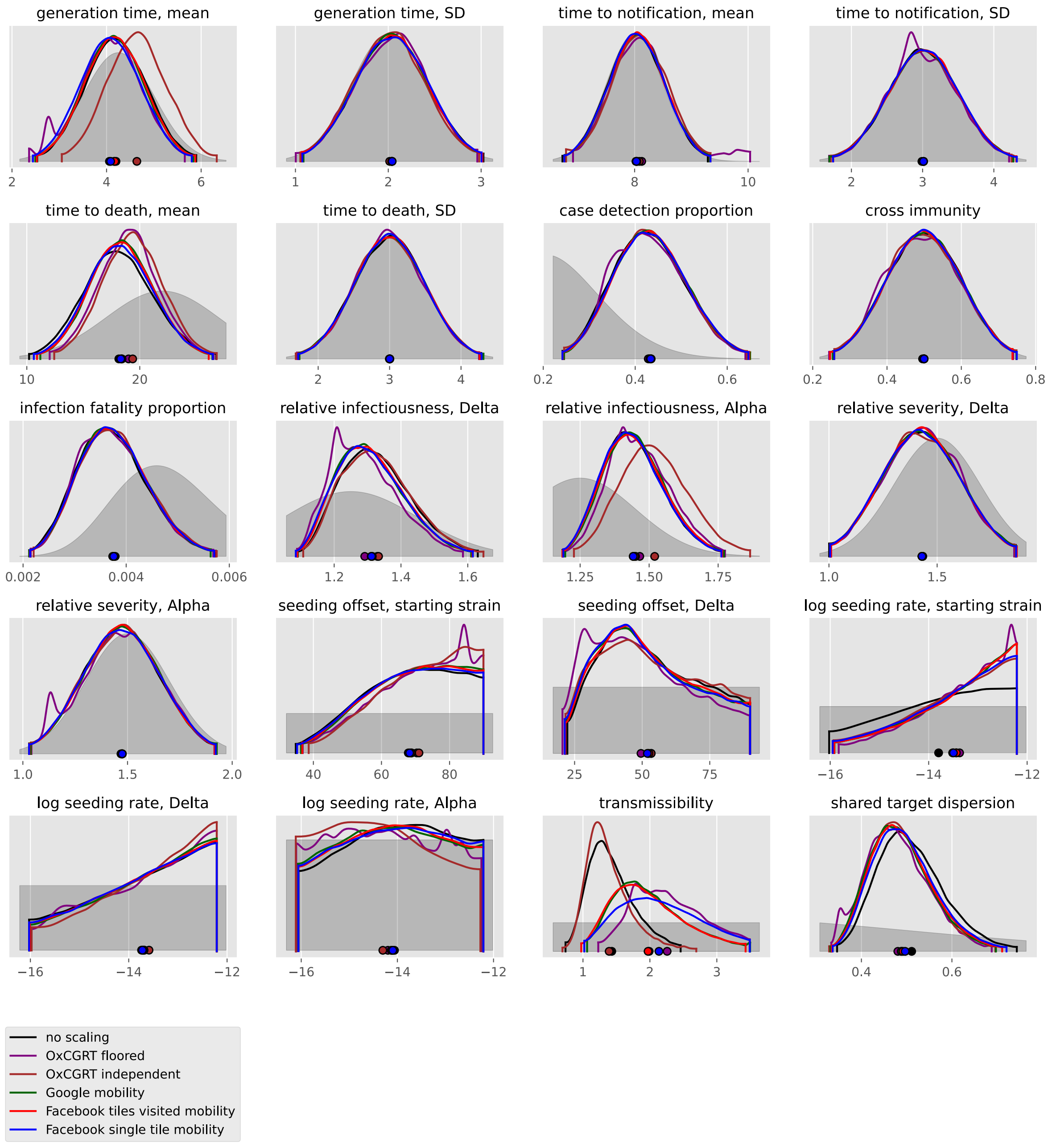
generation time, mean
generation time, SD
time to notification, mean
time to notification, SD
time to death, mean
time to death, SD
case detection proportion
cross immunity
infection fatality proportion
relative infectiousness, Delta
relative infectiousness, Alpha
relative severity, Delta
relative severity, Alpha
seeding offset, starting strain
seeding offset, Delta
log seeding rate, starting strain
log seeding rate, Delta
log seeding rate, Alpha
transmissibility
shared target dispersion
no scaling
OxCGRT floored
OxCGRT independent
Google mobility
Facebook tiles visited mobility
Facebook single tile mobility

#### Oman

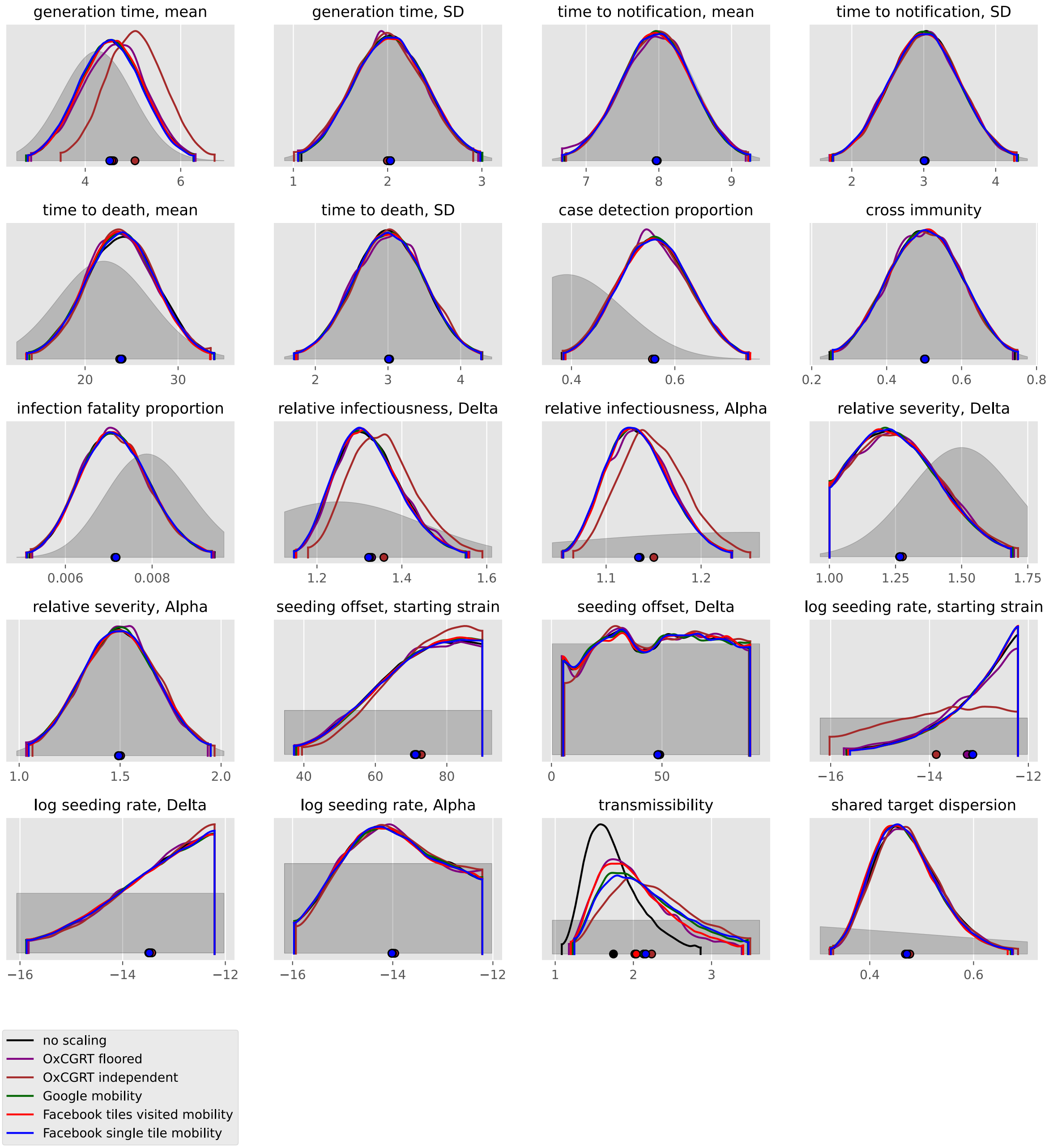

generation time, mean
generation time, SD
time to notification, mean
time to notification, SD
time to death, mean
time to death, SD
case detection proportion
cross immunity
infection fatality proportion
relative infectiousness, Delta
relative infectiousness, Alpha
relative severity, Delta
relative severity, Alpha
seeding offset, starting strain
seeding offset, Delta
log seeding rate, starting strain
log seeding rate, Delta
log seeding rate, Alpha
transmissibility
shared target dispersion
no scaling
OxCGRT floored
OxCGRT independent
Google mobility
Facebook tiles visited mobility
Facebook single tile mobility

#### Pakistan

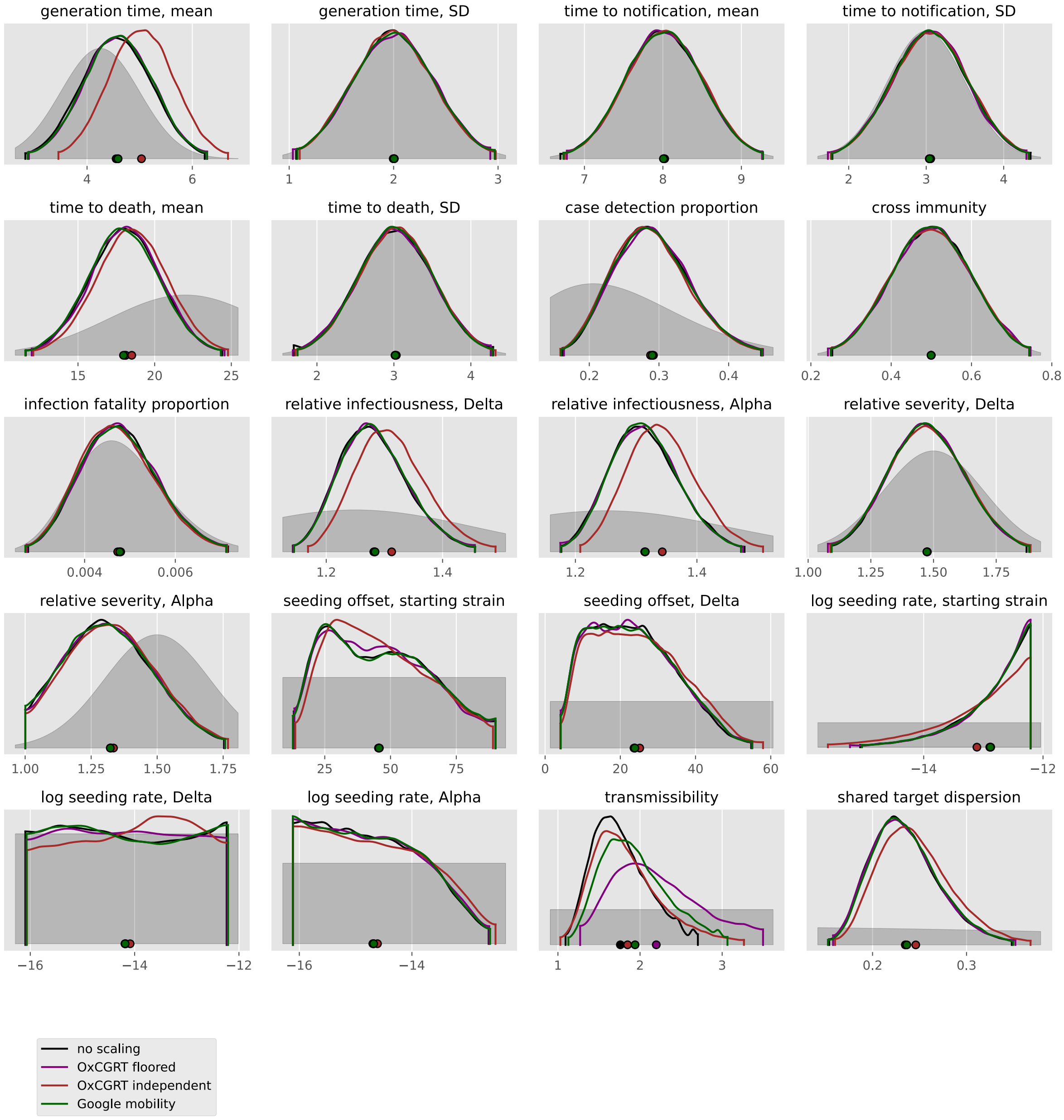
generation time, mean
generation time, SD
time to notification, mean
time to notification, SD
time to death, mean
time to death, SD
case detection proportion
cross immunity
infection fatality proportion
relative infectiousness, Delta
relative infectiousness, Alpha
relative severity, Delta
relative severity, Alpha
seeding offset, starting strain
seeding offset, Delta
log seeding rate, starting strain
log seeding rate, Delta
log seeding rate, Alpha
transmissibility
shared target dispersion
no scaling
OxCGRT floored
OxCGRT independent
Google mobility

#### Palestine, State of

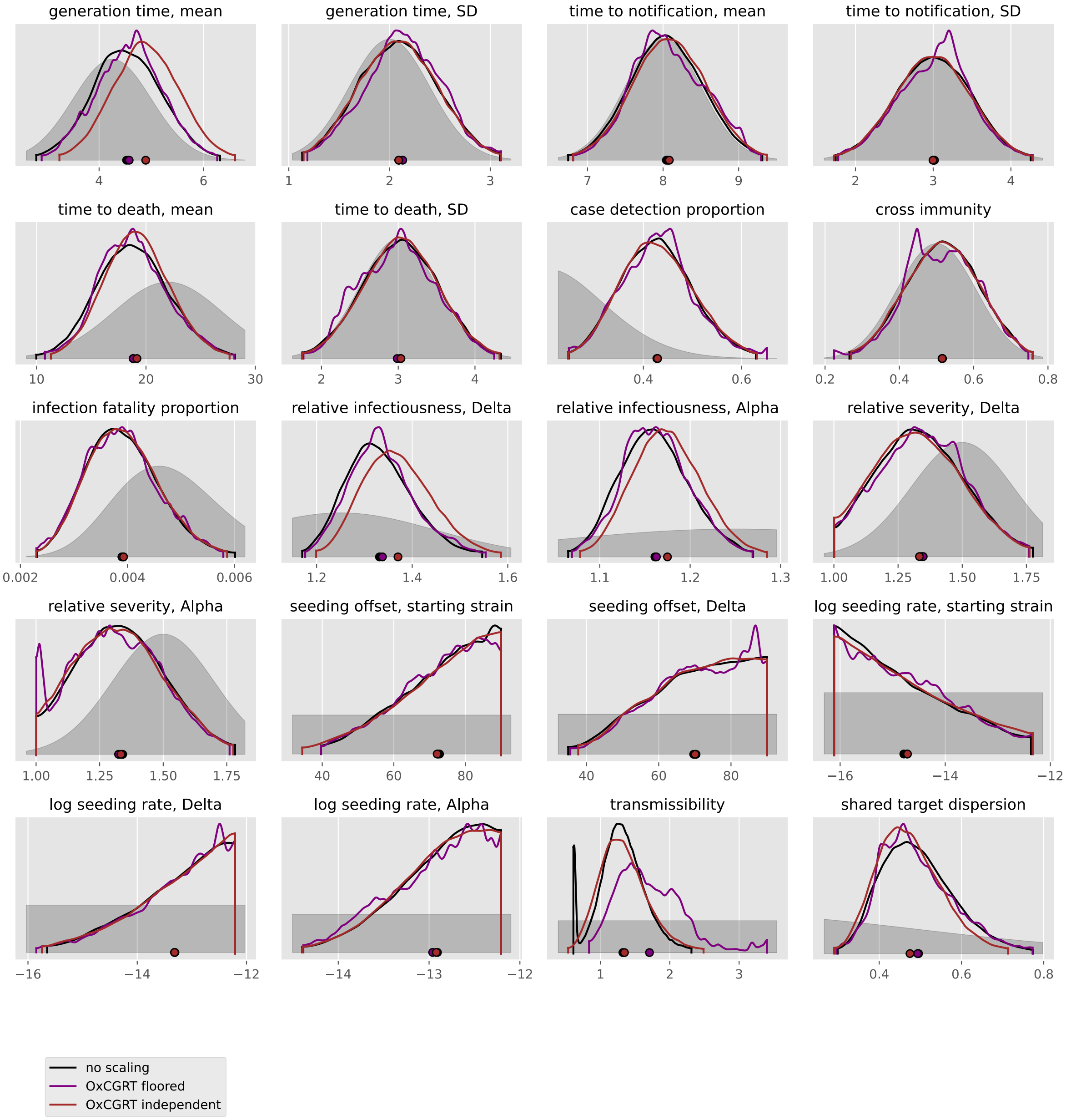

#### Philippines

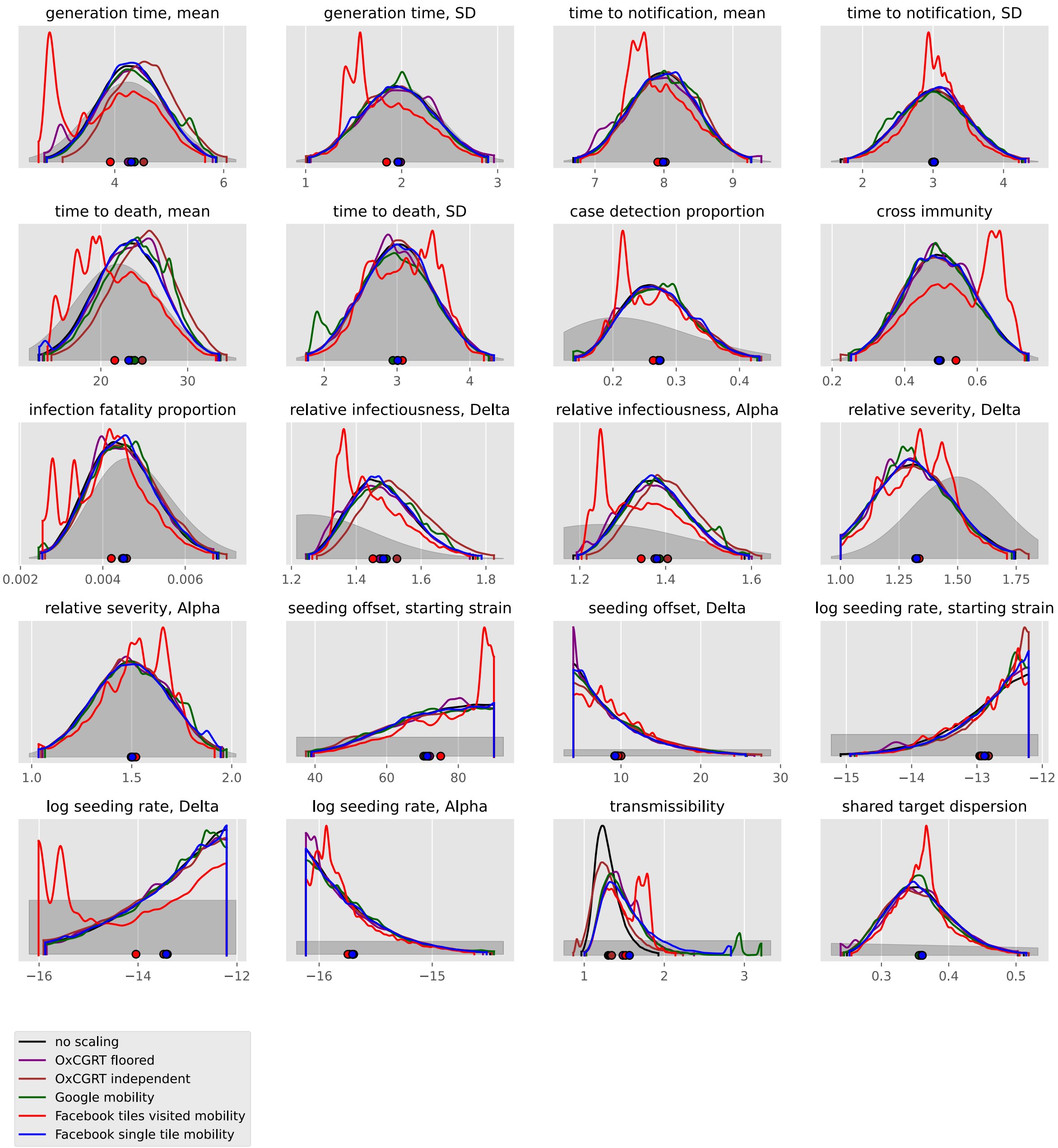

#### Qatar

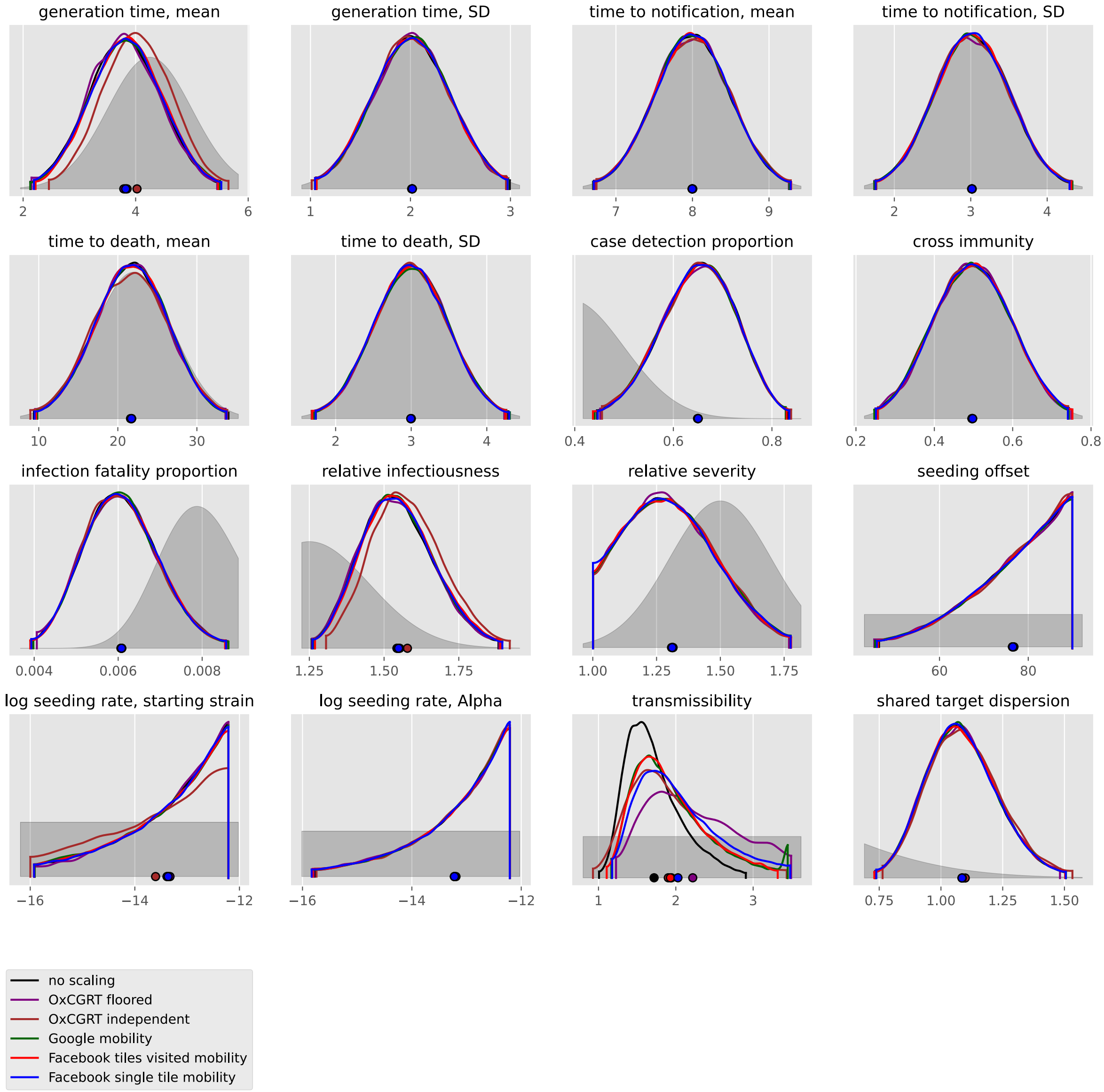

generation time, mean
generation time, SD
time to notification, mean
time to notification, SD
time to death, mean
time to death, SD
case detection proportion
cross immunity
infection fatality proportion
relative infectiousness
relative severity
seeding offset
log seeding rate, starting strain
log seeding rate, Alpha
transmissibility
shared target dispersion
no scaling
OxCGRT floored
OxCGRT independent
Google mobility
Facebook tiles visited mobility
Facebook single tile mobility

#### Saudi Arabia

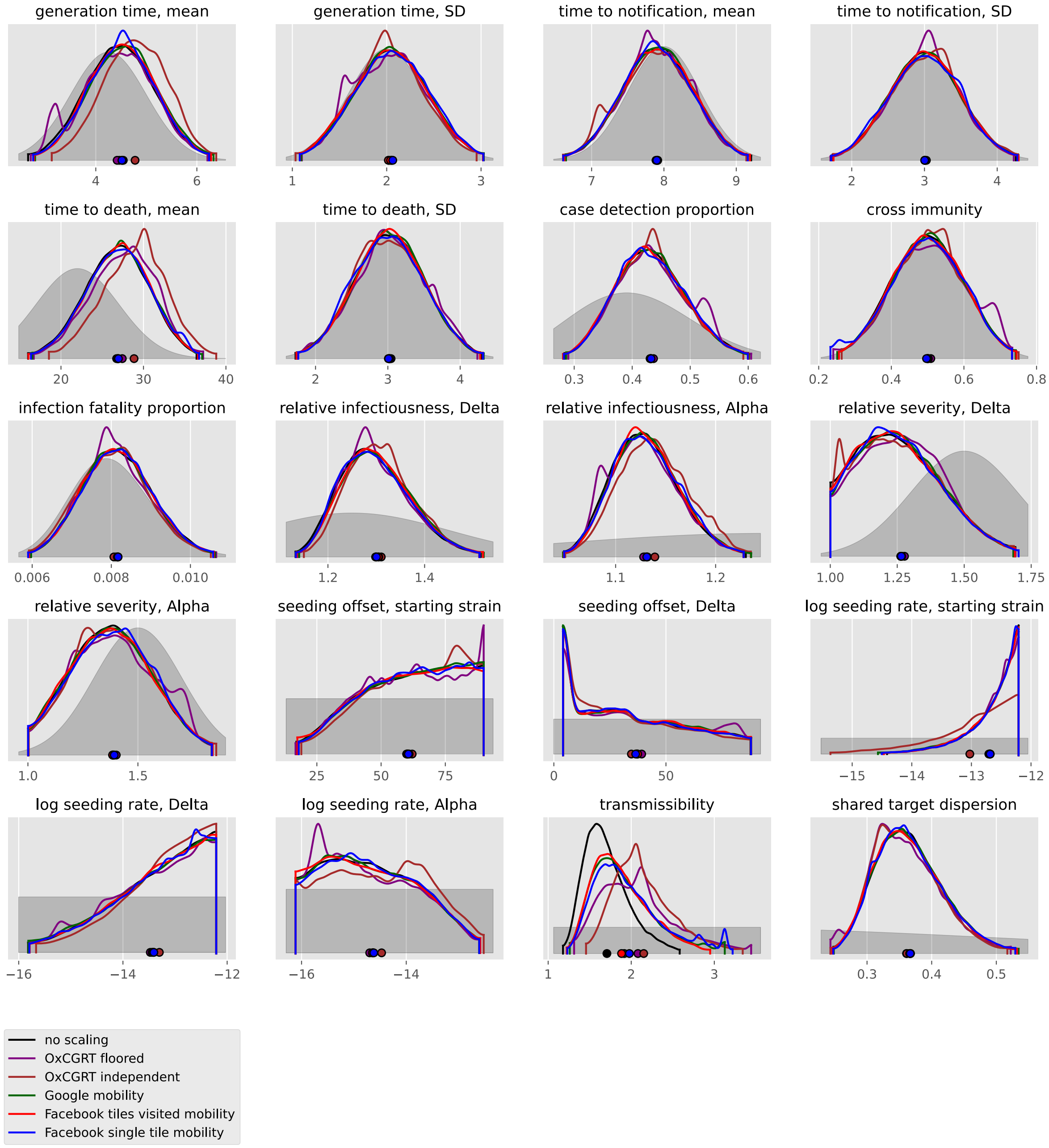

#### Sri Lanka

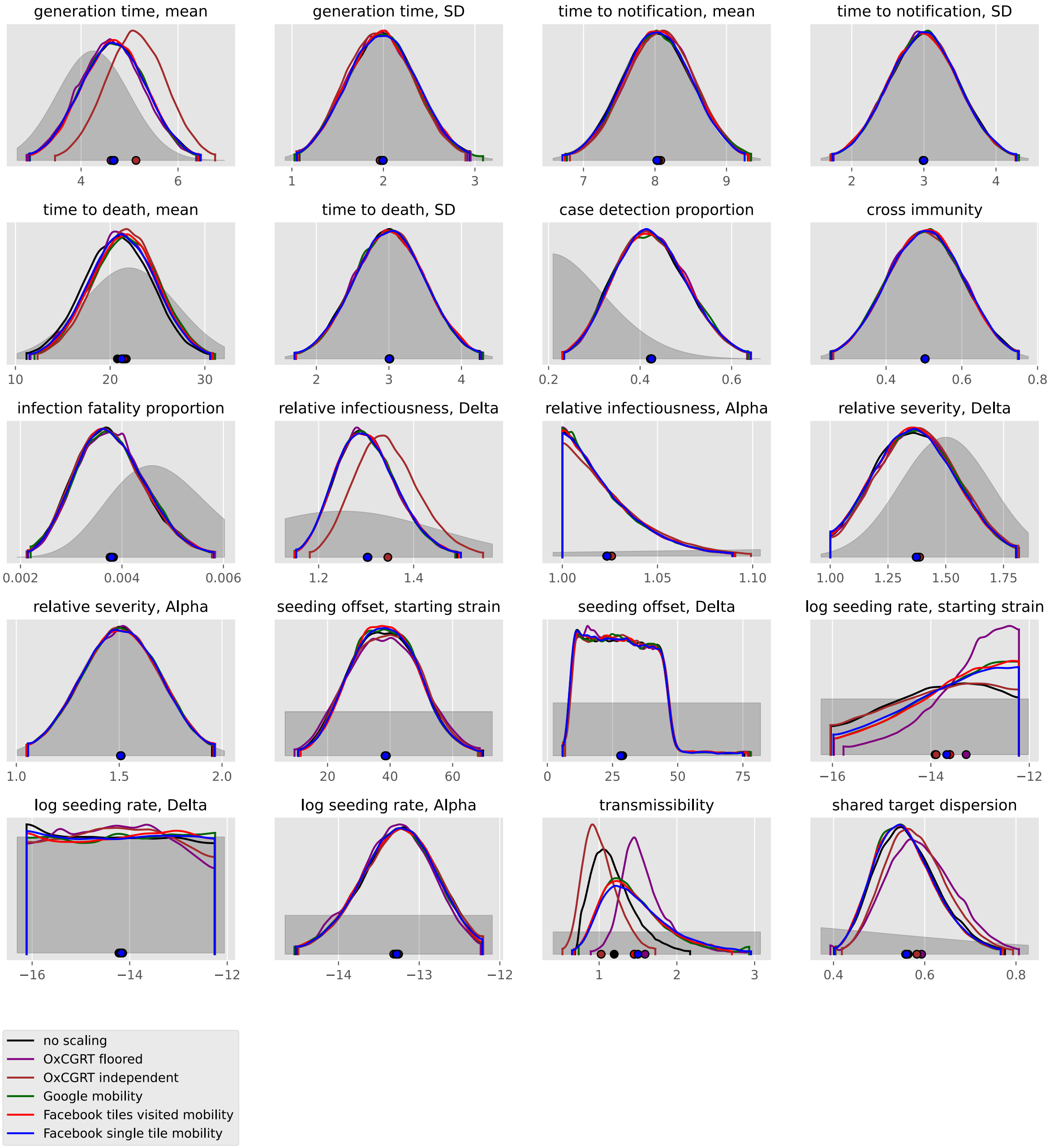
generation time, mean
generation time, SD
time to notification, mean
time to notification, SD
time to death, mean
time to death, SD
case detection proportion
cross immunity
infection fatality proportion
relative infectiousness, Delta
relative infectiousness, Alpha
relative severity, Delta
relative severity, Alpha
seeding offset, starting strain
seeding offset, Delta
log seeding rate, starting strain
log seeding rate, Delta
log seeding rate, Alpha
transmissibility
shared target dispersion
no scaling
OxCGRT floored
OxCGRT independent
Google mobility
Facebook tiles visited mobility
Facebook single tile mobility

#### Syrian Arab Republic

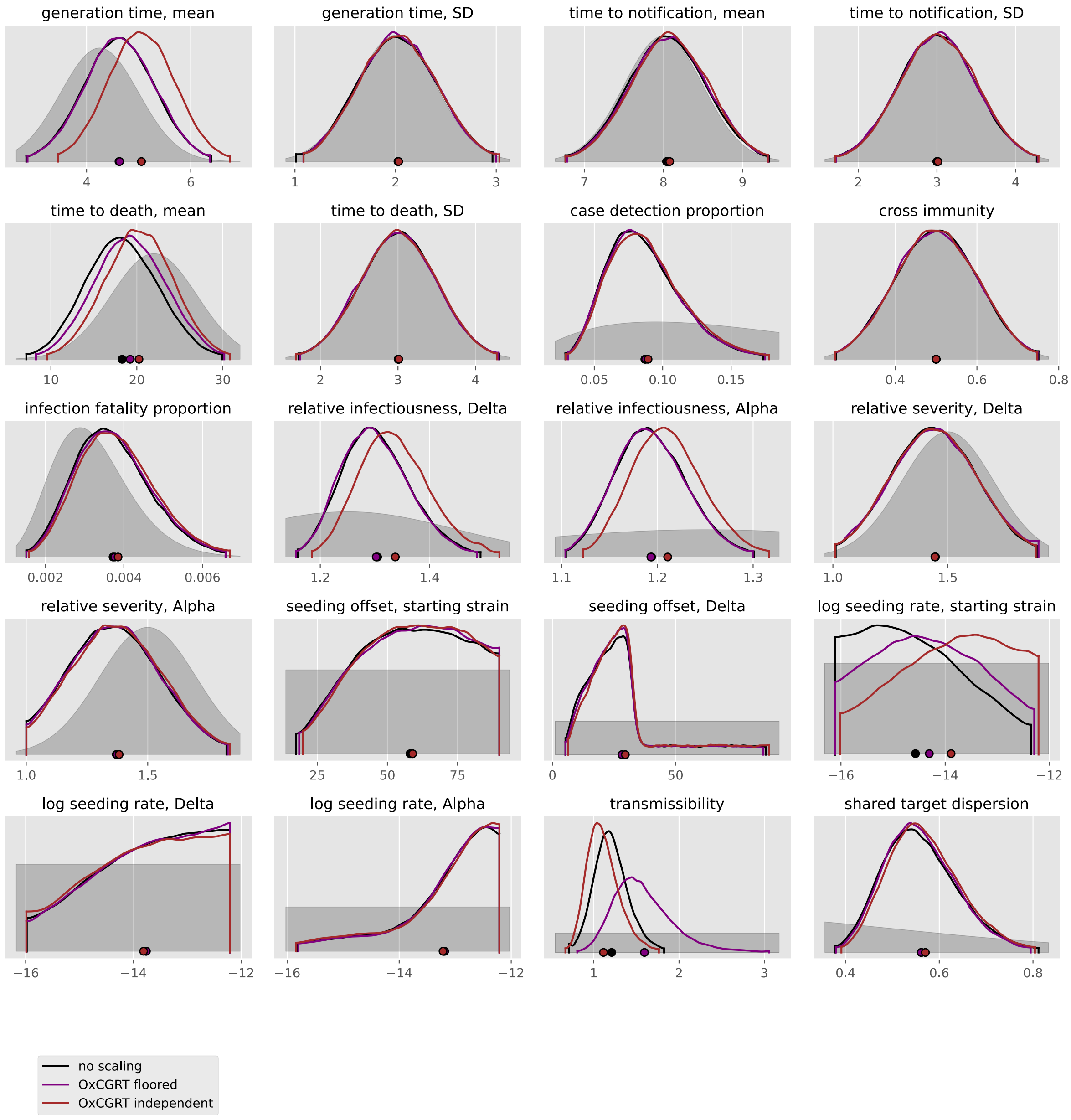

#### Türkiye

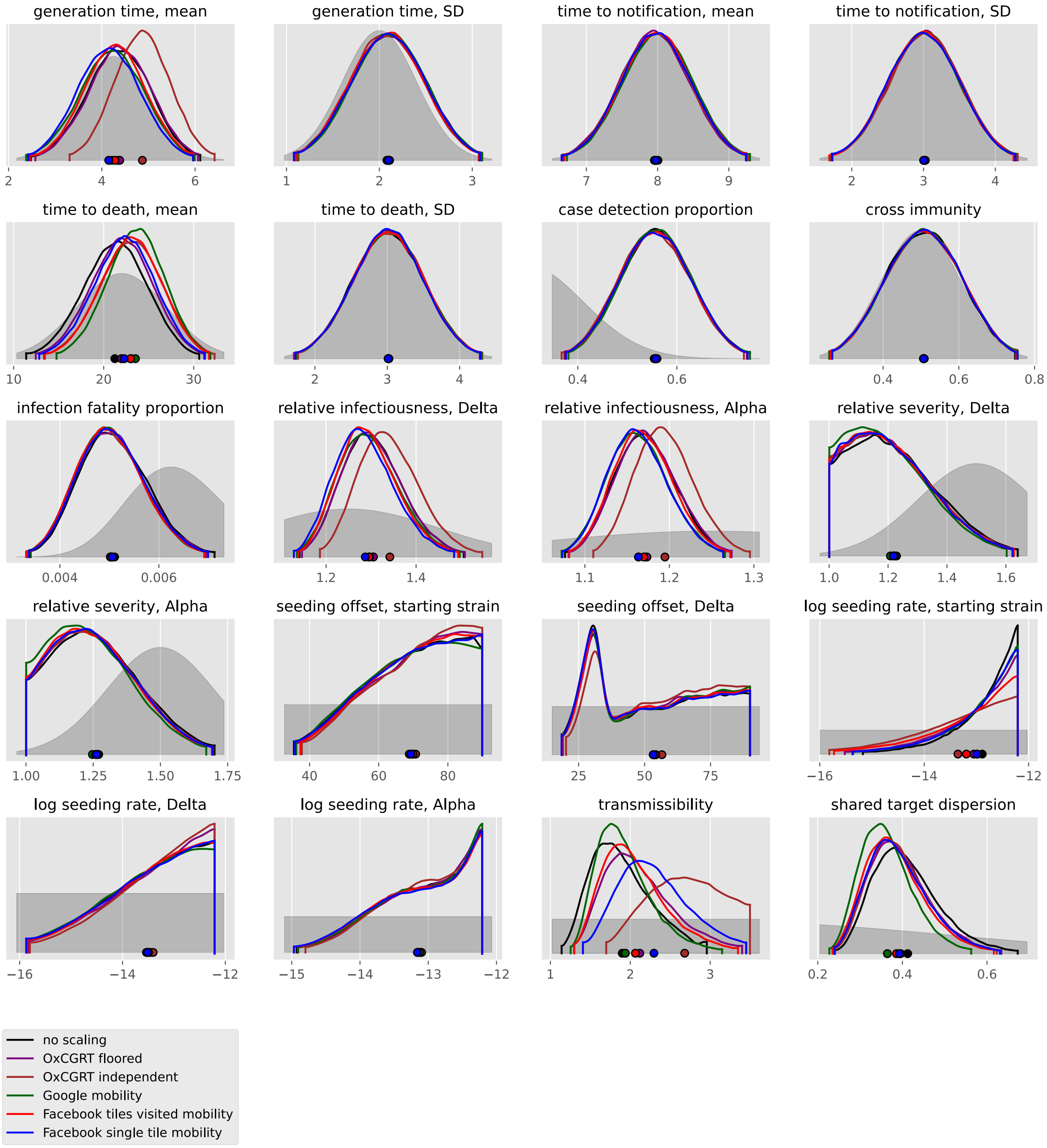
generation time, mean
2
4
6
generation time, SD
1
2
3
time to notification, mean
7
8
9
time to notification, SD
2
3
4
time to death, mean
10
20
30
time to death, SD
2
3
4
case detection proportion
0.4
0.6
cross immunity
0.4
0.6
0.8
infection fatality proportion
0.004
0.006
relative infectiousness, Delta
1.2
1.4
relative infectiousness, Alpha
1.1
1.2
1.3
relative severity, Delta
1.0
1.2
1.4
1.6
relative severity, Alpha
1.00
1.25
1.50
1.75
seeding offset, starting strain
40
60
80
seeding offset, Delta
25
50
75
log seeding rate, starting strain
−16
−14
−12
log seeding rate, Delta
−16
−14
−12
log seeding rate, Alpha
−15
−14
−13
−12
transmissibility
1
2
3
shared target dispersion
0.2
0.4
0.6
no scaling
OxCGRT floored
OxCGRT independent
Google mobility
Facebook tiles visited mobility
Facebook single tile mobility

#### United Arab Emirates

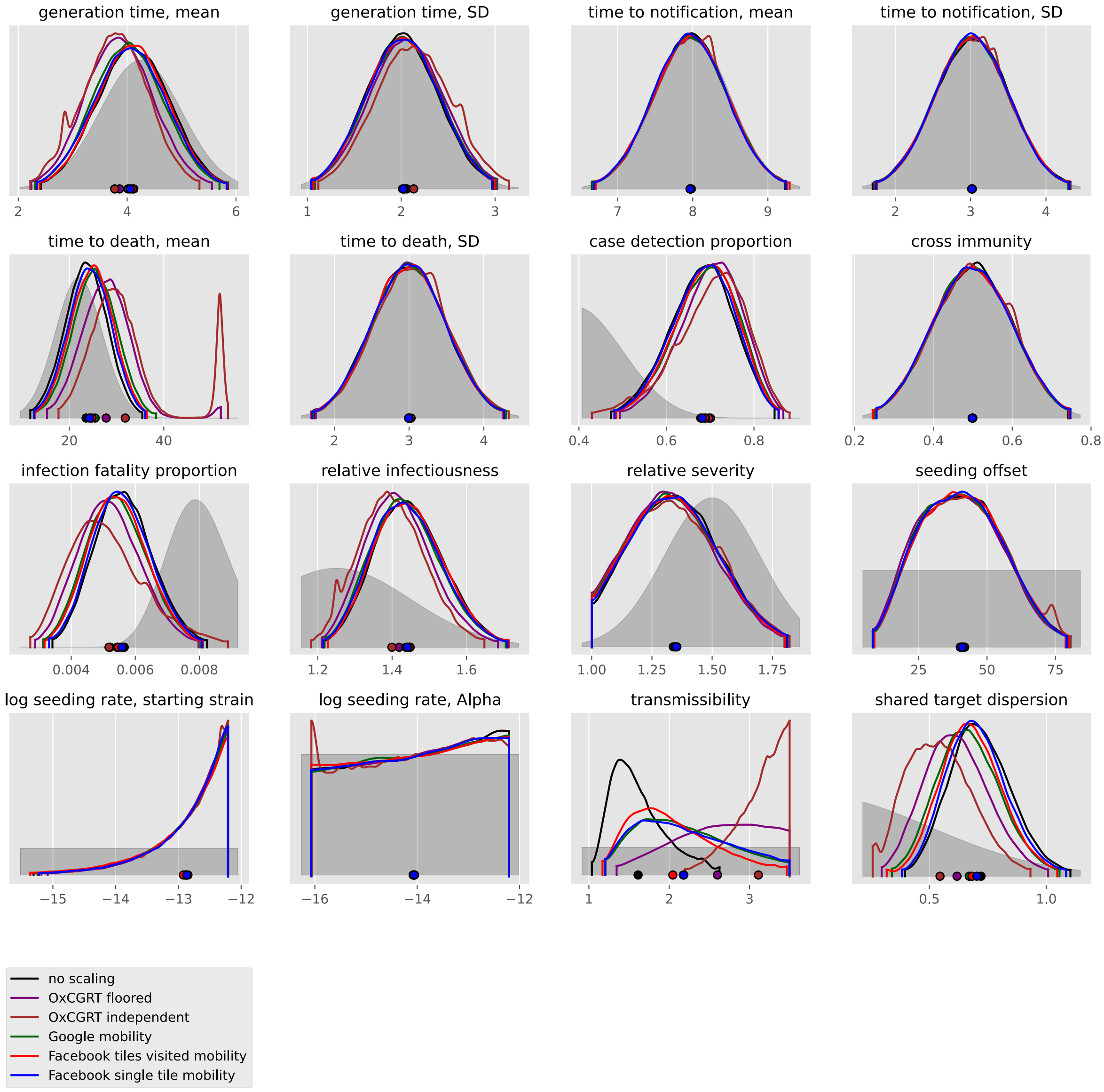

#### Yemen

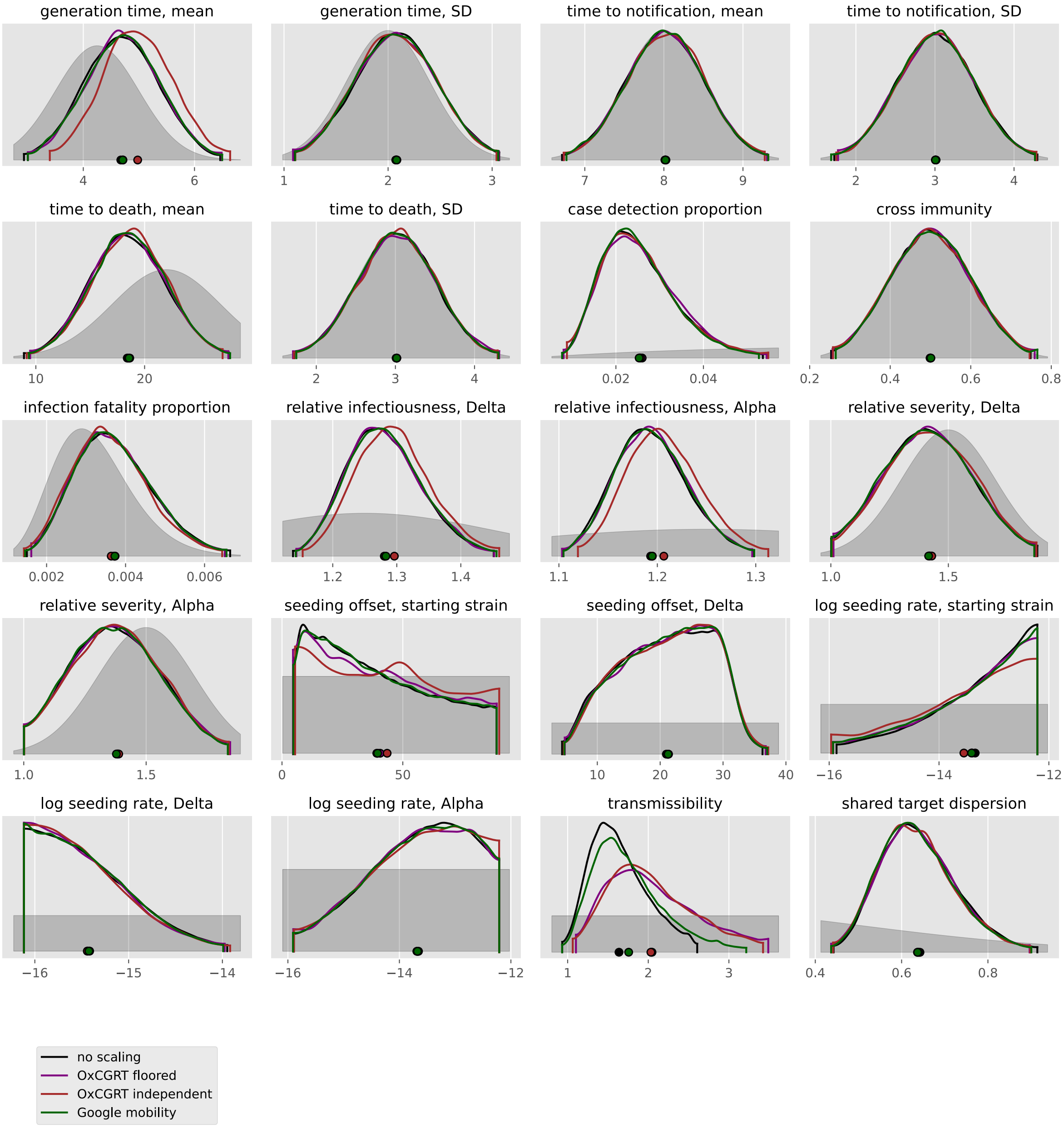

generation time, mean
generation time, SD
time to notification, mean
time to notification, SD
time to death, mean
time to death, SD
case detection proportion
cross immunity
infection fatality proportion
relative infectiousness, Delta
relative infectiousness, Alpha
relative severity, Delta
relative severity, Alpha
seeding offset, starting strain
seeding offset, Delta
log seeding rate, starting strain
log seeding rate, Delta
log seeding rate, Alpha
transmissibility
shared target dispersion
no scaling
OxCGRT floored
OxCGRT independent
Google mobility

### Europe

#### Albania

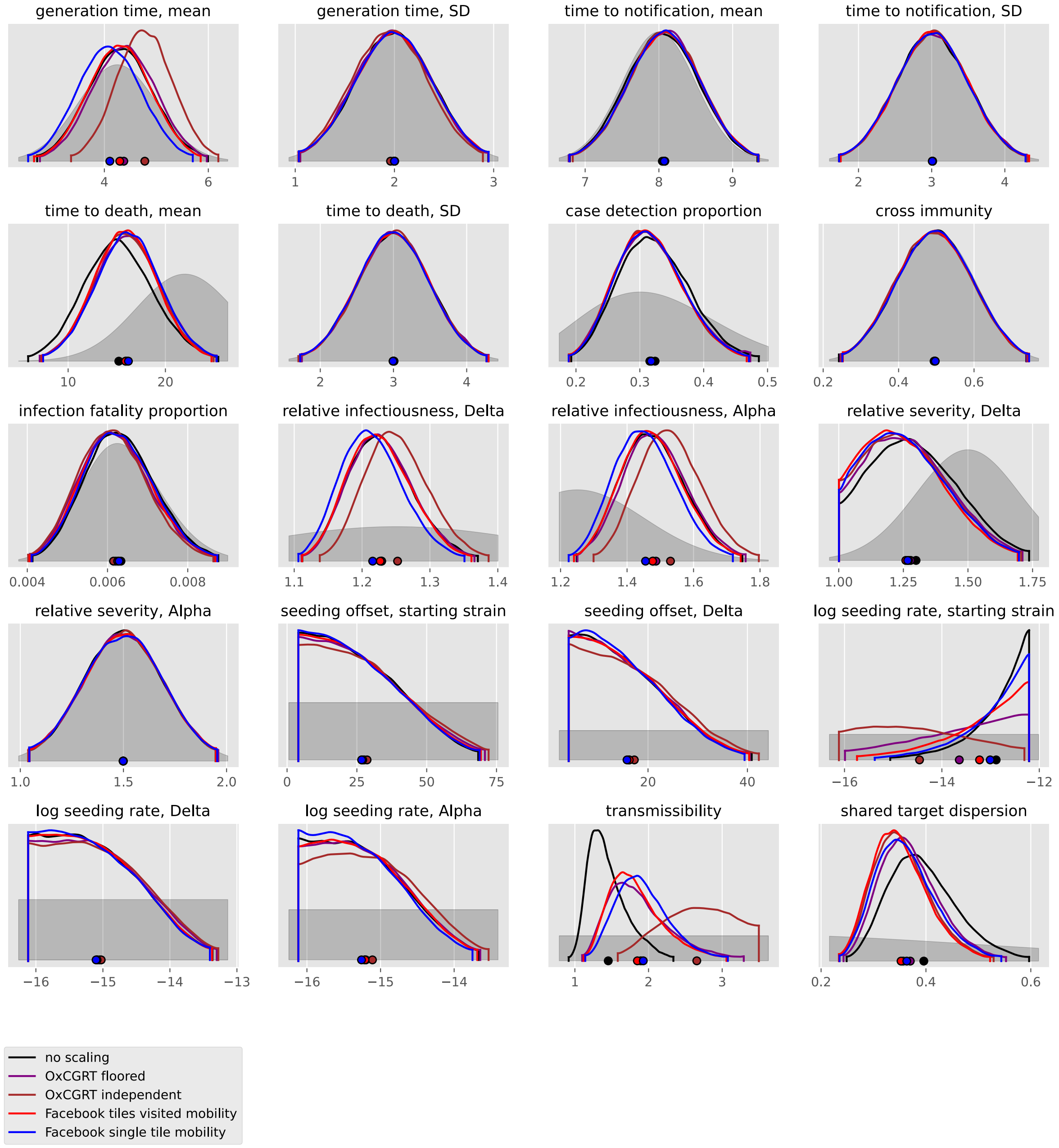

#### Andorra

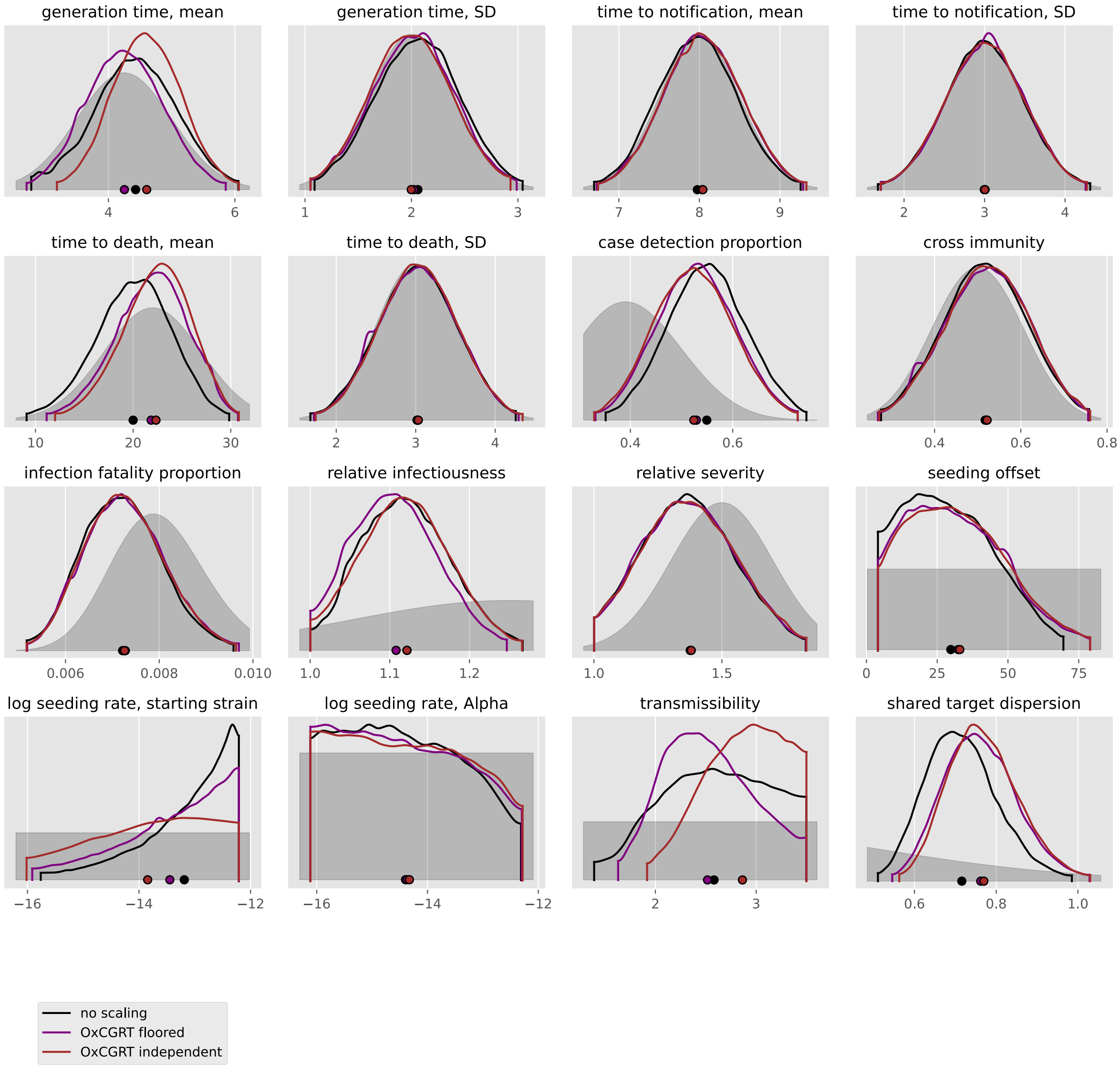

generation time, mean
4
6
generation time, SD
1
2
3
time to notification, mean
7
8
9
time to notification, SD
2
3
4
time to death, mean
10
20
30
time to death, SD
2
3
4
case detection proportion
0.4
0.6
cross immunity
0.4
0.6
0.8
infection fatality proportion
0.006
0.008
0.010
relative infectiousness
1.0
1.1
1.2
relative severity
1.0
1.5
seeding offset
0
25
50
75
log seeding rate, starting strain
−16
−14
−12
log seeding rate, Alpha
−16
−14
−12
transmissibility
2
3
shared target dispersion
0.6
0.8
1.0
no scaling
OxCGRT floored
OxCGRT independent

#### Austria

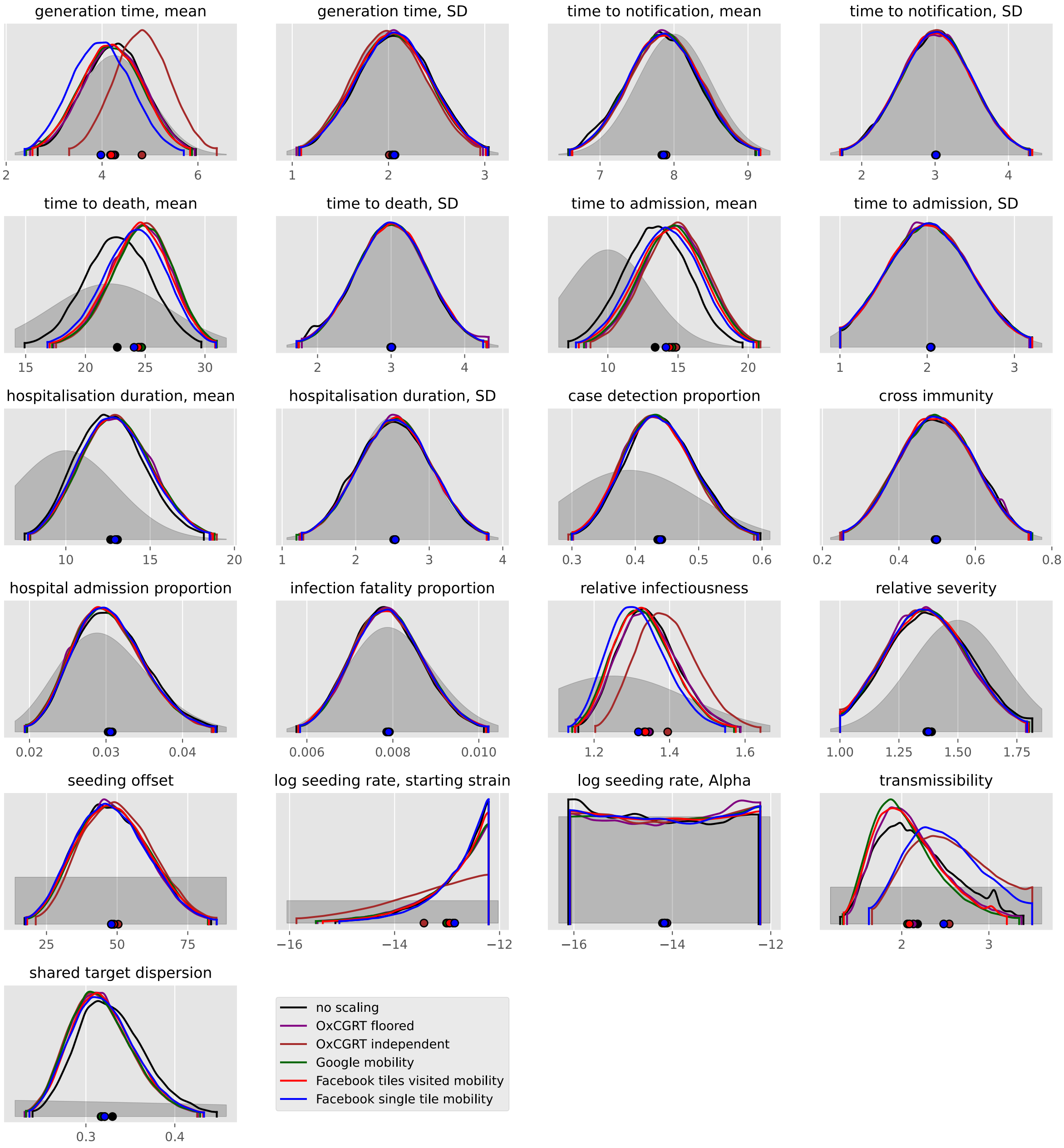
generation time, mean
generation time, SD
time to notification, mean
time to notification, SD
time to death, mean
time to death, SD
time to admission, mean
time to admission, SD
hospitalisation duration, mean
hospitalisation duration, SD
case detection proportion
cross immunity
hospital admission proportion
infection fatality proportion
relative infectiousness
relative severity
seeding offset
log seeding rate, starting strain
log seeding rate, Alpha
transmissibility
shared target dispersion
no scaling
OxCGRT floored
OxCGRT independent
Google mobility
Facebook tiles visited mobility
Facebook single tile mobility

#### Belarus

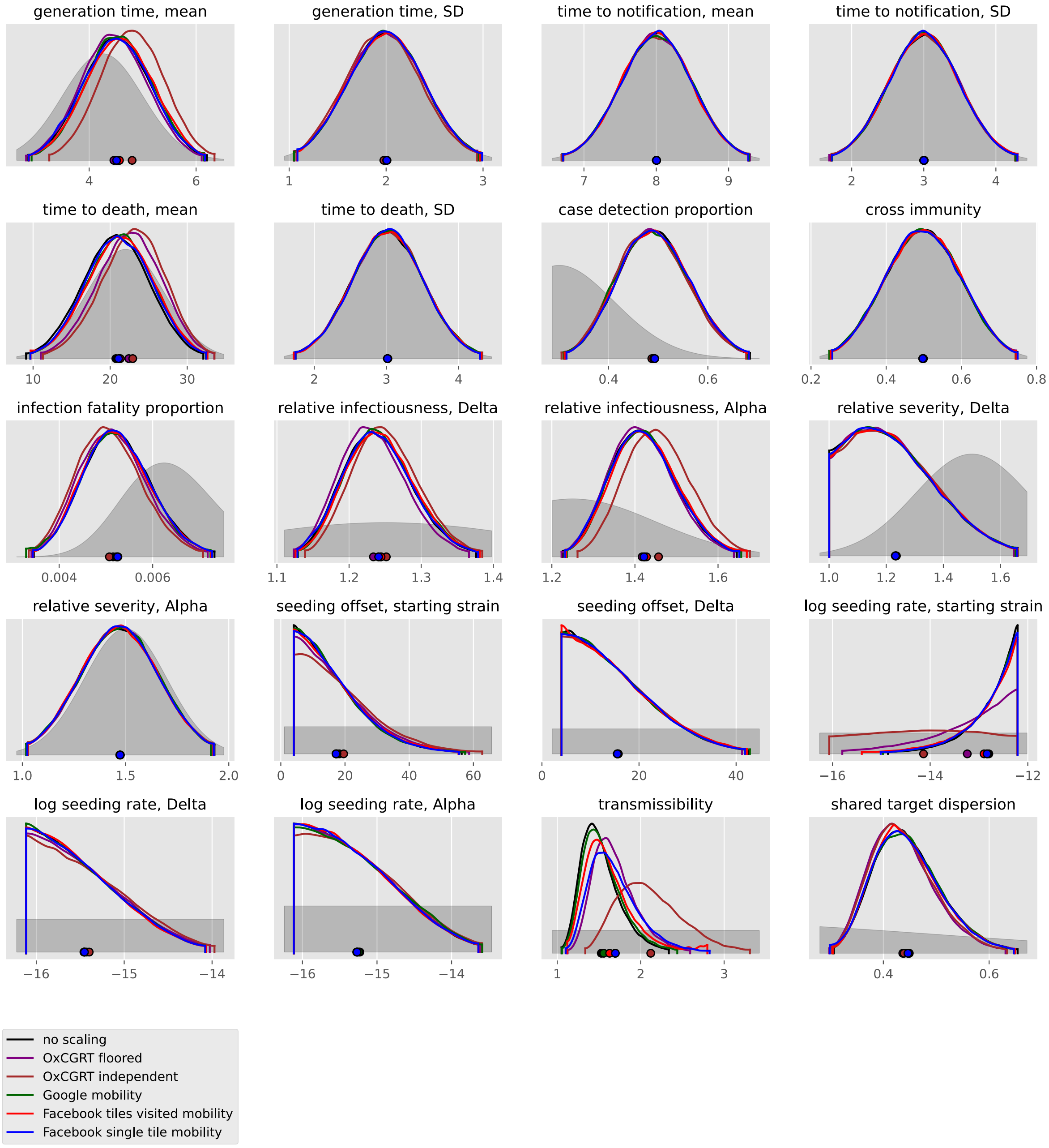

generation time, mean
generation time, SD
time to notification, mean
time to notification, SD
time to death, mean
time to death, SD
case detection proportion
cross immunity
infection fatality proportion
relative infectiousness, Delta
relative infectiousness, Alpha
relative severity, Delta
relative severity, Alpha
seeding offset, starting strain
seeding offset, Delta
log seeding rate, starting strain
log seeding rate, Delta
log seeding rate, Alpha
transmissibility
shared target dispersion
no scaling
OxCGRT floored
OxCGRT independent
Google mobility
Facebook tiles visited mobility
Facebook single tile mobility

#### Belgium

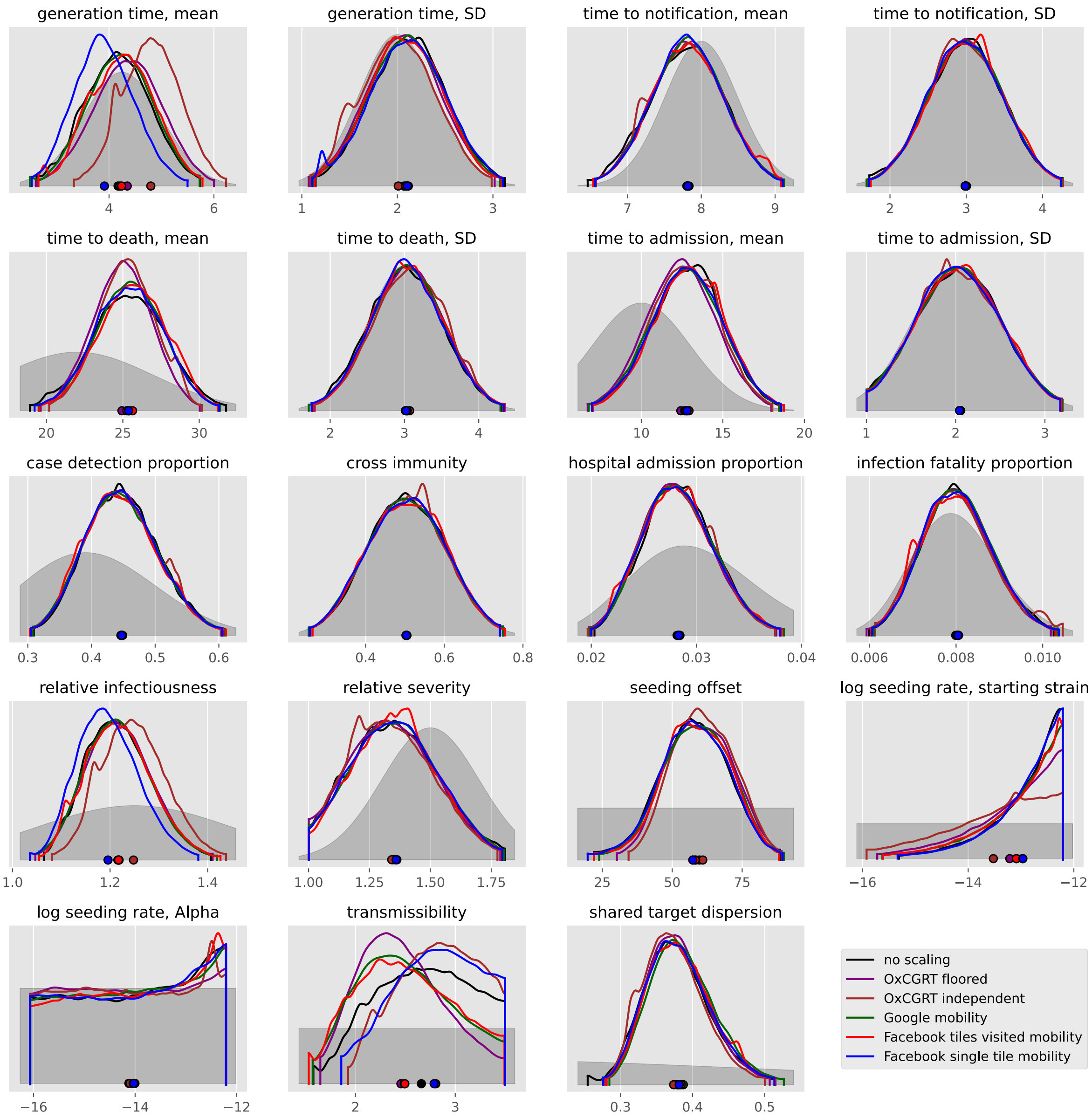

generation time, mean
generation time, SD
time to notification, mean
time to notification, SD
time to death, mean
time to death, SD
time to admission, mean
time to admission, SD
case detection proportion
cross immunity
hospital admission proportion
infection fatality proportion
relative infectiousness
relative severity
seeding offset
log seeding rate, starting strain
log seeding rate, Alpha
transmissibility
shared target dispersion
no scaling
OxCGRT floored
OxCGRT independent
Google mobility
Facebook tiles visited mobility
Facebook single tile mobility

#### Bosnia and Herzegovina

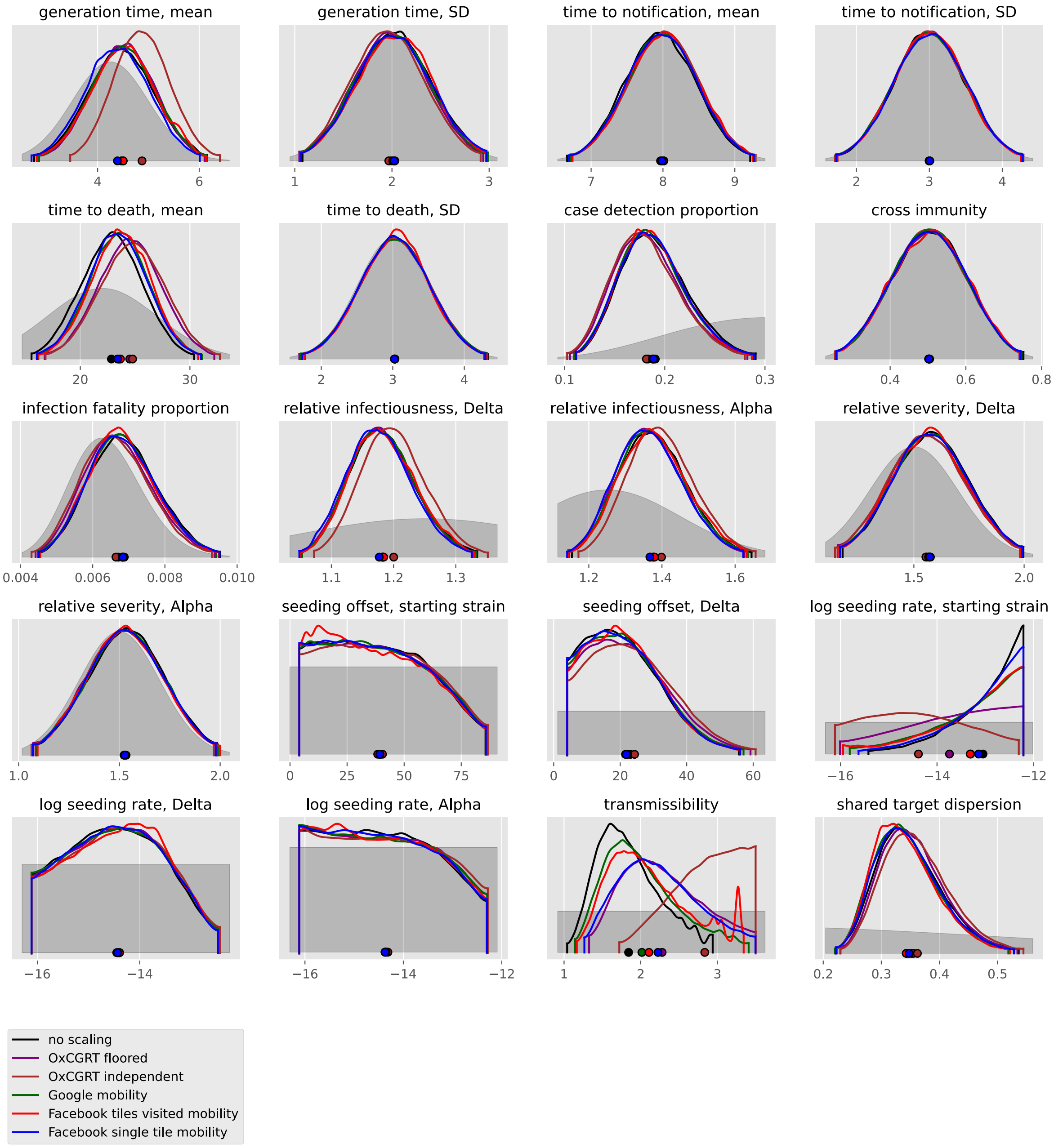

#### Bulgaria

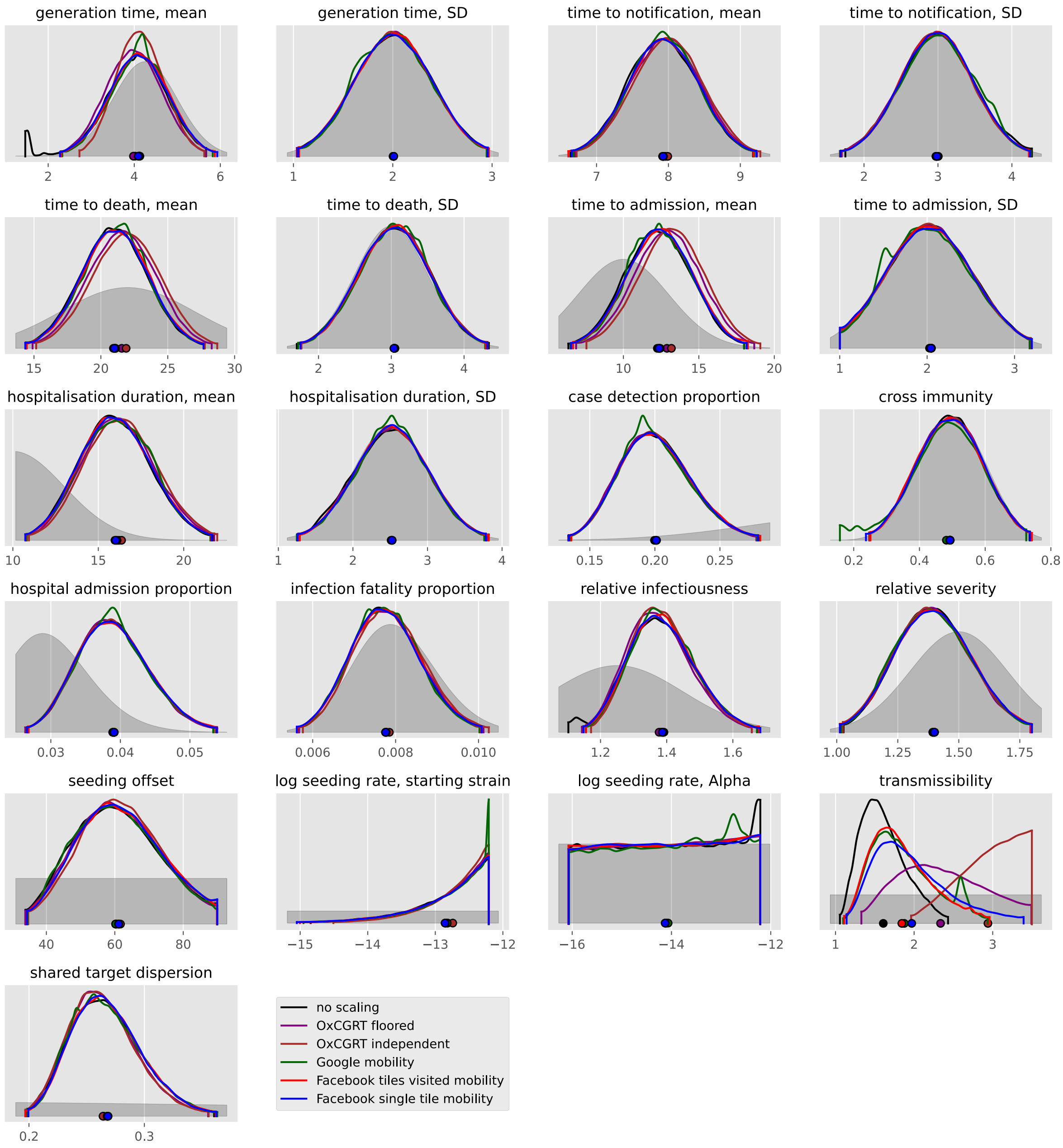

generation time, mean
2 4 6
generation time, SD
1 2 3
time to notification, mean
7 8 9
time to notification, SD
2 3 4
time to death, mean
15 20 25 30
time to death, SD
2 3 4
time to admission, mean
10 15 20
time to admission, SD
1 2 3
hospitalisation duration, mean
10 15 20
hospitalisation duration, SD
1 2 3 4
case detection proportion
0.15 0.20 0.25
cross immunity
0.2 0.4 0.6 0.8
hospital admission proportion
0.03 0.04 0.05
infection fatality proportion
0.006 0.008 0.010
relative infectiousness
1.2 1.4 1.6
relative severity
1.00 1.25 1.50 1.75
seeding offset
40 60 80
log seeding rate, starting strain
−15 −14 −13 −12
log seeding rate, Alpha
−16 −14 −12
transmissibility
1 2 3
shared target dispersion
0.2 0.3
no scaling
OxCGRT floored
OxCGRT independent
Google mobility
Facebook tiles visited mobility
Facebook single tile mobility

#### Croatia

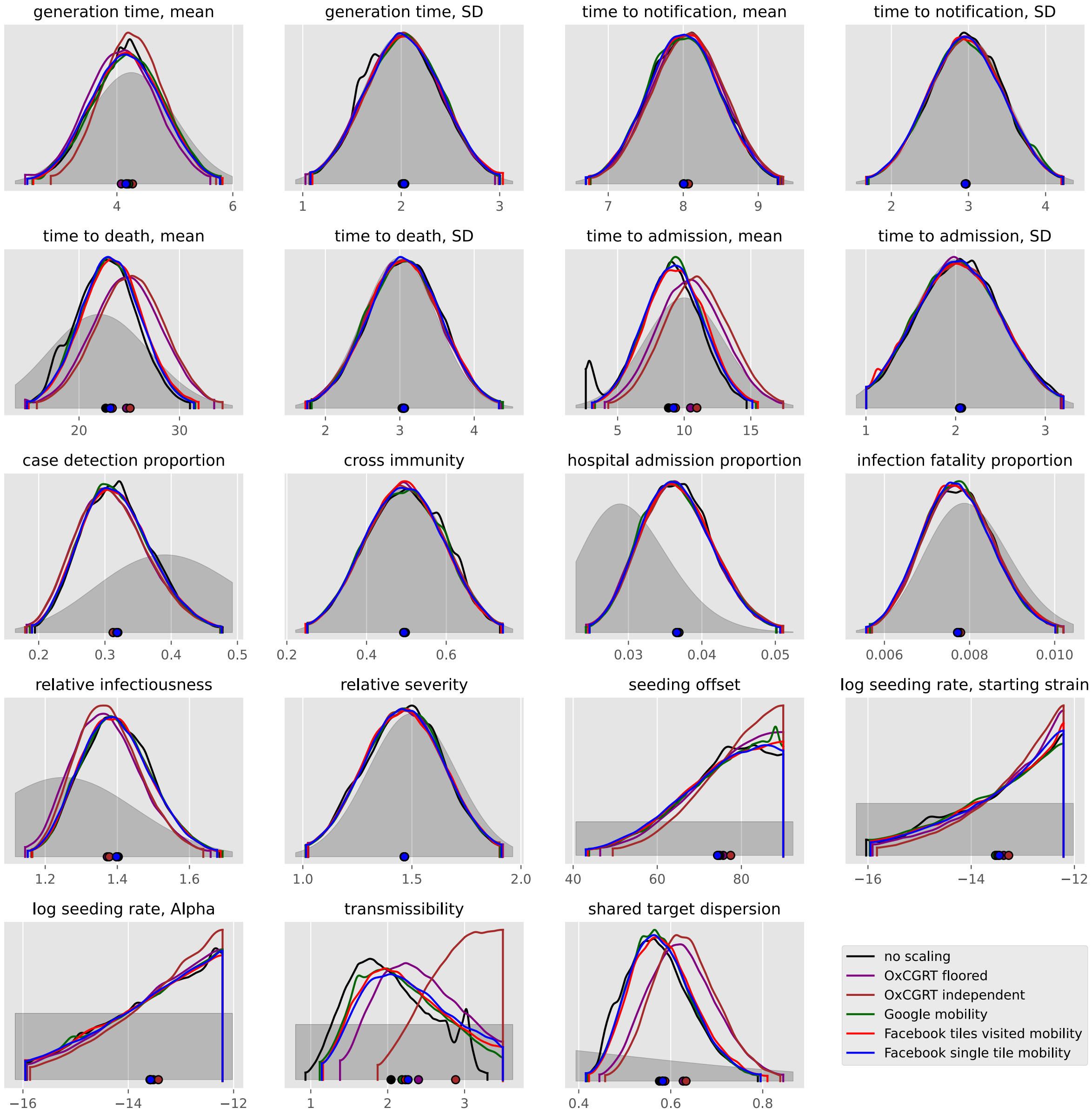
generation time, mean
4
6
generation time, SD
1
2
3
time to notification, mean
7
8
9
time to notification, SD
2
3
4
time to death, mean
20
30
time to death, SD
2
3
4
time to admission, mean
5
10
15
time to admission, SD
1
2
3
case detection proportion
0.2
0.3
0.4
0.5
cross immunity
0.2
0.4
0.6
hospital admission proportion
0.03
0.04
0.05
infection fatality proportion
0.006
0.008
0.010
relative infectiousness
1.2
1.4
1.6
relative severity
1.0
1.5
2.0
seeding offset
40
60
80
log seeding rate, starting strain
−16
−14
−12
log seeding rate, Alpha
−16
−14
−12
transmissibility
1
2
3
shared target dispersion
0.4
0.6
0.8
no scaling
OxCGRT floored
OxCGRT independent
Google mobility
Facebook tiles visited mobility
Facebook single tile mobility

#### Czechia

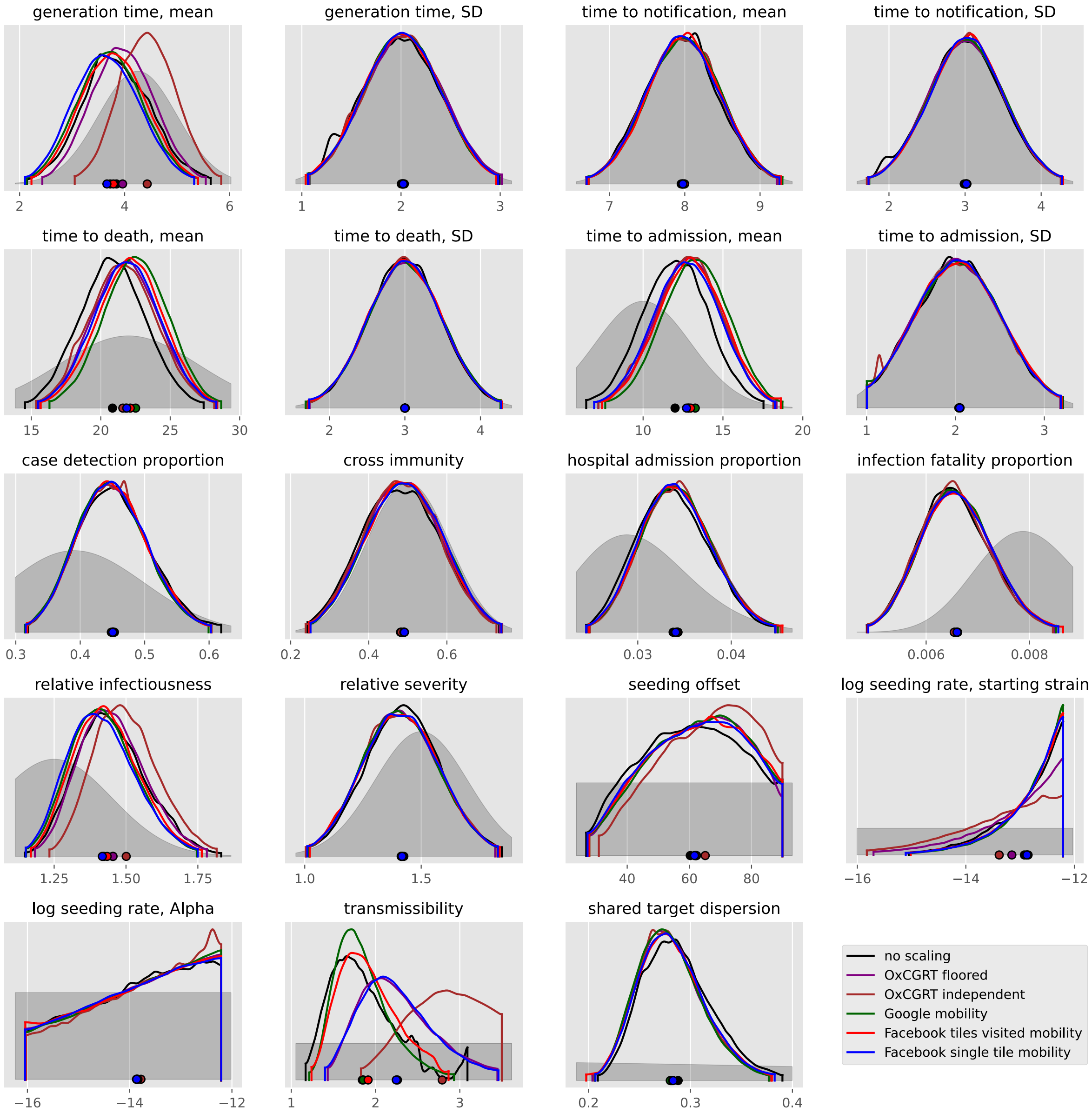
generation time, mean
generation time, SD
time to notification, mean
time to notification, SD
time to death, mean
time to death, SD
time to admission, mean
time to admission, SD
case detection proportion
cross immunity
hospital admission proportion
infection fatality proportion
relative infectiousness
relative severity
seeding offset
log seeding rate, starting strain
log seeding rate, Alpha
transmissibility
shared target dispersion
no scaling
OxCGRT floored
OxCGRT independent
Google mobility
Facebook tiles visited mobility
Facebook single tile mobility

#### Denmark

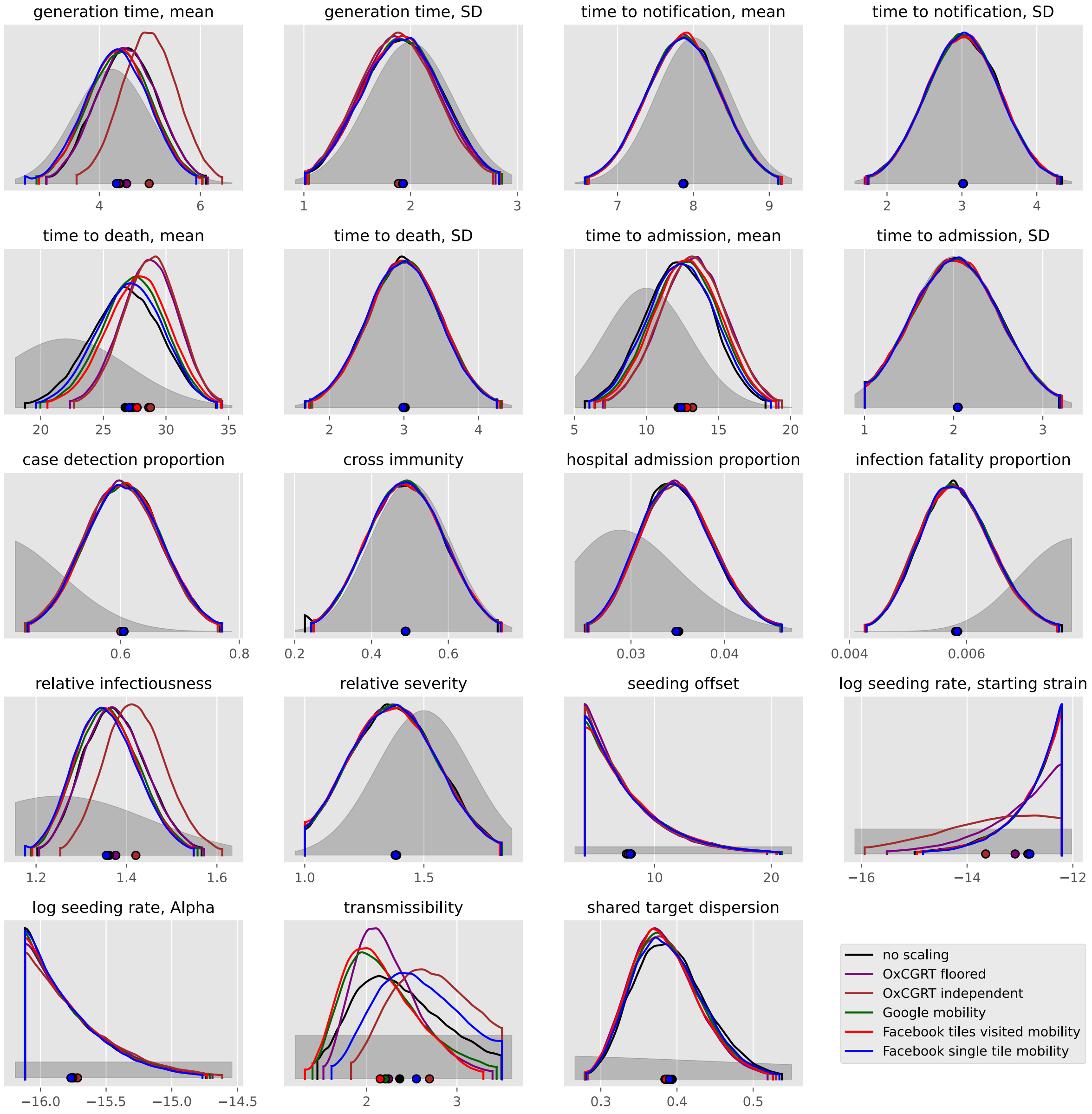
generation time, mean
generation time, SD
time to notification, mean
time to notification, SD
time to death, mean
time to death, SD
time to admission, mean
time to admission, SD
case detection proportion
cross immunity
hospital admission proportion
infection fatality proportion
relative infectiousness
relative severity
seeding offset
log seeding rate, starting strain
log seeding rate, Alpha
transmissibility
shared target dispersion
no scaling
OxCGRT floored
OxCGRT independent
Google mobility
Facebook tiles visited mobility
Facebook single tile mobility

#### Estonia

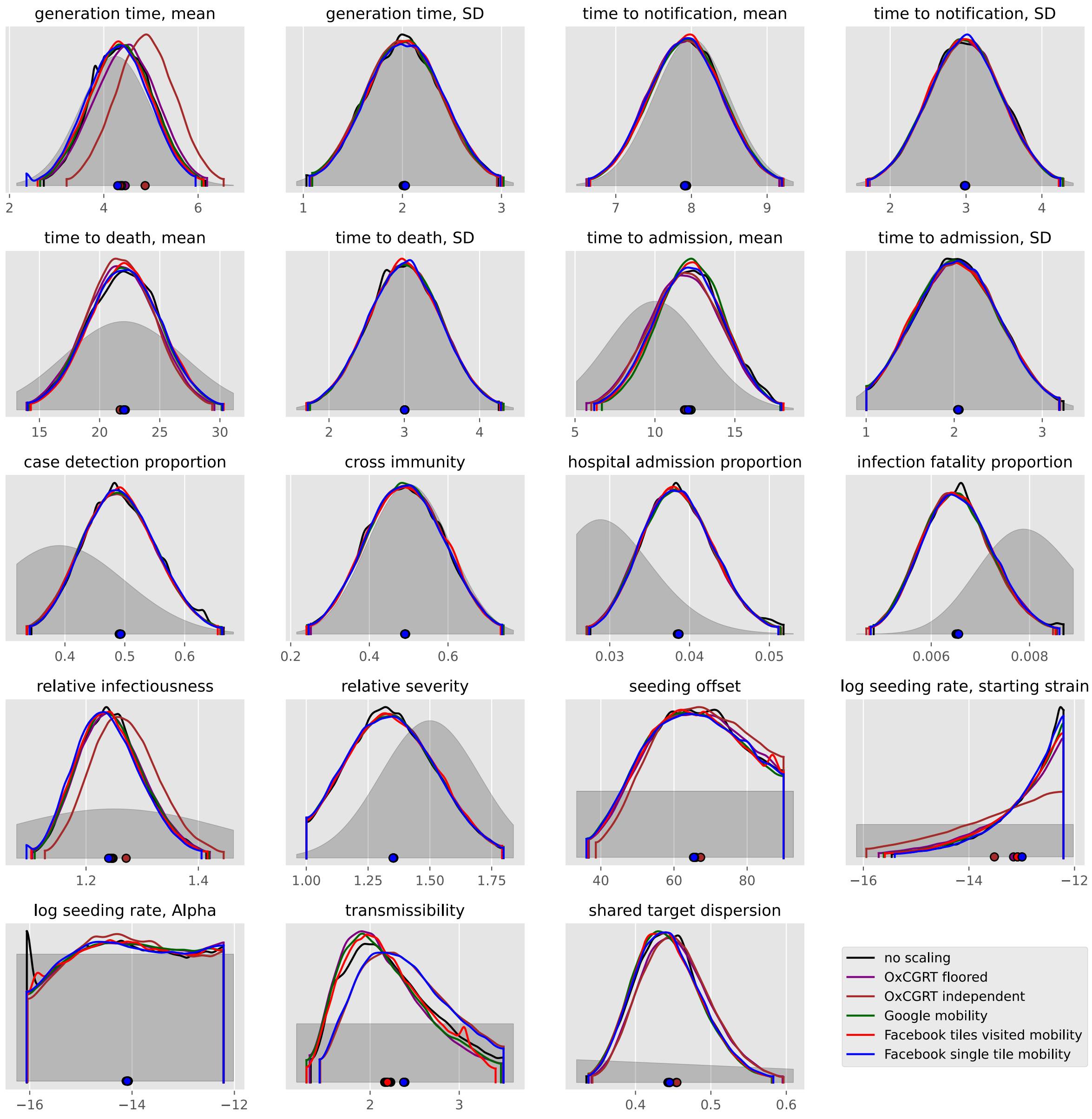
generation time, mean
generation time, SD
time to notification, mean
time to notification, SD
time to death, mean
time to death, SD
time to admission, mean
time to admission, SD
case detection proportion
cross immunity
hospital admission proportion
infection fatality proportion
relative infectiousness
relative severity
seeding offset
log seeding rate, starting strain
log seeding rate, Alpha
transmissibility
shared target dispersion
no scaling
OxCGRT floored
OxCGRT independent
Google mobility
Facebook tiles visited mobility
Facebook single tile mobility

#### Finland

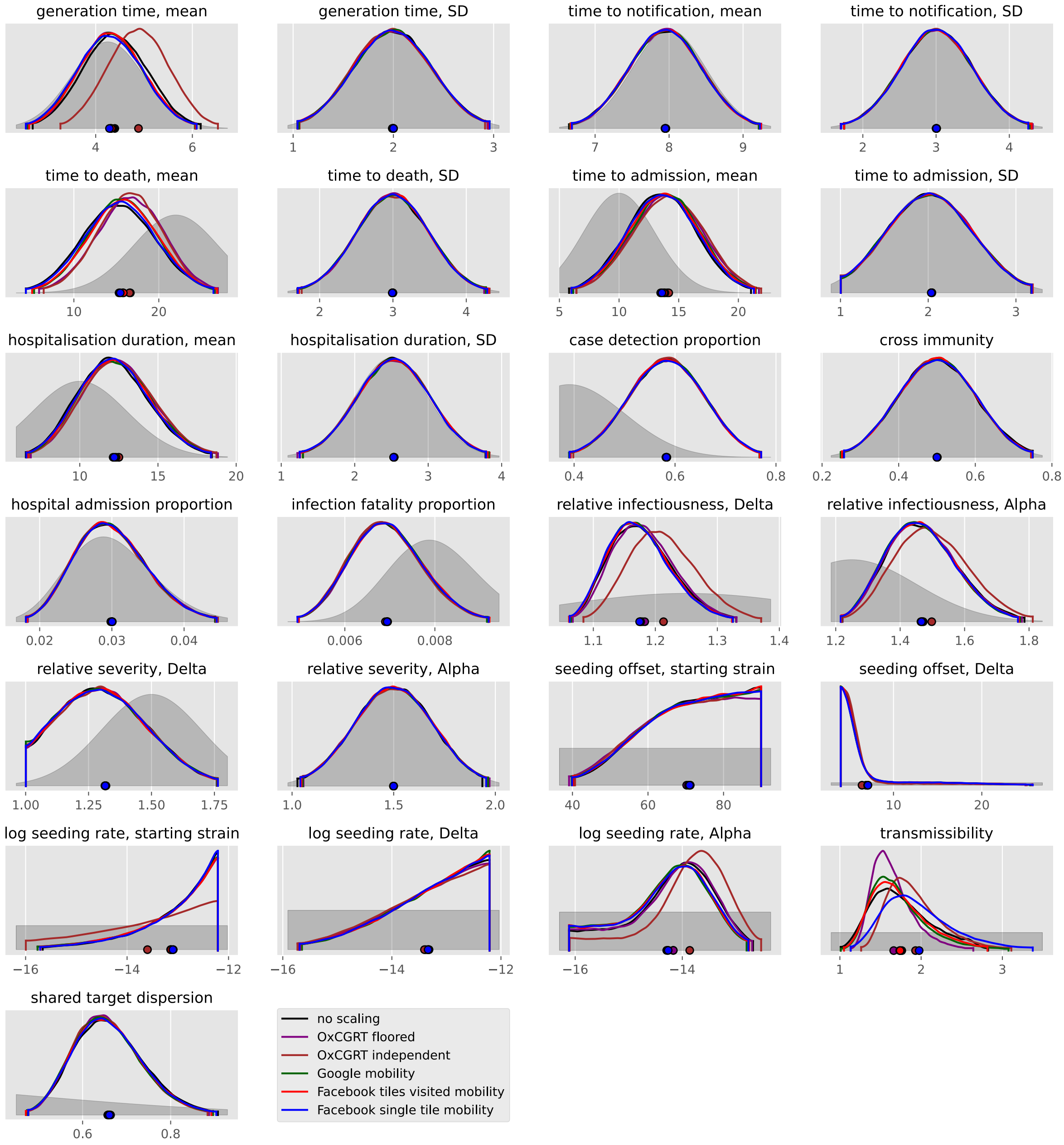
generation time, mean
generation time, SD
time to notification, mean
time to notification, SD
time to death, mean
time to death, SD
time to admission, mean
time to admission, SD
hospitalisation duration, mean
hospitalisation duration, SD
case detection proportion
cross immunity
hospital admission proportion
infection fatality proportion
relative infectiousness, Delta
relative infectiousness, Alpha
relative severity, Delta
relative severity, Alpha
seeding offset, starting strain
seeding offset, Delta
log seeding rate, starting strain
log seeding rate, Delta
log seeding rate, Alpha
transmissibility
shared target dispersion
no scaling
OxCGRT floored
OxCGRT independent
Google mobility
Facebook tiles visited mobility
Facebook single tile mobility

#### France

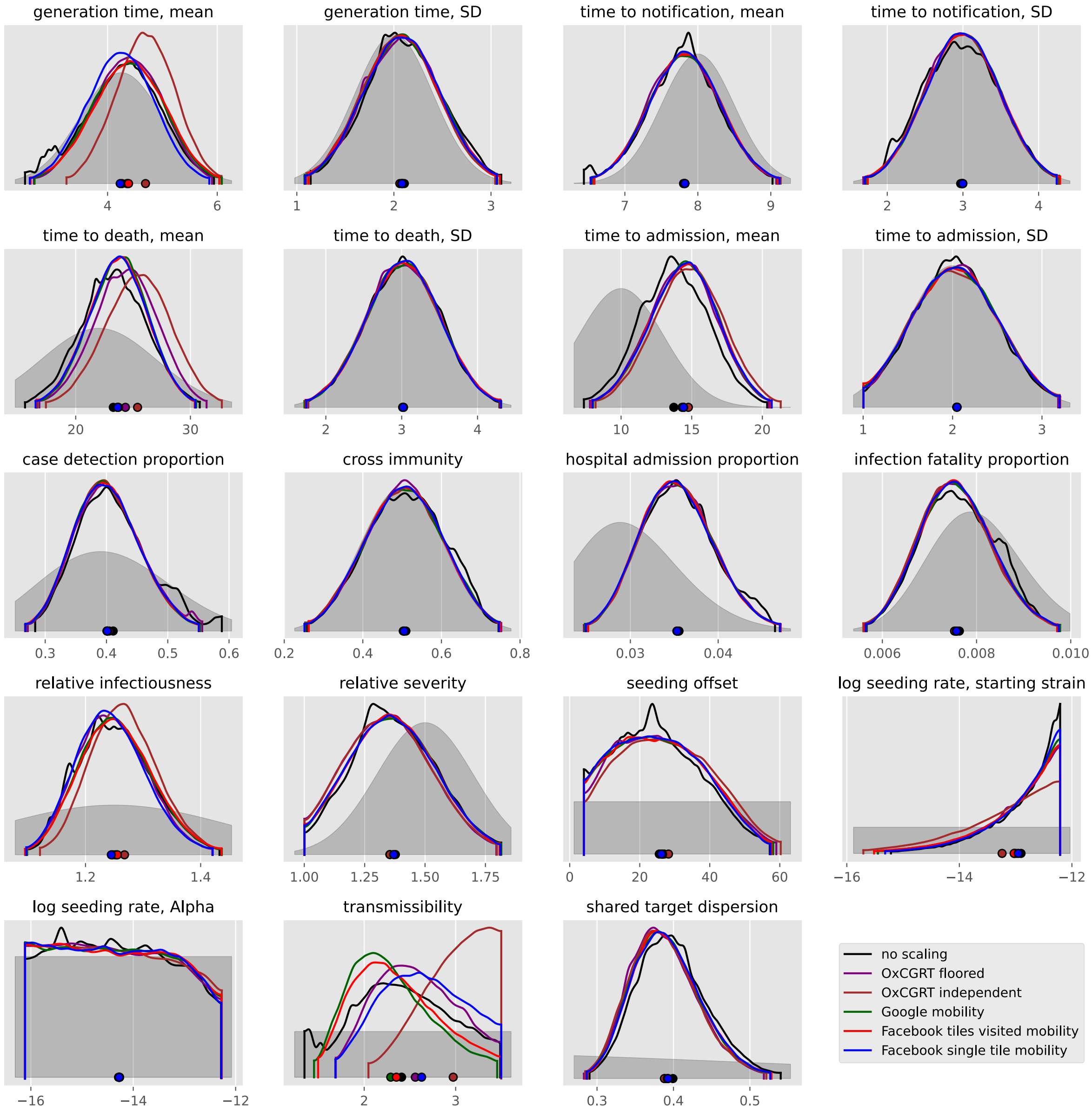
generation time, mean
4
6
generation time, SD
1
2
3
time to notification, mean
7
8
9
time to notification, SD
2
3
4
time to death, mean
20
30
time to death, SD
2
3
4
time to admission, mean
10
15
20
time to admission, SD
1
2
3
case detection proportion
0.3
0.4
0.5
0.6
cross immunity
0.2
0.4
0.6
0.8
hospital admission proportion
0.03
0.04
infection fatality proportion
0.006
0.008
0.010
relative infectiousness
1.2
1.4
relative severity
1.00
1.25
1.50
1.75
seeding offset
0
20
40
60
log seeding rate, starting strain
−16
−14
−12
log seeding rate, Alpha
−16
−14
−12
transmissibility
2
3
shared target dispersion
0.3
0.4
0.5
no scaling
OxCGRT floored
OxCGRT independent
Google mobility
Facebook tiles visited mobility
Facebook single tile mobility

#### Germany

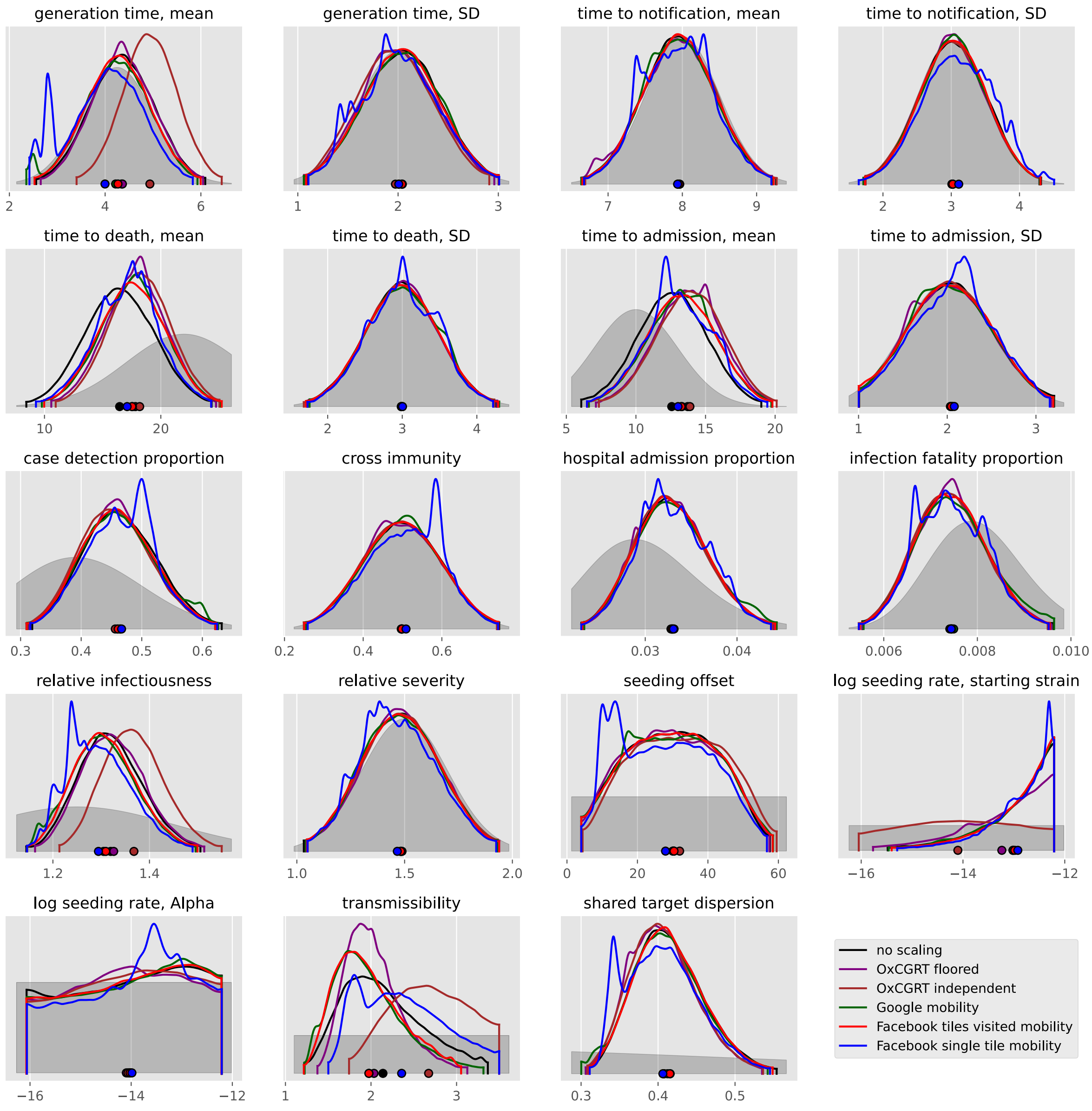

generation time, mean
generation time, SD
time to notification, mean
time to notification, SD
time to death, mean
time to death, SD
time to admission, mean
time to admission, SD
case detection proportion
cross immunity
hospital admission proportion
infection fatality proportion
relative infectiousness
relative severity
seeding offset
log seeding rate, starting strain
log seeding rate, Alpha
transmissibility
shared target dispersion
no scaling
OxCGRT floored
OxCGRT independent
Google mobility
Facebook tiles visited mobility
Facebook single tile mobility

#### Greece

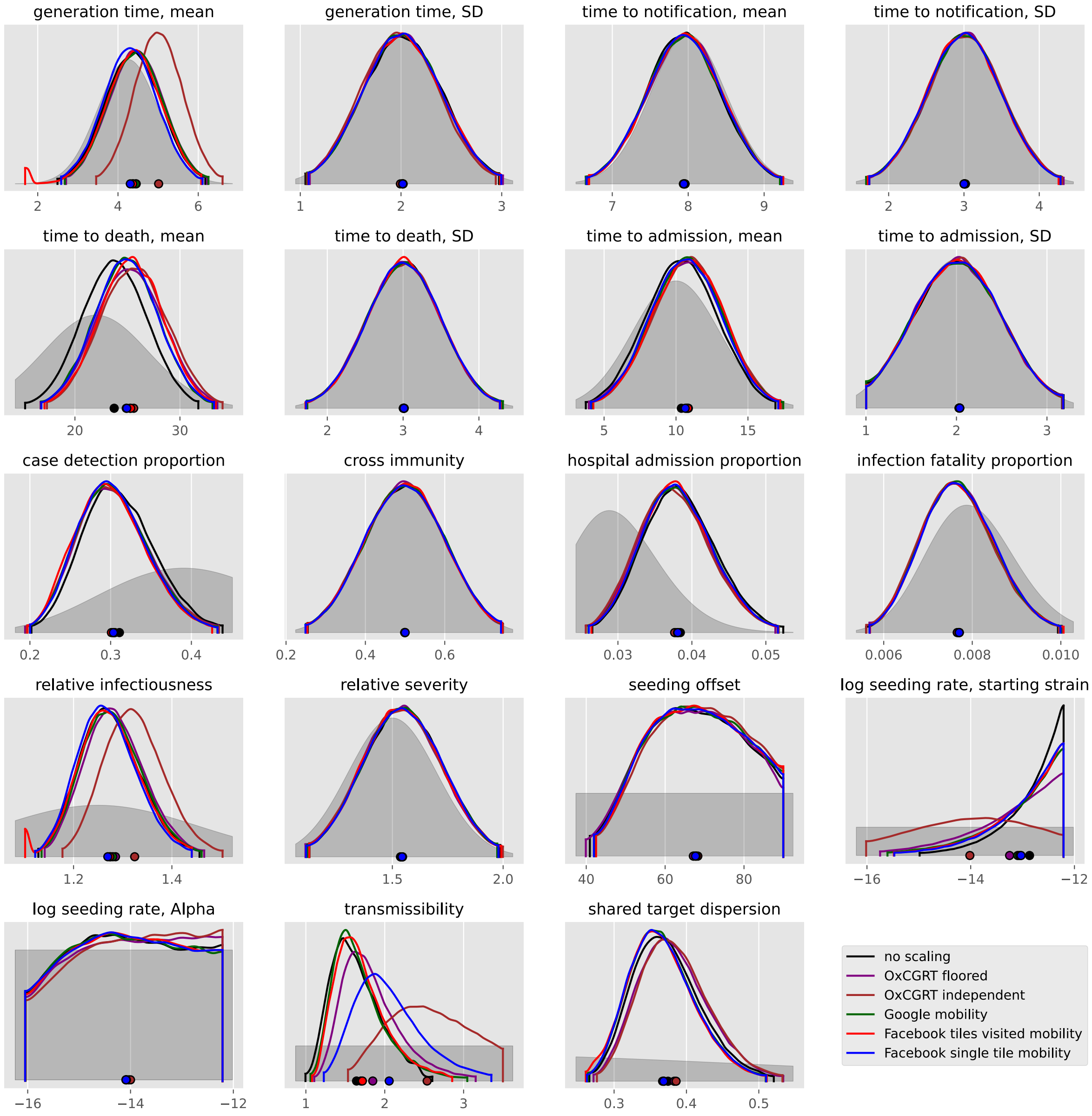

generation time, mean
generation time, SD
time to notification, mean
time to notification, SD
time to death, mean
time to death, SD
time to admission, mean
time to admission, SD
case detection proportion
cross immunity
hospital admission proportion
infection fatality proportion
relative infectiousness
relative severity
seeding offset
log seeding rate, starting strain
log seeding rate, Alpha
transmissibility
shared target dispersion
no scaling
OxCGRT floored
OxCGRT independent
Google mobility
Facebook tiles visited mobility
Facebook single tile mobility

#### Hungary

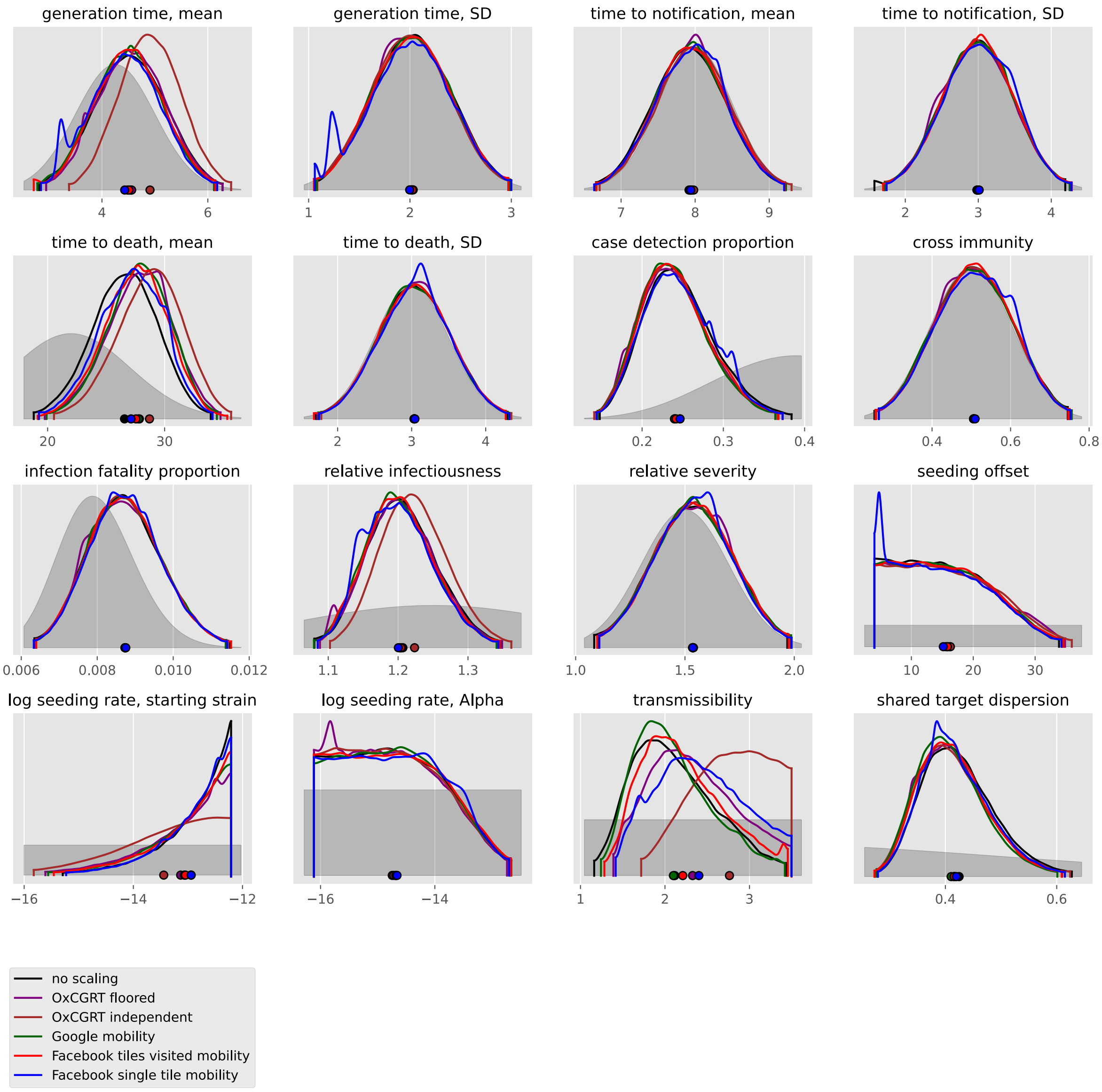
generation time, mean
generation time, SD
time to notification, mean
time to notification, SD
time to death, mean
time to death, SD
case detection proportion
cross immunity
infection fatality proportion
relative infectiousness
relative severity
seeding offset
log seeding rate, starting strain
log seeding rate, Alpha
transmissibility
shared target dispersion
no scaling
OxCGRT floored
OxCGRT independent
Google mobility
Facebook tiles visited mobility
Facebook single tile mobility

#### Ireland

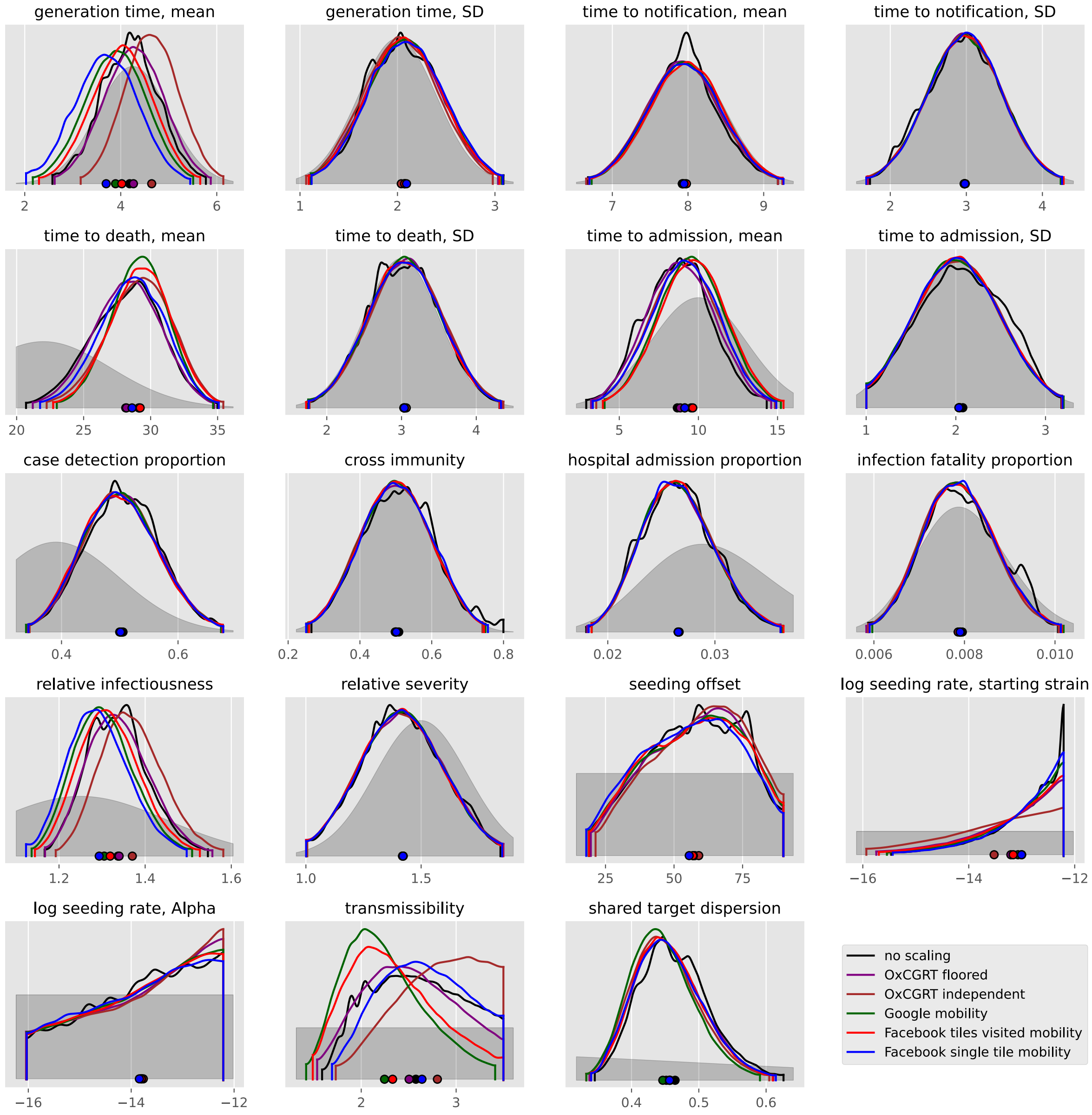
generation time, mean
generation time, SD
time to notification, mean
time to notification, SD
time to death, mean
time to death, SD
time to admission, mean
time to admission, SD
case detection proportion
cross immunity
hospital admission proportion
infection fatality proportion
relative infectiousness
relative severity
seeding offset
log seeding rate, starting strain
log seeding rate, Alpha
transmissibility
shared target dispersion
no scaling
OxCGRT floored
OxCGRT independent
Google mobility
Facebook tiles visited mobility
Facebook single tile mobility

#### Italy

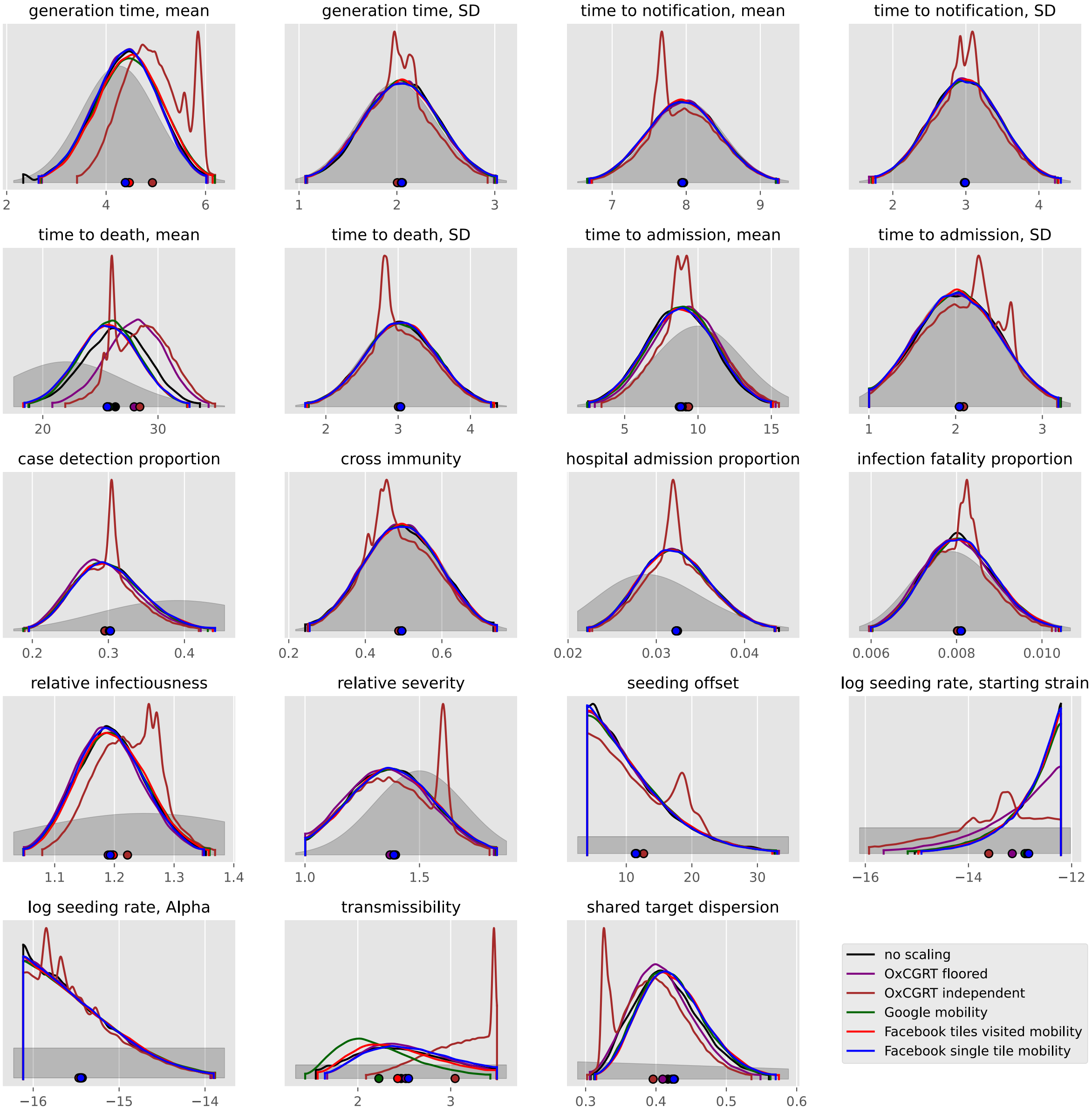

generation time, mean
generation time, SD
time to notification, mean
time to notification, SD
time to death, mean
time to death, SD
time to admission, mean
time to admission, SD
case detection proportion
cross immunity
hospital admission proportion
infection fatality proportion
relative infectiousness
relative severity
seeding offset
log seeding rate, starting strain
log seeding rate, Alpha
transmissibility
shared target dispersion
no scaling
OxCGRT floored
OxCGRT independent
Google mobility
Facebook tiles visited mobility
Facebook single tile mobility

#### Kosovo

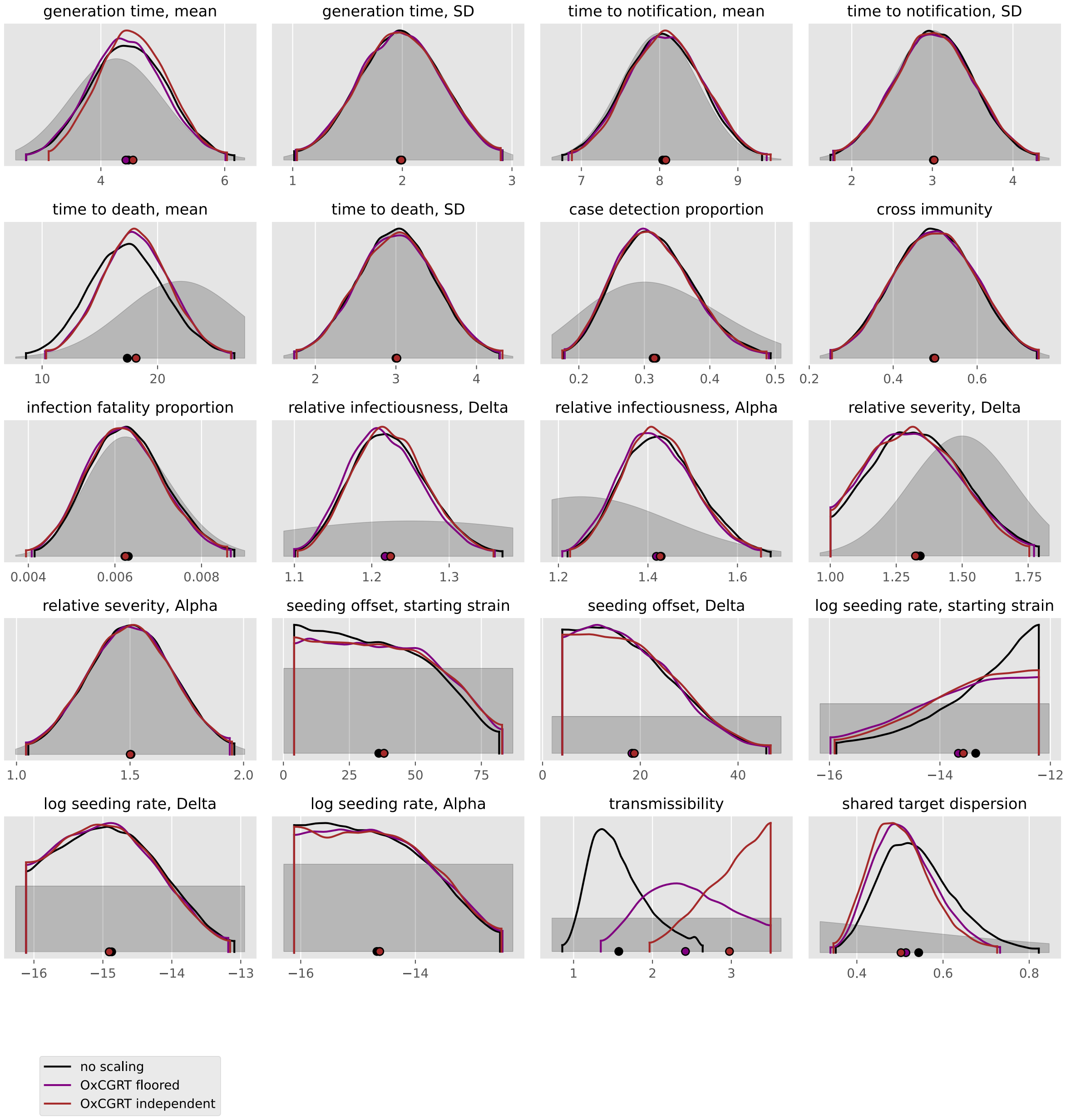

generation time, mean
generation time, SD
time to notification, mean
time to notification, SD
time to death, mean
time to death, SD
case detection proportion
cross immunity
infection fatality proportion
relative infectiousness, Delta
relative infectiousness, Alpha
relative severity, Delta
relative severity, Alpha
seeding offset, starting strain
seeding offset, Delta
log seeding rate, starting strain
log seeding rate, Delta
log seeding rate, Alpha
transmissibility
shared target dispersion
no scaling
OxCGRT floored
OxCGRT independent

#### Latvia

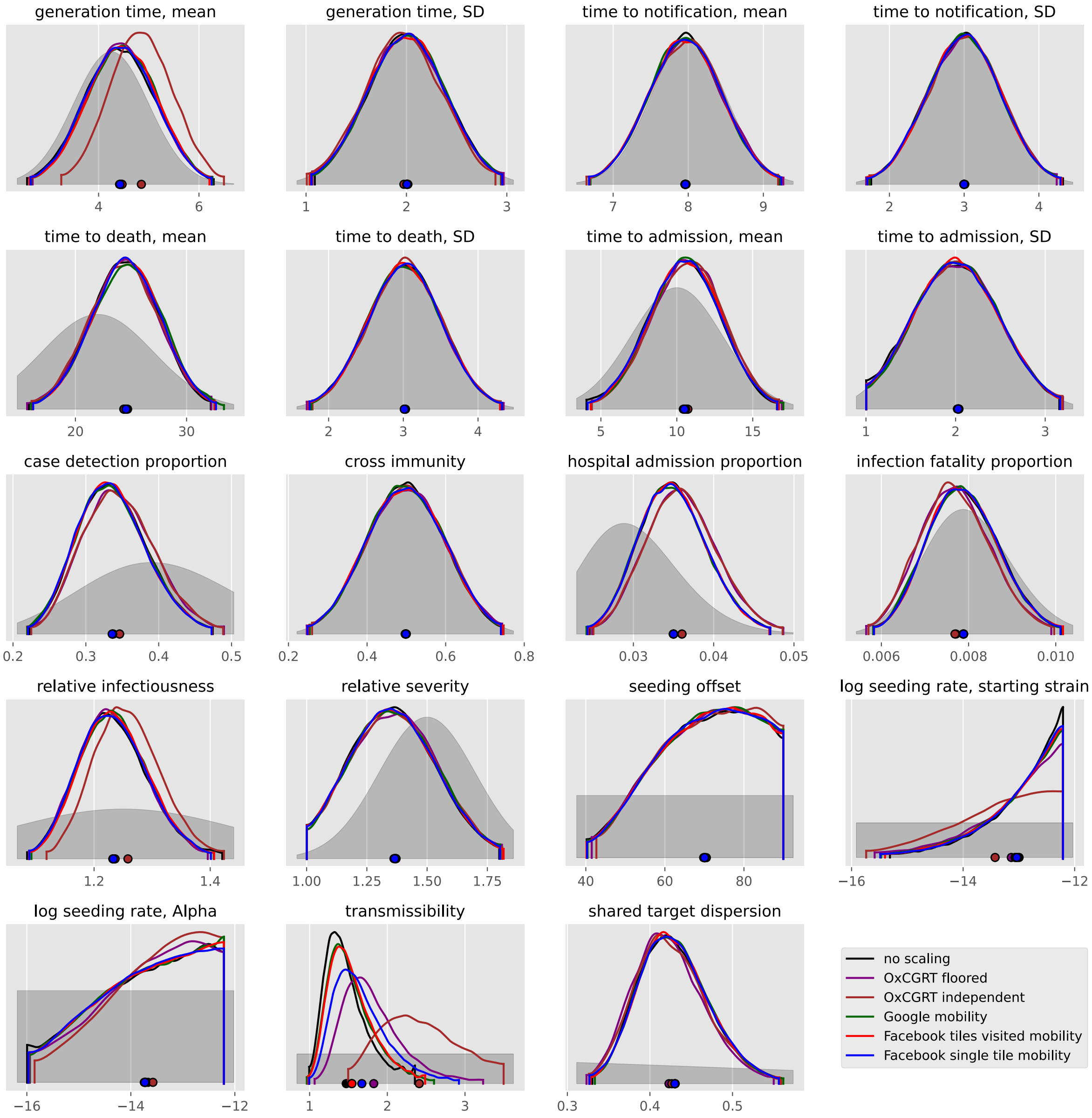

generation time, mean
generation time, SD
time to notification, mean
time to notification, SD
time to death, mean
time to death, SD
time to admission, mean
time to admission, SD
case detection proportion
cross immunity
hospital admission proportion
infection fatality proportion
relative infectiousness
relative severity
seeding offset
log seeding rate, starting strain
log seeding rate, Alpha
transmissibility
shared target dispersion
no scaling
OxCGRT floored
OxCGRT independent
Google mobility
Facebook tiles visited mobility
Facebook single tile mobility

#### Lithuania

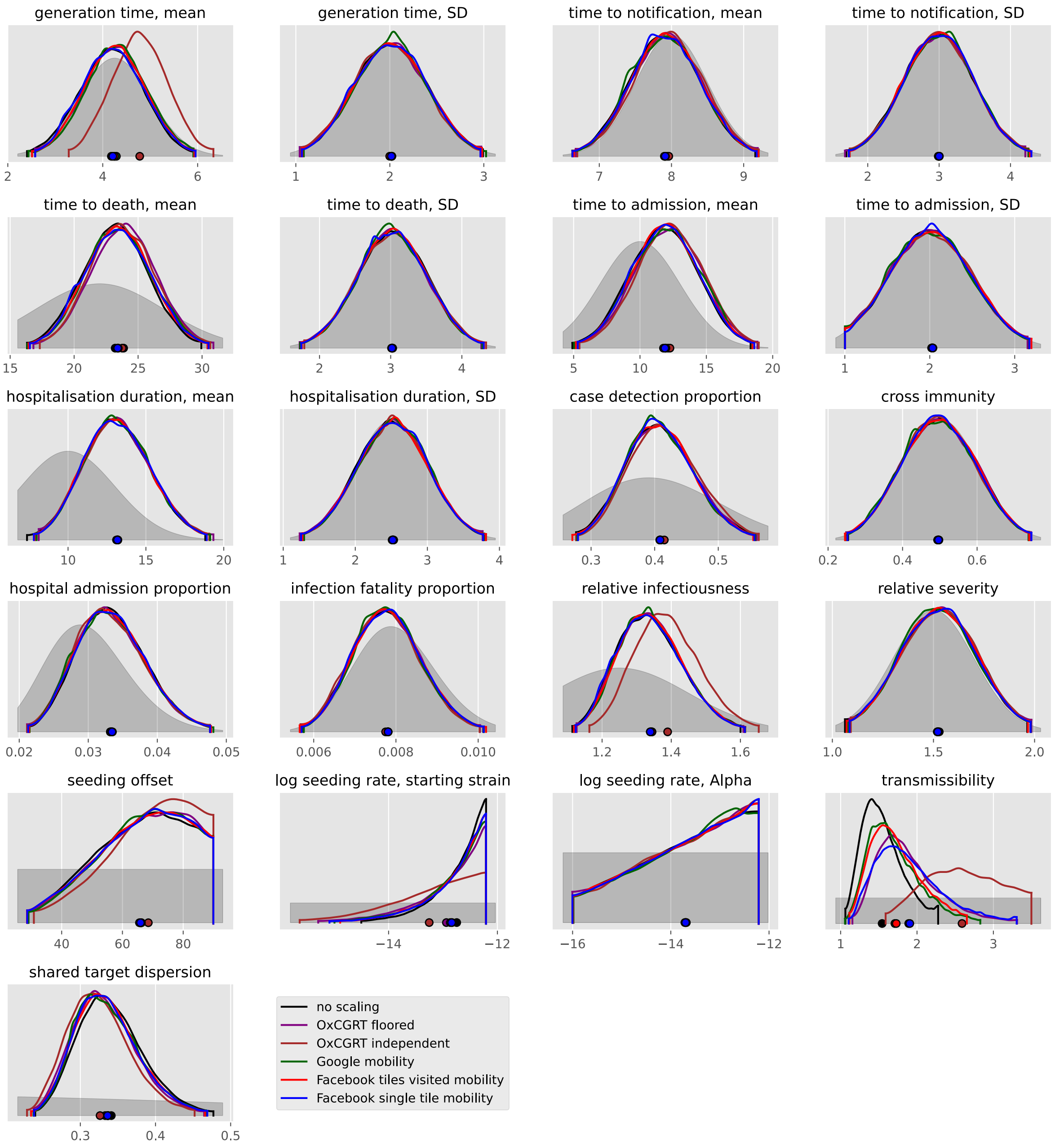
generation time, mean
generation time, SD
time to notification, mean
time to notification, SD
time to death, mean
time to death, SD
time to admission, mean
time to admission, SD
hospitalisation duration, mean
hospitalisation duration, SD
case detection proportion
cross immunity
hospital admission proportion
infection fatality proportion
relative infectiousness
relative severity
seeding offset
log seeding rate, starting strain
log seeding rate, Alpha
transmissibility
shared target dispersion
no scaling
OxCGRT floored
OxCGRT independent
Google mobility
Facebook tiles visited mobility
Facebook single tile mobility

#### Luxembourg

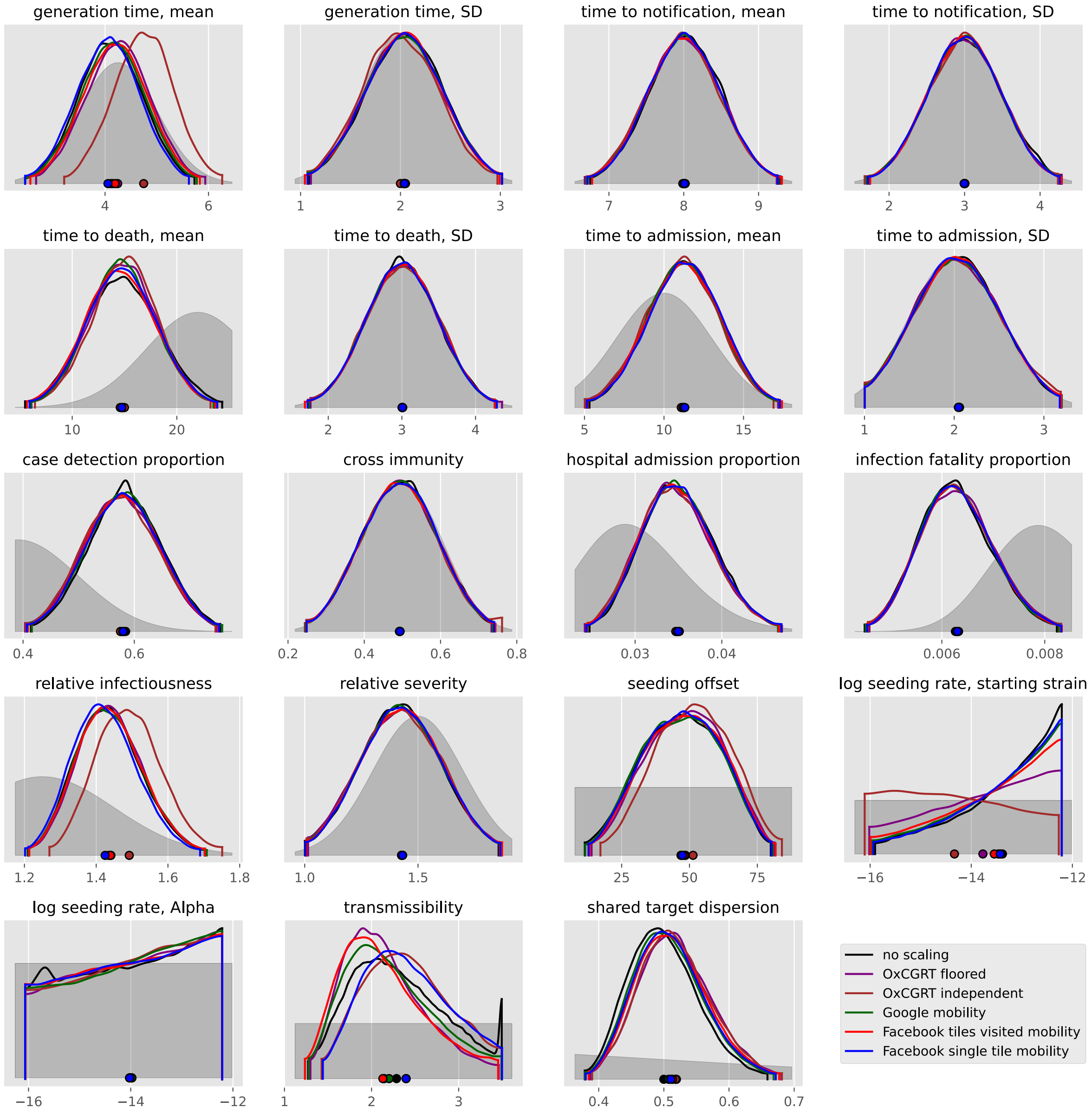
generation time, mean
generation time, SD
time to notification, mean
time to notification, SD
time to death, mean
time to death, SD
time to admission, mean
time to admission, SD
case detection proportion
cross immunity
hospital admission proportion
infection fatality proportion
relative infectiousness
relative severity
seeding offset
log seeding rate, starting strain
log seeding rate, Alpha
transmissibility
shared target dispersion
no scaling
OxCGRT floored
OxCGRT independent
Google mobility
Facebook tiles visited mobility
Facebook single tile mobility

#### Malta

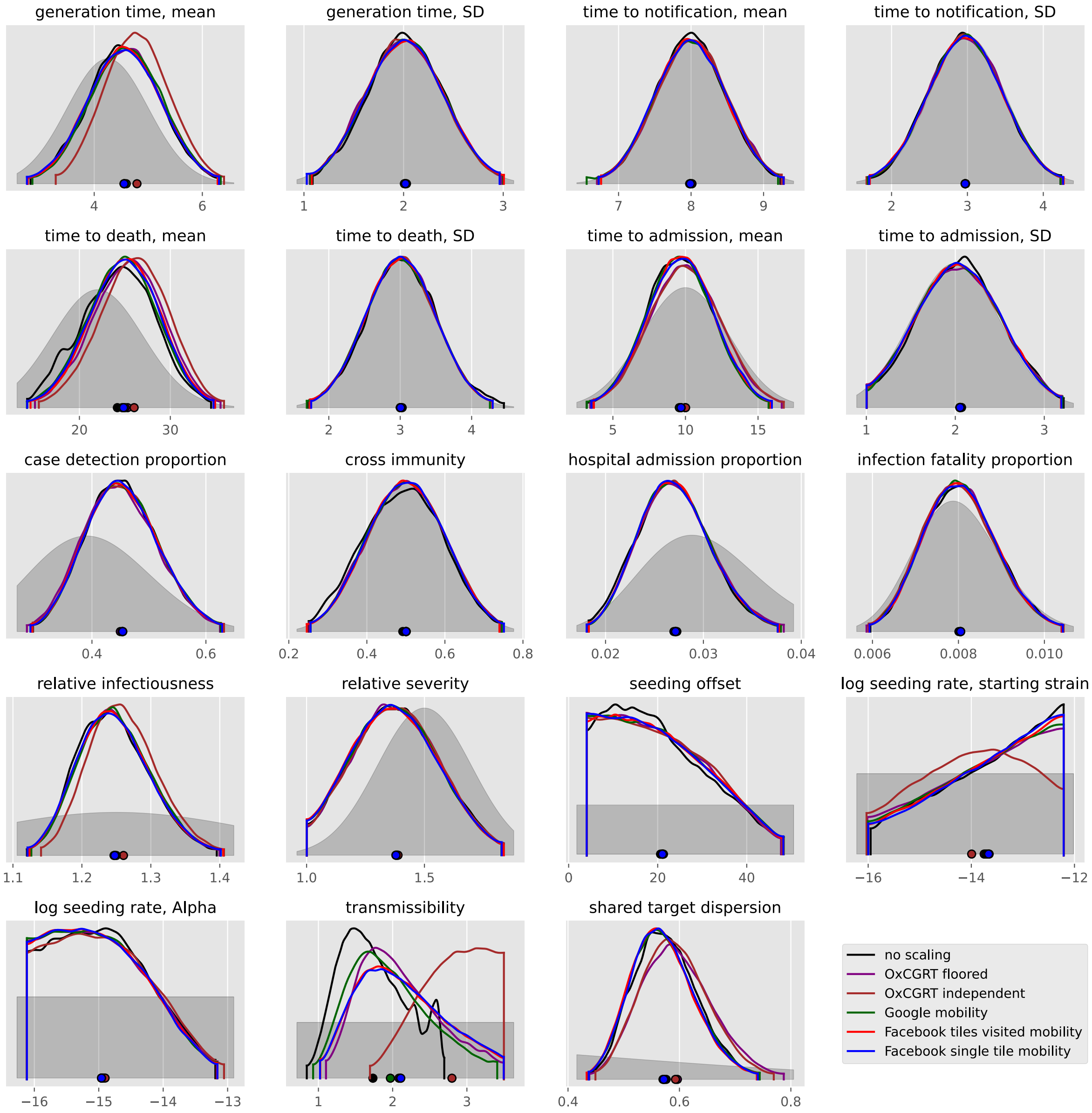
generation time, mean
generation time, SD
time to notification, mean
time to notification, SD
time to death, mean
time to death, SD
time to admission, mean
time to admission, SD
case detection proportion
cross immunity
hospital admission proportion
infection fatality proportion
relative infectiousness
relative severity
seeding offset
log seeding rate, starting strain
log seeding rate, Alpha
transmissibility
shared target dispersion
no scaling
OxCGRT floored
OxCGRT independent
Google mobility
Facebook tiles visited mobility
Facebook single tile mobility

#### Moldova, Republic of

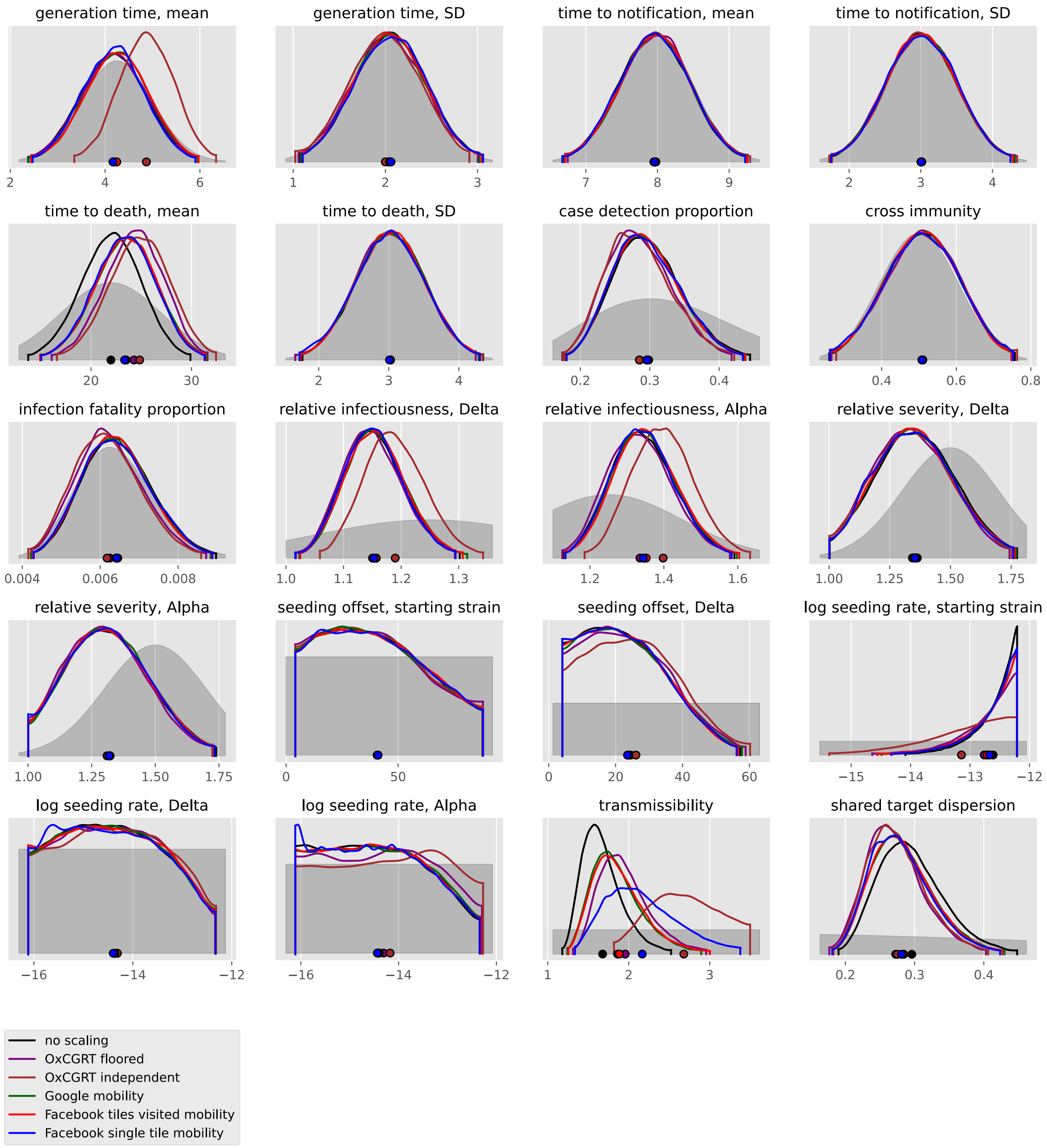

#### Netherlands

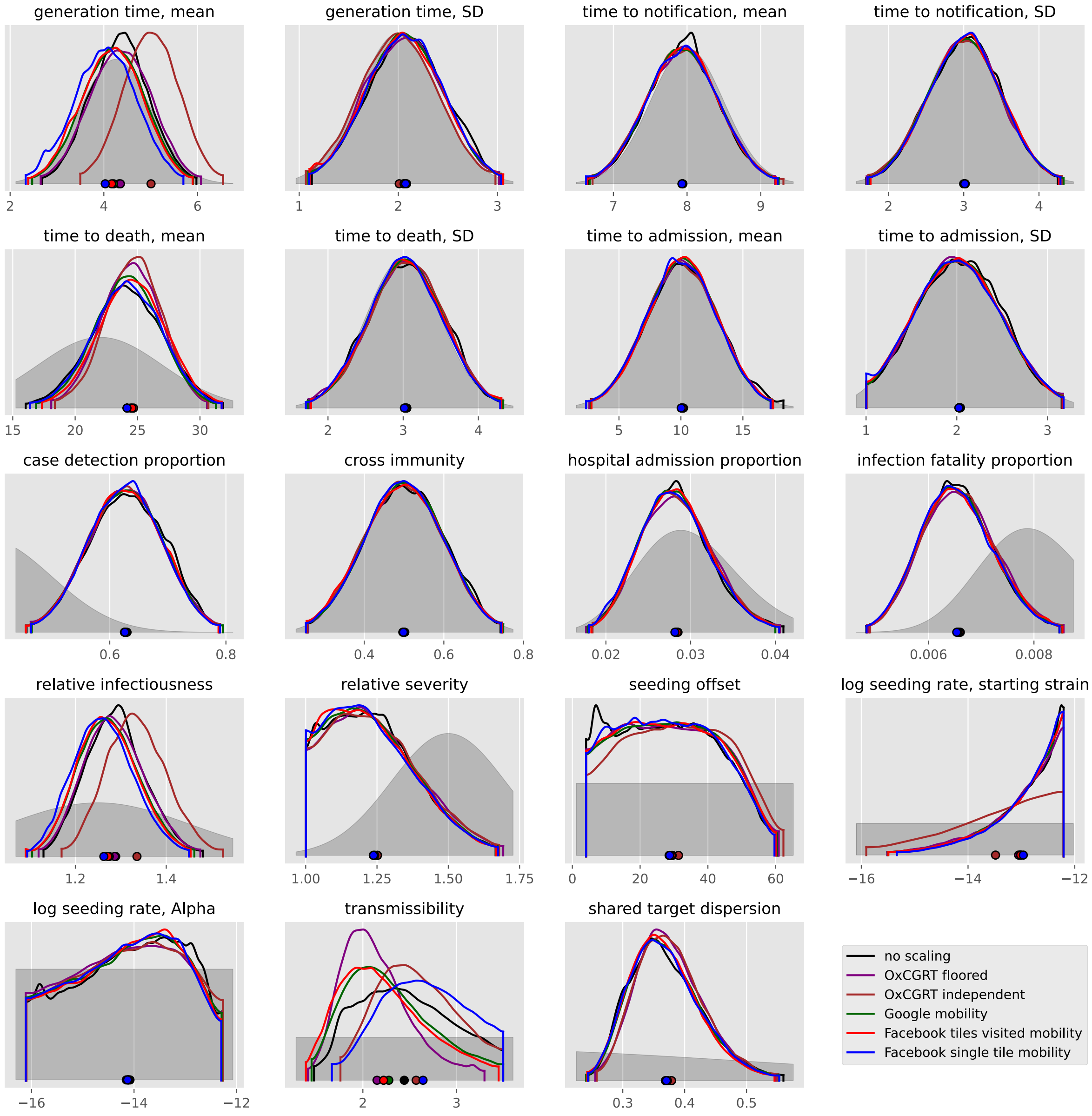

#### Norway

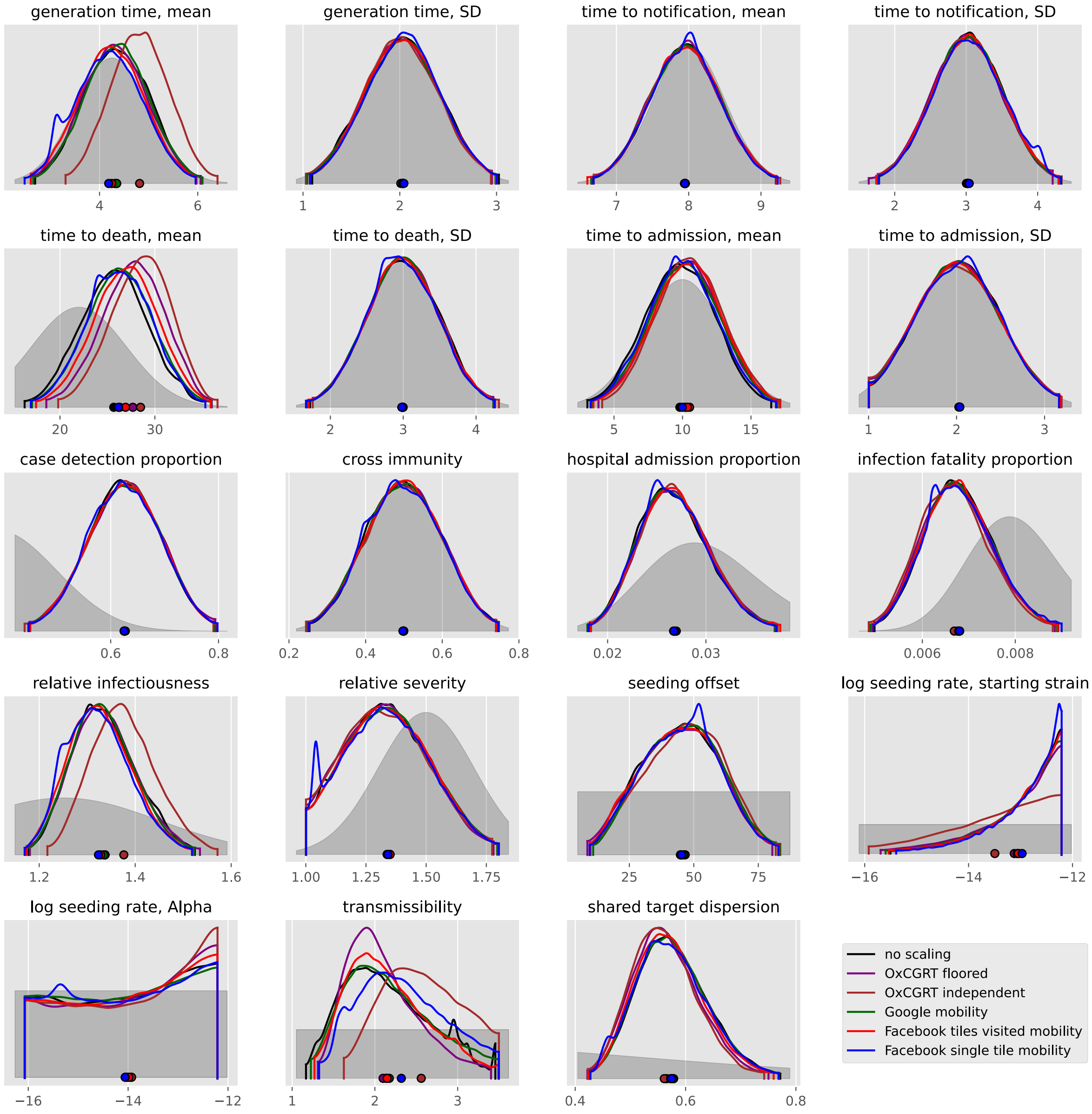
generation time, mean
generation time, SD
time to notification, mean
time to notification, SD
time to death, mean
time to death, SD
time to admission, mean
time to admission, SD
case detection proportion
cross immunity
hospital admission proportion
infection fatality proportion
relative infectiousness
relative severity
seeding offset
log seeding rate, starting strain
log seeding rate, Alpha
transmissibility
shared target dispersion
no scaling
OxCGRT floored
OxCGRT independent
Google mobility
Facebook tiles visited mobility
Facebook single tile mobility

### Poland

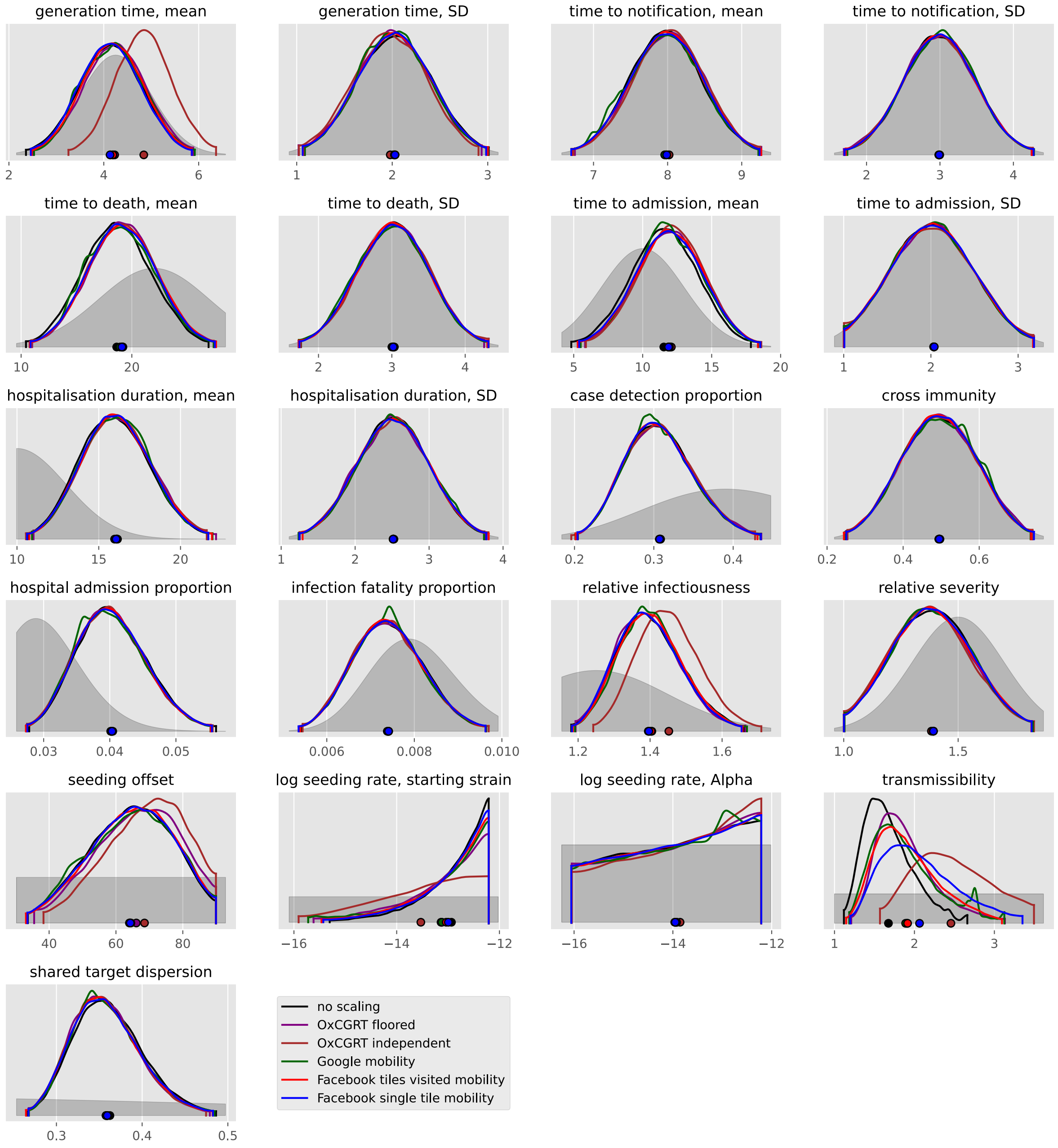

generation time, mean
generation time, SD
time to notification, mean
time to notification, SD
time to death, mean
time to death, SD
time to admission, mean
time to admission, SD
hospitalisation duration, mean
hospitalisation duration, SD
case detection proportion
cross immunity
hospital admission proportion
infection fatality proportion
relative infectiousness
relative severity
seeding offset
log seeding rate, starting strain
log seeding rate, Alpha
transmissibility
shared target dispersion
no scaling
OxCGRT floored
OxCGRT independent
Google mobility
Facebook tiles visited mobility
Facebook single tile mobility

#### Portugal

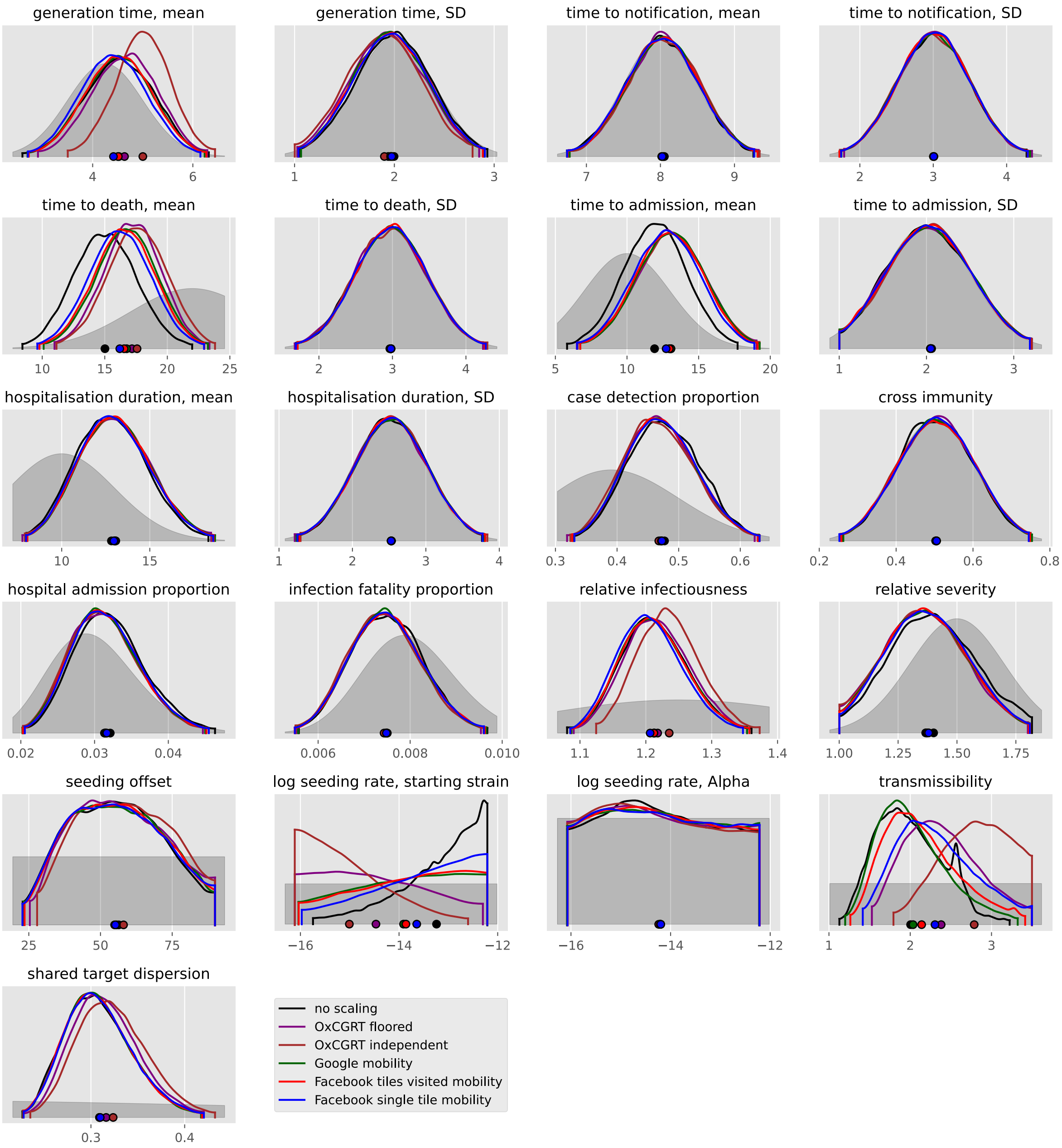

generation time, mean
generation time, SD
time to notification, mean
time to notification, SD
time to death, mean
time to death, SD
time to admission, mean
time to admission, SD
hospitalisation duration, mean
hospitalisation duration, SD
case detection proportion
cross immunity
hospital admission proportion
infection fatality proportion
relative infectiousness
relative severity
seeding offset
log seeding rate, starting strain
log seeding rate, Alpha
transmissibility
shared target dispersion
no scaling
OxCGRT floored
OxCGRT independent
Google mobility
Facebook tiles visited mobility
Facebook single tile mobility

#### Romania

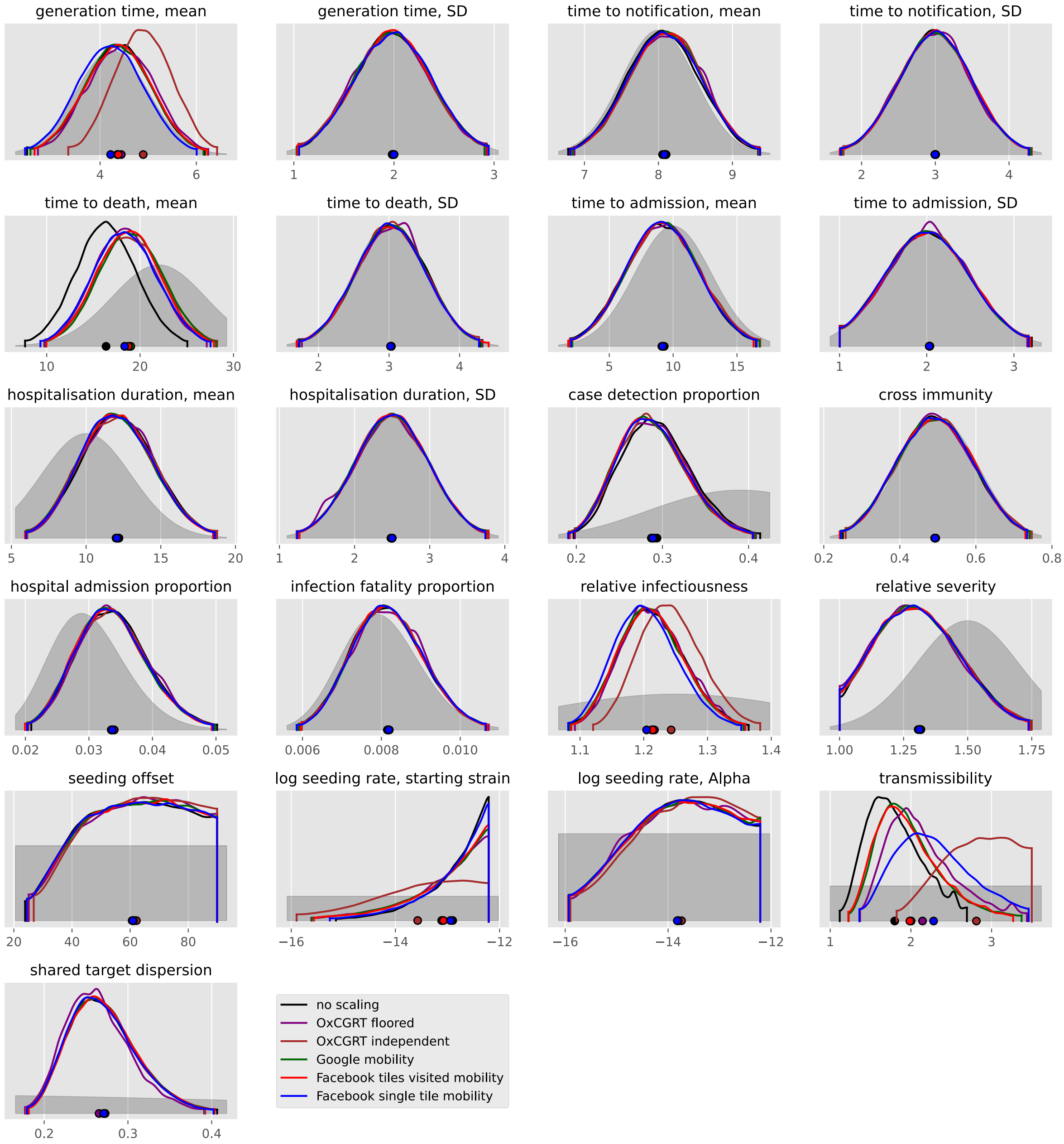
generation time, mean
generation time, SD
time to notification, mean
time to notification, SD
time to death, mean
time to death, SD
time to admission, mean
time to admission, SD
hospitalisation duration, mean
hospitalisation duration, SD
case detection proportion
cross immunity
hospital admission proportion
infection fatality proportion
relative infectiousness
relative severity
seeding offset
log seeding rate, starting strain
log seeding rate, Alpha
transmissibility
shared target dispersion
no scaling
OxCGRT floored
OxCGRT independent
Google mobility
Facebook tiles visited mobility
Facebook single tile mobility

#### Russian Federation

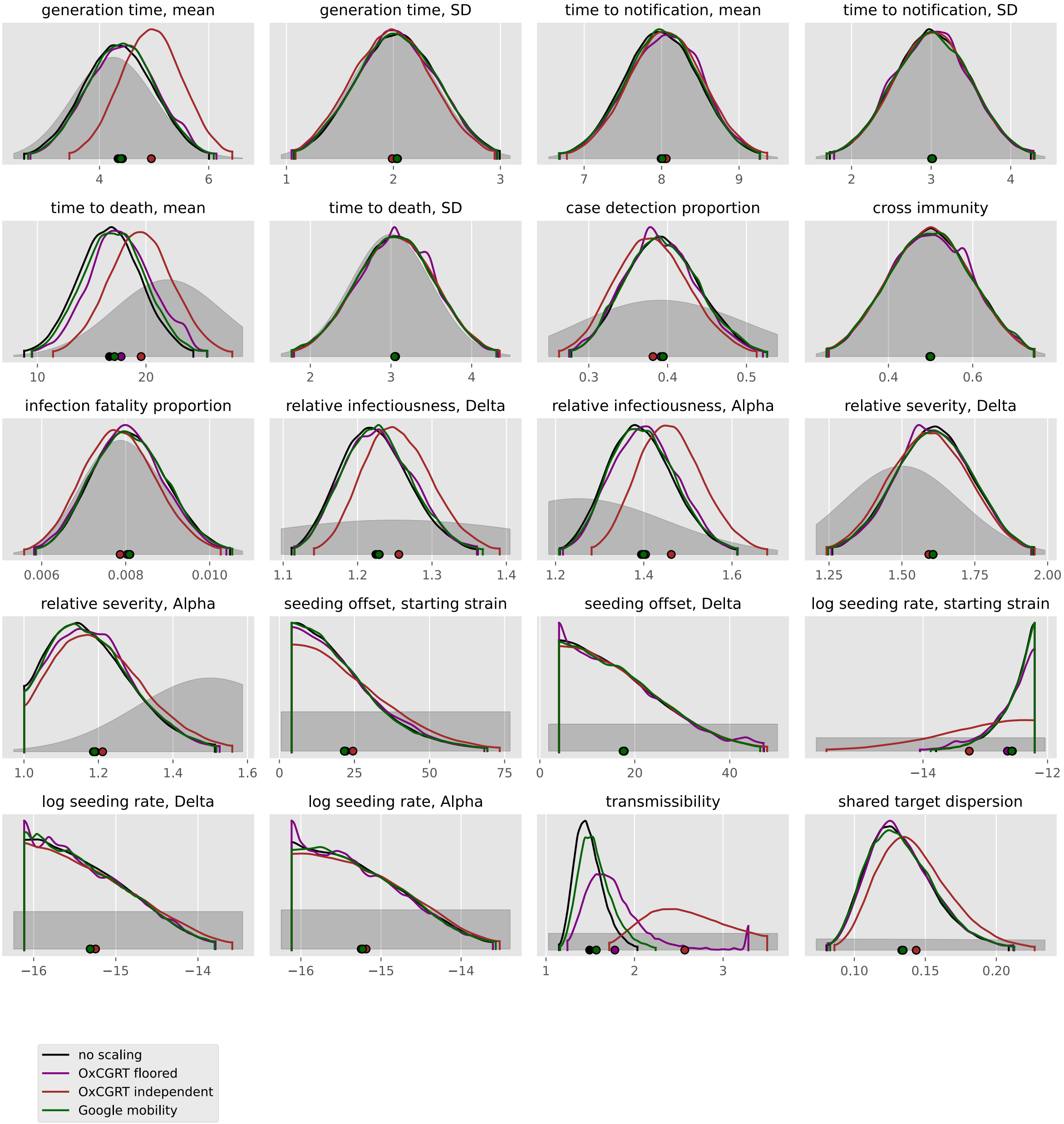

#### Serbia

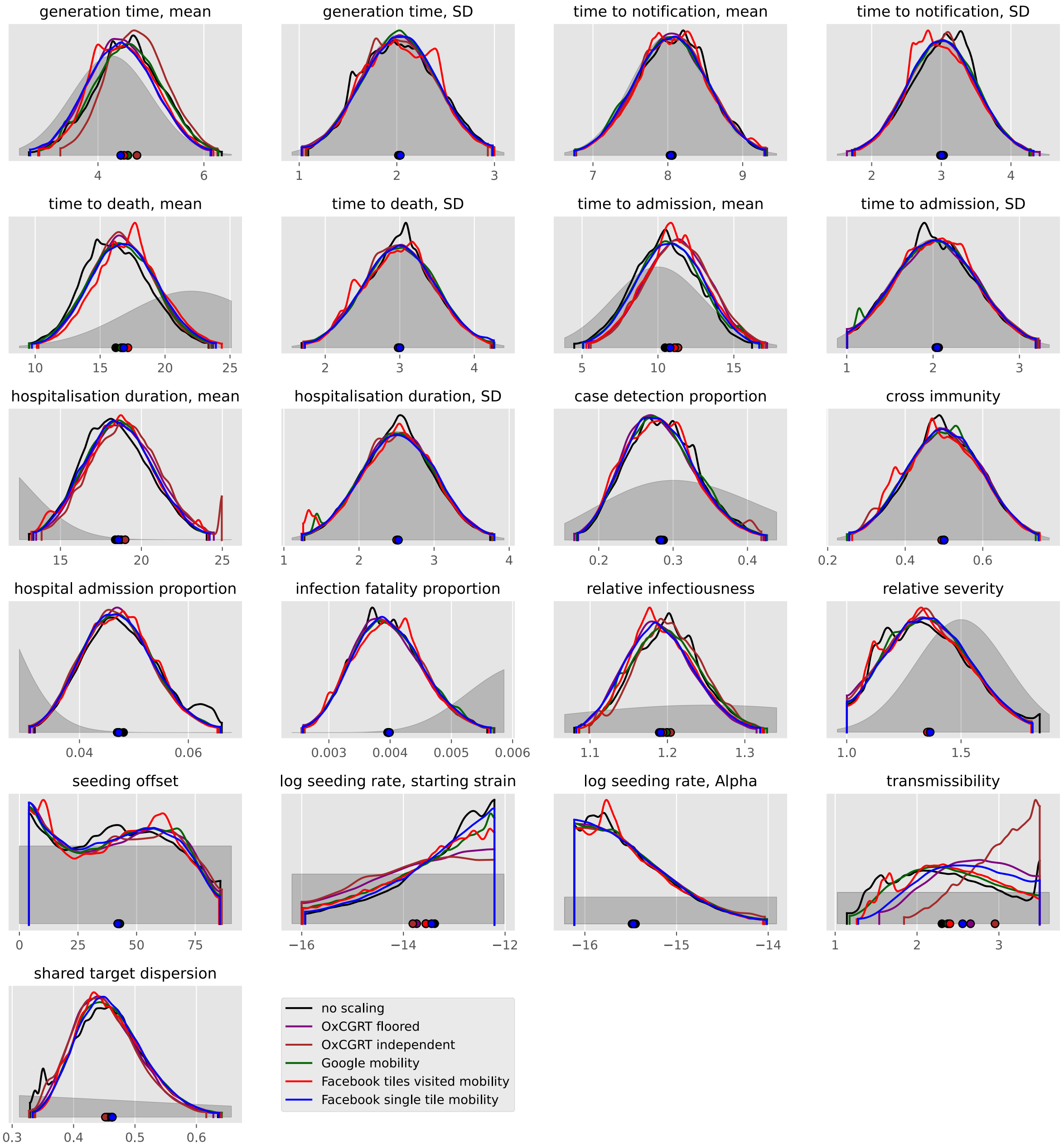
generation time, mean
generation time, SD
time to notification, mean
time to notification, SD
time to death, mean
time to death, SD
time to admission, mean
time to admission, SD
hospitalisation duration, mean
hospitalisation duration, SD
case detection proportion
cross immunity
hospital admission proportion
infection fatality proportion
relative infectiousness
relative severity
seeding offset
log seeding rate, starting strain
log seeding rate, Alpha
transmissibility
shared target dispersion
no scaling
OxCGRT floored
OxCGRT independent
Google mobility
Facebook tiles visited mobility
Facebook single tile mobility

#### Slovakia

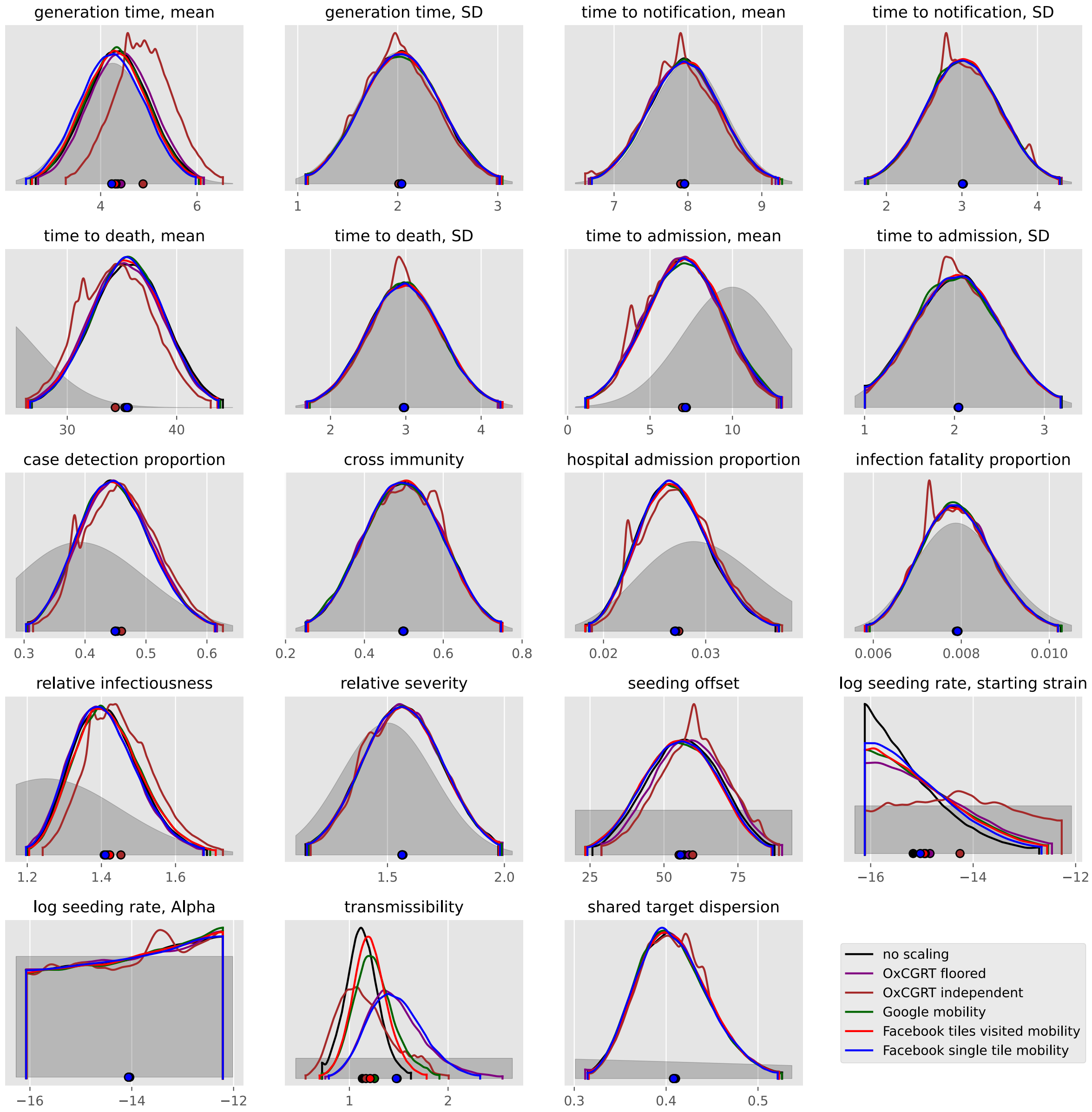
generation time, mean
generation time, SD
time to notification, mean
time to notification, SD
time to death, mean
time to death, SD
time to admission, mean
time to admission, SD
case detection proportion
cross immunity
hospital admission proportion
infection fatality proportion
relative infectiousness
relative severity
seeding offset
log seeding rate, starting strain
log seeding rate, Alpha
transmissibility
shared target dispersion
no scaling
OxCGRT floored
OxCGRT independent
Google mobility
Facebook tiles visited mobility
Facebook single tile mobility

#### Slovenia

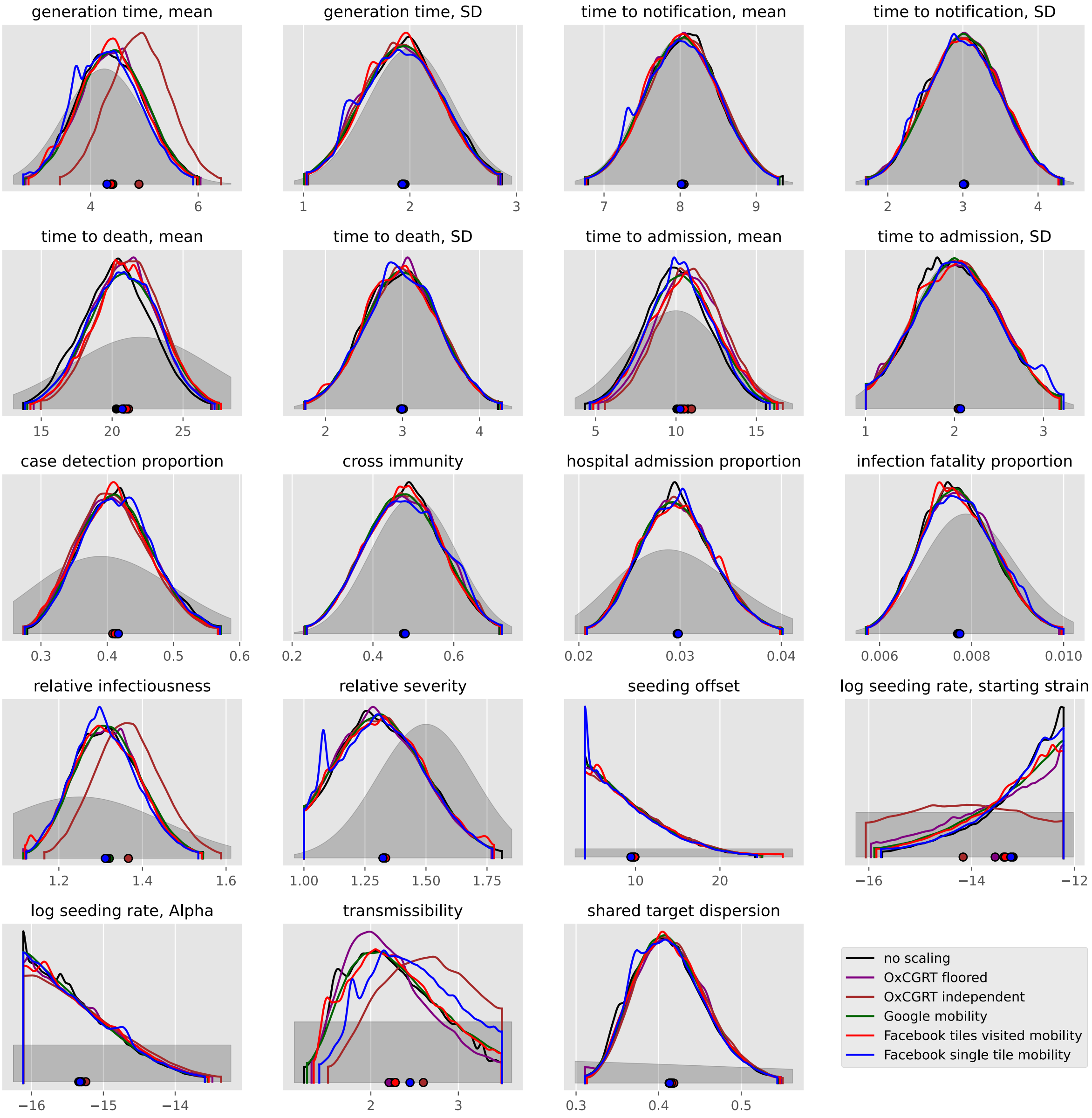
generation time, mean
generation time, SD
time to notification, mean
time to notification, SD
time to death, mean
time to death, SD
time to admission, mean
time to admission, SD
case detection proportion
cross immunity
hospital admission proportion
infection fatality proportion
relative infectiousness
relative severity
seeding offset
log seeding rate, starting strain
log seeding rate, Alpha
transmissibility
shared target dispersion
no scaling
OxCGRT floored
OxCGRT independent
Google mobility
Facebook tiles visited mobility
Facebook single tile mobility

#### Spain

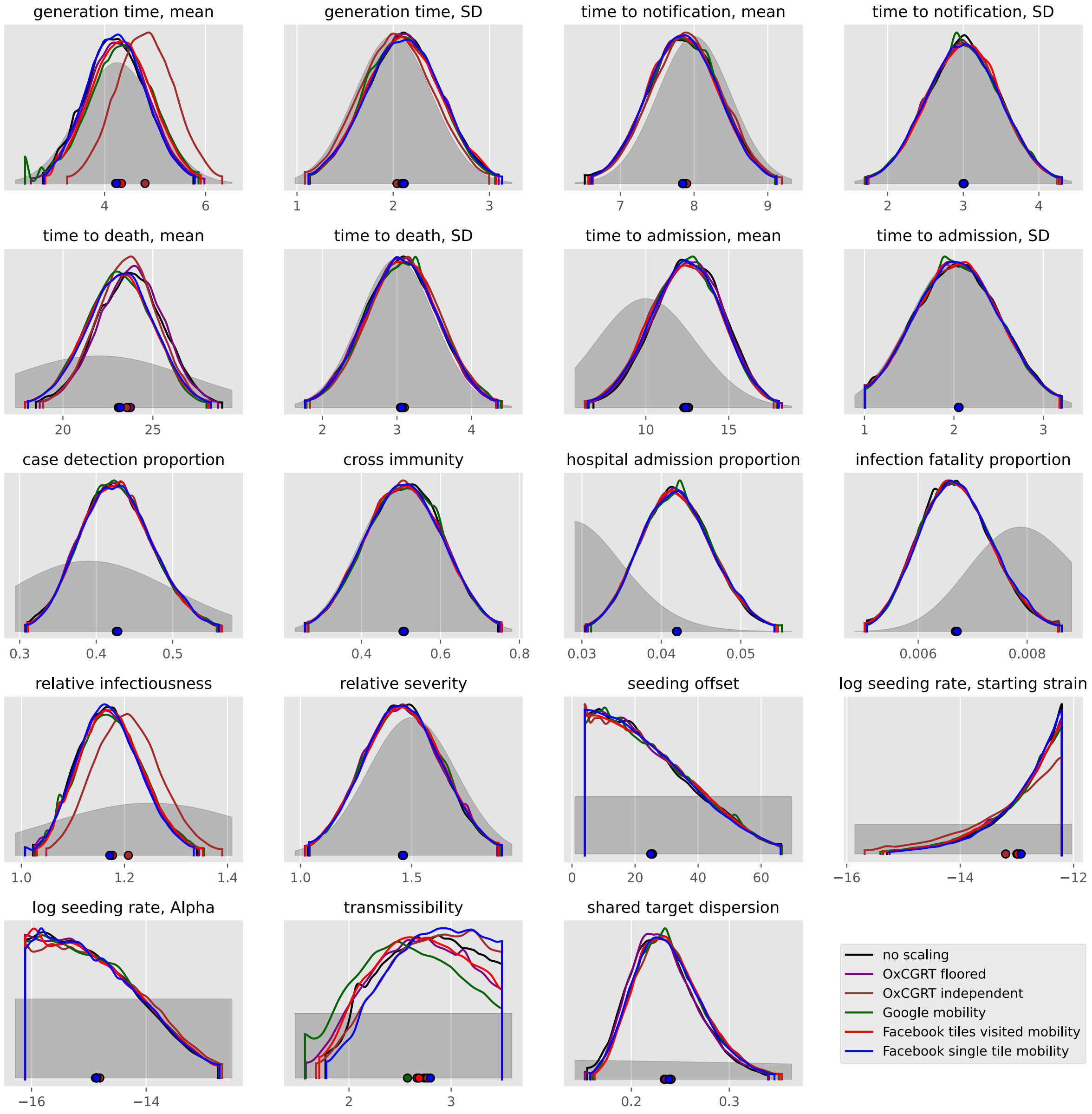

generation time, mean
4
6
generation time, SD
1
2
3
time to notification, mean
7
8
9
time to notification, SD
2
3
4
time to death, mean
20
25
time to death, SD
2
3
4
time to admission, mean
10
15
time to admission, SD
1
2
3
case detection proportion
0.3
0.4
0.5
cross immunity
0.4
0.6
0.8
hospital admission proportion
0.03
0.04
0.05
infection fatality proportion
0.006
0.008
relative infectiousness
1.0
1.2
1.4
relative severity
1.0
1.5
seeding offset
0
20
40
60
log seeding rate, starting strain
−16
−14
−12
log seeding rate, Alpha
−16
−14
transmissibility
2
3
shared target dispersion
0.2
0.3
no scaling
OxCGRT floored
OxCGRT independent
Google mobility
Facebook tiles visited mobility
Facebook single tile mobility

### Sweden

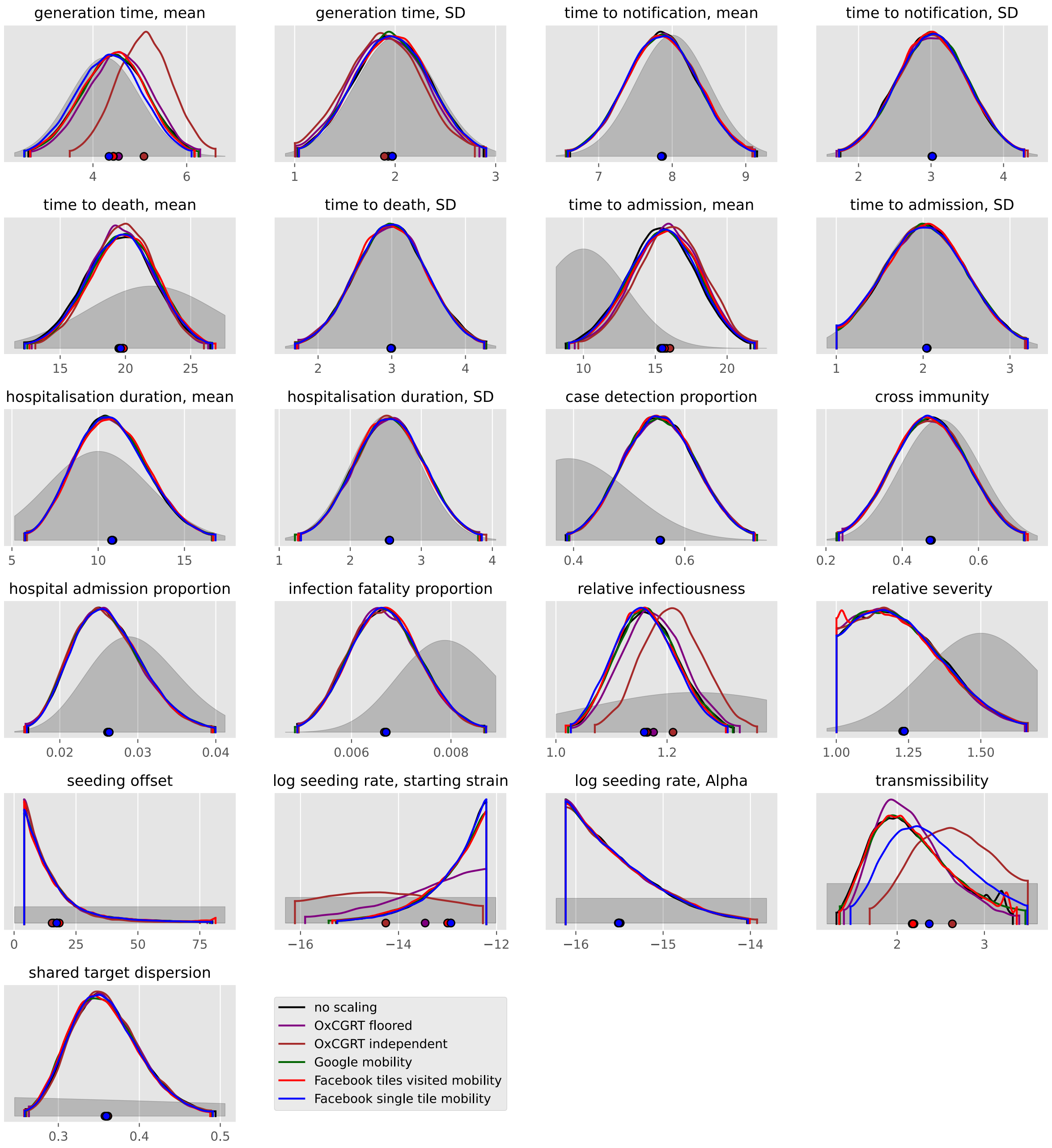
generation time, mean
generation time, SD
time to notification, mean
time to notification, SD
time to death, mean
time to death, SD
time to admission, mean
time to admission, SD
hospitalisation duration, mean
hospitalisation duration, SD
case detection proportion
cross immunity
hospital admission proportion
infection fatality proportion
relative infectiousness
relative severity
seeding offset
log seeding rate, starting strain
log seeding rate, Alpha
transmissibility
shared target dispersion
no scaling
OxCGRT floored
OxCGRT independent
Google mobility
Facebook tiles visited mobility
Facebook single tile mobility

#### Switzerland

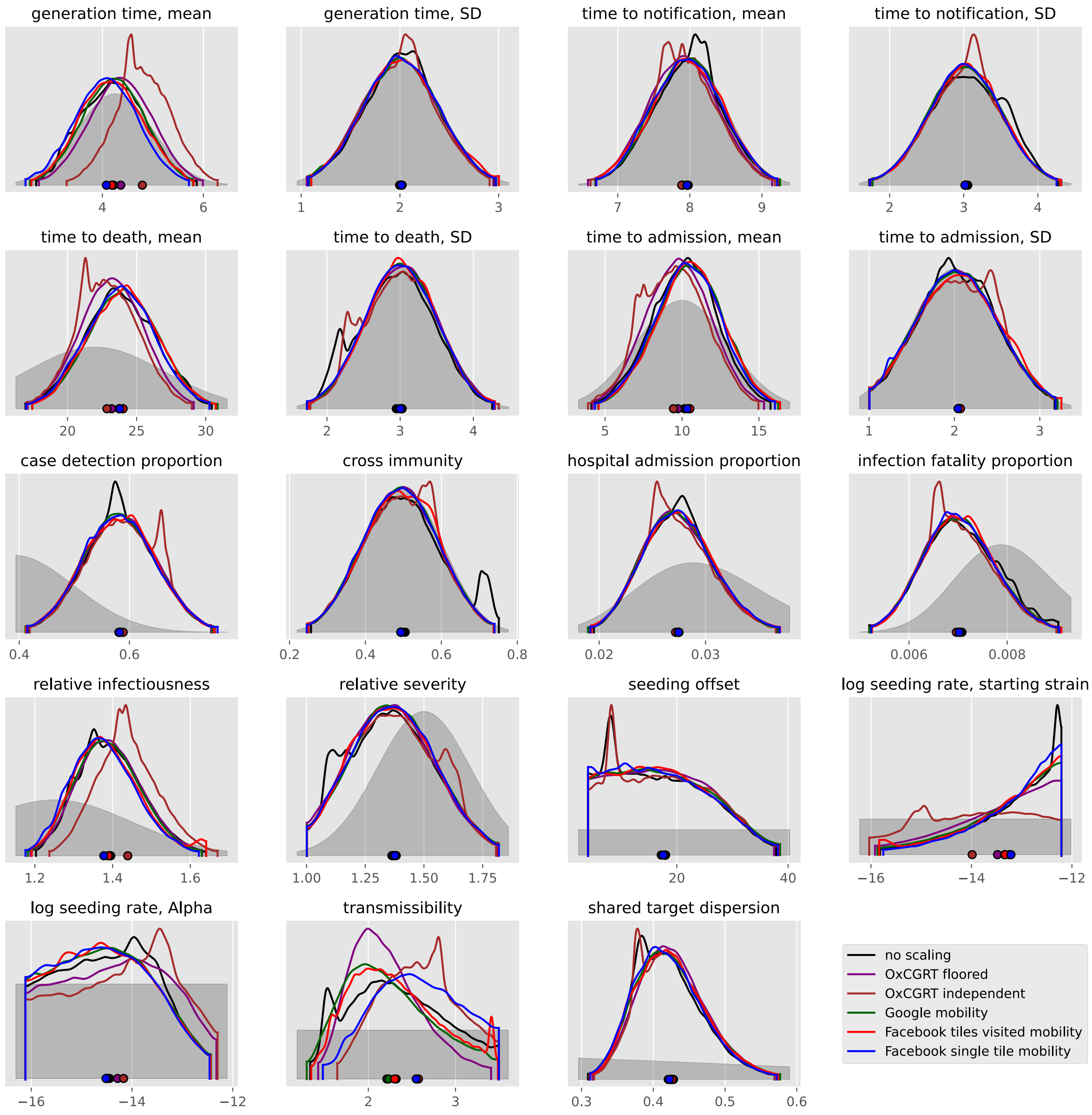

#### Ukraine

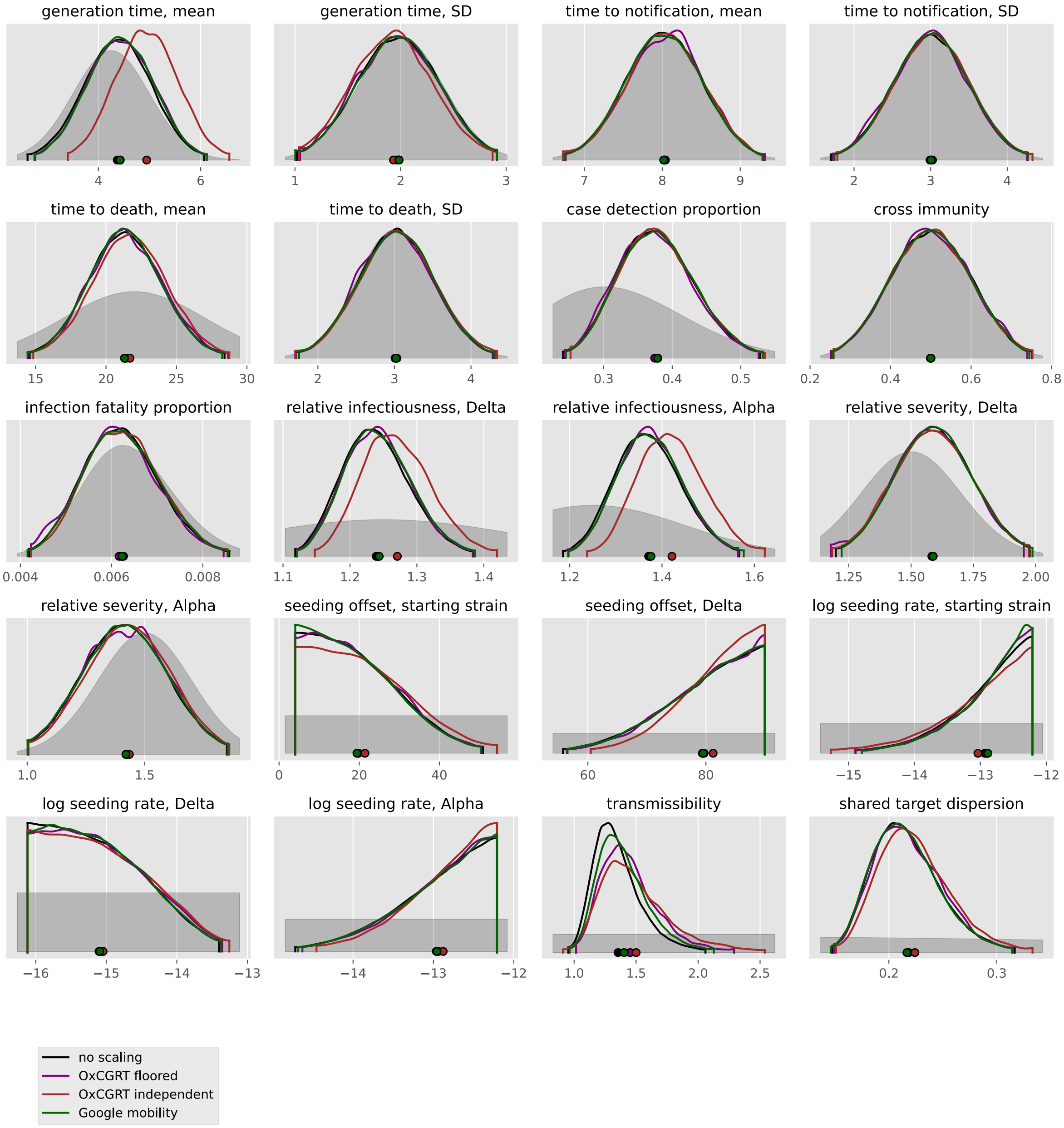

generation time, mean
generation time, SD
time to notification, mean
time to notification, SD
time to death, mean
time to death, SD
case detection proportion
cross immunity
infection fatality proportion
relative infectiousness, Delta
relative infectiousness, Alpha
relative severity, Delta
relative severity, Alpha
seeding offset, starting strain
seeding offset, Delta
log seeding rate, starting strain
log seeding rate, Delta
log seeding rate, Alpha
transmissibility
shared target dispersion
no scaling
OxCGRT floored
OxCGRT independent
Google mobility

#### United Kingdom

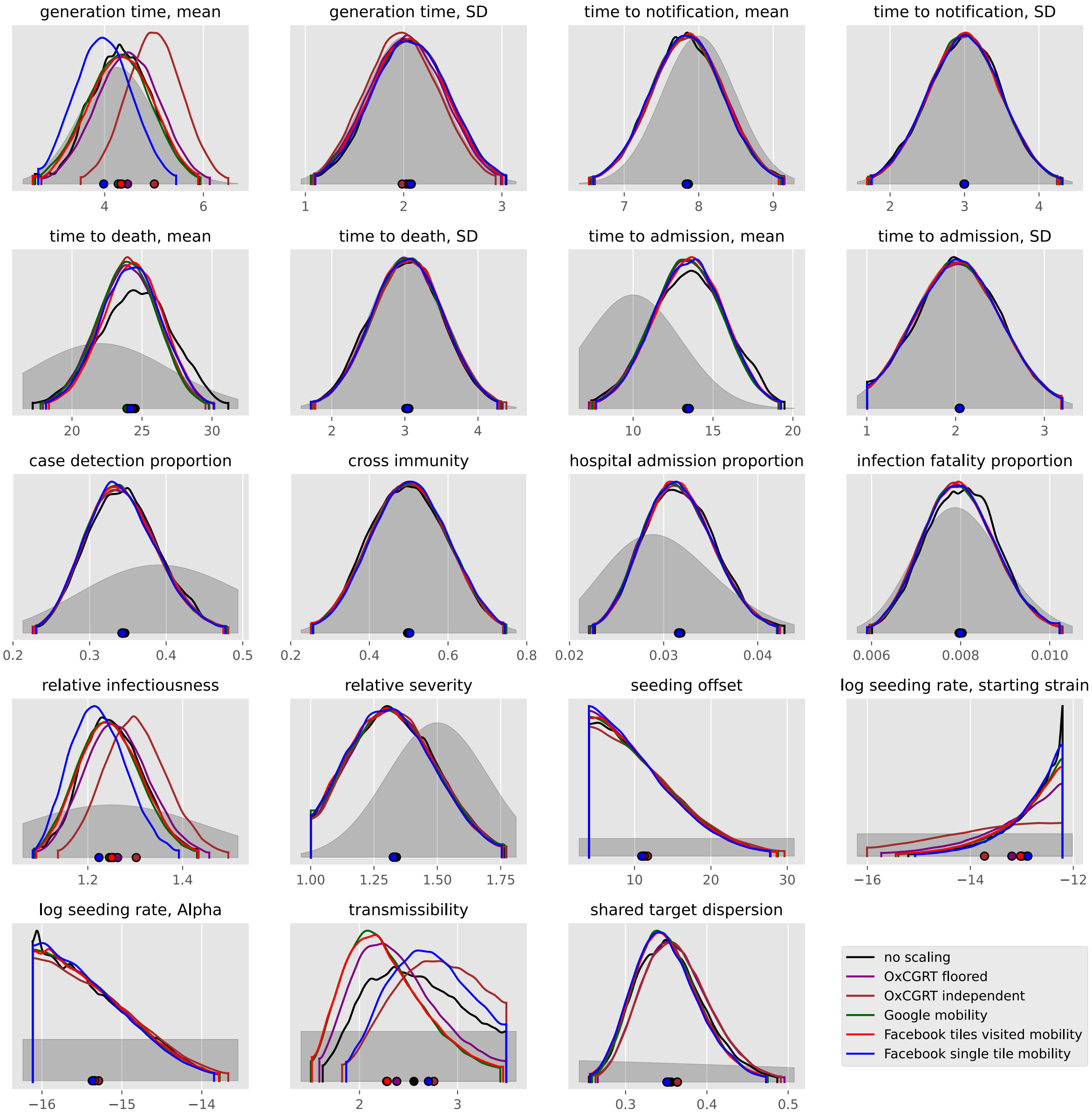

### North America

#### Aruba

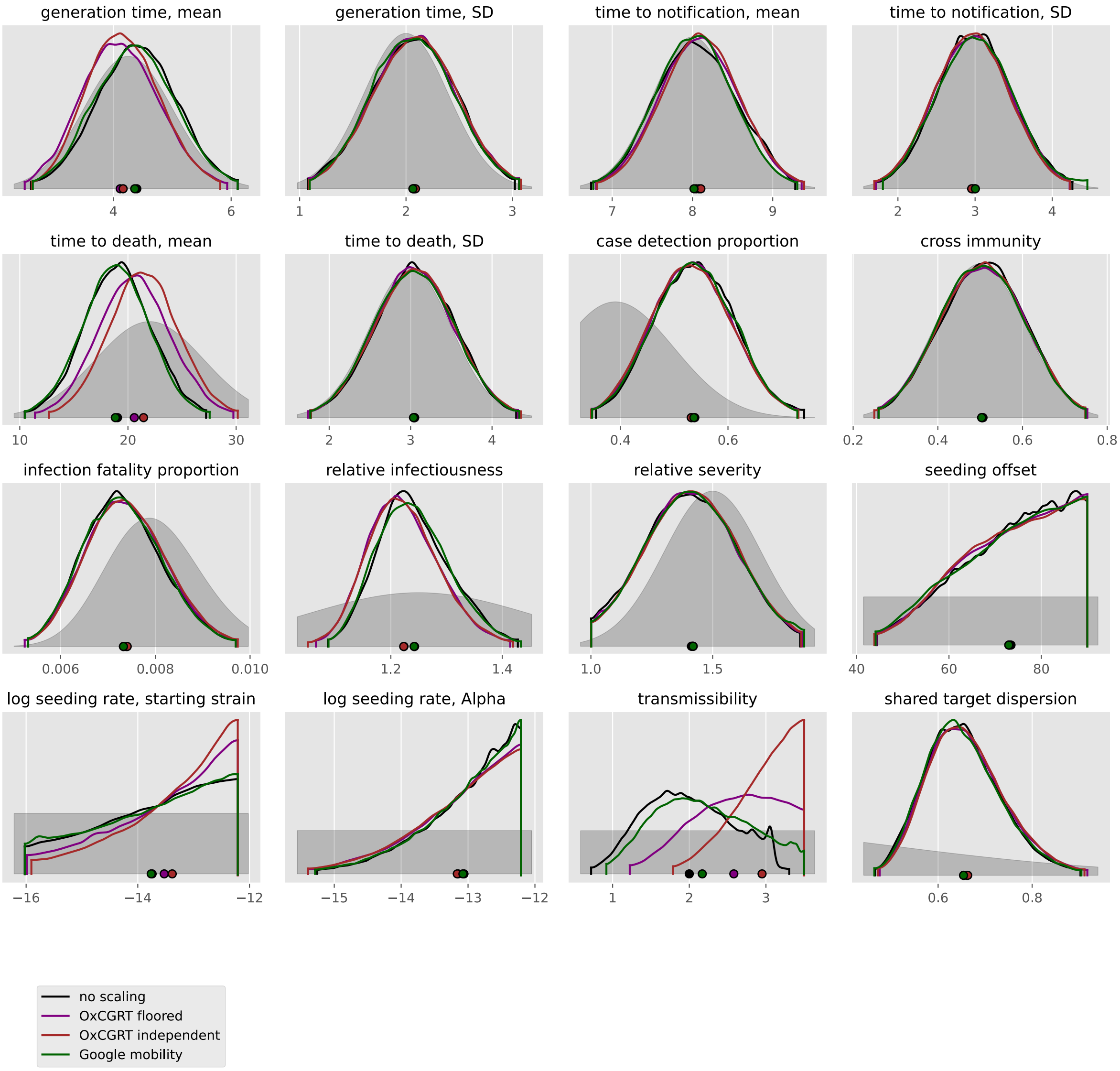

#### Bahamas

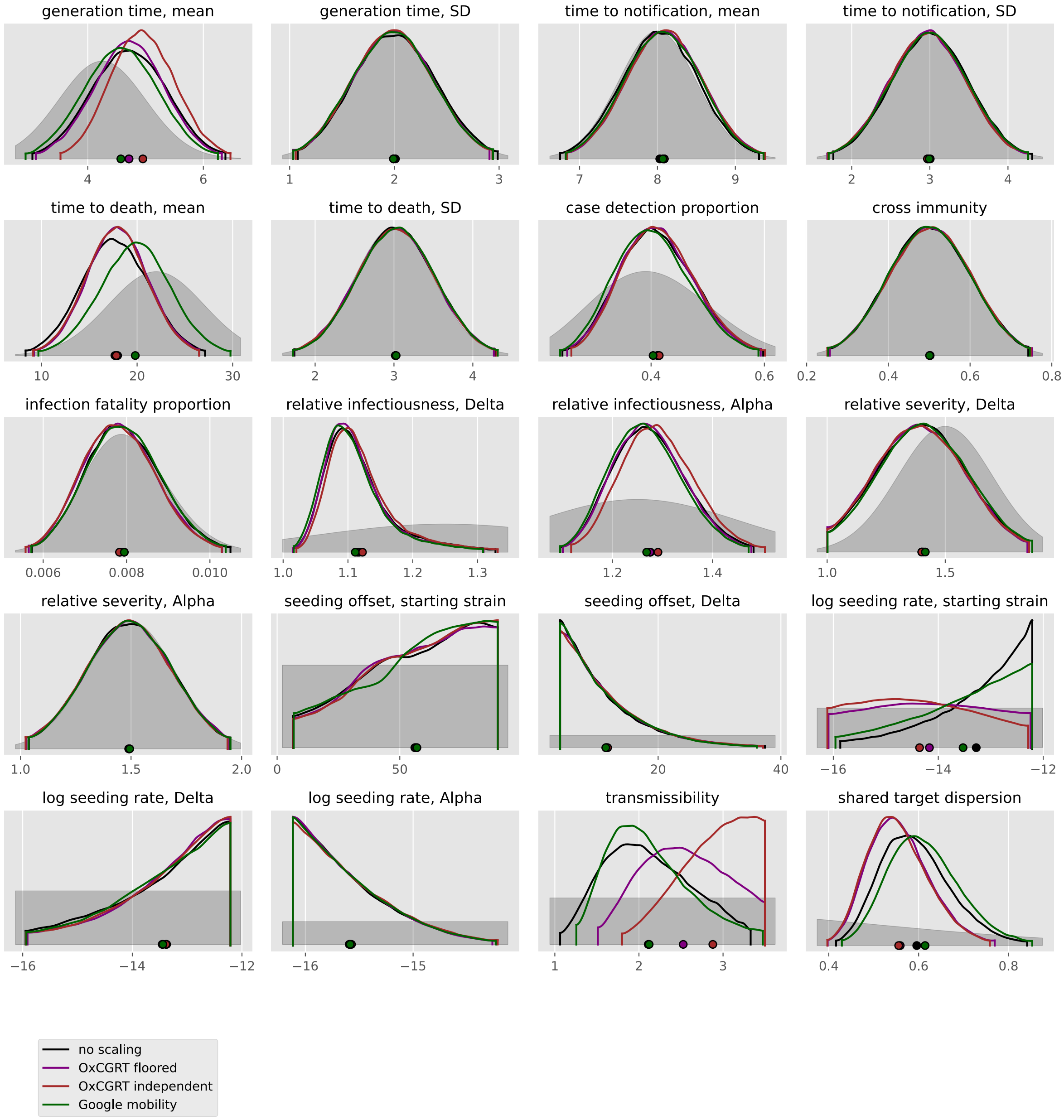

generation time, mean
4
6
generation time, SD
1
2
3
time to notification, mean
7
8
9
time to notification, SD
2
3
4
time to death, mean
10
20
30
time to death, SD
2
3
4
case detection proportion
0.4
0.6
cross immunity
0.2
0.4
0.6
0.8
infection fatality proportion
0.006
0.008
0.010
relative infectiousness, Delta
1.0
1.1
1.2
1.3
relative infectiousness, Alpha
1.2
1.4
relative severity, Delta
1.0
1.5
relative severity, Alpha
1.0
1.5
2.0
seeding offset, starting strain
0
50
seeding offset, Delta
20
40
log seeding rate, starting strain
−16
−14
−12
log seeding rate, Delta
−16
−14
−12
log seeding rate, Alpha
−16
−15
transmissibility
1
2
3
shared target dispersion
0.4
0.6
0.8
no scaling
OxCGRT floored
OxCGRT independent
Google mobility

#### Canada

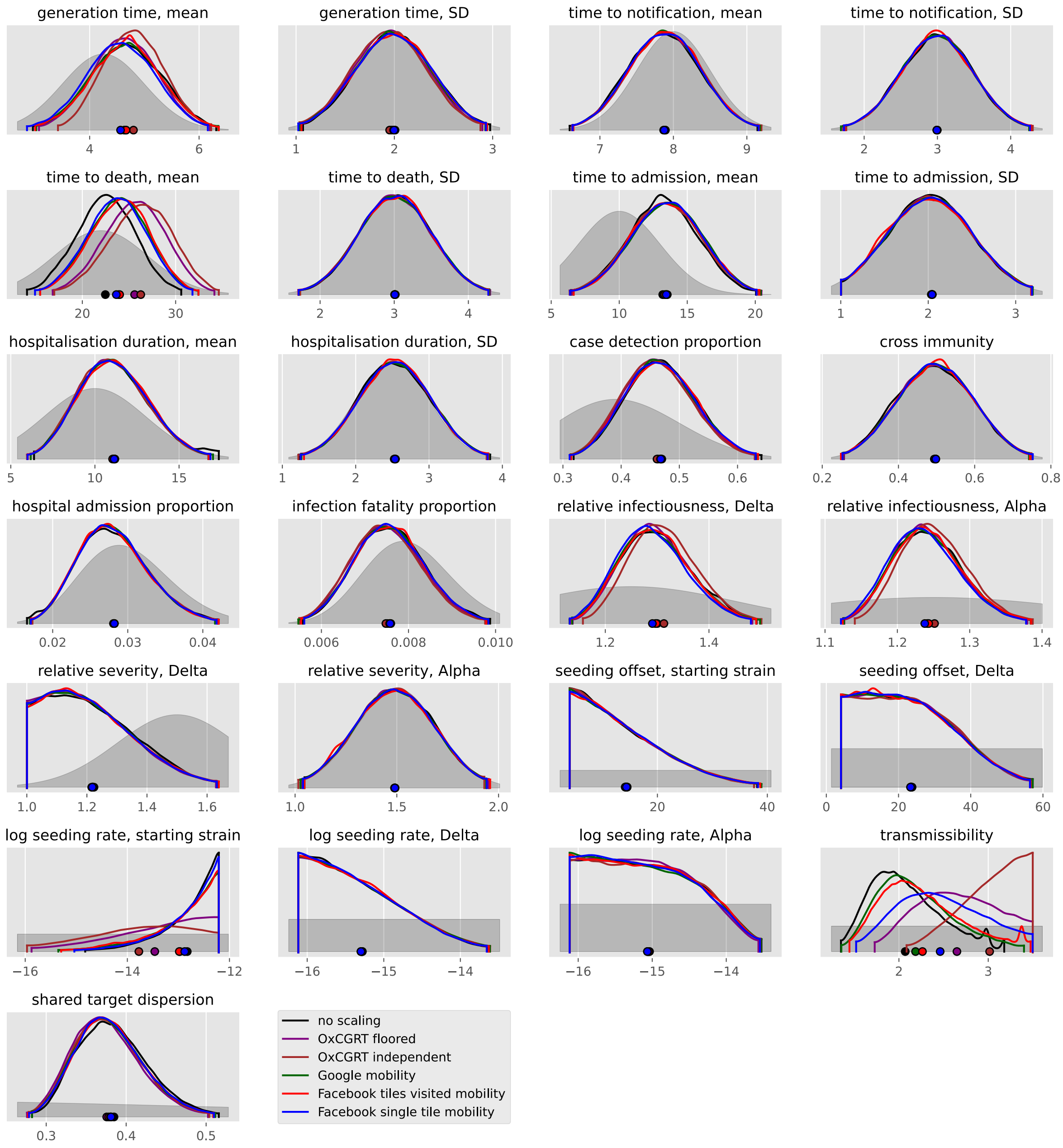
generation time, mean
generation time, SD
time to notification, mean
time to notification, SD
time to death, mean
time to death, SD
time to admission, mean
time to admission, SD
hospitalisation duration, mean
hospitalisation duration, SD
case detection proportion
cross immunity
hospital admission proportion
infection fatality proportion
relative infectiousness, Delta
relative infectiousness, Alpha
relative severity, Delta
relative severity, Alpha
seeding offset, starting strain
seeding offset, Delta
log seeding rate, starting strain
log seeding rate, Delta
log seeding rate, Alpha
transmissibility
shared target dispersion
no scaling
OxCGRT floored
OxCGRT independent
Google mobility
Facebook tiles visited mobility
Facebook single tile mobility

#### Costa Rica

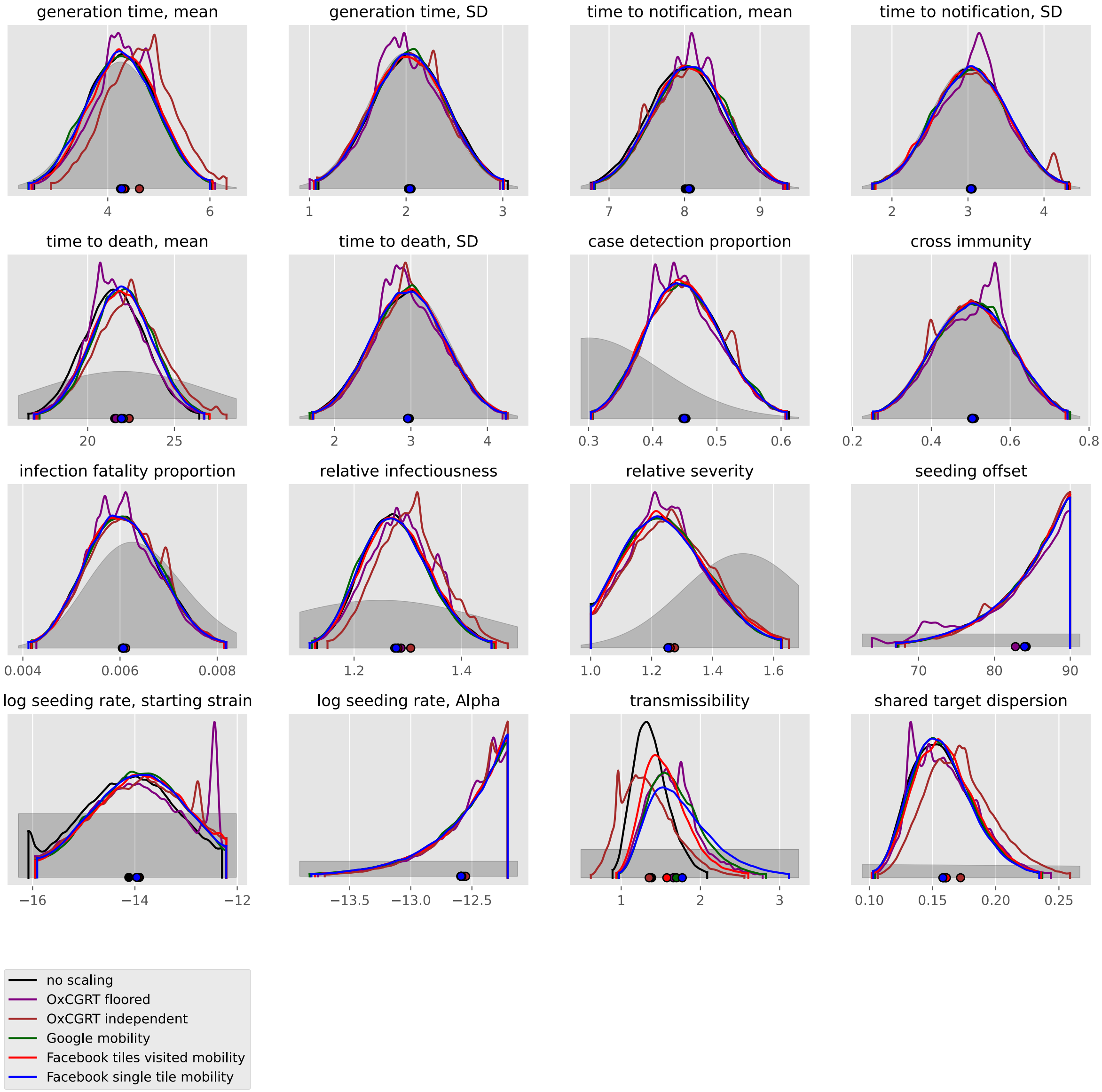

generation time, mean
generation time, SD
time to notification, mean
time to notification, SD
time to death, mean
time to death, SD
case detection proportion
cross immunity
infection fatality proportion
relative infectiousness
relative severity
seeding offset
log seeding rate, starting strain
log seeding rate, Alpha
transmissibility
shared target dispersion
no scaling
OxCGRT floored
OxCGRT independent
Google mobility
Facebook tiles visited mobility
Facebook single tile mobility

#### Cuba

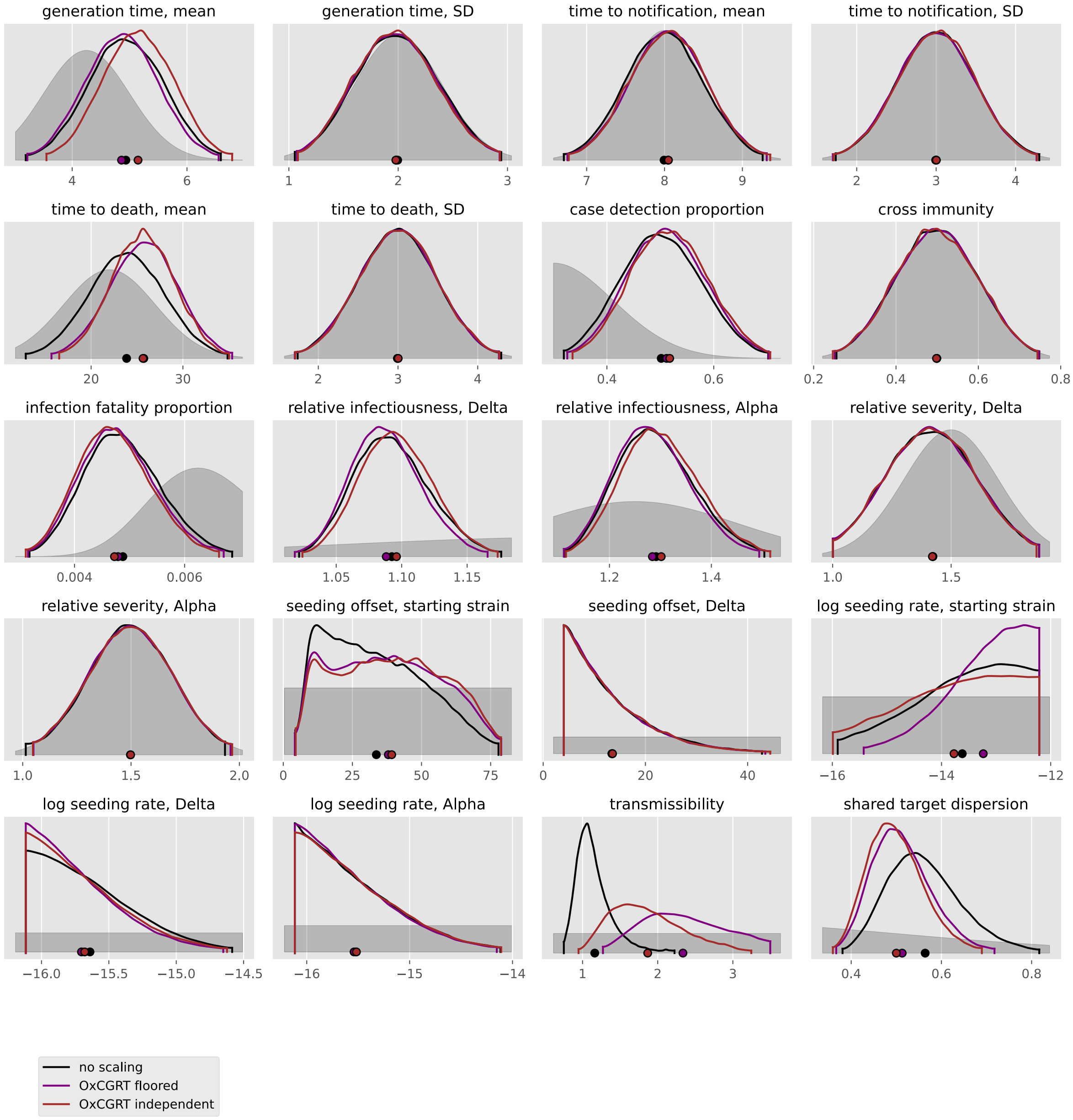

generation time, mean
generation time, SD
time to notification, mean
time to notification, SD
time to death, mean
time to death, SD
case detection proportion
cross immunity
infection fatality proportion
relative infectiousness, Delta
relative infectiousness, Alpha
relative severity, Delta
relative severity, Alpha
seeding offset, starting strain
seeding offset, Delta
log seeding rate, starting strain
log seeding rate, Delta
log seeding rate, Alpha
transmissibility
shared target dispersion
no scaling
OxCGRT floored
OxCGRT independent

#### Dominican Republic

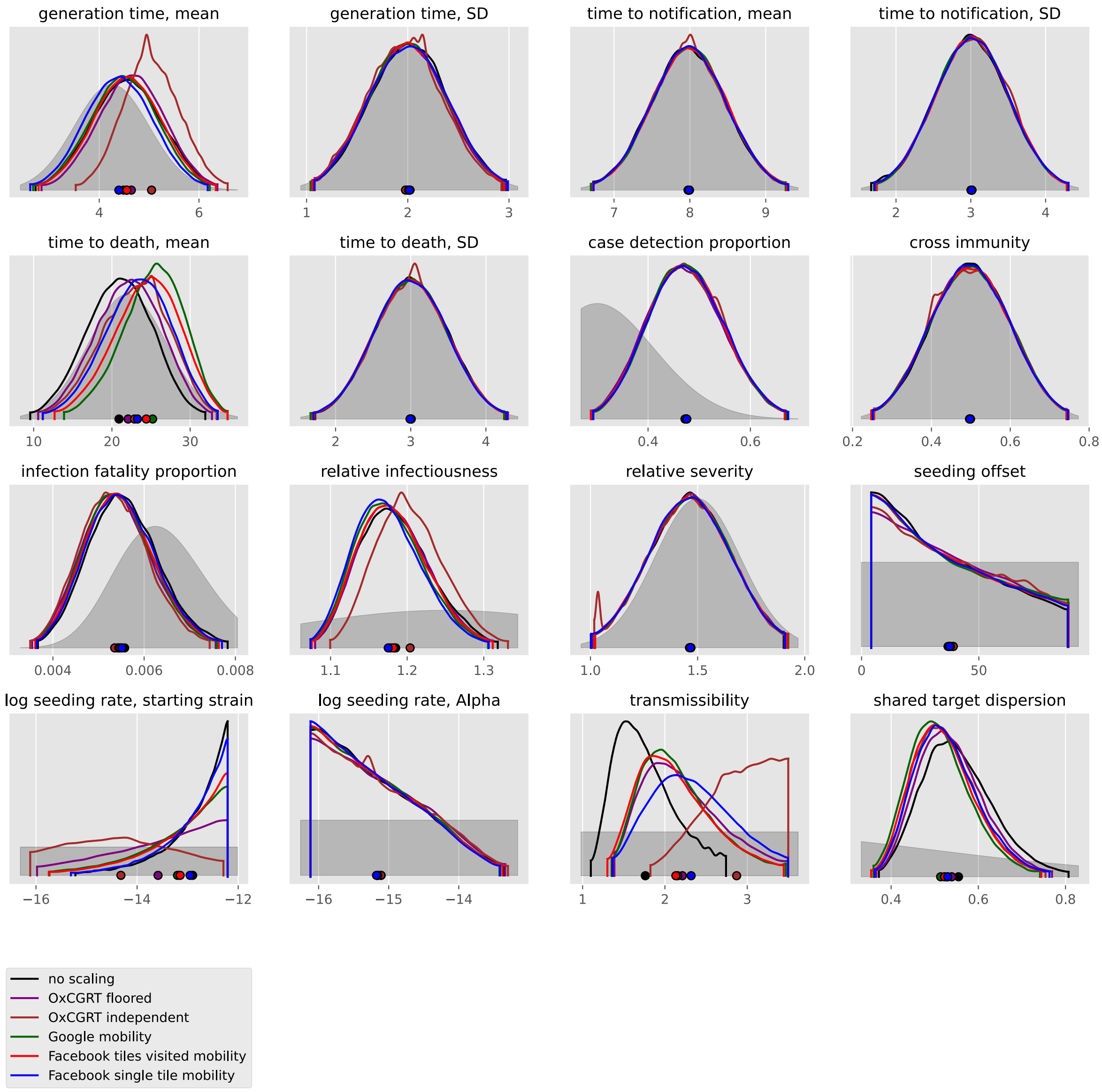

#### El Salvador

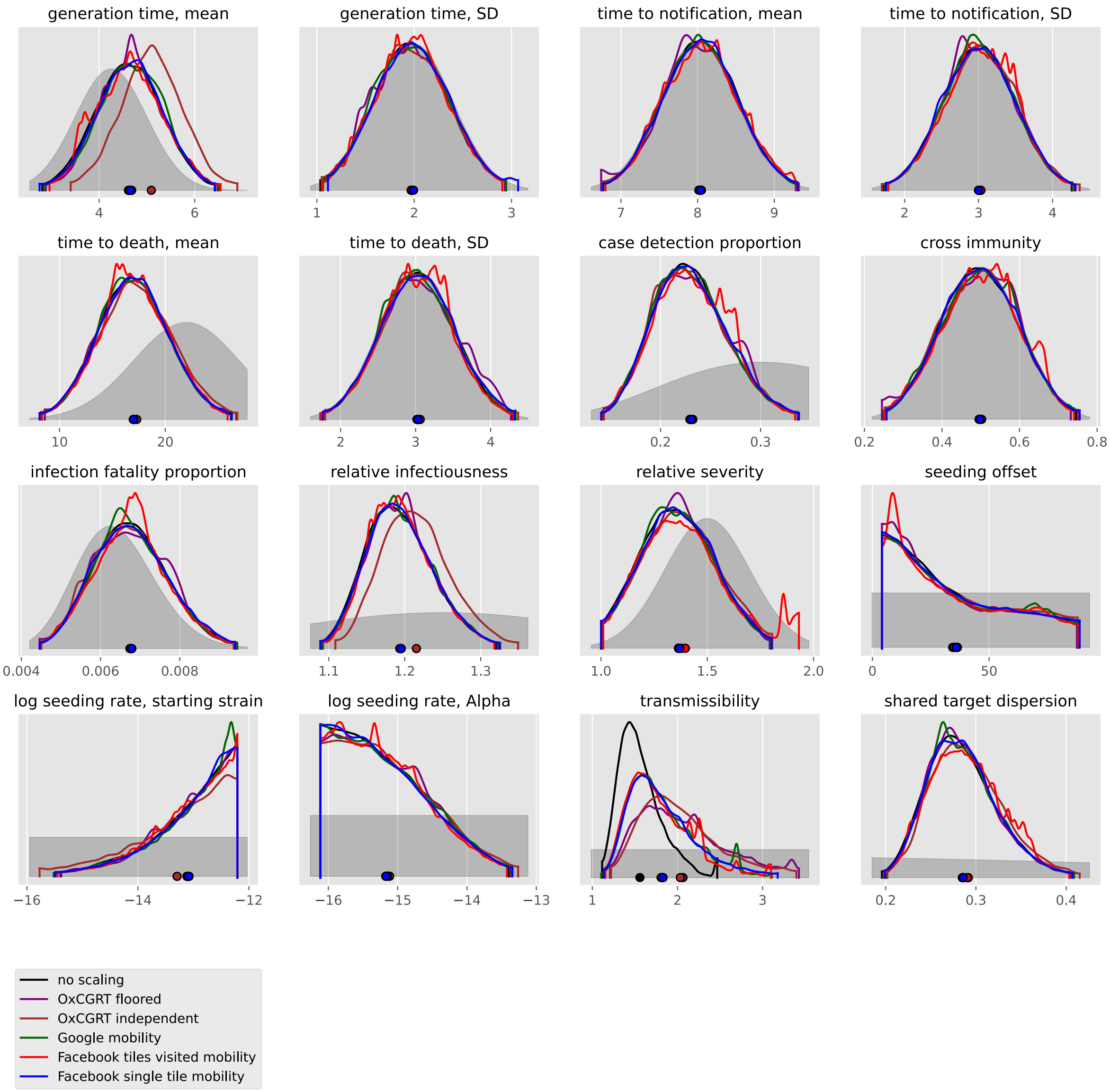

generation time, mean
generation time, SD
time to notification, mean
time to notification, SD
time to death, mean
time to death, SD
case detection proportion
cross immunity
infection fatality proportion
relative infectiousness
relative severity
seeding offset
log seeding rate, starting strain
log seeding rate, Alpha
transmissibility
shared target dispersion
no scaling
OxCGRT floored
OxCGRT independent
Google mobility
Facebook tiles visited mobility
Facebook single tile mobility

#### Haiti

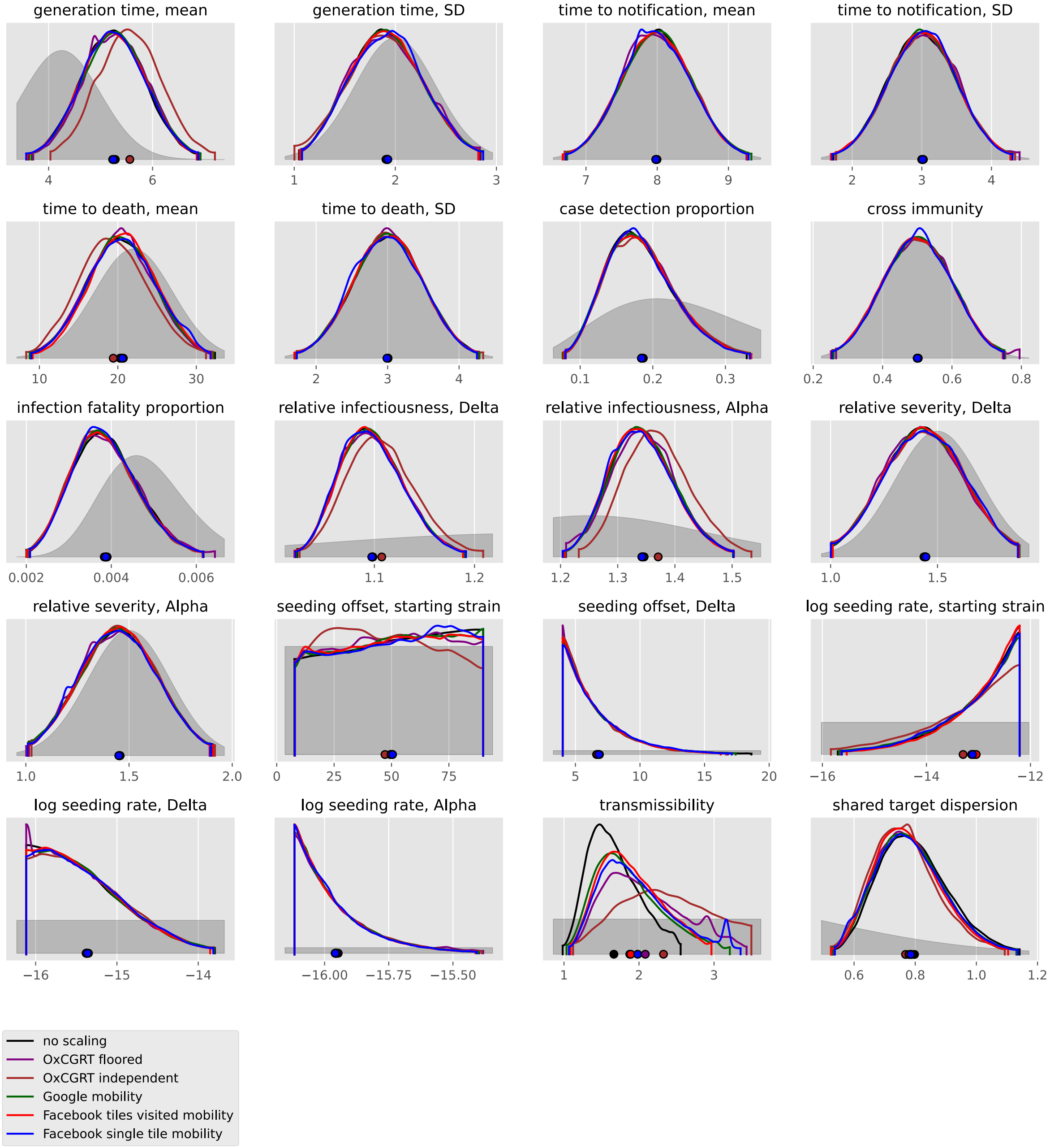
generation time, mean
4
6
generation time, SD
1
2
3
time to notification, mean
7
8
9
time to notification, SD
2
3
4
time to death, mean
10
20
30
time to death, SD
2
3
4
case detection proportion
0.1
0.2
0.3
cross immunity
0.2
0.4
0.6
0.8
infection fatality proportion
0.002
0.004
0.006
relative infectiousness, Delta
1.1
1.2
relative infectiousness, Alpha
1.2
1.3
1.4
1.5
relative severity, Delta
1.0
1.5
relative severity, Alpha
1.0
1.5
2.0
seeding offset, starting strain
0
25
50
75
seeding offset, Delta
5
10
15
20
log seeding rate, starting strain
−16
−14
−12
log seeding rate, Delta
−16
−15
−14
log seeding rate, Alpha
−16.00
−15.75
−15.50
transmissibility
1
2
3
shared target dispersion
0.6
0.8
1.0
1.2
no scaling
OxCGRT floored
OxCGRT independent
Google mobility
Facebook tiles visited mobility
Facebook single tile mobility

#### Honduras

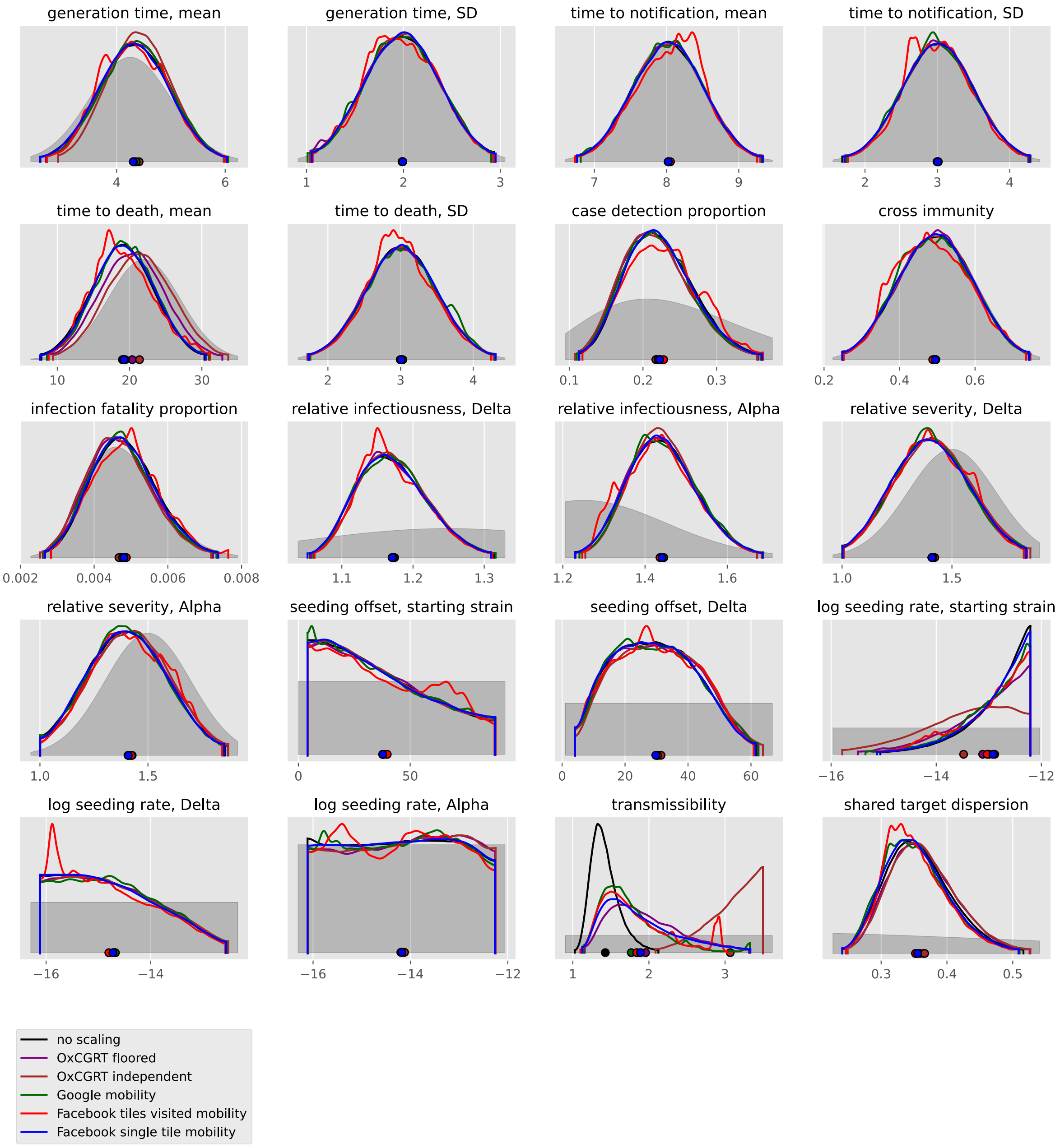

generation time, mean
generation time, SD
time to notification, mean
time to notification, SD
time to death, mean
time to death, SD
case detection proportion
cross immunity
infection fatality proportion
relative infectiousness, Delta
relative infectiousness, Alpha
relative severity, Delta
relative severity, Alpha
seeding offset, starting strain
seeding offset, Delta
log seeding rate, starting strain
log seeding rate, Delta
log seeding rate, Alpha
transmissibility
shared target dispersion
no scaling
OxCGRT floored
OxCGRT independent
Google mobility
Facebook tiles visited mobility
Facebook single tile mobility

#### Jamaica

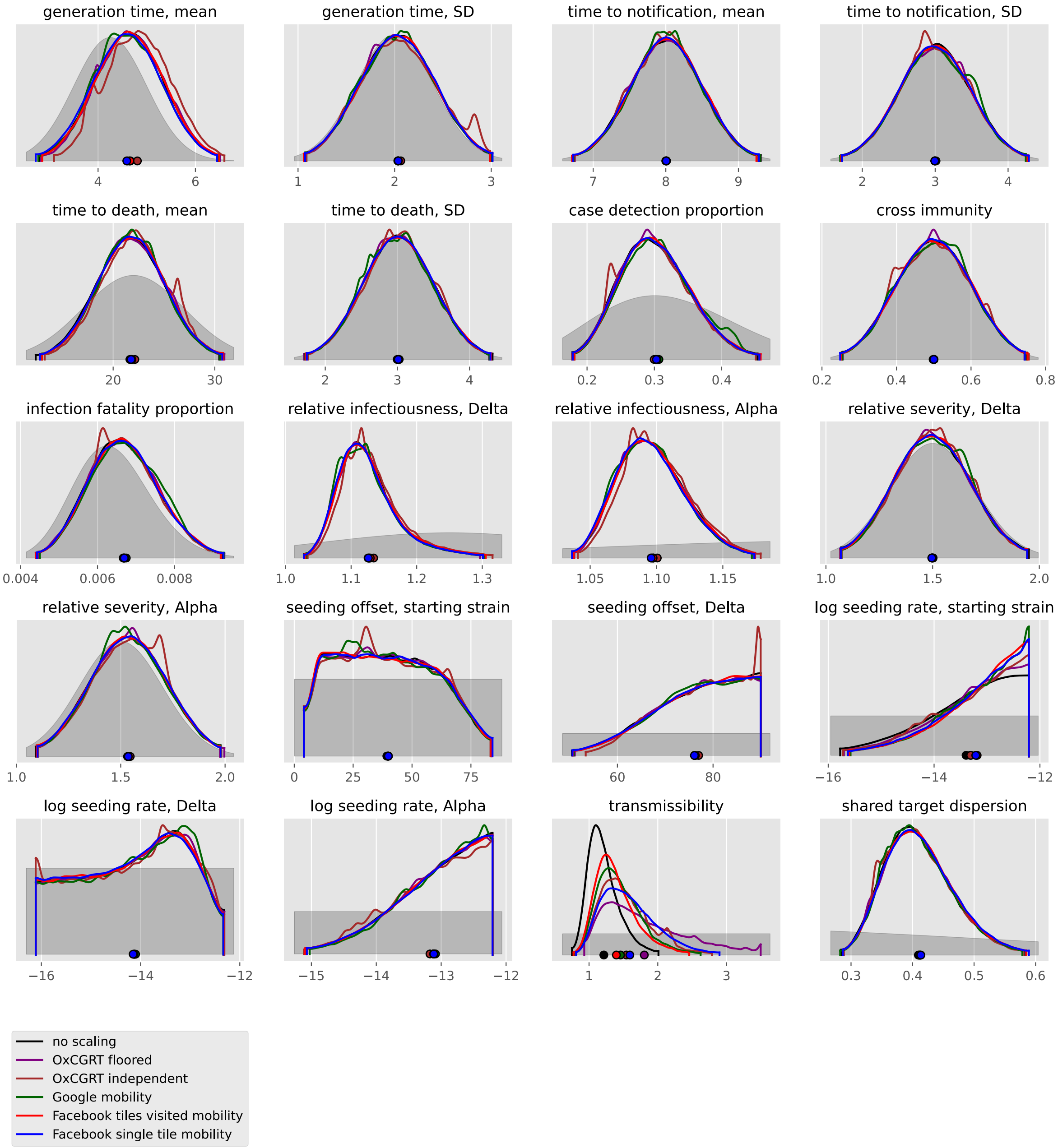
generation time, mean
generation time, SD
time to notification, mean
time to notification, SD
time to death, mean
time to death, SD
case detection proportion
cross immunity
infection fatality proportion
relative infectiousness, Delta
relative infectiousness, Alpha
relative severity, Delta
relative severity, Alpha
seeding offset, starting strain
seeding offset, Delta
log seeding rate, starting strain
log seeding rate, Delta
log seeding rate, Alpha
transmissibility
shared target dispersion
no scaling
OxCGRT floored
OxCGRT independent
Google mobility
Facebook tiles visited mobility
Facebook single tile mobility

#### Mexico

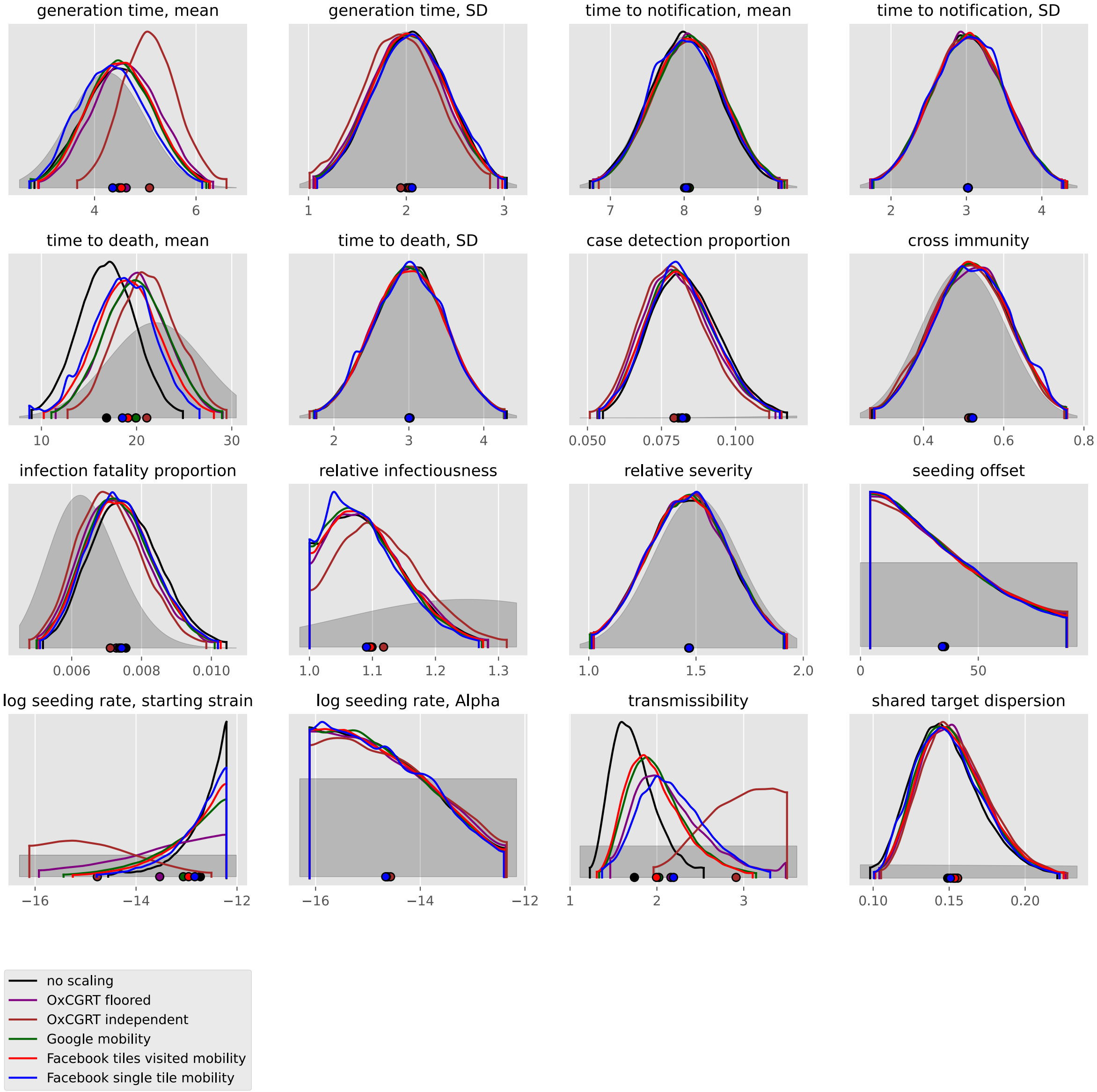

generation time, mean
generation time, SD
time to notification, mean
time to notification, SD
time to death, mean
time to death, SD
case detection proportion
cross immunity
infection fatality proportion
relative infectiousness
relative severity
seeding offset
log seeding rate, starting strain
log seeding rate, Alpha
transmissibility
shared target dispersion
no scaling
OxCGRT floored
OxCGRT independent
Google mobility
Facebook tiles visited mobility
Facebook single tile mobility

#### Panama

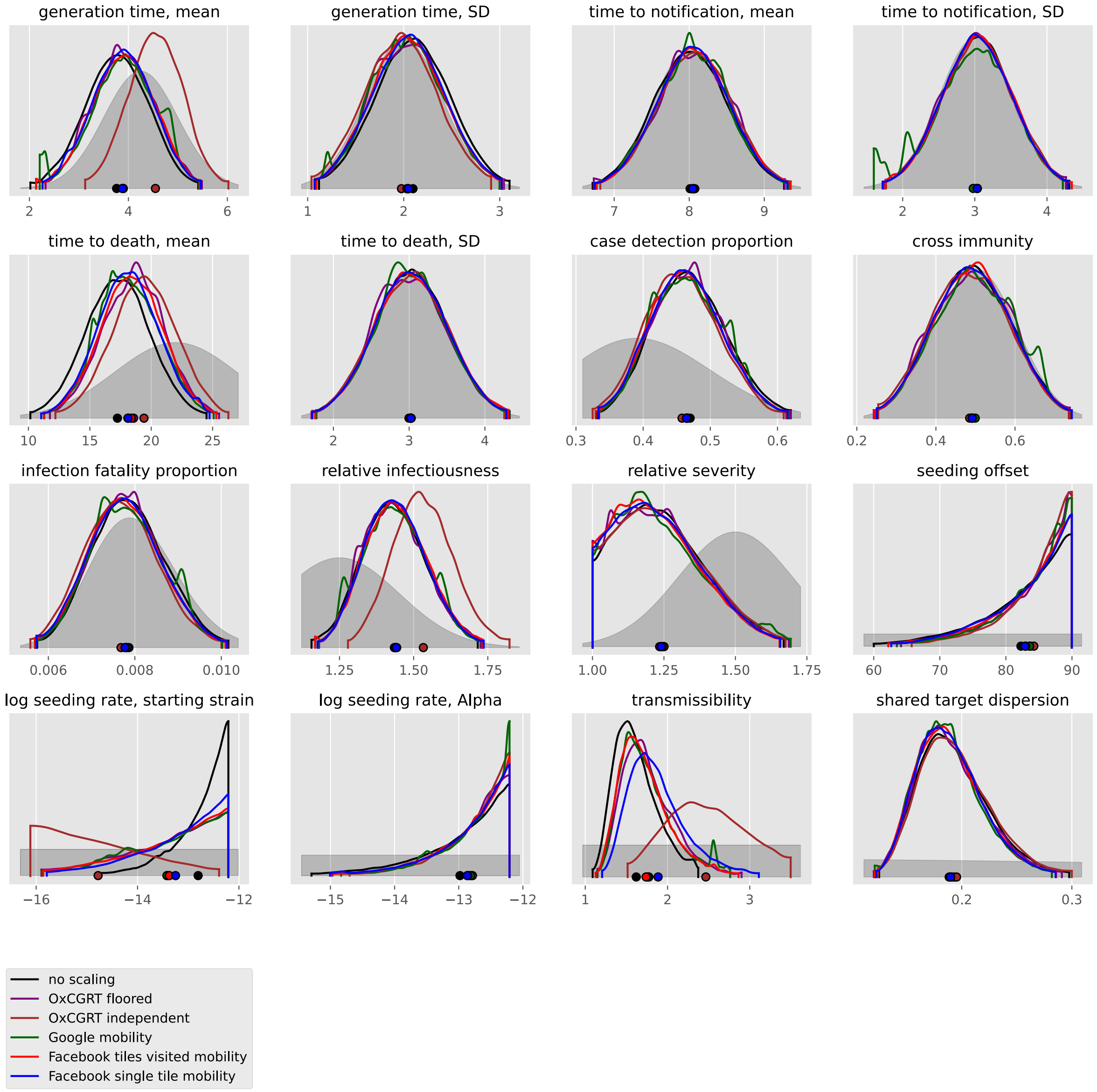

generation time, mean
generation time, SD
time to notification, mean
time to notification, SD
time to death, mean
time to death, SD
case detection proportion
cross immunity
infection fatality proportion
relative infectiousness
relative severity
seeding offset
log seeding rate, starting strain
log seeding rate, Alpha
transmissibility
shared target dispersion
no scaling
OxCGRT floored
OxCGRT independent
Google mobility
Facebook tiles visited mobility
Facebook single tile mobility

#### United States

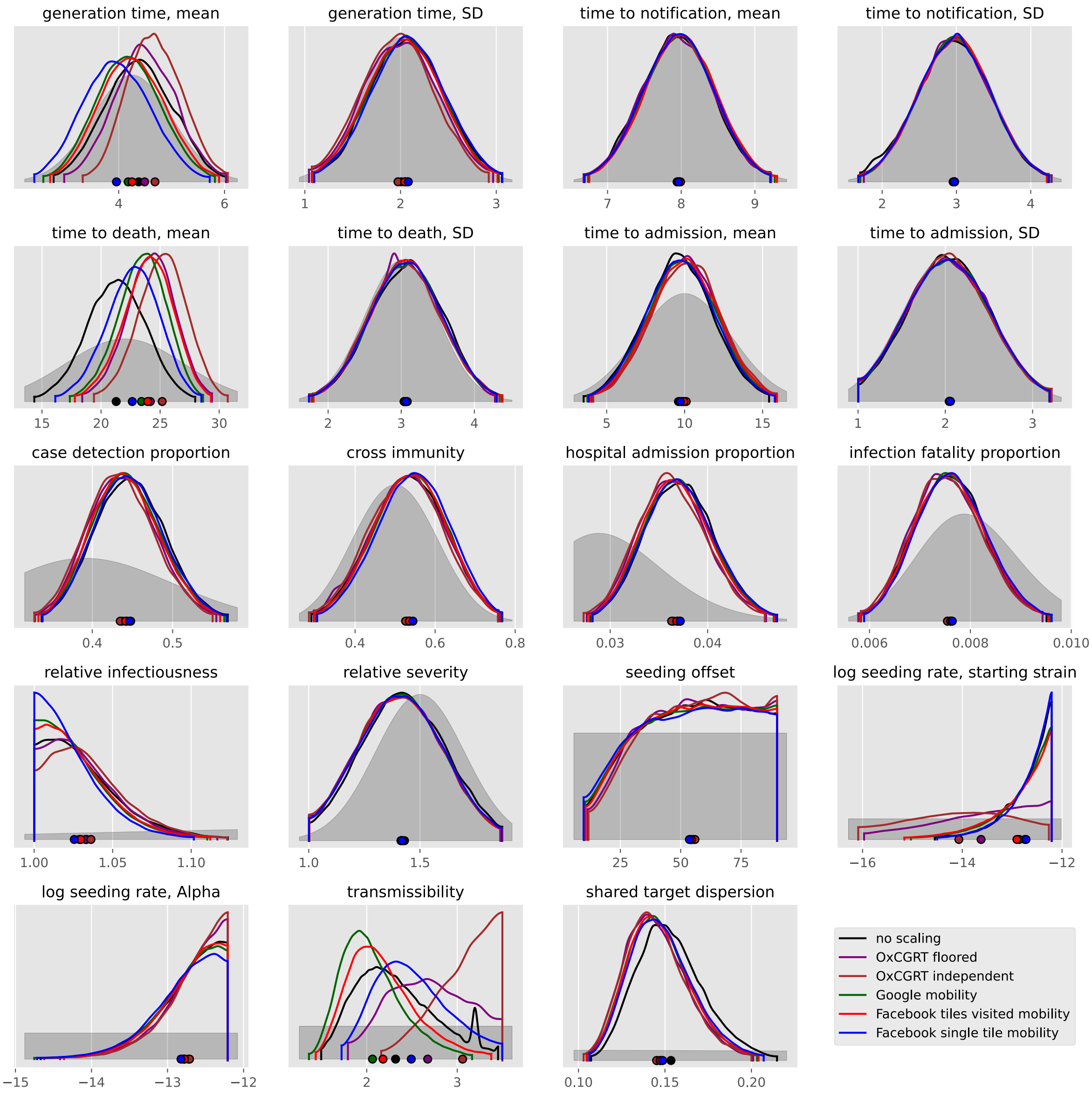

### Oceania

#### Australia

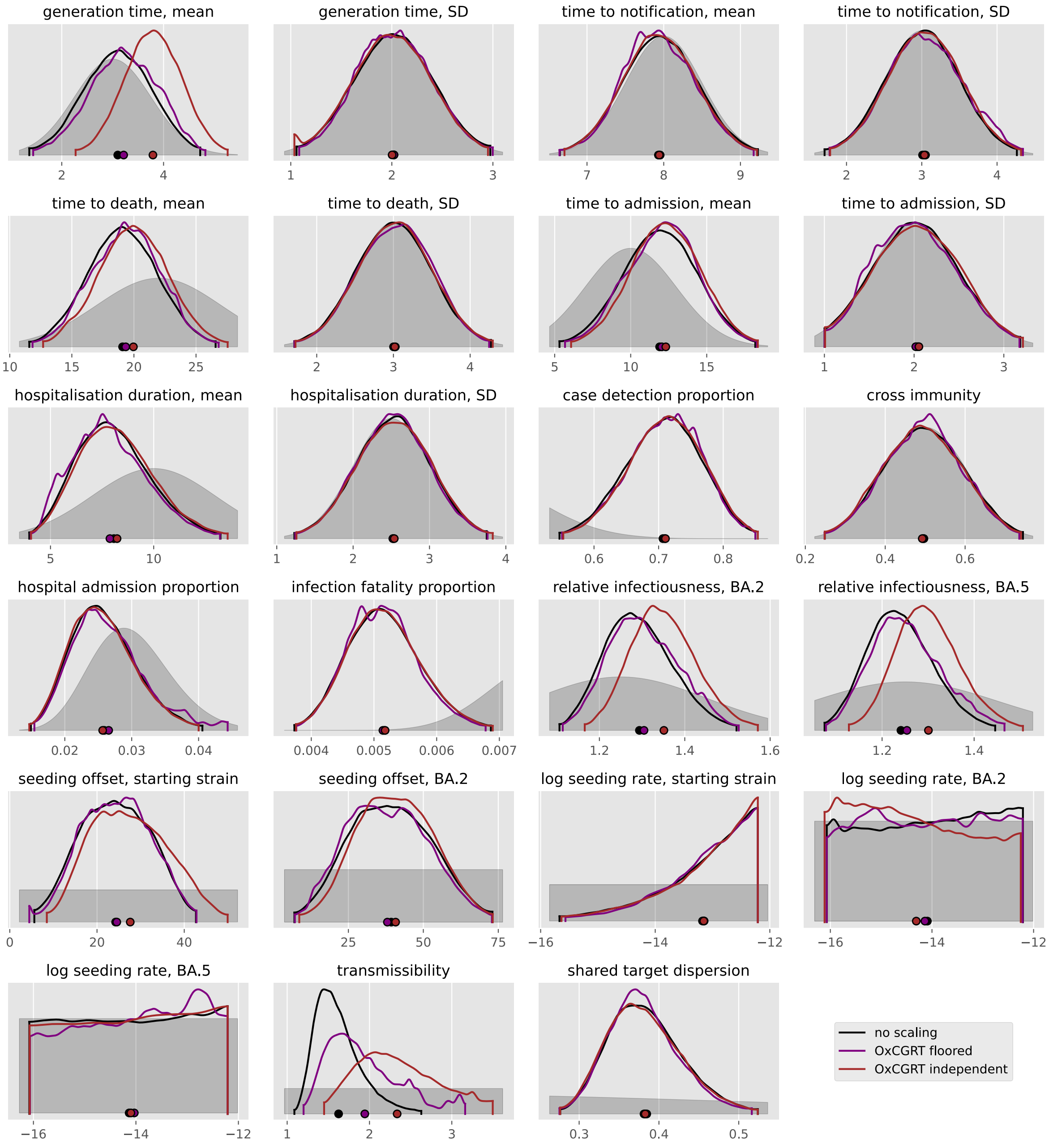

#### New Zealand

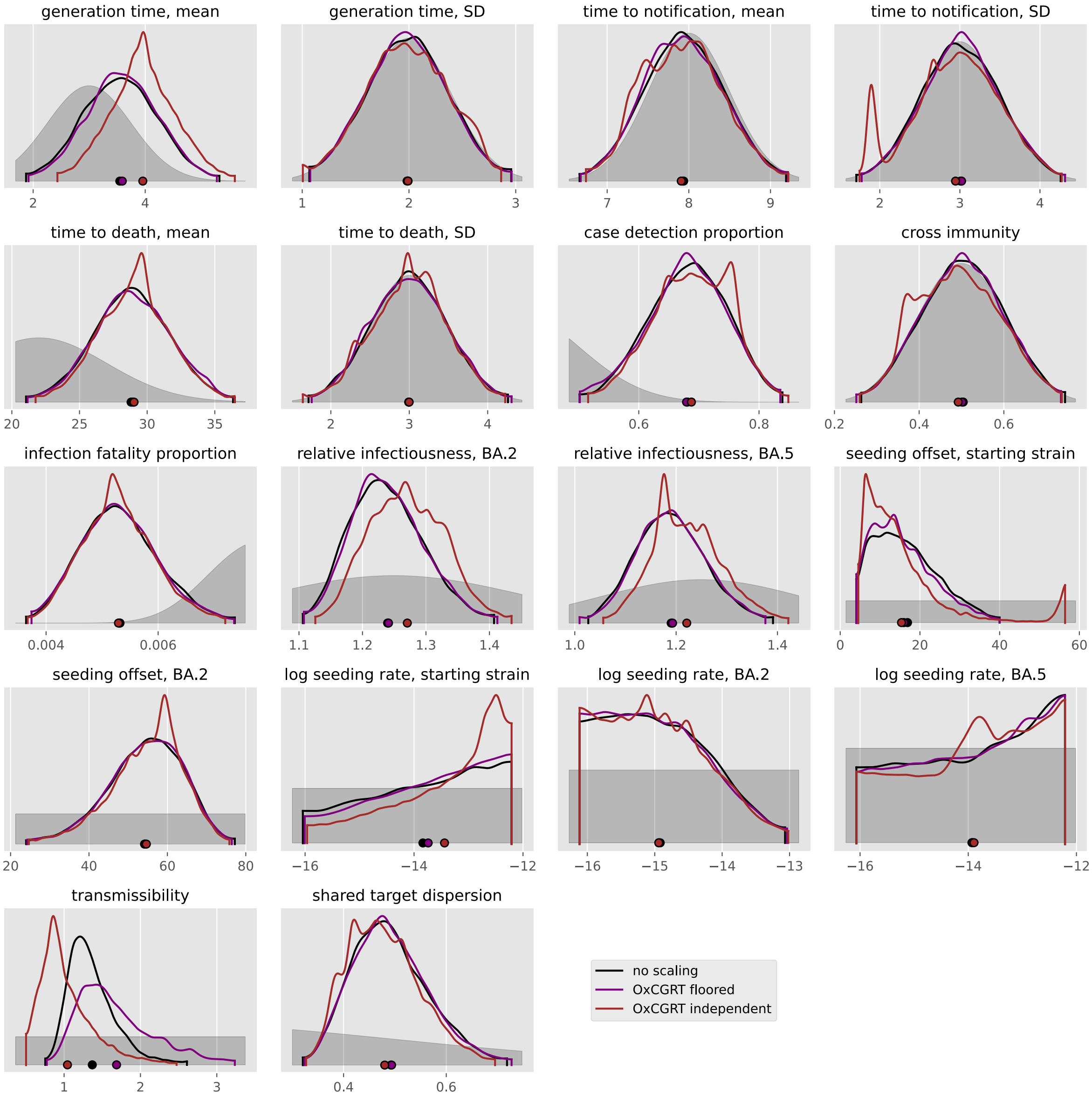

generation time, mean
generation time, SD
time to notification, mean
time to notification, SD
time to death, mean
time to death, SD
case detection proportion
cross immunity
infection fatality proportion
relative infectiousness, BA.2
relative infectiousness, BA.5
seeding offset, starting strain
seeding offset, BA.2
log seeding rate, starting strain
log seeding rate, BA.2
log seeding rate, BA.5
transmissibility
shared target dispersion
no scaling
OxCGRT floored
OxCGRT independent

#### Singapore

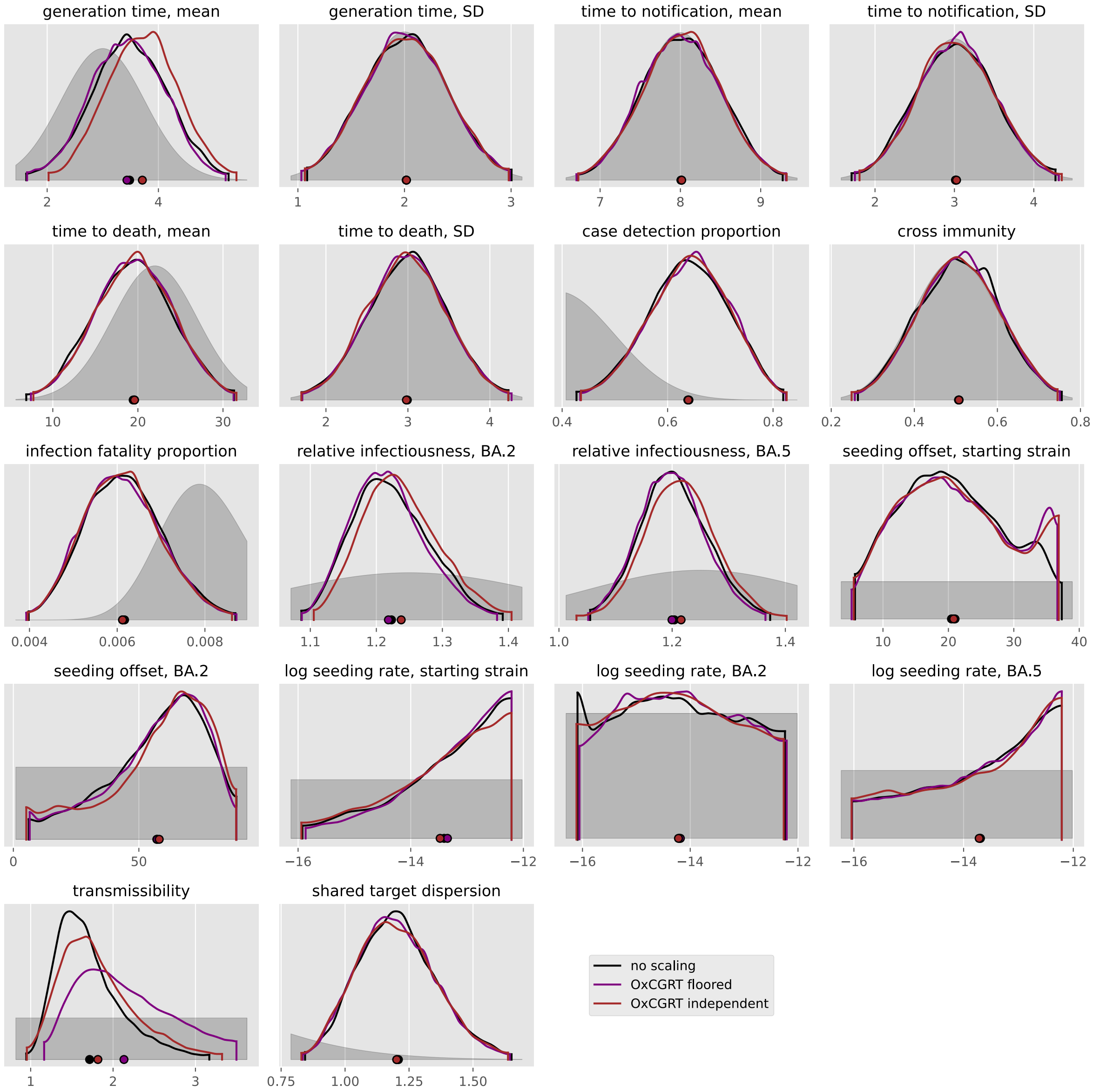
generation time, mean
generation time, SD
time to notification, mean
time to notification, SD
time to death, mean
time to death, SD
case detection proportion
cross immunity
infection fatality proportion
relative infectiousness, BA.2
relative infectiousness, BA.5
seeding offset, starting strain
seeding offset, BA.2
log seeding rate, starting strain
log seeding rate, BA.2
log seeding rate, BA.5
transmissibility
shared target dispersion
no scaling
OxCGRT floored
OxCGRT independent

### South America

#### Argentina

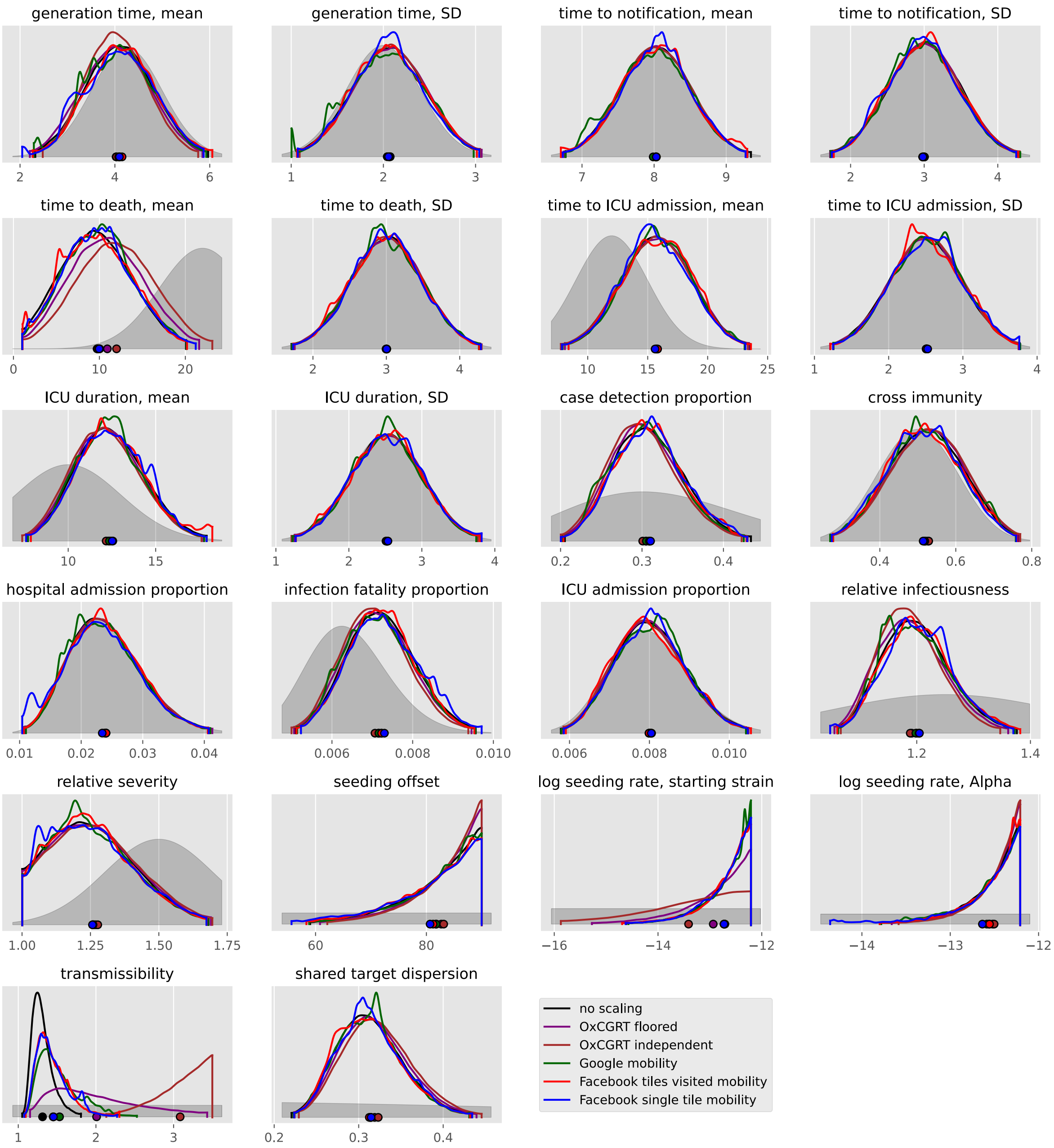

#### Brazil

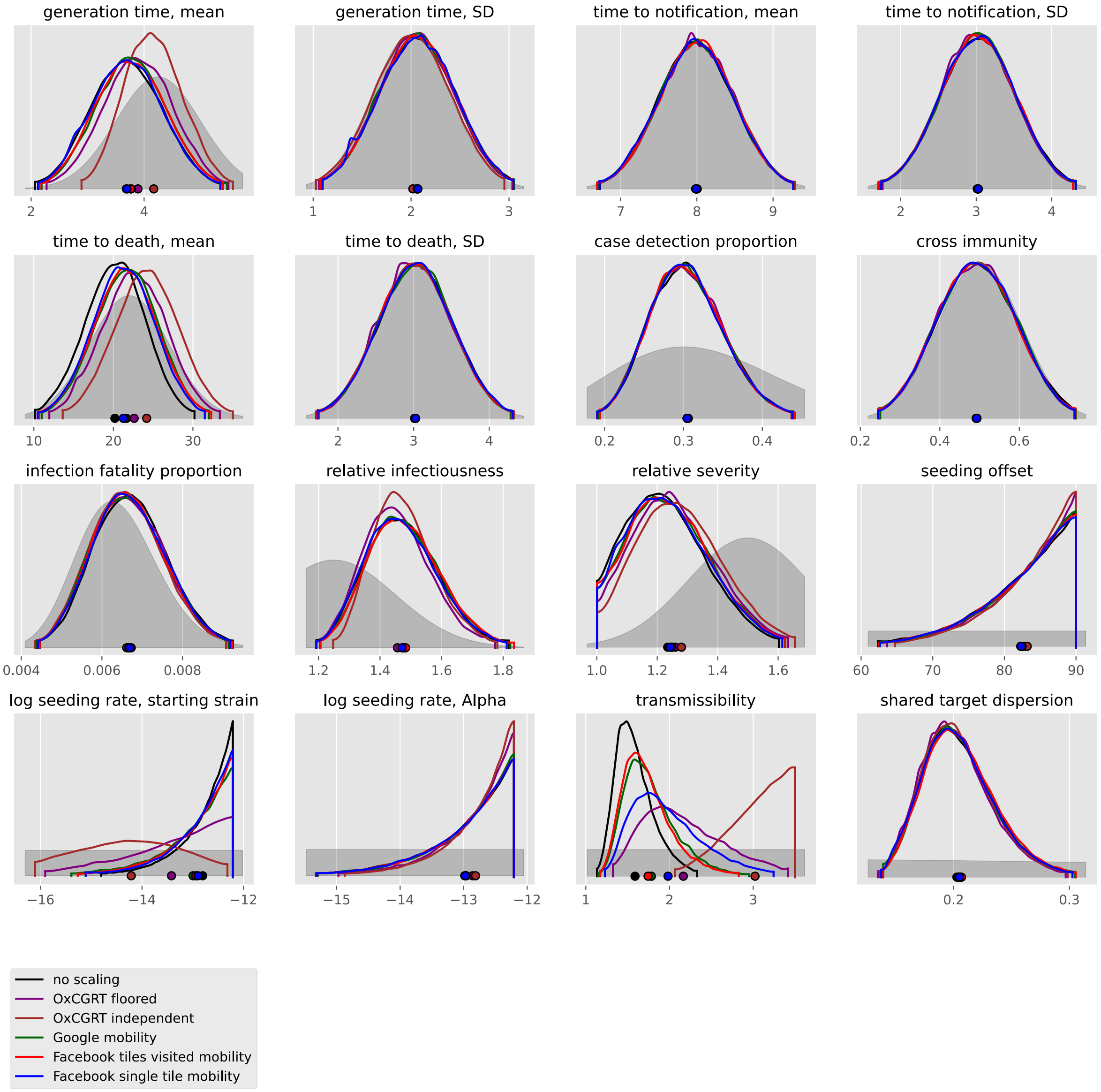
generation time, mean
generation time, SD
time to notification, mean
time to notification, SD
time to death, mean
time to death, SD
case detection proportion
cross immunity
infection fatality proportion
relative infectiousness
relative severity
seeding offset
log seeding rate, starting strain
log seeding rate, Alpha
transmissibility
shared target dispersion
no scaling
OxCGRT floored
OxCGRT independent
Google mobility
Facebook tiles visited mobility
Facebook single tile mobility

#### Chile

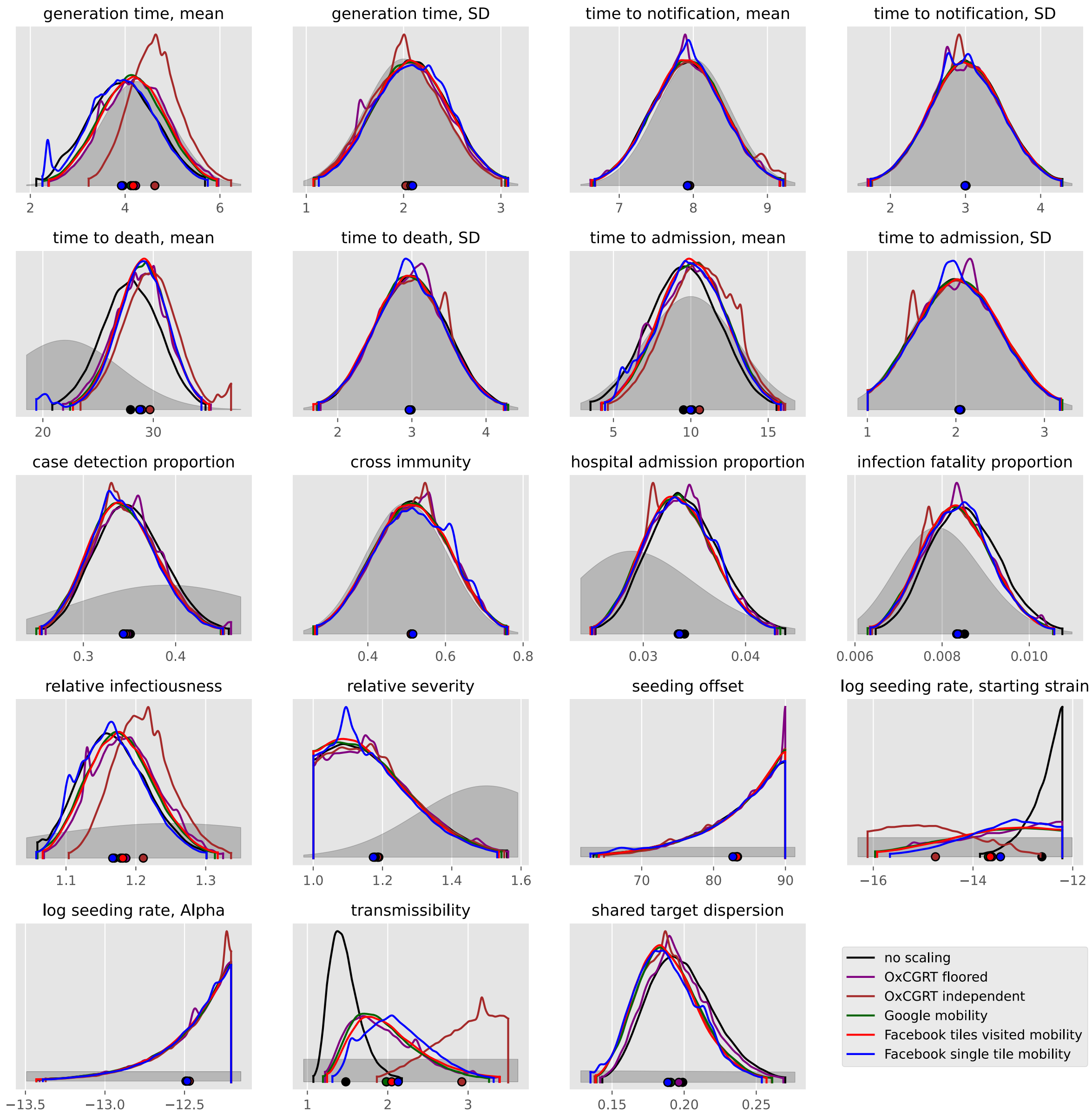
generation time, mean
generation time, SD
time to notification, mean
time to notification, SD
time to death, mean
time to death, SD
time to admission, mean
time to admission, SD
case detection proportion
cross immunity
hospital admission proportion
infection fatality proportion
relative infectiousness
relative severity
seeding offset
log seeding rate, starting strain
log seeding rate, Alpha
transmissibility
shared target dispersion
no scaling
OxCGRT floored
OxCGRT independent
Google mobility
Facebook tiles visited mobility
Facebook single tile mobility

#### Colombia

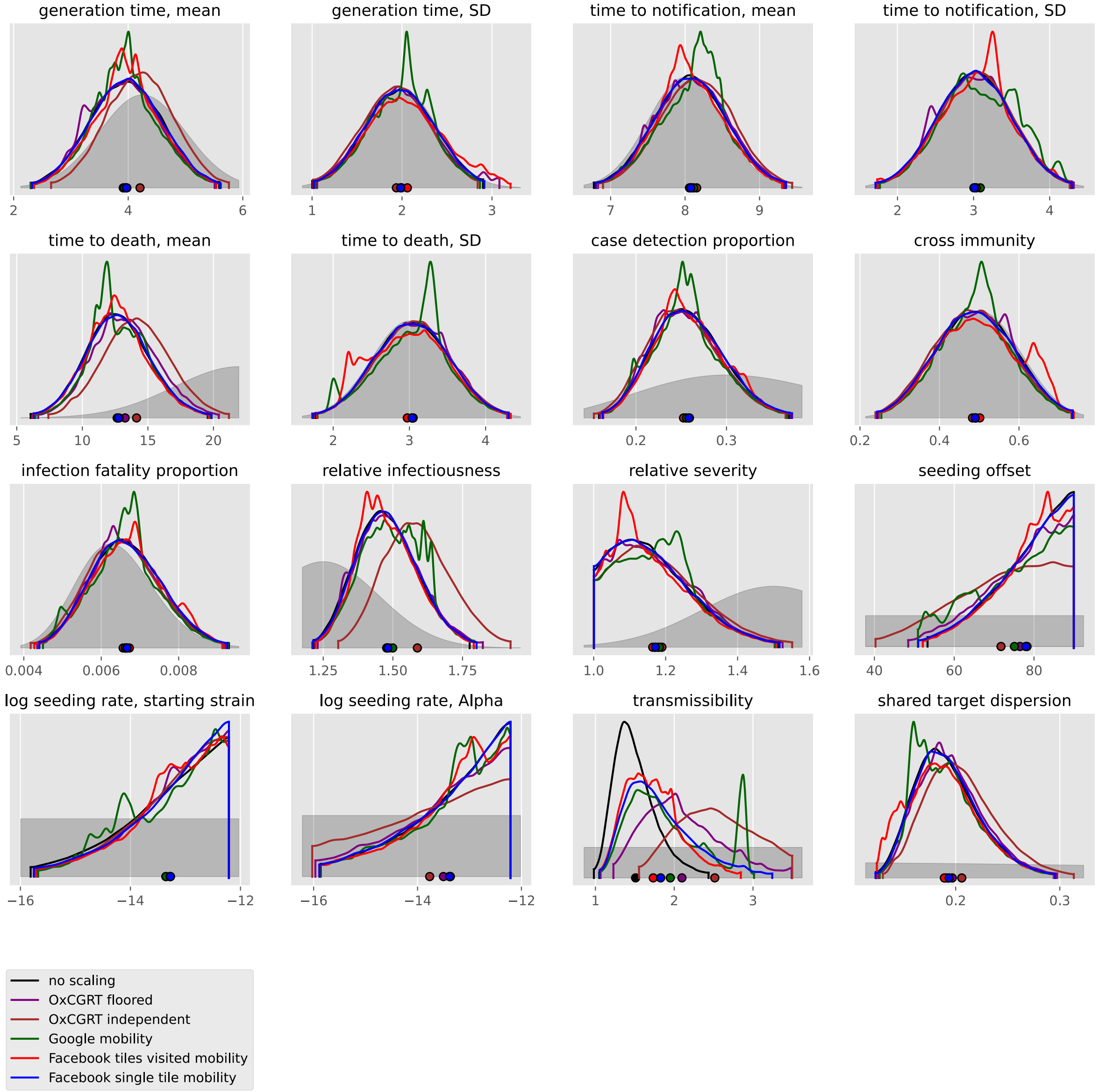

generation time, mean
generation time, SD
time to notification, mean
time to notification, SD
time to death, mean
time to death, SD
case detection proportion
cross immunity
infection fatality proportion
relative infectiousness
relative severity
seeding offset
log seeding rate, starting strain
log seeding rate, Alpha
transmissibility
shared target dispersion
no scaling
OxCGRT floored
OxCGRT independent
Google mobility
Facebook tiles visited mobility
Facebook single tile mobility

#### Guyana

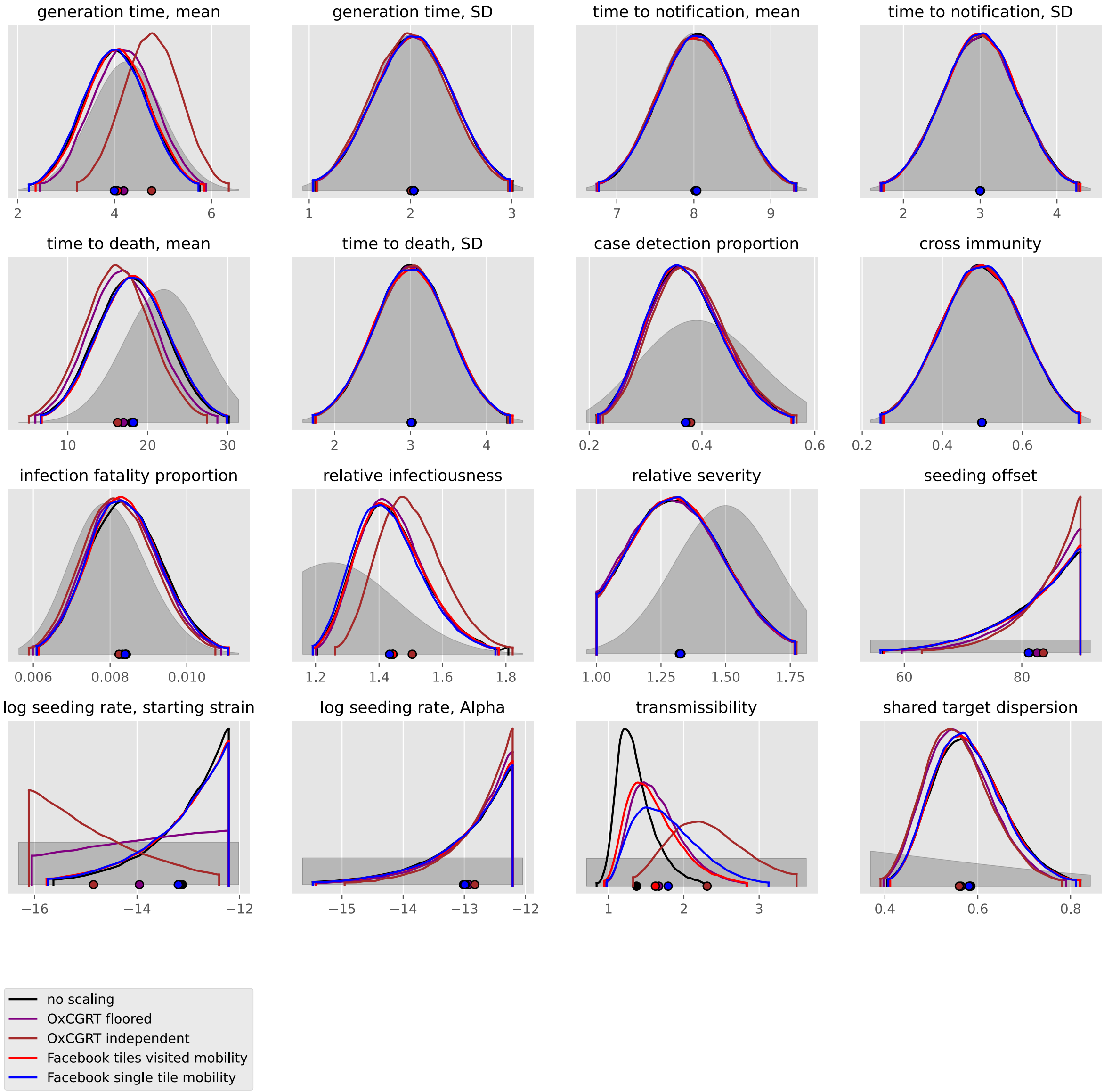

generation time, mean
2
4
6
generation time, SD
1
2
3
time to notification, mean
7
8
9
time to notification, SD
2
3
4
time to death, mean
10
20
30
time to death, SD
2
3
4
case detection proportion
0.2
0.4
0.6
cross immunity
0.2
0.4
0.6
infection fatality proportion
0.006
0.008
0.010
relative infectiousness
1.2
1.4
1.6
1.8
relative severity
1.00
1.25
1.50
1.75
seeding offset
60
80
log seeding rate, starting strain
−16
−14
−12
log seeding rate, Alpha
−15
−14
−13
−12
transmissibility
1
2
3
shared target dispersion
0.4
0.6
0.8
no scaling
OxCGRT floored
OxCGRT independent
Facebook tiles visited mobility
Facebook single tile mobility

#### Paraguay

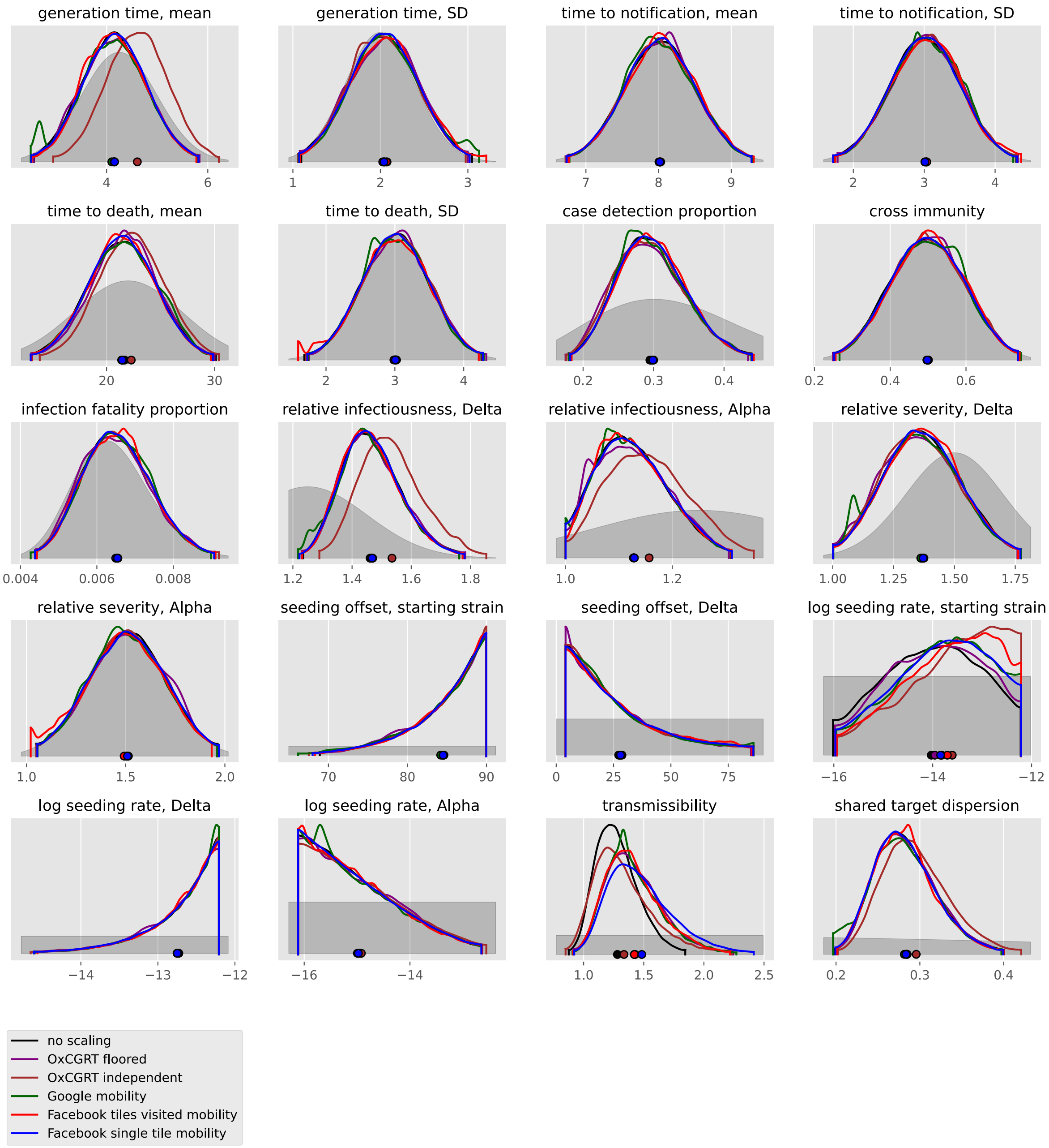

generation time, mean
generation time, SD
time to notification, mean
time to notification, SD
time to death, mean
time to death, SD
case detection proportion
cross immunity
infection fatality proportion
relative infectiousness, Delta
relative infectiousness, Alpha
relative severity, Delta
relative severity, Alpha
seeding offset, starting strain
seeding offset, Delta
log seeding rate, starting strain
log seeding rate, Delta
log seeding rate, Alpha
transmissibility
shared target dispersion
no scaling
OxCGRT floored
OxCGRT independent
Google mobility
Facebook tiles visited mobility
Facebook single tile mobility

#### Peru

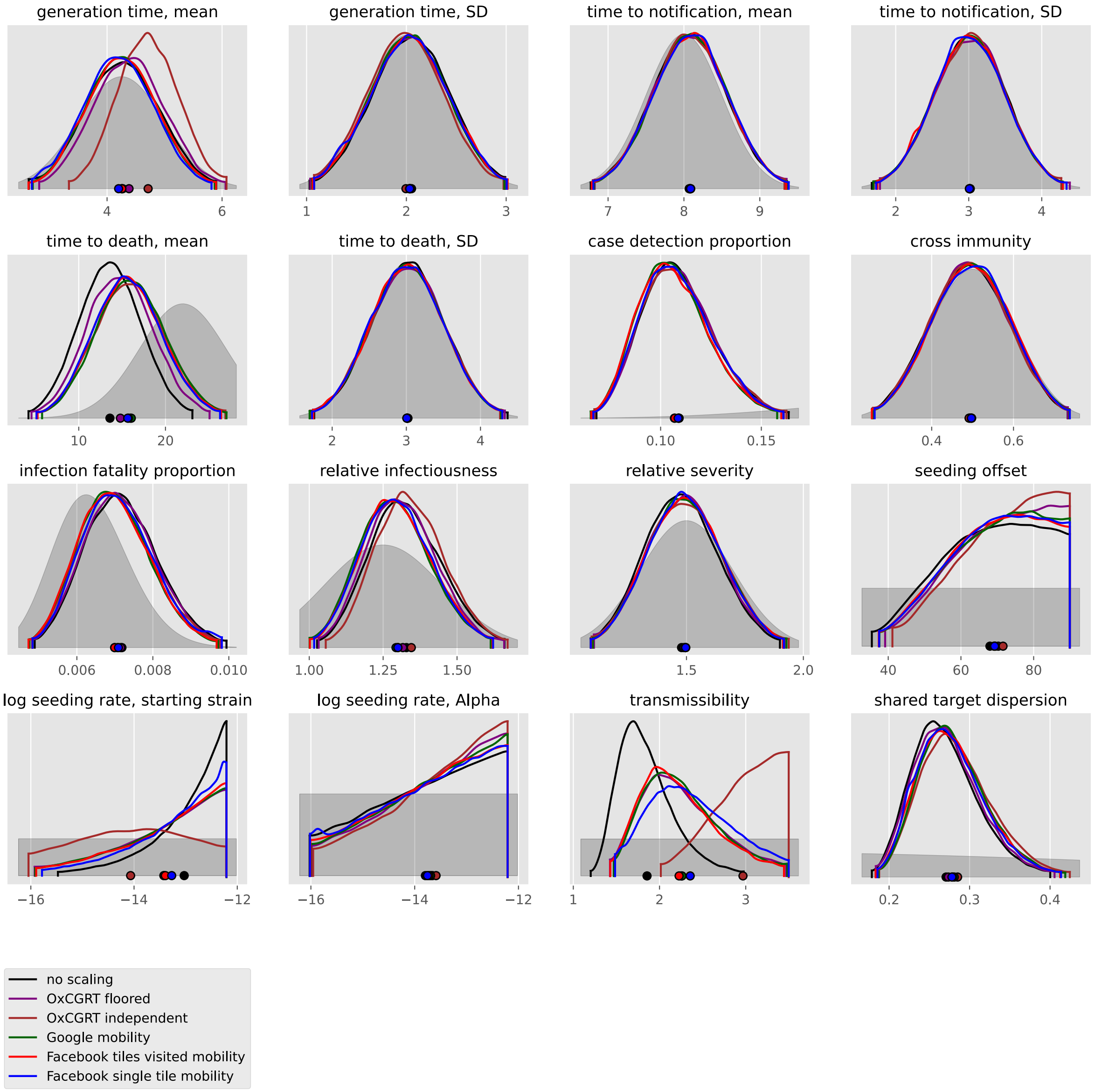
generation time, mean
generation time, SD
time to notification, mean
time to notification, SD
time to death, mean
time to death, SD
case detection proportion
cross immunity
infection fatality proportion
relative infectiousness
relative severity
seeding offset
log seeding rate, starting strain
log seeding rate, Alpha
transmissibility
shared target dispersion
no scaling
OxCGRT floored
OxCGRT independent
Google mobility
Facebook tiles visited mobility
Facebook single tile mobility

#### Suriname

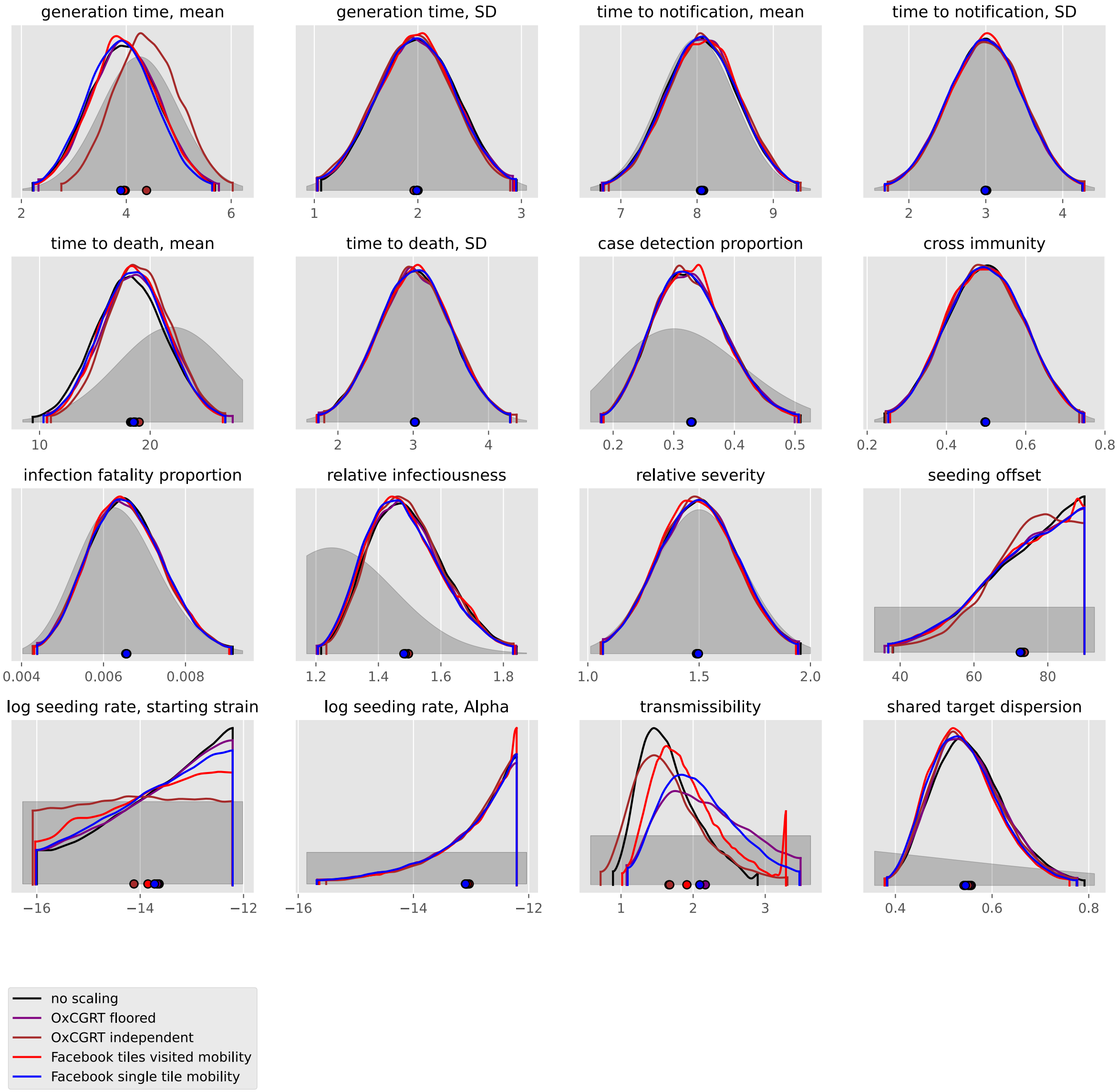

generation time, mean
generation time, SD
time to notification, mean
time to notification, SD
time to death, mean
time to death, SD
case detection proportion
cross immunity
infection fatality proportion
relative infectiousness
relative severity
seeding offset
log seeding rate, starting strain
log seeding rate, Alpha
transmissibility
shared target dispersion
no scaling
OxCGRT floored
OxCGRT independent
Facebook tiles visited mobility
Facebook single tile mobility

#### Uruguay

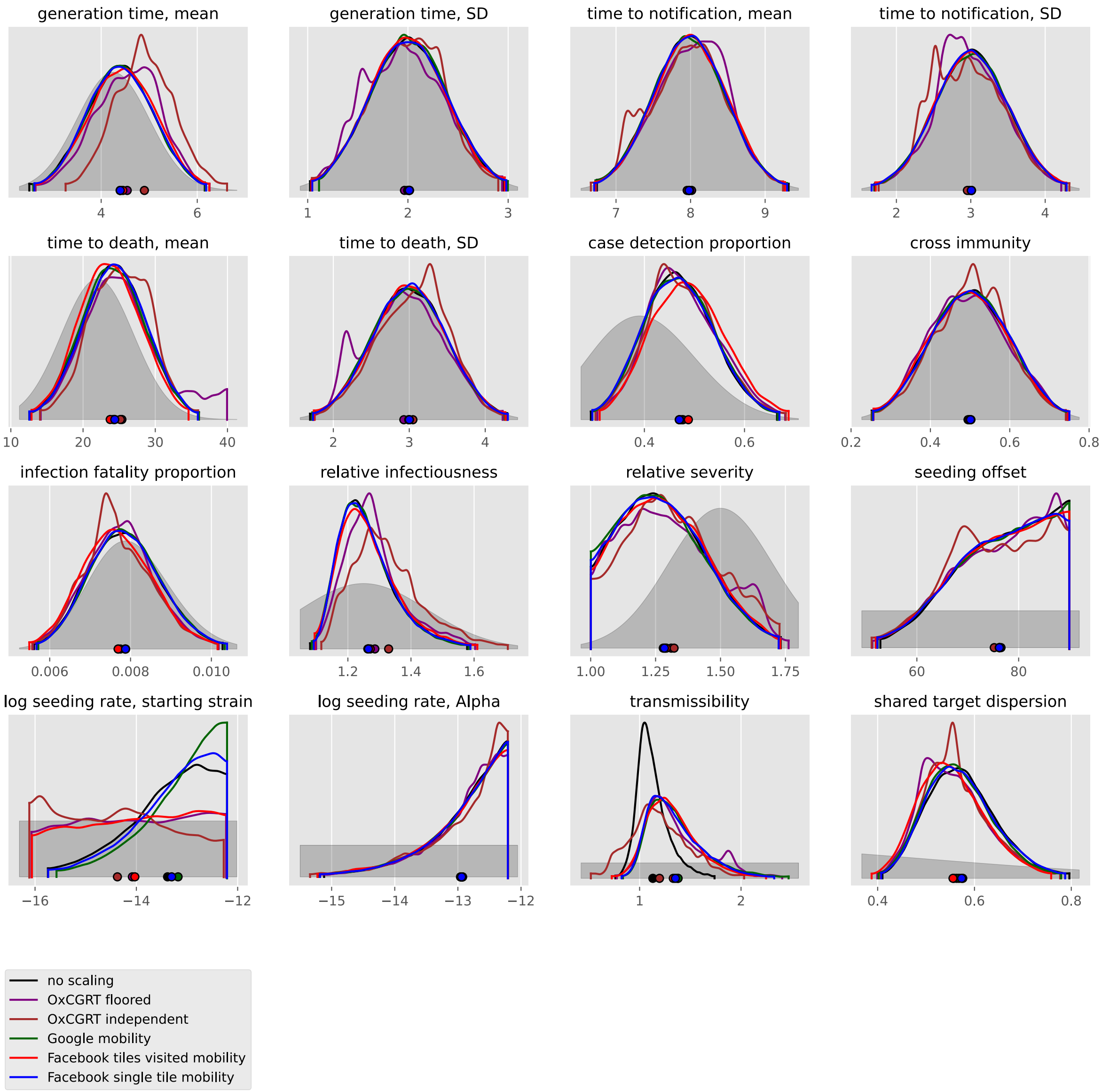

generation time, mean
generation time, SD
time to notification, mean
time to notification, SD
time to death, mean
time to death, SD
case detection proportion
cross immunity
infection fatality proportion
relative infectiousness
relative severity
seeding offset
log seeding rate, starting strain
log seeding rate, Alpha
transmissibility
shared target dispersion
no scaling
OxCGRT floored
OxCGRT independent
Google mobility
Facebook tiles visited mobility
Facebook single tile mobility

#### Venezuela, Bolivarian Republic of

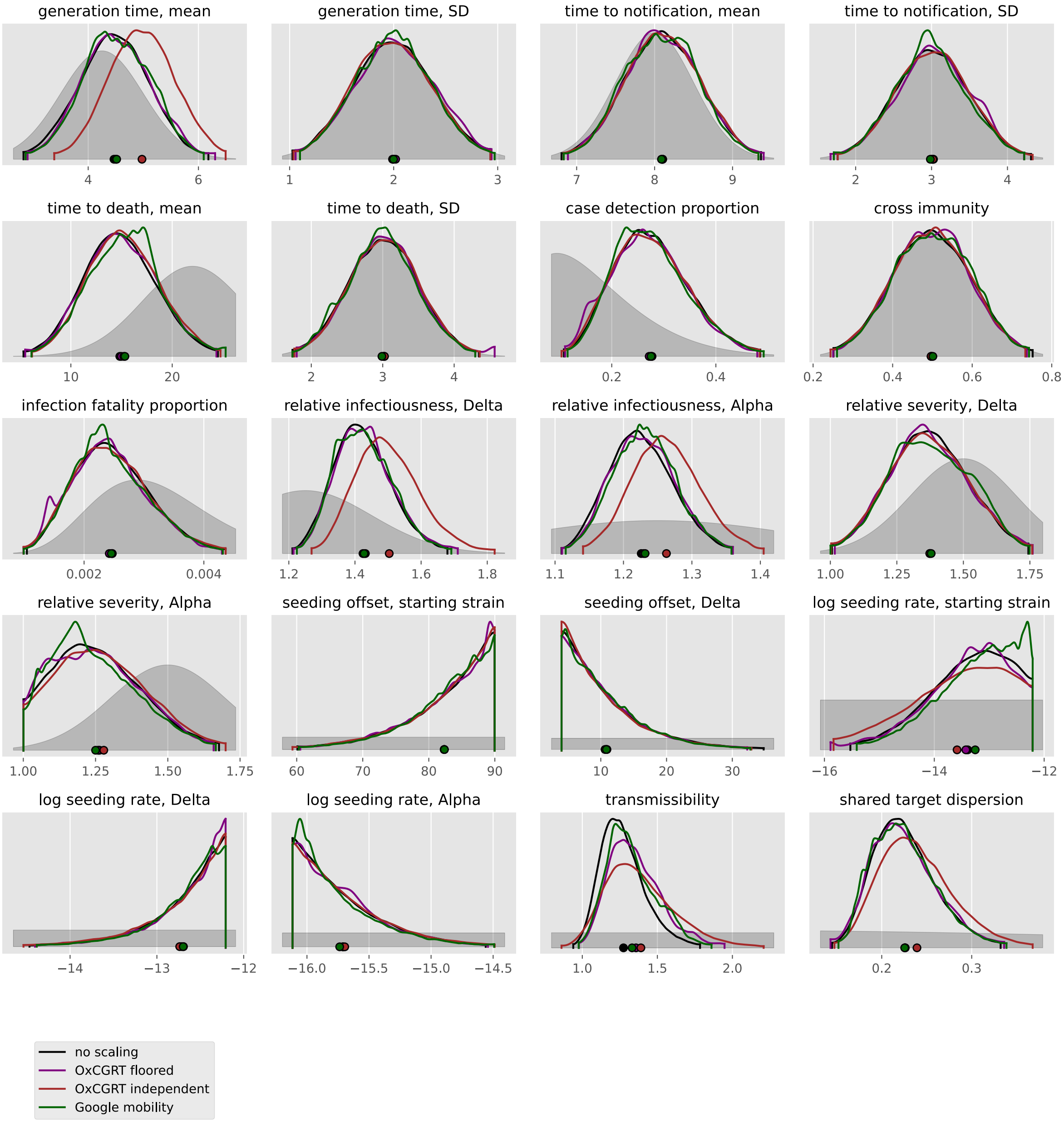

#### Commits used for analyses

For reproducibility, the following table gives the (short) commit SHA for each analysis.

| | no scaling | OxCGRT floored | OxCGRT independent | Google mobility | Facebook tiles visited mobility | Facebook single tile mobility |
|---|---|---|---|---|---|---|
| Afghanistan | 4f57571 | b934196 | b934196 | b934196 | no analysis | no analysis |
| Albania | 4f57571 | b934196 | b934196 | no analysis | b934196 | b934196 |
| Algeria | 4f57571 | b934196 | b934196 | no analysis | b934196 | b934196 |
| Andorra | 4f57571 | b934196 | b934196 | no analysis | no analysis | no analysis |
| Angola | 4f57571 | b934196 | b934196 | cb8324a | cb8324a | cb8324a |
| Argentina | 4f57571 | b934196 | b934196 | b934196 | b934196 | b934196 |
| Aruba | 4f57571 | b934196 | b934196 | 4f57571 | no analysis | no analysis |
| Australia | b934196 | b934196 | b934196 | no analysis | no analysis | no analysis |
| Austria | 4f57571 | b934196 | b934196 | b934196 | b934196 | b934196 |
| Azerbaijan | 4f57571 | b934196 | b934196 | no analysis | no analysis | no analysis |
| Bahamas | 4f57571 | b934196 | b934196 | b934196 | no analysis | no analysis |
| Bahrain | 4f57571 | b934196 | b934196 | 4f57571 | no analysis | no analysis |
| Bangladesh | 4f57571 | b934196 | b934196 | b934196 | b934196 | b934196 |
| Belarus | 4f57571 | b934196 | b934196 | cb8324a | cb8324a | b934196 |
| Belgium | 4f57571 | b934196 | b934196 | 4f57571 | 4f57571 | 4f57571 |
| Benin | 4f57571 | b934196 | b934196 | 4f57571 | 4f57571 | b934196 |
| Bosnia and Herzegovina | 4f57571 | b934196 | b934196 | b934196 | b934196 | b934196 |
| Brazil | 4f57571 | b934196 | b934196 | 4f57571 | 4f57571 | 4f57571 |
| Bulgaria | 4f57571 | b934196 | b934196 | b934196 | b934196 | b934196 |
| Burkina Faso | 4f57571 | b934196 | b934196 | 4f57571 | 4f57571 | 4f57571 |
| Cabo Verde | 4f57571 | b934196 | b934196 | 4f57571 | no analysis | no analysis |
| Cameroon | 4f57571 | b934196 | b934196 | b934196 | b934196 | b934196 |
| Canada | 4f57571 | b934196 | b934196 | b934196 | b934196 | b934196 |
| Chile | 4f57571 | b934196 | b934196 | 4f57571 | b934196 | b934196 |
| Colombia | 4f57571 | b934196 | b934196 | 4f57571 | 4f57571 | b934196 |
| Congo | 4f57571 | b934196 | b934196 | no analysis | 4f57571 | b934196 |
| Costa Rica | 4f57571 | b934196 | b934196 | 4f57571 | b934196 | b934196 |
| Croatia | 4f57571 | b934196 | b934196 | b934196 | b934196 | b934196 |

| | no scaling | OxCGRT floored | OxCGRT independent | Google mobility | Facebook tiles visited mobility | Facebook single tile mobility |
|---|---|---|---|---|---|---|
| Cuba | 4f57571 | b934196 | b934196 | no analysis | no analysis | no analysis |
| Cyprus | 4f57571 | b934196 | b934196 | no analysis | no analysis | no analysis |
| Czechia | 4f57571 | b934196 | b934196 | 4f57571 | b934196 | b934196 |
| Côte d'Ivoire | 4f57571 | b934196 | b934196 | b934196 | b934196 | b934196 |
| Denmark | 4f57571 | b934196 | b934196 | 4f57571 | 4f57571 | 4f57571 |
| Dominican Republic | 4f57571 | b934196 | b934196 | 4f57571 | b934196 | b934196 |
| Egypt | 4f57571 | b934196 | b934196 | b934196 | b934196 | b934196 |
| El Salvador | 4f57571 | 4c513a7 | 4c513a7 | 4f57571 | 4f57571 | 4f57571 |
| Estonia | 4f57571 | b934196 | b934196 | 4f57571 | 4f57571 | 4f57571 |
| Eswatini | 4f57571 | 4c513a7 | missing git information | no analysis | no analysis | no analysis |
| Ethiopia | 4f57571 | b934196 | b934196 | no analysis | b934196 | b934196 |
| Finland | 4f57571 | b934196 | b934196 | b934196 | b934196 | b934196 |
| France | 4f57571 | b934196 | b934196 | b934196 | b934196 | b934196 |
| Gabon | 4f57571 | b934196 | b934196 | b934196 | b934196 | b934196 |
| Georgia | 4f57571 | b934196 | b934196 | 4f57571 | b934196 | b934196 |
| Germany | 4f57571 | b934196 | b934196 | b934196 | b934196 | b934196 |
| Greece | 4f57571 | b934196 | b934196 | 4f57571 | 4f57571 | b934196 |
| Guinea | 4f57571 | 4c513a7 | b934196 | no analysis | b934196 | b934196 |
| Guyana | 4f57571 | b934196 | b934196 | no analysis | 4f57571 | 4f57571 |
| Haiti | 4f57571 | b934196 | 4c513a7 | b934196 | b934196 | b934196 |
| Honduras | 4f57571 | b934196 | b934196 | b934196 | b934196 | b934196 |
| Hungary | 4f57571 | b934196 | b934196 | 4f57571 | 4f57571 | b934196 |
| India | 4f57571 | b934196 | b934196 | b934196 | b934196 | b934196 |
| Indonesia | 4f57571 | b934196 | b934196 | b934196 | b934196 | b934196 |
| Iran, Islamic Republic of | 4f57571 | b934196 | b934196 | no analysis | no analysis | no analysis |
| Iraq | 4f57571 | b934196 | b934196 | 4f57571 | 4f57571 | 4f57571 |
| Ireland | 4f57571 | b934196 | b934196 | b934196 | b934196 | b934196 |
| Israel | 4f57571 | b934196 | b934196 | b934196 | no analysis | no analysis |
| Italy | 4f57571 | b934196 | b934196 | 4f57571 | 4f57571 | b934196 |
| Jamaica | 4f57571 | b934196 | cb8324a | b934196 | b934196 | b934196 |

| | no scaling | OxCGRT floored | OxCGRT independent | Google mobility | Facebook tiles visited mobility | Facebook single tile mobility |
|---|---|---|---|---|---|---|
| Japan | cb8324a | cb8324a | cb8324a | cb8324a | b934196 | cb8324a |
| Jordan | 4f57571 | b934196 | b934196 | 4f57571 | 4f57571 | 4f57571 |
| Kenya | 4f57571 | b934196 | b934196 | b934196 | b934196 | b934196 |
| Korea, Republic of | 4f57571 | b934196 | cb8324a | b934196 | b934196 | cb8324a |
| Kosovo | 4f57571 | 4c513a7 | 4c513a7 | no analysis | no analysis | no analysis |
| Kuwait | 4f57571 | 4c513a7 | 4c513a7 | 4f57571 | no analysis | no analysis |
| Latvia | 4f57571 | 4c513a7 | 4c513a7 | 4f57571 | 4f57571 | 4f57571 |
| Lebanon | 4f57571 | 4c513a7 | b934196 | 4f57571 | 4f57571 | 4f57571 |
| Lesotho | 4f57571 | b934196 | 4c513a7 | no analysis | 4f57571 | 4f57571 |
| Liberia | 4f57571 | 4c513a7 | 4c513a7 | no analysis | 4f57571 | 4f57571 |
| Libya | cb8324a | b934196 | cb8324a | cb8324a | no analysis | no analysis |
| Lithuania | 4f57571 | 4c513a7 | 4c513a7 | 4f57571 | 4f57571 | 4f57571 |
| Luxembourg | 4f57571 | 4c513a7 | 4c513a7 | 4f57571 | cb8324a | cb8324a |
| Madagascar | 4f57571 | 4c513a7 | 4c513a7 | no analysis | 4f57571 | cb8324a |
| Malawi | 4f57571 | 4c513a7 | cb8324a | no analysis | cb8324a | cb8324a |
| Malaysia | 4f57571 | 1cab74a | 4c513a7 | 4f57571 | 4f57571 | 4f57571 |
| Mali | 4f57571 | 4c513a7 | 4c513a7 | 4f57571 | 4f57571 | 4f57571 |
| Malta | cb8324a | cb8324a | cb8324a | cb8324a | cb8324a | cb8324a |
| Mexico | 4f57571 | 4c513a7 | 4c513a7 | 4f57571 | 4f57571 | 4f57571 |
| Moldova, Republic of | 4f57571 | 4c513a7 | 4c513a7 | cb8324a | cb8324a | cb8324a |
| Morocco | 4f57571 | 4c513a7 | 4c513a7 | 4f57571 | no analysis | no analysis |
| Mozambique | 4f57571 | 4c513a7 | 4c513a7 | 4f57571 | cb8324a | cb8324a |
| Nepal | 4f57571 | 4c513a7 | 4c513a7 | cb8324a | cb8324a | cb8324a |
| Netherlands | 4f57571 | 4c513a7 | 4c513a7 | 4f57571 | cb8324a | cb8324a |
| New Zealand | 4c513a7 | 4c513a7 | 4c513a7 | no analysis | no analysis | no analysis |
| Nigeria | 4f57571 | 4c513a7 | 4c513a7 | 4f57571 | 4f57571 | 4f57571 |
| Norway | 4f57571 | 4c513a7 | 4c513a7 | cb8324a | cb8324a | cb8324a |
| Oman | 4f57571 | 4c513a7 | 4c513a7 | 4f57571 | 4f57571 | 4f57571 |
| Pakistan | 4f57571 | 4c513a7 | cb8324a | cb8324a | no analysis | no analysis |
| Palestine, State of | cb8324a | cb8324a | cb8324a | no analysis | no analysis | no analysis |

| | no scaling | OxCGRT floored | OxCGRT independent | Google mobility | Facebook tiles visited mobility | Facebook single tile mobility |
|---|---|---|---|---|---|---|
| Panama | 4f57571 | 4c513a7 | 4c513a7 | 4f57571 | cb8324a | cb8324a |
| Paraguay | 4f57571 | 4c513a7 | cb8324a | cb8324a | cb8324a | cb8324a |
| Peru | 4f57571 | 4c513a7 | 4c513a7 | 4f57571 | 4f57571 | 4f57571 |
| Philippines | 4f57571 | 4c513a7 | cb8324a | cb8324a | cb8324a | cb8324a |
| Poland | 4f57571 | 4c513a7 | 4c513a7 | 4f57571 | 4f57571 | cb8324a |
| Portugal | 4f57571 | 4c513a7 | 4c513a7 | cb8324a | cb8324a | cb8324a |
| Qatar | 4f57571 | 4c513a7 | 4c513a7 | 4f57571 | 4f57571 | 4f57571 |
| Romania | 4f57571 | 4c513a7 | 4c513a7 | 4f57571 | cb8324a | cb8324a |
| Russian Federation | 4f57571 | 4c513a7 | cb8324a | cb8324a | no analysis | no analysis |
| Rwanda | 4f57571 | 4c513a7 | 4c513a7 | 4f57571 | cb8324a | cb8324a |
| Saudi Arabia | 4f57571 | 4c513a7 | 4c513a7 | 4f57571 | 4f57571 | cb8324a |
| Senegal | 4f57571 | 4c513a7 | 4c513a7 | 4f57571 | 4f57571 | 4f57571 |
| Serbia | 4f57571 | 4c513a7 | 4c513a7 | cb8324a | cb8324a | cb8324a |
| Singapore | 4c513a7 | cb8324a | 4c513a7 | no analysis | no analysis | no analysis |
| Slovakia | cb8324a | cb8324a | cb8324a | cb8324a | cb8324a | cb8324a |
| Slovenia | cb8324a | cb8324a | cb8324a | cb8324a | cb8324a | cb8324a |
| Somalia | 4f57571 | 4c513a7 | 4c513a7 | no analysis | no analysis | no analysis |
| South Africa | 4f57571 | 4c513a7 | 4c513a7 | 4f57571 | 4f57571 | cb8324a |
| South Sudan | 4f57571 | cb8324a | cb8324a | no analysis | no analysis | no analysis |
| Spain | cb8324a | b934196 | b934196 | b934196 | b934196 | b934196 |
| Sri Lanka | cb8324a | cb8324a | cb8324a | cb8324a | cb8324a | b934196 |
| Sudan | 4f57571 | 4c513a7 | 4c513a7 | no analysis | no analysis | no analysis |
| Suriname | 4f57571 | 4c513a7 | 4c513a7 | no analysis | 4f57571 | 4f57571 |
| Sweden | 4f57571 | 4c513a7 | 4c513a7 | 4f57571 | 4f57571 | 4f57571 |
| Switzerland | 4f57571 | b934196 | b934196 | 4f57571 | b934196 | b934196 |
| Syrian Arab Republic | 4f57571 | cb8324a | cb8324a | no analysis | no analysis | no analysis |
| Togo | 4f57571 | 4c513a7 | 4c513a7 | 4f57571 | 4f57571 | 4f57571 |
| Tunisia | 4f57571 | 4c513a7 | 4c513a7 | no analysis | 4f57571 | 4f57571 |
| Türkiye | cb8324a | cb8324a | cb8324a | cb8324a | cb8324a | cb8324a |

| | no scaling | OxCGRT floored | OxCGRT independent | Google mobility | Facebook tiles visited mobility | Facebook single tile mobility |
|---|---|---|---|---|---|---|
| Ukraine | 4f57571 | missing git information | 4c513a7 | 4f57571 | no analysis | no analysis |
| United Arab Emirates | 4f57571 | b934196 | b934196 | 4f57571 | 4f57571 | 4f57571 |
| United Kingdom | 4f57571 | b934196 | b934196 | 4f57571 | b934196 | b934196 |
| United States | 4f57571 | 4c513a7 | 4c513a7 | cb8324a | cb8324a | cb8324a |
| Uruguay | 4f57571 | 4c513a7 | 4c513a7 | 4f57571 | 4c513a7 | 4f57571 |
| Venezuela, Bolivarian Republic of | 4f57571 | 4c513a7 | cb8324a | cb8324a | no analysis | no analysis |
| Yemen | 4f57571 | 4c513a7 | 4c513a7 | 4f57571 | no analysis | no analysis |
| Zambia | cb8324a | cb8324a | cb8324a | cb8324a | cb8324a | cb8324a |
| Zimbabwe | 4f57571 | 4c513a7 | 4c513a7 | 4f57571 | cb8324a | cb8324a |
