## Supplemental File 4 for "Quantifying the contribution of policy measures to COVID-19 dynamics across 126 countries"

### Comparison between baseline residual transmission scaling and OxCGRT policy flat averages

#### Purpose

This document presents the residual transmission scaling (with associated uncertainty) implemented under the baseline analysis (without scaling according to policy data for each country analysed). It is intended to provide a comparison between the residual non-mechanistic transmission that needed to be applied for each analysis in the absence of scaling and the OxCGRT time series data, with each included policy domain weighted equally, with various combinations of policies included in the overall metrics. That is, this gives a naive comparison of the estimated fluctuation in transmission not captured through the mechanistic model and the raw OxCGRT data.

#### Included policies

The table below lists the OxCGRT indicators included in each composite policy map.

| indicator | policy | custom | stringency | containment |
|---|---|---|---|---|
| C1 | school closing | yes | yes | yes |
| C2 | workplace closing | yes | yes | yes |
| C3 | cancel public events | yes | yes | yes |
| C4 | gathering restrictions | yes | yes | yes |
| C5 | close public transport | yes | yes | yes |
| C6 | stay at home | yes | yes | yes |
| C7 | movement restrictions | yes | yes | yes |
| C8 | international travel | | yes | yes |
| H1 | public information | | yes | yes |
| H2 | testing policy | | | yes |
| H3 | contact tracing | | | yes |
| H6 | face coverings | yes | | yes |
| H7 | vaccination policy | | | yes |
| H8 | elderly protection | | | yes |

#### Policy profile comparisons

#### Africa

Estimated transmission scaling (without scaling) versus policy scaling

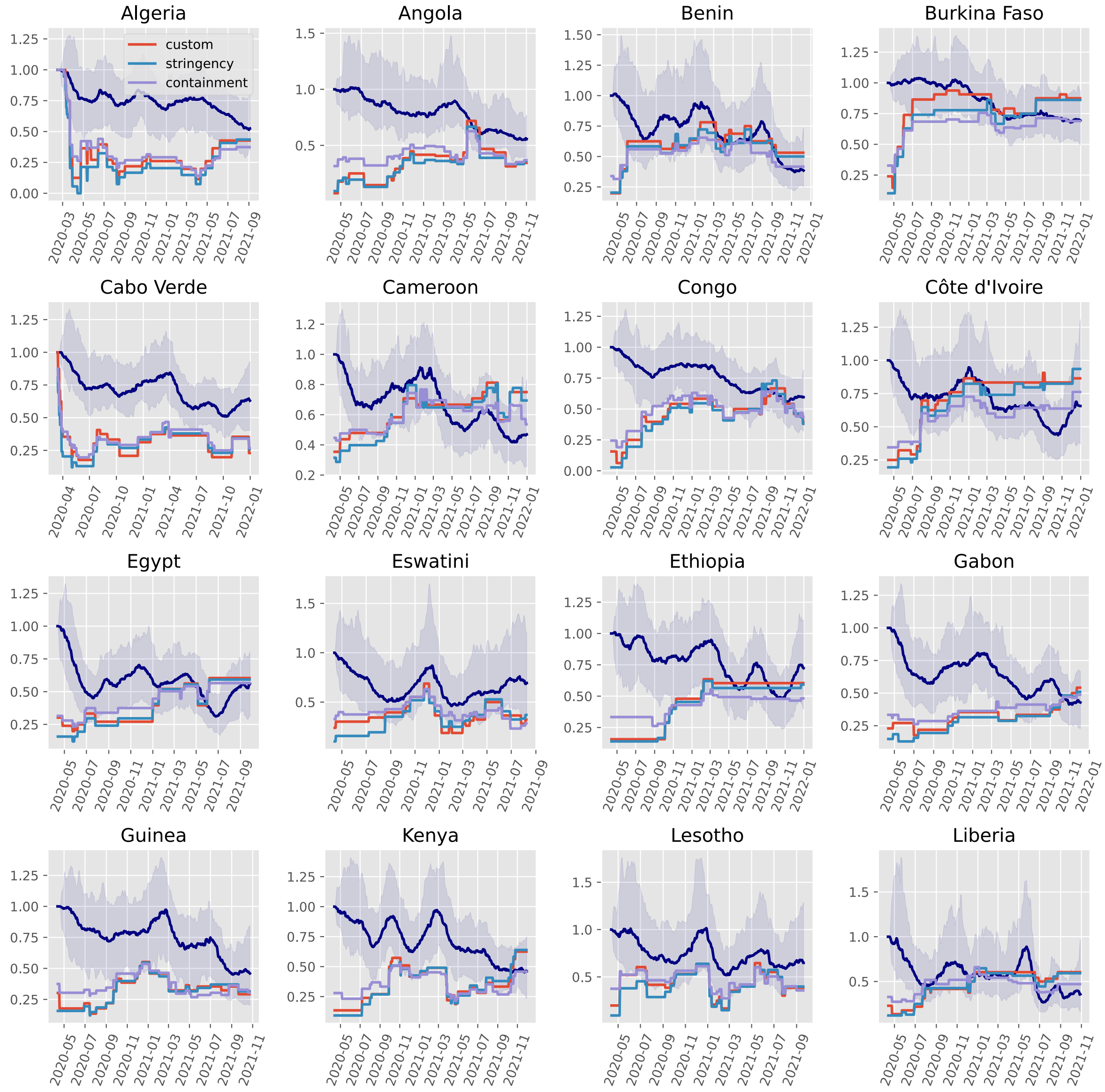

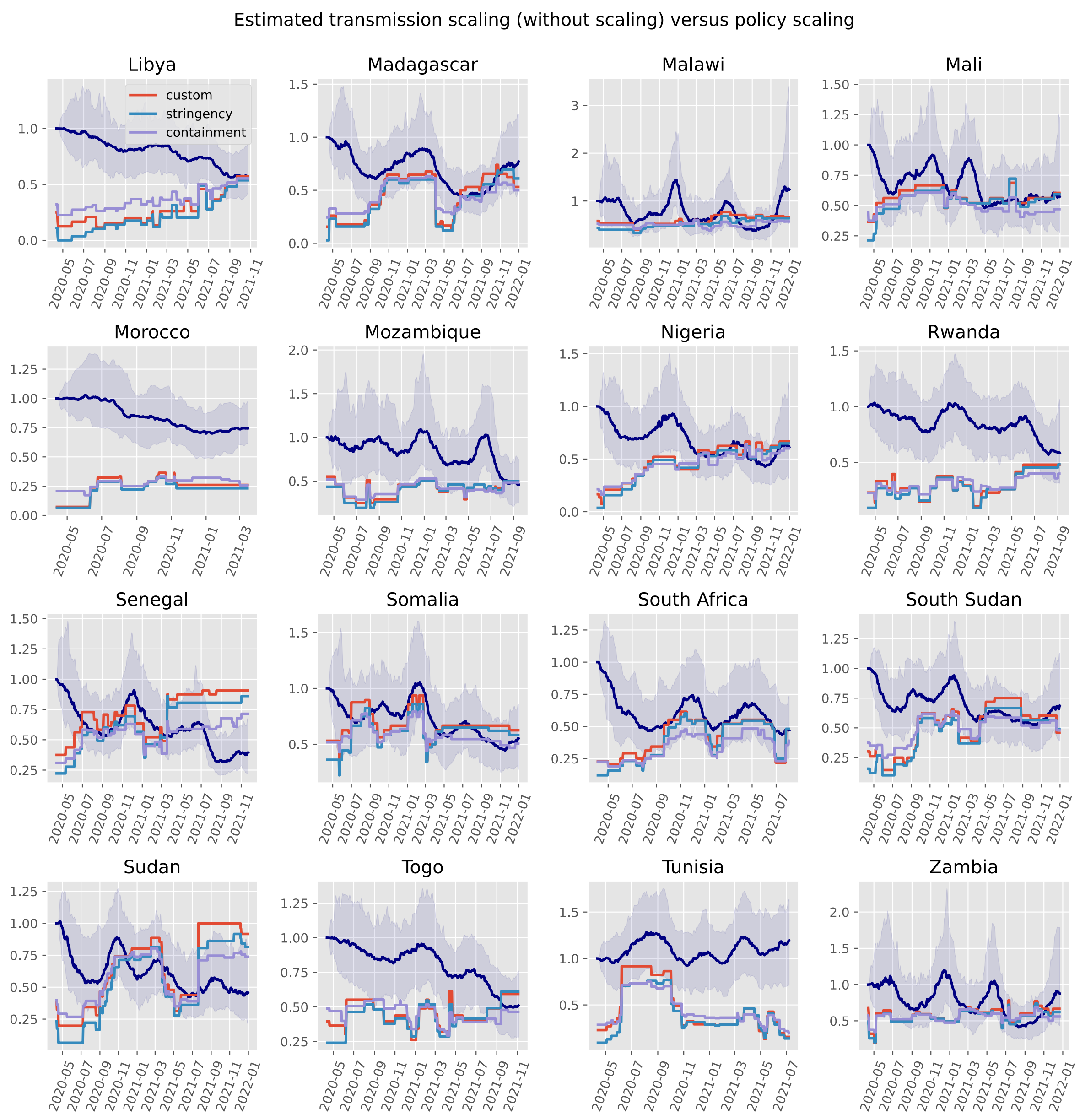
Estimated transmission scaling (without scaling) versus policy scaling
Libya
custom
stringency
containment
Madagascar
Malawi
Mali
Morocco
Mozambique
Nigeria
Rwanda
Senegal
Somalia
South Africa
South Sudan
Sudan
Togo
Tunisia
Zambia

Estimated transmission scaling (without scaling) versus policy scaling

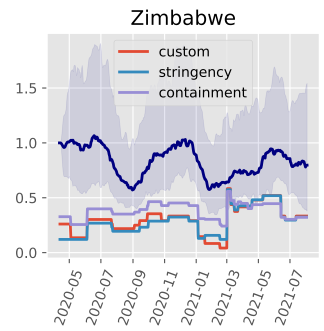

#### Asia

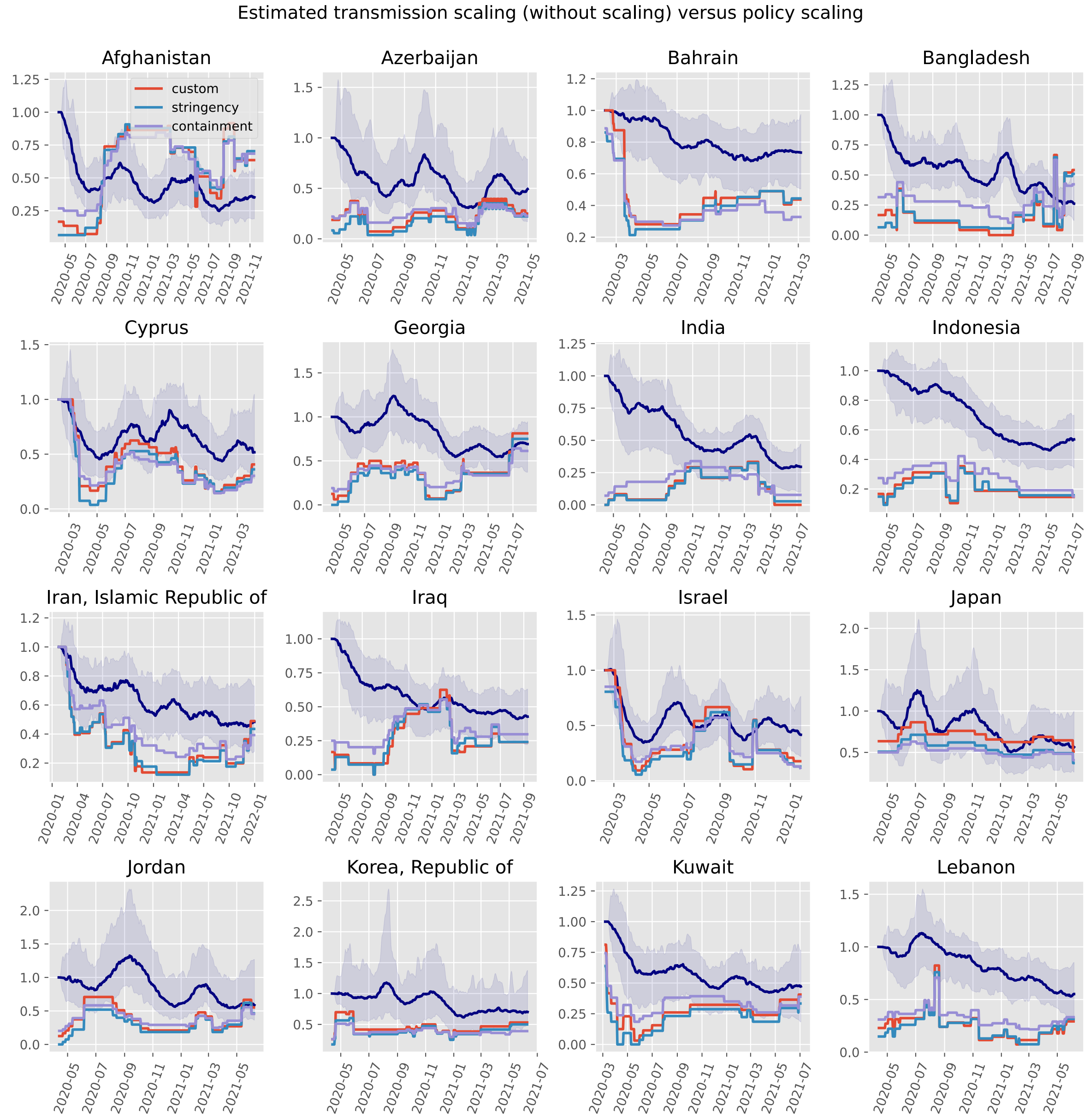

Estimated transmission scaling (without scaling) versus policy scaling
Afghanistan
custom
stringency
containment
Azerbaijan
Bahrain
Bangladesh
Cyprus
Georgia
India
Indonesia
Iran, Islamic Republic of
Iraq
Israel
Japan
Jordan
Korea, Republic of
Kuwait
Lebanon

Estimated transmission scaling (without scaling) versus policy scaling

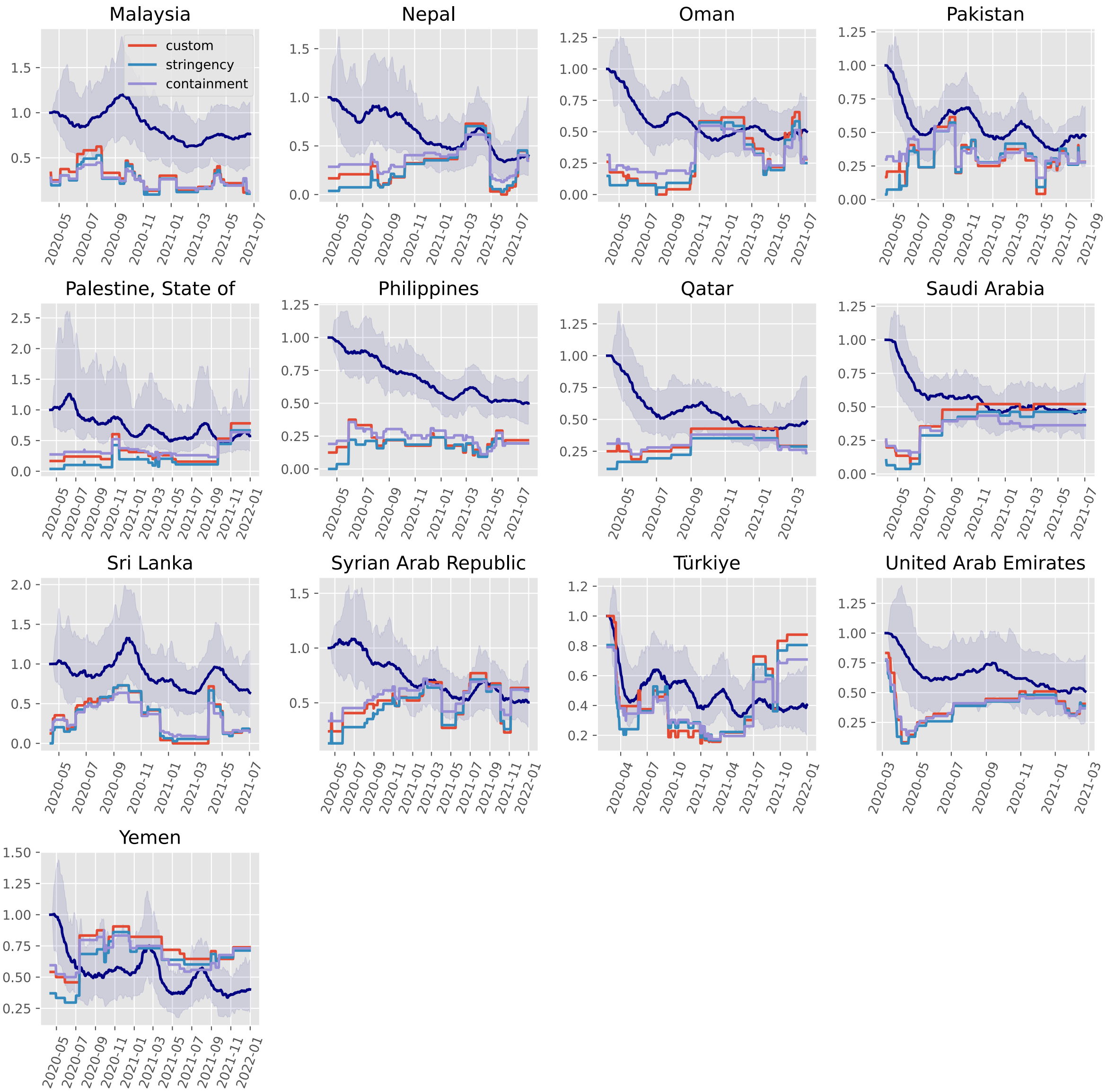

#### Europe

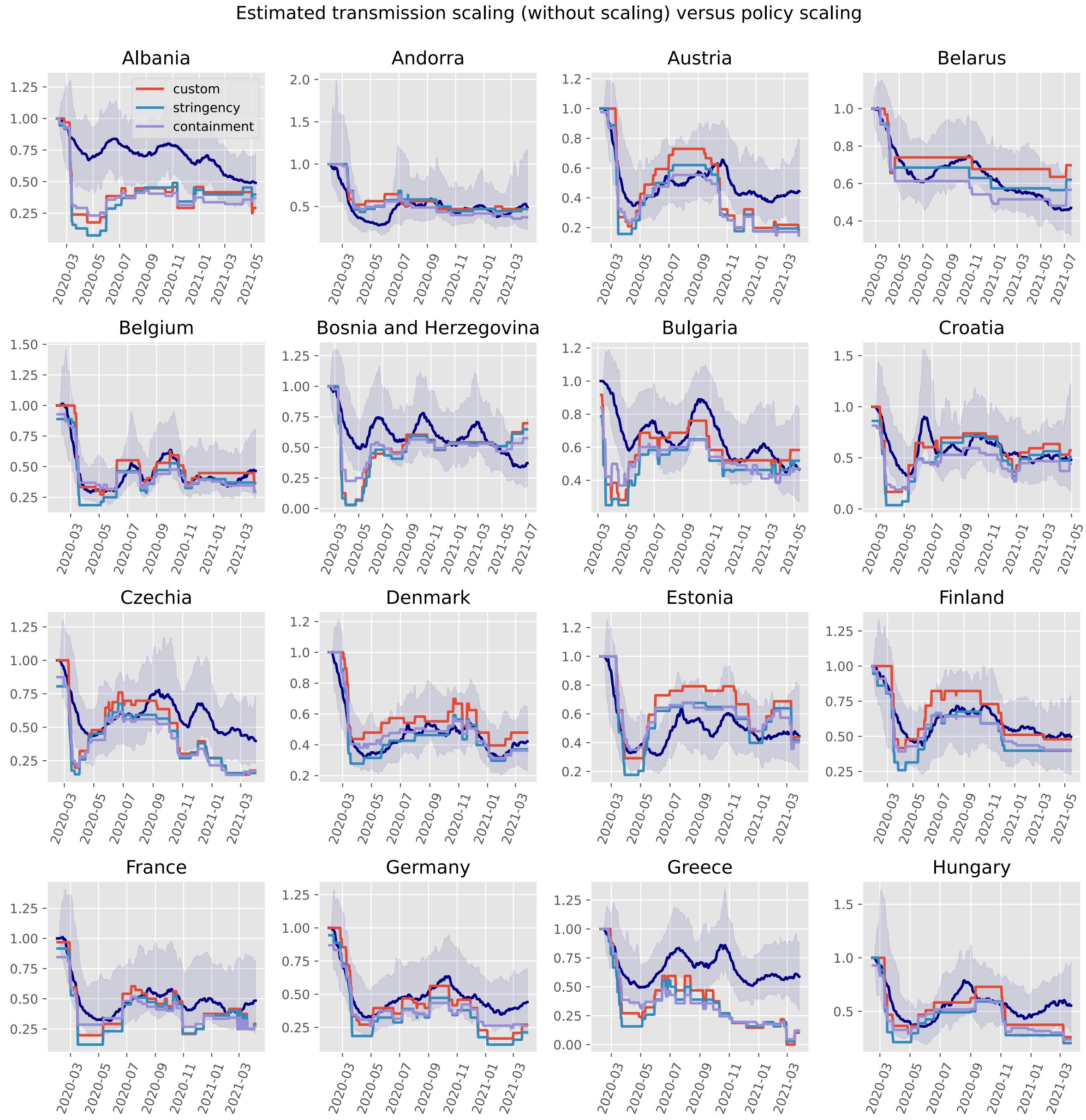
Estimated transmission scaling (without scaling) versus policy scaling
Albania
Andorra
Austria
Belarus
Belgium
Bosnia and Herzegovina
Bulgaria
Croatia
Czechia
Denmark
Estonia
Finland
France
Germany
Greece
Hungary
custom
stringency
containment

Estimated transmission scaling (without scaling) versus policy scaling

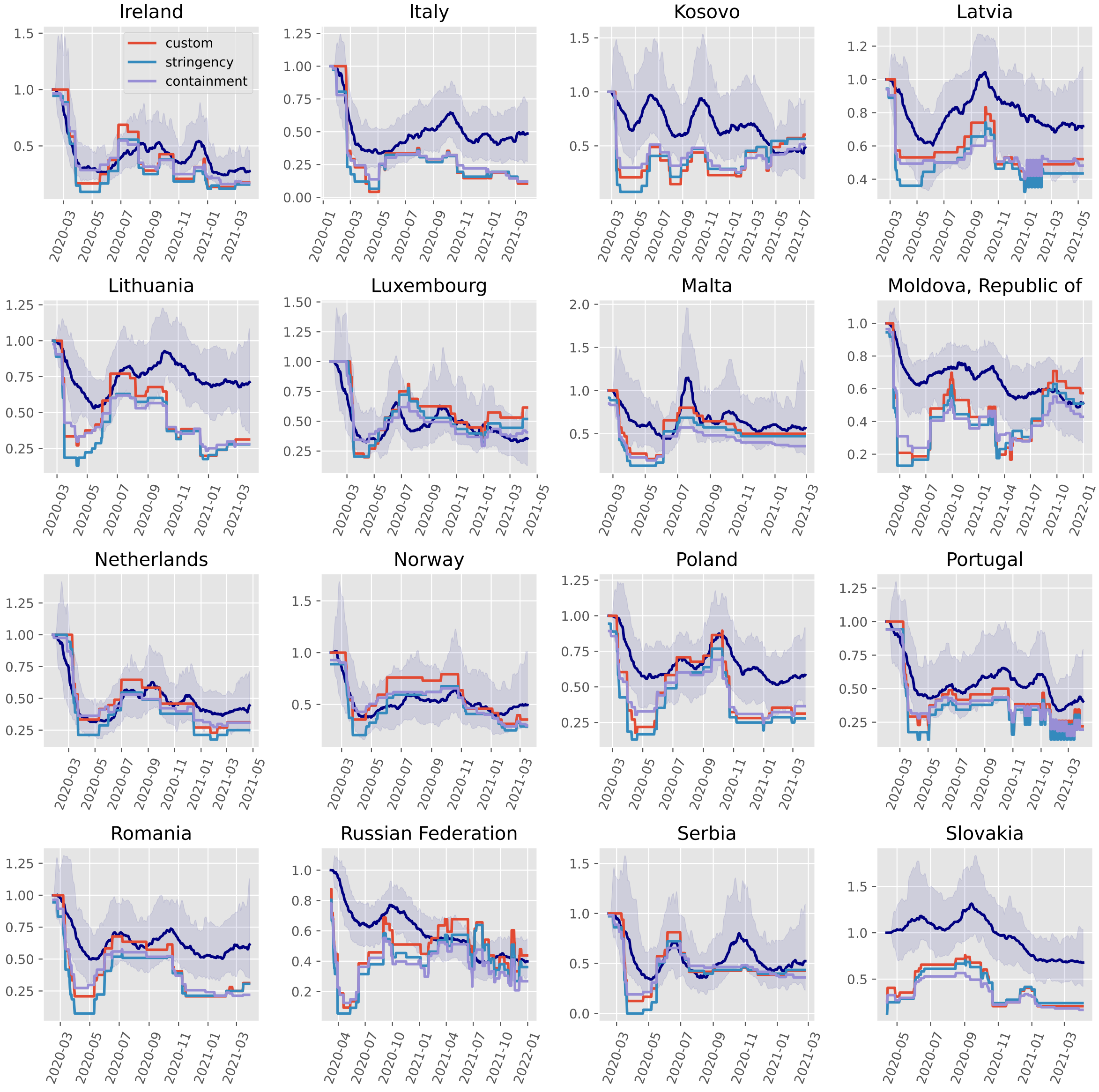

Estimated transmission scaling (without scaling) versus policy scaling

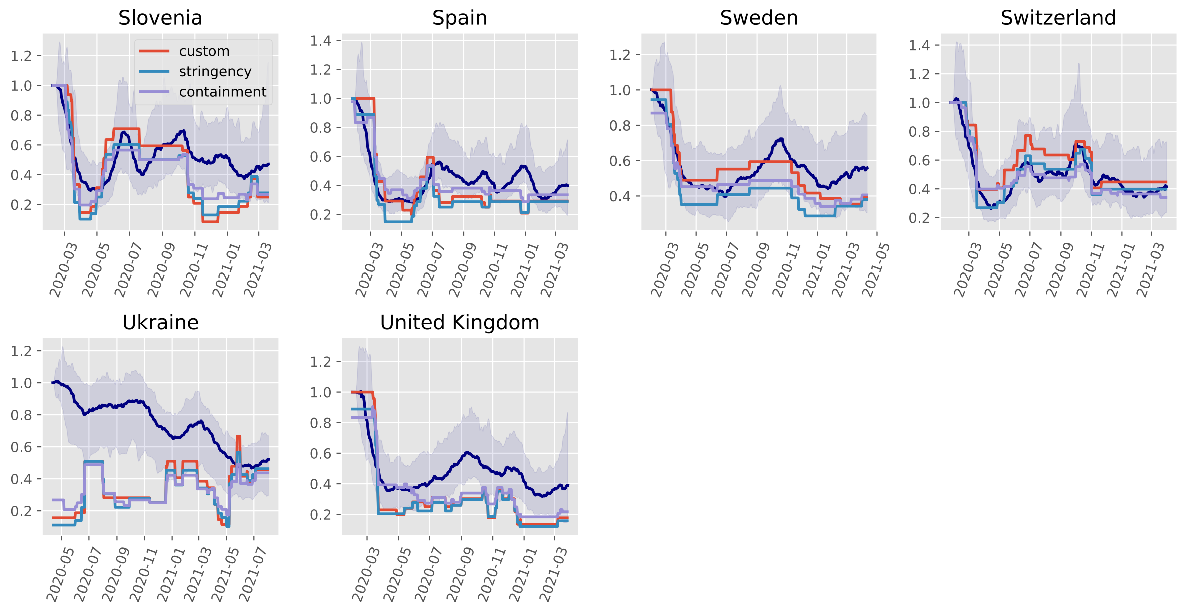

#### North America

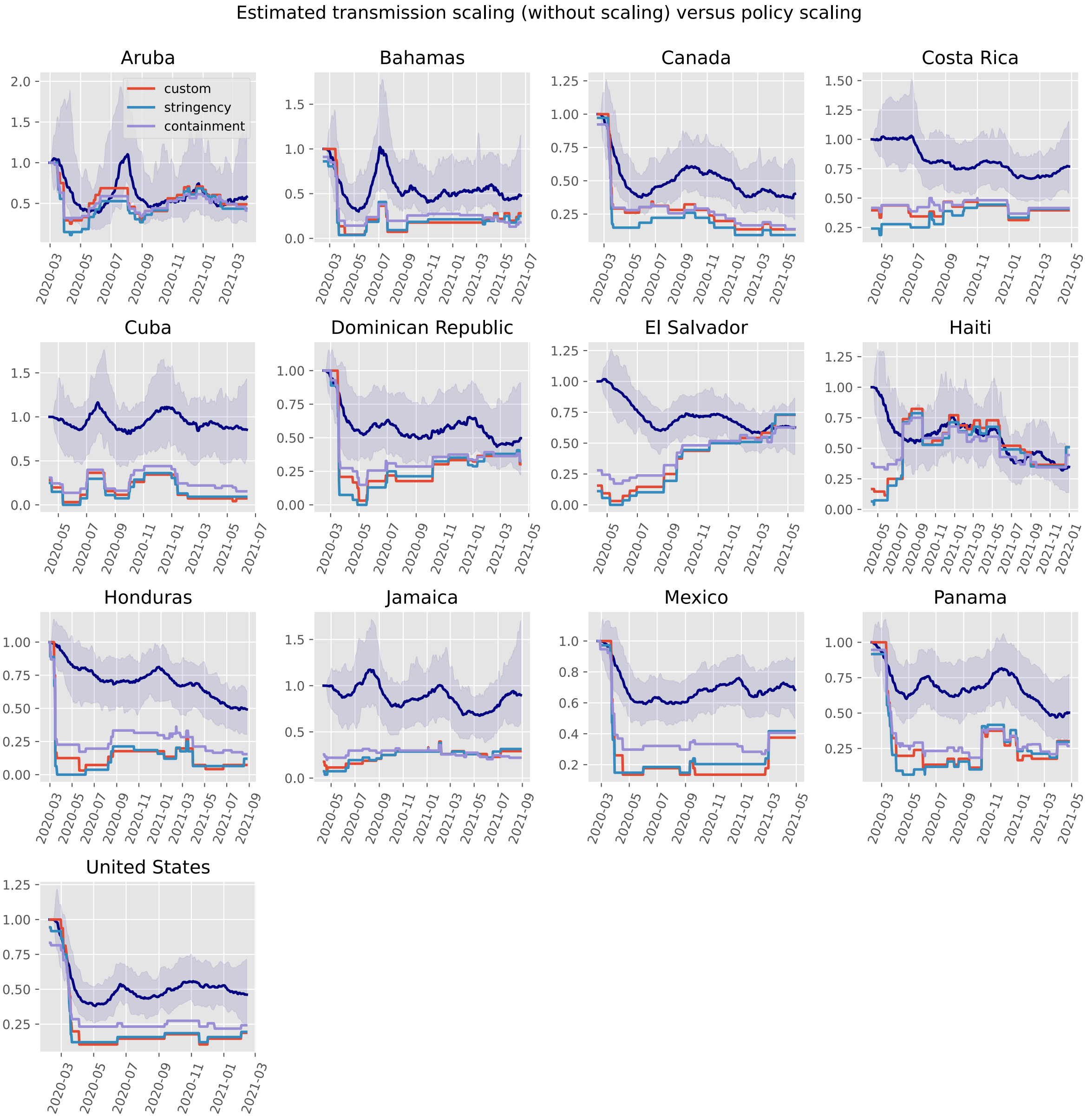

#### Oceania

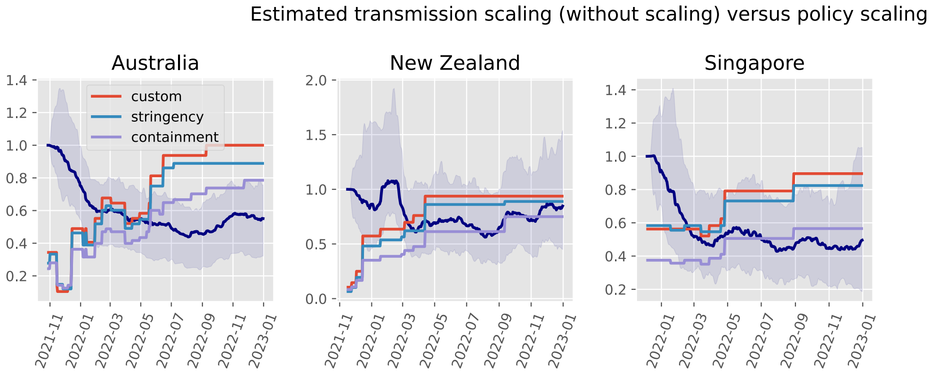

#### South America

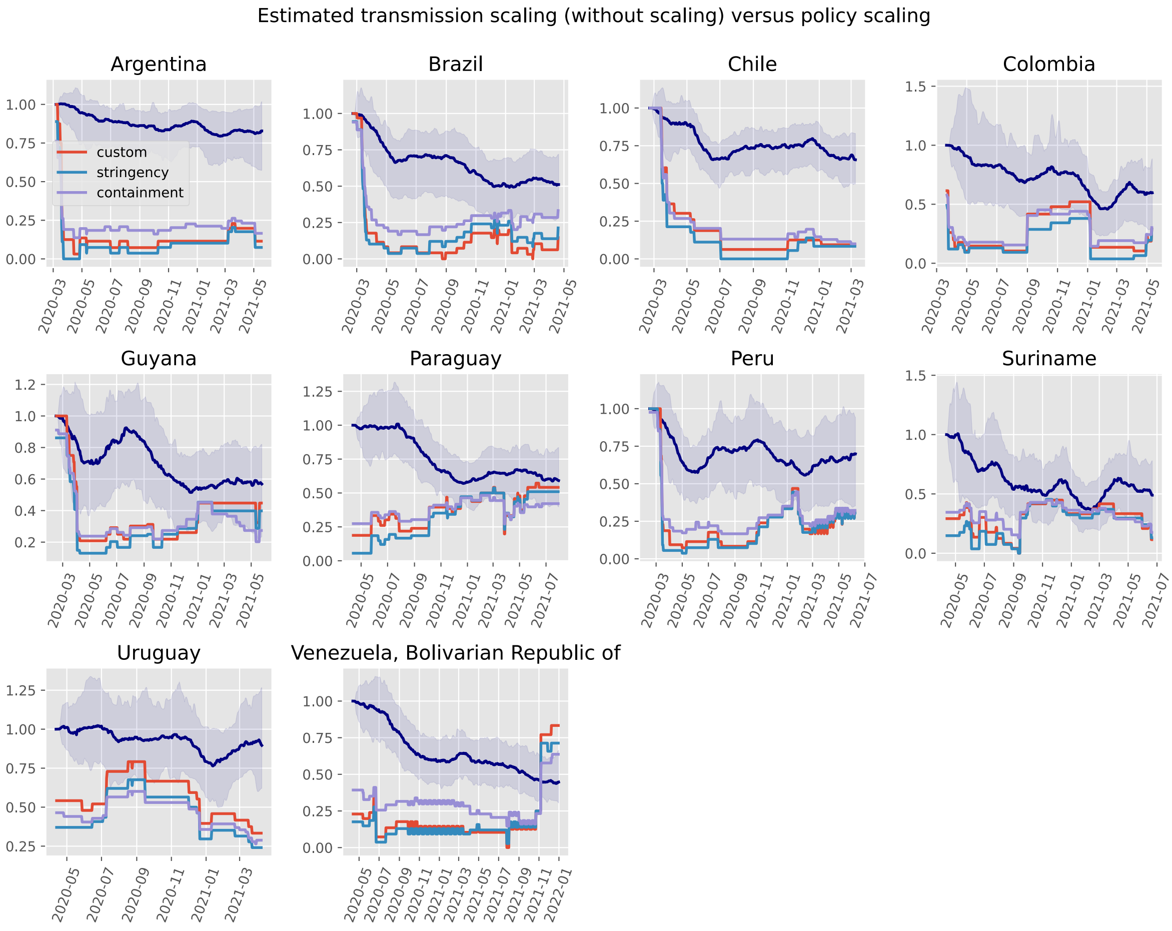

| | no scaling | OxCGRT floored | OxCGRT independent | Google mobility | Facebook tiles visited mobility | Facebook single tile mobility |
|---|---|---|---|---|---|---|
| Afghanistan | 4f57571 | b934196 | b934196 | b934196 | no analysis | no analysis |
| Albania | 4f57571 | b934196 | b934196 | no analysis | b934196 | b934196 |
| Algeria | 4f57571 | b934196 | b934196 | no analysis | b934196 | b934196 |
| Andorra | 4f57571 | b934196 | b934196 | no analysis | no analysis | no analysis |
| Angola | 4f57571 | b934196 | b934196 | cb8324a | cb8324a | cb8324a |
| Argentina | 4f57571 | b934196 | b934196 | b934196 | b934196 | b934196 |
| Aruba | 4f57571 | b934196 | b934196 | 4f57571 | no analysis | no analysis |
| Australia | b934196 | b934196 | b934196 | no analysis | no analysis | no analysis |
| Austria | 4f57571 | b934196 | b934196 | b934196 | b934196 | b934196 |
| Azerbaijan | 4f57571 | b934196 | b934196 | no analysis | no analysis | no analysis |
| Bahamas | 4f57571 | b934196 | b934196 | b934196 | no analysis | no analysis |
| Bahrain | 4f57571 | b934196 | b934196 | 4f57571 | no analysis | no analysis |
| Bangladesh | 4f57571 | b934196 | b934196 | b934196 | b934196 | b934196 |
| Belarus | 4f57571 | b934196 | b934196 | cb8324a | cb8324a | b934196 |
| Belgium | 4f57571 | b934196 | b934196 | 4f57571 | 4f57571 | 4f57571 |
| Benin | 4f57571 | b934196 | b934196 | 4f57571 | 4f57571 | b934196 |
| Bosnia and Herzegovina | 4f57571 | b934196 | b934196 | b934196 | b934196 | b934196 |
| Brazil | 4f57571 | b934196 | b934196 | 4f57571 | 4f57571 | 4f57571 |
| Bulgaria | 4f57571 | b934196 | b934196 | b934196 | b934196 | b934196 |
| Burkina Faso | 4f57571 | b934196 | b934196 | 4f57571 | 4f57571 | 4f57571 |
| Cabo Verde | 4f57571 | b934196 | b934196 | 4f57571 | no analysis | no analysis |
| Cameroon | 4f57571 | b934196 | b934196 | b934196 | b934196 | b934196 |
| Canada | 4f57571 | b934196 | b934196 | b934196 | b934196 | b934196 |
| Chile | 4f57571 | b934196 | b934196 | 4f57571 | b934196 | b934196 |
| Colombia | 4f57571 | b934196 | b934196 | 4f57571 | 4f57571 | b934196 |
| Congo | 4f57571 | b934196 | b934196 | no analysis | 4f57571 | b934196 |
| Costa Rica | 4f57571 | b934196 | b934196 | 4f57571 | b934196 | b934196 |
| Croatia | 4f57571 | b934196 | b934196 | b934196 | b934196 | b934196 |
| Cuba | 4f57571 | b934196 | b934196 | no analysis | no analysis | no analysis |
| Cyprus | 4f57571 | b934196 | b934196 | no analysis | no analysis | no analysis |
