## Supplemental File 5 for "Quantifying the contribution of policy measures to COVID-19 dynamics across 126 countries"

### Comparisons of the transmission scaling dispersion parameter posterior by analysis approach

**Proportion better than baseline** 25

**Proportion floored better than independent** 29

**Commits used for analyses** 33

#### Purpose

This document presents results based on the posterior distribution of the residual transmission scaling dispersion parameter. This quantity governs the distribution in the change in residual scaling for transmission from one value in the process series to the subsequent update (in log space). As such, smaller values imply that smaller updates could still result in good calibrations. Lower values for this parameter can therefore represent less dramatic changes needing to be applied through the non-mechanistic component of the model. As such, we interpret mobility analysis approaches for which this posterior distribution of this parameter was lower as being a more plausible representation of reality. This is intended as a more formal quantification of the differences in variation in the non-mechanistic transmission scaling presented in the previous document.

#### Comparison of posterior distributions

##### Policy analyses

Comparisons between baseline and the two OxCGRT scaling configurations.

### Africa

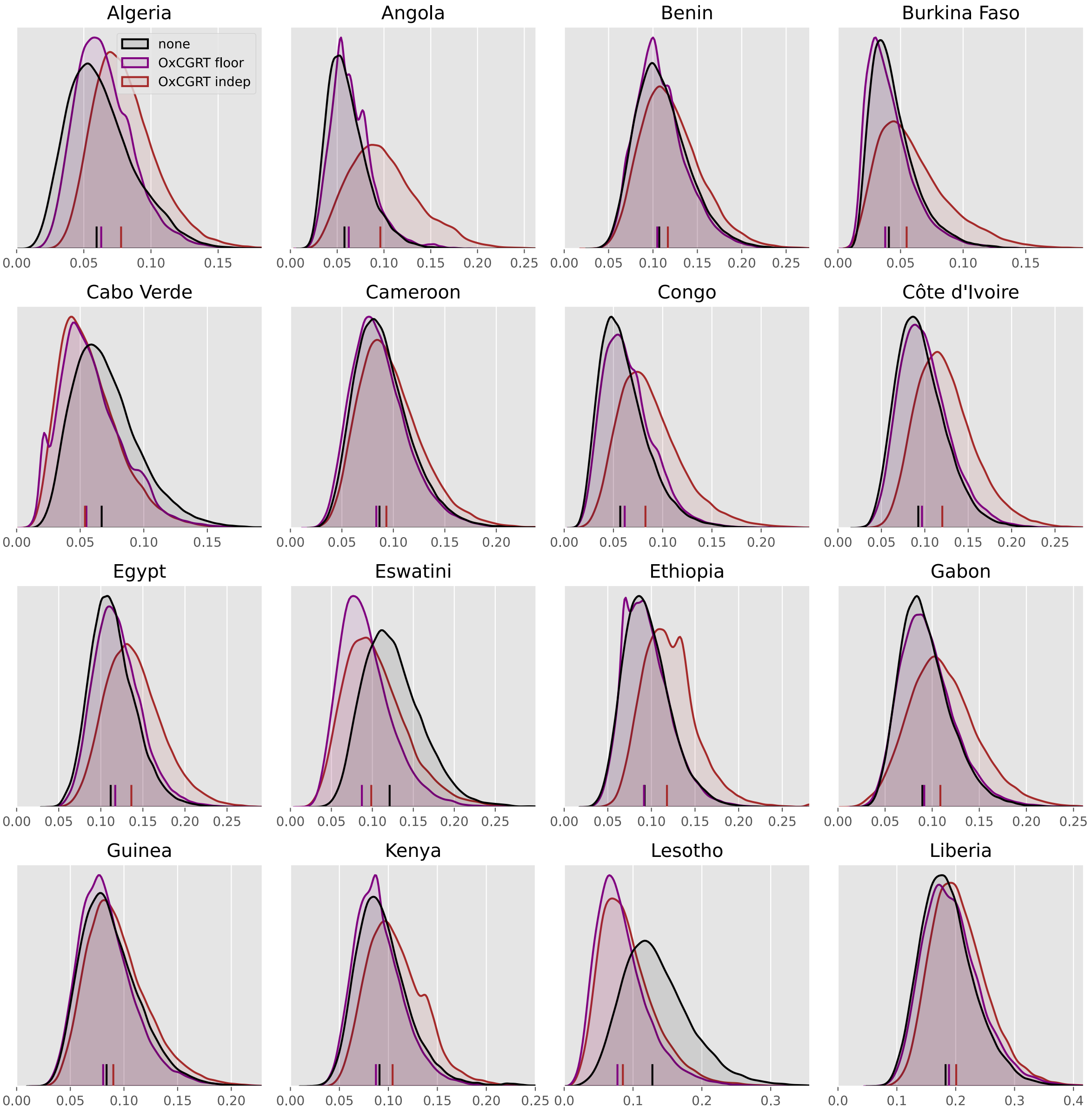
Algeria
none
OxCGRT floor
OxCGRT indep
Angola
Benin
Burkina Faso
Cabo Verde
Cameroon
Congo
Côte d'Ivoire
Egypt
Eswatini
Ethiopia
Gabon
Guinea
Kenya
Lesotho
Liberia

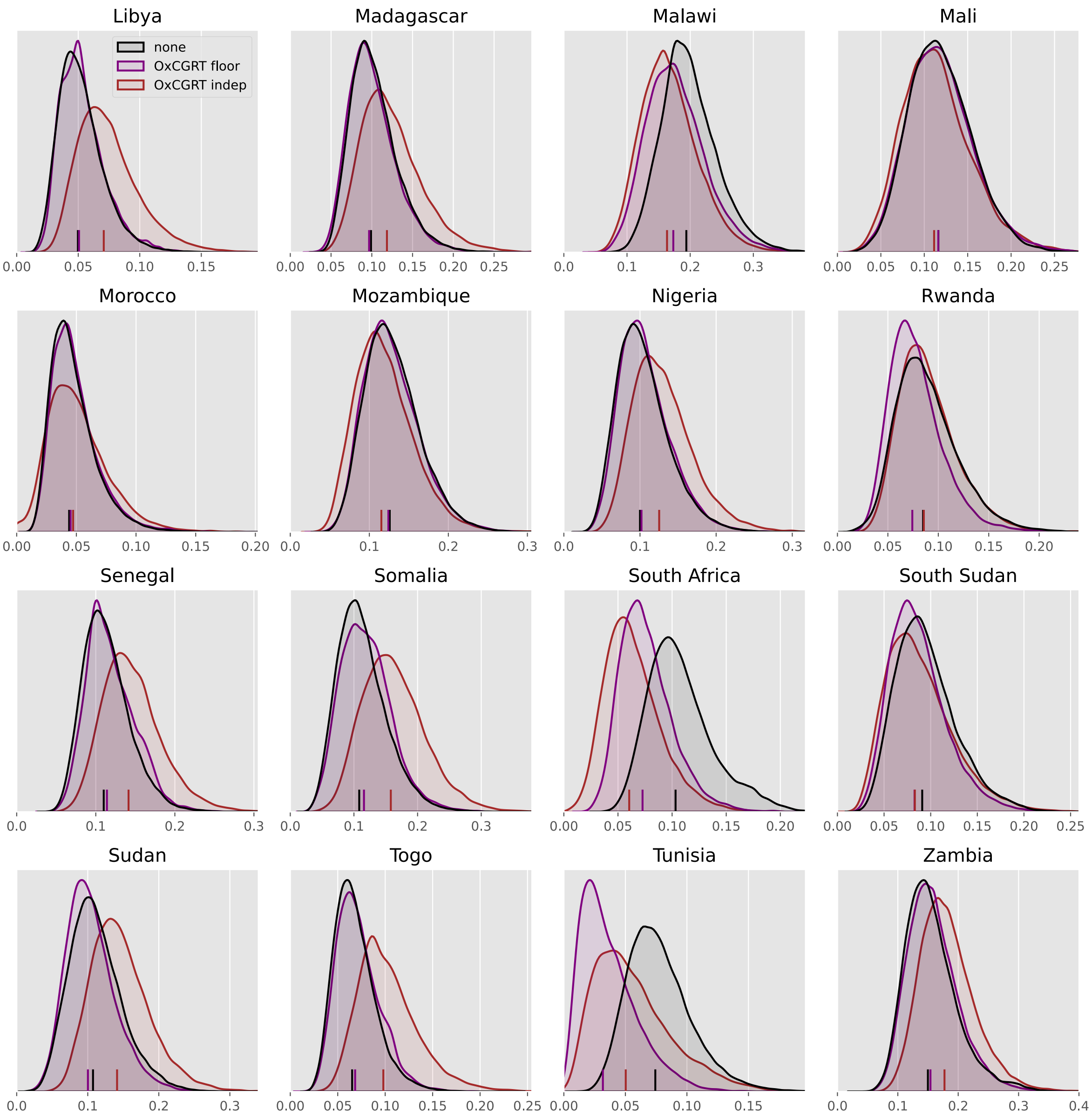
Libya
Madagascar
Malawi
Mali
none
OxCGRT floor
OxCGRT indep
Morocco
Mozambique
Nigeria
Rwanda
Senegal
Somalia
South Africa
South Sudan
Sudan
Togo
Tunisia
Zambia

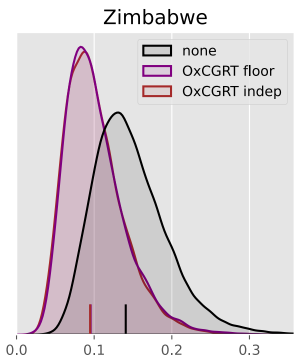
Zimbabwe
none
OxCGRT floor
OxCGRT indep
0.0
0.1
0.2
0.3

#### Asia

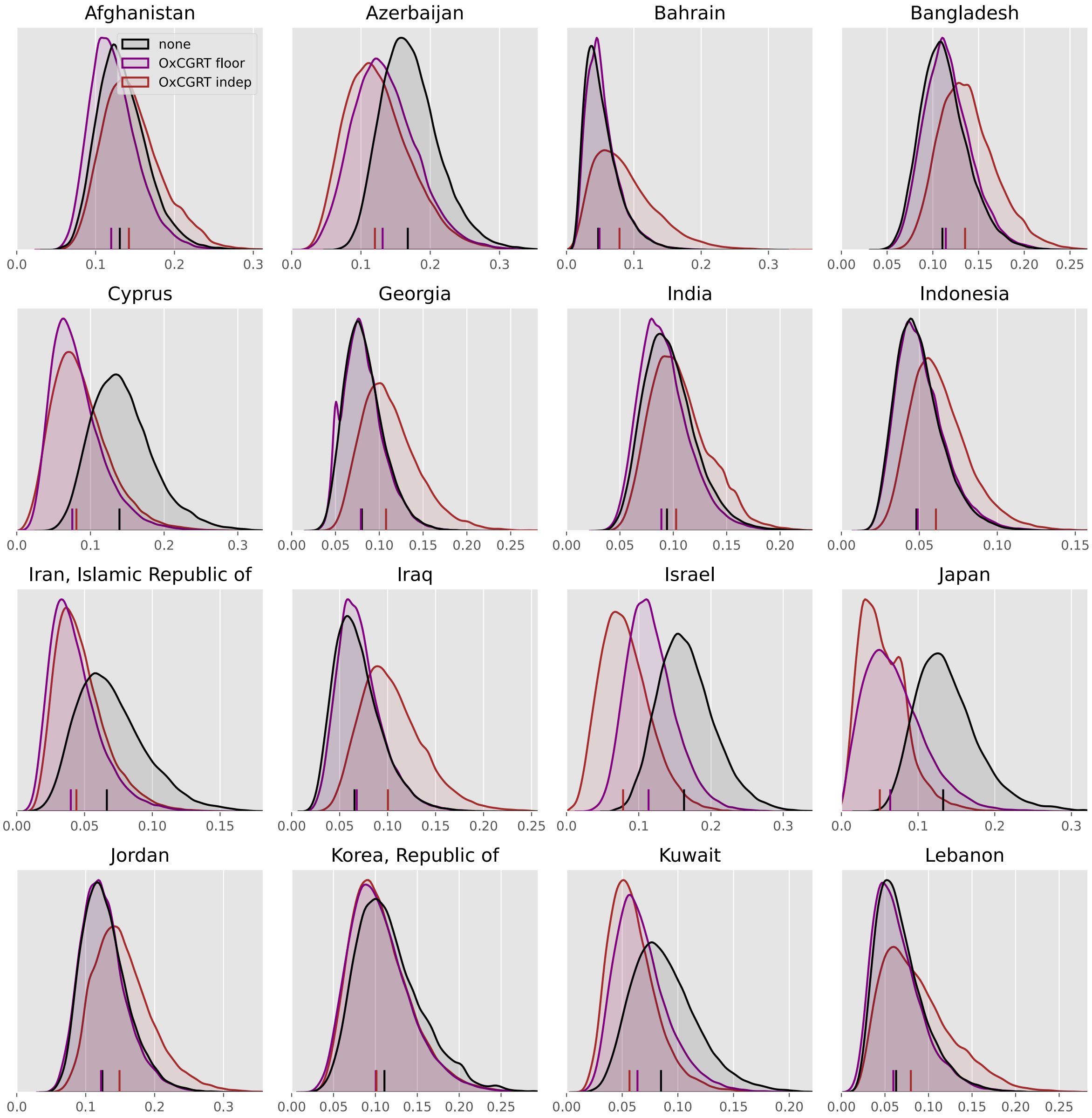
Afghanistan
none
OxCGRT floor
OxCGRT indep
Azerbaijan
Bahrain
Bangladesh
Cyprus
Georgia
India
Indonesia
Iran, Islamic Republic of
Iraq
Israel
Japan
Jordan
Korea, Republic of
Kuwait
Lebanon

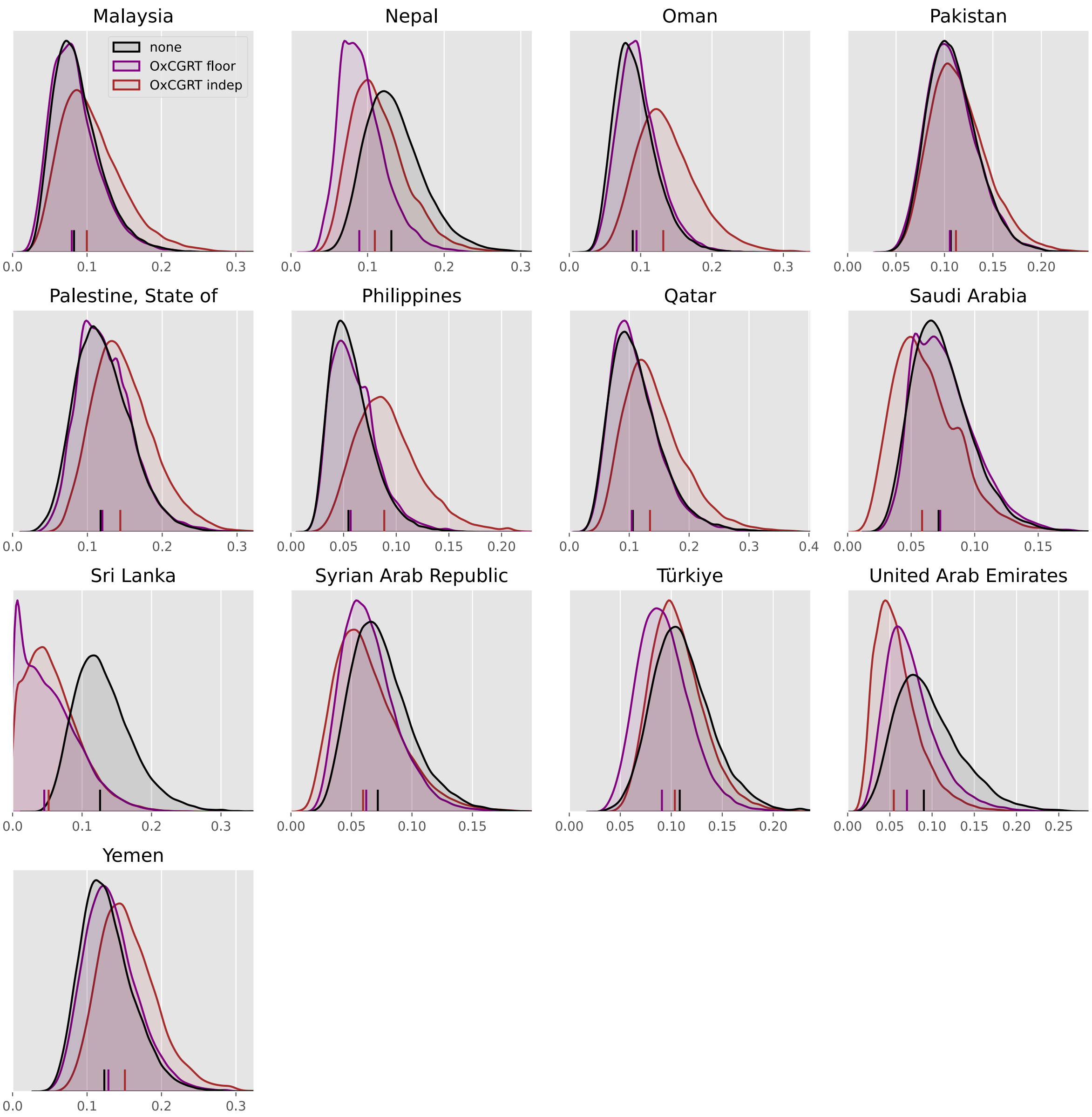
Malaysia
none
OxCGRT floor
OxCGRT indep
0.0 0.1 0.2 0.3
Nepal
0.0 0.1 0.2 0.3
Oman
0.0 0.1 0.2 0.3
Pakistan
0.00 0.05 0.10 0.15 0.20
Palestine, State of
0.0 0.1 0.2 0.3
Philippines
0.00 0.05 0.10 0.15 0.20
Qatar
0.0 0.1 0.2 0.3 0.4
Saudi Arabia
0.00 0.05 0.10 0.15
Sri Lanka
0.0 0.1 0.2 0.3
Syrian Arab Republic
0.00 0.05 0.10 0.15
Türkiye
0.00 0.05 0.10 0.15 0.20
United Arab Emirates
0.00 0.05 0.10 0.15 0.20 0.25
Yemen
0.0 0.1 0.2 0.3

#### Europe

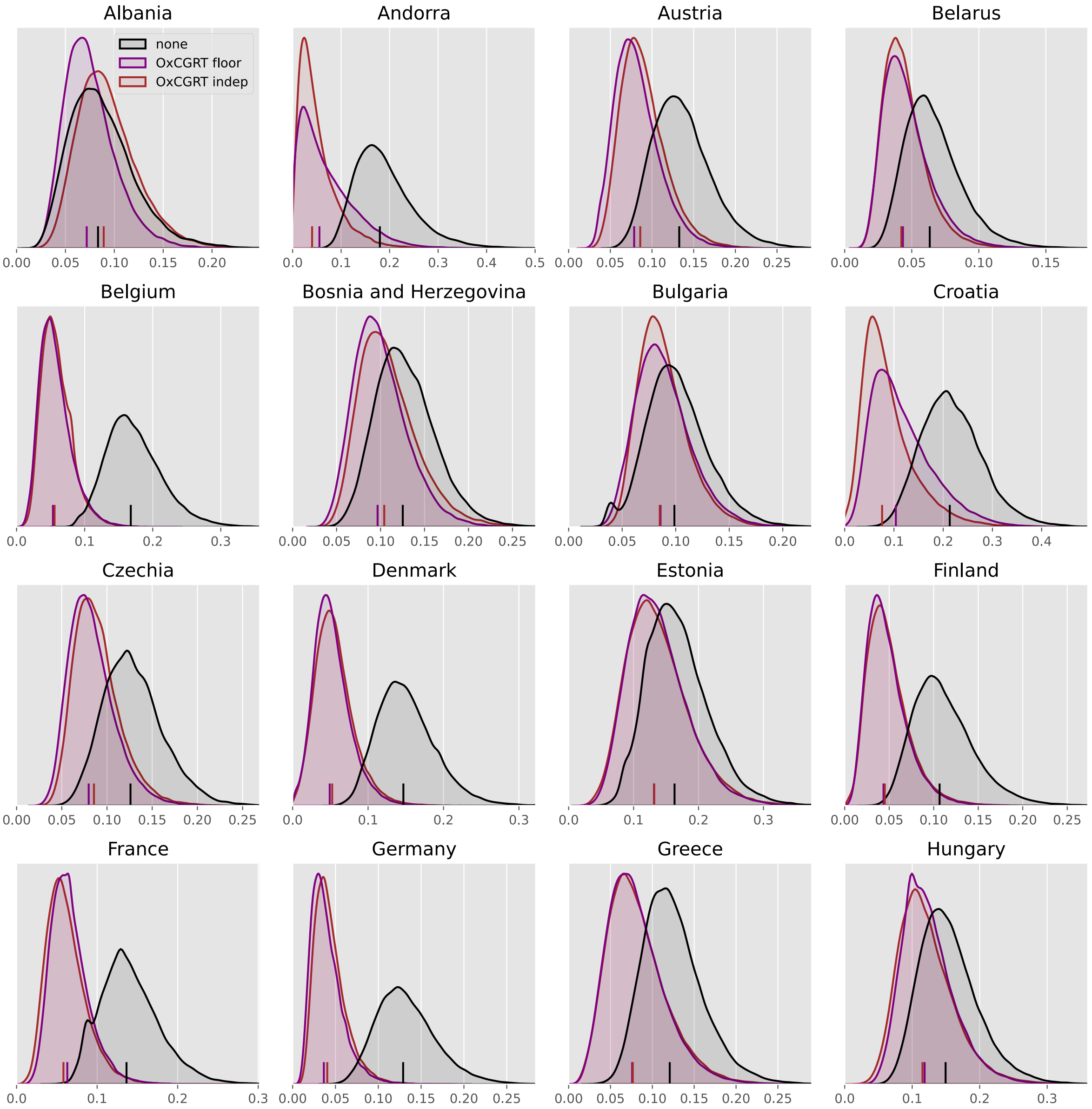

Albania
Andorra
Austria
Belarus
Belgium
Bosnia and Herzegovina
Bulgaria
Croatia
Czechia
Denmark
Estonia
Finland
France
Germany
Greece
Hungary
none
OxCGRT floor
OxCGRT indep

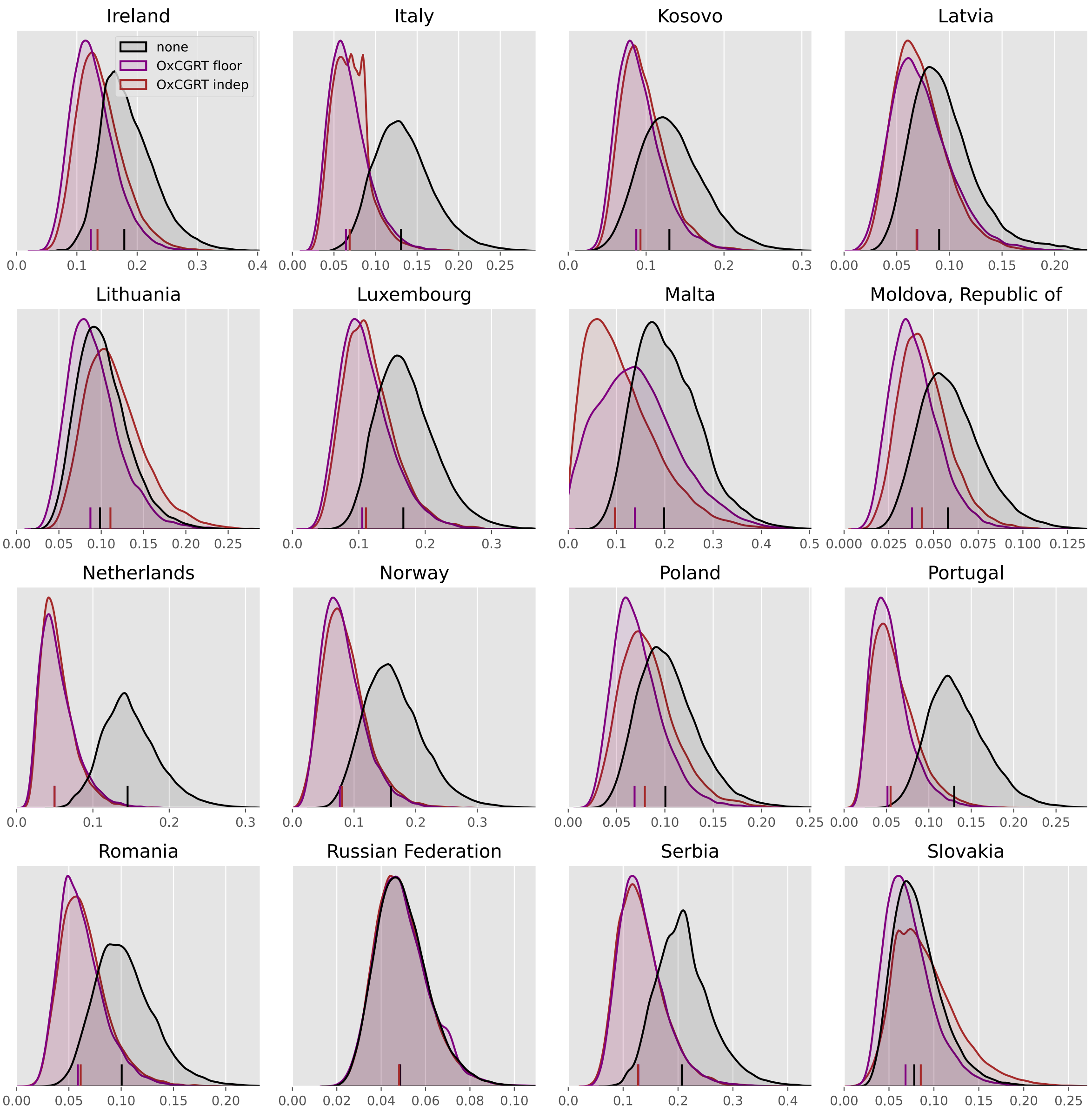

Ireland
Italy
Kosovo
Latvia
none
OxCGRT floor
OxCGRT indep
Lithuania
Luxembourg
Malta
Moldova, Republic of
Netherlands
Norway
Poland
Portugal
Romania
Russian Federation
Serbia
Slovakia

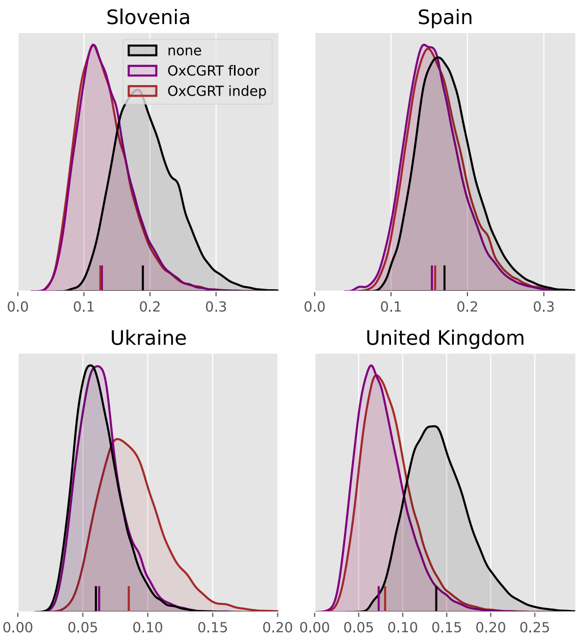

Slovenia
none
OxCGRT floor
OxCGRT indep
0.0
0.1
0.2
0.3
Spain
0.0
0.1
0.2
0.3
Sweden
0.00
0.05
0.10
0.15
0.20
Switzerland
0.0
0.1
0.2
0.3
Ukraine
0.00
0.05
0.10
0.15
0.20
United Kingdom
0.00
0.05
0.10
0.15
0.20
0.25

#### North America

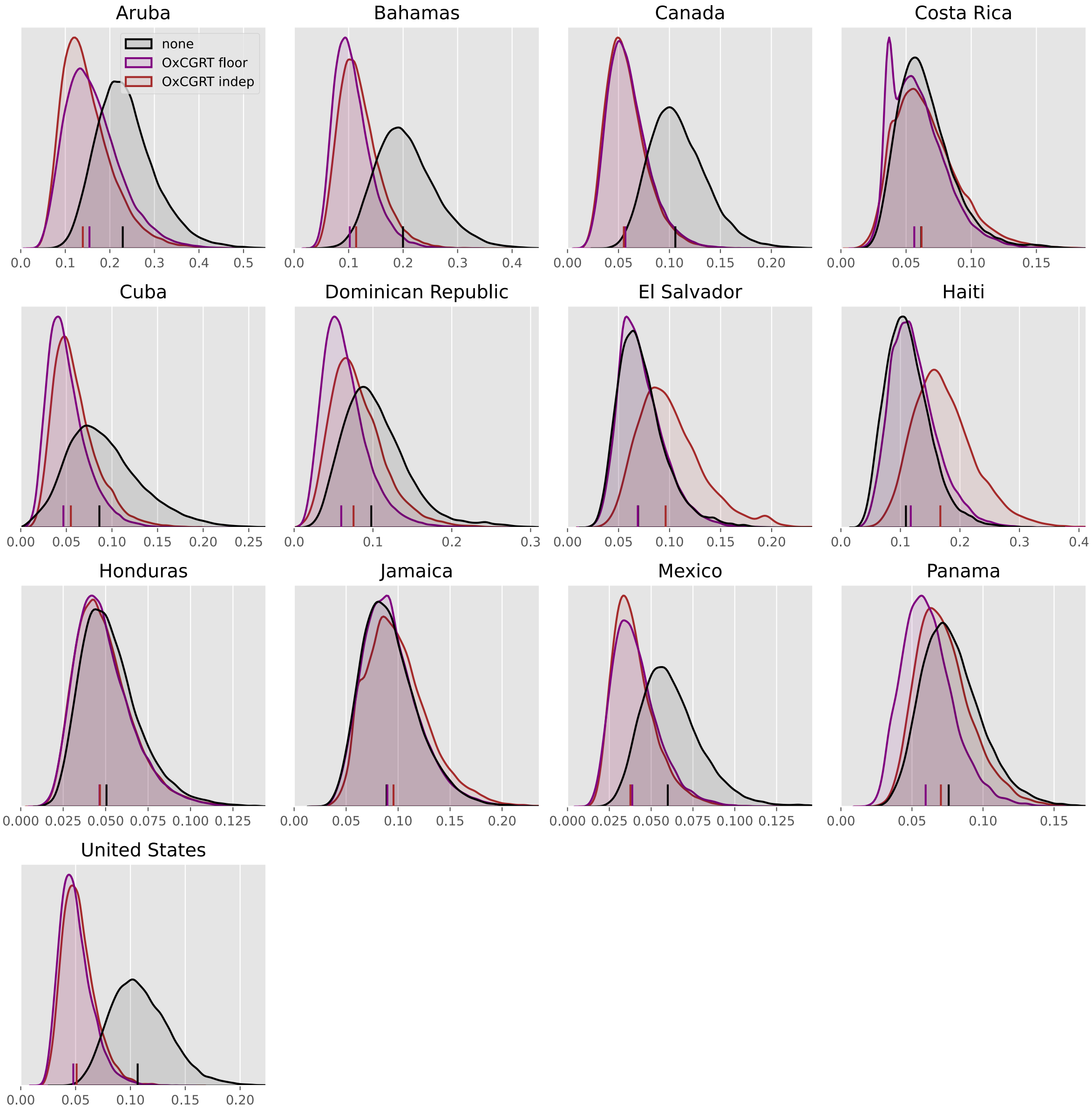

#### Oceania

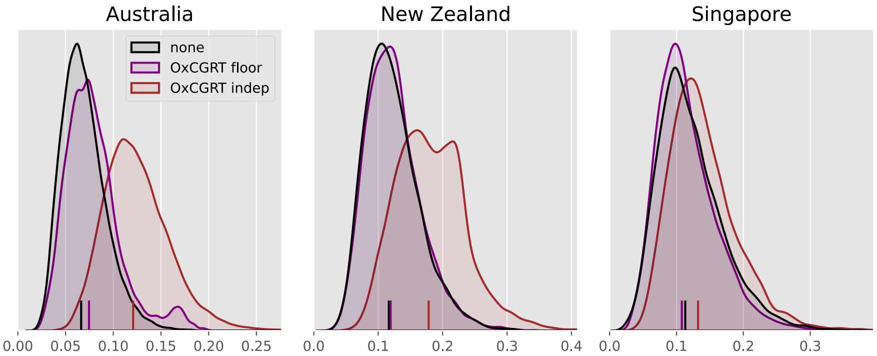
Australia
New Zealand
Singapore
none
OxCGRT floor
OxCGRT indep
0.00
0.05
0.10
0.15
0.20
0.25
0.0
0.1
0.2
0.3
0.4
0.0
0.1
0.2
0.3

#### South America

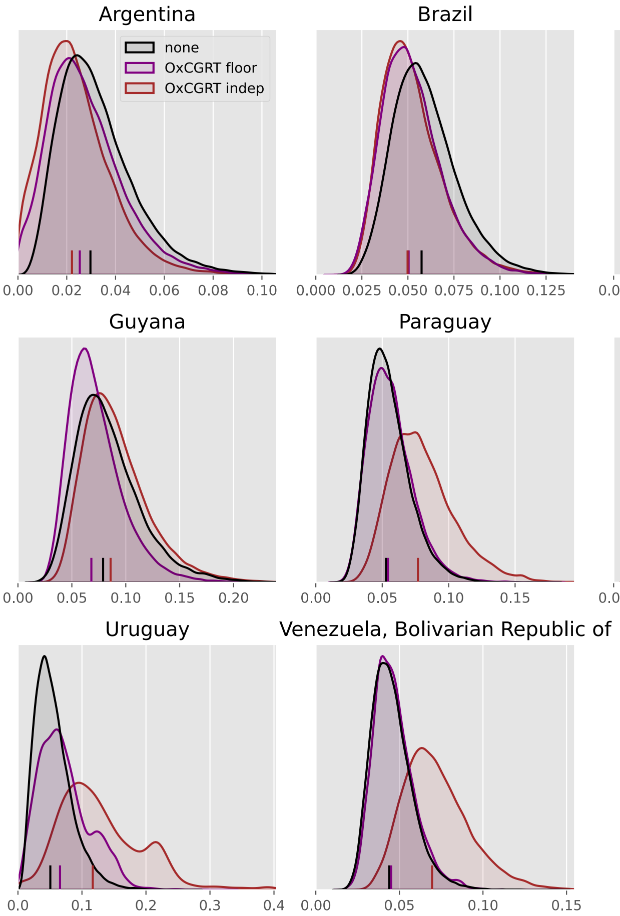

#### All analyses

Comparisons between baseline and all policy and mobility-scaled analyses.

#### Africa

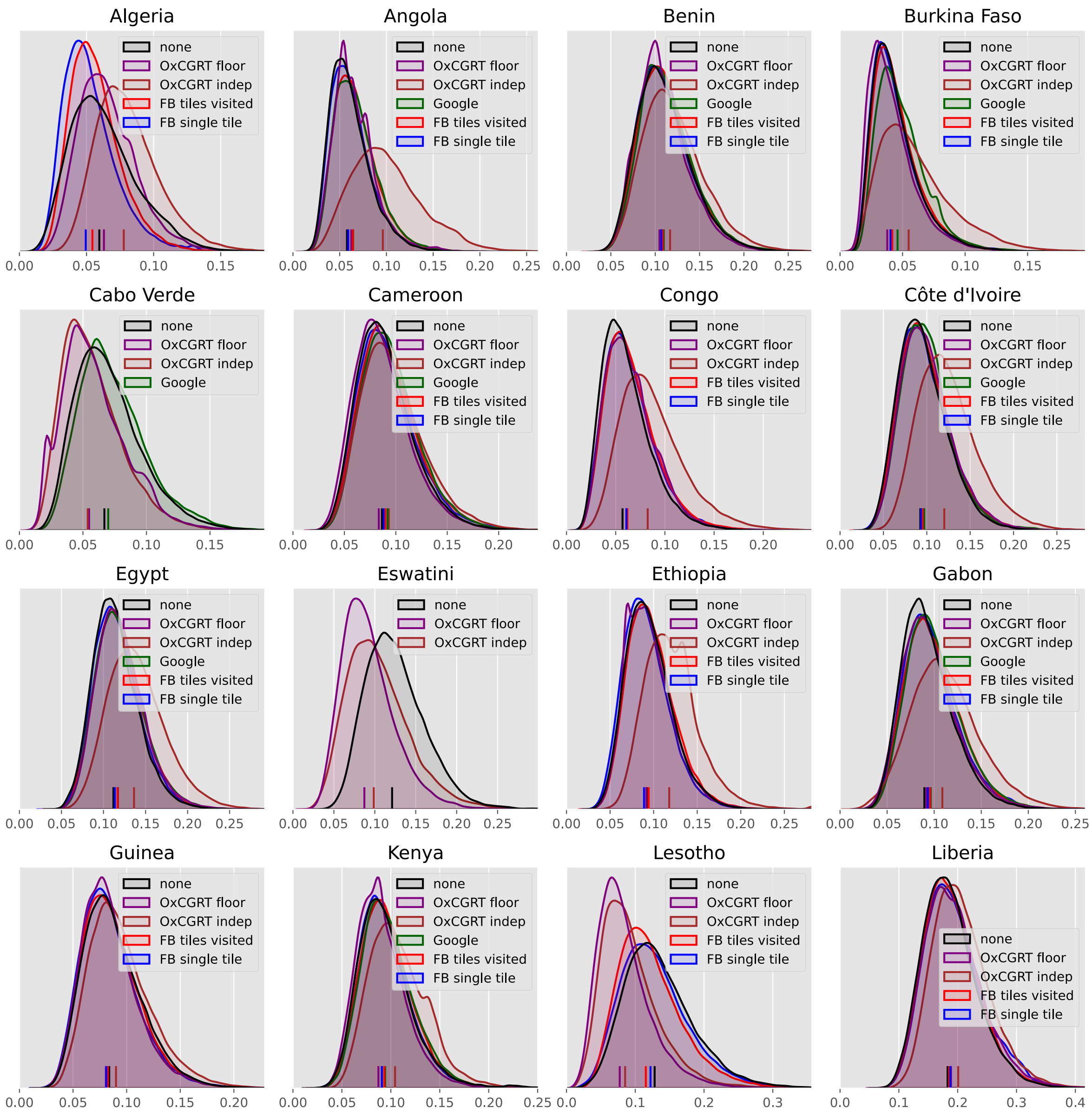

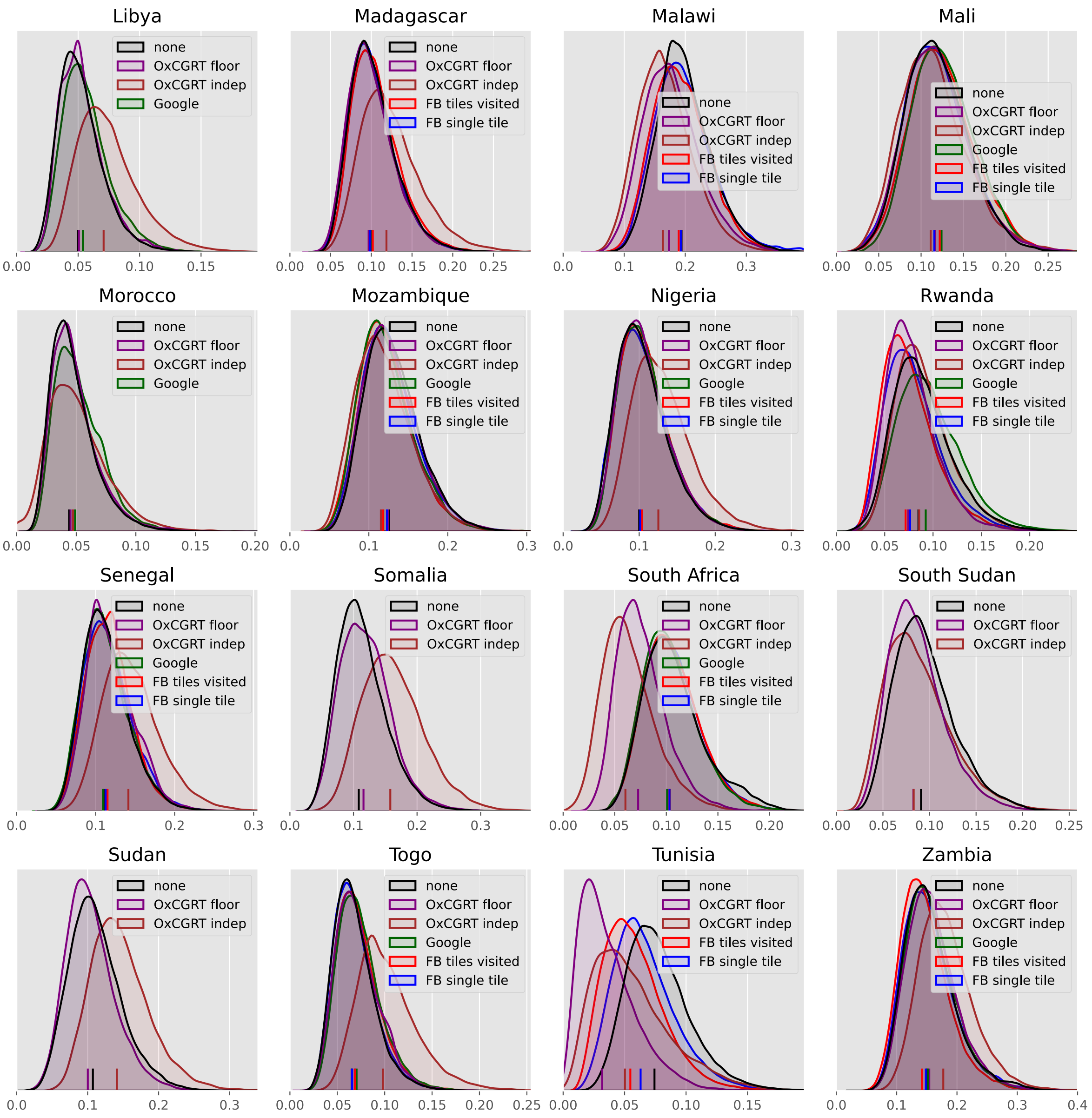

Libya
none
OxCGRT floor
OxCGRT indep
Google
0.00 0.05 0.10 0.15
Madagascar
none
OxCGRT floor
OxCGRT indep
FB tiles visited
FB single tile
0.00 0.05 0.10 0.15 0.20 0.25
Malawi
none
OxCGRT floor
OxCGRT indep
FB tiles visited
FB single tile
0.0 0.1 0.2 0.3
Mali
none
OxCGRT floor
OxCGRT indep
Google
FB tiles visited
FB single tile
0.00 0.05 0.10 0.15 0.20 0.25
Morocco
none
OxCGRT floor
OxCGRT indep
Google
0.00 0.05 0.10 0.15 0.20
Mozambique
none
OxCGRT floor
OxCGRT indep
Google
FB tiles visited
FB single tile
0.0 0.1 0.2 0.3
Nigeria
none
OxCGRT floor
OxCGRT indep
Google
FB tiles visited
FB single tile
0.0 0.1 0.2 0.3
Rwanda
none
OxCGRT floor
OxCGRT indep
Google
FB tiles visited
FB single tile
0.00 0.05 0.10 0.15 0.20
Senegal
none
OxCGRT floor
OxCGRT indep
Google
FB tiles visited
FB single tile
0.0 0.1 0.2 0.3
Somalia
none
OxCGRT floor
OxCGRT indep
0.0 0.1 0.2 0.3
South Africa
none
OxCGRT floor
OxCGRT indep
Google
FB tiles visited
FB single tile
0.00 0.05 0.10 0.15 0.20
South Sudan
none
OxCGRT floor
OxCGRT indep
0.00 0.05 0.10 0.15 0.20 0.25
Sudan
none
OxCGRT floor
OxCGRT indep
0.0 0.1 0.2 0.3
Togo
none
OxCGRT floor
OxCGRT indep
Google
FB tiles visited
FB single tile
0.00 0.05 0.10 0.15 0.20 0.25
Tunisia
none
OxCGRT floor
OxCGRT indep
FB tiles visited
FB single tile
0.00 0.05 0.10 0.15
Zambia
none
OxCGRT floor
OxCGRT indep
Google
FB tiles visited
FB single tile
0.0 0.1 0.2 0.3 0.4

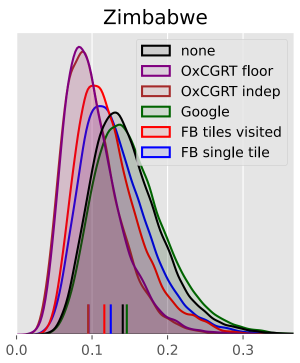

Zimbabwe
none
OxCGRT floor
OxCGRT indep
Google
FB tiles visited
FB single tile
0.0
0.1
0.2
0.3

#### Asia

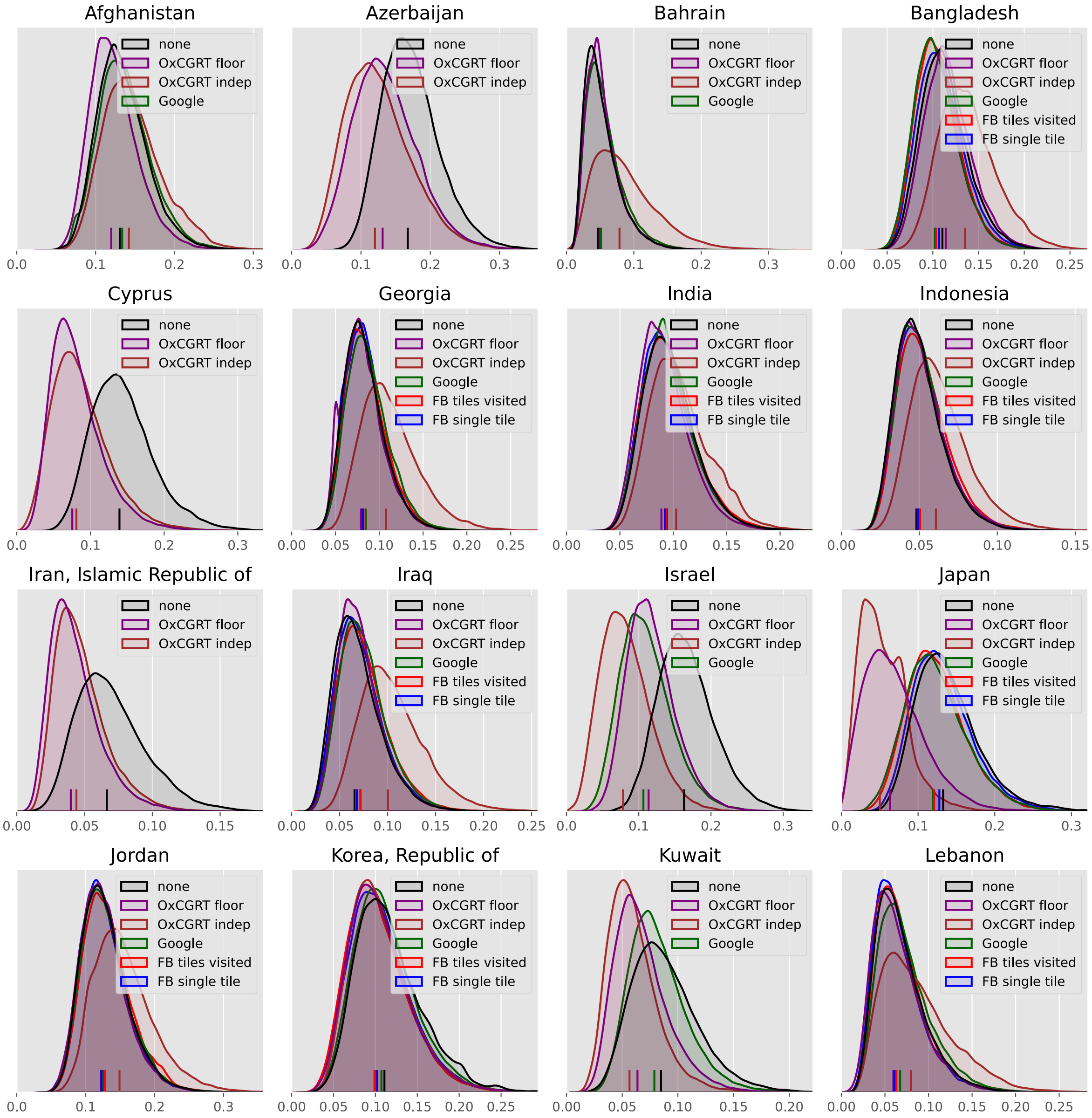

Afghanistan
none
OxCGRT floor
OxCGRT indep
Google
Azerbaijan
none
OxCGRT floor
OxCGRT indep
Bahrain
none
OxCGRT floor
OxCGRT indep
Google
Bangladesh
none
OxCGRT floor
OxCGRT indep
Google
FB tiles visited
FB single tile
Cyprus
none
OxCGRT floor
OxCGRT indep
Georgia
none
OxCGRT floor
OxCGRT indep
Google
FB tiles visited
FB single tile
India
none
OxCGRT floor
OxCGRT indep
Google
FB tiles visited
FB single tile
Indonesia
none
OxCGRT floor
OxCGRT indep
Google
FB tiles visited
FB single tile
Iran, Islamic Republic of
none
OxCGRT floor
OxCGRT indep
Iraq
none
OxCGRT floor
OxCGRT indep
Google
FB tiles visited
FB single tile
Israel
none
OxCGRT floor
OxCGRT indep
Google
Japan
none
OxCGRT floor
OxCGRT indep
Google
FB tiles visited
FB single tile
Jordan
none
OxCGRT floor
OxCGRT indep
Google
FB tiles visited
FB single tile
Korea, Republic of
none
OxCGRT floor
OxCGRT indep
Google
FB tiles visited
FB single tile
Kuwait
none
OxCGRT floor
OxCGRT indep
Google
Lebanon
none
OxCGRT floor
OxCGRT indep
Google
FB tiles visited
FB single tile

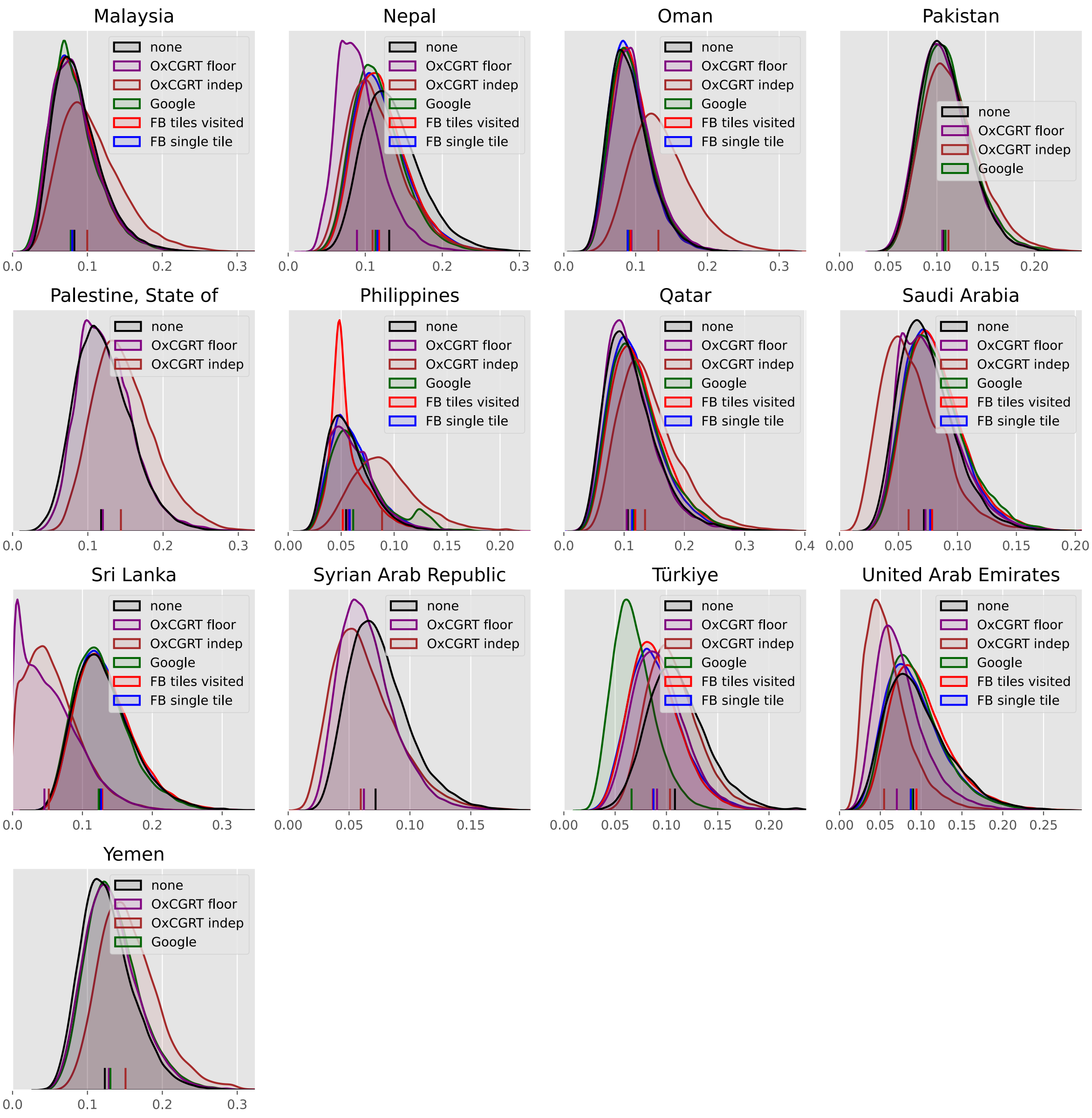
Malaysia
none
OxCGRT floor
OxCGRT indep
Google
FB tiles visited
FB single tile
0.0 0.1 0.2 0.3
Nepal
none
OxCGRT floor
OxCGRT indep
Google
FB tiles visited
FB single tile
0.0 0.1 0.2 0.3
Oman
none
OxCGRT floor
OxCGRT indep
Google
FB tiles visited
FB single tile
0.0 0.1 0.2 0.3
Pakistan
none
OxCGRT floor
OxCGRT indep
Google
0.00 0.05 0.10 0.15 0.20
Palestine, State of
none
OxCGRT floor
OxCGRT indep
0.0 0.1 0.2 0.3
Philippines
none
OxCGRT floor
OxCGRT indep
Google
FB tiles visited
FB single tile
0.00 0.05 0.10 0.15 0.20
Qatar
none
OxCGRT floor
OxCGRT indep
Google
FB tiles visited
FB single tile
0.0 0.1 0.2 0.3 0.4
Saudi Arabia
none
OxCGRT floor
OxCGRT indep
Google
FB tiles visited
FB single tile
0.00 0.05 0.10 0.15 0.20
Sri Lanka
none
OxCGRT floor
OxCGRT indep
Google
FB tiles visited
FB single tile
0.0 0.1 0.2 0.3
Syrian Arab Republic
none
OxCGRT floor
OxCGRT indep
0.00 0.05 0.10 0.15
Türkiye
none
OxCGRT floor
OxCGRT indep
Google
FB tiles visited
FB single tile
0.00 0.05 0.10 0.15 0.20
United Arab Emirates
none
OxCGRT floor
OxCGRT indep
Google
FB tiles visited
FB single tile
0.00 0.05 0.10 0.15 0.20 0.25
Yemen
none
OxCGRT floor
OxCGRT indep
Google
0.0 0.1 0.2 0.3

#### Europe

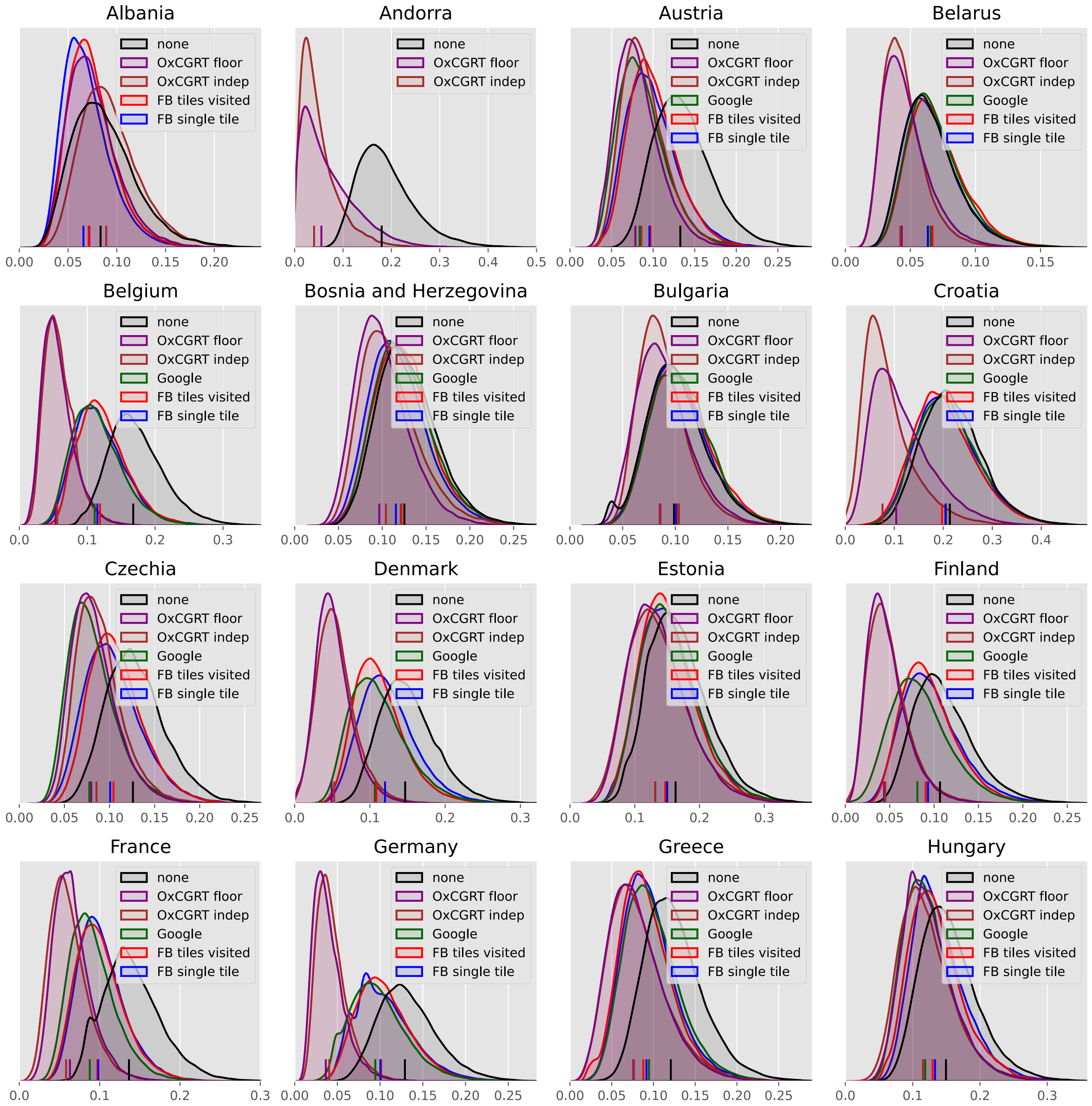
Albania
none
OxCGRT floor
OxCGRT indep
FB tiles visited
FB single tile
0.00 0.05 0.10 0.15 0.20
Andorra
none
OxCGRT floor
OxCGRT indep
0.0 0.1 0.2 0.3 0.4 0.5
Austria
none
OxCGRT floor
OxCGRT indep
Google
FB tiles visited
FB single tile
0.00 0.05 0.10 0.15 0.20 0.25
Belarus
none
OxCGRT floor
OxCGRT indep
Google
FB tiles visited
FB single tile
0.00 0.05 0.10 0.15
Belgium
none
OxCGRT floor
OxCGRT indep
Google
FB tiles visited
FB single tile
0.0 0.1 0.2 0.3
Bosnia and Herzegovina
none
OxCGRT floor
OxCGRT indep
Google
FB tiles visited
FB single tile
0.00 0.05 0.10 0.15 0.20 0.25
Bulgaria
none
OxCGRT floor
OxCGRT indep
Google
FB tiles visited
FB single tile
0.00 0.05 0.10 0.15 0.20
Croatia
none
OxCGRT floor
OxCGRT indep
Google
FB tiles visited
FB single tile
0.0 0.1 0.2 0.3 0.4
Czechia
none
OxCGRT floor
OxCGRT indep
Google
FB tiles visited
FB single tile
0.00 0.05 0.10 0.15 0.20 0.25
Denmark
none
OxCGRT floor
OxCGRT indep
Google
FB tiles visited
FB single tile
0.0 0.1 0.2 0.3
Estonia
none
OxCGRT floor
OxCGRT indep
Google
FB tiles visited
FB single tile
0.0 0.1 0.2 0.3
Finland
none
OxCGRT floor
OxCGRT indep
Google
FB tiles visited
FB single tile
0.00 0.05 0.10 0.15 0.20 0.25
France
none
OxCGRT floor
OxCGRT indep
Google
FB tiles visited
FB single tile
0.0 0.1 0.2 0.3
Germany
none
OxCGRT floor
OxCGRT indep
Google
FB tiles visited
FB single tile
0.00 0.05 0.10 0.15 0.20 0.25
Greece
none
OxCGRT floor
OxCGRT indep
Google
FB tiles visited
FB single tile
0.00 0.05 0.10 0.15 0.20 0.25
Hungary
none
OxCGRT floor
OxCGRT indep
Google
FB tiles visited
FB single tile
0.0 0.1 0.2 0.3

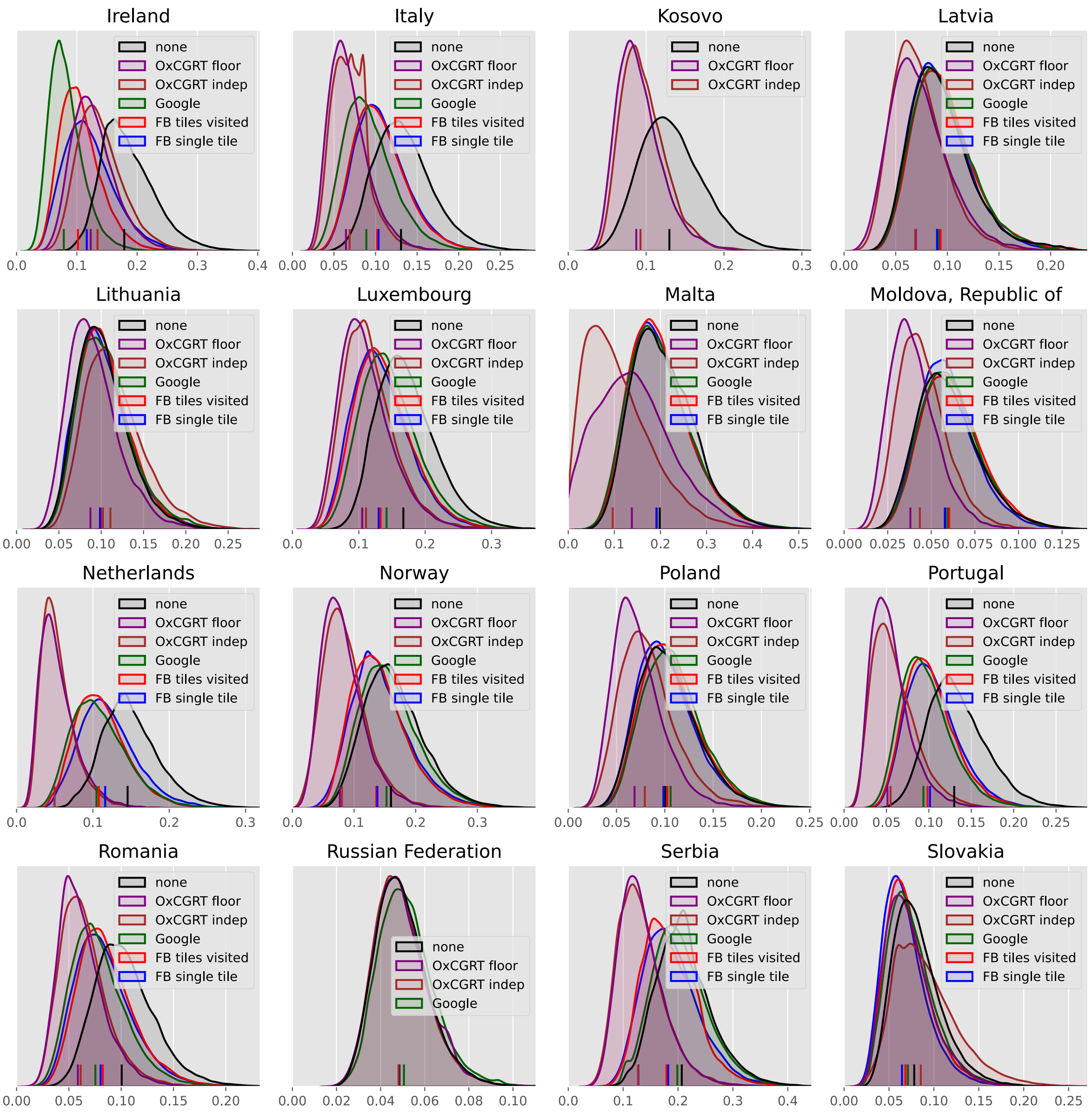
Ireland
Italy
Kosovo
Latvia
Lithuania
Luxembourg
Malta
Moldova, Republic of
Netherlands
Norway
Poland
Portugal
Romania
Russian Federation
Serbia
Slovakia
none
OxCGRT floor
OxCGRT indep
Google
FB tiles visited
FB single tile

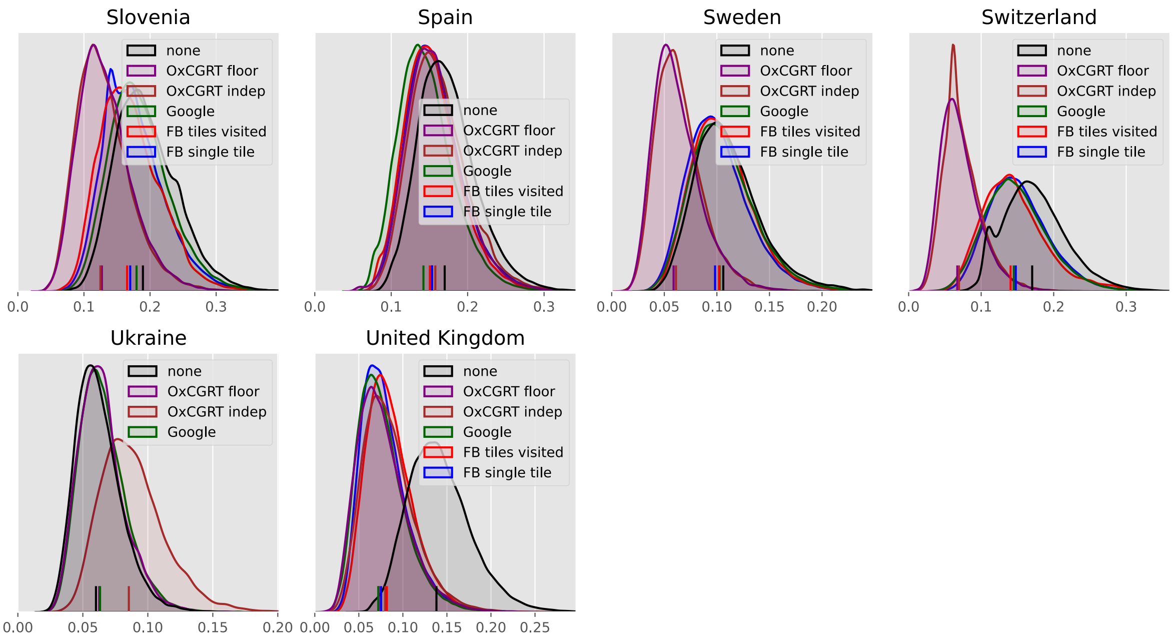

Slovenia
none
OxCGRT floor
OxCGRT indep
Google
FB tiles visited
FB single tile
0.0
0.1
0.2
0.3
Spain
none
OxCGRT floor
OxCGRT indep
Google
FB tiles visited
FB single tile
0.0
0.1
0.2
0.3
Sweden
none
OxCGRT floor
OxCGRT indep
Google
FB tiles visited
FB single tile
0.00
0.05
0.10
0.15
0.20
Switzerland
none
OxCGRT floor
OxCGRT indep
Google
FB tiles visited
FB single tile
0.0
0.1
0.2
0.3
Ukraine
none
OxCGRT floor
OxCGRT indep
Google
0.00
0.05
0.10
0.15
0.20
United Kingdom
none
OxCGRT floor
OxCGRT indep
Google
FB tiles visited
FB single tile
0.00
0.05
0.10
0.15
0.20
0.25

#### North America

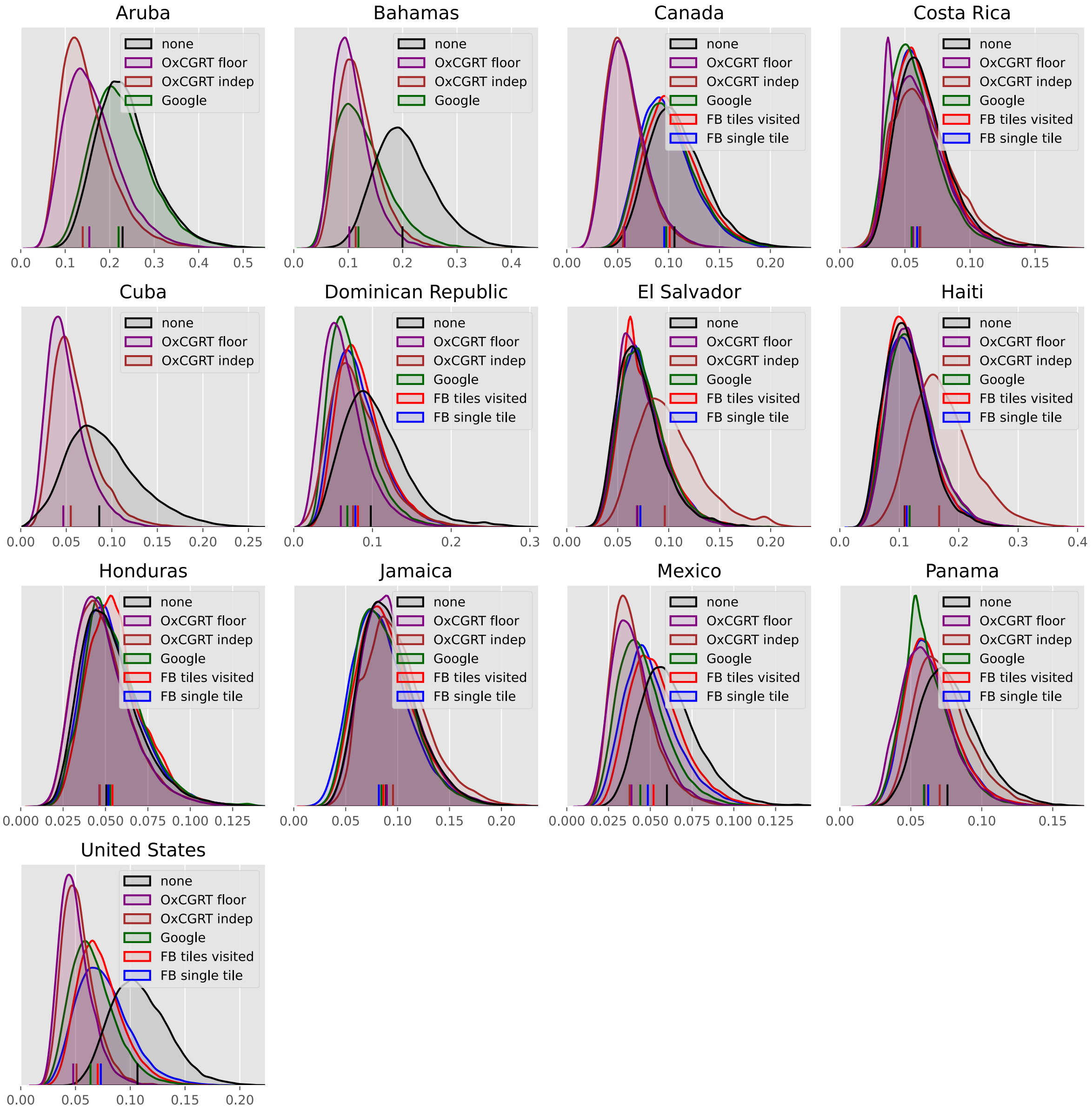

#### Oceania

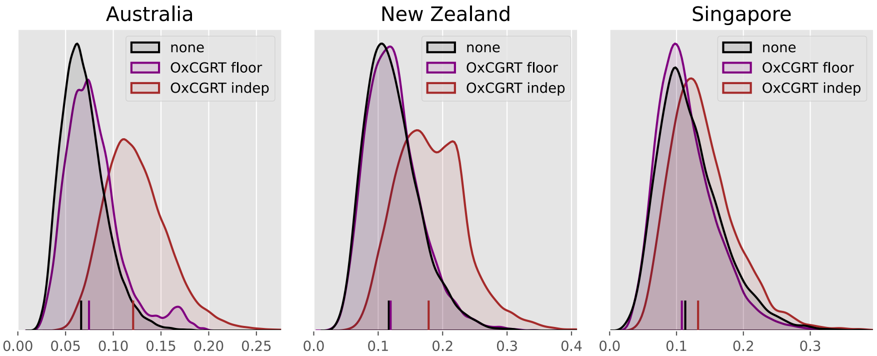

Australia
New Zealand
Singapore
none
OxCGRT floor
OxCGRT indep
0.00
0.05
0.10
0.15
0.20
0.25
0.0
0.1
0.2
0.3
0.4

#### South America

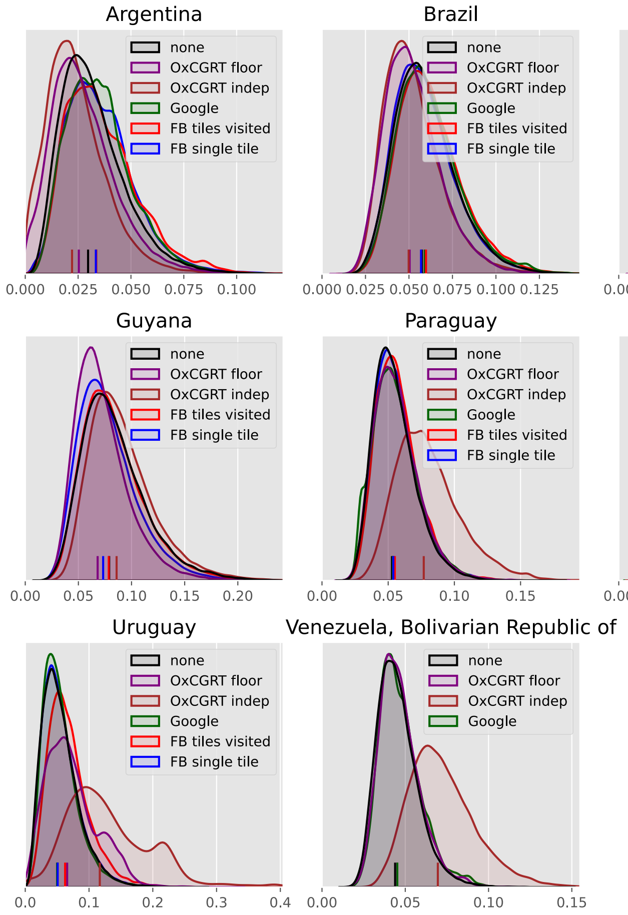

#### Proportion better than baseline

For each country and scaled analysis, we present a summary metric for the probability that inclusion of transmission scaling with policy changes or mobility improved the explanatory power of the model. This metric was created by randomly shuffling the baseline (`no_scaling`) posterior draws and then asking what proportion of comparison runs from the analysis considered have a lower dispersion than under baseline conditions. As such, values above 0.5 favour the analysis with scaling included, values below favour baseline.

| | OxCGRT floored | OxCGRT independent | Facebook tiles visited mobility | Facebook single tile mobility | Google mobility |
|---|---|---|---|---|---|
| Algeria | 0.461 | 0.293 | 0.568 | 0.625 | no analysis |
| Angola | 0.441 | 0.177 | 0.419 | 0.484 | 0.417 |
| Benin | 0.517 | 0.414 | 0.487 | 0.502 | 0.472 |
| Burkina Faso | 0.553 | 0.327 | 0.483 | 0.505 | 0.420 |
| Cabo Verde | 0.633 | 0.652 | no analysis | no analysis | 0.461 |
| Cameroon | 0.532 | 0.432 | 0.462 | 0.482 | 0.442 |
| Congo | 0.452 | 0.251 | 0.440 | 0.458 | no analysis |
| Côte d'Ivoire | 0.459 | 0.258 | 0.472 | 0.491 | 0.457 |
| Egypt | 0.444 | 0.281 | 0.448 | 0.480 | 0.444 |
| Eswatini | 0.769 | 0.670 | no analysis | no analysis | no analysis |
| Ethiopia | 0.508 | 0.260 | 0.480 | 0.534 | no analysis |
| Gabon | 0.484 | 0.345 | 0.441 | 0.464 | 0.433 |
| Guinea | 0.538 | 0.433 | 0.516 | 0.530 | no analysis |
| Kenya | 0.540 | 0.367 | 0.475 | 0.502 | 0.464 |
| Lesotho | 0.800 | 0.752 | 0.577 | 0.538 | no analysis |
| Liberia | 0.464 | 0.387 | 0.483 | 0.465 | no analysis |
| Libya | 0.483 | 0.231 | no analysis | no analysis | 0.430 |
| Madagascar | 0.529 | 0.332 | 0.473 | 0.509 | no analysis |
| Malawi | 0.631 | 0.691 | 0.534 | 0.510 | no analysis |
| Mali | 0.509 | 0.531 | 0.456 | 0.505 | 0.439 |
| Morocco | 0.483 | 0.464 | no analysis | no analysis | 0.426 |

| | OxCGRT floored | OxCGRT independent | Facebook tiles visited mobility | Facebook single tile mobility | Google mobility |
|---|---|---|---|---|---|
| Mozambique | 0.513 | 0.587 | 0.563 | 0.525 | 0.559 |
| Nigeria | 0.480 | 0.306 | 0.468 | 0.489 | 0.475 |
| Rwanda | 0.608 | 0.485 | 0.628 | 0.585 | 0.435 |
| Senegal | 0.450 | 0.247 | 0.462 | 0.477 | 0.506 |
| Somalia | 0.459 | 0.212 | no analysis | no analysis | no analysis |
| South Africa | 0.799 | 0.874 | 0.509 | 0.507 | 0.527 |
| South Sudan | 0.578 | 0.566 | no analysis | no analysis | no analysis |
| Sudan | 0.549 | 0.264 | no analysis | no analysis | no analysis |
| Togo | 0.455 | 0.169 | 0.452 | 0.494 | 0.431 |
| Tunisia | 0.892 | 0.721 | 0.719 | 0.625 | no analysis |
| Zambia | 0.477 | 0.323 | 0.557 | 0.511 | 0.484 |
| Zimbabwe | 0.788 | 0.789 | 0.655 | 0.601 | 0.466 |
| Afghanistan | 0.600 | 0.406 | no analysis | no analysis | 0.477 |
| Azerbaijan | 0.725 | 0.776 | no analysis | no analysis | no analysis |
| Bahrain | 0.474 | 0.256 | no analysis | no analysis | 0.452 |
| Bangladesh | 0.461 | 0.266 | 0.583 | 0.543 | 0.596 |
| Cyprus | 0.883 | 0.850 | no analysis | no analysis | no analysis |
| Georgia | 0.521 | 0.226 | 0.477 | 0.484 | 0.450 |
| India | 0.562 | 0.399 | 0.501 | 0.520 | 0.510 |
| Indonesia | 0.481 | 0.291 | 0.453 | 0.485 | 0.481 |
| Iran, Islamic Republic of | 0.810 | 0.767 | no analysis | no analysis | no analysis |
| Iraq | 0.470 | 0.187 | 0.432 | 0.467 | 0.427 |
| Israel | 0.832 | 0.946 | no analysis | no analysis | 0.859 |
| Japan | 0.891 | 0.957 | 0.592 | 0.539 | 0.596 |
| Jordan | 0.520 | 0.314 | 0.469 | 0.511 | 0.494 |
| Korea, Republic of | 0.584 | 0.575 | 0.595 | 0.572 | 0.528 |
| Kuwait | 0.714 | 0.788 | no analysis | no analysis | 0.557 |
| Lebanon | 0.537 | 0.360 | 0.497 | 0.526 | 0.454 |
| Malaysia | 0.524 | 0.368 | 0.527 | 0.523 | 0.540 |
| Nepal | 0.814 | 0.656 | 0.602 | 0.614 | 0.639 |
| Oman | 0.443 | 0.191 | 0.459 | 0.499 | 0.470 |

| | OxCGRT floored | OxCGRT independent | Facebook tiles visited mobility | Facebook single tile mobility | Google mobility |
|---|---|---|---|---|---|
| Pakistan | 0.511 | 0.441 | no analysis | no analysis | 0.476 |
| Palestine, State of | 0.480 | 0.310 | no analysis | no analysis | no analysis |
| Philippines | 0.468 | 0.169 | 0.508 | 0.450 | 0.402 |
| Qatar | 0.501 | 0.315 | 0.418 | 0.454 | 0.435 |
| Saudi Arabia | 0.483 | 0.651 | 0.417 | 0.440 | 0.424 |
| Sri Lanka | 0.906 | 0.902 | 0.483 | 0.494 | 0.520 |
| Syrian Arab Republic | 0.614 | 0.630 | no analysis | no analysis | no analysis |
| Türkiye | 0.678 | 0.544 | 0.715 | 0.704 | 0.889 |
| United Arab Emirates | 0.661 | 0.789 | 0.470 | 0.519 | 0.508 |
| Yemen | 0.460 | 0.284 | no analysis | no analysis | 0.440 |
| Albania | 0.613 | 0.443 | 0.617 | 0.669 | no analysis |
| Andorra | 0.925 | 0.973 | no analysis | no analysis | no analysis |
| Austria | 0.897 | 0.861 | 0.789 | 0.789 | 0.861 |
| Belarus | 0.778 | 0.805 | 0.448 | 0.499 | 0.464 |
| Belgium | 0.994 | 0.995 | 0.827 | 0.834 | 0.855 |
| Bosnia and Herzegovina | 0.739 | 0.670 | 0.535 | 0.580 | 0.533 |
| Bulgaria | 0.629 | 0.644 | 0.456 | 0.478 | 0.457 |
| Croatia | 0.878 | 0.943 | 0.569 | 0.542 | 0.534 |
| Czechia | 0.871 | 0.840 | 0.686 | 0.712 | 0.873 |
| Denmark | 0.988 | 0.983 | 0.784 | 0.697 | 0.776 |
| Estonia | 0.690 | 0.685 | 0.602 | 0.579 | 0.579 |
| Finland | 0.949 | 0.943 | 0.644 | 0.619 | 0.715 |
| France | 0.962 | 0.967 | 0.805 | 0.797 | 0.860 |
| Germany | 0.992 | 0.991 | 0.728 | 0.726 | 0.772 |
| Greece | 0.826 | 0.815 | 0.766 | 0.739 | 0.712 |
| Hungary | 0.714 | 0.725 | 0.637 | 0.607 | 0.715 |
| Ireland | 0.853 | 0.789 | 0.928 | 0.853 | 0.979 |
| Italy | 0.944 | 0.944 | 0.729 | 0.716 | 0.816 |
| Kosovo | 0.804 | 0.769 | no analysis | no analysis | no analysis |
| Latvia | 0.704 | 0.715 | 0.469 | 0.502 | 0.478 |
| Lithuania | 0.608 | 0.392 | 0.473 | 0.496 | 0.473 |
| Luxembourg | 0.851 | 0.840 | 0.713 | 0.729 | 0.662 |

| | OxCGRT floored | OxCGRT independent | Facebook tiles visited mobility | Facebook single tile mobility | Google mobility |
|---|---|---|---|---|---|
| Malta | 0.722 | 0.835 | 0.530 | 0.528 | 0.525 |
| Moldova, Republic of | 0.825 | 0.750 | 0.463 | 0.506 | 0.477 |
| Netherlands | 0.984 | 0.987 | 0.782 | 0.719 | 0.786 |
| Norway | 0.923 | 0.916 | 0.644 | 0.627 | 0.544 |
| Poland | 0.791 | 0.692 | 0.479 | 0.516 | 0.450 |
| Portugal | 0.970 | 0.959 | 0.763 | 0.740 | 0.793 |
| Romania | 0.885 | 0.868 | 0.679 | 0.698 | 0.743 |
| Russian Federation | 0.500 | 0.508 | no analysis | no analysis | 0.454 |
| Serbia | 0.884 | 0.891 | 0.649 | 0.632 | 0.540 |
| Slovakia | 0.610 | 0.433 | 0.609 | 0.652 | 0.576 |
| Slovenia | 0.847 | 0.857 | 0.647 | 0.622 | 0.562 |
| Spain | 0.629 | 0.591 | 0.650 | 0.625 | 0.708 |
| Sweden | 0.900 | 0.895 | 0.542 | 0.578 | 0.539 |
| Switzerland | 0.980 | 0.977 | 0.688 | 0.648 | 0.662 |
| Ukraine | 0.467 | 0.185 | no analysis | no analysis | 0.446 |
| United Kingdom | 0.927 | 0.900 | 0.902 | 0.925 | 0.935 |
| Aruba | 0.800 | 0.848 | no analysis | no analysis | 0.540 |
| Bahamas | 0.941 | 0.908 | no analysis | no analysis | 0.861 |
| Canada | 0.926 | 0.934 | 0.547 | 0.603 | 0.583 |
| Costa Rica | 0.579 | 0.500 | 0.530 | 0.533 | 0.594 |
| Cuba | 0.818 | 0.751 | no analysis | no analysis | no analysis |
| Dominican Republic | 0.797 | 0.672 | 0.629 | 0.652 | 0.739 |
| El Salvador | 0.497 | 0.230 | 0.456 | 0.463 | 0.463 |
| Haiti | 0.431 | 0.162 | 0.481 | 0.477 | 0.438 |
| Honduras | 0.570 | 0.565 | 0.441 | 0.473 | 0.458 |
| Jamaica | 0.493 | 0.436 | 0.527 | 0.575 | 0.548 |
| Mexico | 0.842 | 0.857 | 0.637 | 0.689 | 0.764 |
| Panama | 0.729 | 0.574 | 0.692 | 0.693 | 0.727 |
| United States | 0.979 | 0.973 | 0.862 | 0.824 | 0.899 |
| Australia | 0.403 | 0.079 | no analysis | no analysis | no analysis |
| New Zealand | 0.486 | 0.184 | no analysis | no analysis | no analysis |
| Singapore | 0.534 | 0.381 | no analysis | no analysis | no analysis |

| | OxCGRT floored | OxCGRT independent | Facebook tiles visited mobility | Facebook single tile mobility | Google mobility |
|---|---|---|---|---|---|
| Argentina | 0.591 | 0.657 | 0.429 | 0.436 | 0.438 |
| Brazil | 0.620 | 0.626 | 0.462 | 0.509 | 0.473 |
| Chile | 0.561 | 0.442 | 0.548 | 0.678 | 0.580 |
| Colombia | 0.632 | 0.546 | 0.508 | 0.485 | 0.506 |
| Guyana | 0.615 | 0.429 | 0.503 | 0.560 | no analysis |
| Paraguay | 0.476 | 0.188 | 0.459 | 0.481 | 0.492 |
| Peru | 0.621 | 0.561 | 0.673 | 0.680 | 0.723 |
| Suriname | 0.503 | 0.381 | 0.617 | 0.601 | no analysis |
| Uruguay | 0.372 | 0.134 | 0.381 | 0.492 | 0.506 |
| Venezuela, Bolivarian Republic of | 0.468 | 0.114 | no analysis | no analysis | 0.465 |

#### Proportion floored better than independent

For each country and scaled analysis, we present the same summary metric, but for the probability that the OxCGRT floored configuration performed better than the OxCGRT independent approach for the same country.

For 76.2% of countries, the floored configuration performed better in most calibration iterations.

| | Probability OxCGRT floored better than independent |
|---|---|
| Algeria | 0.686 |
| Angola | 0.793 |
| Benin | 0.605 |
| Burkina Faso | 0.709 |
| Cabo Verde | 0.488 |
| Cameroon | 0.599 |
| Congo | 0.708 |
| Côte d'Ivoire | 0.709 |
| Egypt | 0.670 |
| Eswatini | 0.591 |
| Ethiopia | 0.747 |
| Gabon | 0.638 |

| | Probability OxCGRT floored better than independent |
|---|---|
| Guinea | 0.602 |
| Kenya | 0.678 |
| Lesotho | 0.559 |
| Liberia | 0.575 |
| Libya | 0.759 |
| Madagascar | 0.693 |
| Malawi | 0.438 |
| Mali | 0.467 |
| Morocco | 0.512 |
| Mozambique | 0.430 |
| Nigeria | 0.680 |
| Rwanda | 0.624 |
| Senegal | 0.720 |
| Somalia | 0.755 |
| South Africa | 0.352 |
| South Sudan | 0.503 |
| Sudan | 0.789 |
| Togo | 0.800 |
| Tunisia | 0.695 |
| Zambia | 0.655 |
| Zimbabwe | 0.492 |
| Afghanistan | 0.684 |
| Azerbaijan | 0.434 |
| Bahrain | 0.730 |
| Bangladesh | 0.704 |
| Cyprus | 0.539 |
| Georgia | 0.784 |
| India | 0.656 |
| Indonesia | 0.692 |
| Iran, Islamic Republic of | 0.570 |
| Iraq | 0.804 |
| Israel | 0.225 |
| Japan | 0.388 |
| Jordan | 0.701 |
| Korea, Republic of | 0.511 |
| Kuwait | 0.398 |
| Lebanon | 0.667 |
| Malaysia | 0.665 |
| Nepal | 0.673 |
| Oman | 0.782 |
| Pakistan | 0.565 |

| | Probability OxCGRT floored better than independent |
|---|---|
| Palestine, State of | 0.673 |
| Philippines | 0.813 |
| Qatar | 0.687 |
| Saudi Arabia | 0.341 |
| Sri Lanka | 0.547 |
| Syrian Arab Republic | 0.473 |
| Türkiye | 0.645 |
| United Arab Emirates | 0.337 |
| Yemen | 0.675 |
| Albania | 0.674 |
| Andorra | 0.410 |
| Austria | 0.583 |
| Belarus | 0.477 |
| Belgium | 0.529 |
| Bosnia and Herzegovina | 0.577 |
| Bulgaria | 0.494 |
| Croatia | 0.352 |
| Czechia | 0.568 |
| Denmark | 0.536 |
| Estonia | 0.496 |
| Finland | 0.518 |
| France | 0.442 |
| Germany | 0.568 |
| Greece | 0.506 |
| Hungary | 0.473 |
| Ireland | 0.588 |
| Italy | 0.539 |
| Kosovo | 0.558 |
| Latvia | 0.494 |
| Lithuania | 0.708 |
| Luxembourg | 0.540 |
| Malta | 0.373 |
| Moldova, Republic of | 0.616 |
| Netherlands | 0.502 |
| Norway | 0.530 |
| Poland | 0.607 |
| Portugal | 0.547 |
| Romania | 0.536 |
| Russian Federation | 0.495 |
| Serbia | 0.489 |
| Slovakia | 0.658 |

| | Probability OxCGRT floored better than independent |
|---|---|
| Slovenia | 0.487 |
| Spain | 0.534 |
| Sweden | 0.523 |
| Switzerland | 0.538 |
| Ukraine | 0.790 |
| United Kingdom | 0.573 |
| Aruba | 0.434 |
| Bahamas | 0.598 |
| Canada | 0.477 |
| Costa Rica | 0.576 |
| Cuba | 0.614 |
| Dominican Republic | 0.647 |
| El Salvador | 0.770 |
| Haiti | 0.789 |
| Honduras | 0.503 |
| Jamaica | 0.566 |
| Mexico | 0.482 |
| Panama | 0.664 |
| United States | 0.560 |
| Australia | 0.862 |
| New Zealand | 0.808 |
| Singapore | 0.654 |
| Argentina | 0.435 |
| Brazil | 0.494 |
| Chile | 0.615 |
| Colombia | 0.587 |
| Guyana | 0.689 |
| Paraguay | 0.799 |
| Peru | 0.567 |
| Suriname | 0.610 |
| Uruguay | 0.775 |
| Venezuela, Bolivarian Republic of | 0.882 |
