## Supplemental File 6 for "Quantifying the contribution of policy measures to COVID-19 dynamics across 126 countries"

### Comparison between baseline residual transmission scaling and fitted OxCGRT scaling

**Commits used for analyses** 12

#### Purpose

This document presents the residual transmission scaling (with uncertainty) implemented without scaling for policy effects for each country. It is intended to provide a raw comparison between the residual non-mechanistic transmission that needed to be applied for each analysis in the absence of mobility scaling and the various mobility data fields available from both Google and Facebook. We compare this against the policy scaling function, incorporating the fitted weights, flooring and exponentiation (with associated credible intervals).

### Weighted scaling process versus residual comparison

#### Africa

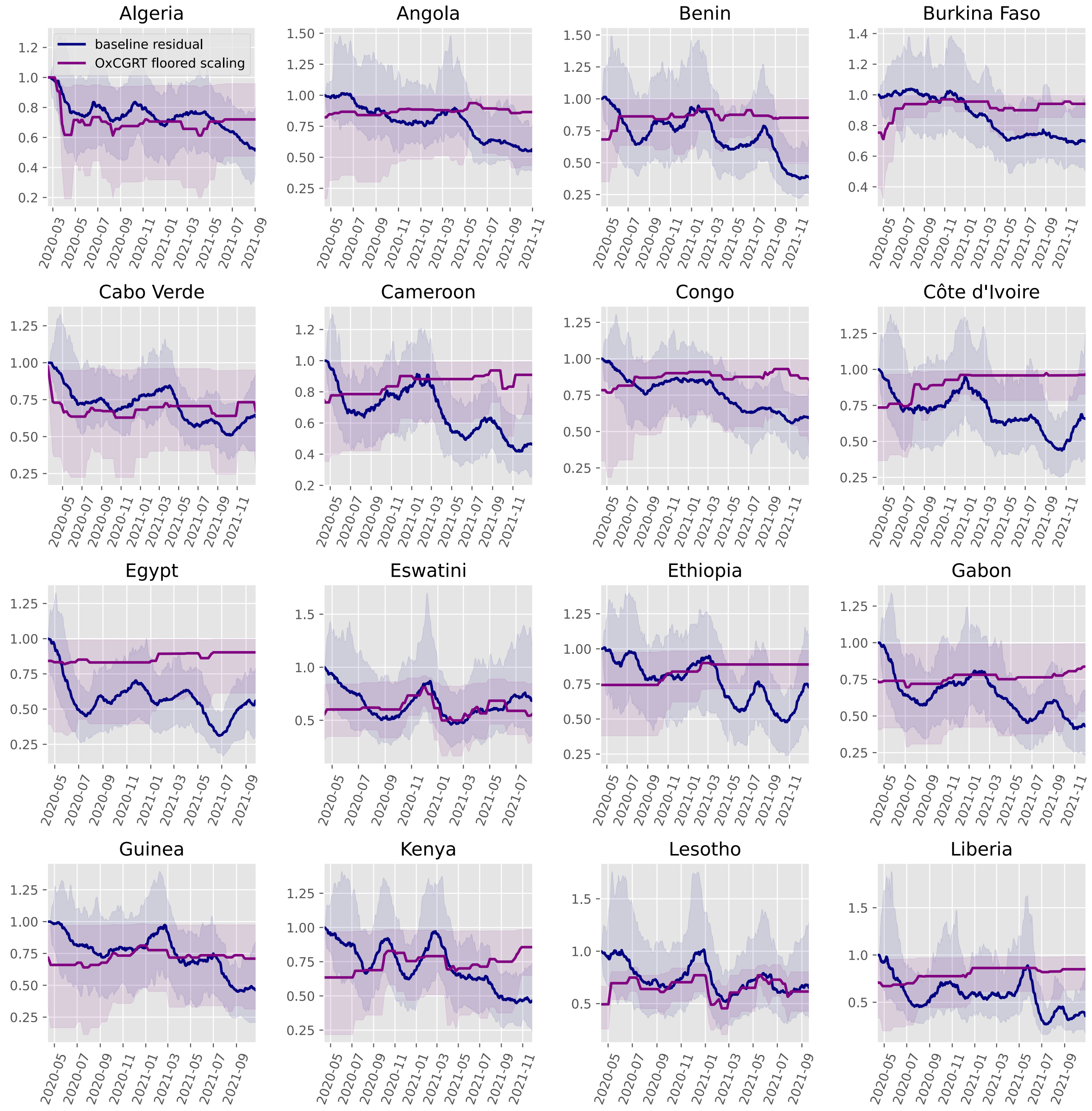

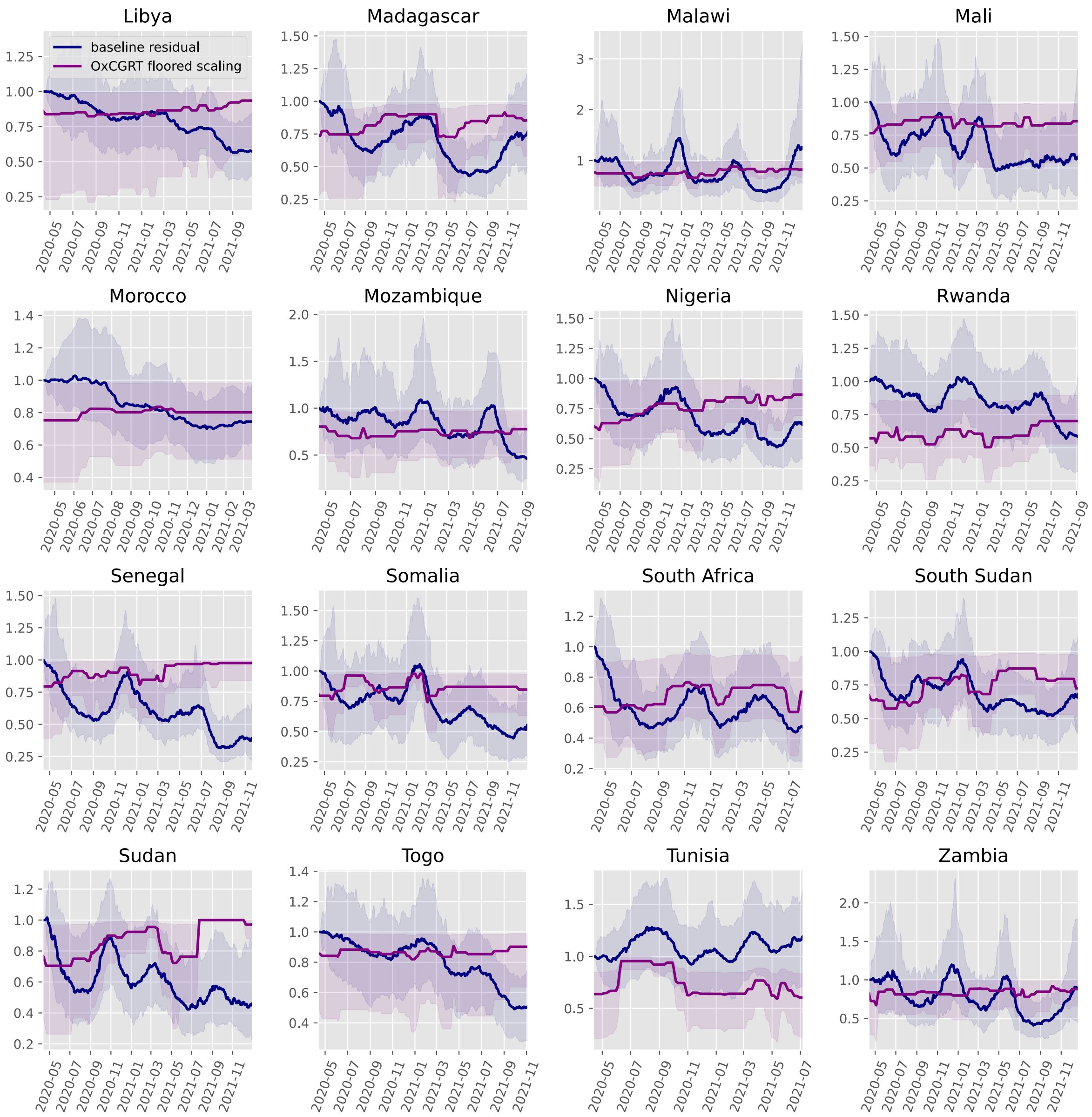
Libya
baseline residual
OxCGRT floored scaling
Madagascar
Malawi
Mali
Morocco
Mozambique
Nigeria
Rwanda
Senegal
Somalia
South Africa
South Sudan
Sudan
Togo
Tunisia
Zambia

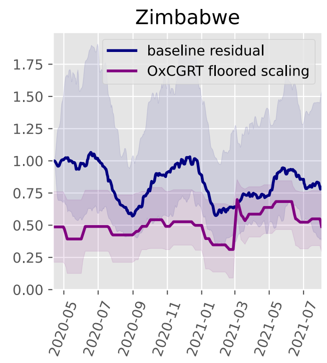

Zimbabwe
baseline residual
OxCGRT floored scaling
1.75
1.50
1.25
1.00
0.75
0.50
0.25
0.00
2020-05
2020-07
2020-09
2020-11
2021-01
2021-03
2021-05
2021-07

#### Asia

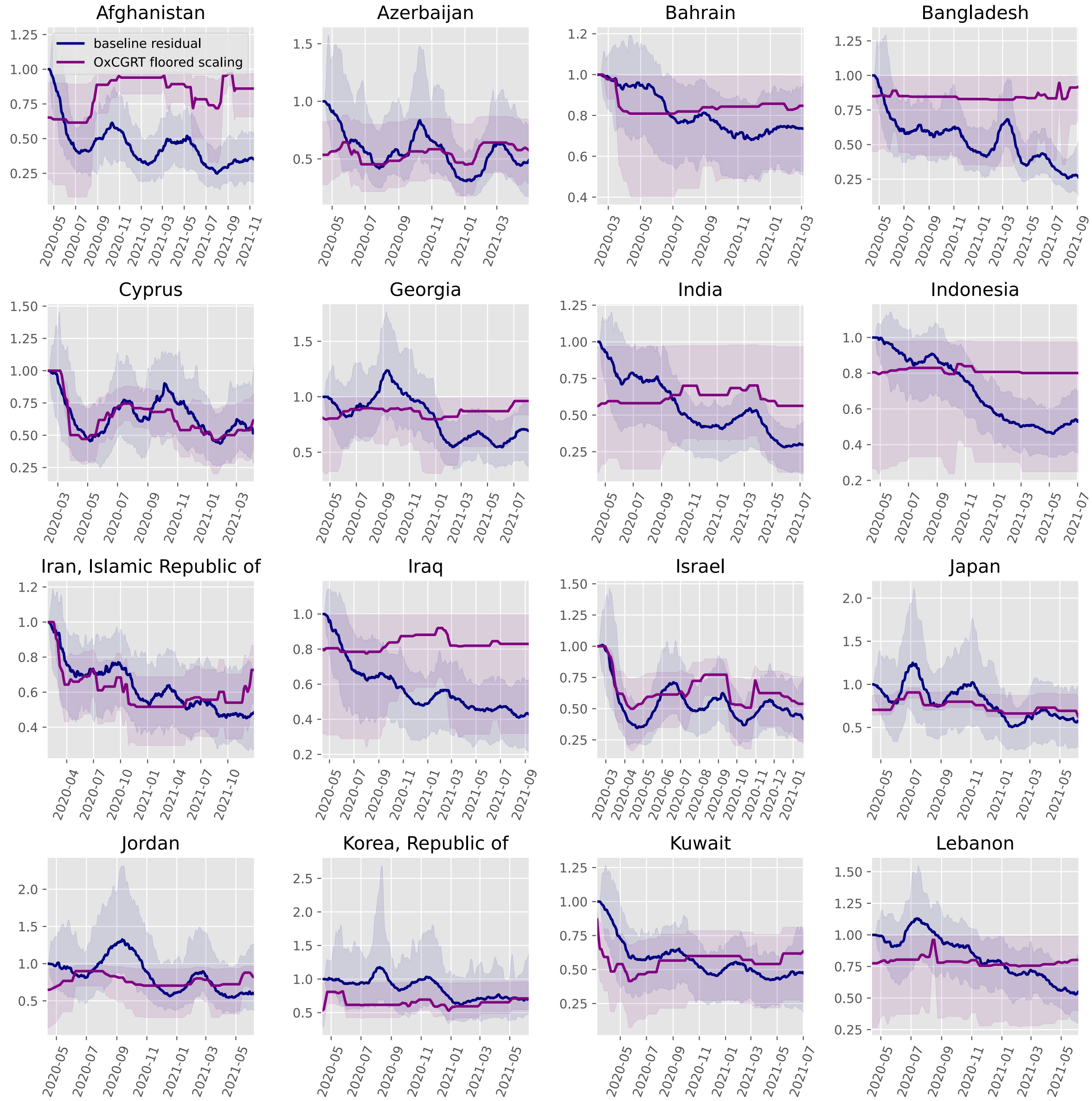

Afghanistan
baseline residual
OxCGRT floored scaling
Azerbaijan
Bahrain
Bangladesh
Cyprus
Georgia
India
Indonesia
Iran, Islamic Republic of
Iraq
Israel
Japan
Jordan
Korea, Republic of
Kuwait
Lebanon

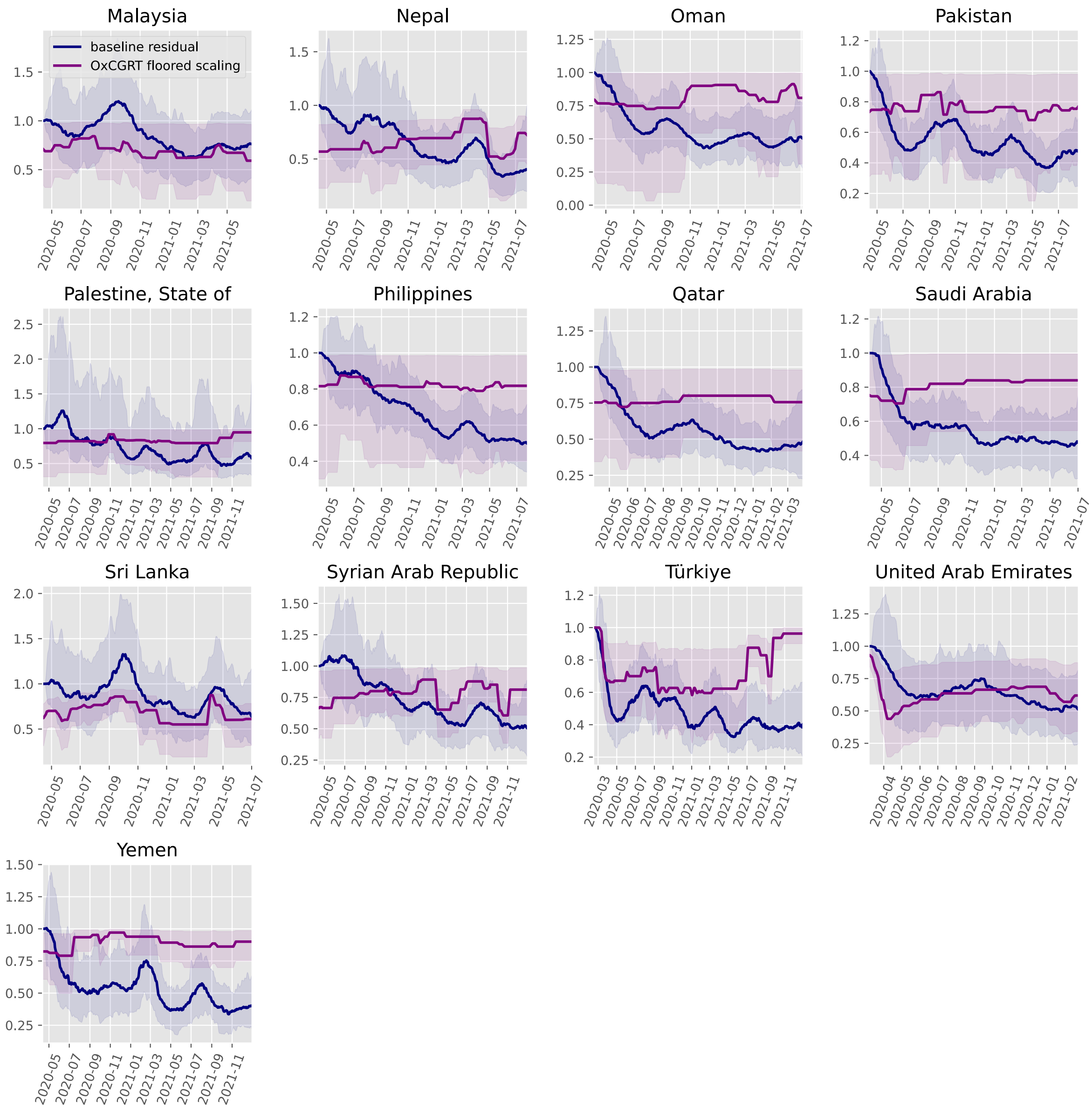
Malaysia
baseline residual
OxCGRT floored scaling
Nepal
Oman
Pakistan
Palestine, State of
Philippines
Qatar
Saudi Arabia
Sri Lanka
Syrian Arab Republic
Türkiye
United Arab Emirates
Yemen

#### Europe

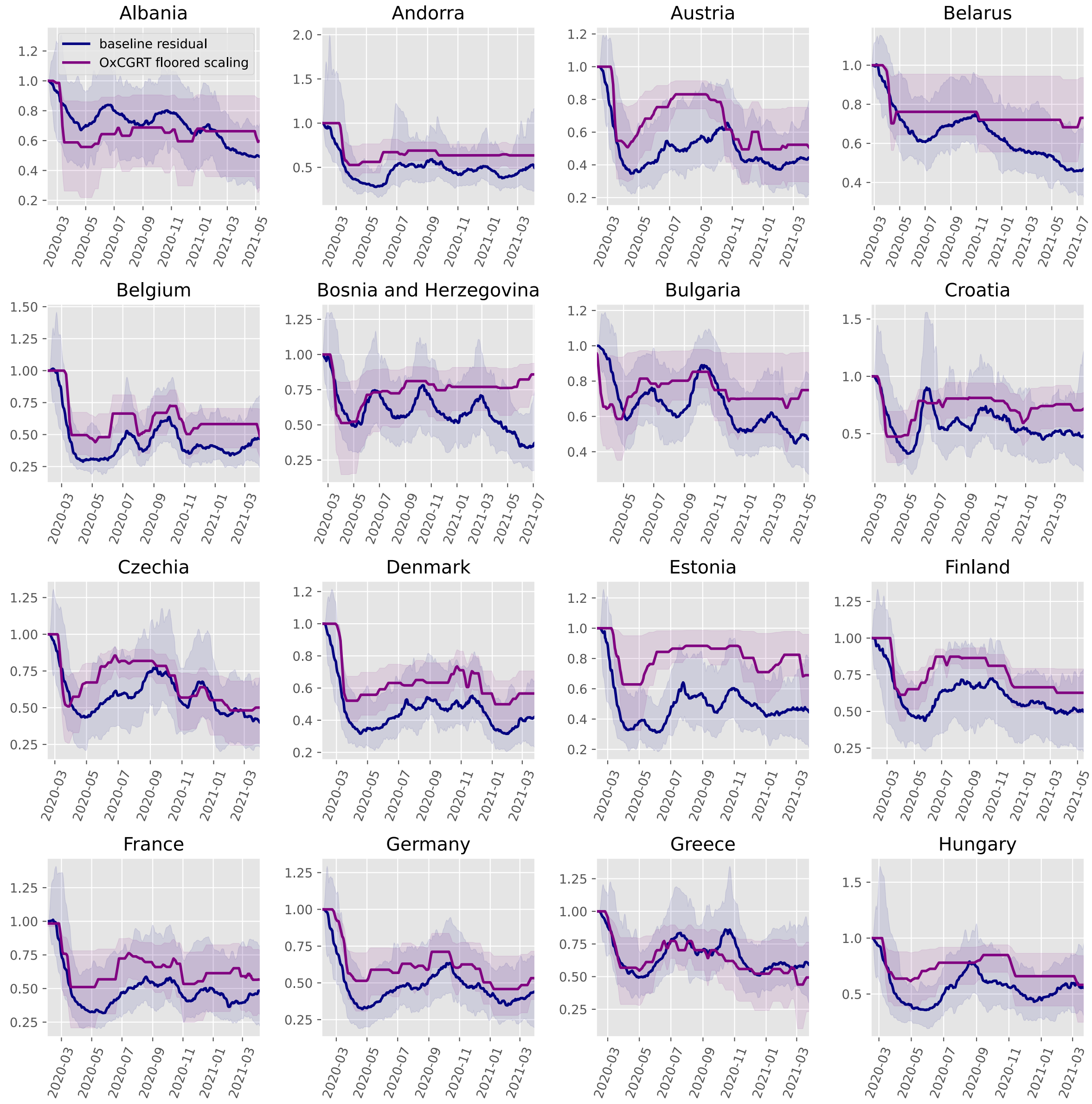
Albania
Andorra
Austria
Belarus
baseline residual
OxCGRT floored scaling
Belgium
Bosnia and Herzegovina
Bulgaria
Croatia
Czechia
Denmark
Estonia
Finland
France
Germany
Greece
Hungary

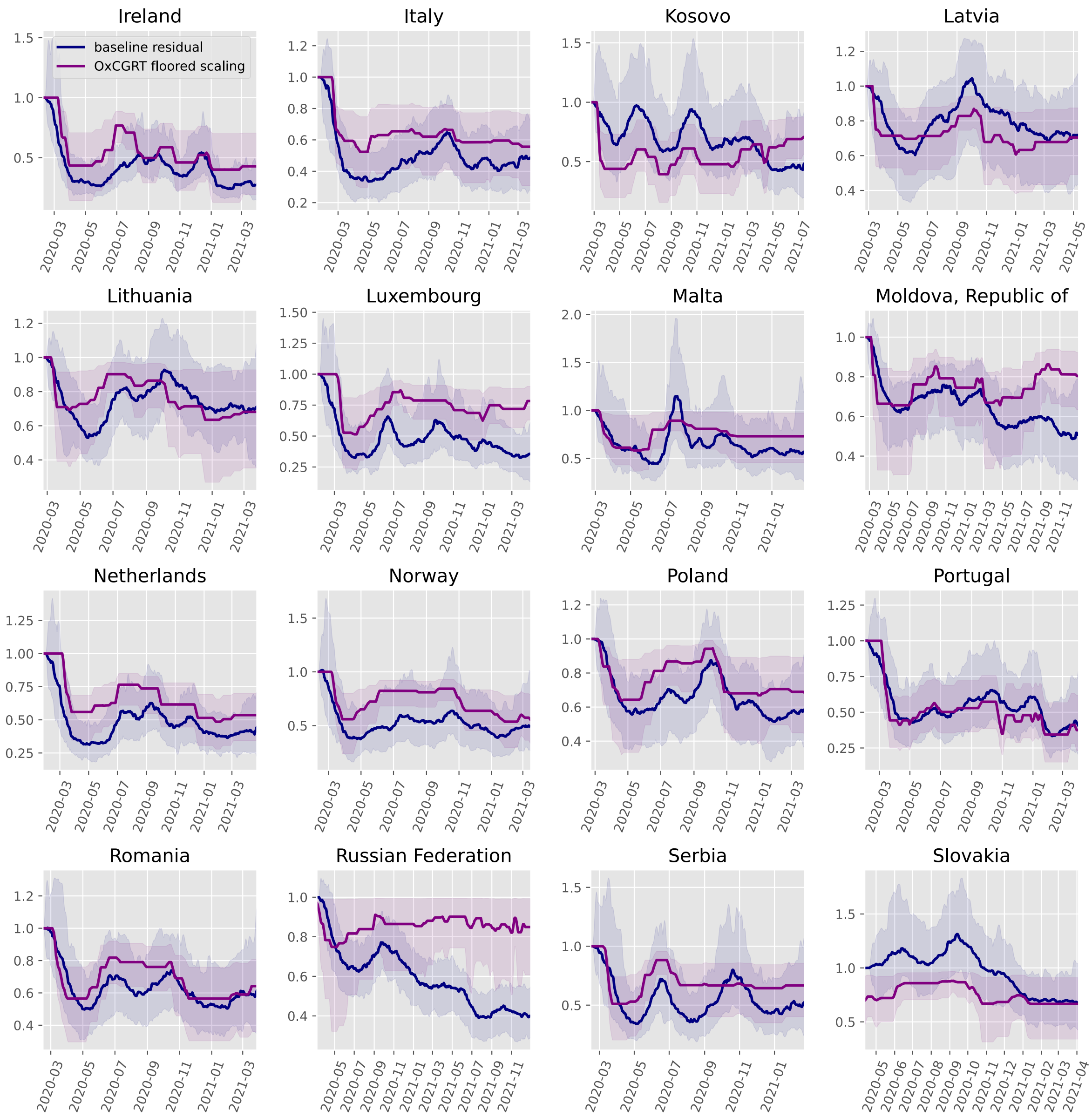
Ireland
Italy
Kosovo
Latvia
Lithuania
Luxembourg
Malta
Moldova, Republic of
Netherlands
Norway
Poland
Portugal
Romania
Russian Federation
Serbia
Slovakia
baseline residual
OxCGRT floored scaling

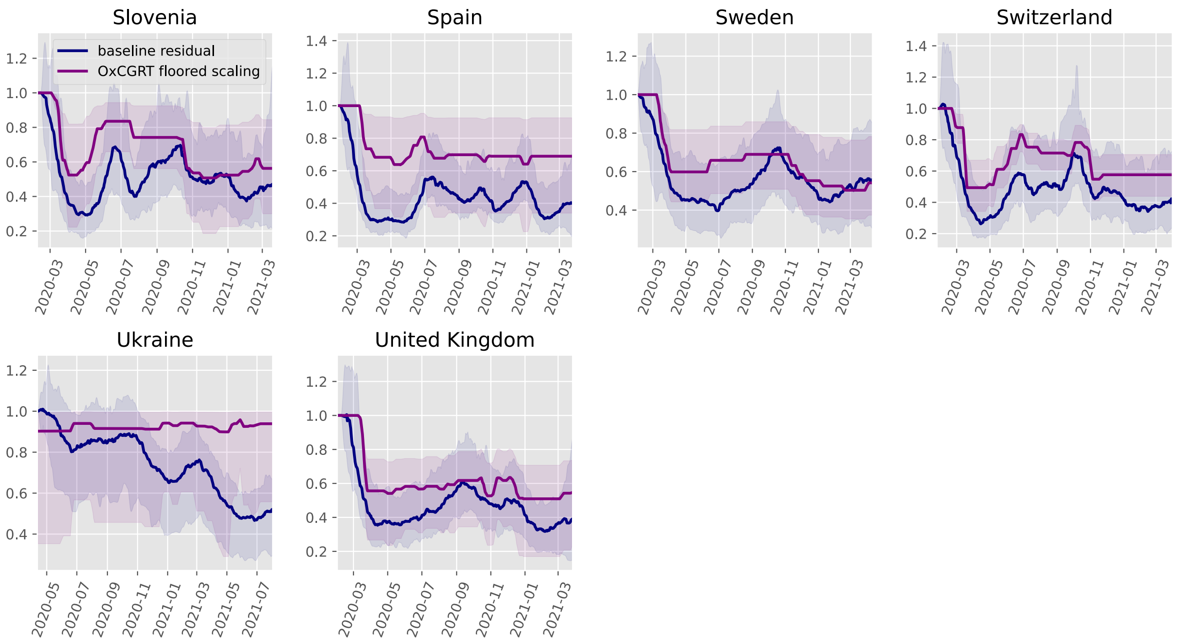
Slovenia
baseline residual
OxCGRT floored scaling
Spain
Sweden
Switzerland
Ukraine
United Kingdom

#### North America

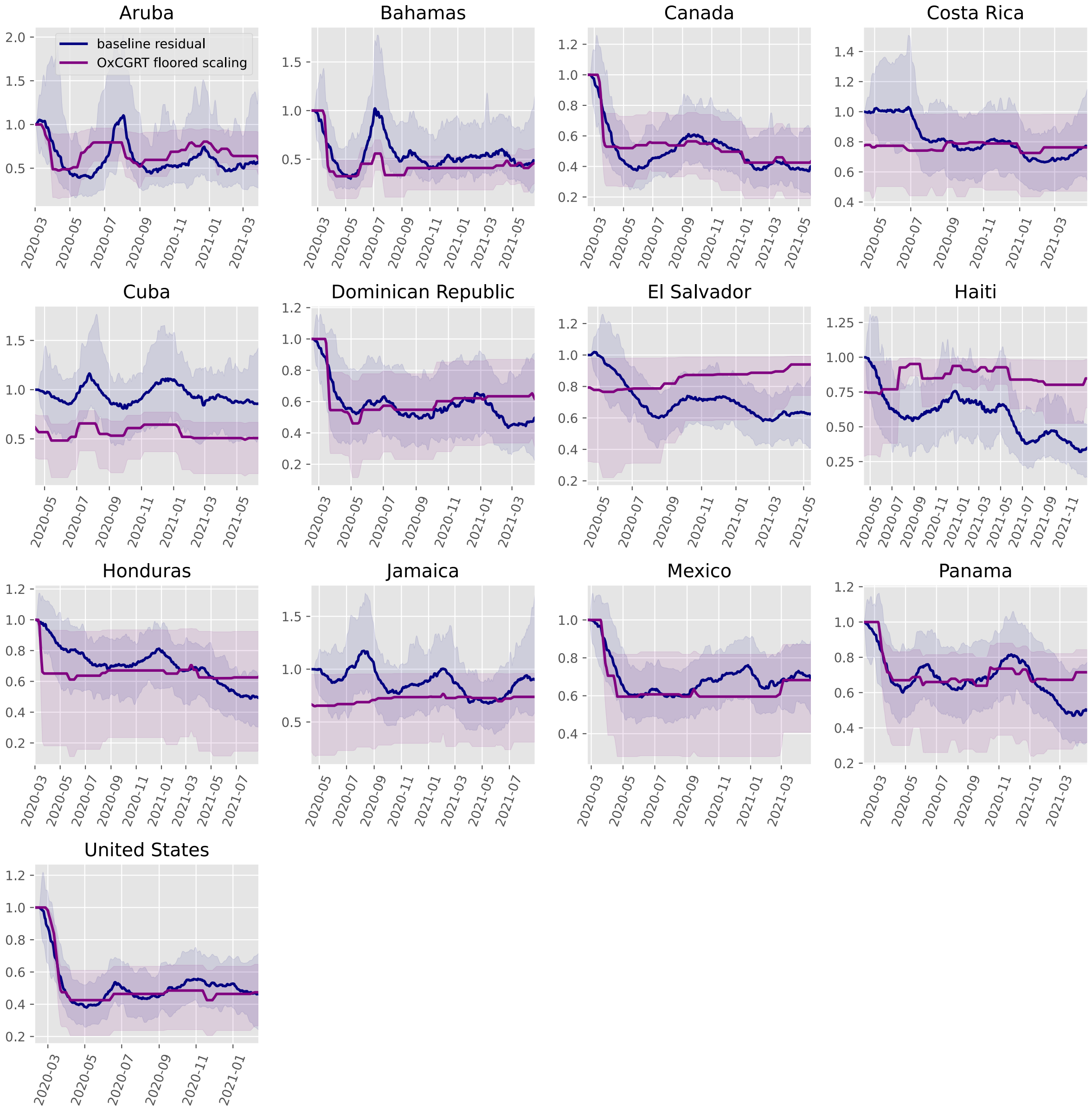

#### Oceania

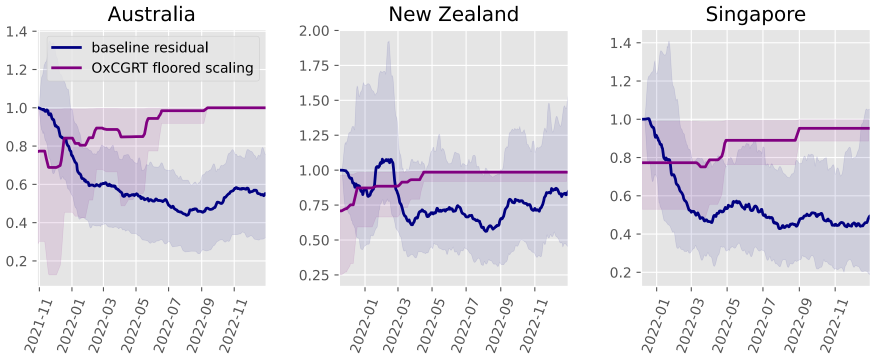

#### South America

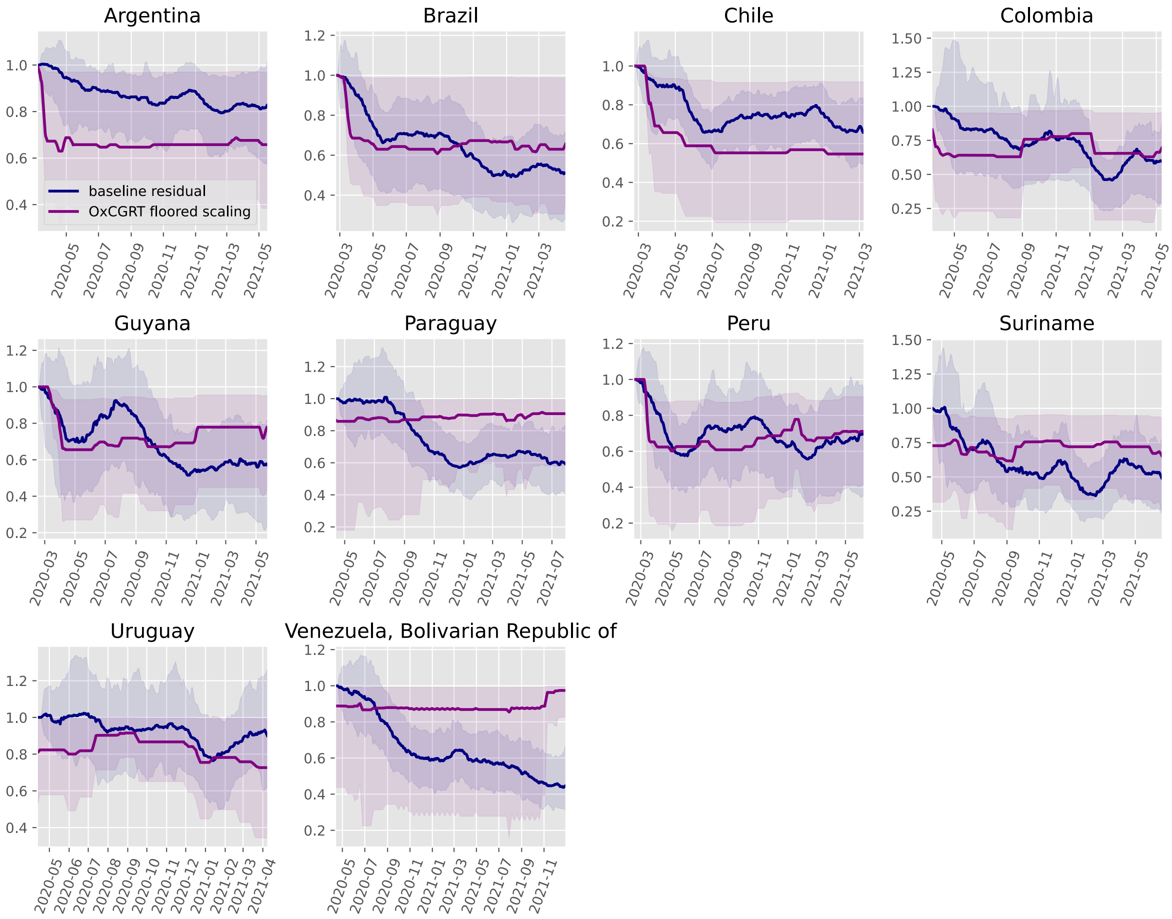

#### Commits used for analyses

For reproducibility, the following table gives the (short) commit SHA for each analysis.

| | no scaling | OxCGRT floored | OxCGRT independent | Google mobility | Facebook tiles visited mobility | Facebook single tile mobility |
|---|---|---|---|---|---|---|
| Afghanistan | 4f57571 | b934196 | b934196 | b934196 | no analysis | no analysis |
| Albania | 4f57571 | b934196 | b934196 | no analysis | b934196 | b934196 |
| Algeria | 4f57571 | b934196 | b934196 | no analysis | b934196 | b934196 |
| Andorra | 4f57571 | b934196 | b934196 | no analysis | no analysis | no analysis |
| Angola | 4f57571 | b934196 | b934196 | cb8324a | cb8324a | cb8324a |
| Argentina | 4f57571 | b934196 | b934196 | b934196 | b934196 | b934196 |
| Aruba | 4f57571 | b934196 | b934196 | 4f57571 | no analysis | no analysis |
| Australia | b934196 | b934196 | b934196 | no analysis | no analysis | no analysis |
| Austria | 4f57571 | b934196 | b934196 | b934196 | b934196 | b934196 |
| Azerbaijan | 4f57571 | b934196 | b934196 | no analysis | no analysis | no analysis |
| Bahamas | 4f57571 | b934196 | b934196 | b934196 | no analysis | no analysis |
| Bahrain | 4f57571 | b934196 | b934196 | 4f57571 | no analysis | no analysis |
| Bangladesh | 4f57571 | b934196 | b934196 | b934196 | b934196 | b934196 |
| Belarus | 4f57571 | b934196 | b934196 | cb8324a | cb8324a | b934196 |
| Belgium | 4f57571 | b934196 | b934196 | 4f57571 | 4f57571 | 4f57571 |
| Benin | 4f57571 | b934196 | b934196 | 4f57571 | 4f57571 | b934196 |
| Bosnia and Herzegovina | 4f57571 | b934196 | b934196 | b934196 | b934196 | b934196 |
| Brazil | 4f57571 | b934196 | b934196 | 4f57571 | 4f57571 | 4f57571 |
| Bulgaria | 4f57571 | b934196 | b934196 | b934196 | b934196 | b934196 |
| Burkina Faso | 4f57571 | b934196 | b934196 | 4f57571 | 4f57571 | 4f57571 |
| Cabo Verde | 4f57571 | b934196 | b934196 | 4f57571 | no analysis | no analysis |
| Cameroon | 4f57571 | b934196 | b934196 | b934196 | b934196 | b934196 |
| Canada | 4f57571 | b934196 | b934196 | b934196 | b934196 | b934196 |
| Chile | 4f57571 | b934196 | b934196 | 4f57571 | b934196 | b934196 |
| Colombia | 4f57571 | b934196 | b934196 | 4f57571 | 4f57571 | b934196 |
| Congo | 4f57571 | b934196 | b934196 | no analysis | 4f57571 | b934196 |
| Costa Rica | 4f57571 | b934196 | b934196 | 4f57571 | b934196 | b934196 |
| Croatia | 4f57571 | b934196 | b934196 | b934196 | b934196 | b934196 |
