## Supplemental File 7 for "Quantifying the contribution of policy measures to COVID-19 dynamics across 126 countries"

### Comparisons of residual transmission scaling under each OxCGRT configuration

**Commits used for analyses** 20

#### Purpose

This document presents comparisons of residual transmission scaling across each policy analysis approach for each country, along with the 95% credible interval associated with each. We would expect that approaches to analysis that if applying policy metrics captured some part of the variation in transmission that would otherwise need to be included through residual variation, then that analysis approach would be associated with less dramatic changes in the residual transmission scaling.

### Results by continent

#### Africa

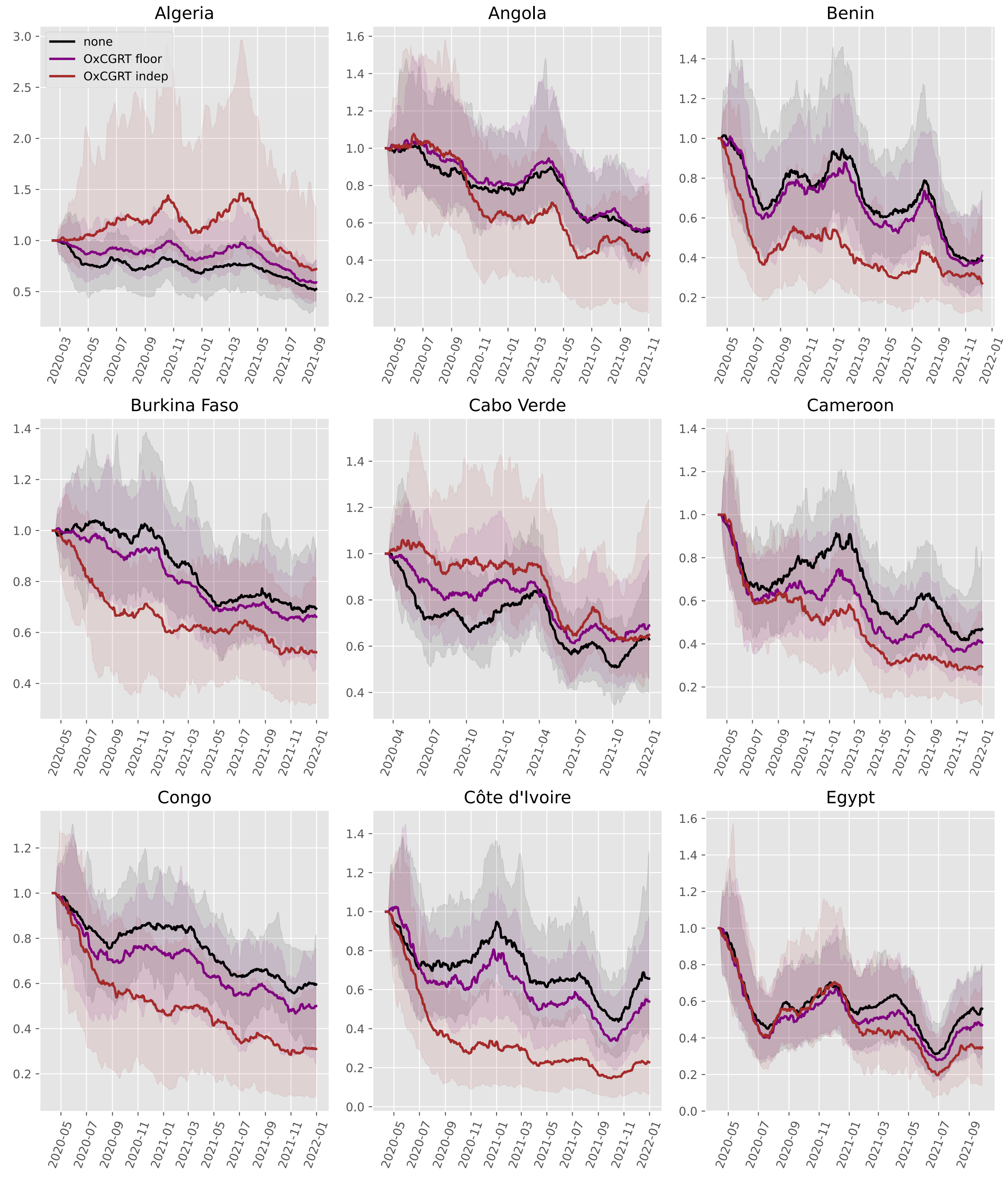

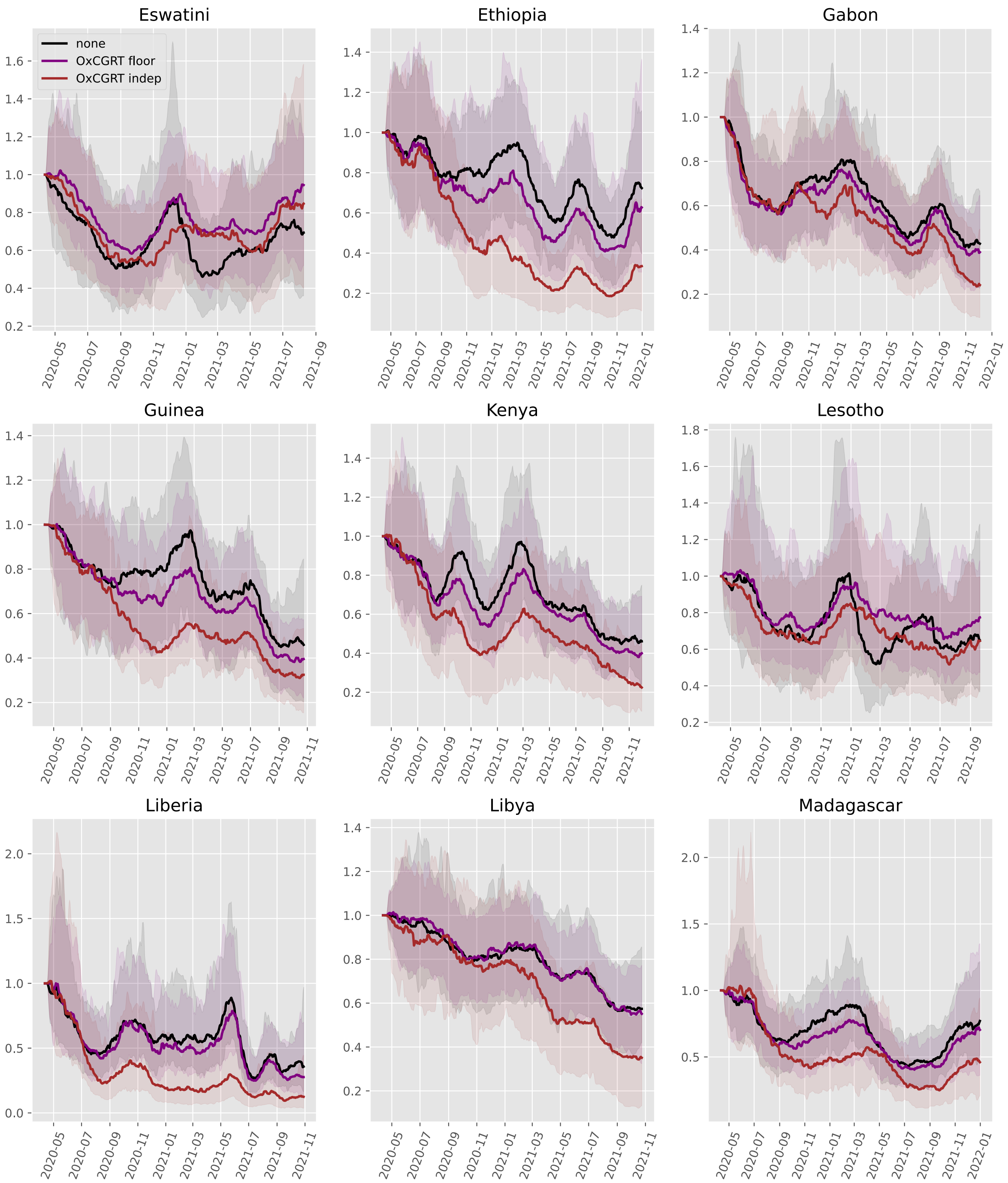

Eswatini
Ethiopia
Gabon
Guinea
Kenya
Lesotho
Liberia
Libya
Madagascar
none
OxCGRT floor
OxCGRT indep

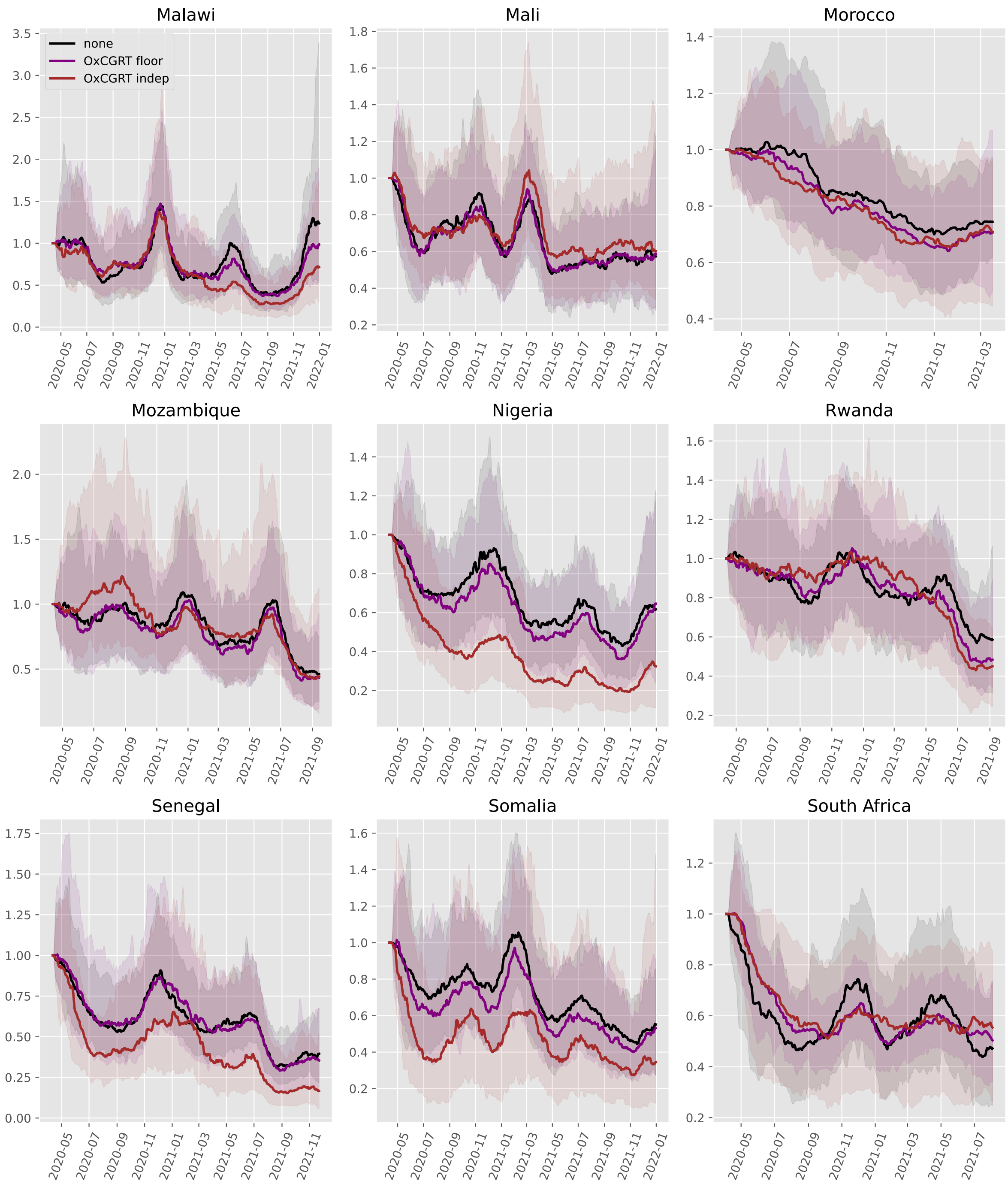

Malawi
Mali
Morocco
Mozambique
Nigeria
Rwanda
Senegal
Somalia
South Africa
none
OxCGRT floor
OxCGRT indep

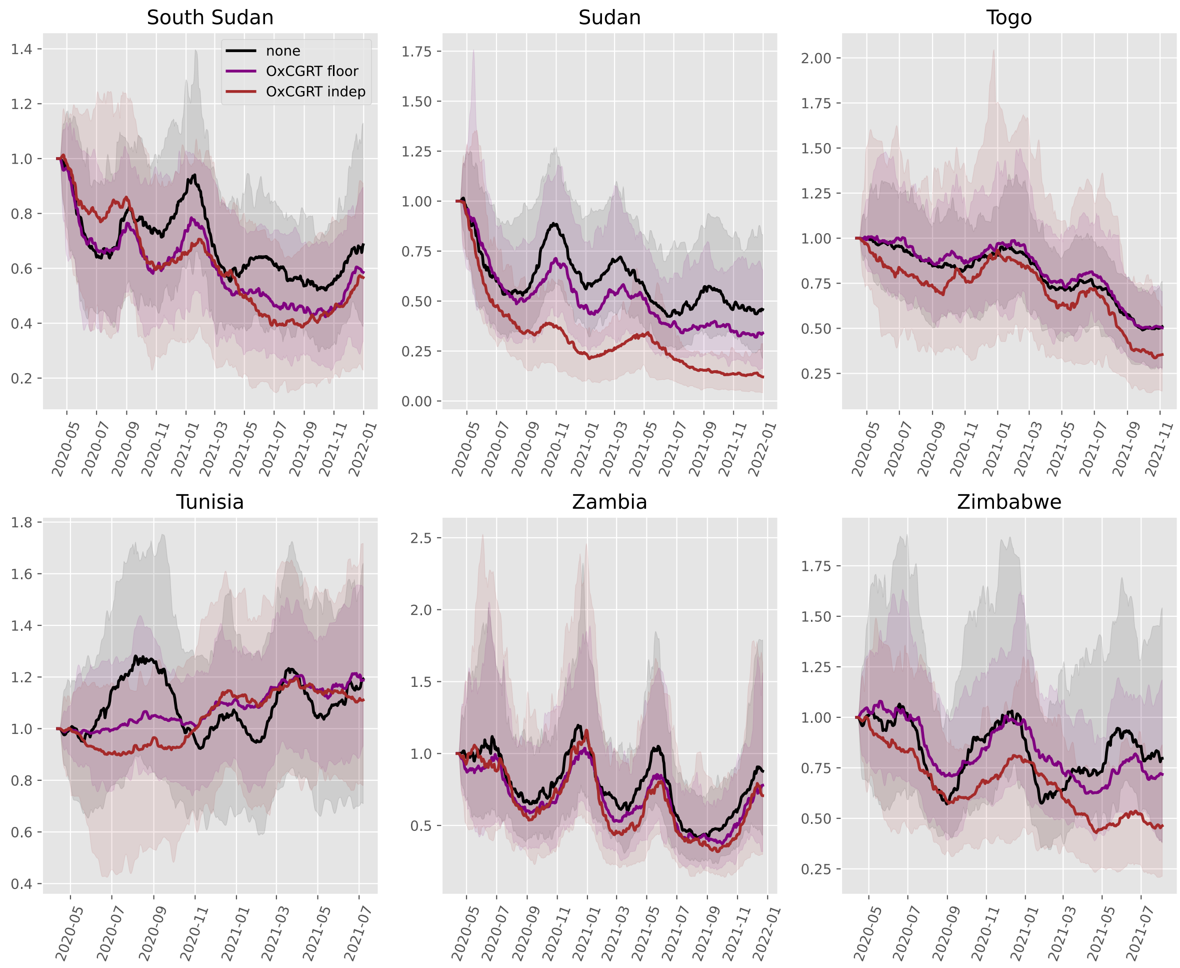
South Sudan
Sudan
Togo
Tunisia
Zambia
Zimbabwe
none
OxCGRT floor
OxCGRT indep

### Asia

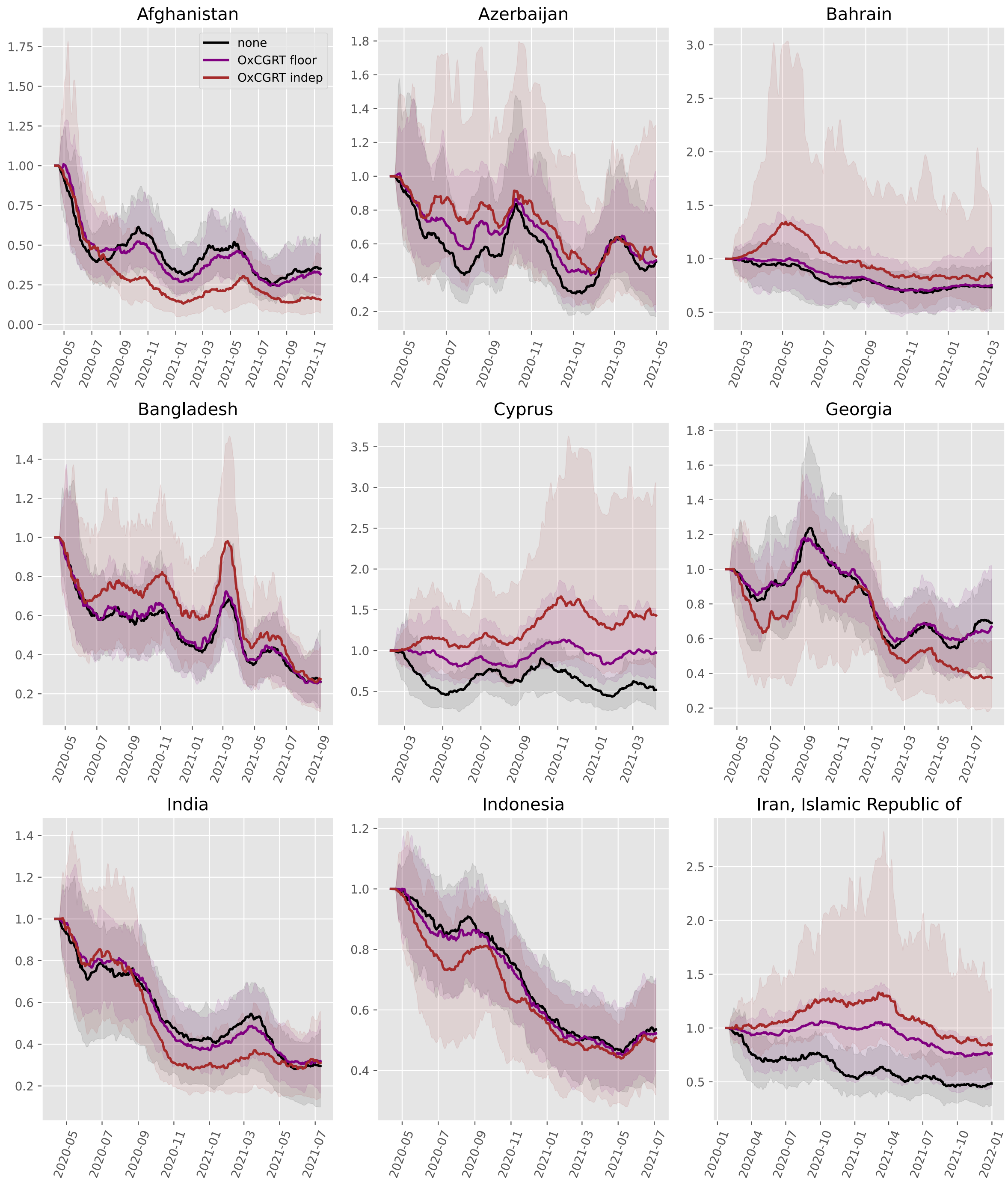

Afghanistan
none
OxCGRT floor
OxCGRT indep
Azerbaijan
Bahrain
Bangladesh
Cyprus
Georgia
India
Indonesia
Iran, Islamic Republic of

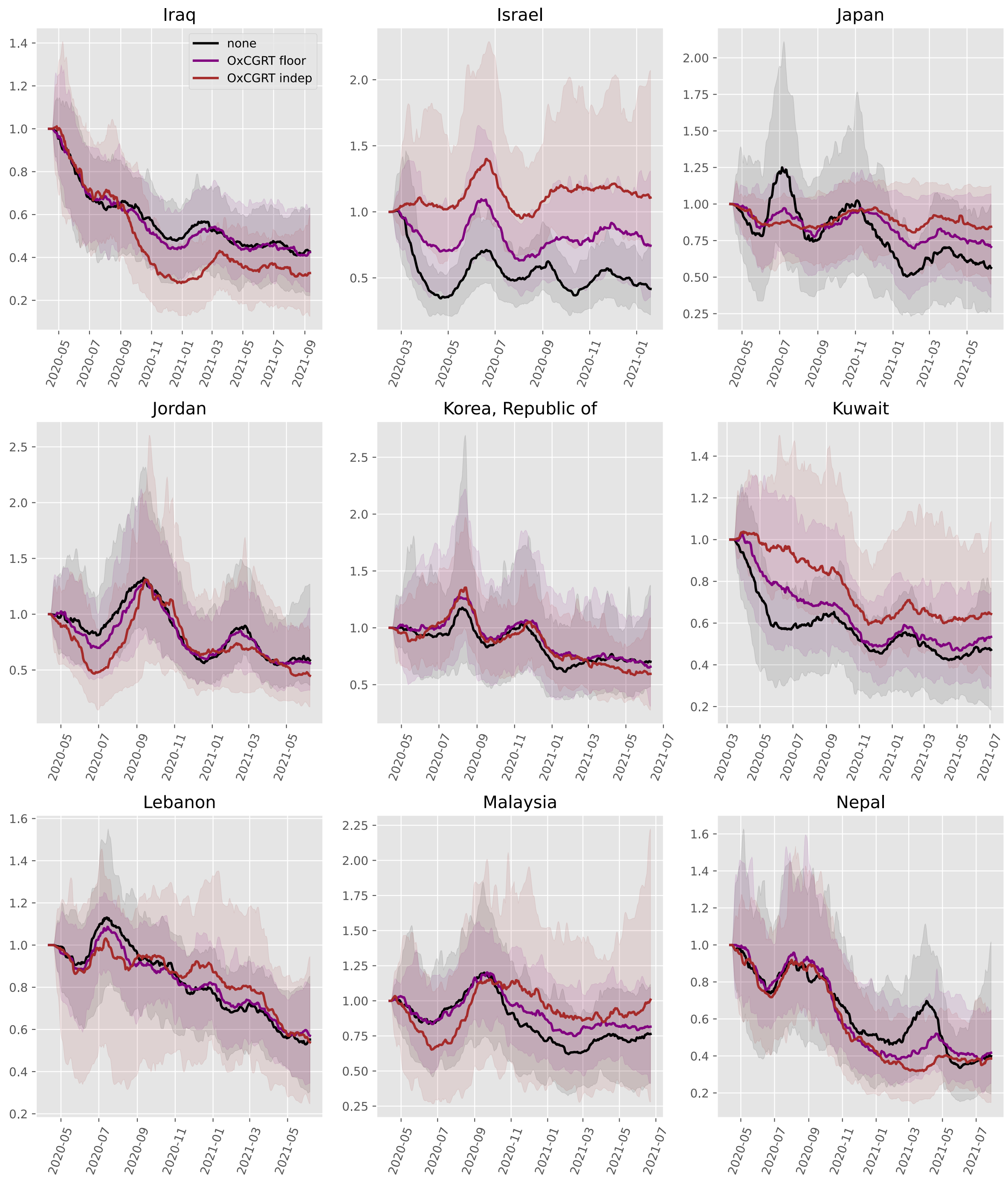
Iraq
none
OxCGRT floor
OxCGRT indep
Israel
Japan
Jordan
Korea, Republic of
Kuwait
Lebanon
Malaysia
Nepal

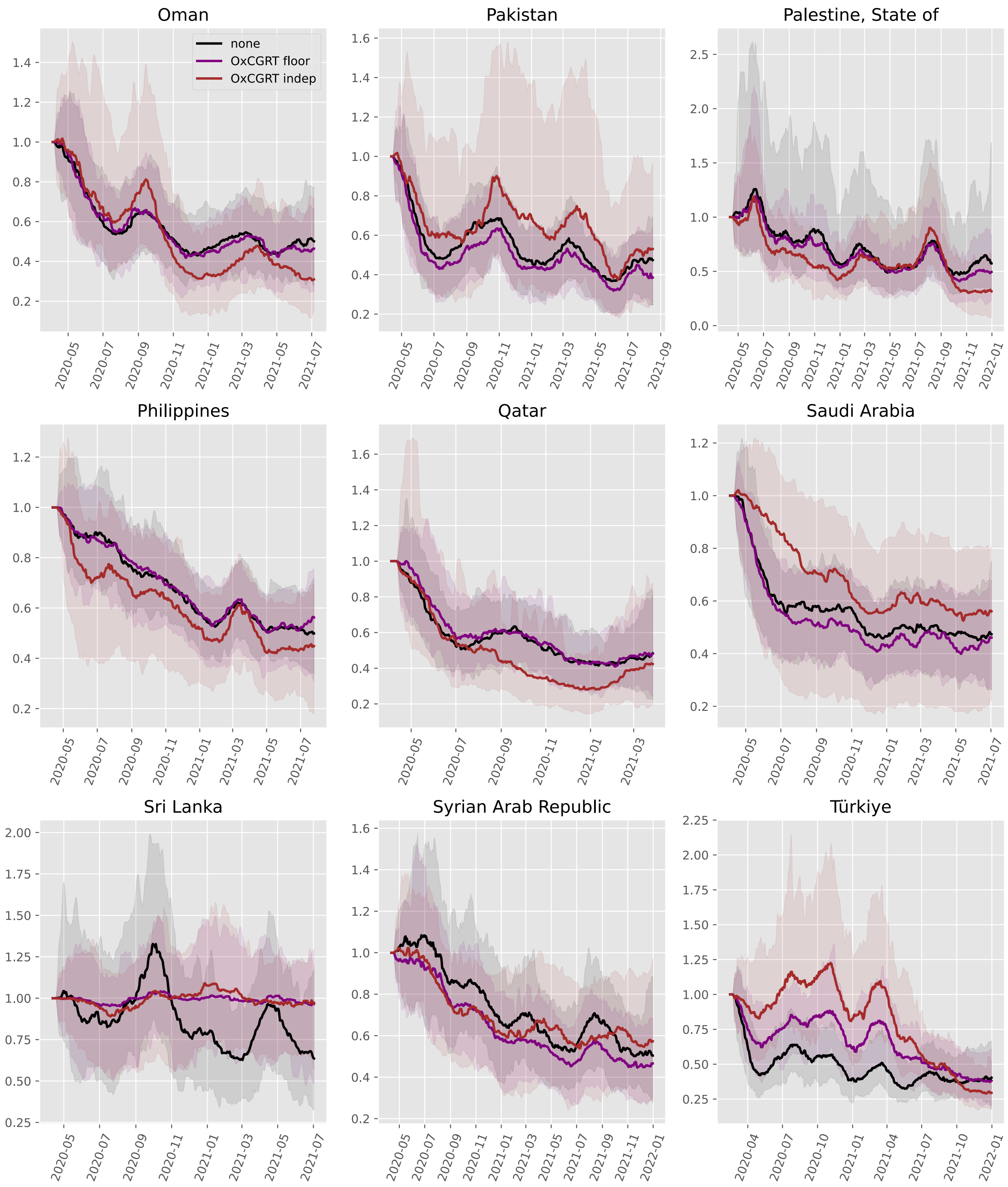

Oman
Pakistan
Palestine, State of
Philippines
Qatar
Saudi Arabia
Sri Lanka
Syrian Arab Republic
Türkiye
none
OxCGRT floor
OxCGRT indep

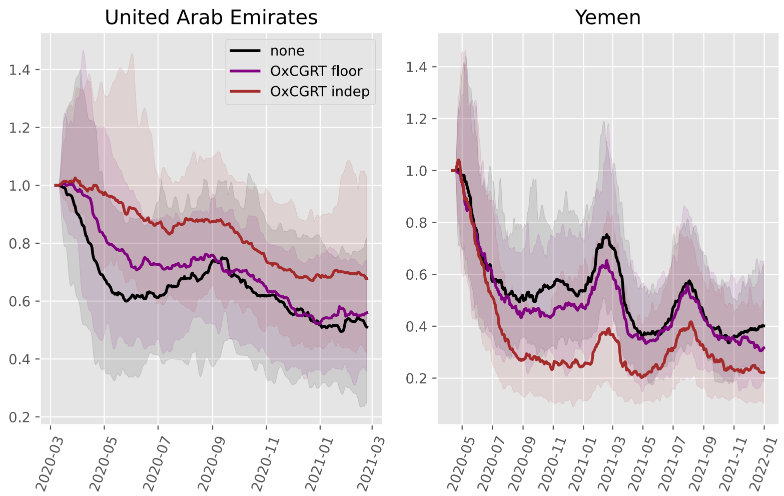
United Arab Emirates
Yemen
none
OxCGRT floor
OxCGRT indep
1.4
1.2
1.0
0.8
0.6
0.4
0.2
2020-03
2020-05
2020-07
2020-09
2020-11
2021-01
2021-03
2021-05
2021-07
2021-09
2021-11
2022-01

#### Europe

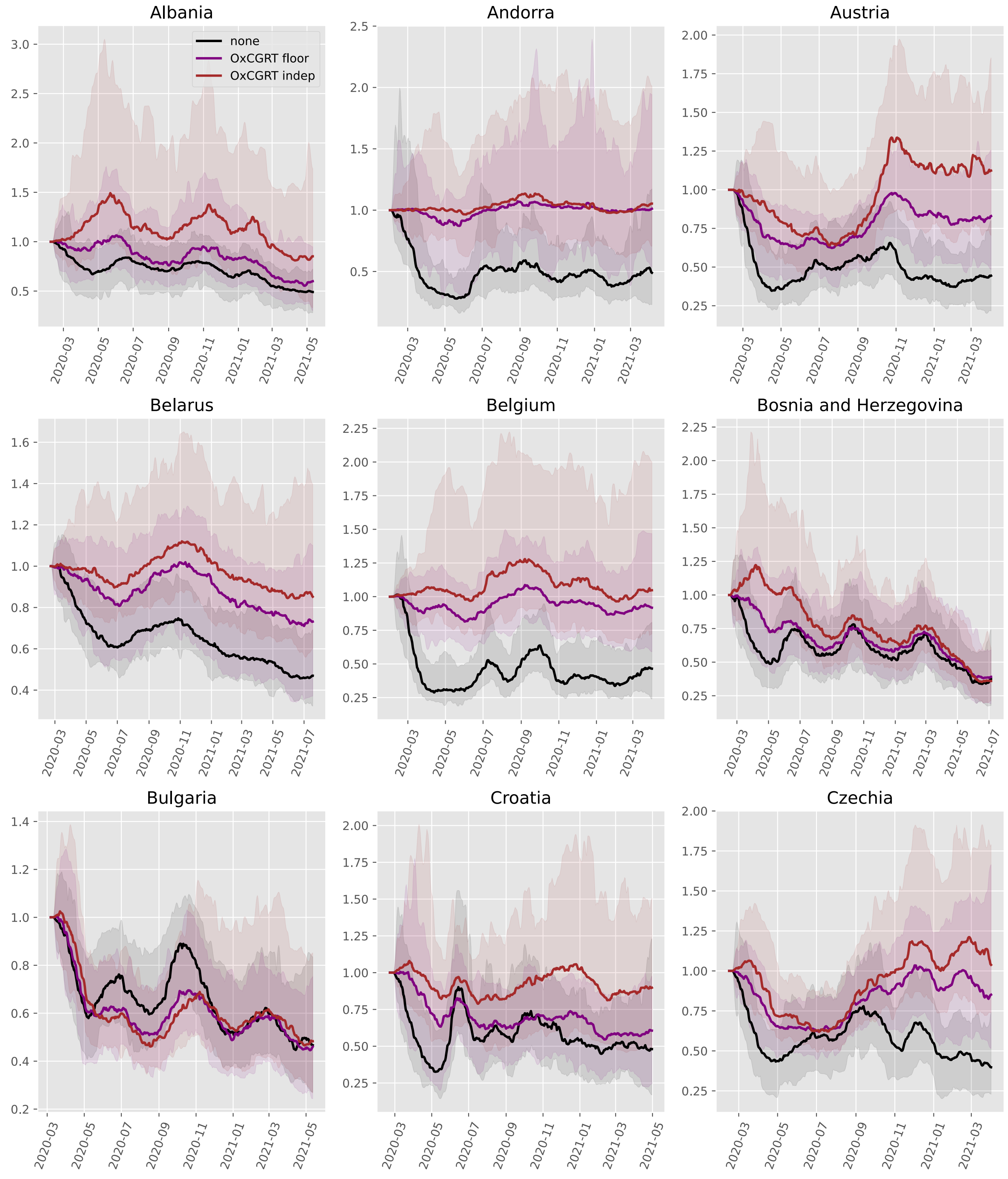
Albania
none
OxCGRT floor
OxCGRT indep
Andorra
Austria
Belarus
Belgium
Bosnia and Herzegovina
Bulgaria
Croatia
Czechia

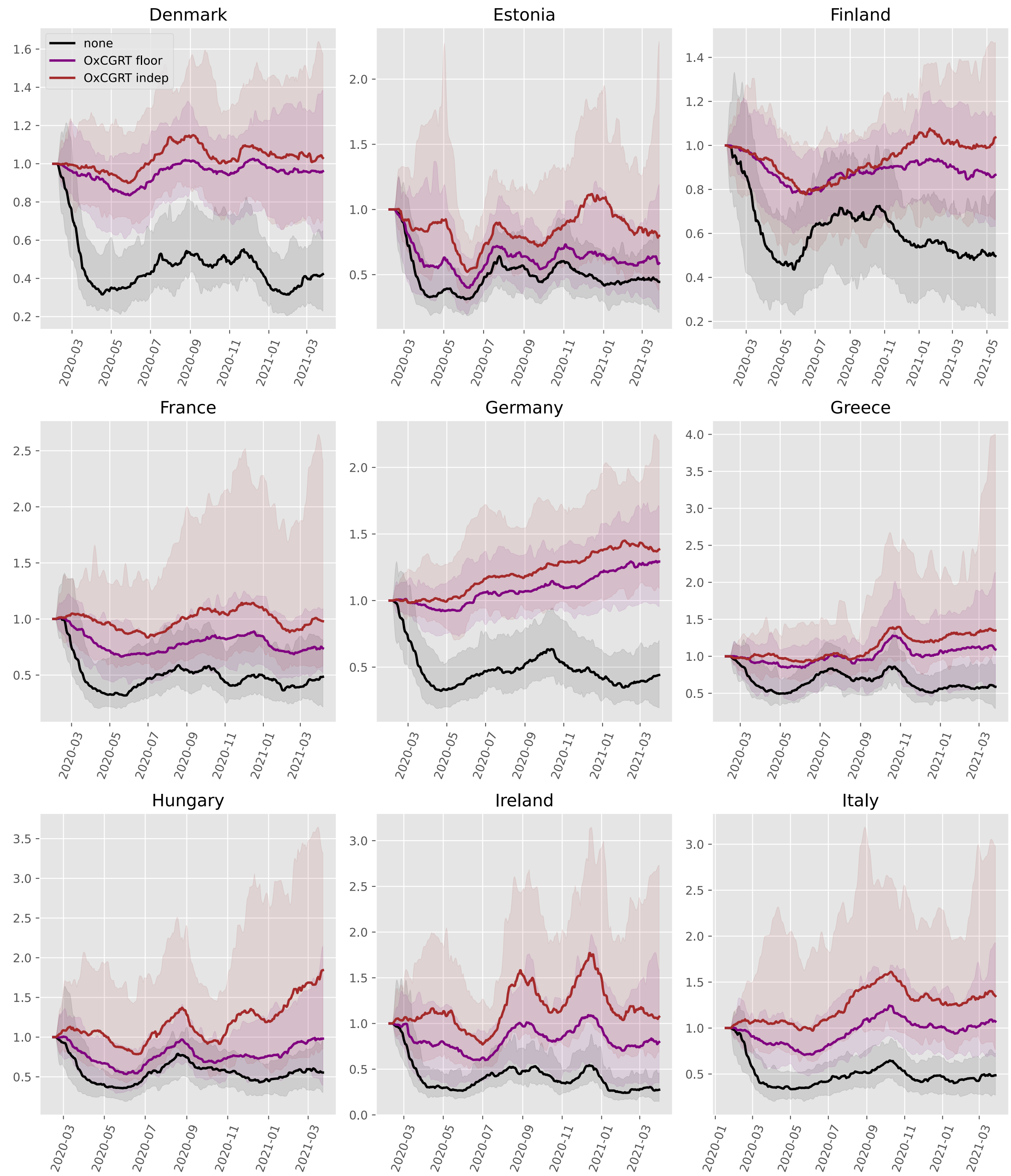
Denmark
Estonia
Finland
France
Germany
Greece
Hungary
Ireland
Italy
none
OxCGRT floor
OxCGRT indep

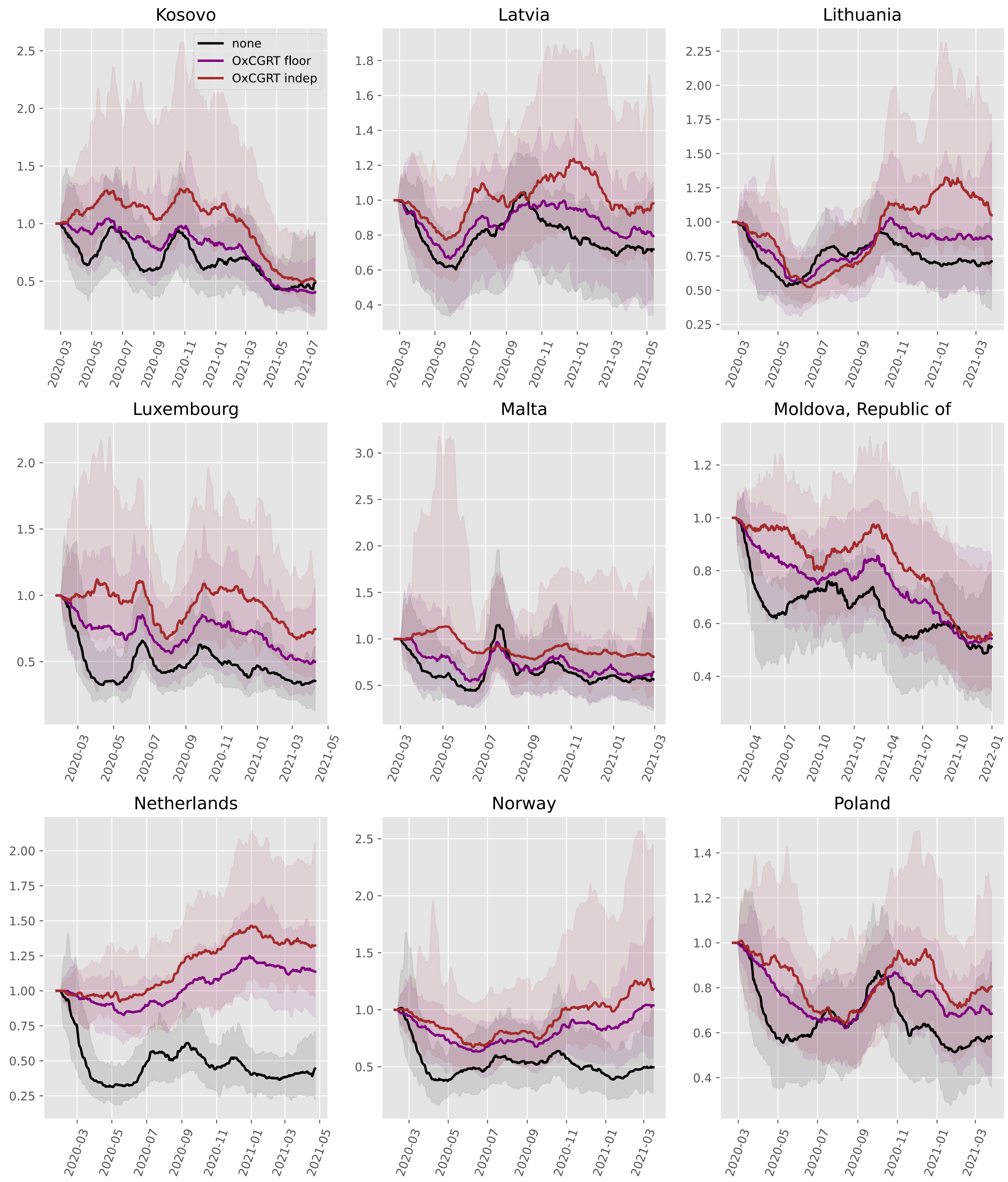
Kosovo
Latvia
Lithuania
Luxembourg
Malta
Moldova, Republic of
Netherlands
Norway
Poland
none
OxCGRT floor
OxCGRT indep

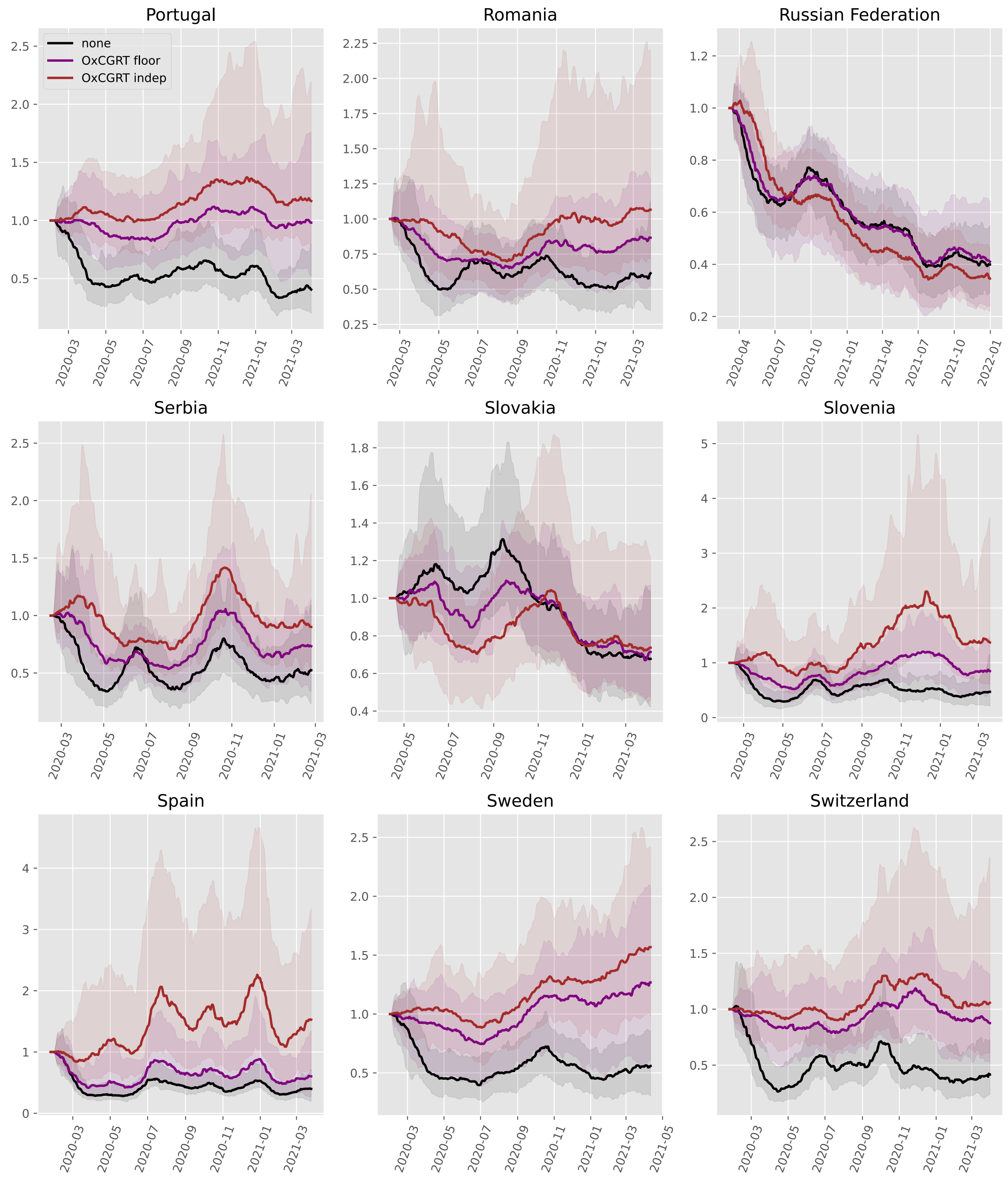
Portugal
none
OxCGRT floor
OxCGRT indep
Romania
Russian Federation
Serbia
Slovakia
Slovenia
Spain
Sweden
Switzerland

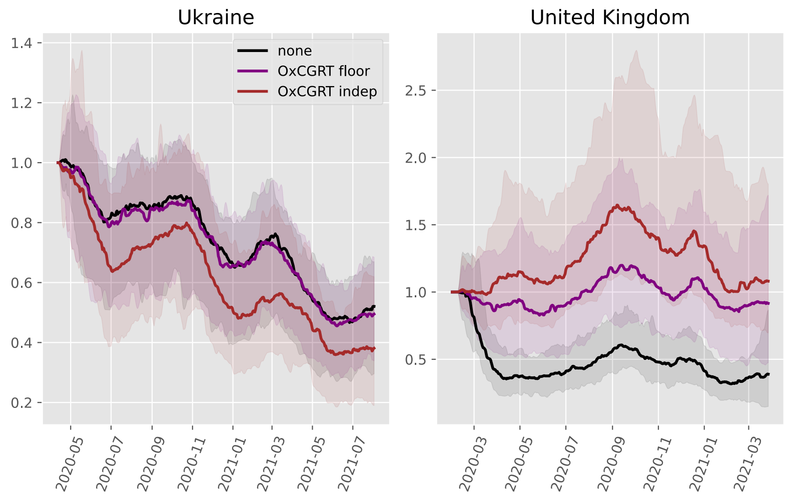
Ukraine
United Kingdom
none
OxCGRT floor
OxCGRT indep
1.4
1.2
1.0
0.8
0.6
0.4
0.2
2.5
2.0
1.5
1.0
0.5
2020-05
2020-07
2020-09
2020-11
2021-01
2021-03
2021-05
2021-07
2020-03
2020-05
2020-07
2020-09
2020-11
2021-01
2021-03

#### North America

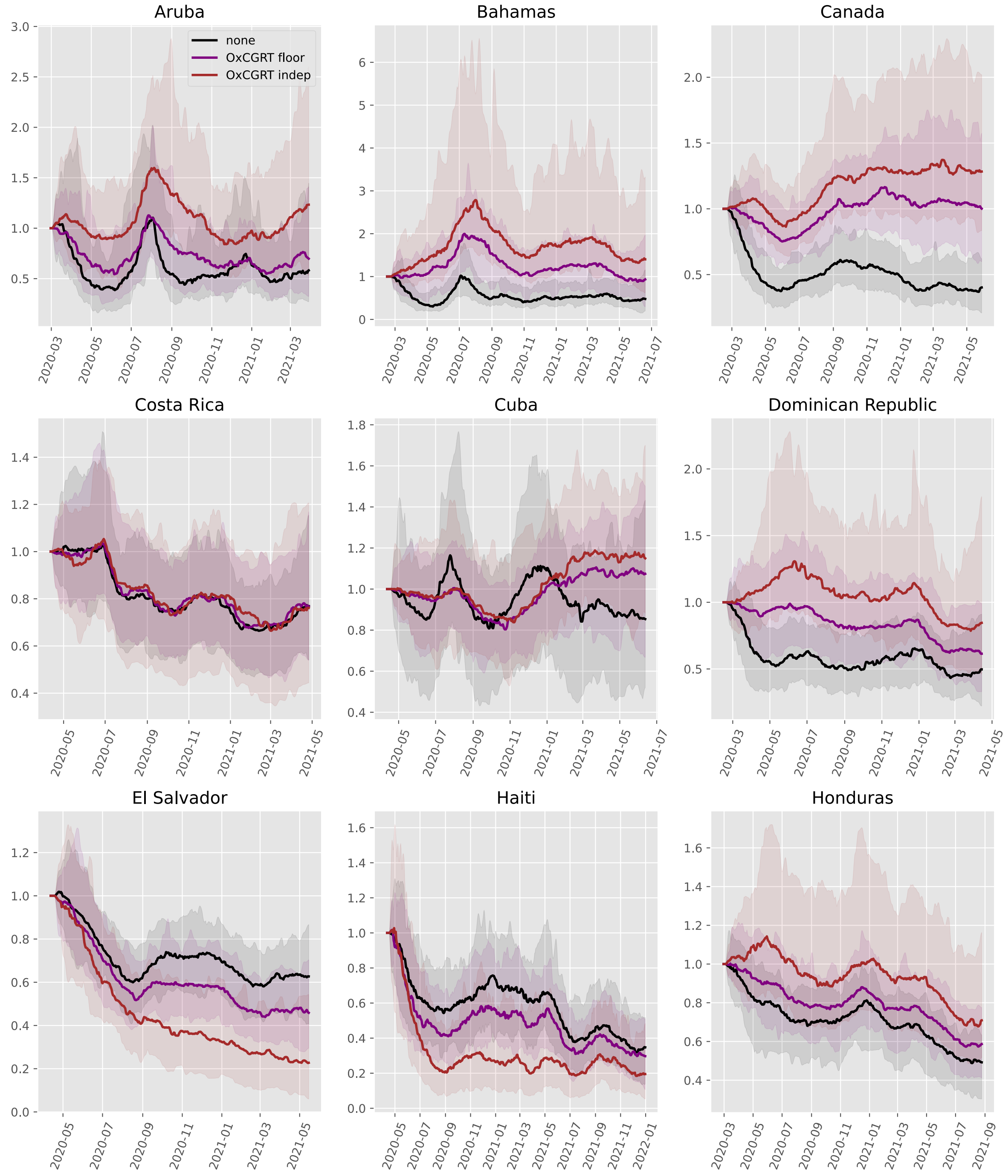

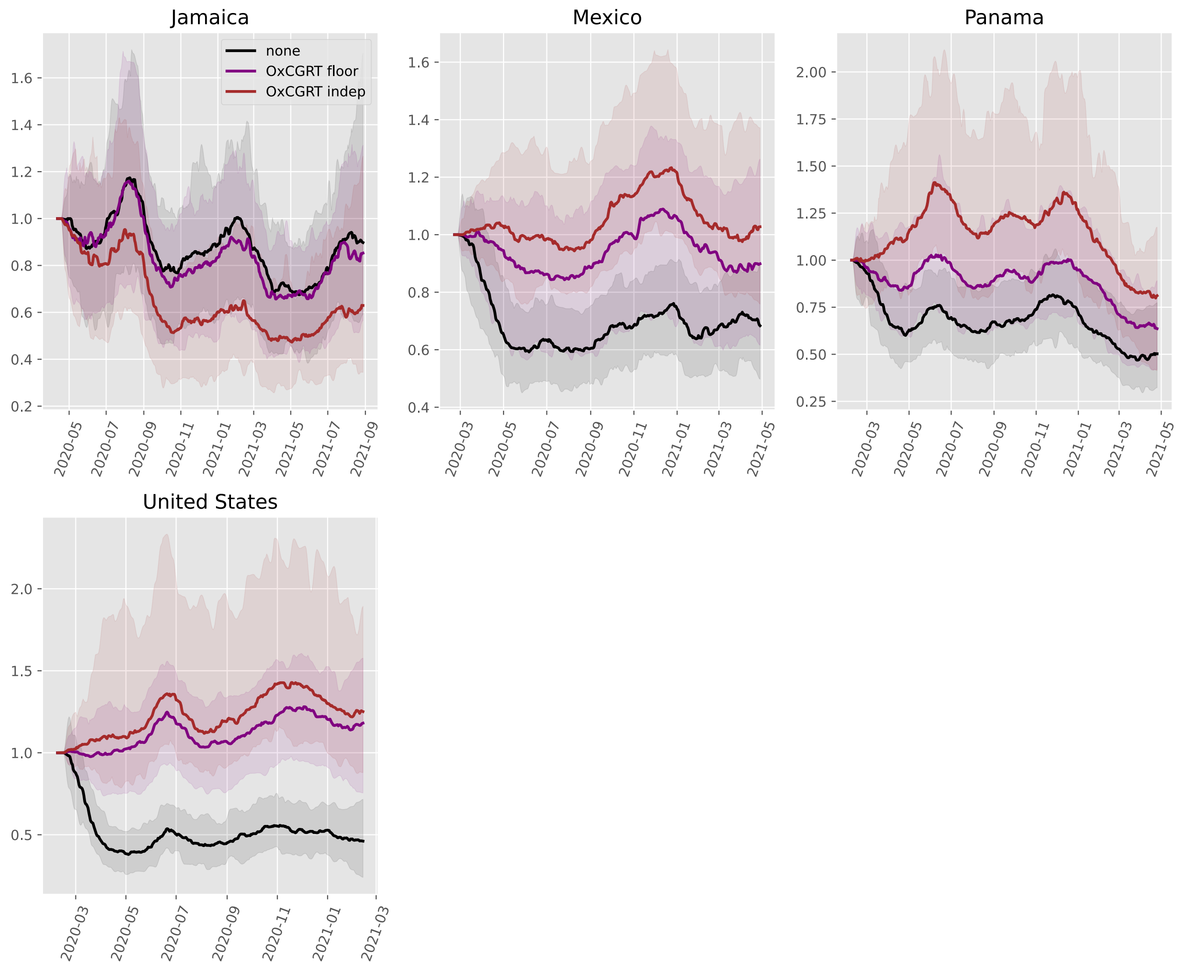

Jamaica
none
OxCGRT floor
OxCGRT indep
Mexico
Panama
United States

#### Oceania

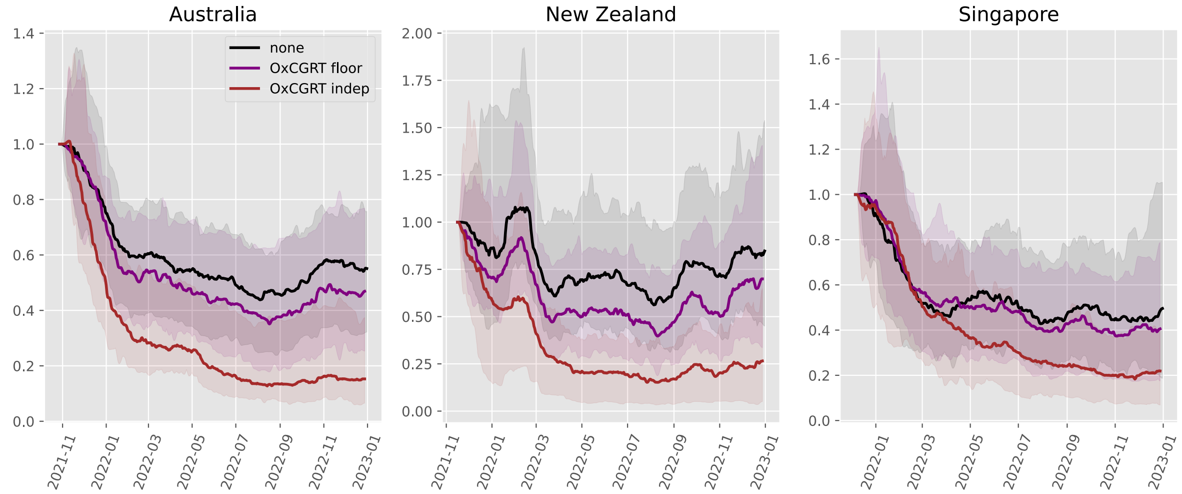

Australia
New Zealand
Singapore
none
OxCGRT floor
OxCGRT indep

#### South America

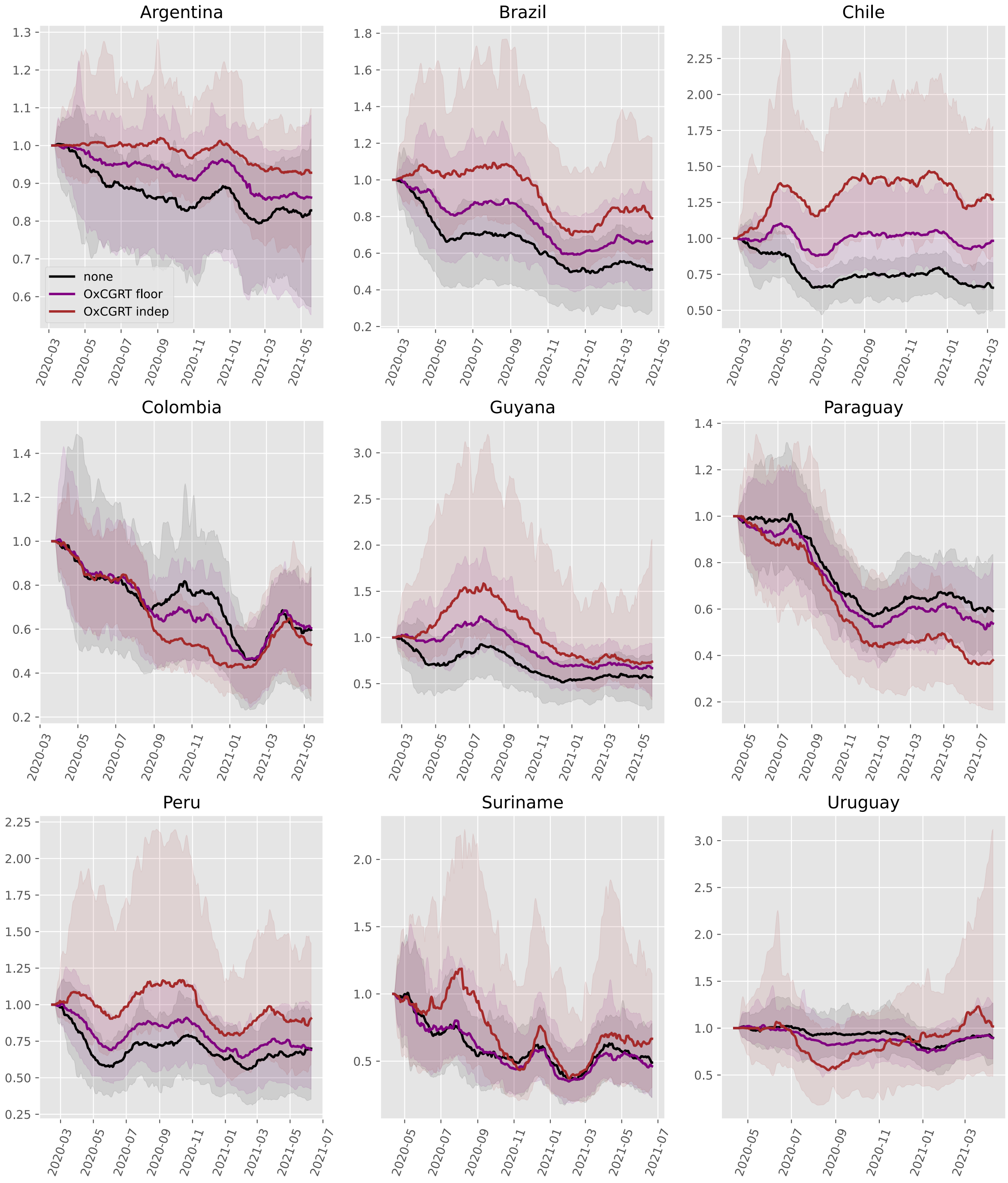

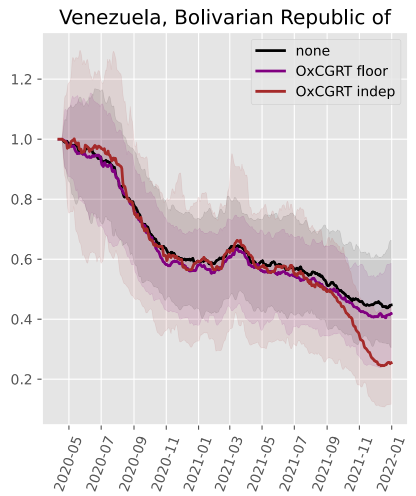

Venezuela, Bolivarian Republic of
none
OxCGRT floor
OxCGRT indep
1.2
1.0
0.8
0.6
0.4
0.2
2020-05
2020-07
2020-09
2020-11
2021-01
2021-03
2021-05
2021-07
2021-09
2021-11
2022-01
