## Supplemental File 8 for "Quantifying the contribution of policy measures to COVID-19 dynamics across 126 countries"

### Transmission scaling parameter posteriors

#### Table of contents

**Purpose** 1

**Scaling floor parameter** 3
- Africa 3
- Asia 5
- Europe 7
- North America 10
- Oceania 11
- South America 11

**Scaling exponent parameter** 12
- Africa 12
- Asia 14
- Europe 16
- North America 19
- Oceania 20
- South America 20

#### Purpose

This document presents the posterior distributions of the two parameters that define the floored scaling family of analyses used for the floored OxCGRT and mobility approaches: the scaling floor $f$ and the scaling exponent $m$. In these analysis approaches:

$$M_t = \left[f + (1 - f)\, S_t\right]^m,$$

where $S_t$ is the weighted policy or complement-of-restriction signal (one when unrestricted or at its reference level, zero at maximum restriction or zero mobility). The floor parameter $f$ is the residual level of the population interaction proxy under the strongest possible intervention measures: even with policies at their most restrictive (or mobility at zero), the signal cannot be reduced below $f$, such a fraction $f$ of unrestricted contact remains before the effect of the exponent parameter is applied. The exponent parameter $m$ then governs how strongly reductions in the signal translate into reductions in transmission ($m = 1$ leaves the floored signal unchanged; $m > 1$ deepens reductions as the signal drops further below one).

Priors on these parameters were both set to be uniform over plausible bounds ($f \sim \text{Beta}(1, 1)$ and $m \sim \text{Uniform}(0, 2)$). Because $f$ and $m$ trade off, their marginal posteriors are not fully separable: similar realised $M_t$ could arise from a higher floor with a larger exponent or a lower floor with a smaller exponent. We therefore take our primary overall metric to be $(1 - f)^m$, results from which are presented in the following document. Note that the independent OxCGRT analysis is omitted, because it does not use $f$ or $m$.

Applying a floor was necessary because the OxCGRT metrics could reach full restrictions across all domains, implying $S_t = 0$, which did not support epidemiological fitting to data. To match this approach, our analyses considering the effects of mobility were adapted from the approach in our previous paper to align structurally with this policy-focused approach.

In countries for which effects of policies and mobility are clear, mobility floors often sit lower than policy floors. This matches our intuition that zero mobility could conceptually come close to halting transmission through reduced community interactions, whereas maximum recorded policy intensity reaches a ceiling effect prior to this point.

### Scaling floor parameter

#### Africa

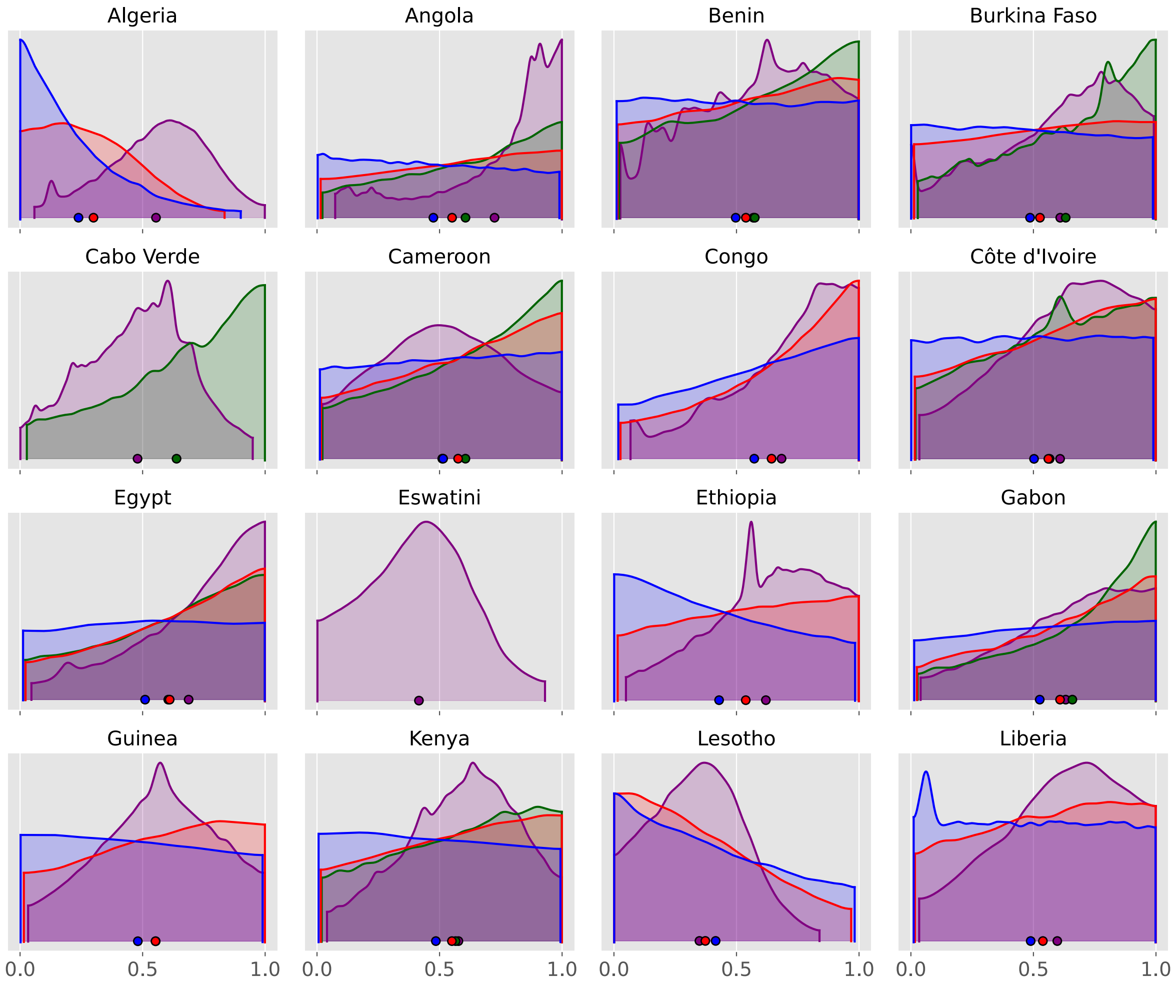

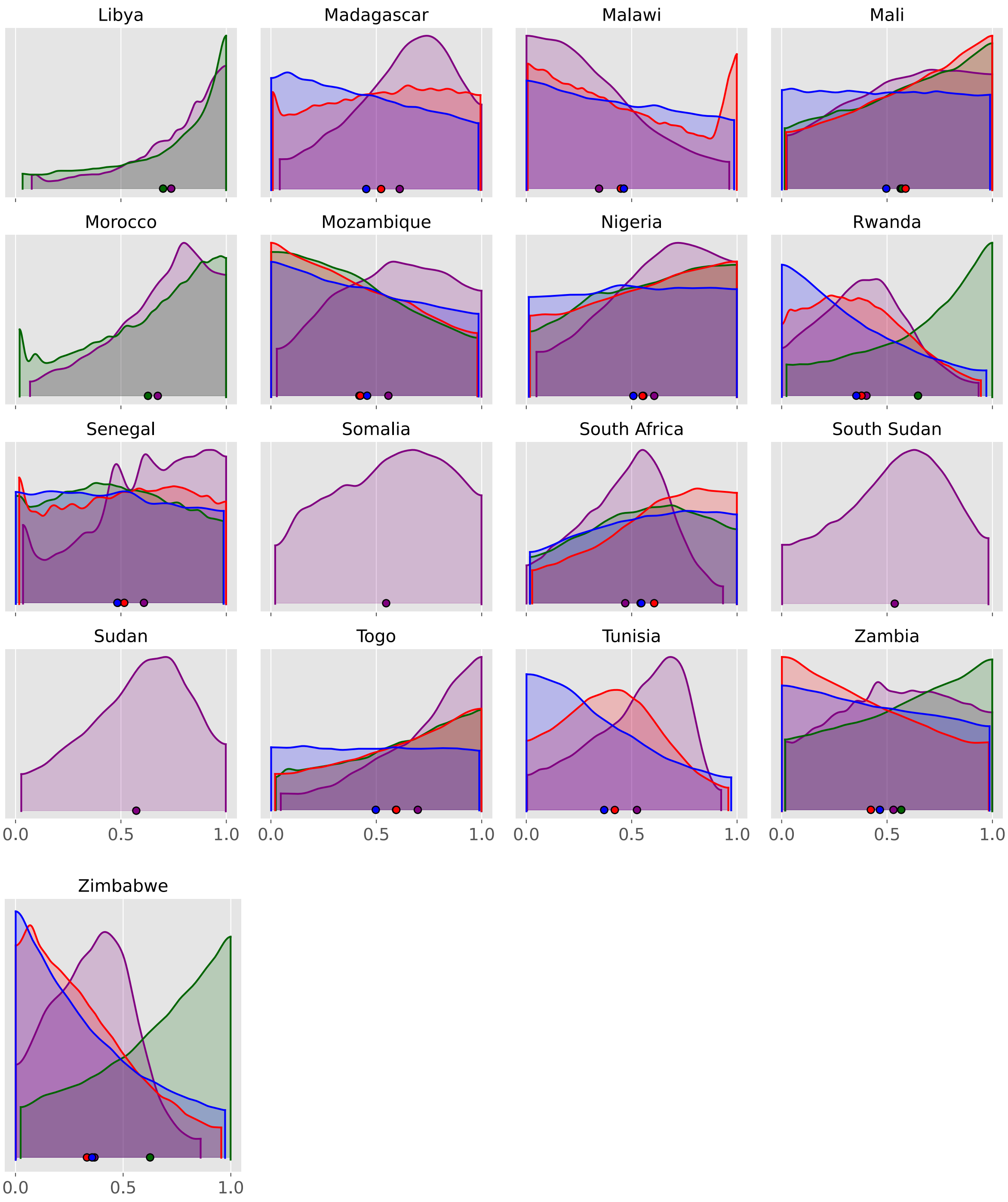
Libya
Madagascar
Malawi
Mali
Morocco
Mozambique
Nigeria
Rwanda
Senegal
Somalia
South Africa
South Sudan
Sudan
Togo
Tunisia
Zambia
Zimbabwe
0.0
0.5
1.0

#### Asia

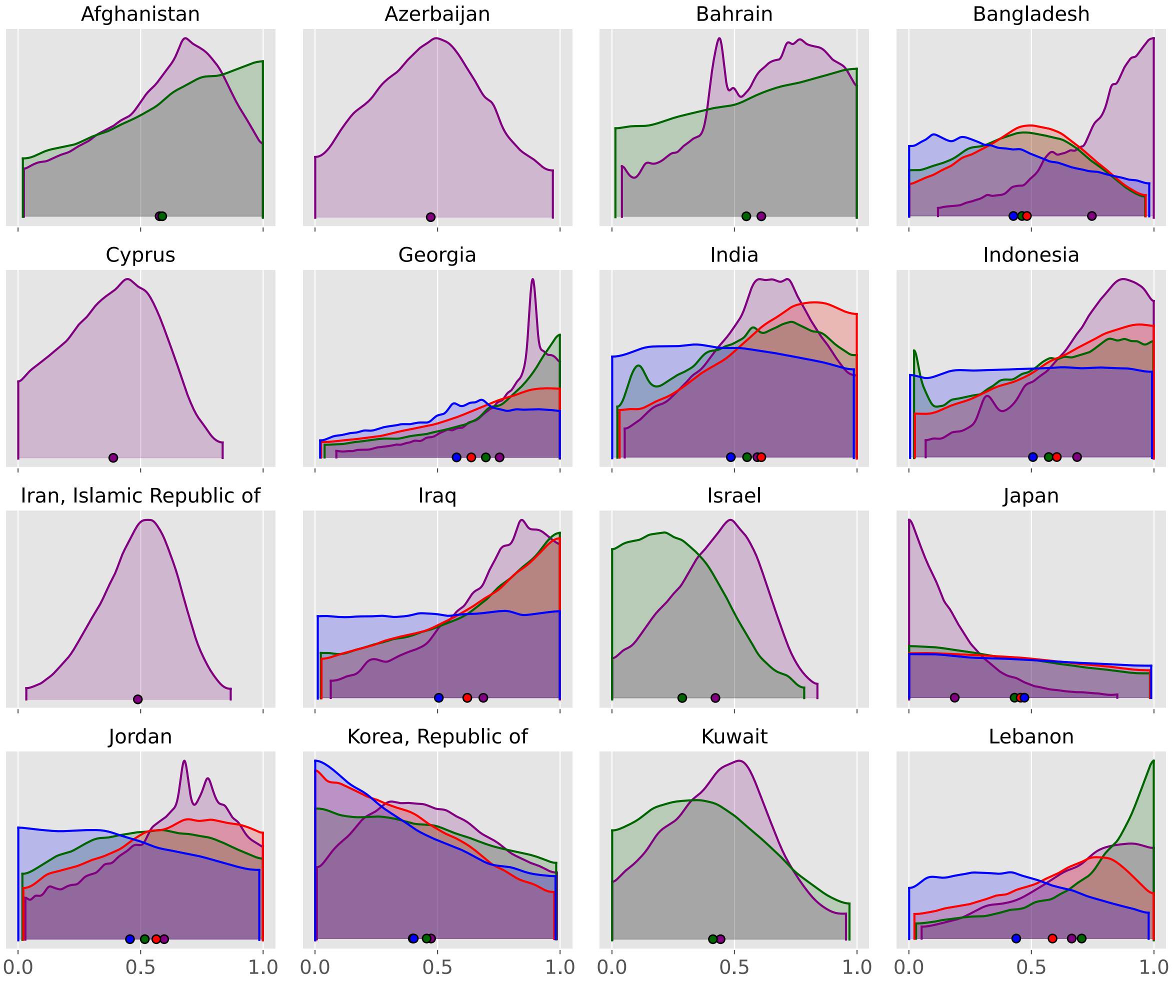

Afghanistan
Azerbaijan
Bahrain
Bangladesh
Cyprus
Georgia
India
Indonesia
Iran, Islamic Republic of
Iraq
Israel
Japan
Jordan
Korea, Republic of
Kuwait
Lebanon
0.0
0.5
1.0

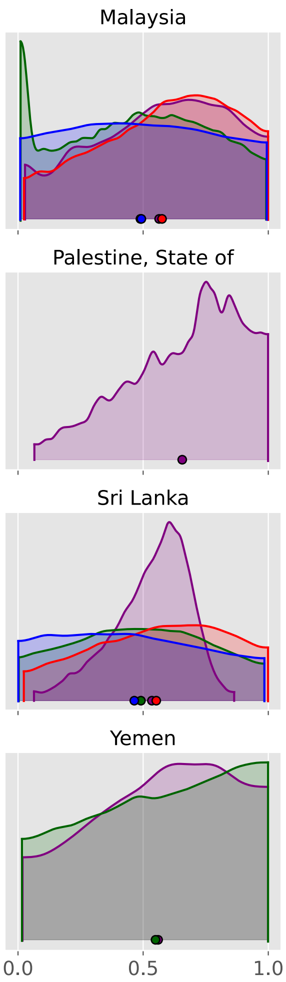
Malaysia
Nepal
Oman
Pakistan
Palestine, State of
Philippines
Qatar
Saudi Arabia
Sri Lanka
Syrian Arab Republic
Türkiye
United Arab Emirates
Yemen
0.0
0.5
1.0

#### Europe

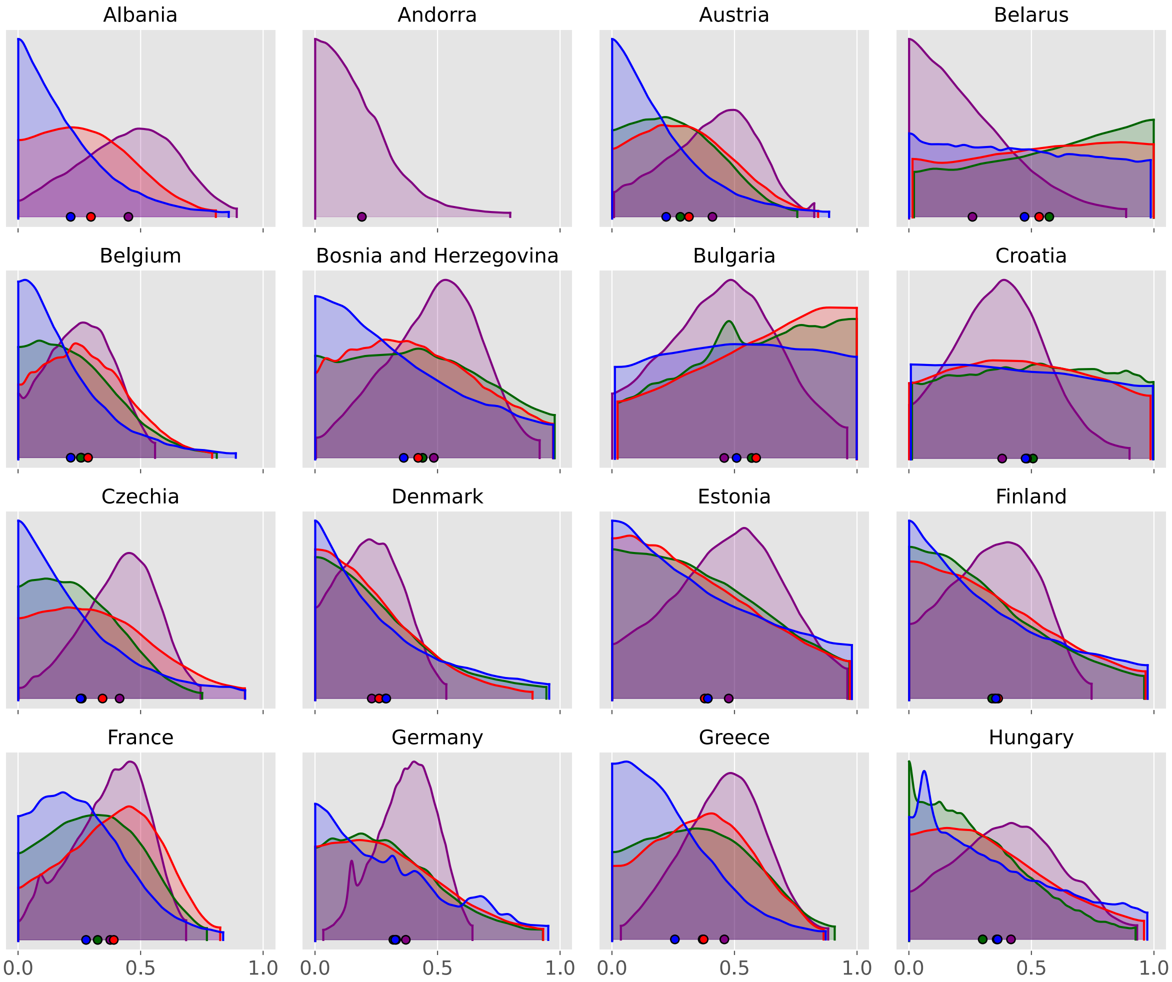

Albania
Andorra
Austria
Belarus
Belgium
Bosnia and Herzegovina
Bulgaria
Croatia
Czechia
Denmark
Estonia
Finland
France
Germany
Greece
Hungary
0.0
0.5
1.0

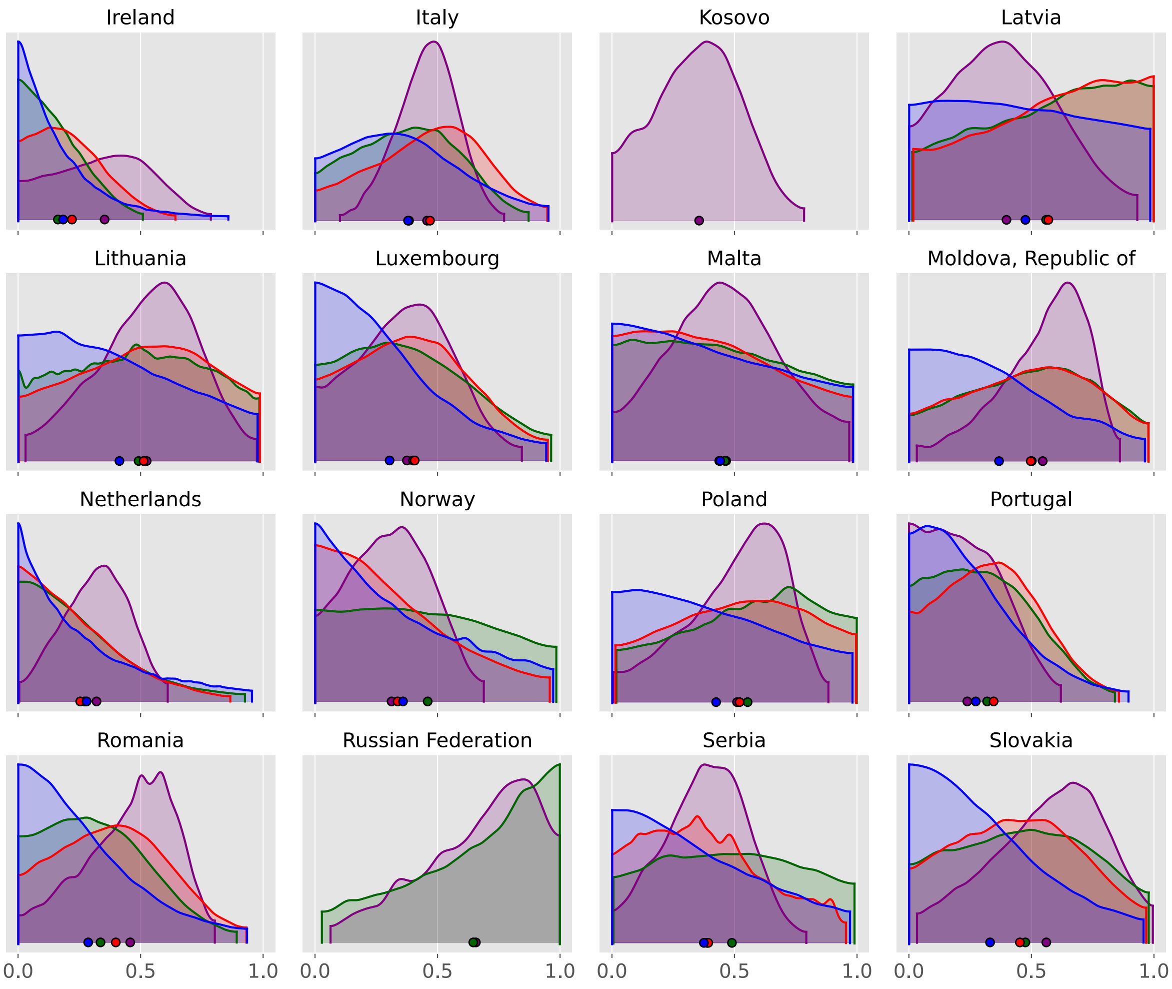
Ireland
Italy
Kosovo
Latvia
Lithuania
Luxembourg
Malta
Moldova, Republic of
Netherlands
Norway
Poland
Portugal
Romania
Russian Federation
Serbia
Slovakia
0.0
0.5
1.0

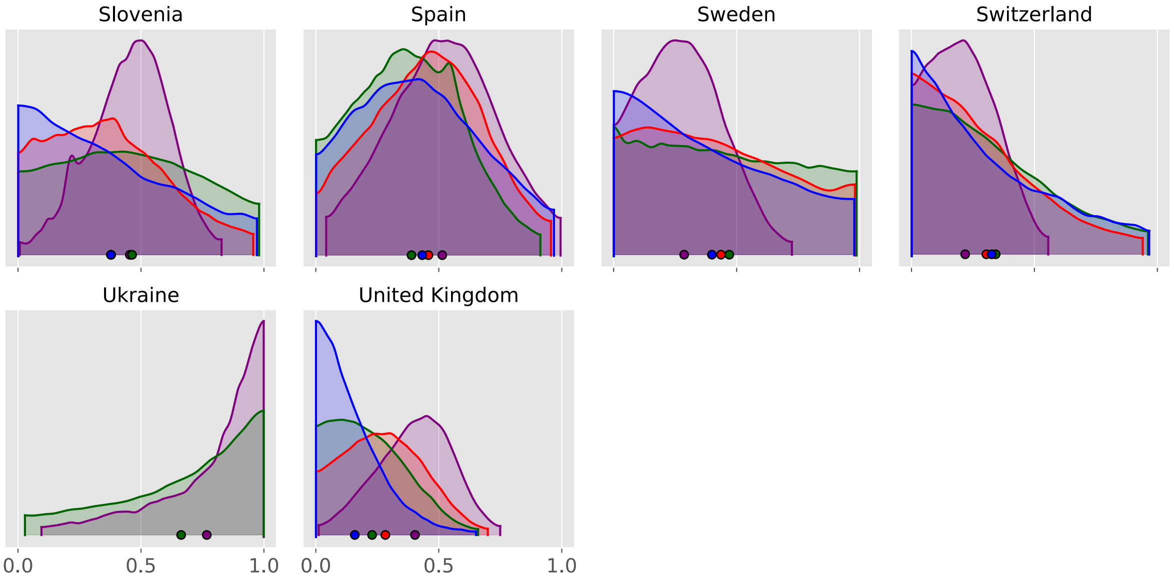
Slovenia
Spain
Sweden
Switzerland
Ukraine
United Kingdom
0.0
0.5
1.0
0.0
0.5
1.0

#### North America

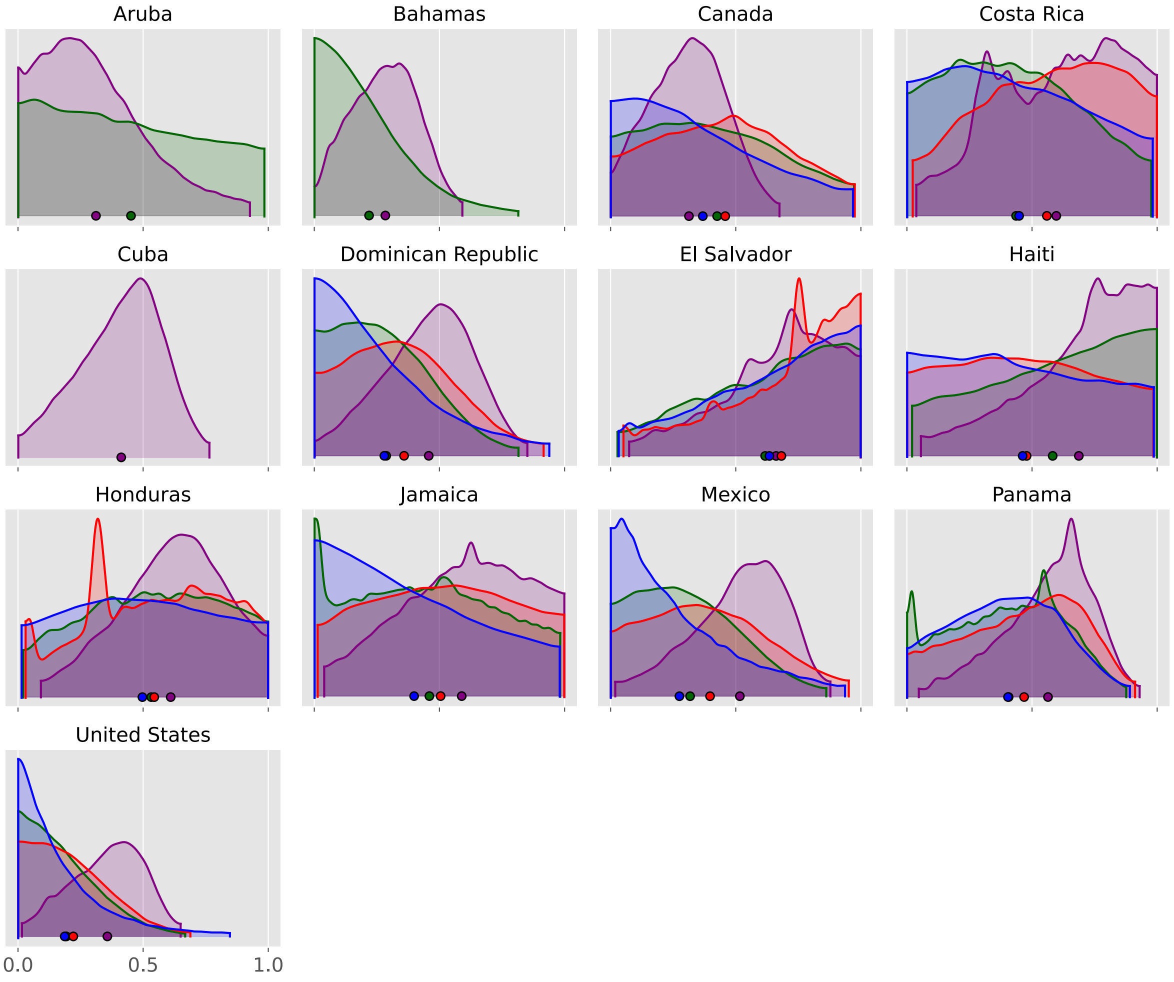

#### Oceania

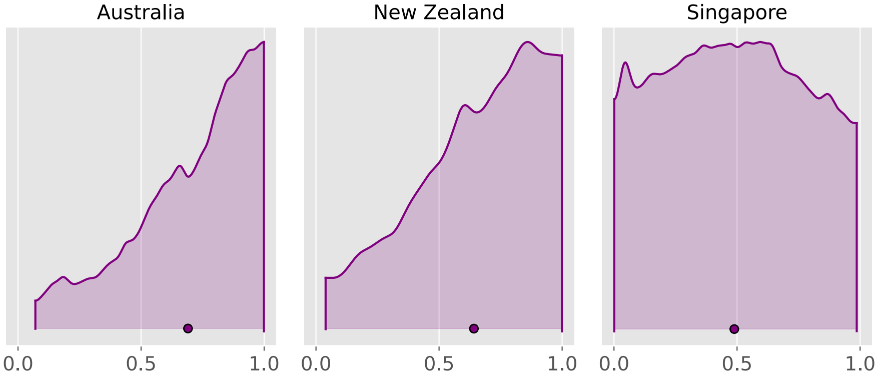


#### South America

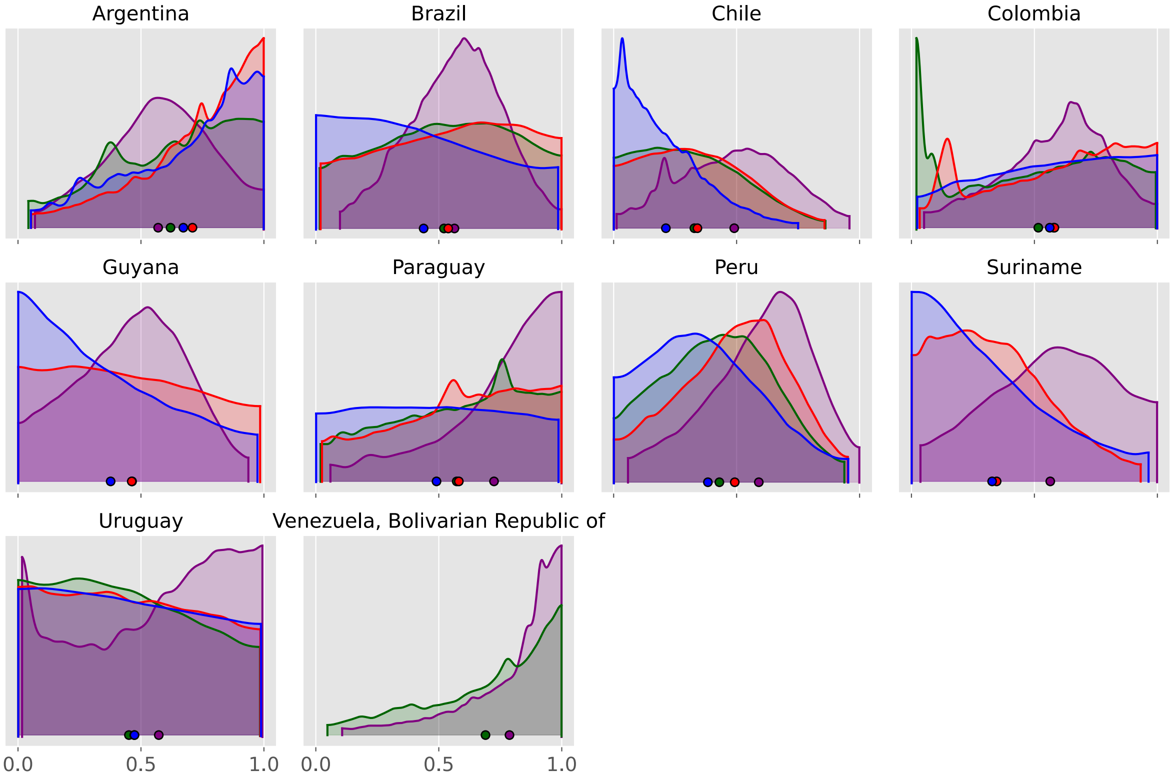

### Scaling exponent parameter

#### Africa

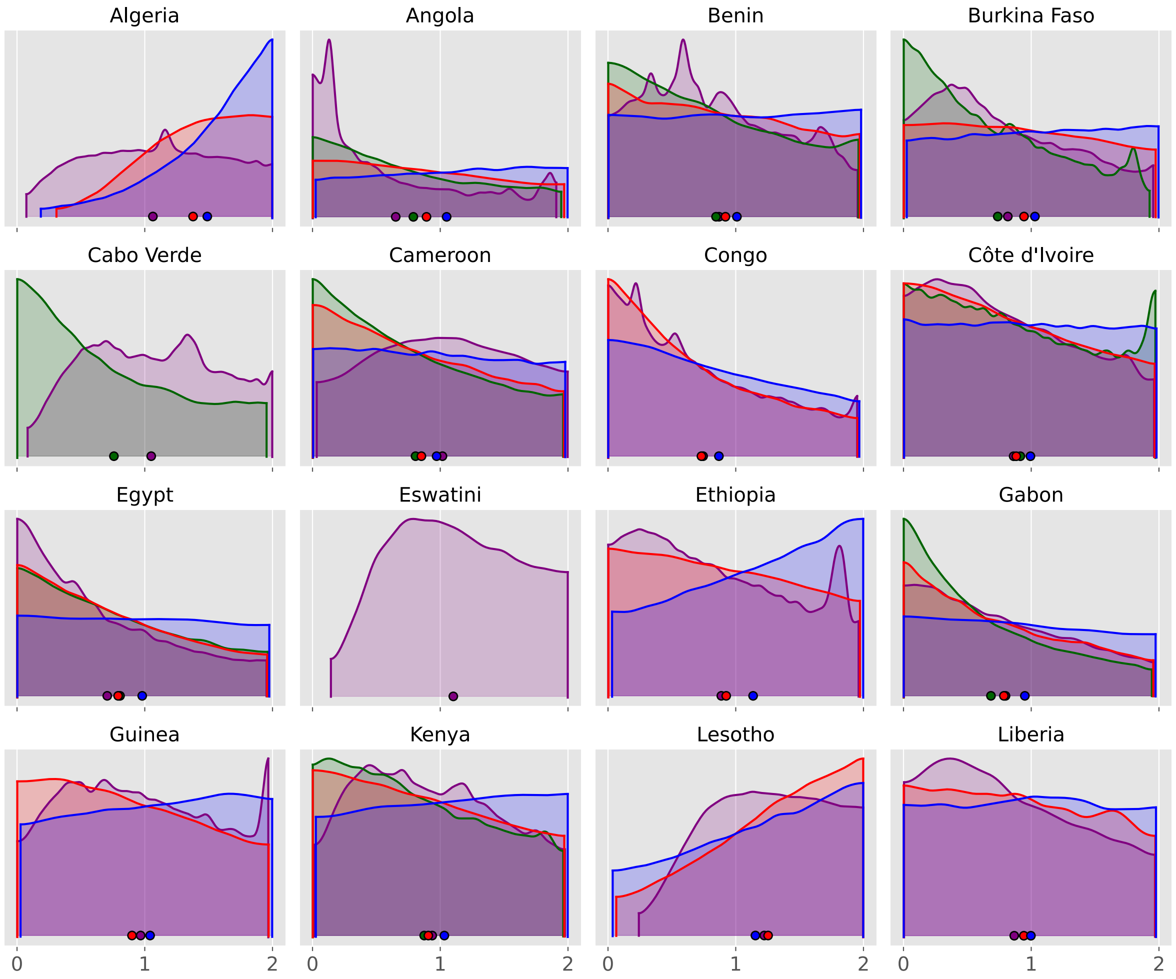

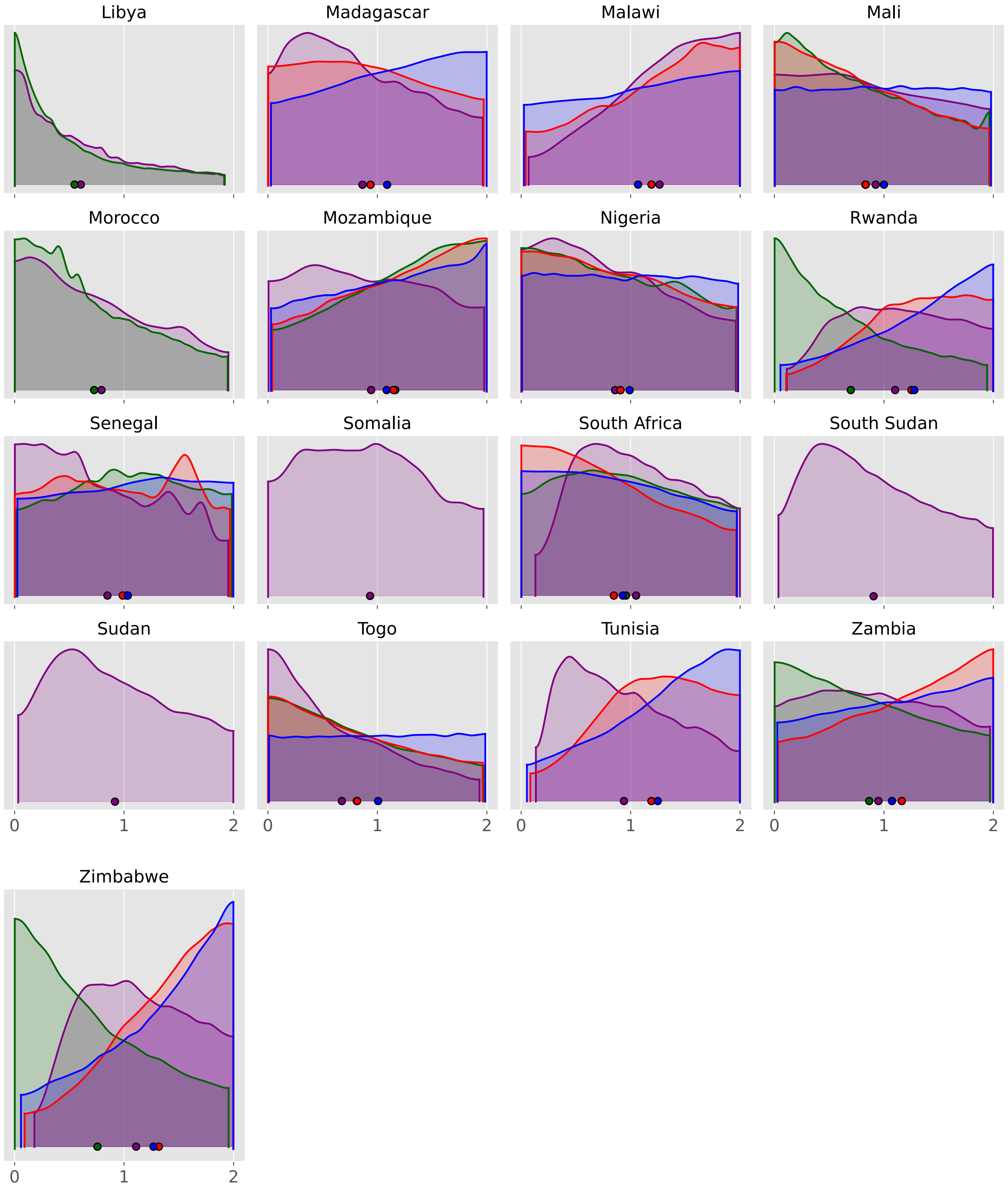
Libya
Madagascar
Malawi
Mali
Morocco
Mozambique
Nigeria
Rwanda
Senegal
Somalia
South Africa
South Sudan
Sudan
Togo
Tunisia
Zambia
Zimbabwe
0
1
2

#### Asia

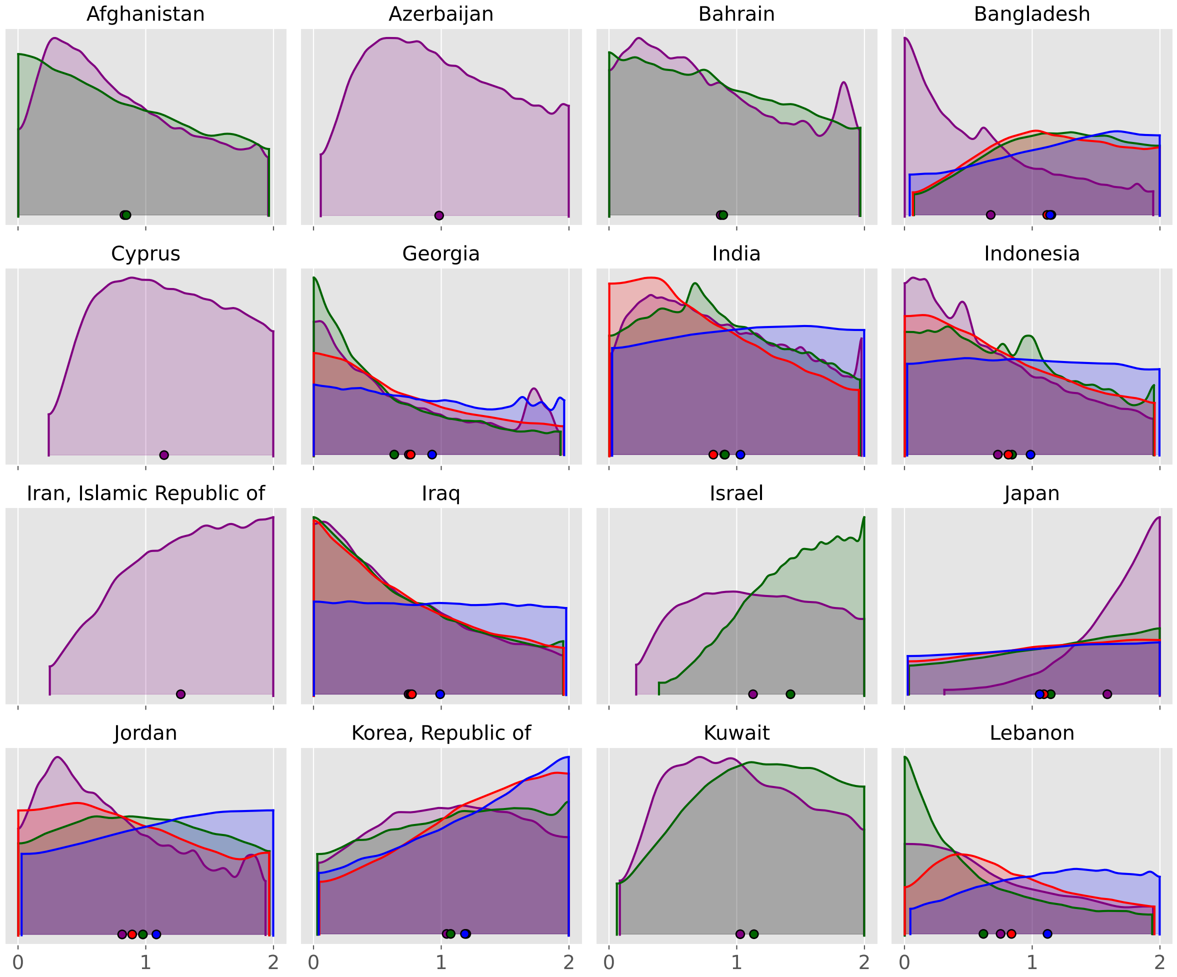
Afghanistan
Azerbaijan
Bahrain
Bangladesh
Cyprus
Georgia
India
Indonesia
Iran, Islamic Republic of
Iraq
Israel
Japan
Jordan
Korea, Republic of
Kuwait
Lebanon
0
1
2

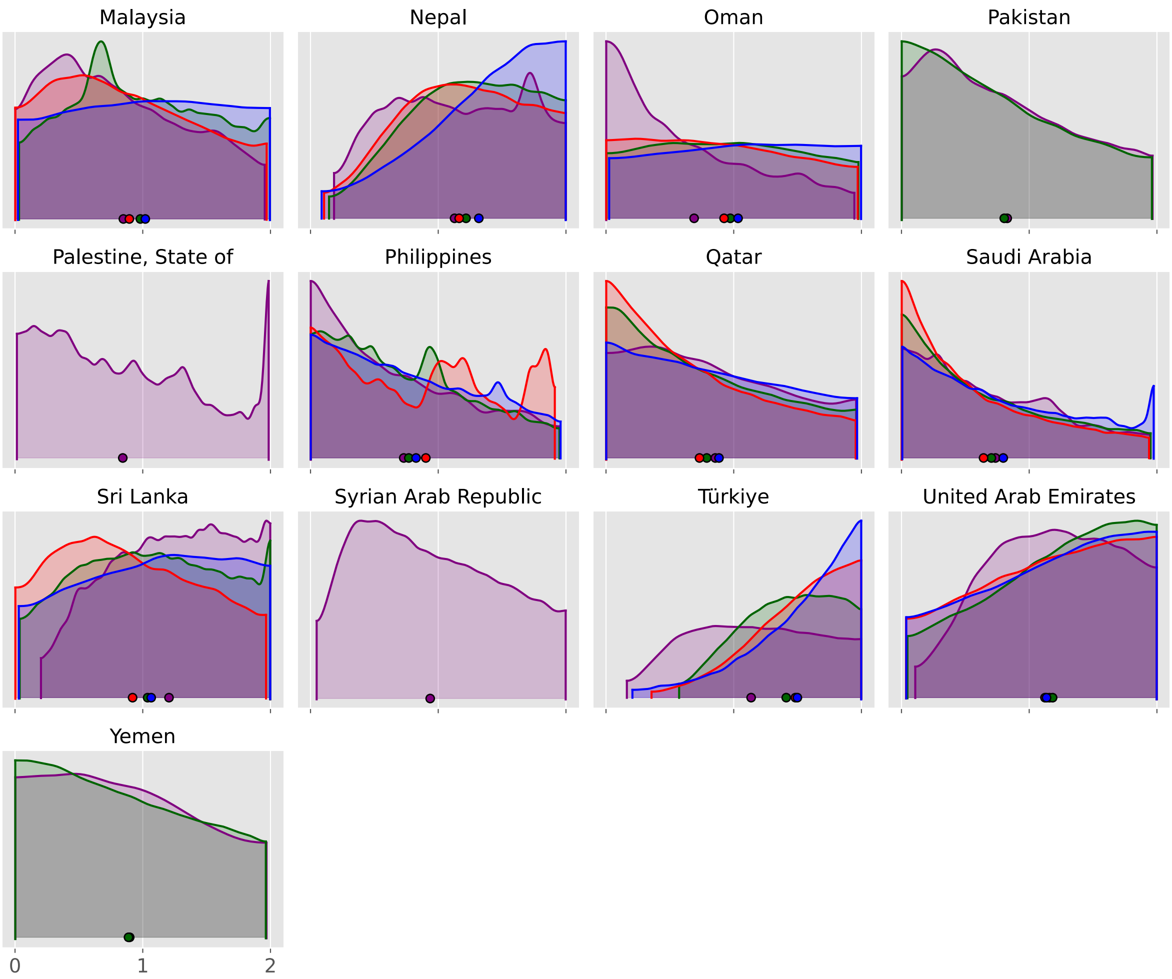
Malaysia
Nepal
Oman
Pakistan
Palestine, State of
Philippines
Qatar
Saudi Arabia
Sri Lanka
Syrian Arab Republic
Türkiye
United Arab Emirates
Yemen
0
1
2

#### Europe

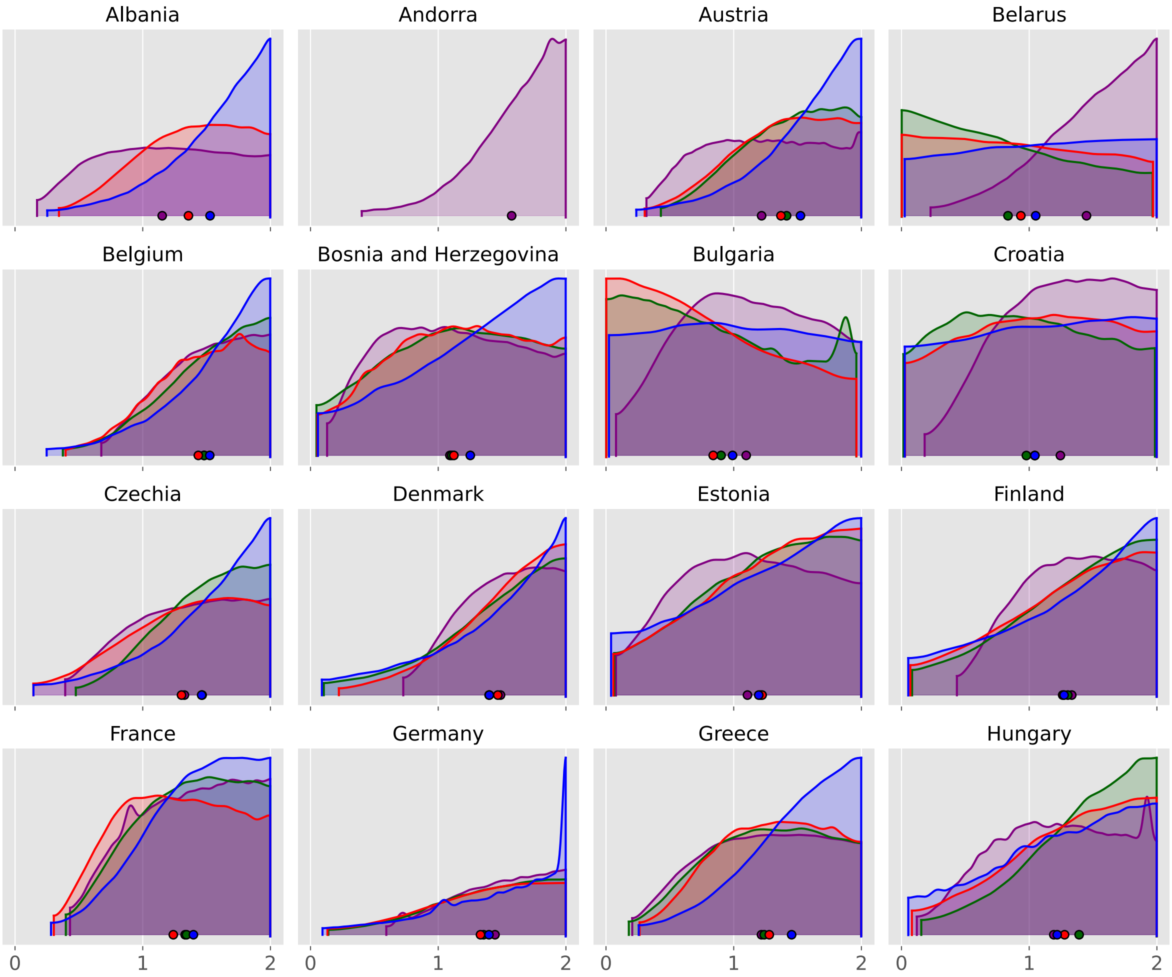
Albania
Andorra
Austria
Belarus
Belgium
Bosnia and Herzegovina
Bulgaria
Croatia
Czechia
Denmark
Estonia
Finland
France
Germany
Greece
Hungary
0
1
2
0
1
2
0
1
2
0
1
2

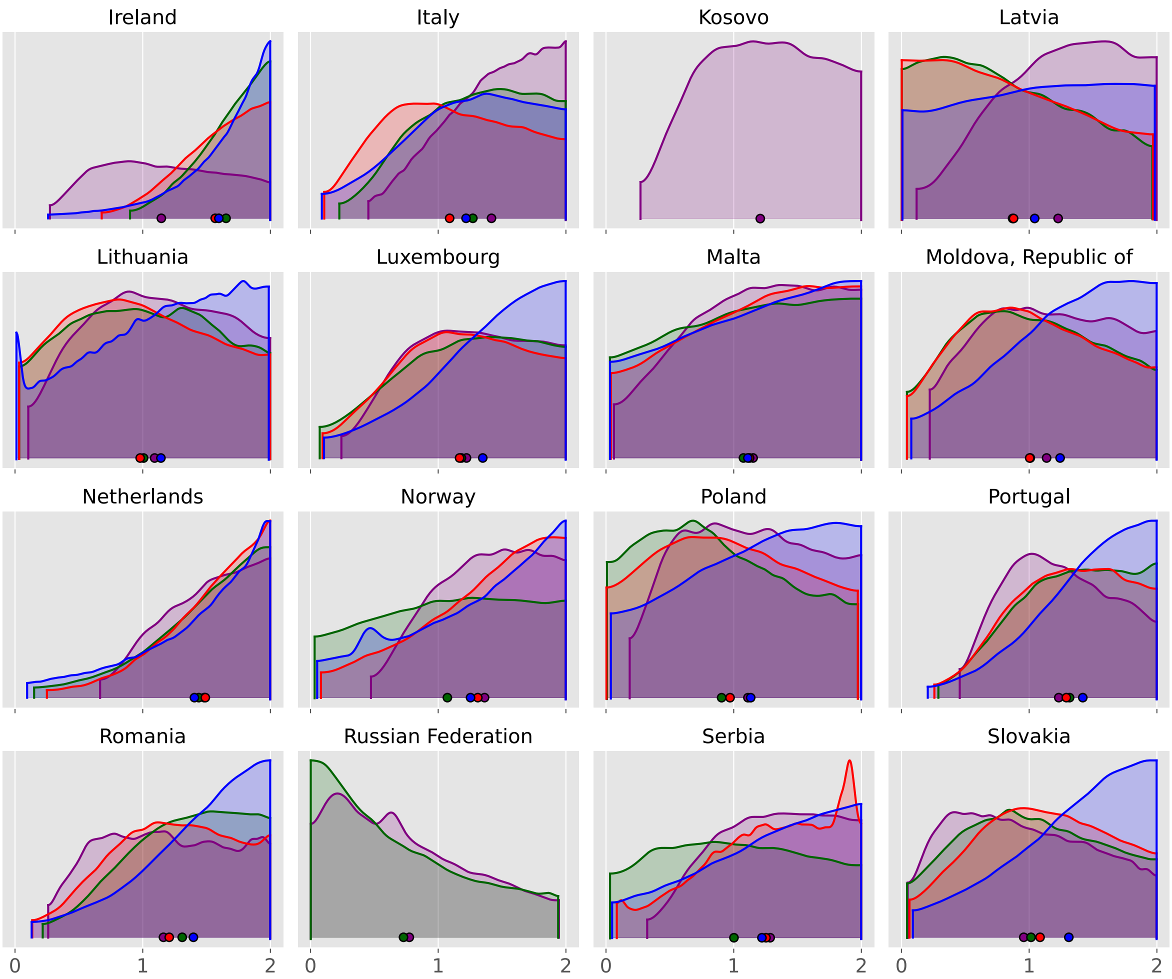
Ireland
Italy
Kosovo
Latvia
Lithuania
Luxembourg
Malta
Moldova, Republic of
Netherlands
Norway
Poland
Portugal
Romania
Russian Federation
Serbia
Slovakia
0
1
2

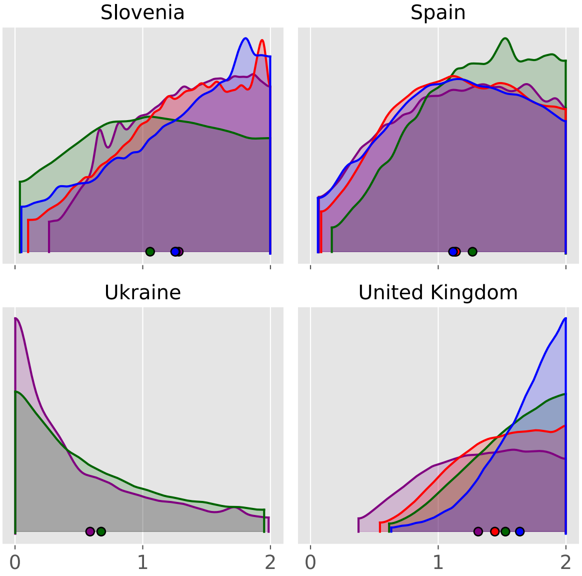
Slovenia
Spain
Sweden
Switzerland
Ukraine
United Kingdom
0
1
2
0
1
2

#### North America

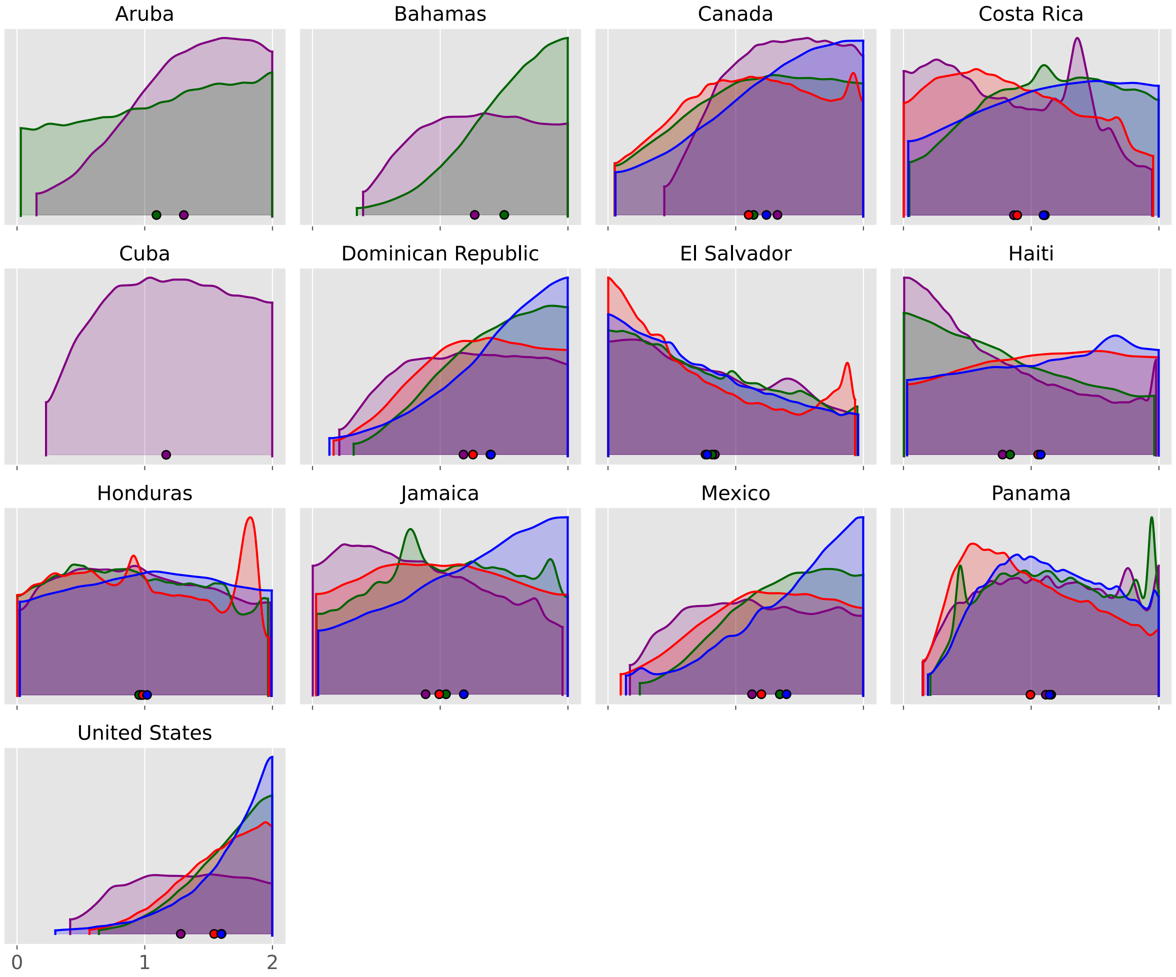

#### Oceania

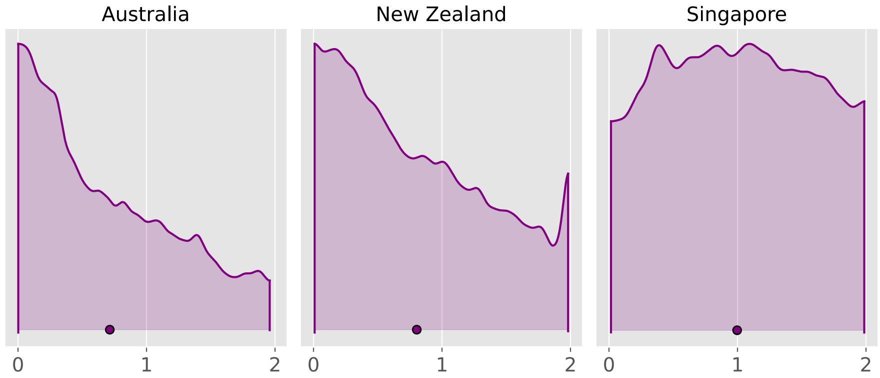


#### South America

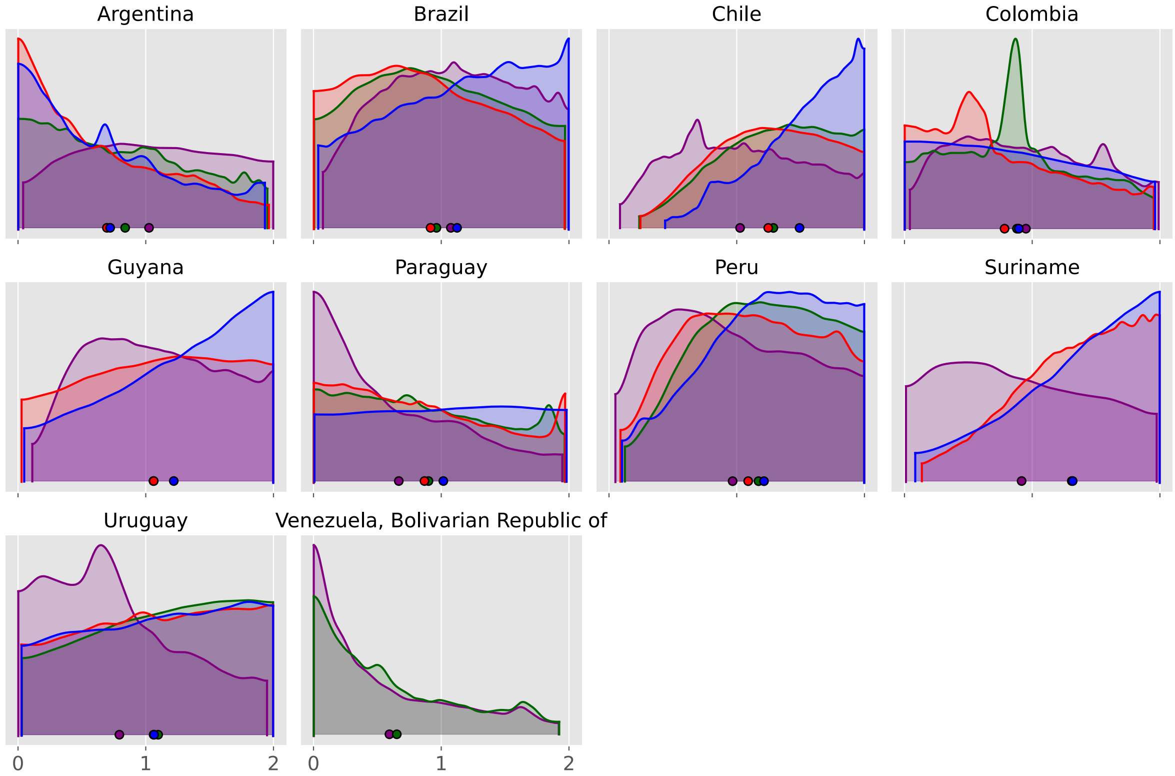

| | no scaling | OxCGRT floored | OxCGRT independent | Google mobility | Facebook tiles visited mobility | Facebook single tile mobility |
|---|---|---|---|---|---|---|
| Afghanistan | 4f57571 | b934196 | b934196 | b934196 | no analysis | no analysis |
| Albania | 4f57571 | b934196 | b934196 | no analysis | b934196 | b934196 |
| Algeria | 4f57571 | b934196 | b934196 | no analysis | b934196 | b934196 |
| Andorra | 4f57571 | b934196 | b934196 | no analysis | no analysis | no analysis |
| Angola | 4f57571 | b934196 | b934196 | cb8324a | cb8324a | cb8324a |
| Argentina | 4f57571 | b934196 | b934196 | b934196 | b934196 | b934196 |
| Aruba | 4f57571 | b934196 | b934196 | 4f57571 | no analysis | no analysis |
| Australia | b934196 | b934196 | b934196 | no analysis | no analysis | no analysis |
| Austria | 4f57571 | b934196 | b934196 | b934196 | b934196 | b934196 |
| Azerbaijan | 4f57571 | b934196 | b934196 | no analysis | no analysis | no analysis |
| Bahamas | 4f57571 | b934196 | b934196 | b934196 | no analysis | no analysis |
| Bahrain | 4f57571 | b934196 | b934196 | 4f57571 | no analysis | no analysis |
| Bangladesh | 4f57571 | b934196 | b934196 | b934196 | b934196 | b934196 |
| Belarus | 4f57571 | b934196 | b934196 | cb8324a | cb8324a | b934196 |
| Belgium | 4f57571 | b934196 | b934196 | 4f57571 | 4f57571 | 4f57571 |
| Benin | 4f57571 | b934196 | b934196 | 4f57571 | 4f57571 | b934196 |
| Bosnia and Herzegovina | 4f57571 | b934196 | b934196 | b934196 | b934196 | b934196 |
| Brazil | 4f57571 | b934196 | b934196 | 4f57571 | 4f57571 | 4f57571 |
| Bulgaria | 4f57571 | b934196 | b934196 | b934196 | b934196 | b934196 |
| Burkina Faso | 4f57571 | b934196 | b934196 | 4f57571 | 4f57571 | 4f57571 |
| Cabo Verde | 4f57571 | b934196 | b934196 | 4f57571 | no analysis | no analysis |
| Cameroon | 4f57571 | b934196 | b934196 | b934196 | b934196 | b934196 |
| Canada | 4f57571 | b934196 | b934196 | b934196 | b934196 | b934196 |
| Chile | 4f57571 | b934196 | b934196 | 4f57571 | b934196 | b934196 |
| Colombia | 4f57571 | b934196 | b934196 | 4f57571 | 4f57571 | b934196 |
| Congo | 4f57571 | b934196 | b934196 | no analysis | 4f57571 | b934196 |
| Costa Rica | 4f57571 | b934196 | b934196 | 4f57571 | b934196 | b934196 |
| Croatia | 4f57571 | b934196 | b934196 | b934196 | b934196 | b934196 |
| Cuba | 4f57571 | b934196 | b934196 | no analysis | no analysis | no analysis |
| Cyprus | 4f57571 | b934196 | b934196 | no analysis | no analysis | no analysis |
