## Supplemental File 9 for "Quantifying the contribution of policy measures to COVID-19 dynamics across 126 countries"

### Scaling strength

#### Table of contents

**Purpose** 1

**Map of scaling strengths** 1

**Numerical results** 2

#### Purpose

This document presents the composite scaling strength for the floored OxCGRT analysis,

$$1 - f^m,$$

the fractional reduction in $M_t$ from unrestricted ($S_t = 1$) to full restriction ($S_t = 0$) under

$$M_t = [f + (1 - f)\, S_t]^m.$$

The table also records whether residual-process dispersion improved relative to `no_scaling` (as in the dispersion comparisons document; yes if that probability exceeds 0.5). The map colours countries by strength: red where dispersion improved, grey where it did not. Countries without an estimate are hatched.

#### Map of scaling strengths

The following map presents the median scaling strength value ($1 - f^m$) for each country analysed as intensity of red colouring. Countries for which there was no evidence of an improvement in the ease of fitting to data following the inclusion of policy effects are still displayed, but the shading is shown in grey instead of red.

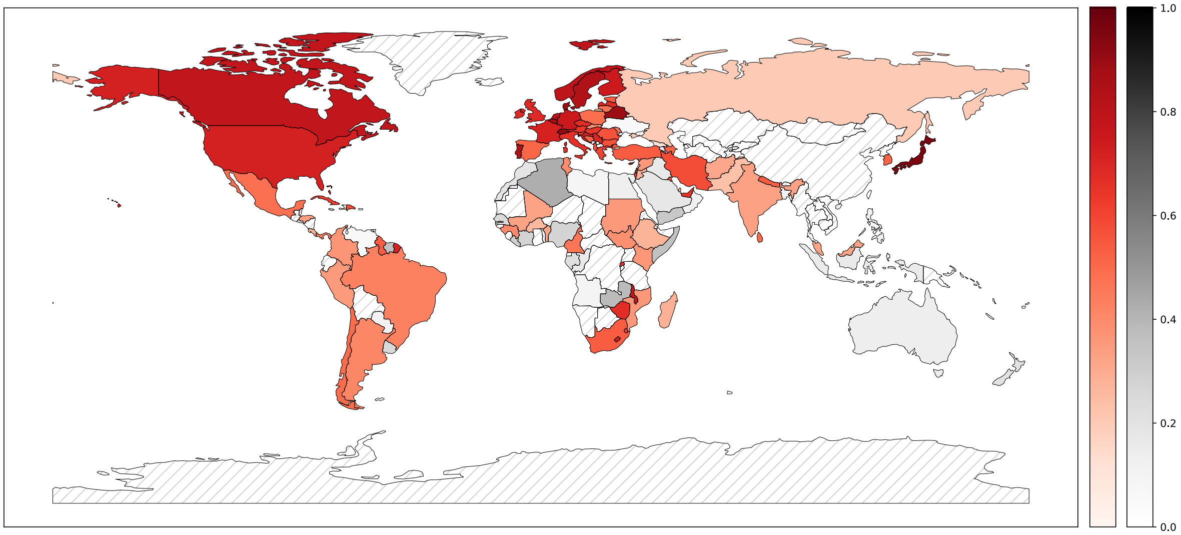


#### Numerical results

The same numerical results as displayed in the above map are presented in the following table, alongside an indication of whether ease of fitting improved. This is indicated as “yes” or “no” based on whether more than 50% of baseline runs showed a lower dispersion value than a randomly selected run from the OxCGRT floored analysis approach.

| | effect strength | dispersion improved |
|---|---|---|
| Algeria | 0.423 | no |
| Angola | 0.090 | no |
| Benin | 0.324 | yes |
| Burkina Faso | 0.256 | yes |
| Cabo Verde | 0.515 | yes |
| Cameroon | 0.463 | yes |
| Congo | 0.158 | no |
| Côte d’Ivoire | 0.271 | no |
| Egypt | 0.132 | no |
| Eswatini | 0.619 | yes |
| Ethiopia | 0.271 | yes |
| Gabon | 0.220 | no |
| Guinea | 0.401 | yes |
| Kenya | 0.361 | yes |
| Lesotho | 0.727 | yes |
| Liberia | 0.295 | no |

| | effect strength | dispersion improved |
|---|---|---|
| Libya | 0.084 | no |
| Madagascar | 0.284 | yes |
| Malawi | 0.779 | yes |
| Mali | 0.327 | yes |
| Morocco | 0.191 | no |
| Mozambique | 0.357 | yes |
| Nigeria | 0.272 | no |
| Rwanda | 0.637 | yes |
| Senegal | 0.249 | no |
| Somalia | 0.356 | no |
| South Africa | 0.533 | yes |
| South Sudan | 0.386 | yes |
| Sudan | 0.352 | yes |
| Togo | 0.124 | no |
| Tunisia | 0.397 | yes |
| Zambia | 0.383 | no |
| Zimbabwe | 0.675 | yes |
| Afghanistan | 0.309 | yes |
| Azerbaijan | 0.513 | yes |
| Bahrain | 0.263 | no |
| Bangladesh | 0.097 | no |
| Cyprus | 0.642 | yes |
| Georgia | 0.110 | yes |
| India | 0.329 | yes |
| Indonesia | 0.157 | no |
| Iran, Islamic Republic of | 0.578 | yes |
| Iraq | 0.160 | no |
| Israel | 0.598 | yes |
| Japan | 0.961 | yes |
| Jordan | 0.280 | yes |
| Korea, Republic of | 0.517 | yes |
| Kuwait | 0.565 | yes |
| Lebanon | 0.178 | yes |
| Malaysia | 0.322 | yes |
| Nepal | 0.535 | yes |
| Oman | 0.117 | no |
| Pakistan | 0.231 | yes |
| Palestine, State of | 0.222 | no |
| Philippines | 0.125 | no |
| Qatar | 0.272 | yes |
| Saudi Arabia | 0.173 | no |

| | effect strength | dispersion improved |
|---|---|---|
| Sri Lanka | 0.497 | yes |
| Syrian Arab Republic | 0.354 | yes |
| Türkiye | 0.533 | yes |
| United Arab Emirates | 0.655 | yes |
| Yemen | 0.325 | no |
| Albania | 0.577 | yes |
| Andorra | 0.949 | yes |
| Austria | 0.642 | yes |
| Belarus | 0.889 | yes |
| Belgium | 0.862 | yes |
| Bosnia and Herzegovina | 0.522 | yes |
| Bulgaria | 0.569 | yes |
| Croatia | 0.704 | yes |
| Czechia | 0.676 | yes |
| Denmark | 0.888 | yes |
| Estonia | 0.538 | yes |
| Finland | 0.743 | yes |
| France | 0.714 | yes |
| Germany | 0.751 | yes |
| Greece | 0.599 | yes |
| Hungary | 0.646 | yes |
| Ireland | 0.683 | yes |
| Italy | 0.661 | yes |
| Kosovo | 0.710 | yes |
| Latvia | 0.678 | yes |
| Lithuania | 0.474 | yes |
| Luxembourg | 0.693 | yes |
| Malta | 0.587 | yes |
| Moldova, Republic of | 0.449 | yes |
| Netherlands | 0.810 | yes |
| Norway | 0.791 | yes |
| Poland | 0.487 | yes |
| Portugal | 0.834 | yes |
| Romania | 0.561 | yes |
| Russian Federation | 0.203 | yes |
| Serbia | 0.697 | yes |
| Slovakia | 0.377 | yes |
| Slovenia | 0.623 | yes |
| Spain | 0.508 | yes |
| Sweden | 0.834 | yes |
| Switzerland | 0.900 | yes |

| | effect strength | dispersion improved |
|---|---|---|
| Ukraine | 0.062 | no |
| United Kingdom | 0.684 | yes |
| Aruba | 0.814 | yes |
| Bahamas | 0.793 | yes |
| Canada | 0.785 | yes |
| Costa Rica | 0.271 | yes |
| Cuba | 0.623 | yes |
| Dominican Republic | 0.582 | yes |
| El Salvador | 0.223 | no |
| Haiti | 0.162 | no |
| Honduras | 0.339 | yes |
| Jamaica | 0.315 | no |
| Mexico | 0.483 | yes |
| Panama | 0.430 | yes |
| United States | 0.720 | yes |
| Australia | 0.136 | no |
| New Zealand | 0.203 | no |
| Singapore | 0.458 | yes |
| Argentina | 0.407 | yes |
| Brazil | 0.428 | yes |
| Chile | 0.495 | yes |
| Colombia | 0.366 | yes |
| Guyana | 0.532 | yes |
| Paraguay | 0.107 | no |
| Peru | 0.352 | yes |
| Suriname | 0.355 | no |
| Uruguay | 0.254 | no |
| Venezuela, Bolivarian Republic of | 0.056 | no |
