## Supplemental File 10 for "Quantifying the contribution of policy measures to COVID-19 dynamics across 126 countries"

### Policy weight posteriors

**Commits used for analyses** **16**

#### Purpose

This document presents results for the weights for each policy domain of OxCGRT. To create an overall time series of policy restrictions, we weighted each contribution according to a calibrated parameter ranging between zero and one, which was then normalised to sum to one. The distribution of the parameter for each policy is presented as follows.

### Results by continent

#### Africa

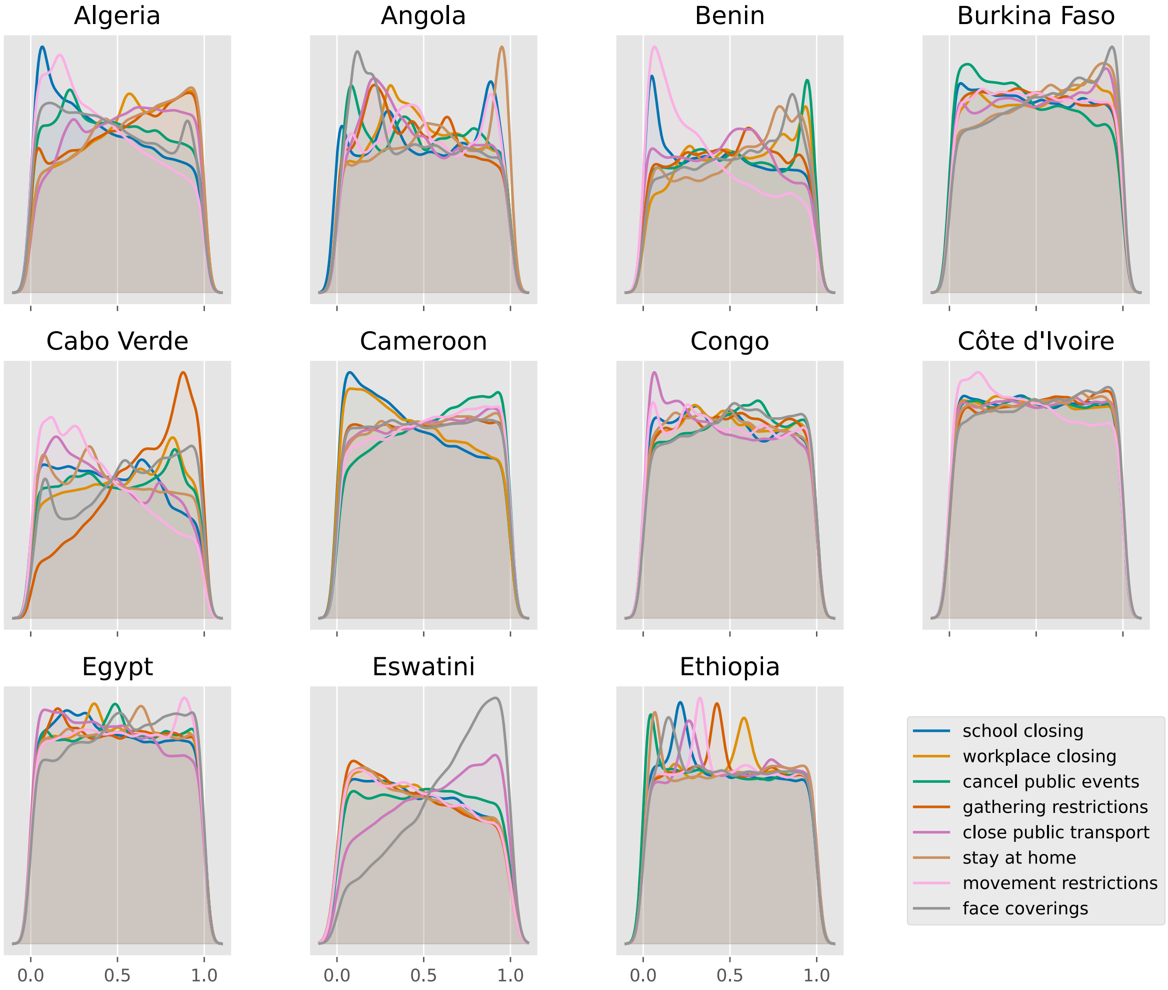

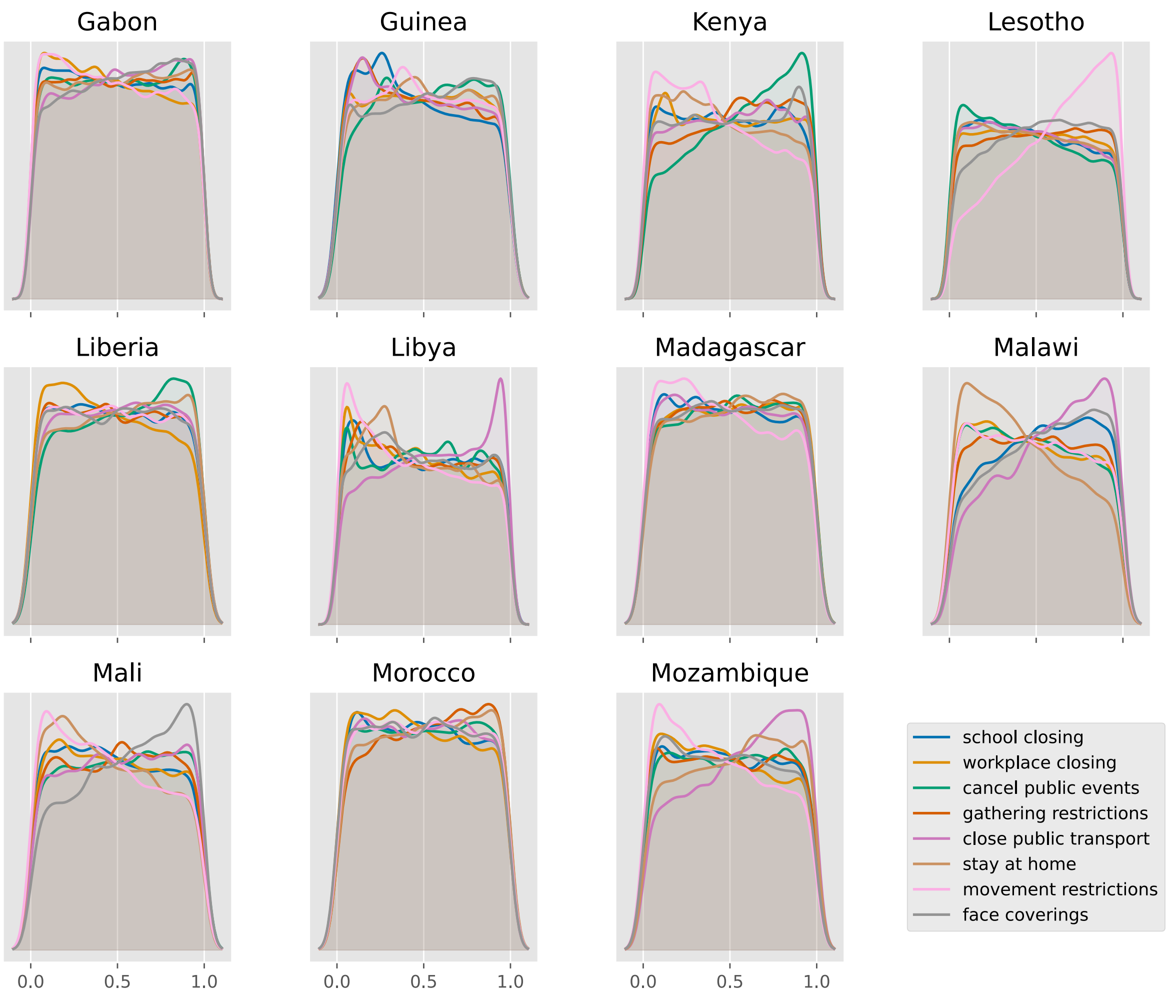
Gabon
Guinea
Kenya
Lesotho
Liberia
Libya
Madagascar
Malawi
Mali
Morocco
Mozambique
0.0
0.5
1.0
school closing
workplace closing
cancel public events
gathering restrictions
close public transport
stay at home
movement restrictions
face coverings

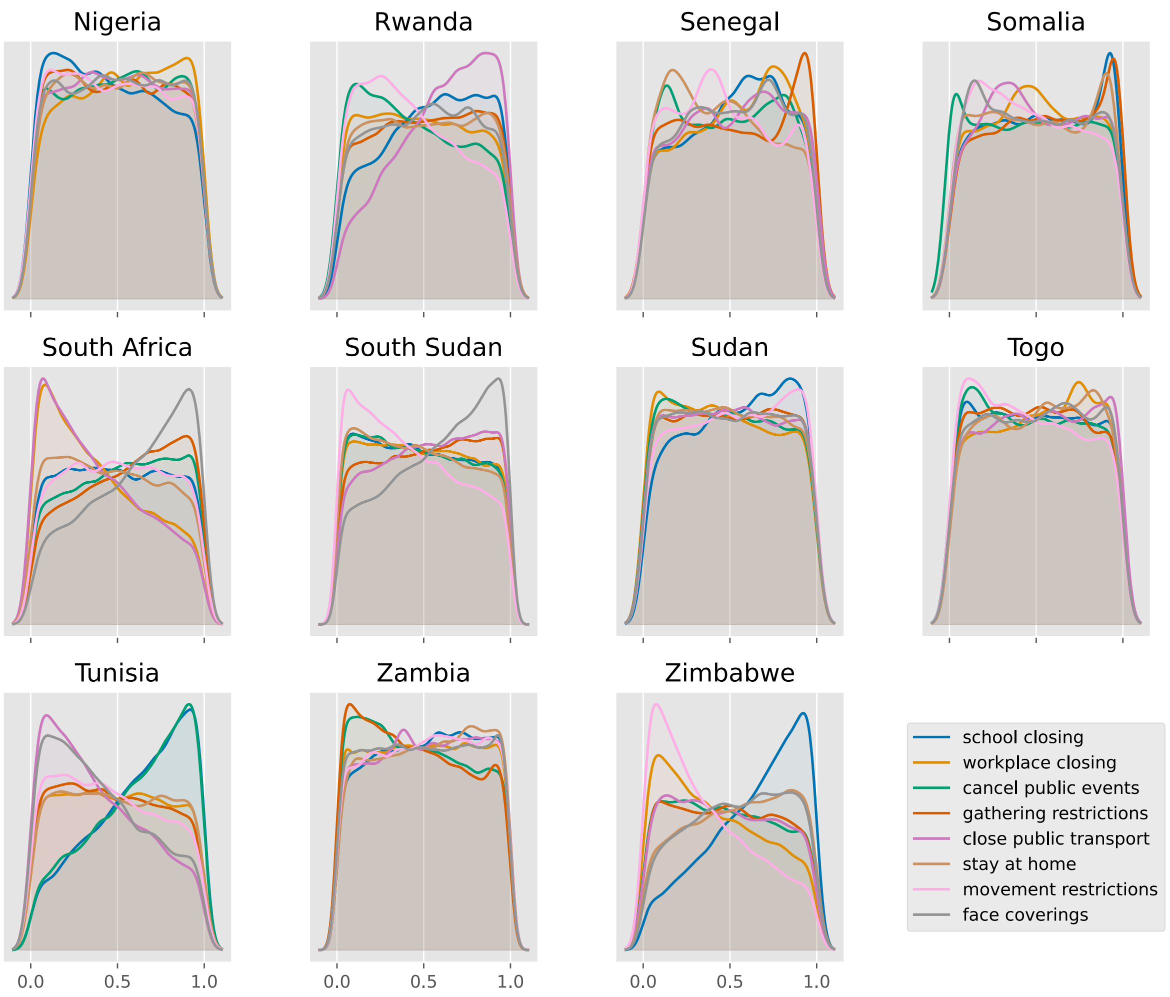
Nigeria
Rwanda
Senegal
Somalia
South Africa
South Sudan
Sudan
Togo
Tunisia
Zambia
Zimbabwe
0.0
0.5
1.0
school closing
workplace closing
cancel public events
gathering restrictions
close public transport
stay at home
movement restrictions
face coverings

#### Asia

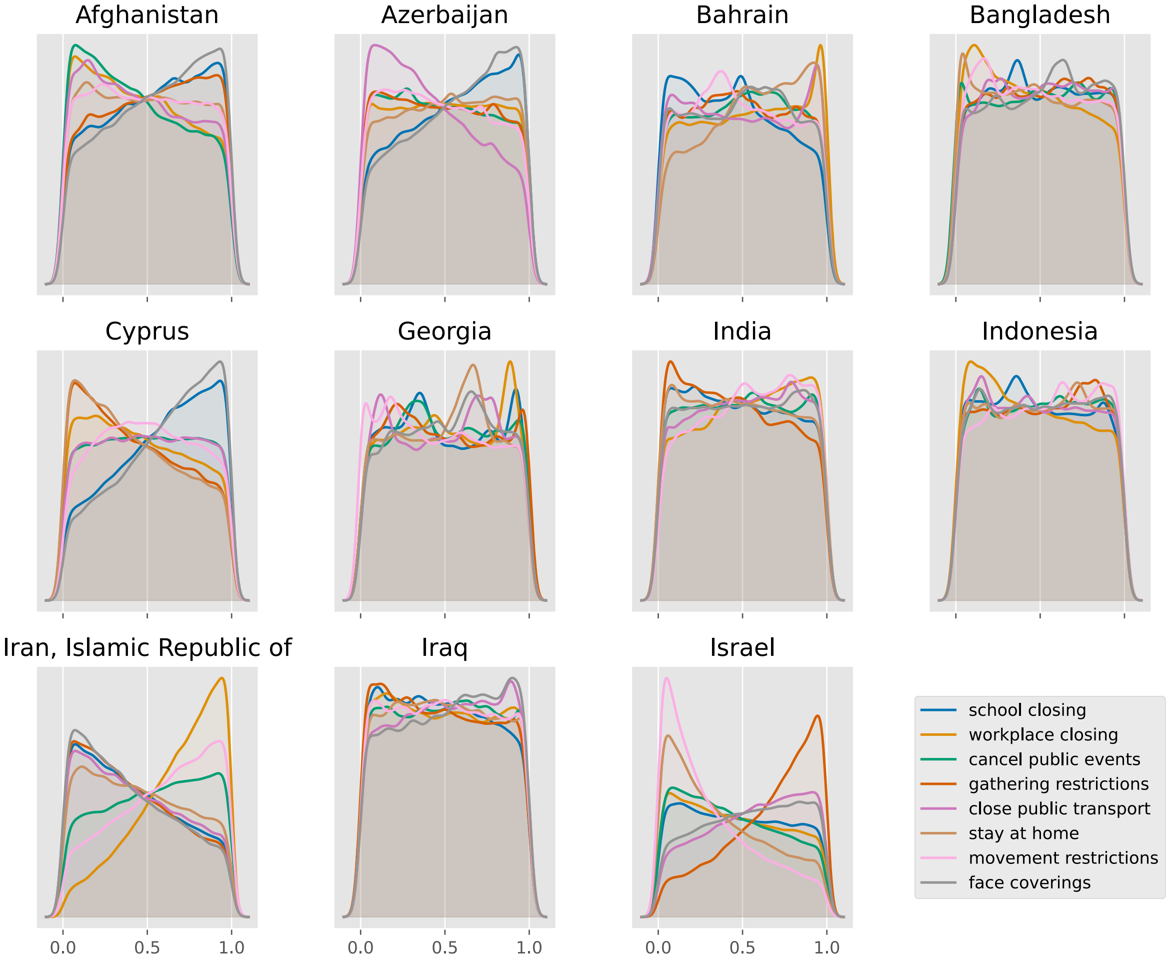
Afghanistan
Azerbaijan
Bahrain
Bangladesh
Cyprus
Georgia
India
Indonesia
Iran, Islamic Republic of
Iraq
Israel
0.0
0.5
1.0
school closing
workplace closing
cancel public events
gathering restrictions
close public transport
stay at home
movement restrictions
face coverings

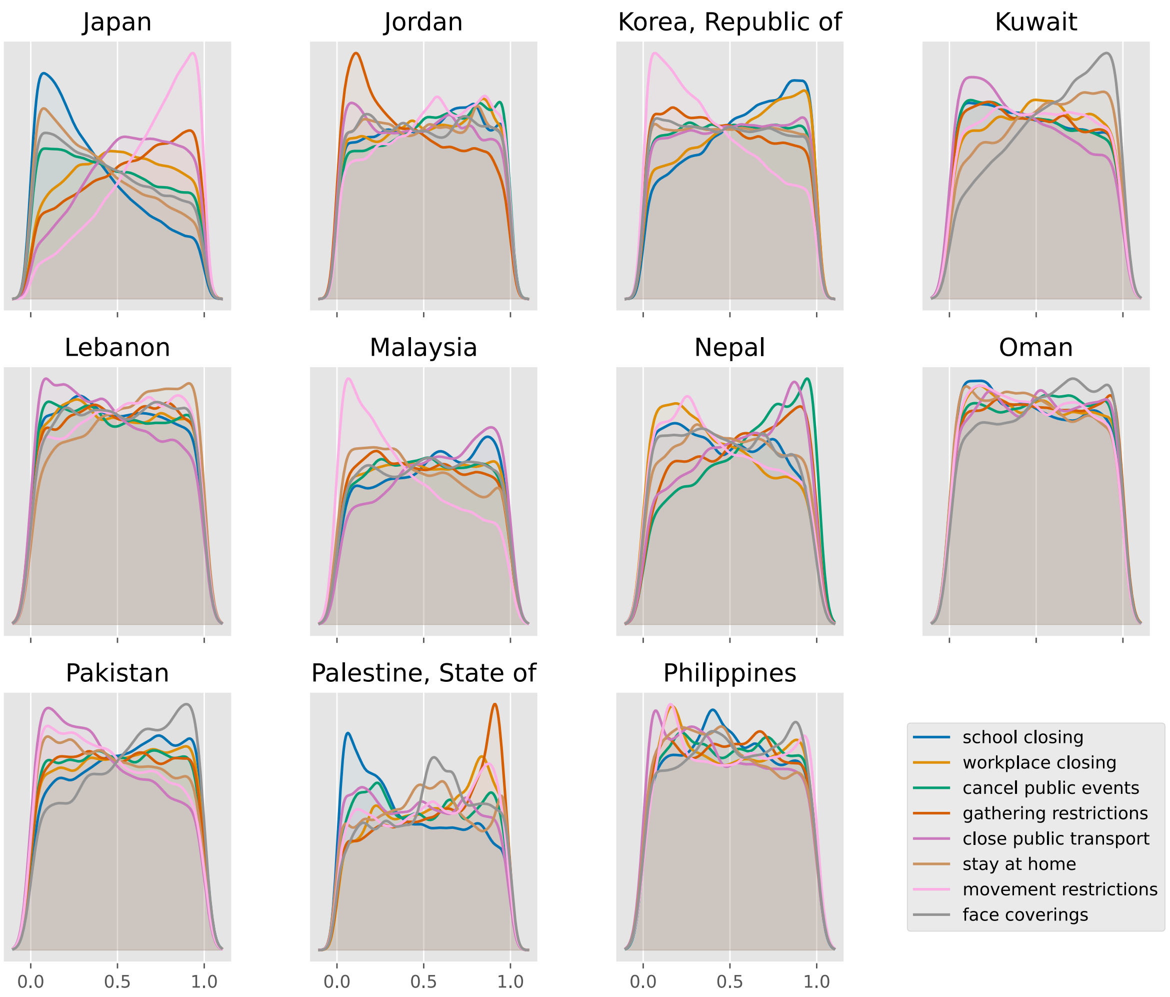
Japan
Jordan
Korea, Republic of
Kuwait
Lebanon
Malaysia
Nepal
Oman
Pakistan
Palestine, State of
Philippines
school closing
workplace closing
cancel public events
gathering restrictions
close public transport
stay at home
movement restrictions
face coverings
0.0
0.5
1.0

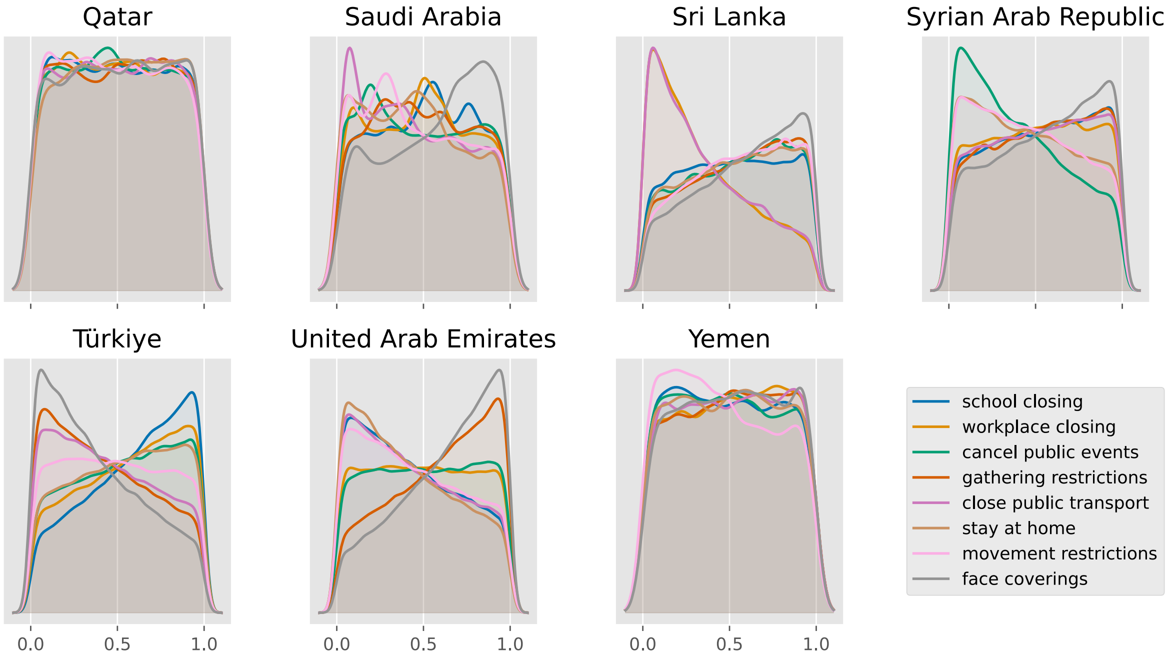
Qatar
Saudi Arabia
Sri Lanka
Syrian Arab Republic
Türkiye
United Arab Emirates
Yemen
0.0
0.5
1.0
school closing
workplace closing
cancel public events
gathering restrictions
close public transport
stay at home
movement restrictions
face coverings

#### Europe

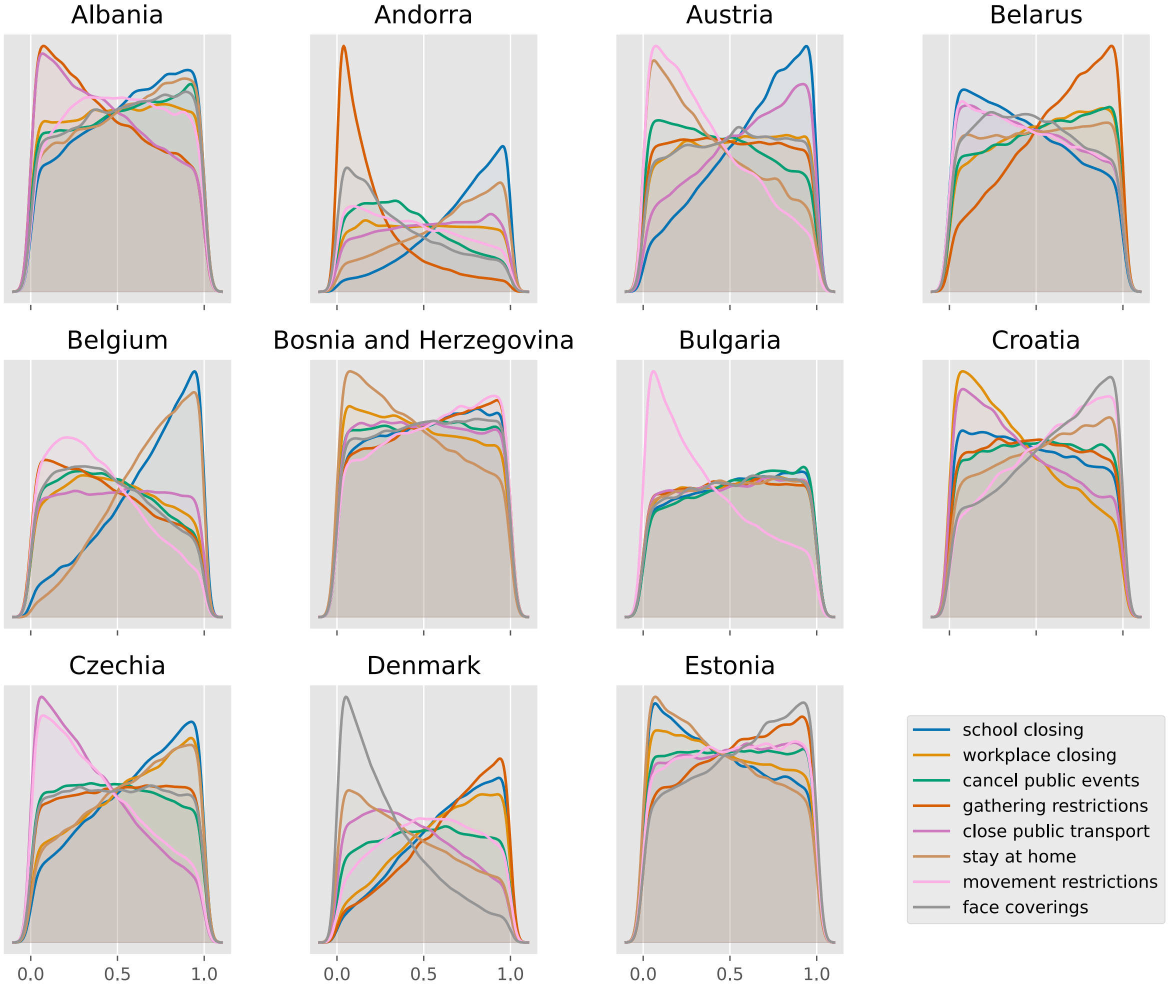

Albania
Andorra
Austria
Belarus
Belgium
Bosnia and Herzegovina
Bulgaria
Croatia
Czechia
Denmark
Estonia
0.0
0.5
1.0
school closing
workplace closing
cancel public events
gathering restrictions
close public transport
stay at home
movement restrictions
face coverings

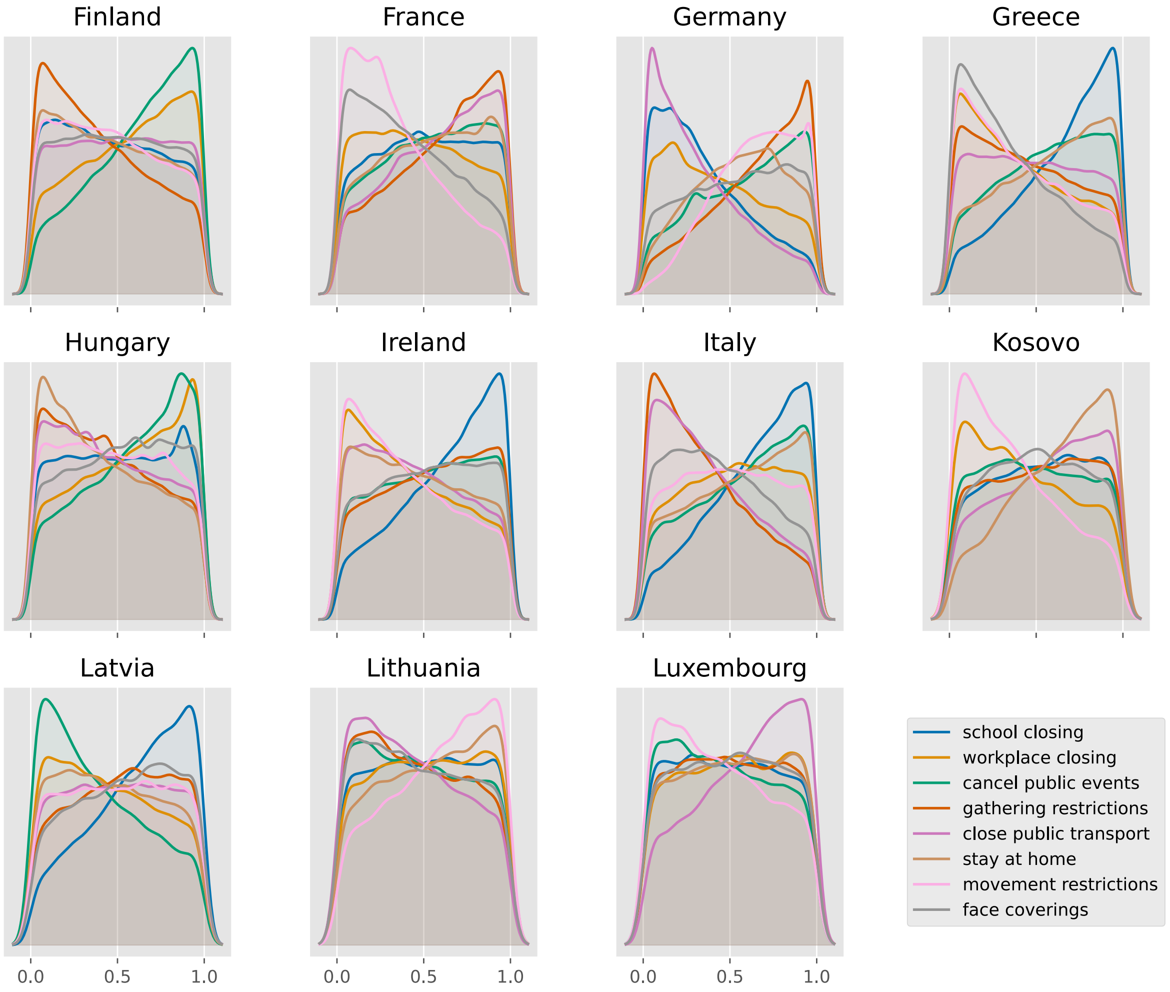
Finland
France
Germany
Greece
Hungary
Ireland
Italy
Kosovo
Latvia
Lithuania
Luxembourg
0.0
0.5
1.0
school closing
workplace closing
cancel public events
gathering restrictions
close public transport
stay at home
movement restrictions
face coverings

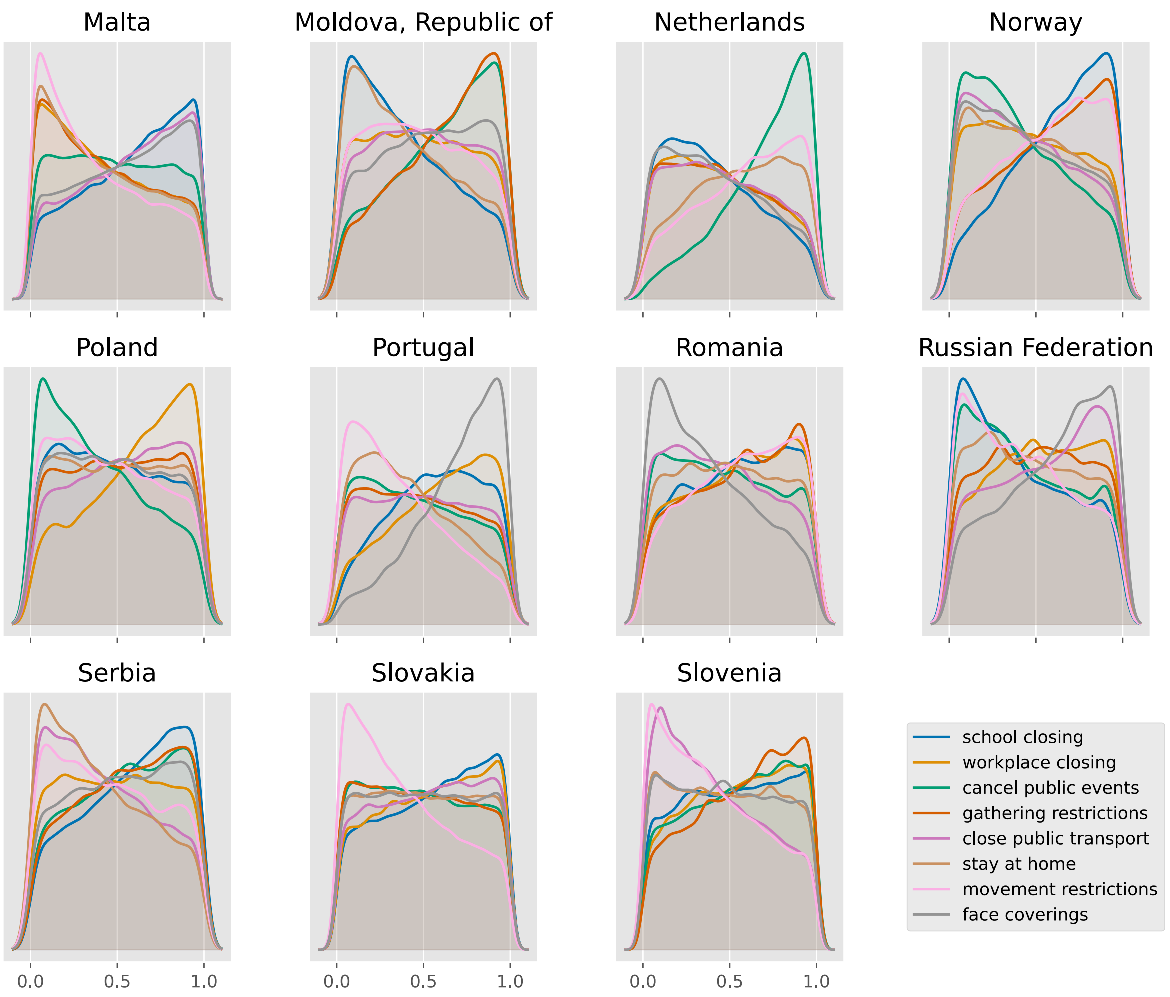
Malta
Moldova, Republic of
Netherlands
Norway
Poland
Portugal
Romania
Russian Federation
Serbia
Slovakia
Slovenia
0.0
0.5
1.0
school closing
workplace closing
cancel public events
gathering restrictions
close public transport
stay at home
movement restrictions
face coverings

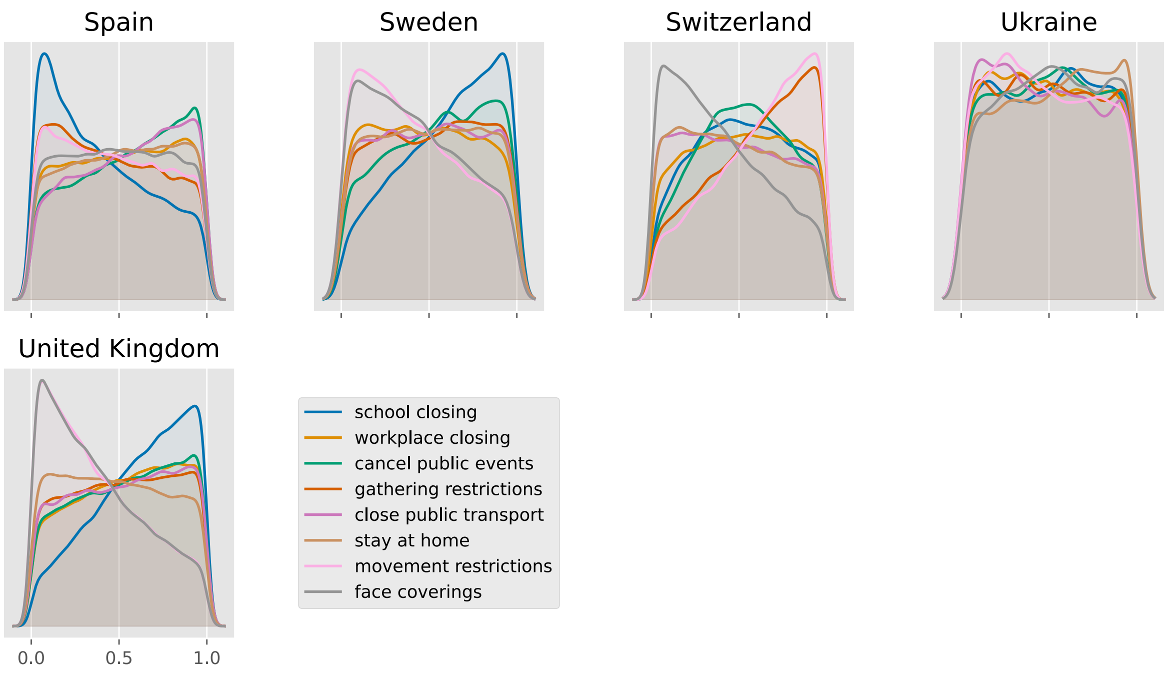
Spain
Sweden
Switzerland
Ukraine
United Kingdom
0.0
0.5
1.0
school closing
workplace closing
cancel public events
gathering restrictions
close public transport
stay at home
movement restrictions
face coverings

#### North America

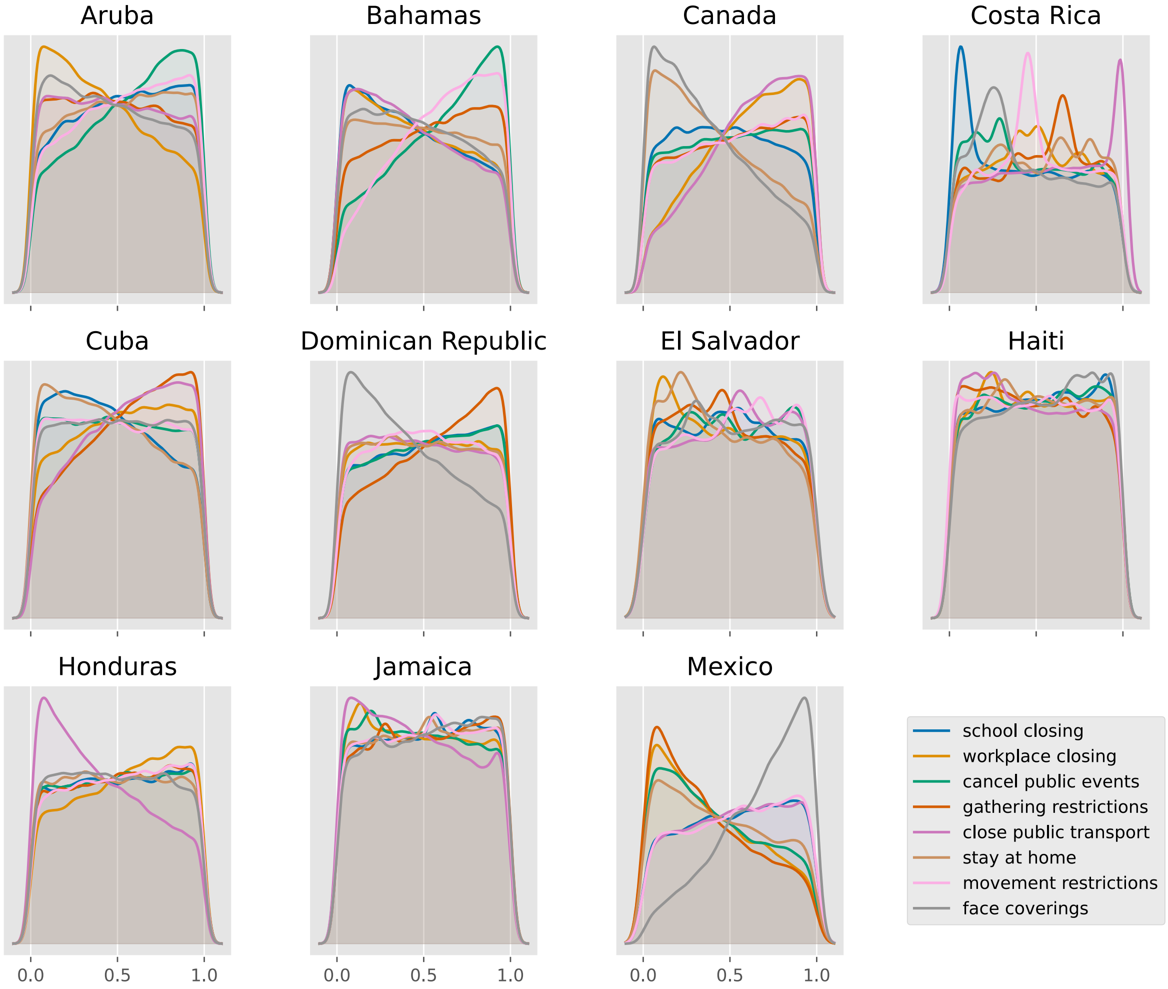

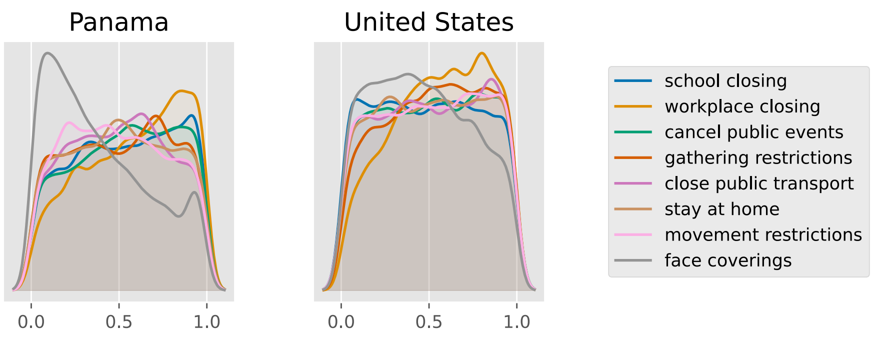
Panama
United States
0.0
0.5
1.0
0.0
0.5
1.0
school closing
workplace closing
cancel public events
gathering restrictions
close public transport
stay at home
movement restrictions
face coverings

#### Oceania

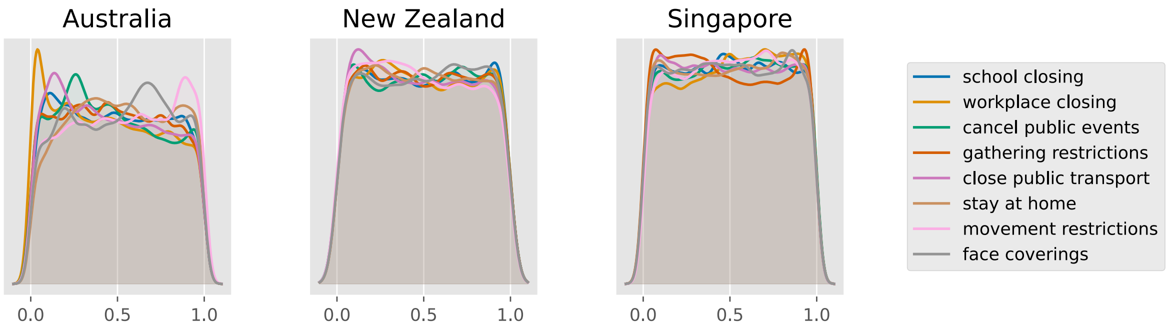

Australia
New Zealand
Singapore
0.0
0.5
1.0
school closing
workplace closing
cancel public events
gathering restrictions
close public transport
stay at home
movement restrictions
face coverings

#### South America

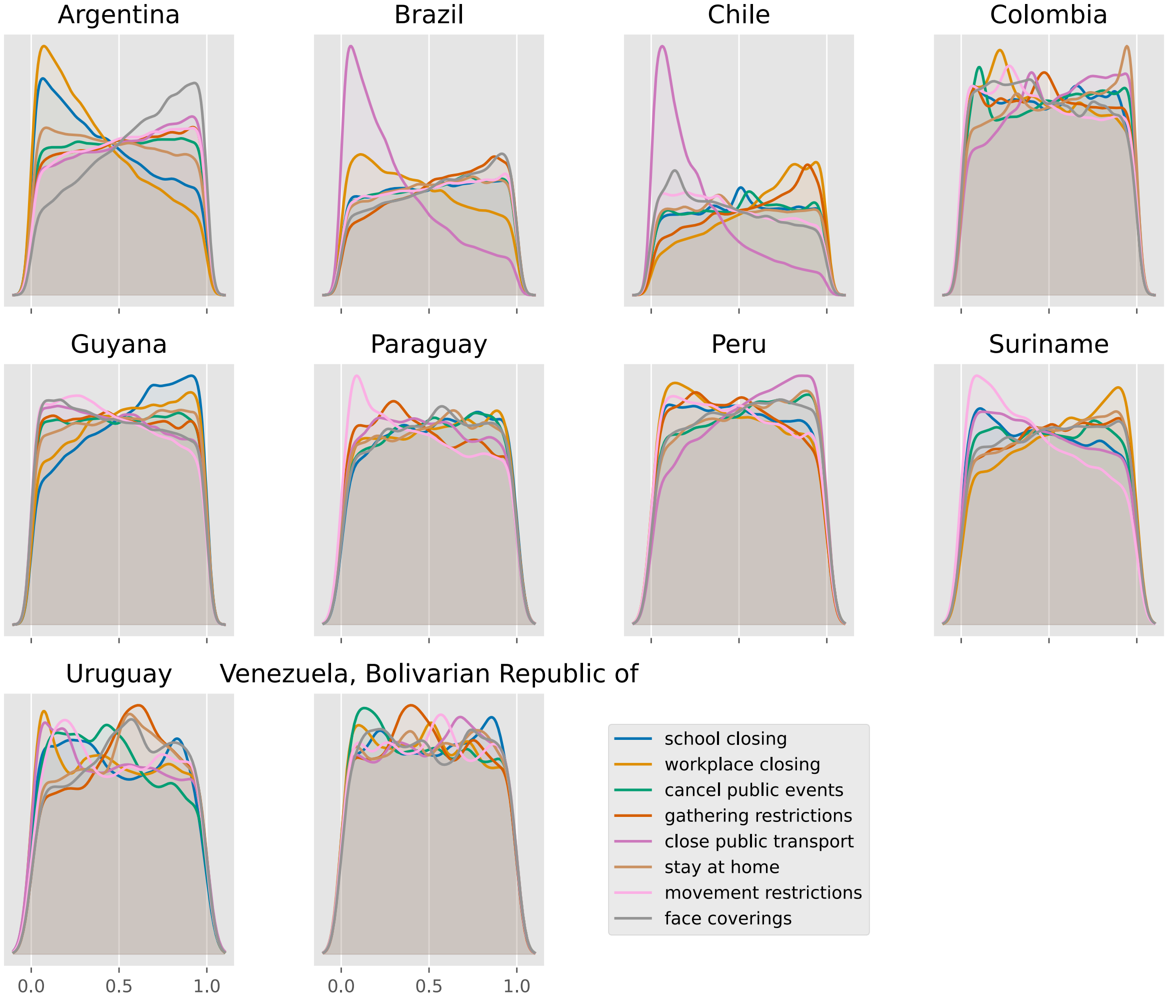

#### Commits used for analyses

For reproducibility, the following table gives the (short) commit SHA for each analysis.

| | no scaling | OxCGRT floored | OxCGRT independent | Google mobility | Facebook tiles visited mobility | Facebook single tile mobility |
|---|---|---|---|---|---|---|
| Afghanistan | 4f57571 | b934196 | b934196 | b934196 | no analysis | no analysis |
| Albania | 4f57571 | b934196 | b934196 | no analysis | b934196 | b934196 |
| Algeria | 4f57571 | b934196 | b934196 | no analysis | b934196 | b934196 |
| Andorra | 4f57571 | b934196 | b934196 | no analysis | no analysis | no analysis |
| Angola | 4f57571 | b934196 | b934196 | cb8324a | cb8324a | cb8324a |
| Argentina | 4f57571 | b934196 | b934196 | b934196 | b934196 | b934196 |
| Aruba | 4f57571 | b934196 | b934196 | 4f57571 | no analysis | no analysis |
| Australia | b934196 | b934196 | b934196 | no analysis | no analysis | no analysis |
| Austria | 4f57571 | b934196 | b934196 | b934196 | b934196 | b934196 |
| Azerbaijan | 4f57571 | b934196 | b934196 | no analysis | no analysis | no analysis |
| Bahamas | 4f57571 | b934196 | b934196 | b934196 | no analysis | no analysis |
| Bahrain | 4f57571 | b934196 | b934196 | 4f57571 | no analysis | no analysis |
| Bangladesh | 4f57571 | b934196 | b934196 | b934196 | b934196 | b934196 |
| Belarus | 4f57571 | b934196 | b934196 | cb8324a | cb8324a | b934196 |
| Belgium | 4f57571 | b934196 | b934196 | 4f57571 | 4f57571 | 4f57571 |
| Benin | 4f57571 | b934196 | b934196 | 4f57571 | 4f57571 | b934196 |
| Bosnia and Herzegovina | 4f57571 | b934196 | b934196 | b934196 | b934196 | b934196 |
| Brazil | 4f57571 | b934196 | b934196 | 4f57571 | 4f57571 | 4f57571 |
| Bulgaria | 4f57571 | b934196 | b934196 | b934196 | b934196 | b934196 |
| Burkina Faso | 4f57571 | b934196 | b934196 | 4f57571 | 4f57571 | 4f57571 |
| Cabo Verde | 4f57571 | b934196 | b934196 | 4f57571 | no analysis | no analysis |
| Cameroon | 4f57571 | b934196 | b934196 | b934196 | b934196 | b934196 |
| Canada | 4f57571 | b934196 | b934196 | b934196 | b934196 | b934196 |
| Chile | 4f57571 | b934196 | b934196 | 4f57571 | b934196 | b934196 |
| Colombia | 4f57571 | b934196 | b934196 | 4f57571 | 4f57571 | b934196 |
| Congo | 4f57571 | b934196 | b934196 | no analysis | 4f57571 | b934196 |
| Costa Rica | 4f57571 | b934196 | b934196 | 4f57571 | b934196 | b934196 |
| Croatia | 4f57571 | b934196 | b934196 | b934196 | b934196 | b934196 |
