## Supplemental File 11 for "Quantifying the contribution of policy measures to COVID-19 dynamics across 126 countries"

### Policy weight matrix

**Countries omitted from the matrix** 9

#### Purpose

This document presents the calibrated weight for each OxCGRT policy domain of the floored OxCGRT analysis as a country by policy matrix.

Only the normalised weights $\omega_p / \sum_q \omega_q$ enter the likelihood, such that the overall scale of the weight vector is non-identified and the posterior median of each unnormalised weight is uninformative. By contrast, the data are able to identify the within-country tilt among domains for certain countries. We therefore plot the boundary-mass log-odds:

$$B_p = \log \frac{\widehat{P}(\omega_p > 1 - \tau)}{\widehat{P}(\omega_p < \tau)}, \qquad \tau = 0.1,$$

for a given country and policy domain $p$. Under the $U[0,1]$ prior both tails carry mass $\tau$, so $B = 0$ is the prior value. Positive $B$ (red) indicates mass piled toward the upper boundary (domain up-weighted relative to the country's other policies), whereas negative $B$ (blue) indicates down-weighting.

Using the same convention as in the preceding documents, only results for countries for which residual-process dispersion improved relative to `no_scaling` are presented. For the remaining countries, the analysis provides no evidence that policy contributed to transmission, such that the weights should not be interpreted and so are not displayed.

Singapore is shown with Asia here, although the epidemiological implementation is aligned with Oceania otherwise. This is because it is the only country of this group for which the dispersion posterior criterion was reached (Australia and New Zealand excluded).

$B$ is a relative within-country statement: a country in which all eight domains contributed equally gives $B \approx 0$ across the row and is indistinguishable from a country in which the calibration learned nothing. It should therefore be read alongside the dispersion evidence and the composite scaling strength $1 - f^m$ shown in the final column (presented in greater detail in Supplement 09). Supplement 10 provides the full distribution of the policy weights.

### Results by continent

#### Africa

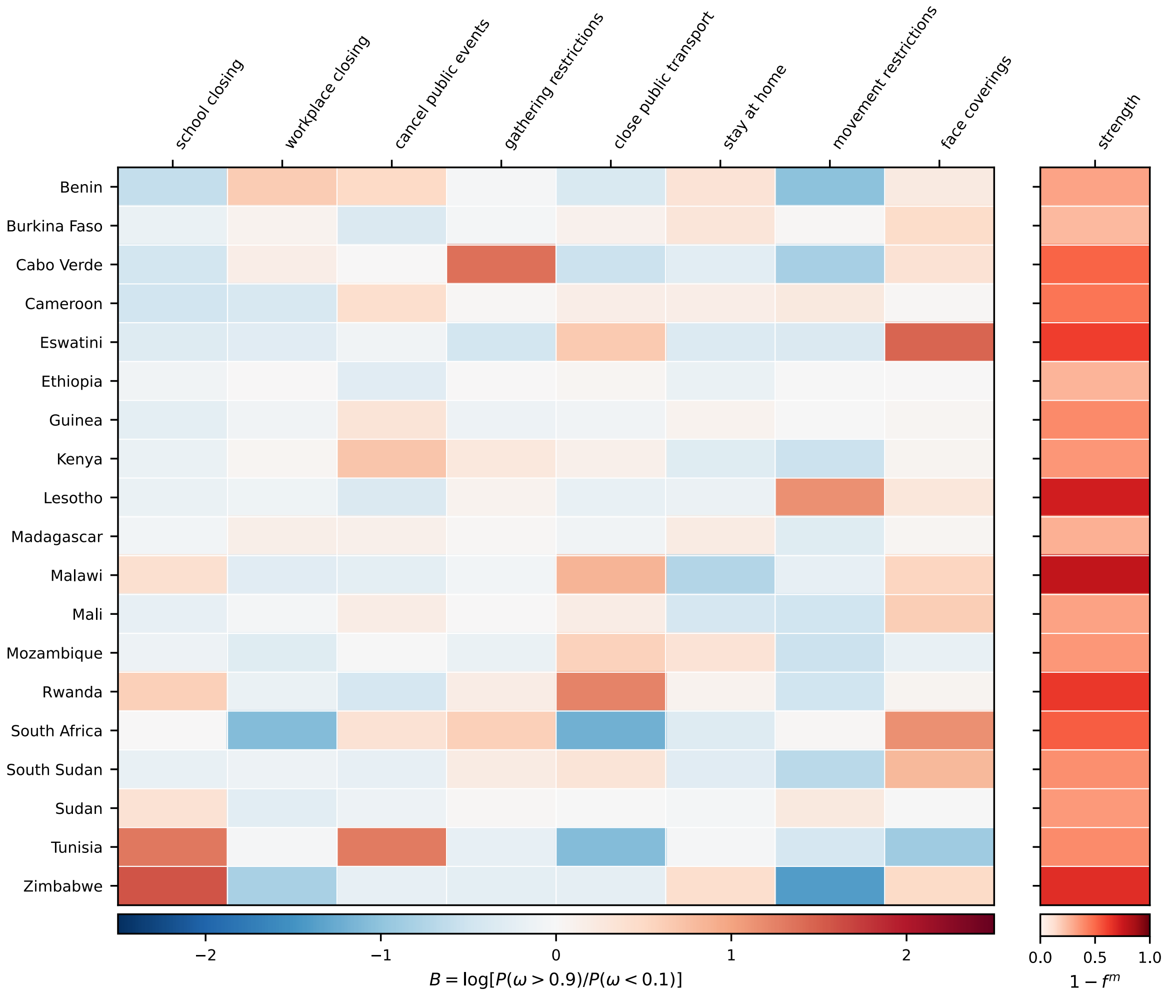

#### Asia

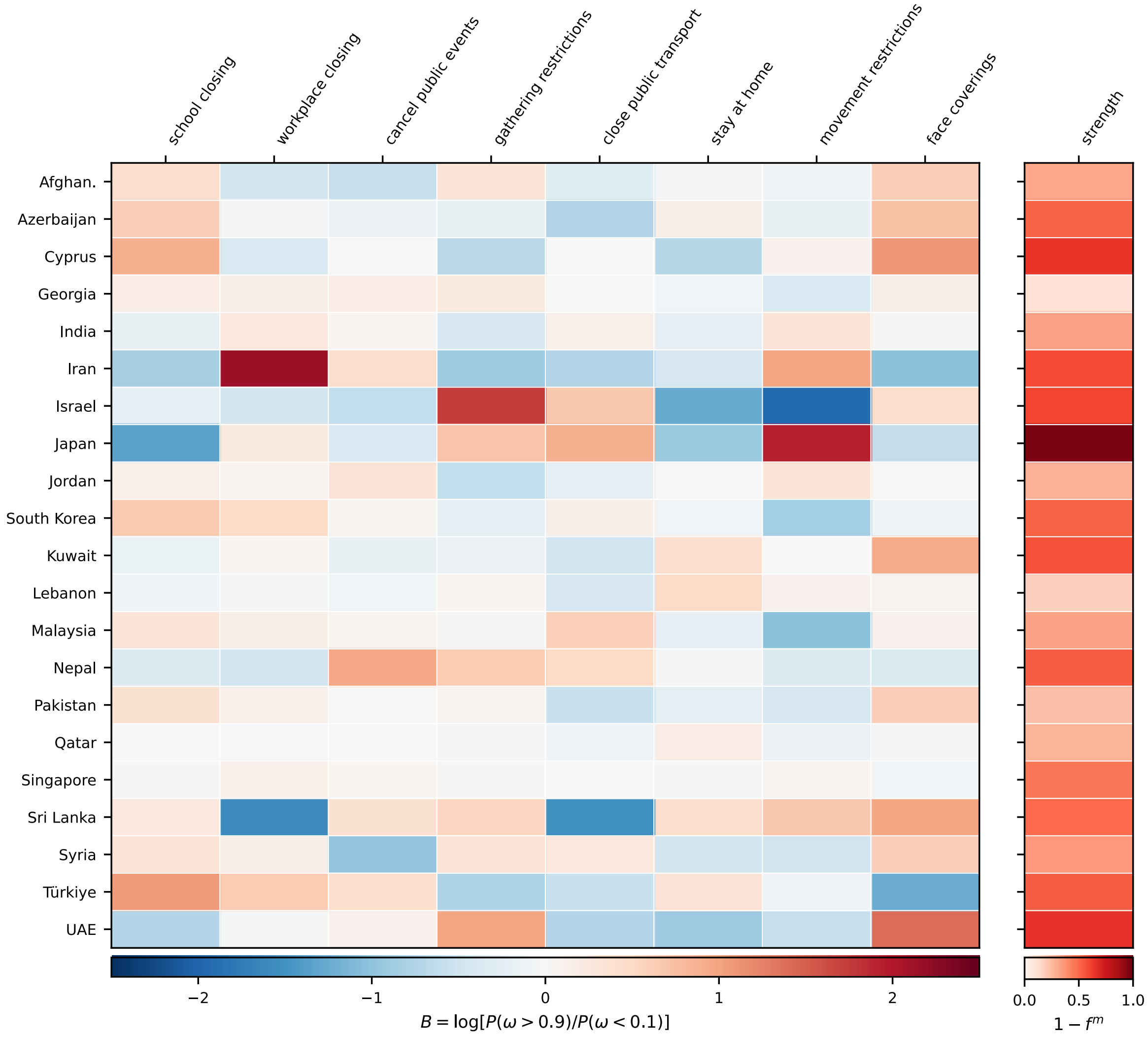
school closing
workplace closing
cancel public events
gathering restrictions
close public transport
stay at home
movement restrictions
face coverings
strength
Afghan.
Azerbaijan
Cyprus
Georgia
India
Iran
Israel
Japan
Jordan
South Korea
Kuwait
Lebanon
Malaysia
Nepal
Pakistan
Qatar
Singapore
Sri Lanka
Syria
Türkiye
UAE
−2
−1
0
1
2
$B = \log[P(\omega > 0.9)/P(\omega < 0.1)]$
0.0
0.5
1.0
$1 - f^m$

#### Europe

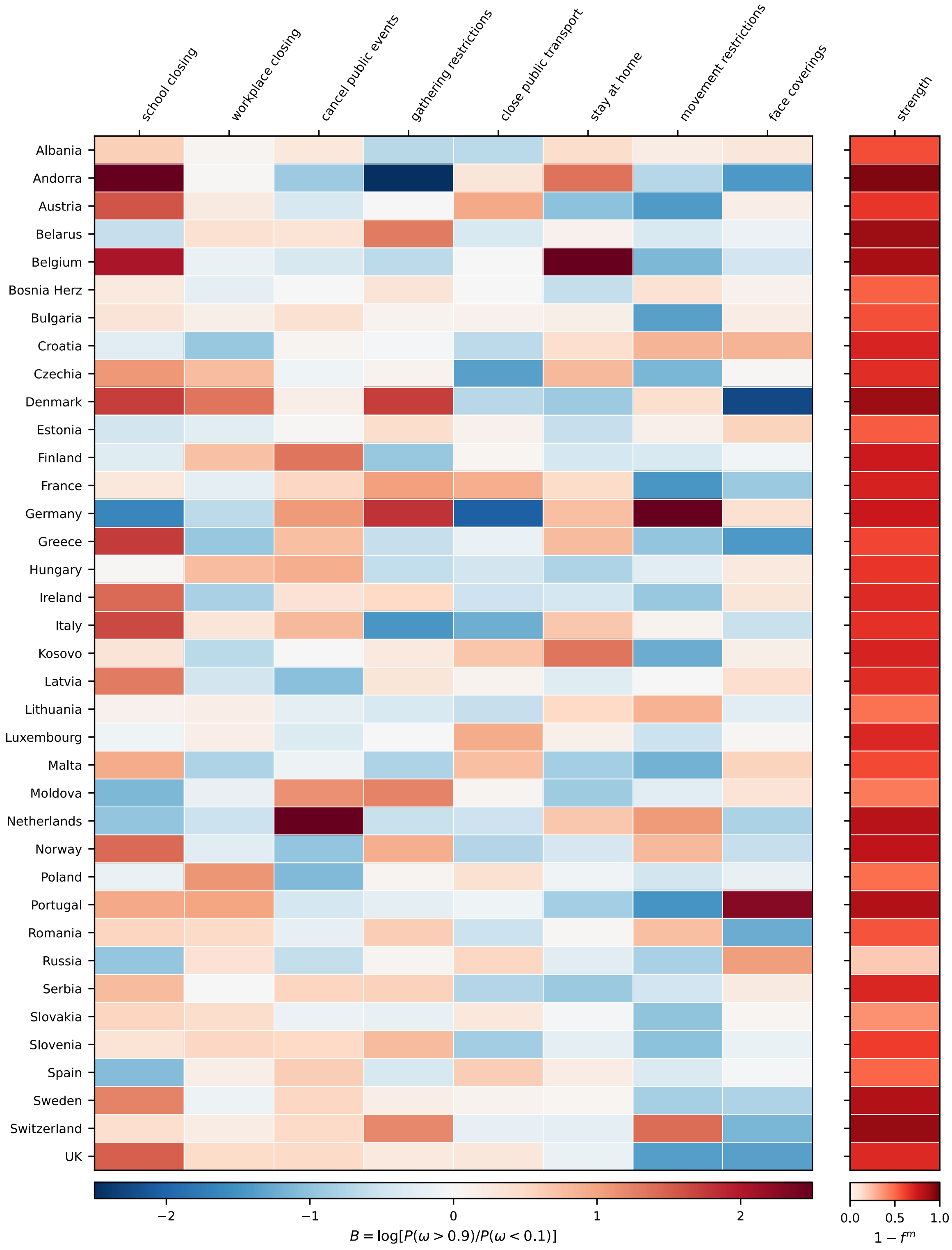

school closing
workplace closing
cancel public events
gathering restrictions
close public transport
stay at home
movement restrictions
face coverings
strength
Albania
Andorra
Austria
Belarus
Belgium
Bosnia Herz
Bulgaria
Croatia
Czechia
Denmark
Estonia
Finland
France
Germany
Greece
Hungary
Ireland
Italy
Kosovo
Latvia
Lithuania
Luxembourg
Malta
Moldova
Netherlands
Norway
Poland
Portugal
Romania
Russia
Serbia
Slovakia
Slovenia
Spain
Sweden
Switzerland
UK
−2
−1
0
1
2
$B = \log[P(\omega > 0.9)/P(\omega < 0.1)]$
0.0
0.5
1.0
$1 - f^m$

#### North America

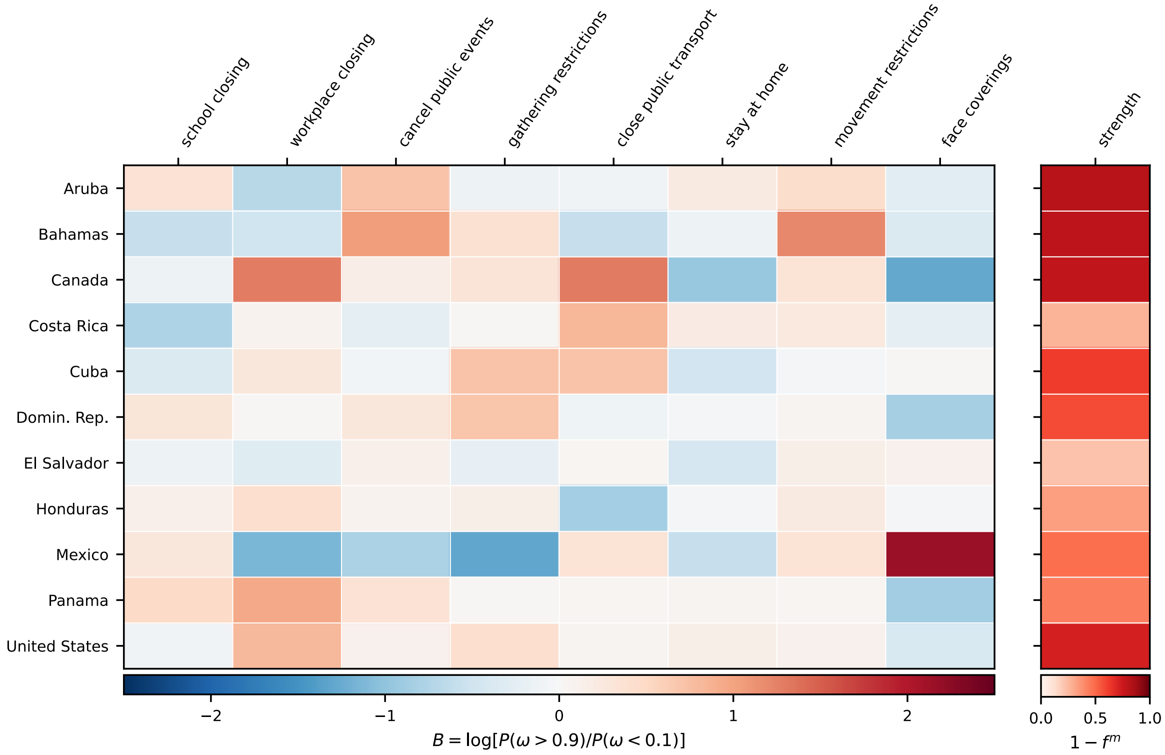

#### South America

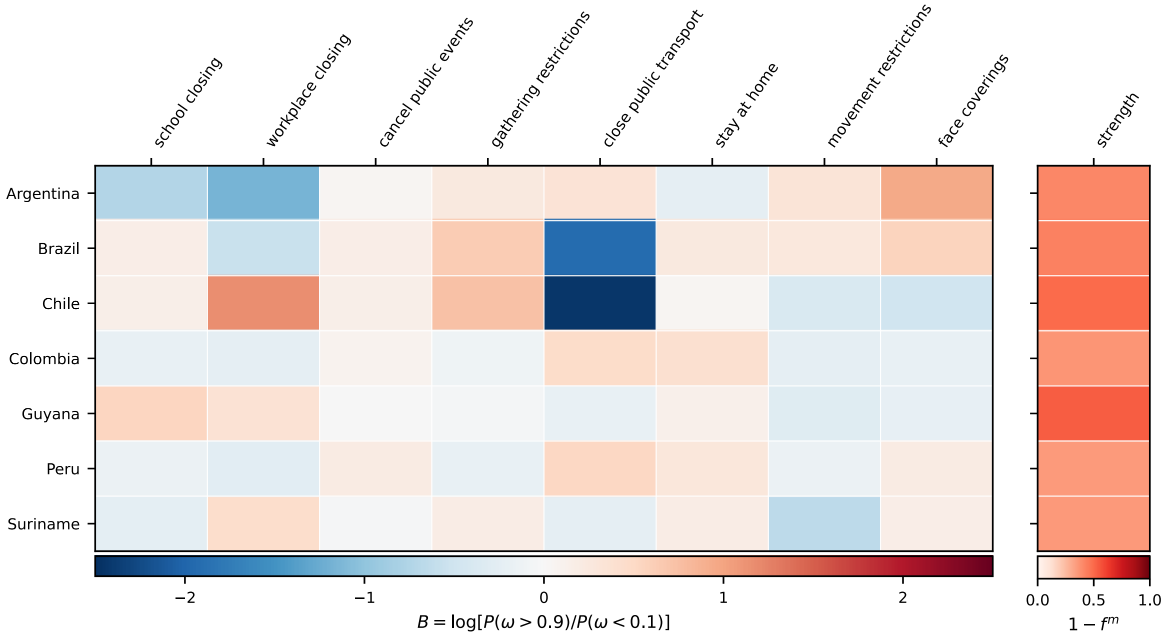

#### Countries omitted from the matrix

The following countries were analysed under the floored OxCGRT approach, but did not show an improvement in the dispersion of the residual transmission process relative to the analysis without scaling. Their policy weights are omitted above, and the proportion of paired runs with lower dispersion than baseline is given for reference.

| | effect strength | proportion better |
|---|---|---|
| Algeria | 0.423 | 0.456 |
| Angola | 0.09 | 0.438 |
| Australia | 0.136 | 0.403 |
| Bahrain | 0.263 | 0.475 |
| Bangladesh | 0.097 | 0.457 |
| Congo | 0.158 | 0.45 |
| Côte d'Ivoire | 0.271 | 0.456 |
| Egypt | 0.132 | 0.443 |
| Gabon | 0.22 | 0.484 |
| Haiti | 0.162 | 0.43 |
| Indonesia | 0.157 | 0.482 |
| Iraq | 0.16 | 0.472 |
| Jamaica | 0.315 | 0.496 |
| Liberia | 0.295 | 0.467 |
| Libya | 0.084 | 0.484 |
| Morocco | 0.191 | 0.475 |
| New Zealand | 0.203 | 0.49 |
| Nigeria | 0.272 | 0.48 |
| Oman | 0.117 | 0.454 |
| Palestine, State of | 0.222 | 0.478 |
| Paraguay | 0.107 | 0.484 |
| Philippines | 0.125 | 0.471 |
| Saudi Arabia | 0.173 | 0.486 |
| Senegal | 0.249 | 0.453 |
| Somalia | 0.356 | 0.45 |
| Togo | 0.124 | 0.45 |
| Ukraine | 0.062 | 0.463 |
| Uruguay | 0.254 | 0.378 |
| Venezuela, Bolivarian Republic of | 0.056 | 0.468 |
| Yemen | 0.325 | 0.454 |
| Zambia | 0.383 | 0.472 |
